# Increased brain volume from higher cereal and lower coffee intake: Shared genetic determinants and impacts on cognition and metabolism

**DOI:** 10.1101/2020.10.11.20210781

**Authors:** Jujiao Kang, Tianye Jia, Zeyu Jiao, Chun Shen, Chao Xie, Wei Cheng, Barbara J Sahakian, David Waxman, Jianfeng Feng

**Affiliations:** Shanghai Center for Mathematical Sciences, Fudan University, Shanghai, China; Institute of Science and Technology for Brain-Inspired Intelligence, Fudan University, Shanghai, China; Key Laboratory of Computational Neuroscience and Brain-Inspired Intelligence (Fudan University), Ministry of Education, China; Centre for Population Neuroscience and Precision Medicine (PONS), Institute of Psychiatry, Psychology & Neuroscience, SGDP Centre, King’s College London, United Kingdom, SE5 8AF; Department of Psychiatry, University of Cambridge School of Clinical Medicine, Cambridge, United Kingdom; Department of the Behavioural and Clinical Neuroscience Institute, University of Cambridge, Cambridge, United Kingdom; Department of Computer Science, University of Warwick, Coventry, United Kingdom; School of Mathematical Sciences and Centre for Computational Systems Biology, Fudan University, Shanghai, China

**Keywords:** Mendel randomization, CPLX3 gene, genetic correlation, pattern correlation, gene expression in the brain

## Abstract

It is unclear how different diets may affect human brain development and if genetic and environmental factors play a part. We investigated diet effects in the UK Biobank data from 18,879 healthy adults and discovered anti-correlated brain-wide grey matter volume (GMV)-association patterns between coffee and cereal intake, coincidence with their anti-correlated genetic constructs. The Mendelian randomisation approach further indicated a causal effect of higher coffee intake on reduced total GMV, which is likely through regulating the expression of genes responsible for synaptic development in the brain. The identified genetic factors may further affect people’s lifestyle habits and body/blood fat levels through the mediation of cereal/coffee intake, and the brain-wide expression pattern of gene CPLX3, a dedicated marker of subplate neurons that regulate cortical development and plasticity, may underlie the shared GMV-association patterns among the coffee/cereal intake and cognitive functions. All the main findings were successfully replicated in the newly-released independent UK Biobank data from 16,412 healthy adults. Our findings thus revealed that high-cereal and low-coffee diets shared similar brain and genetic constructs, leading to long-term beneficial associations regarding cognitive, BMI and other metabolic measures. This study has important implications for public health, especially during the pandemic, given the poorer outcomes of COVID-19 patients with greater BMIs.

**Significance statement:** We investigated diet effects on the brain structure and its genetic constructs using the UK Biobank data and discovered a causal effect of higher coffee intake on reduced total grey matter volume (GMV) and replicable anti-correlated brain-wide association GMV patterns between cereal and coffee intake. Further, the high-cereal and low-coffee diets shared similar brain and genetic constructs, leading to long-term beneficial associations regarding cognitive, BMI, and other metabolic indicators. Our study has important implications for public health, especially during the pandemic, given the poorer outcomes of COVID-19 patients with greater BMIs.

## Introduction

Increases in human brain volume, due to growth, begin at an early stage of embryonic development and continue until late adolescence(1). After this, the brain experiences a persistent but slow decrease in size throughout adulthood(2). Generally, development is tissue-specific but systematically organised across the brain(2, 3) and may be susceptible to both genetic and environmental influences(2-6), as well as their interactions, e.g. through epigenetic modifications(7). Diet is a common environmental factor that can influence the trajectory of brain size. For example, a lack of nutrients over an extended period of time causes both structural and functional damage to the brain(8), and improved diet quality is associated with larger brain volumes(9). Furthermore, evidence suggested that ingested substances (both food and drink) in well-fed and healthy adults may also cause changes in brain size. For example, in a small-scale study, an increase in the size of the hippocampus was inferred to have occurred as an effect of both low and high coffee consumption(10).

While there are extensive studies of the degree to which different diets affect the body(11-14), there is an absence of systematic investigation into how different diets may affect the human brain in both the short and long term. Thus, it is not known if impacts of different diets on brain structures follow similar patterns, or whether different brain regions exhibit differential sensitivity to diet and other environmental factors. In addition, there is a lack of knowledge about whether genetic factors play any role in the sensitivity of the brain to environmental factors. In the present research, consisting of an original study of 18,879 individuals and a replication study of 16,412 adults, we provide a detailed analysis of brain-size changes that occur in healthy adults due to the ingestion of different common foods and drinks. We investigated whether these influences from diets were systematically organised across the brain, whether these dietary influences have underlying genetic factors, and whether these genetic factors have further implications in people’s daily activities, metabolism and cognitive functions.

## Results

### Association between grey matter volume and diets

We first investigated the relationship between grey matter volume (GMV) and 17 different diet phenotypes, which were both measured at the second visit (i.e. at follow-up) of participants to a research center(15). We found that the *total grey matter volume of the brain* (TGMV) is affected by diet. Some dietary items were negatively correlated with the TGMV, thus decreasing consumption of these items had the tendency to increase the TGMV, while other items were positively correlated, and had the opposite tendency on the TGMV. With a statistically significant correlation (P<0.05 Bonferroni corrected), intake of coffee, water, processed meat, beef, lamb/mutton and pork were found to be negatively correlated with TGMV, while intake of cereal and dried fruit were positively correlated with TGMV (see Table S2). We note that predated measurements (i.e., baseline measurements) of cereal and coffee intake were also related to the follow-up values of TGMV (Table S3), and these remained significant even after controlling for the corresponding follow-up intakes (Table S4). This indicates a persistent, rather than a short-term connection between diet and GMV. We then validated the above results in the newly-released additional 16412 UK Biobank individuals and confirmed the persistent positive associations between TGMV and cereal intake and negative associations between TGMV and coffee intake (Table S21 A).

Correlations were further investigated between 17 diet phenotypes and the volumes of 166 brain regions defined by the automated anatomical labelling 3 (AAL3) atlas(16). A total of 454 statistically significant correlations (Bonferroni correction: P<0.05/166/17) were found, again mainly between GMV and intake of cereal, coffee, water, dried fruit, processed meat, beef, pork and lamb/mutton (Fig.1.A and Table S2). It is interesting to note that the GMV-association pattern of cereal intake highly resembles, although in the opposite direction, the GMV-association pattern of coffee intake (pattern correlation across the whole brain: r=-0.6177, P < 1E-04 based on 10000-permutation; Fig.1.B), the same negative pattern correlation could also be observed in the newly-released UKB data (r=-0.45, P_one-tailed_ = 0.0116 based on 10000 permutation).

**Figure 1.**
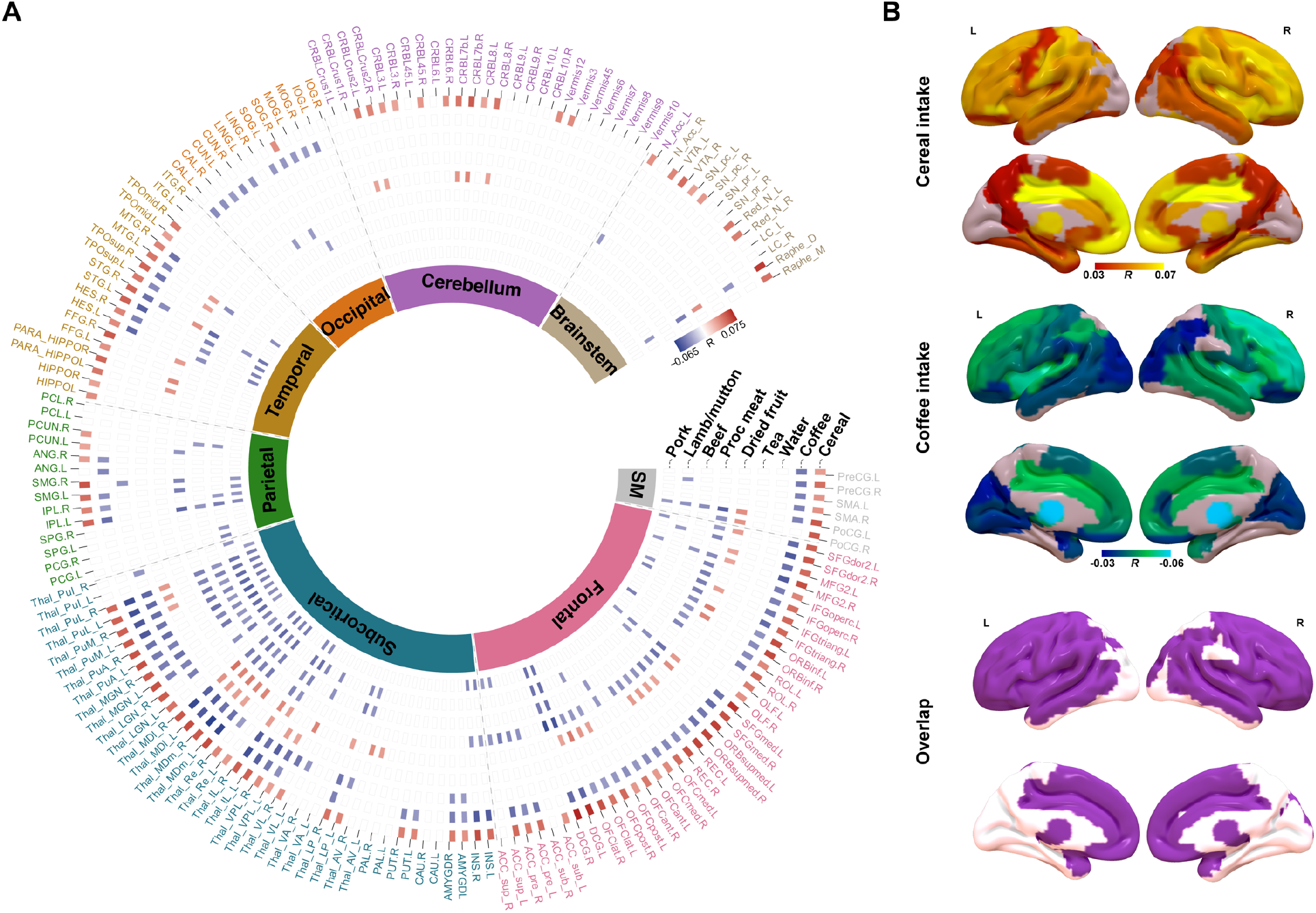
Correlations between grey matter volume (GMV) and different daily diets. (A) Circular heatmap of correlations between GMVs of 166 brain regions from AAL3 (the outer layer) and different diets (along radius). As indicated by the colour bar, positive correlations were highlighted in red while negative correlations were highlighted in blue. The inner layer indicates the lobes that brain regions belong to. (B) Brain regions with significant correlations between their GMVs and the intake of cereal (upper) and coffee (middle), as well as the overlapped significant regions (bottom). SM: Sensorimotor.

### Genome-wide association studies for the intake of cereal and coffee

We conducted genome-wide association studies (GWAS) for the intake of both cereal (n=335696) and coffee (n=335068) at baseline and identified 21 and 45 independent lead genome-wide significant variants with P<5E-08 (i.e., the lead SNPs, see methods for details) respectively (Fig.2, Table S7&S8 and fig.S1). A linkage disequilibrium (LD) score regression(17) analysis indicates that both findings were free from systematically inflated false-positive rates, e.g., due to population stratification, with intercepts of 1.013 (cereal intake) and 1.005 (coffee intake), and the corresponding SNP-based heritabilities were estimated as 0.0652 (se=0.0038) and 0.0618 (se=0.007) respectively. Furthermore, we observed a significant negative genetic correlation(18) between intake of cereal and coffee (r_g_=-0.233, se=0.052, z-score=-4.49, P=7.1E-06), i.e., the alleles associated with higher cereal intake were likely to be in association with reduced coffee intake, which is in line with the above GWAS findings, where the three shared lead SNPs, i.e. rs2504706, rs4410790 and rs2472297, were found in associations with both cereal and coffee intake, again in opposite directions (Table S9).

**Figure 2.**
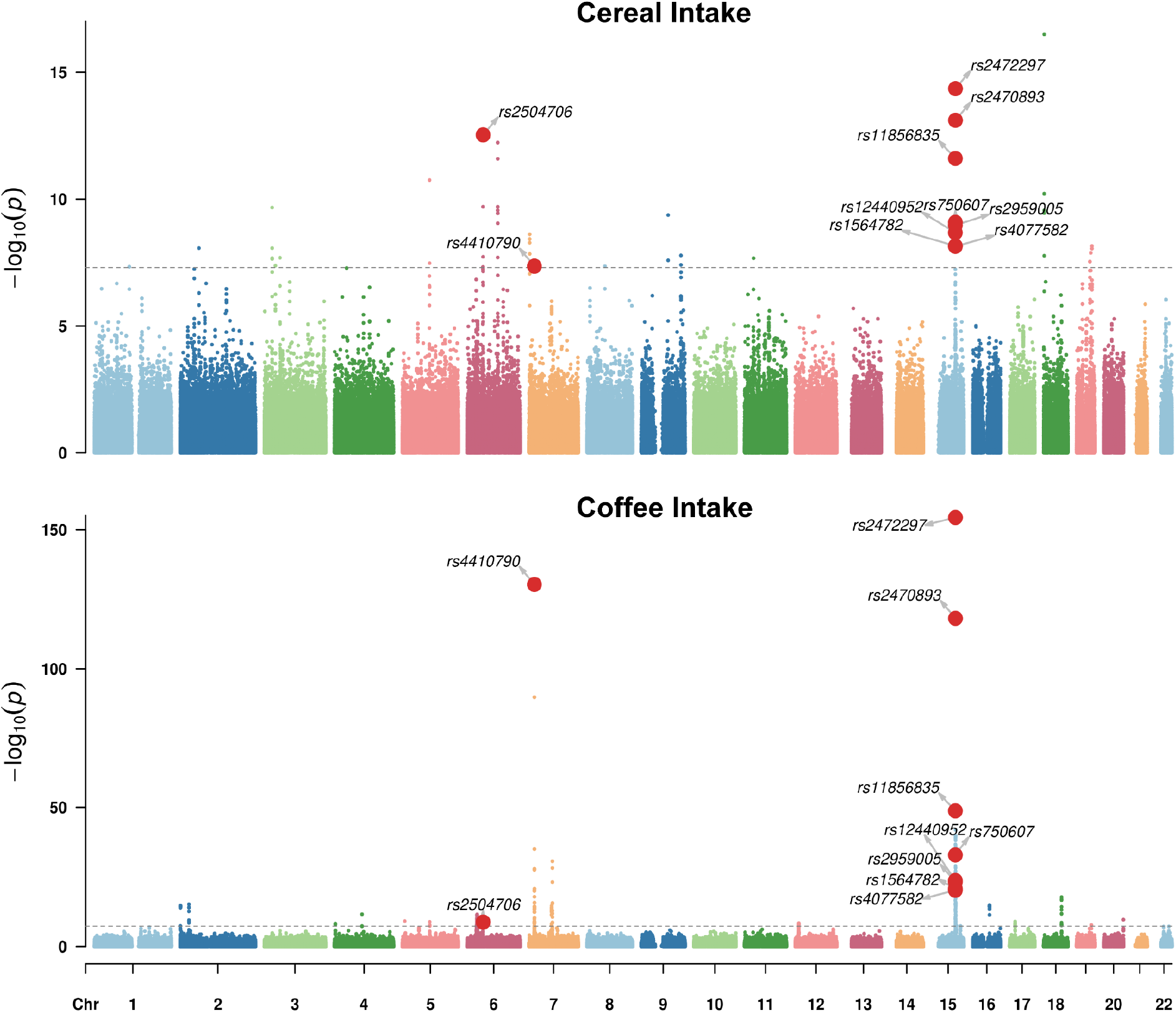
Manhattan plots of the genome-wide association results for the intake of cereal (upper) and coffee (bottom). The gray line indicates the genome-wide significance level (i.e. P-value=5E-08). Variants with significant associations with both cereal and coffee intake were highlighted with red dots.

The minor C-allele at rs2504706 was associated with a higher intake of cereal (beta=0.058, 95% CIs=[0.042, 0.073], T_df=334441_=7.30, P=2.94E-13) and a lower intake of coffee (beta=-0.034, 95% CIs=[-0.045, -0.023], T_df=*333816*_=-6.03, P=1.67E-09). The minor C-allele at rs4410790 was associated with lower intake of cereal (beta=-0.038, 95% CIs=[-0.052, -0.024], T_df=*334331*_=-5.47, P=4.40E-08) and higher intake of coffee (beta=0.120, 95% CIs=[0.110, 0.130], T_df=*333705*_=24.37, P=4.71E-131). The minor T-allele, at rs2472297, was associated with lower intake of cereal (beta=-0.059, 95% CIs=[-0.074, -0.044], T_df=*334951*_=-7.84, P=4.38E-15) and higher intake of coffee (beta=0.142, 95% CIs=[0.131, 0.152], T_df=*334321*_=26.55, P=4.20E-155). While rs4410790 and rs2472297 have both been previously associated with coffee/caffeine consumption(19-21), caffeine metabolism(22), and alcohol consumption(23), this is the first study to identify an association with cereal intake. It is notable that SNPs rs4410790 (the C-allele) and rs2472297 (the T-allele) were also strongly associated with higher intake of tea (beta=0.111, 95% CIs=[0.098, 0.123], T_df=*332509*_=17.04, P=4.58E-65 for rs4410790; beta=0.148, 95% CIs=[0.134, 0.162], T_df=*333124*_=21.03, P=3.82E-98 for rs2472297, respectively) and lower intake of water (beta=-0.075, 95% CIs=[-0.085, -0.065], T_df=*331879*_=-14.64, P=1.65E-48; beta=-0.086, 95% CIs=[-0.097, -0.076], T_df=*332497*_=-15.62, P=5.53E-55, respectively) (Fig.3.A and Table S10), although both intakes were not observed with significant long term impacts on the TGMV (Table S4). This result is remarkable because there is a median to large anti-correlation between the intake of coffee and tea (r=-0.359, beta=-0.472, 95% CIs= [-0.477, -0.468], T_df=*332711*_=-221.65, P<1.0E-256), which is likely due to the seesaw effect given the limited amount of beverages one may consume each day. Thus, individuals with both SNPs (i.e., C-allele of rs4410790 and T-allele of rs2472297) might generally prefer flavoured beverages to the water.

**Figure 3.**
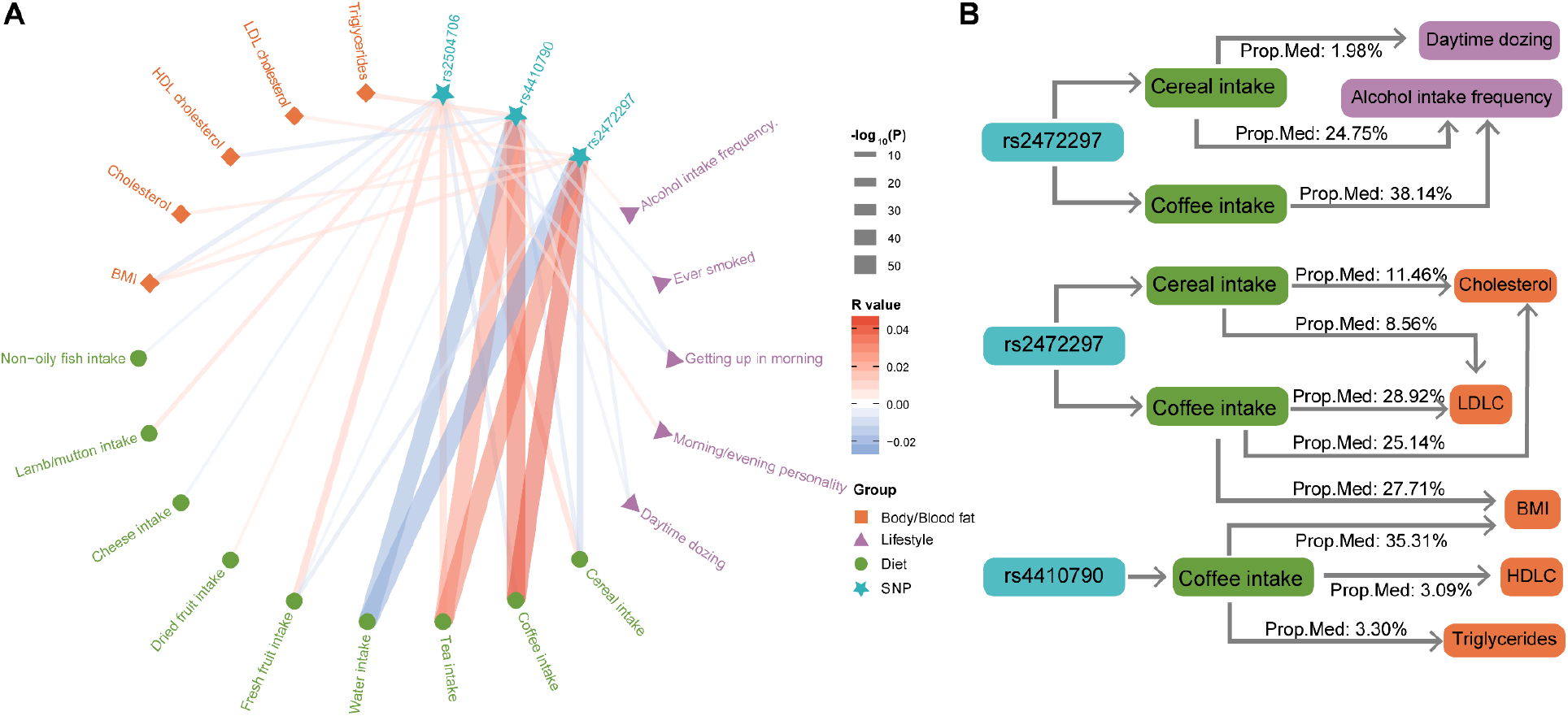
Relationships between the lead SNPs and diets, lifestyle, and body/blood fat. (A) Associations between the three lead SNPs (of both cereal and coffee intake) and other diets, lifestyle, and body/blood fat levels. The colour of each line represents the correlation coefficient (positive in red and negative in blue), and the thickness of each line represents the -log_10_ P-value (capped at 50) of the corresponding correlation. (B) Proposed mediation models of genetic variants, body/blood fat levels, lifestyles, and the intake of cereal and coffee. Prop. Med: proportion of mediation. BMI: Body mass index, HDL: High-density lipoprotein, LDL: Low-density lipoprotein.

### Causal effects of cereal/coffee intake on TGMV based on mendelian randomisation

Above, we have established and verified the associations of cereal/coffee intake with TGMV (Table S2 and S3). Here, we further implemented a modified Mendelian randomisation approach to investigate potential causal relationship by introducing non-pleiotropic polygenic risk scores (i.e. the valid-PRS of cereal/coffee intake or TGMV) as instrument variables of randomised experiments, which only affect the explanatory variables, but not the outcome variables if not through the explanatory variables (see Methods for more details)(24).

Using the availability of neuroimaging data as a random stratification of UK Biobank data, we reconducted GWAS of cereal/coffee intake in the large discovery sample without neuroimaging information (N=307750), as well as conducting a new GWAS of TGMV in the first batch of neuroimaging data (N=14764). In the independent second batch of neuroimaging data (N=12783), we then calculated the PRSs of cereal/coffee intake and TGMV (using PRSice v1.25(25) with the P-value threshold of 0.05), which all showed consistent associations with the corresponding phenotypes (PRS_cereal_ with cereal intake: R=0.081, P_one-tailed_ = 2.13E-20; PRS_coffee_ with coffee intake: R=0.075, P_one-tailed_ = 1.31E-17; PRS_TGMV_ with TGMV: R=0.097, P_one-tailed_ = 4.39E-28). Interestingly, while PRS_cereal_ and PRS_coffee_ were found in significant associations with TGMV (R=0.021, P_one-tailed_ = 8.20E-3 and R=-0.026, P_one-tailed_ = 1.48E-3, respectively), PRS_TGMV_ was not associated with the intake of either cereal (R=0.007, P_one-tailed_ = 0.215) or coffee (R=-0.006, P_one-tailed_ = 0.244), indicating that genetically TGMV might be secondary to the intake of cereal/coffee. Further, we removed potential pleiotropic SNPs from the above PRSs with a stepwise approach (i.e. stepwise removing cross-associated SNPs based on their P-values with the confounding phenotypes from 0.05 to 0.50 with a step of 0.05), and found these valid-PRSs maintained strong correlations with the corresponding main phenotypes (valid-PRS_cereal_ with cereal intake: R>0.057, P_one-tailed_ < 6.16E-11; valid-PRS_coffee_ with coffee intake: R>0.059, P_one-tailed_ < 1.07E-11; valid-PRS_TGMV_ with TGMV: R>0.059, P_one-tailed_ < 1.26E-11; Table S22.A&B&C), hence as qualified instrumental variables of random experiments. However, while valid-PRS_coffee_ maintained consistently significant negative associations with TGMV across all 10 thresholds (R < -0.015, P_one-tailed_ <0.046; Table S22.D), valid-PRS_cereal_ lost the significance with TGMV in 7 out of 10 thresholds although maintaining consistent positive associations throughout (R > 0.012, P_one-tailed_ <0.095; Table S22.D). Nevertheless, the associations of valid-PRS_TGMV_ with the intake of cereal (−0.001<R<0.003, P_one-tailed_>0.370; Table S22.E) and coffee (−0.006<R<-0.002, P_one-tailed_>0.266; Table S22.F) further diminished. Thus, following the argument of Mendelian randomisation, we could reach a conclusion that randomly increased coffee intake will cause reduced TGMV, but not the other way around.

### Transcriptome-wide whole brain pattern associations with the GMV-association patterns of the coffee intake

To explore the potential neurobiological mechanisms underlying the causal effects of coffee intake on the TGMV, we next investigate if coffee intake may regulate gene expression pertaining to synaptic development. Particularly, we evaluated the pattern correlation between the GMV-association pattern of the coffee intake and the spatial gene expression pattern from the Allen Institute for Brain Science (AIBS) (26), and identified 1737 validated significant genes (P_FDR,perm_ <0.05 in the discovery sample and P_perm_ < 0.05 in the validation sample; see materials and methods for details). Out of these 1737 genes, 15 were overlapped with the 199 candidate genes identified through the coffee intake GWAS (using FUMA(27) software; see materials and methods for details) and were mainly enriched in the perception of bitter taste (P_FDR_ < 0.001) based on a follow-up Gene Ontology (GO) analysis (28), which might highlight a potential complicated gene-environmental interaction that the same gene may influence its own expression through regulating the coffee intake. Further, the remaining non-overlapped 1722 genes were highly enriched in biological pathways related to synapse organizations (best P_FDR_ = 4.18E-4, Table S23B; see materials and methods for details), thus highlighting that coffee intake could regulate the expression of these synaptic genes in the brain, which may explain the biological mechanism underlying the causal effect of coffee intake on the TGMV.

### Association between genetic variants, diets and lifestyle

As both cereal and coffee intake, as well as their shared lead SNPs, were associated with different lifestyles, such as the frequency of physical activity (R=0.016, P=2.52E-17 for cereal and R=-0.011, P=3.23E-09 for coffee), being a morning/evening person (R=-0.040, P=2.57E-104 for cereal and R=0.032, P=3.05E-69 for coffee) and the frequency of alcohol use (R=-0.101, P<1.0E-256 for cereal and R=0.050, P=5.77E-184 for coffee) (Fig.3A and Table S11 & S12), we then investigated possible mediation roles of diet or/and lifestyles on their associations with SNPs. As no prior assumptions about whether diet or lifestyle should serve as the mediator for their associations with the lead SNPs, we evaluated the most likely mediator, based on the corresponding proportion of mediation (PM) that they are responsible for (see the Supplementary Material for more details). We found the following:

1. Both intake of cereal and coffee were likely to mediate the positive association of the frequency of alcohol intake with the T-allele of rs2472297 (PM=24.75%, P_bootstrap_=5.47E-15 and PM=38.14%, P_bootstrap_=1.37E-82 respectively; Table S13); these were superior to alternative mediation models with the frequency of alcohol intake as the mediator (excess PM>20% with P_bootstrap_<0.002 for both alternative models, Fig.3.B and Table S13; see supplementary materials for detailed analyses);
2. The association between higher T-alleles of rs2472297 and less daytime sleeping was mediated by cereal intake (PM=1.98%, P_bootstrap_=2.82E-6; Fig.3.B and Table S13), which was superior to the alternative mediation model with daytime sleeping as the mediator (excess PM=1.39% with P_bootstrap_=0.018, Table S13; see supplementary materials for detailed analyses);
3. Both difficult in rising and less daytime sleeping were found to mediate the negative association of cereal intake with the C-allele of rs4410790, so did the alternative mediation models with the cereal intake as the mediator. However, neither group of mediation models was superior to the other (Table S13);
4. Interestingly, while individuals with rs2504706 (the C-allele) were more likely to be an ‘evening person’ and experience difficulties in rising, both lifestyle traits did not mediate the associations of the SNP with higher cereal intake or lower coffee intake (nor did the alternative mediation models), which was mainly due to nonconcordant correlations, e.g., a positive correlation was observed between ease in rising and higher cereal intake while their associations with SNP rs2504706 would indicate a negative correlation instead (Fig.3.A and Table S9, S11 & S12).

### Association between genetic variants, diets and metabolic measures

In addition to lifestyle, both cereal and coffee intake, as well as their shared lead SNPs, were also associated with blood (for example with total cholesterol, R=-0.066, P<1.0E-256 for cereal and R=0.045, P=1.89E-139 for coffee) and body fat levels (for example with the body mass index (BMI), R=-0.076, P<1.0E-256 for cereal and R=0.053, P=3.84E-206 for coffee) (Table S14, Table S15). Therefore, we further explored possible mediator roles of fat levels and the intake of cereal and coffee. We found the following:

1. Associations between rs4410790 (C-allele) and: an increased body mass index (BMI), triglycerides and decreased HDL cholesterol, were mediated by increased coffee intake (PM=35.31%, P_bootstrap_=1.41E-81, PM=3.30%, P_bootstrap_=1.04E-5, and PM=3.09%, P_bootstrap_=2.28E-4, respectively), which were superior to the alternative mediation models with corresponding fat levels as mediators (excess PMs=34.48%, 3.10% and 2.95%, respectively; all corresponding P_bootstrap_<0.002) (Table S16);
2. Associations between rs2472297 (T-allele) and higher body mass index (BMI), total cholesterol, and LDL cholesterol, were mediated by higher coffee intake (PM=27.71%, P_bootstrap_=6.72E-83, PM=25.14%, P_bootstrap_=6.82E-68 and PM=28.92%, P_bootstrap_=4.65E-75, respectively), as well as by lower cereal intake, to a lesser extent (PM=11.46% for total cholesterol, P_bootstrap_=6.51E-14 and PM=8.56% for LDL cholesterol, P_bootstrap_=1.80E-13). The above models were superior to alternative mediation models with corresponding fat levels as mediators (for the coffee intake: excess PMs=26.66%, 24.38% and 28.13%, respectively, with all corresponding P_bootstrap_<0.002; for the cereal intake: excess PMs=7.54% and 5.83%, respectively, with all corresponding P_bootstrap_<0.05) (Table S16).

Related to the current COVID-19 pandemic, using the UK Biobank data we found that individuals who tested positive of COVID-19 (n=639, inpatients only) had higher BMIs (Cohen’s D=0.27, t=6.72, P=1.86E-11) and lower cereal intake (Cohen’s D=-0.09, t=-2.36, P=0.019) than the rest population (n=314982, either tested negative or not tested). This further highlights the importance of our finding for public health that cereal intake is associated with lower BMIs.

### Associations between the GMV-association patterns of cognitive functions and the GMV-association patterns of the intake of cereal and coffee

To further characterise the negatively correlated brain-wide GMV-association patterns for cereal and coffee intakes, we further investigated if such similarities have any implications for cognitive functions, and we found that brain-wide GMV-association patterns of most cognitive functions were significantly correlated with those of both cereal and coffee intake, although in opposite directions, at both baseline and follow-up (GMV were measured at follow-up only). In particular, performance in tasks of matrix pattern completion, symbol digit substitution, and numeric and alphabet-numeric trail making showed similar brain-wide GMV-association patterns with both cereal (in positive correlation) and coffee (in negative correlation) intake at both baseline and follow-up (|R|_min_=0.5945, all P_FDR_<0.05; Fig.4 and Table S17&S18), while the fluid intelligence score only showed a similar brain-wide GMV association pattern with the cereal intake (at both baseline and follow-up; R_min_=0.62, all P_FDR_<0.05; Table S17&S18). The same findings could be replicated in the recently-released 16412 additional individuals (|R|_min_ =0.40, P_one-tailed_ <0.01) (Table S21.B). In line with the above findings, higher risk of Alzheimer’s disease (estimated as the proxy-AD (29)), characterised by reduced cognitive functions, was associated with reduced cereal intake (R=-0.009, P=3.42E-6), as well as increased coffee intake to a much lesser extent (R=0.004, P=0.024), in contrast to previous findings of either protective(30) or non-significant(31) effect of high coffee intake on Alzheimer’s disease.

**Figure 4.**
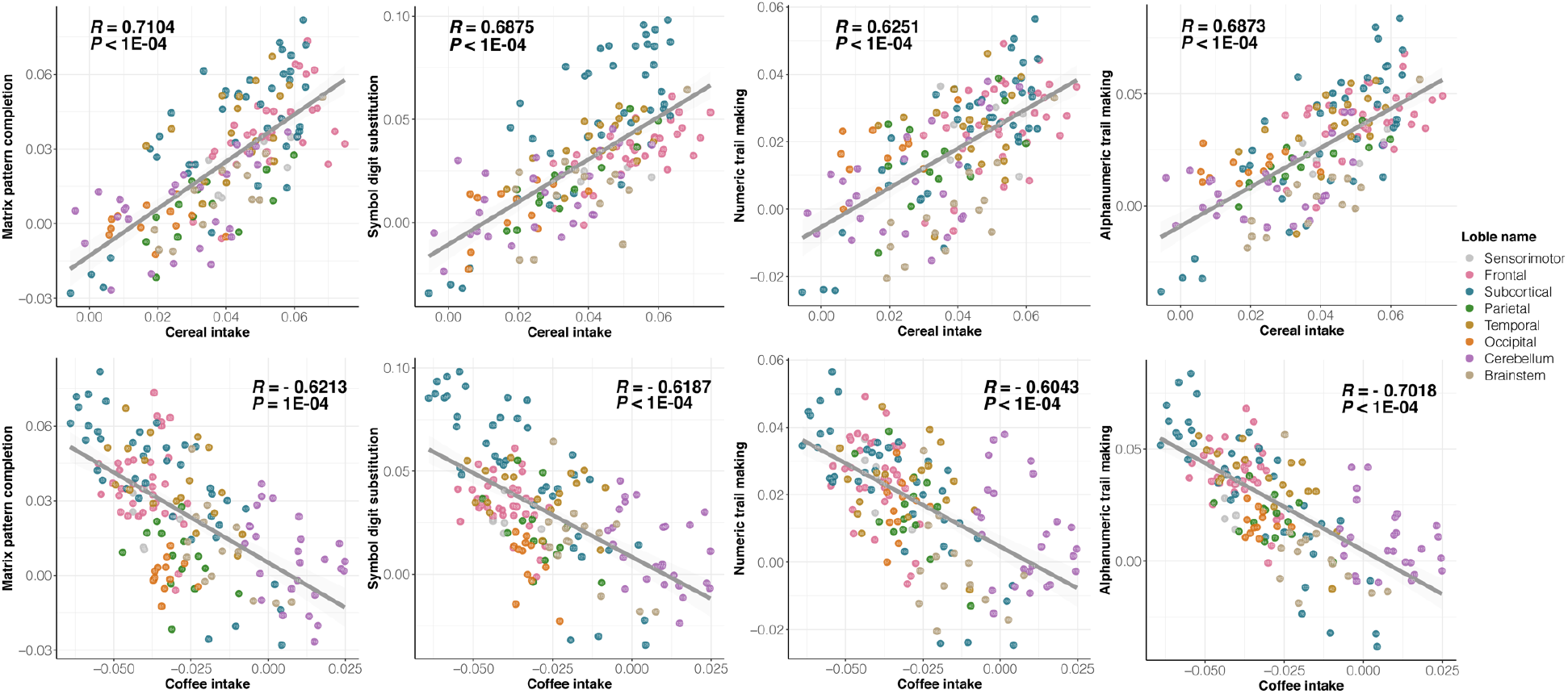
Scatter plots of brain-wide GMV-association patterns of cognitive functions and the intake of cereal (upper) and coffee (bottom). Each dot represents one of 166 AAL3 brain regions, where the colours indicate at which lobes the brain regions were located.

### Associations between the GMV-association patterns of the cereal/coffee intake and the gene-expression patterns of eQTL genes

Finally, we investigated if the identified putative genetic variants associated with both cereal and coffee intake may also contribute to observed similarities of brain-wide GMV association patterns between diet and cognitive function. We first performed eQTL mapping of the 3 shared lead SNPs using FUMA(27) software and identified 31 candidate protein-coding genes (Table S19) that also have brain-wise gene expression information from the Allen Institute for Brain Science (AIBS)(26). After also mapping to the AAL3 atlas, the brain-wide expression pattern for each candidate gene (i.e. the mean expression level across all AIBS individuals for each brain region) was then correlated with the brain-wide GMV association patterns for cereal and coffee intakes. While multiple candidate genes had their brain-wide expression pattern in significant correlation with brain-wide GMV associations patterns for the coffee intake (Table S19), only gene CPLX3 showed significant ‘gene-expression vs GMV-association’ pattern similarity with both intakes of cereal (R=0.47, P_perm_=2.9E-3, P_FDR-corrected_=0.033) and coffee (R=-0.44, P_perm_=7.2E-3, P_FDR-corrected_=0.046). Both findings could be reproduced in the recently-released independent data (R=0.40, P_one-tailed_ = 0.011 for cereal intake; R=-0.36, P_one-tailed_ = 0.020 for coffee intake; 10000 permutation; Table S21.C). It is of particular interest that the gene-expression of CPLX3 (a known prominent marker specific for subplate neurons in the brain that regulate cortical development and plasticity across the brain (32-34)) also showed significant pattern correlations with almost all cognitive functions (i.e., R=0.42 for fluid intelligence, R=0.49 for numerical memory, R=0.44 for prospective memory, R=0.46 for matrix pattern completion, R=0.39 for symbol digit substitution, and R=0.44/R=0.55 for both trail making tasks; all corresponding P_FDR-corrected_<0.05; Table S20). The above findings were again fully reproducible in the recently-released data (R_min_ = 0.39, P_one-tailed_ < 0.011) (Table S21.D).

## Discussion

In the large-scale imaging/genetics analysis presented in this work, we have: (i) gained insights into long-term associations between brain-wide GMV and diets, especially the anti-correlated impacts from cereal and coffee intake, (ii) identified shared genetic constructs for both higher cereal and lower coffee intake and confirmed the causal effect of coffee intake on the brain volume, which is likely through regulating the expression of genes responsible for synaptic development, (iii) explored the complex relationship among cereal/coffee intake, their genetics constructs, lifestyle, and body/blood fat level, (iv) revealed shared brain-wide GMV-association patterns between cognitive function and the intake of cereal and coffee and further showed that such similarity might be underlaid by the brain-wide expression patterns of gene CPLX3, a shared genetic determinant identified for the intake of both cereal and coffee. These novel findings hence suggest the existence of a brain-wide systematic organisation of GMV that is susceptible to both genetic and environmental influences, as well as their complex interaction, which may have further impacts on people’s lifestyles, cognitive functions, and metabolic measures (e.g. BMI and blood cholesterol level).

Two lead SNPs shared by the intake of coffee and cereal, i.e. rs4410790 and rs2472297, have been previously associated with coffee/caffeine consumption(19-21), caffeine metabolism(22), and alcohol consumption(23). However, this is the first study to identify their associations with cereal intake. Moreover, while CPLX3, within the LD complexity around the lead SNP rs2472297, has previously been proposed as a candidate gene for both coffee consumption(35) and blood pressure(36), it is the first time that this gene links to multiple cognitive functions, such as intelligence, and cereal intake. Remarkably, the expression of CPLX3 is a highly specific marker of subplate neurons (32) that regulate cortical development and neuronal plasticity across the brain (33, 34). Specifically, while most subplate neurons were short-lived during the development of the brain, previous studies have shown that the Cplx3-positive subplate neurons could survive into adulthood in mice (32). Therefore, our findings were not only congruent with the role of subplate neurons in the cortical development, but with further implications that these CPLX3-positive subplate neurons might mark the dynamic system of GMV in the brain that is susceptible to environmental factors. Such a hypothesis could be supported by previous findings that Cplx3 protein’s regulation of exocytosis in mice retinal neurons could be altered by both light and electrical stimuli (37, 38).

Overall, since high cereal diets, but low coffee diets, have long-term beneficial associations regarding the brain, cognition, BMI and other metabolic measures, this study has significant implications for public health. Our findings highlight the importance of a ‘cereal’ breakfast across the life span, but perhaps especially for children and adolescents whose brains are still in development and for reducing the risk of Alzheimer’s disease and poor outcomes due to high BMIs in patients with COVID-19 (39, 40).

## Materials and Methods

### Study participants

Study samples were from the UK Biobank study, a prospective epidemiological study that involves over 500,000 individuals in 22 centres across the UK(15). Between 2006 and 2010, participants were recruited to collect a range of questionnaires about detailed phenotypic information including diet, lifestyle, anthropometric and cognitive function assessments, biological samples, including blood and medical records obtained from the NHS registries. Since 2014, a subsample of the original population has been invited back to collect magnetic resonance imaging of body and brain, and questionnaires about diet, lifestyle, and cognitive function assessments.

In the current study, we used data collected at both recruitment and MRI scan. The original sample comprised 488289 individuals (56.54 ±8.09 years; 54.21% women). We included 431039 white British individuals and then excluded 810 individuals who were diagnosed with Alzheimer’s or dementia defined by codes G30/F00 in the 10th edition of the International Classification of Diseases (ICD-10). Of the 430228 individuals, 336517 individuals had quality-controlled genetic data, and 18879 individuals had available brain MRI data as the discovery sample, and the newly-released (41) 16412 individuals were used as an independent replication. Table S1 summarised relevant demographic information. Behavioural and neuroimaging data collection and protocol are publicly available on (15, 42). All participants provided written informed consent to UK Biobank. The UK Biobank study received ethical approval from the NHS National Research Ethics Service North West (reference number: 16/NW/0274). Data access permission was granted under UKB application 19542 (PI Jianfeng Feng).

### Assessment of the intake of cereal and coffee

Dietary information was obtained from the touchscreen questionnaire at the baseline and the MRI scan appointment. Cereal intake was the number of bowls of cereal the participants consumed per week. The types of cereal included bran cereal, biscuit cereal, oat cereal, muesli, and other types (e.g., cornflakes, Frosties). Coffee intake was the number of cups of coffee the participants drank per day. The types of coffee included decaffeinated coffee, instant coffee, ground coffee, other types of coffee. Detailed information can be found in supplementary materials.

### Assessment of lifestyle

Lifestyle phenotypes included physical activity, sleep, smoking and alcohol and were obtained from the touchscreen questionnaire at the baseline appointment. Physical activities were assessed using MET (Metabolic Equivalent Task) scores derived based on International Physical Activity Questionnaire) of total physical activity (including walking, moderate, and vigorous activity) and usual walking pace. The time spent watching television was also included to reflect physical activity. Sleep data included information for sleep duration, morningness or eveningness type, insomnia symptoms, daytime dozing, getting up in morning, and nap during day. Smoking status included smoking history and the number of cigarettes currently smoked daily. Alcohol intake was examined using frequency and amounts of alcohol drinking. Detailed description can be found in supplementary materials.

### Assessment of cognitive functions

Cognitive function performances were examined at the baseline and the MRI scan appointment. The cognitive tests included fluid intelligence score, reaction time, numeric memory, pairs matching, prospective memory, matrix pattern completion, symbol digit substitution and trail making. Detailed descriptions of procedures can be found in supplementary materials.

### Assessment of body size and blood cholesterol

Body mass index was calculated from the participant’s measured weight (kg)/height (m2). Cholesterol, high-density lipoprotein (HDL) cholesterol, low-density lipoprotein (LDL) cholesterol, and triglycerides were measured in the blood sample collected at recruitment.

### Assessment of the Alzheimer’s disease risk

We used a proxy phenotype for Alzheimer’s disease (AD) case-control status derived from the genetic risk index for AD based on parents’ diagnoses as suggested in a previous study(29). The proxy phenotype ranged approximately from 0 to 2, with values near zero when both parents were unaffected (lower for older parents and possible values below zero if both parents were over age 100) and values of two when both parents were affected.

### COVID-19 test

COVID-19 test results data are linked to UK Biobank by Public Health England (PHE). Data were available for the period 16th March 2020 to 3rd August 2020. Data provided included specimen origin (hospital inpatient indicating severe COVID-19 vs. other settings). Detailed information is available on the website (http://biobank.ndph.ox.ac.uk/ukb/exinfo.cgi?src=COVID19_tests). To focus on the COVID-19, we excluded individuals passed away except for those who had positive test results. There were 13145 unique test results available, of which 1649 (12.54%) were positive; 10098 (76.82%) tests were conducted on inpatients; 1069 (639 had available data on BMI and diet) were inpatients and positive.

### Structural MRI preprocessing

Detailed structural MRI data collection and acquisition procedures can be found in supplementary materials. All UK Biobank structural MRI data were preprocessed in the Statistical Parametric Mapping package(43) (SPM12) using the VBM8 toolbox with default settings, including the usage of high-dimensional spatial normalisation with an already integrated Dartel template in Montreal Neurological Institute (MNI) space. All images were subjected to nonlinear modulations and corrected for each individual head size. Images were then smoothed with a 6 mm full-width at half-maximum Gaussian kernel with the resulting voxel size 1.5mm3. The estimated total intracranial volume (TIV) covariate, were calculated as the summation of the grey matter, white matter, and CSF volumes in native space. The automated anatomical labelling 3 (AAL3) atlas(16), which partitioned the brain into 166 regions of interest, was employed to obtain the total brain grey matter volume and region-wise grey matter volume. 18879 discovery and 16412 replication T1 images were successfully preprocessed, and the grey matter volumes of the AAL3 atlas of the discovery sample were extracted. The majority of discovery samples were assessed in the Cheadle MRI site (84.49%) and the rest in the Newcastle site (15.51%). In comparison, 37.17% of replication samples were assessed in the Cheadle MRI site, 37.87% were tested in the Newcastle site, and the remaining 24.96% were in the Reading site.

### Genetic data quality control

Detailed genotyping and quality control procedures of the UK Biobank can be found in supplementary materials or http://biobank.ctsu.ox.ac.uk/. In this study, we performed stringent QC standards by PLINK 1.90(44). Single-nucleotide polymorphisms (SNPs) with call rates <95%, minor allele frequency <0.1%, deviation from the Hardy–Weinberg equilibrium with p<1E-10 were excluded from the analysis. In addition, we selected subjects that were estimated to have recent British ancestry and have no more than ten putative third-degree relatives in the kinship table using the sample quality control information provided by UKB. For more details, we refer to the official document for genetic data of the UKB (http://www.ukbiobank.ac.uk/scientists-3/genetic-data/). After the quality control procedures, we obtained a total of 616,339 SNPs and 336517 participants.

### Preprocessing of the Allen Human Brain Atlas data

We followed the AHBA preprocessing pipeline suggested by Arnatkevičiūtė et al.(45) and using the same pipeline as Shen et al.(46), including probe-to-gene re-annotation, data filtering, probe selection. In the next step, we separated the samples into the areas based on their MNI coordinates, using the automated anatomical labelling 3 (AAL3) atlas(16) and excluding the samples located outside of the grey matter defined by this atlas. To control for the inter-individual differences, we conducted two within-donor normalisations. The expression data were first normalised within-sample and across-gene and then normalised across samples. One gene failed the normalisation and therefore was deleted, resulting in 15,408 genes. We used the mean expression of samples located in the brain region and the mean expression in the brain region of all subjects as the gene expression in each brain region defined by AAL3 atlas(16).

### Association analysis

We conducted linear regression analysis to test the pairwise associations between the diet phenotypes and the total and regional grey matter volume (GMV), respectively. The covariate variables included were age at imaging scan, sex, imaging sites (dummy variable), and total intracranial volume (TIV).

To further understand the biological insights of the shared variants of cereal intake and coffee intake, we performed linear regression analysis to examine the pairwise associations between the independent lead SNPs and other diets, lifestyle, and body/blood fat covarying age, sex, the top 40 genetic principal components. We also performed linear regression analysis to examine the pairwise associations between the cereal/coffee intakes, other diets, lifestyle, and cholesterol covarying age, sex.

### Genome-wide association analysis and annotation of significant variants

We performed genome-wide association analysis adjusting for age, sex, and the top 40 ancestry principal components using PLINK 1.90(44) to assess the association between phenotype and genotype on cereal intake and coffee intake separately. After association analysis, we employed the FUMA(27) online platform (version 1.3.6, http://fuma.ctglab.nl/) to define genomic risk loci. The GWAS summary statistics was submitted as input. FUMA identifies significant variants with P value less than 5E-8 that were largely independent of each other (r^2^ < 0.6). Based on the clumping of the independent significant variants (r^2^ < 0.1), independent lead variants were obtained.

The independent significant SNPs were mapped to genes based on positional, eQTL and chromatin interaction mapping using FUMA(27). Positional mapping map SNPs to genes based on physical distances (within a 10-kb window). The eQTL mapping map SNPs to genes based on eQTL associations that SNP was significant (false discovery rate (FDR)≤ 0.05) expression level of gene using information on eQTLs of 44 tissue type in GTEX(47) v8 and BRAINEAC(48). The eQTL mapping was based on cis-eQTLs and could map SNPs to genes up to 1Mb apart. The chromatin interaction mapping map SNPs to genes using Hi-C data of 14 tissue types from GSE87112(49).

Shared lead SNPs of cereal and coffee were mapped to genes based on cis-eQTL (p value≤ 0.05) in the brain using database GTEX(47) v8 with FUMA(27). The eQTL mapping assigned SNPs to genes up to 1Mb apart.

### Heritability and genetic correlation estimation

The LDSC software (https://github.com/bulik/ldsc) was employed to estimate the heritability of cereal intake and coffee intake as well as their genome-wide genetic correlation(18). We used the pre-calculated LD scores using 1000 Genomes European data. We used the overlap of summary statistics variants and HapMap variants as recommended(18).

### Inference of causality based on a modified Mendelian randomisation approach

Mendelian randomisation has been shown as a powerful tool to establish random experiments based on genetic variants that are valid as instrumental variables that are directly linked to the independent variables only. An instrumental variable could represent different ‘trials’ of a random experiment (of the independent variable) and hence leading to a causal inference on the outcome (i.e. the dependent variable). However, due to the lack of knowledge about which SNPs could serve as instrumental variables that only directly link to the independent variable but not the dependent variable, it is generally very difficult to perform the Mendelian randomisation on quantitative phenotypes without the confounding of genetic pleiotropy (the same genetic variant may influence both phenotypes of questions independently) and the risk of reverse causality (the dependent variable may reversely also have causal influence on the independent variable). Therefore, in many occasions, the interpretation of Mendelian randomisation results heavily relied on widely accepted hypothetic causal relationship, thus paradoxically, these applications of Mendelian randomisation do not serve its original purpose as a tool to infer causality without any pre-knowledge.

Nevertheless, as the influence of genetic variants on the dependent variable (as an indirect effect through the independent variable) should always be lower than that on the directly influenced independent variable, by gradually removing SNPs that cross-associated with both independent and dependent variables based on varied threshold, we could eventually keep only SNPs influencing the independent variables, thus valid as instrumental variables. Further, this approach is unbiased in that instrumental variables for both dependent and independent variables could be acquired simultaneously, and therefore a bi-directional causal inference could be conducted, which directly addresses the issue of reverse causal inference in traditional Mendelian randomisation approaches.

Finally, we employed the polygenic risk score (PRS), in combination with the above process to remove pleiotropy, as the instrumental variable to integrate contribution from multiple genetic variants, instead of using a single SNP, to enlarge the difference between ‘trials’ randomised by the instrumental variable.

### Mediation analysis

Mediation effects were examined using Baron and Kenny’s (1986)(50) causal steps approach. The causal steps approach involved four steps to establishing mediation. Firstly, a significant relation of the independent variable to the dependent variable is required in *Y* = *k*_1_ + *τX* + *ε*_1_ (*reject H*_0_: *τ* = 0). Secondly, a significant relation of the independent variable to the hypothesised mediating variable is required in *Z* = *k*_2_ + *αX* + *ε*_2_ (*reject H*_0_: *α* = 0). Thirdly, the mediating variable must be significantly related to the dependent variable when both the independent variable and mediating variable are predictors of the dependent variable in *Y* = *k*_3_ + *τ*′*X* + *βZ* + *ε*_3_ (*reject H*_0_: *β* = 0) Fourthly, the coefficient relating the independent variable to the dependent variable must be larger (in absolute value) than the coefficient relating the independent variable to the dependent variable in the regression model with both the independent variable and the mediating variable predicting the dependent variable (*i. e*. |*τ*| > |*τ*′|). To further evaluate the p-value of the significant mediation identified by the above process, we performed 1000 times bootstrap of the individuals to obtain the distribution of the proportion of the mediation, i.e., *PM* = (*τ* − *τ*′)/*τ*, under the alternative hypothesis. Thus, the PM was expected to be positive by definition, and the corresponding p-value could be calculated as the doubled chance of observing the PM less than zero during the 1000 bootstrap procedure. As no priory assumption about whether diet or lifestyle/ blood and body fat levels should serve as the mediator for their associations with the lead SNPs, we, therefore, identified the most likely mediator with an excess PM, i.e., the model showing higher PM, of which the significance level was again evaluated through a 1000-bootstrap process.

### Pattern similarity analysis

We examined the similarity among the brain-wide GMV-association patterns of cereal/coffee intake and cognitive functions. Specifically, we first performed association analyses between region-wide GMV and each phenotype. Then, we calculated the Pearson correlation coefficient (similarity) between the GMV-association patterns of a pair of phenotypes of interest, of which the significance level was evaluated through 10000 times permutation that shuffled the individual’s IDs of the GMV data at each iteration.

The similarity between brain-wide GMV-association pattern of a given phenotype and the brain-wide gene-expression pattern was also examined through their pattern correlation, of which the null distribution was established through 10000 times permutation that at each iteration, the pattern correlation was re-calculated with the GMV-association patterns been regenerated with shuffled IDs of the GMV data. The corresponding p-values were hence calculated as the chance of randomly getting a higher pattern correlation than the observed one in terms of their absolute value based on the established null distribution. The above permutation process was employed to ensure that the potential oversampling of brain regions will not inflate the false positive rate.

### Functional enrichment analysis

Functional properties of these genes were characterized by gene ontology (GO) terms including molecular function (MF), cellular component (CC), and biological process (BP) using “clusterProfiler” (28) R package. Adjusted p values were acquired using Benjamini-Hochberg method.

## Data Availability

All UK Biobank data used in this work were obtained under Data Access Application 19542 and are available to eligible researchers through the UK Biobank (www.biobank.ac.uk). Gene expression data from the Allen Institute for Brain Science are freely available at https://human.brain-map.org/static/download. Custom code that supports the findings of this study is available from the corresponding author upon request.

## Author Contributions

<u>Conception or Design of the Study:</u> T.J., B.J.S. and J.F.. <u>Manuscript Writing and Editing:</u> J.K., T.J. and D.W. wrote the manuscript; B.J.S. and J.F. edited the first draft; all authors critically reviewed the manuscript. <u>Imaging Data Preprocessing:</u> J.K., Z.J. and W.C.. <u>Visualisation:</u> J.K., T.J. and C.X.. <u>Data Analysis:</u> J.K. conducted all the statistical analyses, under the instruction of T.J.. <u>Results Interpretation:</u> T.J., B.J.S. and J.F.. <u>Supervision of the Study:</u> T.J. and J.F.. <u>Funding Acquisition:</u> T.J. and J.F..

## Data Availability

All UK Biobank data used in this work were obtained under Data Access Application 19542 and are available to eligible researchers through the UK Biobank (www.biobank.ac.uk). Gene expression data from the Allen Institute for Brain Science are freely available at https://human.brain-map.org/static/download.

## Code Availability

Custom code that supports the findings of this study is available from the corresponding author upon request.

