## Supplementary Materials for "Increased brain volume from higher cereal and lower coffee intake: Shared genetic determinants and impacts on cognition and metabolism"

**This PDF file includes:**

Supplementary Methods
Supplementary Figure
Table S1 to S23

### Supplementary Methods

#### Assessment of diet

Dietary data were obtained from the touchscreen questionnaire at the baseline and the MRI scan appointment. Participants were asked about their frequency of intake of a range of common food and drink items [UK Biobank Category ID: 100052]. The questionnaires were described as below. Cereal intake was obtained from the touchscreen questionnaire. Participants were asked, "How many bowls of cereal do you eat a week?" Either answer < 0 or > 99 was then rejected. If answer > 14, then the participant was asked to confirm. Participants who indicated that they ate cereal at least one bowl of cereal each week were further asked, "What type of cereal do you mainly eat?" The answers included Bran cereal (e.g., All Bran, Branflakes), Biscuit cereal (e.g., Weetabix), Oat cereal (e.g., Ready Brek, porridge), Muesli, and Other (e.g., Cornflakes, Frosties). For coffee intake, participants were asked, "How many cups of coffee do you drink each day? (Include decaffeinated coffee)". Participants who indicated that they drank at least one glass of coffee each day were further asked, "What type of coffee do you usually drink?" The answers included Decaffeinated coffee (any type), Instant coffee, Ground coffee (including espresso, filter, etc.), Other types of coffee. Participants were asked, "How many glasses of water do you drink a day?" for water intake. For tea intake, participants were asked, "How many cups of tea do you drink each day? (Include black and green tea)". Cooked vegetable intake was obtained from the question, "On average how many heaped tablespoons of cooked vegetables would you eat per day? (Do not include potatoes)". Salad / raw vegetable intake assessed using the following question "On average, how many heaped tablespoons of salad or raw vegetables would you eat per day? (Include lettuce, tomato in sandwiches)". Fresh fruit intake was obtained from the question "About how many pieces of fresh fruit would you eat per day? (Count one apple, one banana, ten grapes, etc. as one piece)". Dried fruit intake was obtained from the question "About how many pieces of dried fruit would you eat per day? (Count one prune, one dried apricot, ten raisins as one piece)". Bread intake was obtained from the question, "How many slices of bread do you eat each week?". For food and drink items above, the answer "less than one" was set to 0.5. Participants were asked about their intake frequency of oil fish/ non-oily fish, processed meat, poultry, beef lamb/mutton, pork, cheese, which was reported as "never", "less than once a week", "once a week", "2-4 times a week", "5-6 times a week", and "Once or more daily" and coded from 0 to 5.

In this study, we excluded individuals beyond four times the standard deviation from the mean for each kind of diet phenotype.

**Assessment of physical activity**

We used self-reported physical activity measured using a modified version of the International Physical Activity Questionnaire (IPAQ), which assessed total physical activity (TPA), including walking, moderate, and vigorous activity undertaken over the last seven days. MET (Metabolic Equivalent Task) scores data were derived based on IPAQ (International Physical Activity Questionnaire) guidelines. Detailed information referred to the website (http://biobank.ndph.ox.ac.uk/showcase/label.cgi?id=54). We used total Metabolic Equivalent Task (MET) minutes per week for all activity, including walking, moderate, and vigorous activity. Usual walking pace was characterized as "slow pace", "steady average pace", "brisk pace", and coded to 1 to 3. To assess the time spent watching television, participants were asked, "In a typical day, how many hours do you spend watching TV?"

**Assessment of sleep**

Sleep duration was recorded as the number of reported hours to the following question "About how many hours sleep do you get in every 24 hours? (please include naps)". Participants were asked "on an average day, how easy do you find getting up in the morning?" with four responses provided ("not at all easy", "not very easy", "fairly easy", "very easy") and coded to 1 to 4. Participants were asked, "Do you consider yourself to be..." to assess morningness or eveningness (chronotype). Four options were provided under this question: "definitely a 'morning' person", "more a 'morning' than 'evening' person", "more an 'evening' than 'morning' person", "definitely an 'evening' person" and coded 1 to 4. To assess insomnia symptoms, participants were asked, "Do you have trouble falling asleep at night, or do you wake up in the middle of the night?" with three options "never/rarely", "sometimes", and "usually" and codes 1,2,3. Participants were asked, "Do you have a nap during the day?" with the same responses to insomnia symptoms. Finally, daytime dozing/sleeping (narcolepsy) was coded based on answers to the following question: "How likely are you to doze off or fall asleep during the daytime when you don't mean to", with the following options: "never/rarely", "sometimes", "often", "all of the time", coded to 0 to 3.

**Assessment of smoking**

Smoking data were obtained from the touchscreen questionnaire at the baseline appointment. Participants were asked about their smoking history [UK Biobank Field ID: 20160]. The number of cigarettes currently smoked daily (current cigarette smokers) was acquired from the question "About how many cigarettes do you smoke on average each day?".

**Assessment of alcohol intake**

Alcohol intake data were obtained from the touchscreen questionnaire at the baseline appointment. Participants were asked about their alcohol intake frequency [UK Biobank Field ID: 1558] was reported as "never", "special occasions only", "one to three times a month", "once or twice a week", "three or four times a week", and "daily or almost daily" and coded from 0 to 5. Average weekly alcohol consumption of a range of drink types: red wine, champagne /white wine, beer/cider, spirits, and fortified wine were collected from participants who indicated they drink alcohol more often than once or twice a week. The measurement units for each of the five alcoholic drink types: measures for spirits, glasses for wines, and pints for beer/cider, which were estimated to be equivalent to 1, 2, and 2.5 units, respectively. Individuals reporting current intake frequency of "one to three times a month", "special occasions only" or "never" (for whom this phenotype was not collected), were assumed to have a weekly alcohol consumption volume of 0. From these measures, we calculated an average intake of alcoholic units per week, derived by combining the self-reported estimated intakes of the different alcoholic beverage consumptions across the five drink types, as in a previous study[1].

**Assessment of cognitive function**

Fluid intelligence score was obtained from a simple unweighted sum of the number of correct answers given to the 13 fluid intelligence questions. Participants who did not answer all of the questions within the allotted two min limit are scored as zero for each of the unattempted questions.

Reaction time: First, the participants were presented with 2 cards in the middle of the screen. Second, the participants were asked to press the button box as soon as possible when two cards matched. The variable was the mean duration (milliseconds) to the first press of snap-button summed over rounds in which both cards matched.

Numeric memory: Firstly, the participants were presented with a string of numbers on the screen, which subsequently disappeared. Secondly, the participants were asked to enter the number in reverse order. If the answer was correct, there would be a longer number on the screen to remember. The test would end when two mistakes are made or five successive incorrect responses at string length = 2. The variable was maximum string length recalled correctly (range: 0-12), with higher scores indicating better performance.

Pairs matching: Firstly, participants were shown six card-pairs of symbols randomly scrambled on screen. Secondly, cards will then turn face down on the screen, and the participants were asked to select the pairs by touching the screen. As pairs were found correctly, the cards will disappear. The variable was the number of errors made during pairs matching (range 0-146).

Prospective memory: In this test, the participants were asked to remember to carry out a pre-planned instruction after a filled interval. First, at the beginning of the test, they were shown the following instruction: "At the end of the games, we will show you four colored symbols and ask you to touch the blue square. However, to test your memory, we want you to actually touch the orange circle instead." The variable was whether or not the participants remembered to touch the orange circle on the first attempt.

Matrix pattern completion: First, the participants were shown a series of matrix pattern blocks with missing elements. Second, the participants were asked to choose the element that could best complete the pattern from a series of given choices. The variable was the number of puzzles for which the participant gave the correct solution.

Symbol digit substitution: First, the participants were shown a grid linking symbols to a single integer, and another grid containing only symbols. Second, the participants were required to use the symbol-integer pair in the first grid as a key to indicate the number attached to each symbol in the second grid. The variable is the number of symbols correctly matched to digits by the participants.

Tower rearranging: First, the participants were shown illustrations of three piles (towers) on which three iron rings of different colors were placed. Second, the participants were asked to indicate how many steps it takes to rearrange the hoop to another specific position. The variable was the number of puzzles correctly answered.

Trail making: First, the participants were shown 25 circles distributed on the screen, which contained 1-25 numbers (digital trails) or a mixture of numbers and letters (alphanumeric trails). Second, the participants are required to connect the circle from the smallest number to the largest number in one task (number tracking), or alternate between numbers and letters in ascending alphabetical order in another task (alphanumeric tracking). The variable was the duration (deciseconds) to complete the trail. Please note that we reverse the sign of the variable to ensure that the higher the score, the better the performance.

**Structural MRI**

Structural MRIs were collected across three imaging centers that were equipped with identical scanners (Siemens Skyra 3T running VD13A SP4 with a Siemens 32-channel RF receive head coil, Munich, Germany). For each scan, Siemens auto-align software determined the field-of-view, which aligns a scout scan to an atlas. If auto-align failed, the radiographer set the alignment. Structural images were acquired with straight sagittal orientation (i.e., with the field of view aligned to the scanner axes), with a resolution of 1 × 1 × 1 mm and a field of view of 208 × 256 × 256 matrix, over a duration of 5 minutes, and with 1-mm isotropic resolution using a 3-dimensional magnetization-prepared rapid-acquisition gradient echo. The MRI protocols have been described in detail elsewhere[2].

**Genetic Data**

Genotype data are available for all 500,000 participants in the UKB cohort. All blood samples were genotyped using the UK BiLEVE array and the UK Biobank axiom array. The two single nucleotide polymorphism (SNP) arrays are very similar, with over 95% common marker content. Quality control was performed by Affymetrix, the Wellcome Trust Centre for Human Genetics, and by the present authors; this included removal of participants based on missingness, relatedness, gender mismatch, non-British ancestry and other criteria, details of the array design, genotyping, quality control and imputation are available in a previous publication[3].

#### Supplementary Figure

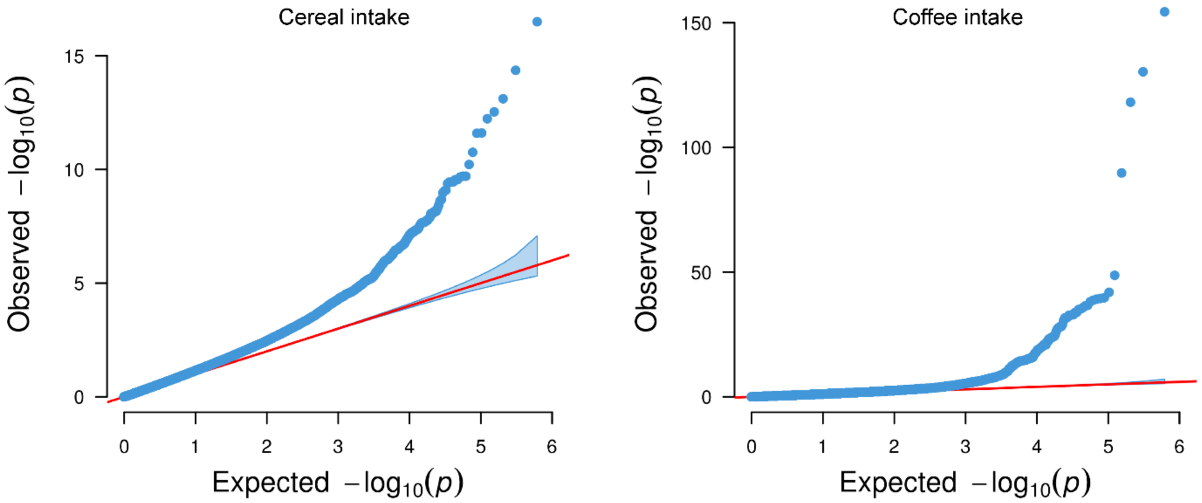


**Fig. S1.** QQ-plots for the genome-wide association studies of cereal intake (left) and coffee intake (right). Inflations of the QQ-plots were not due to population stratification, with intercepts from LD regression estimated as 1.013 (cereal intake) and 1.005 (coffee intake).

Table S1: Demographic characteristics.

| | Baseline (2006-2010) | | | Baseline (2006-2010) | | | | | | | | | Follow-up (2014+) | | | | | | | | |
|---|---|---|---|---|---|---|---|---|---|---|---|---|---|---|---|---|---|---|---|---|---|
| | | | | Discovery sample | | | Replication sample | | | T test | | | Discovery sample | | | Replication sample | | | T test | | |
| N.genotyped,non-AD | 336517 | | | 18879 | | | 16412 | | | Cohen's D | T | P | 18879 | | | 16412 | | | Cohen's D | T | P |
| N. of females (%) | 180737(53.71%) | | | 9890(52.39%) | | | 8655(52.74%) | | | NA | | | 9890(52.39%) | | | 8655(52.74%) | | | 0.007 | 0.66 | 5.12E-01 |
| | N | mean | std | N | mean | std | N | mean | std | | | | N | mean | std | N | mean | std | | | |
| Age at baseline (years) | 336517 | 56.87 | 7.99 | 18879 | 55.10 | 7.46 | 16412 | 54.90 | 7.37 | NA | | | 18879 | 55.10 | 7.46 | 16412 | 54.90 | 7.37 | 0.026 | 2.45 | 1.44E-02 |
| Age at MRI scan (years) | | | | | NA | | | NA | | NA | | | 18879 | 63.24 | 7.45 | 16412 | 64.98 | 7.40 | -0.234 | -21.94 | 6.12E-106 |
| **Diet** | | | | | | | | | | | | | | | | | | | | | |
| Cereal intake | 335696 | 4.63 | 2.75 | 18858 | 4.85 | 2.66 | 16395 | 4.78 | 2.69 | 0.027 | 2.52 | 1.19E-02 | 18727 | 4.58 | 2.78 | 16311 | 4.42 | 2.84 | 0.058 | 5.45 | 5.11E-08 |
| Coffee intake | 335068 | 2.04 | 1.94 | 18840 | 2.07 | 1.92 | 16314 | 2.00 | 1.80 | 0.038 | 3.53 | 4.14E-04 | 18657 | 2.03 | 1.75 | 16251 | 2.00 | 1.67 | 0.018 | 1.64 | 1.01E-01 |
| Water intake | 333236 | 2.56 | 2.03 | 18756 | 2.42 | 1.98 | 16317 | 2.56 | 2.00 | -0.069 | -6.45 | 1.17E-10 | 18643 | 2.38 | 1.89 | 16227 | 2.61 | 1.93 | -0.120 | -11.16 | 7.29E-29 |
| Tea intake | 333868 | 3.38 | 2.55 | 18781 | 3.40 | 2.54 | 16338 | 3.30 | 2.47 | 0.042 | 3.93 | 8.48E-05 | 18678 | 3.36 | 2.47 | 16220 | 3.15 | 2.33 | 0.086 | 8.03 | 1.02E-15 |
| Cooked vegetable intake | 330285 | 2.59 | 1.37 | 18667 | 2.55 | 1.33 | 16238 | 2.56 | 1.33 | -0.011 | -0.98 | 3.27E-01 | 18587 | 2.72 | 1.49 | 16077 | 2.72 | 1.42 | 0.003 | 0.31 | 7.57E-01 |
| Salad / raw vegetable intake | 331342 | 2.05 | 1.68 | 18707 | 2.06 | 1.69 | 16184 | 2.02 | 1.53 | 0.028 | 2.58 | 9.82E-03 | 18555 | 2.12 | 1.71 | 16171 | 2.23 | 1.73 | -0.063 | -5.87 | 4.30E-09 |
| Fresh fruit intake | 334354 | 2.16 | 1.39 | 18767 | 2.17 | 1.34 | 16323 | 2.20 | 1.34 | -0.021 | -1.94 | 5.27E-02 | 18647 | 2.19 | 1.39 | 16243 | 2.25 | 1.40 | -0.041 | -3.78 | 1.59E-04 |
| Dried fruit intake | 330603 | 0.73 | 1.18 | 18651 | 0.80 | 1.20 | 16191 | 0.80 | 1.20 | -0.001 | -0.11 | 9.10E-01 | 18524 | 0.79 | 1.20 | 16082 | 0.81 | 1.21 | -0.018 | -1.63 | 1.04E-01 |
| Bread intake | 331220 | 12.26 | 8.03 | 18605 | 12.25 | 8.05 | 16249 | 11.92 | 7.82 | 0.041 | 3.85 | 1.19E-04 | 18613 | 10.26 | 7.10 | 16187 | 9.67 | 6.68 | 0.084 | 7.85 | 4.30E-15 |
| Oily fish intake | 335040 | 1.64 | 0.92 | 18848 | 1.64 | 0.89 | 16389 | 1.64 | 0.89 | -0.001 | -0.07 | 9.41E-01 | 18731 | 1.77 | 0.91 | 16304 | 1.77 | 0.93 | -0.004 | -0.38 | 7.04E-01 |
| Non-oily fish intake | 334997 | 1.80 | 0.77 | 18843 | 1.78 | 0.77 | 16375 | 1.77 | 0.76 | 0.014 | 1.27 | 2.05E-01 | 18706 | 1.78 | 0.76 | 16288 | 1.78 | 0.77 | 0.002 | 0.21 | 8.34E-01 |
| Processed meat intake | 336048 | 1.90 | 1.04 | 18871 | 1.86 | 1.05 | 16402 | 1.84 | 1.05 | 0.017 | 1.57 | 1.18E-01 | 18739 | 1.83 | 1.04 | 16314 | 1.77 | 1.06 | 0.053 | 4.92 | 8.59E-07 |
| Poultry intake | 335976 | 2.30 | 0.87 | 18869 | 2.29 | 0.89 | 16404 | 2.29 | 0.90 | 0.002 | 0.17 | 8.65E-01 | 18744 | 2.34 | 0.92 | 16317 | 2.29 | 0.94 | 0.052 | 4.87 | 1.13E-06 |
| Beef intake | 335277 | 1.46 | 0.82 | 18862 | 1.43 | 0.82 | 16398 | 1.45 | 0.82 | -0.018 | -1.67 | 9.45E-02 | 18725 | 1.38 | 0.80 | 16299 | 1.35 | 0.80 | 0.046 | 4.33 | 1.51E-05 |
| Lamb/mutton intake | 334587 | 1.10 | 0.70 | 18834 | 1.08 | 0.67 | 16383 | 1.09 | 0.68 | -0.011 | -1.02 | 3.08E-01 | 18715 | 1.00 | 0.63 | 16283 | 0.95 | 0.64 | 0.065 | 6.04 | 1.59E-09 |
| Pork intake | 334529 | 1.14 | 0.70 | 18833 | 1.12 | 0.68 | 16377 | 1.11 | 0.67 | 0.016 | 1.48 | 1.40E-01 | 18703 | 1.09 | 0.68 | 16276 | 1.03 | 0.67 | 0.081 | 7.59 | 3.17E-14 |
| Cheese intake | 328719 | 2.55 | 1.06 | 18596 | 2.62 | 1.04 | 16126 | 2.68 | 1.05 | -0.054 | -4.99 | 6.20E-07 | 18416 | 2.66 | 1.07 | 15980 | 2.78 | 1.08 | -0.109 | -10.12 | 4.89E-24 |
| **Lifestyle** | | | | | | | | | | | | | | | | | | | | | |
| Time spent watching television (TV) | 334146 | 2.81 | 1.64 | 18825 | 2.46 | 1.44 | 16373 | 2.35 | 1.40 | 0.073 | 6.80 | 1.09E-11 | 18713 | 2.83 | 1.53 | 16284 | 2.78 | 1.53 | 0.031 | 2.87 | 4.05E-03 |
| Usual walking pace | 334676 | 2.33 | 0.61 | 18846 | 2.47 | 0.56 | 16392 | 2.49 | 0.55 | -0.033 | -3.07 | 2.13E-03 | 18723 | 2.42 | 0.57 | 16304 | 2.41 | 0.58 | 0.030 | 2.83 | 4.61E-03 |
| Summed MET minutes per week for all activity | 272702 | 2665.33 | 2709.46 | 16170 | 2475.16 | 2454.39 | 14163 | 2445.76 | 2383.88 | 0.012 | 1.05 | 2.92E-01 | | NA | | | NA | | | NA | |
| MET minutes per week for moderate activity | 272702 | 943.81 | 1232.02 | 16170 | 836.99 | 1124.67 | 14163 | 820.33 | 1107.09 | 0.015 | 1.30 | 1.95E-01 | | NA | | | NA | | | NA | |
| MET minutes per week for vigorous activity | 272702 | 673.03 | 1196.86 | 16170 | 672.17 | 1068.91 | 14163 | 668.95 | 1036.28 | 0.003 | 0.27 | 7.91E-01 | | NA | | | NA | | | NA | |
| MET minutes per week for walking | 272702 | 1048.49 | 1089.74 | 16170 | 966.00 | 1033.92 | 14163 | 956.47 | 1009.89 | 0.009 | 0.81 | 4.19E-01 | | NA | | | NA | | | NA | |
| Sleep duration | 334737 | 7.17 | 1.08 | 18833 | 7.17 | 0.96 | 16388 | 7.18 | 0.95 | -0.012 | -1.13 | 2.57E-01 | 18710 | 7.16 | 1.04 | 16280 | 7.15 | 1.02 | 0.005 | 0.43 | 6.70E-01 |
| Getting up in morning | 335820 | 3.12 | 0.77 | 18765 | 3.10 | 0.77 | 16360 | 3.10 | 0.75 | 0.006 | 0.56 | 5.72E-01 | 18745 | 3.22 | 0.73 | 16313 | 3.26 | 0.73 | -0.053 | -4.92 | 8.76E-07 |
| Morning/evening person (chronotype) | 300556 | 2.19 | 0.93 | 16803 | 2.23 | 0.92 | 14631 | 2.23 | 0.93 | -0.006 | -0.52 | 6.05E-01 | 17072 | 2.16 | 0.94 | 14882 | 2.15 | 0.94 | 0.010 | 0.88 | 3.77E-01 |
| Nap during day | 336393 | 1.49 | 0.60 | 18879 | 1.43 | 0.57 | 16408 | 1.41 | 0.56 | 0.029 | 2.75 | 5.93E-03 | 18750 | 1.48 | 0.60 | 16321 | 1.49 | 0.60 | -0.025 | -2.37 | 1.78E-02 |
| Sleeplessness / insomnia | 336284 | 2.05 | 0.72 | 18871 | 1.99 | 0.72 | 16403 | 1.96 | 0.73 | 0.046 | 4.30 | 1.71E-05 | 18743 | 2.09 | 0.72 | 16308 | 2.12 | 0.73 | -0.035 | -3.31 | 9.49E-04 |
| Daytime dozing / sleeping (narcolepsy) | 335404 | 0.26 | 0.49 | 18859 | 0.22 | 0.46 | 16393 | 0.22 | 0.46 | 0.002 | 0.16 | 8.69E-01 | 18730 | 0.25 | 0.48 | 16307 | 0.25 | 0.49 | -0.015 | -1.41 | 1.59E-01 |
| Ever smoked | 335394 | 0.60 | 0.49 | 18840 | 0.57 | 0.49 | 16390 | 0.56 | 0.50 | 0.018 | 1.73 | 8.44E-02 | 18696 | 0.54 | 0.50 | 16270 | 0.53 | 0.50 | 0.022 | 2.05 | 4.00E-02 |
| Number of cigarettes currently smoked daily (current cigarette smokers) | 23173 | 15.75 | 8.42 | 687 | 14.64 | 8.83 | 542 | 13.93 | 7.93 | 0.084 | 1.47 | 1.42E-01 | 377 | 12.62 | 7.48 | 255 | 13.18 | 8.57 | -0.071 | -0.88 | 3.81E-01 |
| Alcohol intake frequency. | 336285 | 3.16 | 1.48 | 18877 | 3.34 | 1.36 | 16409 | 3.37 | 1.37 | -0.021 | -1.99 | 4.70E-02 | 18750 | 3.13 | 1.40 | 16313 | 3.14 | 1.42 | -0.004 | -0.38 | 7.06E-01 |
| Average weekly alcohol intake | 332474 | 16.49 | 19.58 | 18768 | 17.18 | 18.05 | 16310 | 17.37 | 17.87 | -0.010 | -0.97 | 3.31E-01 | 18649 | 14.65 | 16.69 | 16219 | 14.36 | 16.36 | 0.018 | 1.64 | 1.01E-01 |
| **Body/blood fat** | | | | | | | | | | | | | | | | | | | | | |
| Body mass index (BMI) | 335428 | 27.39 | 4.75 | 18879 | 26.65 | 4.27 | 16412 | 26.49 | 4.16 | 0.037 | 3.45 | 5.52E-04 | | NA | | | NA | | | NA | |
| Cholesterol | 320830 | 5.71 | 1.14 | 17857 | 5.73 | 1.10 | 15703 | 5.75 | 1.07 | -0.018 | -1.64 | 1.01E-01 | | NA | | | NA | | | NA | |
| HDL cholesterol | 293704 | 1.45 | 0.38 | 16269 | 1.47 | 0.37 | 14330 | 1.49 | 0.38 | -0.040 | -3.50 | 4.59E-04 | | NA | | | NA | | | NA | |
| LDL direct | 320224 | 3.57 | 0.87 | 17820 | 3.58 | 0.84 | 15671 | 3.59 | 0.82 | -0.015 | -1.36 | 1.73E-01 | | NA | | | NA | | | NA | |
| Triglycerides | 320566 | 1.76 | 1.02 | 17843 | 1.66 | 0.98 | 15691 | 1.63 | 0.94 | 0.037 | 3.41 | 6.49E-04 | | NA | | | NA | | | NA | |
| **Cognitive function** | | | | | | | | | | | | | | | | | | | | | |
| Fluid intelligence score | 108659 | 6.18 | 2.10 | 6745 | 6.75 | 1.98 | 4999 | 6.87 | 2.03 | -0.060 | -3.23 | 1.24E-03 | 17045 | 6.80 | 2.06 | 15536 | 6.57 | 2.03 | 0.116 | 10.43 | 2.00E-25 |
| Maximum digits remembered correctly | 34880 | 6.72 | 1.32 | 1868 | 7.00 | 1.21 | 1737 | 6.98 | 1.25 | 0.014 | 0.43 | 6.67E-01 | 8184 | 6.80 | 1.24 | 15355 | 6.78 | 1.26 | 0.019 | 1.38 | 1.68E-01 |
| Prospective memory result | 110912 | 0.80 | 0.40 | 6791 | 0.88 | 0.33 | 5020 | 0.88 | 0.32 | -0.014 | -0.77 | 4.44E-01 | 17459 | 0.87 | 0.33 | 15696 | 0.82 | 0.38 | 0.140 | 12.71 | 6.51E-37 |
| Mean time to correctly identify matches | 334488 | 555.37 | 112.91 | 18849 | 538.20 | 100.14 | 16383 | 533.91 | 97.81 | 0.043 | 4.05 | 5.06E-05 | 17411 | 587.69 | 106.70 | 15595 | 598.72 | 109.33 | -0.102 | -9.27 | 1.96E-20 |
| Number of incorrect matches in round | 336199 | 0.50 | 1.10 | 18778 | 0.36 | 0.86 | 16367 | 0.37 | 0.90 | -0.012 | -1.10 | 2.73E-01 | 17476 | 0.33 | 0.83 | 15698 | 0.35 | 0.86 | -0.024 | -2.19 | 2.84E-02 |
| Number of puzzles correctly solved | | NA | | | NA | | | NA | | | NA | | 7564 | 8.07 | 2.10 | 15378 | 8.00 | 2.12 | 0.032 | 2.25 | 2.42E-02 |
| Number of symbol digit matches made correctly | | NA | | | NA | | | NA | | | NA | | 7584 | 19.35 | 5.21 | 15385 | 18.83 | 5.24 | 0.099 | 7.07 | 1.64E-12 |
| Number of puzzles correct | | NA | | | NA | | | NA | | | NA | | 7495 | 10.09 | 3.22 | 15264 | 9.93 | 3.21 | 0.051 | 3.59 | 3.34E-04 |
| Duration to complete numeric path | | NA | | | NA | | | NA | | | NA | | 7647 | 217.10 | 83.23 | 15520 | 224.39 | 86.74 | -0.085 | -6.10 | 1.10E-09 |
| Duration to complete alphanumeric path | | NA | | | NA | | | NA | | | NA | | 7647 | 531.45 | 255.91 | 15520 | 555.02 | 273.04 | -0.088 | -6.31 | 2.91E-10 |

Table S2: Associations between grey matter volumes and diet phenotypes (follow-up).

| Number | AAL 3 region | beta | 95%CI_l | 95%CI_h | se | t | r | p | df.e | beta | 95%CI_l | 95%CI_h | se | t | r | p | df.e |
|---|---|---|---|---|---|---|---|---|---|---|---|---|---|---|---|---|---|
| | | Coffee intake | | | | | | | | Cereal intake | | | | | | | |
| 0 | Total brain | -340.263 | -415.637 | -264.889 | 38.454 | -8.849 | -0.065 | 9.65E-19 | 18651 | 289.283 | 241.700 | 336.866 | 24.276 | 11.916 | 0.087 | 1.27E-32 | 18721 |
| 1 | Precentral_L | -8.109 | -10.765 | -5.454 | 1.355 | -5.985 | -0.044 | 2.20E-09 | 18651 | 4.094 | 2.414 | 5.774 | 0.857 | 4.778 | 0.035 | 1.79E-06 | 18721 |
| 2 | Precentral_R | -7.425 | -9.859 | -4.991 | 1.242 | -5.980 | -0.044 | 2.27E-09 | 18651 | 5.492 | 3.954 | 7.031 | 0.785 | 7.000 | 0.051 | 2.65E-12 | 18721 |
| 3 | Frontal_Sup_2_L | -9.415 | -12.218 | -6.613 | 1.430 | -6.585 | -0.048 | 4.67E-11 | 18651 | 6.944 | 5.173 | 8.715 | 0.904 | 7.685 | 0.056 | 1.61E-14 | 18721 |
| 4 | Frontal_Sup_2_R | -11.824 | -14.911 | -8.736 | 1.575 | -7.506 | -0.055 | 6.37E-14 | 18651 | 8.035 | 6.083 | 9.986 | 0.996 | 8.068 | 0.059 | 7.56E-16 | 18721 |
| 5 | Frontal_Mid_2_L | -9.467 | -12.411 | -6.523 | 1.502 | -6.303 | -0.046 | 2.98E-10 | 18651 | 6.858 | 4.998 | 8.719 | 0.949 | 7.225 | 0.053 | 5.20E-13 | 18721 |
| 6 | Frontal_Mid_2_R | -12.563 | -15.893 | -9.234 | 1.699 | -7.397 | -0.054 | 1.45E-13 | 18651 | 9.195 | 7.091 | 11.299 | 1.073 | 8.566 | 0.062 | 1.16E-17 | 18721 |
| 7 | Frontal_Inf_Oper_L | -2.543 | -3.413 | -1.673 | 0.444 | -5.729 | -0.042 | 1.03E-08 | 18651 | 1.655 | 1.105 | 2.205 | 0.281 | 5.896 | 0.043 | 3.78E-09 | 18721 |
| 8 | Frontal_Inf_Oper_R | -3.103 | -4.195 | -2.011 | 0.557 | -5.571 | -0.041 | 2.57E-08 | 18651 | 2.438 | 1.748 | 3.127 | 0.352 | 6.930 | 0.051 | 4.33E-12 | 18721 |
| 9 | Frontal_Inf_Tri_L | -5.629 | -7.477 | -3.781 | 0.943 | -5.971 | -0.044 | 2.40E-09 | 18651 | 4.356 | 3.189 | 5.524 | 0.596 | 7.315 | 0.053 | 2.68E-13 | 18721 |
| 10 | Frontal_Inf_Tri_R | -4.926 | -6.377 | -3.474 | 0.741 | -6.651 | -0.049 | 3.00E-11 | 18651 | 3.456 | 2.539 | 4.373 | 0.468 | 7.388 | 0.054 | 1.55E-13 | 18721 |
| 11 | Frontal_Inf_Orb_2_L | -1.331 | -1.883 | -0.779 | 0.282 | -4.723 | -0.035 | 2.34E-06 | 18651 | 1.547 | 1.199 | 1.896 | 0.178 | 8.696 | 0.063 | 3.71E-18 | 18721 |
| 12 | Frontal_Inf_Orb_2_R | -1.488 | -2.066 | -0.910 | 0.295 | -5.047 | -0.037 | 4.53E-07 | 18651 | 1.629 | 1.264 | 1.994 | 0.186 | 8.753 | 0.064 | 2.26E-18 | 18721 |
| 13 | Rolandic_Oper_L | -2.183 | -3.019 | -1.346 | 0.427 | -5.116 | -0.037 | 3.16E-07 | 18651 | 1.564 | 1.035 | 2.092 | 0.270 | 5.800 | 0.042 | 6.72E-09 | 18721 |
| 14 | Rolandic_Oper_R | -3.386 | -4.583 | -2.189 | 0.611 | -5.545 | -0.041 | 2.97E-08 | 18651 | 2.824 | 2.069 | 3.580 | 0.386 | 7.326 | 0.053 | 2.47E-13 | 18721 |
| 15 | Supp_Motor_Area_L | -5.603 | -7.590 | -3.616 | 1.014 | -5.528 | -0.040 | 3.29E-08 | 18651 | 3.311 | 2.055 | 4.567 | 0.641 | 5.168 | 0.038 | 2.40E-07 | 18721 |
| 16 | Supp_Motor_Area_R | -6.147 | -8.343 | -3.952 | 1.120 | -5.489 | -0.040 | 4.09E-08 | 18651 | 3.760 | 2.373 | 5.148 | 0.708 | 5.312 | 0.039 | 1.10E-07 | 18721 |
| 17 | Olfactory_L | -0.537 | -0.833 | -0.242 | 0.151 | -3.564 | -0.026 | 3.66E-04 | 18651 | 0.602 | 0.415 | 0.789 | 0.095 | 6.323 | 0.046 | 2.63E-10 | 18721 |
| 18 | Olfactory_R | -0.565 | -0.861 | -0.269 | 0.151 | -3.736 | -0.027 | 1.87E-04 | 18651 | 0.508 | 0.321 | 0.696 | 0.095 | 5.325 | 0.039 | 1.02E-07 | 18721 |
| 19 | Frontal_Sup_Medial_L | -5.904 | -7.765 | -4.043 | 0.949 | -6.219 | -0.045 | 5.13E-10 | 18651 | 5.745 | 4.570 | 6.920 | 0.599 | 9.584 | 0.070 | 1.05E-21 | 18721 |
| 20 | Frontal_Sup_Medial_R | -4.487 | -5.842 | -3.132 | 0.691 | -6.491 | -0.047 | 8.74E-11 | 18651 | 3.644 | 2.788 | 4.500 | 0.437 | 8.348 | 0.061 | 7.45E-17 | 18721 |
| 21 | Frontal_Med_Orb_L | -2.116 | -2.812 | -1.420 | 0.355 | -5.957 | -0.044 | 2.61E-09 | 18651 | 2.050 | 1.610 | 2.489 | 0.224 | 9.149 | 0.067 | 6.34E-20 | 18721 |
| 22 | Frontal_Med_Orb_R | -2.708 | -3.563 | -1.853 | 0.436 | -6.205 | -0.045 | 5.57E-10 | 18651 | 2.464 | 1.924 | 3.003 | 0.275 | 8.948 | 0.065 | 3.95E-19 | 18721 |
| 23 | Rectus_L | -1.740 | -2.460 | -1.020 | 0.367 | -4.735 | -0.035 | 2.21E-06 | 18651 | 2.094 | 1.639 | 2.548 | 0.232 | 9.030 | 0.066 | 1.88E-19 | 18721 |
| 24 | Rectus_R | -1.668 | -2.334 | -1.002 | 0.340 | -4.911 | -0.036 | 9.16E-07 | 18651 | 1.773 | 1.353 | 2.194 | 0.215 | 8.265 | 0.060 | 1.49E-16 | 18721 |
| 25 | OFCmed_L | -1.109 | -1.526 | -0.691 | 0.213 | -5.208 | -0.038 | 1.92E-07 | 18651 | 0.954 | 0.691 | 1.218 | 0.134 | 7.096 | 0.052 | 1.33E-12 | 18721 |
| 26 | OFCmed_R | -1.268 | -1.754 | -0.781 | 0.248 | -5.109 | -0.037 | 3.26E-07 | 18651 | 0.888 | 0.581 | 1.196 | 0.157 | 5.666 | 0.041 | 1.49E-08 | 18721 |
| 27 | OFCant_L | -1.254 | -1.737 | -0.771 | 0.246 | -5.091 | -0.037 | 3.61E-07 | 18651 | 0.769 | 0.464 | 1.074 | 0.156 | 4.939 | 0.036 | 7.92E-07 | 18721 |
| 28 | OFCant_R | -1.483 | -2.121 | -0.845 | 0.326 | -4.556 | -0.033 | 5.25E-06 | 18651 | 1.351 | 0.948 | 1.754 | 0.205 | 6.573 | 0.048 | 5.05E-11 | 18721 |
| 29 | OFCpost_L | -1.531 | -2.136 | -0.927 | 0.308 | -4.965 | -0.036 | 6.93E-07 | 18651 | 0.977 | 0.595 | 1.359 | 0.195 | 5.016 | 0.037 | 5.33E-07 | 18721 |
| 30 | OFCpost_R | -1.300 | -1.859 | -0.741 | 0.285 | -4.560 | -0.033 | 5.16E-06 | 18651 | 1.302 | 0.949 | 1.655 | 0.180 | 7.233 | 0.053 | 4.91E-13 | 18721 |
| 31 | OFClat_L | -0.459 | -0.663 | -0.255 | 0.104 | -4.415 | -0.032 | 1.02E-05 | 18651 | 0.443 | 0.314 | 0.572 | 0.066 | 6.746 | 0.049 | 1.56E-11 | 18721 |
| 32 | OFClat_R | -0.381 | -0.553 | -0.209 | 0.088 | -4.338 | -0.032 | 1.44E-05 | 18651 | 0.467 | 0.358 | 0.576 | 0.055 | 8.419 | 0.061 | 4.08E-17 | 18721 |
| 33 | Insula_L | -6.983 | -8.849 | -5.117 | 0.952 | -7.335 | -0.054 | 2.31E-13 | 18651 | 4.304 | 3.124 | 5.484 | 0.602 | 7.148 | 0.052 | 9.14E-13 | 18721 |
| 34 | Insula_R | -6.881 | -8.701 | -5.062 | 0.928 | -7.413 | -0.054 | 1.29E-13 | 18651 | 5.083 | 3.934 | 6.232 | 0.586 | 8.668 | 0.063 | 4.75E-18 | 18721 |
| 37 | Cingulate_Mid_L | -5.538 | -7.116 | -3.960 | 0.805 | -6.878 | -0.050 | 6.28E-12 | 18651 | 5.013 | 4.017 | 6.010 | 0.508 | 9.860 | 0.072 | 7.09E-23 | 18721 |
| 38 | Cingulate_Mid_R | -5.675 | -7.359 | -3.992 | 0.859 | -6.608 | -0.048 | 3.99E-11 | 18651 | 5.561 | 4.498 | 6.624 | 0.542 | 10.257 | 0.075 | 1.28E-24 | 18721 |
| 39 | Cingulate_Post_L | -0.761 | -1.244 | -0.278 | 0.246 | -3.087 | -0.023 | 2.02E-03 | 18651 | 0.558 | 0.253 | 0.864 | 0.156 | 3.587 | 0.026 | 3.35E-04 | 18721 |
| 40 | Cingulate_Post_R | -0.178 | -0.446 | 0.090 | 0.137 | -1.299 | -0.010 | 1.94E-01 | 18651 | 0.197 | 0.027 | 0.366 | 0.086 | 2.277 | 0.017 | 2.28E-02 | 18721 |
| 41 | Hippocampus_L | -0.455 | -1.236 | 0.326 | 0.399 | -1.141 | -0.008 | 2.54E-01 | 18651 | 1.382 | 0.889 | 1.875 | 0.252 | 5.491 | 0.040 | 4.04E-08 | 18721 |
| 42 | Hippocampus_R | -0.557 | -1.367 | 0.253 | 0.413 | -1.348 | -0.010 | 1.78E-01 | 18651 | 1.183 | 0.672 | 1.695 | 0.261 | 4.533 | 0.033 | 5.86E-06 | 18721 |
| 43 | ParaHippocampal_L | -0.724 | -1.418 | -0.030 | 0.354 | -2.045 | -0.015 | 4.09E-02 | 18651 | 1.491 | 1.053 | 1.929 | 0.224 | 6.669 | 0.049 | 2.65E-11 | 18721 |
| 44 | ParaHippocampal_R | -1.062 | -1.937 | -0.186 | 0.447 | -2.377 | -0.017 | 1.75E-02 | 18651 | 2.088 | 1.535 | 2.641 | 0.282 | 7.404 | 0.054 | 1.38E-13 | 18721 |
| 45 | Amygdala_L | -0.617 | -0.887 | -0.348 | 0.138 | -4.488 | -0.033 | 7.23E-06 | 18651 | 0.464 | 0.293 | 0.634 | 0.087 | 5.335 | 0.039 | 9.65E-08 | 18721 |

| | | | | | | | | | | | | | | | | | |
|---|---|---|---|---|---|---|---|---|---|---|---|---|---|---|---|---|---|
| 46 | Amygdala_R | -0.798 | -1.073 | -0.522 | 0.141 | -5.676 | -0.042 | 1.40E-08 | 18651 | 0.572 | 0.398 | 0.746 | 0.089 | 6.433 | 0.047 | 1.28E-10 | 18721 |
| 47 | Calcarine_L | -6.668 | -9.238 | -4.097 | 1.312 | -5.084 | -0.037 | 3.73E-07 | 18651 | 0.712 | -0.914 | 2.339 | 0.830 | 0.858 | 0.006 | 3.91E-01 | 18721 |
| 48 | Calcarine_R | -5.356 | -7.505 | -3.207 | 1.096 | -4.886 | -0.036 | 1.04E-06 | 18651 | 0.788 | -0.571 | 2.148 | 0.694 | 1.137 | 0.008 | 2.56E-01 | 18721 |
| 49 | Cuneus_L | -3.575 | -5.188 | -1.961 | 0.823 | -4.343 | -0.032 | 1.42E-05 | 18651 | 1.073 | 0.053 | 2.093 | 0.521 | 2.061 | 0.015 | 3.93E-02 | 18721 |
| 50 | Cuneus_R | -3.156 | -4.391 | -1.922 | 0.630 | -5.013 | -0.037 | 5.41E-07 | 18651 | 1.380 | 0.600 | 2.161 | 0.398 | 3.467 | 0.025 | 5.28E-04 | 18721 |
| 51 | Lingual_L | -3.718 | -5.264 | -2.172 | 0.789 | -4.713 | -0.034 | 2.46E-06 | 18651 | 1.324 | 0.346 | 2.302 | 0.499 | 2.654 | 0.019 | 7.96E-03 | 18721 |
| 52 | Lingual_R | -3.067 | -4.689 | -1.446 | 0.827 | -3.708 | -0.027 | 2.10E-04 | 18651 | 1.699 | 0.674 | 2.724 | 0.523 | 3.249 | 0.024 | 1.16E-03 | 18721 |
| 53 | Occipital_Sup_L | -2.457 | -3.521 | -1.394 | 0.543 | -4.529 | -0.033 | 5.97E-06 | 18651 | 1.407 | 0.735 | 2.079 | 0.343 | 4.103 | 0.030 | 4.10E-05 | 18721 |
| 54 | Occipital_Sup_R | -2.695 | -3.849 | -1.542 | 0.588 | -4.582 | -0.034 | 4.64E-06 | 18651 | 2.033 | 1.305 | 2.762 | 0.372 | 5.473 | 0.040 | 4.49E-08 | 18721 |
| 55 | Occipital_Mid_L | -5.709 | -8.202 | -3.216 | 1.272 | -4.489 | -0.033 | 7.20E-06 | 18651 | 1.848 | 0.272 | 3.424 | 0.804 | 2.299 | 0.017 | 2.15E-02 | 18721 |
| 56 | Occipital_Mid_R | -3.824 | -5.558 | -2.090 | 0.885 | -4.322 | -0.032 | 1.55E-05 | 18651 | 1.957 | 0.861 | 3.053 | 0.559 | 3.499 | 0.026 | 4.69E-04 | 18721 |
| 57 | Occipital_Inf_L | -2.113 | -2.941 | -1.286 | 0.422 | -5.006 | -0.037 | 5.62E-07 | 18651 | 0.236 | -0.287 | 0.760 | 0.267 | 0.885 | 0.006 | 3.76E-01 | 18721 |
| 58 | Occipital_Inf_R | -1.511 | -2.467 | -0.555 | 0.488 | -3.098 | -0.023 | 1.95E-03 | 18651 | 0.245 | -0.360 | 0.849 | 0.308 | 0.793 | 0.006 | 4.28E-01 | 18721 |
| 59 | Fusiform_L | -2.428 | -4.238 | -0.617 | 0.924 | -2.628 | -0.019 | 8.61E-03 | 18651 | 3.107 | 1.962 | 4.251 | 0.584 | 5.322 | 0.039 | 1.04E-07 | 18721 |
| 60 | Fusiform_R | -3.162 | -5.154 | -1.170 | 1.016 | -3.112 | -0.023 | 1.86E-03 | 18651 | 5.141 | 3.884 | 6.398 | 0.641 | 8.016 | 0.058 | 1.16E-15 | 18721 |
| 61 | Postcentral_L | -7.623 | -10.345 | -4.902 | 1.388 | -5.490 | -0.040 | 4.06E-08 | 18651 | 7.094 | 5.375 | 8.814 | 0.877 | 8.085 | 0.059 | 6.57E-16 | 18721 |
| 62 | Postcentral_R | -5.550 | -8.343 | -2.757 | 1.425 | -3.895 | -0.029 | 9.84E-05 | 18651 | 7.144 | 5.381 | 8.907 | 0.899 | 7.942 | 0.058 | 2.10E-15 | 18721 |
| 63 | Parietal_Sup_L | -2.598 | -4.167 | -1.029 | 0.801 | -3.245 | -0.024 | 1.18E-03 | 18651 | 1.790 | 0.798 | 2.782 | 0.506 | 3.536 | 0.026 | 4.07E-04 | 18721 |
| 64 | Parietal_Sup_R | -3.023 | -4.415 | -1.631 | 0.710 | -4.258 | -0.031 | 2.08E-05 | 18651 | 1.204 | 0.324 | 2.084 | 0.449 | 2.680 | 0.020 | 7.36E-03 | 18721 |
| 65 | Parietal_Inf_L | -6.742 | -8.795 | -4.689 | 1.047 | -6.438 | -0.047 | 1.24E-10 | 18651 | 4.878 | 3.580 | 6.176 | 0.662 | 7.366 | 0.054 | 1.83E-13 | 18721 |
| 66 | Parietal_Inf_R | -2.919 | -4.250 | -1.588 | 0.679 | -4.298 | -0.031 | 1.73E-05 | 18651 | 2.570 | 1.729 | 3.412 | 0.429 | 5.989 | 0.044 | 2.15E-09 | 18721 |
| 67 | SupraMarginal_L | -3.021 | -4.224 | -1.817 | 0.614 | -4.920 | -0.036 | 8.74E-07 | 18651 | 2.756 | 1.995 | 3.517 | 0.388 | 7.102 | 0.052 | 1.27E-12 | 18721 |
| 68 | SupraMarginal_R | -3.328 | -5.032 | -1.624 | 0.869 | -3.828 | -0.028 | 1.30E-04 | 18651 | 4.504 | 3.429 | 5.580 | 0.549 | 8.209 | 0.060 | 2.37E-16 | 18721 |
| 69 | Angular_L | -3.233 | -4.416 | -2.051 | 0.603 | -5.360 | -0.039 | 8.42E-08 | 18651 | 1.565 | 0.817 | 2.313 | 0.382 | 4.101 | 0.030 | 4.14E-05 | 18721 |
| 70 | Angular_R | -3.816 | -5.402 | -2.230 | 0.809 | -4.716 | -0.035 | 2.43E-06 | 18651 | 2.409 | 1.406 | 3.411 | 0.512 | 4.708 | 0.034 | 2.52E-06 | 18721 |
| 71 | Precuneus_L | -5.492 | -8.375 | -2.610 | 1.471 | -3.735 | -0.027 | 1.88E-04 | 18651 | 4.042 | 2.220 | 5.864 | 0.929 | 4.349 | 0.032 | 1.38E-05 | 18721 |
| 72 | Precuneus_R | -5.532 | -8.100 | -2.964 | 1.310 | -4.223 | -0.031 | 2.42E-05 | 18651 | 4.085 | 2.462 | 5.708 | 0.828 | 4.934 | 0.036 | 8.13E-07 | 18721 |
| 73 | Paracentral_Lobule_L | -2.569 | -3.890 | -1.248 | 0.674 | -3.812 | -0.028 | 1.39E-04 | 18651 | 1.141 | 0.306 | 1.976 | 0.426 | 2.679 | 0.020 | 7.39E-03 | 18721 |
| 74 | Paracentral_Lobule_R | -1.341 | -2.287 | -0.395 | 0.483 | -2.778 | -0.020 | 5.48E-03 | 18651 | 1.266 | 0.668 | 1.864 | 0.305 | 4.151 | 0.030 | 3.32E-05 | 18721 |
| 75 | Caudate_L | -0.772 | -1.828 | 0.284 | 0.539 | -1.432 | -0.010 | 1.52E-01 | 18651 | 0.029 | -0.638 | 0.696 | 0.340 | 0.086 | 0.001 | 9.32E-01 | 18721 |
| 76 | Caudate_R | -1.609 | -2.804 | -0.414 | 0.610 | -2.640 | -0.019 | 8.30E-03 | 18651 | 0.214 | -0.541 | 0.969 | 0.385 | 0.555 | 0.004 | 5.79E-01 | 18721 |
| 77 | Putamen_L | -3.677 | -5.277 | -2.077 | 0.816 | -4.505 | -0.033 | 6.69E-06 | 18651 | 3.071 | 2.059 | 4.083 | 0.516 | 5.949 | 0.043 | 2.74E-09 | 18721 |
| 78 | Putamen_R | -4.450 | -6.080 | -2.820 | 0.831 | -5.352 | -0.039 | 8.78E-08 | 18651 | 3.302 | 2.271 | 4.332 | 0.526 | 6.280 | 0.046 | 3.46E-10 | 18721 |
| 79 | Pallidum_L | -0.131 | -0.274 | 0.011 | 0.073 | -1.808 | -0.013 | 7.06E-02 | 18651 | 0.112 | 0.022 | 0.202 | 0.046 | 2.433 | 0.018 | 1.50E-02 | 18721 |
| 80 | Pallidum_R | -0.204 | -0.360 | -0.048 | 0.080 | -2.559 | -0.019 | 1.05E-02 | 18651 | 0.141 | 0.043 | 0.240 | 0.050 | 2.806 | 0.021 | 5.02E-03 | 18721 |
| 83 | Heschl_L | -0.994 | -1.283 | -0.706 | 0.147 | -6.754 | -0.049 | 1.48E-11 | 18651 | 0.529 | 0.347 | 0.712 | 0.093 | 5.687 | 0.042 | 1.31E-08 | 18721 |
| 84 | Heschl_R | -0.899 | -1.222 | -0.577 | 0.165 | -5.466 | -0.040 | 4.65E-08 | 18651 | 0.618 | 0.414 | 0.822 | 0.104 | 5.945 | 0.043 | 2.82E-09 | 18721 |
| 85 | Temporal_Sup_L | -7.543 | -9.626 | -5.461 | 1.062 | -7.100 | -0.052 | 1.29E-12 | 18651 | 5.133 | 3.817 | 6.448 | 0.671 | 7.645 | 0.056 | 2.20E-14 | 18721 |
| 86 | Temporal_Sup_R | -7.410 | -9.714 | -5.106 | 1.175 | -6.304 | -0.046 | 2.96E-10 | 18651 | 5.444 | 3.988 | 6.900 | 0.743 | 7.329 | 0.053 | 2.41E-13 | 18721 |
| 87 | Temporal_Pole_Sup_L | -2.564 | -3.551 | -1.577 | 0.503 | -5.093 | -0.037 | 3.56E-07 | 18651 | 2.077 | 1.453 | 2.701 | 0.318 | 6.528 | 0.048 | 6.83E-11 | 18721 |
| 88 | Temporal_Pole_Sup_R | -3.248 | -4.287 | -2.210 | 0.530 | -6.131 | -0.045 | 8.91E-10 | 18651 | 2.469 | 1.813 | 3.126 | 0.335 | 7.372 | 0.054 | 1.75E-13 | 18721 |
| 89 | Temporal_Mid_L | -9.235 | -12.708 | -5.762 | 1.772 | -5.212 | -0.038 | 1.88E-07 | 18651 | 7.469 | 5.274 | 9.664 | 1.120 | 6.669 | 0.049 | 2.64E-11 | 18721 |
| 90 | Temporal_Mid_R | -9.776 | -12.858 | -6.693 | 1.573 | -6.216 | -0.045 | 5.22E-10 | 18651 | 5.979 | 4.029 | 7.928 | 0.994 | 6.012 | 0.044 | 1.87E-09 | 18721 |
| 91 | Temporal_Pole_Mid_L | -1.347 | -2.177 | -0.517 | 0.423 | -3.182 | -0.023 | 1.47E-03 | 18651 | 1.234 | 0.710 | 1.759 | 0.268 | 4.612 | 0.034 | 4.01E-06 | 18721 |
| 92 | Temporal_Pole_Mid_R | -2.092 | -3.100 | -1.084 | 0.514 | -4.068 | -0.030 | 4.77E-05 | 18651 | 2.088 | 1.451 | 2.725 | 0.325 | 6.425 | 0.047 | 1.35E-10 | 18721 |
| 93 | Temporal_Inf_L | -4.237 | -6.626 | -1.848 | 1.219 | -3.477 | -0.025 | 5.09E-04 | 18651 | 2.538 | 1.028 | 4.048 | 0.770 | 3.295 | 0.024 | 9.87E-04 | 18721 |
| 94 | Temporal_Inf_R | -5.124 | -7.728 | -2.519 | 1.329 | -3.856 | -0.028 | 1.15E-04 | 18651 | 1.914 | 0.268 | 3.561 | 0.840 | 2.279 | 0.017 | 2.27E-02 | 18721 |
| 95 | Cerebellum_Crus1_L | -1.480 | -4.316 | 1.355 | 1.447 | -1.023 | -0.007 | 3.06E-01 | 18651 | 3.036 | 1.243 | 4.829 | 0.915 | 3.320 | 0.024 | 9.03E-04 | 18721 |

| | | | | | | | | | | | | | | | | | |
|---|---|---|---|---|---|---|---|---|---|---|---|---|---|---|---|---|---|
| 96 | Cerebellum_Crus1_R | -1.197 | -4.090 | 1.695 | 1.476 | -0.811 | -0.006 | 4.17E-01 | 18651 | 3.521 | 1.693 | 5.349 | 0.933 | 3.776 | 0.028 | 1.60E-04 | 18721 |
| 97 | Cerebellum_Crus2_L | -0.579 | -2.831 | 1.672 | 1.149 | -0.504 | -0.004 | 6.14E-01 | 18651 | 4.651 | 3.231 | 6.072 | 0.725 | 6.416 | 0.047 | 1.43E-10 | 18721 |
| 98 | Cerebellum_Crus2_R | -0.303 | -2.385 | 1.779 | 1.062 | -0.285 | -0.002 | 7.75E-01 | 18651 | 3.972 | 2.658 | 5.287 | 0.671 | 5.922 | 0.043 | 3.23E-09 | 18721 |
| 99 | Cerebellum_3_L | 0.140 | -0.011 | 0.291 | 0.077 | 1.815 | 0.013 | 6.96E-02 | 18651 | 0.237 | 0.142 | 0.333 | 0.049 | 4.879 | 0.036 | 1.08E-06 | 18721 |
| 100 | Cerebellum_3_R | -0.030 | -0.234 | 0.174 | 0.104 | -0.291 | -0.002 | 7.71E-01 | 18651 | 0.290 | 0.162 | 0.419 | 0.066 | 4.421 | 0.032 | 9.87E-06 | 18721 |
| 101 | Cerebellum_4_5_L | 0.401 | -0.821 | 1.624 | 0.623 | 0.644 | 0.005 | 5.20E-01 | 18651 | 1.330 | 0.558 | 2.102 | 0.394 | 3.376 | 0.025 | 7.37E-04 | 18721 |
| 102 | Cerebellum_4_5_R | -0.182 | -1.177 | 0.814 | 0.508 | -0.358 | -0.003 | 7.20E-01 | 18651 | 1.622 | 0.993 | 2.251 | 0.321 | 5.056 | 0.037 | 4.31E-07 | 18721 |
| 103 | Cerebellum_6_L | -0.587 | -2.697 | 1.524 | 1.077 | -0.545 | -0.004 | 5.86E-01 | 18651 | 2.339 | 1.006 | 3.673 | 0.681 | 3.438 | 0.025 | 5.88E-04 | 18721 |
| 104 | Cerebellum_6_R | -0.952 | -3.175 | 1.272 | 1.135 | -0.839 | -0.006 | 4.02E-01 | 18651 | 3.947 | 2.542 | 5.352 | 0.717 | 5.507 | 0.040 | 3.69E-08 | 18721 |
| 105 | Cerebellum_7b_L | 0.069 | -0.619 | 0.758 | 0.351 | 0.198 | 0.001 | 8.43E-01 | 18651 | 1.480 | 1.045 | 1.914 | 0.222 | 6.676 | 0.049 | 2.52E-11 | 18721 |
| 106 | Cerebellum_7b_R | -0.101 | -0.783 | 0.581 | 0.348 | -0.290 | -0.002 | 7.72E-01 | 18651 | 1.747 | 1.317 | 2.177 | 0.220 | 7.957 | 0.058 | 1.86E-15 | 18721 |
| 107 | Cerebellum_8_L | 1.616 | -0.569 | 3.800 | 1.115 | 1.450 | 0.011 | 1.47E-01 | 18651 | 3.119 | 1.740 | 4.499 | 0.704 | 4.432 | 0.032 | 9.39E-06 | 18721 |
| 108 | Cerebellum_8_R | 1.964 | -0.878 | 4.807 | 1.450 | 1.355 | 0.010 | 1.75E-01 | 18651 | 6.173 | 4.378 | 7.967 | 0.916 | 6.741 | 0.049 | 1.62E-11 | 18721 |
| 109 | Cerebellum_9_L | 2.031 | 0.797 | 3.265 | 0.630 | 3.226 | 0.024 | 1.26E-03 | 18651 | 0.449 | -0.332 | 1.229 | 0.398 | 1.126 | 0.008 | 2.60E-01 | 18721 |
| 110 | Cerebellum_9_R | 2.072 | 0.859 | 3.285 | 0.619 | 3.348 | 0.025 | 8.15E-04 | 18651 | 0.611 | -0.157 | 1.378 | 0.392 | 1.560 | 0.011 | 1.19E-01 | 18721 |
| 111 | Cerebellum_10_L | 0.063 | -0.025 | 0.151 | 0.045 | 1.408 | 0.010 | 1.59E-01 | 18651 | -0.016 | -0.071 | 0.040 | 0.028 | -0.559 | -0.004 | 5.76E-01 | 18721 |
| 112 | Cerebellum_10_R | 0.061 | -0.030 | 0.152 | 0.046 | 1.314 | 0.010 | 1.89E-01 | 18651 | 0.036 | -0.022 | 0.093 | 0.029 | 1.218 | 0.009 | 2.23E-01 | 18721 |
| 113 | Vermis_1_2 | 0.030 | -0.027 | 0.087 | 0.029 | 1.035 | 0.008 | 3.01E-01 | 18651 | 0.104 | 0.068 | 0.140 | 0.018 | 5.692 | 0.042 | 1.28E-08 | 18721 |
| 114 | Vermis_3 | 0.149 | -0.078 | 0.375 | 0.116 | 1.288 | 0.009 | 1.98E-01 | 18651 | 0.411 | 0.268 | 0.554 | 0.073 | 5.630 | 0.041 | 1.82E-08 | 18721 |
| 115 | Vermis_4_5 | 0.747 | 0.082 | 1.413 | 0.340 | 2.201 | 0.016 | 2.77E-02 | 18651 | 0.531 | 0.110 | 0.951 | 0.214 | 2.475 | 0.018 | 1.33E-02 | 18721 |
| 116 | Vermis_6 | 0.385 | 0.015 | 0.756 | 0.189 | 2.039 | 0.015 | 4.14E-02 | 18651 | 0.103 | -0.131 | 0.337 | 0.119 | 0.866 | 0.006 | 3.87E-01 | 18721 |
| 117 | Vermis_7 | 0.268 | 0.058 | 0.477 | 0.107 | 2.504 | 0.018 | 1.23E-02 | 18651 | 0.098 | -0.034 | 0.231 | 0.068 | 1.457 | 0.011 | 1.45E-01 | 18721 |
| 118 | Vermis_8 | 0.552 | 0.208 | 0.896 | 0.176 | 3.145 | 0.023 | 1.66E-03 | 18651 | 0.042 | -0.176 | 0.259 | 0.111 | 0.374 | 0.003 | 7.08E-01 | 18721 |
| 119 | Vermis_9 | 0.481 | 0.203 | 0.760 | 0.142 | 3.384 | 0.025 | 7.15E-04 | 18651 | 0.127 | -0.050 | 0.303 | 0.090 | 1.409 | 0.010 | 1.59E-01 | 18721 |
| 120 | Vermis_10 | 0.082 | 0.004 | 0.160 | 0.040 | 2.065 | 0.015 | 3.90E-02 | 18651 | -0.005 | -0.054 | 0.045 | 0.025 | -0.180 | -0.001 | 8.57E-01 | 18721 |
| 121 | Thal_AV_L | -0.034 | -0.065 | -0.003 | 0.016 | -2.156 | -0.016 | 3.11E-02 | 18651 | 0.040 | 0.020 | 0.060 | 0.010 | 3.965 | 0.029 | 7.36E-05 | 18721 |
| 122 | Thal_AV_R | -0.057 | -0.101 | -0.012 | 0.023 | -2.504 | -0.018 | 1.23E-02 | 18651 | 0.061 | 0.033 | 0.089 | 0.014 | 4.248 | 0.031 | 2.17E-05 | 18721 |
| 123 | Thal_LP_L | -0.065 | -0.100 | -0.030 | 0.018 | -3.691 | -0.027 | 2.24E-04 | 18651 | 0.067 | 0.045 | 0.088 | 0.011 | 5.982 | 0.044 | 2.25E-09 | 18721 |
| 124 | Thal_LP_R | -0.127 | -0.172 | -0.082 | 0.023 | -5.529 | -0.040 | 3.27E-08 | 18651 | 0.100 | 0.071 | 0.128 | 0.014 | 6.874 | 0.050 | 6.44E-12 | 18721 |
| 125 | Thal_VA_L | 0.011 | -0.026 | 0.048 | 0.019 | 0.589 | 0.004 | 5.56E-01 | 18651 | -0.009 | -0.032 | 0.014 | 0.012 | -0.761 | -0.006 | 4.47E-01 | 18721 |
| 126 | Thal_VA_R | 0.013 | -0.033 | 0.058 | 0.023 | 0.544 | 0.004 | 5.86E-01 | 18651 | 0.012 | -0.016 | 0.041 | 0.015 | 0.848 | 0.006 | 3.97E-01 | 18721 |
| 127 | Thal_VL_L | -0.053 | -0.157 | 0.052 | 0.053 | -0.986 | -0.007 | 3.24E-01 | 18651 | 0.091 | 0.025 | 0.158 | 0.034 | 2.704 | 0.020 | 6.85E-03 | 18721 |
| 128 | Thal_VL_R | -0.227 | -0.366 | -0.087 | 0.071 | -3.189 | -0.023 | 1.43E-03 | 18651 | 0.244 | 0.156 | 0.332 | 0.045 | 5.434 | 0.040 | 5.59E-08 | 18721 |
| 129 | Thal_VPL_L | -0.313 | -0.438 | -0.189 | 0.064 | -4.927 | -0.036 | 8.43E-07 | 18651 | 0.184 | 0.106 | 0.263 | 0.040 | 4.585 | 0.033 | 4.58E-06 | 18721 |
| 130 | Thal_VPL_R | -0.400 | -0.531 | -0.269 | 0.067 | -5.986 | -0.044 | 2.19E-09 | 18651 | 0.231 | 0.148 | 0.314 | 0.042 | 5.461 | 0.040 | 4.78E-08 | 18721 |
| 131 | Thal_IL_L | -0.197 | -0.251 | -0.143 | 0.028 | -7.174 | -0.052 | 7.57E-13 | 18651 | 0.134 | 0.100 | 0.168 | 0.017 | 7.726 | 0.056 | 1.16E-14 | 18721 |
| 132 | Thal_IL_R | -0.248 | -0.310 | -0.187 | 0.031 | -7.964 | -0.058 | 1.76E-15 | 18651 | 0.154 | 0.115 | 0.192 | 0.020 | 7.793 | 0.057 | 6.90E-15 | 18721 |
| 133 | Thal_Re_L | 0.000 | 0.000 | 0.000 | 0.000 | 0.290 | 0.002 | 7.72E-01 | 18651 | 0.000 | 0.000 | 0.001 | 0.000 | 4.907 | 0.036 | 9.33E-07 | 18721 |
| 134 | Thal_Re_R | -0.002 | -0.004 | 0.000 | 0.001 | -2.476 | -0.018 | 1.33E-02 | 18651 | 0.003 | 0.002 | 0.005 | 0.001 | 5.858 | 0.043 | 4.76E-09 | 18721 |
| 135 | Thal_MDm_L | -1.729 | -2.129 | -1.330 | 0.204 | -8.491 | -0.062 | 2.20E-17 | 18651 | 1.118 | 0.865 | 1.370 | 0.129 | 8.682 | 0.063 | 4.20E-18 | 18721 |
| 136 | Thal_MDm_R | -2.073 | -2.537 | -1.610 | 0.237 | -8.762 | -0.064 | 2.07E-18 | 18651 | 1.206 | 0.912 | 1.499 | 0.150 | 8.057 | 0.059 | 8.30E-16 | 18721 |
| 137 | Thal_MDl_L | -0.263 | -0.332 | -0.193 | 0.035 | -7.439 | -0.054 | 1.06E-13 | 18651 | 0.191 | 0.148 | 0.235 | 0.022 | 8.581 | 0.063 | 1.02E-17 | 18721 |
| 138 | Thal_MDl_R | -0.337 | -0.415 | -0.258 | 0.040 | -8.404 | -0.061 | 4.61E-17 | 18651 | 0.205 | 0.155 | 0.255 | 0.025 | 8.092 | 0.059 | 6.20E-16 | 18721 |
| 139 | Thal_LGN_L | -0.012 | -0.023 | -0.002 | 0.005 | -2.308 | -0.017 | 2.10E-02 | 18651 | 0.023 | 0.016 | 0.029 | 0.003 | 6.752 | 0.049 | 1.50E-11 | 18721 |
| 140 | Thal_LGN_R | -0.039 | -0.061 | -0.016 | 0.011 | -3.406 | -0.025 | 6.60E-04 | 18651 | 0.057 | 0.043 | 0.071 | 0.007 | 7.923 | 0.058 | 2.45E-15 | 18721 |
| 141 | Thal_MGN_L | -0.052 | -0.071 | -0.033 | 0.010 | -5.429 | -0.040 | 5.74E-08 | 18651 | 0.051 | 0.039 | 0.063 | 0.006 | 8.425 | 0.061 | 3.87E-17 | 18721 |
| 142 | Thal_MGN_R | -0.059 | -0.080 | -0.039 | 0.010 | -5.714 | -0.042 | 1.12E-08 | 18651 | 0.052 | 0.039 | 0.065 | 0.007 | 7.903 | 0.058 | 2.88E-15 | 18721 |
| 143 | Thal_PuA_L | -0.066 | -0.089 | -0.044 | 0.011 | -5.890 | -0.043 | 3.92E-09 | 18651 | 0.060 | 0.046 | 0.074 | 0.007 | 8.453 | 0.062 | 3.04E-17 | 18721 |

| | | | | | | | | | | | | | | | | | |
|---|---|---|---|---|---|---|---|---|---|---|---|---|---|---|---|---|---|
| 144 | Thal_PuA_R | -0.147 | -0.193 | -0.101 | 0.023 | -6.271 | -0.046 | 3.68E-10 | 18651 | 0.123 | 0.094 | 0.153 | 0.015 | 8.330 | 0.061 | 8.65E-17 | 18721 |
| 145 | Thal_PuM_L | -1.689 | -2.128 | -1.249 | 0.224 | -7.534 | -0.055 | 5.15E-14 | 18651 | 1.138 | 0.860 | 1.415 | 0.142 | 8.026 | 0.059 | 1.06E-15 | 18721 |
| 146 | Thal_PuM_R | -2.111 | -2.659 | -1.564 | 0.279 | -7.559 | -0.055 | 4.24E-14 | 18651 | 1.360 | 1.014 | 1.706 | 0.177 | 7.700 | 0.056 | 1.42E-14 | 18721 |
| 147 | Thal_PuL_L | -0.326 | -0.406 | -0.246 | 0.041 | -7.968 | -0.058 | 1.71E-15 | 18651 | 0.197 | 0.147 | 0.248 | 0.026 | 7.630 | 0.056 | 2.45E-14 | 18721 |
| 148 | Thal_PuL_R | -0.526 | -0.653 | -0.398 | 0.065 | -8.082 | -0.059 | 6.74E-16 | 18651 | 0.301 | 0.220 | 0.382 | 0.041 | 7.315 | 0.053 | 2.69E-13 | 18721 |
| 149 | Thal_Pul_L | -0.043 | -0.065 | -0.021 | 0.011 | -3.864 | -0.028 | 1.12E-04 | 18651 | 0.023 | 0.009 | 0.037 | 0.007 | 3.286 | 0.024 | 1.02E-03 | 18721 |
| 150 | Thal_Pul_R | -0.062 | -0.091 | -0.033 | 0.015 | -4.224 | -0.031 | 2.41E-05 | 18651 | 0.037 | 0.019 | 0.056 | 0.009 | 3.999 | 0.029 | 6.39E-05 | 18721 |
| 151 | ACC_sub_L | -0.527 | -0.778 | -0.275 | 0.128 | -4.102 | -0.030 | 4.11E-05 | 18651 | 0.430 | 0.271 | 0.589 | 0.081 | 5.298 | 0.039 | 1.18E-07 | 18721 |
| 152 | ACC_sub_R | -0.475 | -0.710 | -0.240 | 0.120 | -3.959 | -0.029 | 7.57E-05 | 18651 | 0.315 | 0.166 | 0.464 | 0.076 | 4.148 | 0.030 | 3.37E-05 | 18721 |
| 153 | ACC_pre_L | -1.970 | -2.615 | -1.325 | 0.329 | -5.983 | -0.044 | 2.23E-09 | 18651 | 1.481 | 1.073 | 1.889 | 0.208 | 7.119 | 0.052 | 1.12E-12 | 18721 |
| 154 | ACC_pre_R | -1.626 | -2.247 | -1.004 | 0.317 | -5.126 | -0.038 | 2.99E-07 | 18651 | 1.268 | 0.875 | 1.660 | 0.200 | 6.326 | 0.046 | 2.57E-10 | 18721 |
| 155 | ACC_sup_L | -2.337 | -3.038 | -1.636 | 0.358 | -6.535 | -0.048 | 6.54E-11 | 18651 | 1.646 | 1.203 | 2.089 | 0.226 | 7.281 | 0.053 | 3.45E-13 | 18721 |
| 156 | ACC_sup_R | -1.553 | -2.280 | -0.827 | 0.371 | -4.191 | -0.031 | 2.79E-05 | 18651 | 0.961 | 0.502 | 1.421 | 0.234 | 4.101 | 0.030 | 4.12E-05 | 18721 |
| 157 | N_Acc_L | -0.306 | -0.517 | -0.095 | 0.108 | -2.840 | -0.021 | 4.51E-03 | 18651 | 0.362 | 0.229 | 0.496 | 0.068 | 5.321 | 0.039 | 1.04E-07 | 18721 |
| 158 | N_Acc_R | -0.331 | -0.513 | -0.148 | 0.093 | -3.546 | -0.026 | 3.92E-04 | 18651 | 0.248 | 0.133 | 0.364 | 0.059 | 4.213 | 0.031 | 2.54E-05 | 18721 |
| 159 | VTA_L | -0.012 | -0.019 | -0.005 | 0.004 | -3.185 | -0.023 | 1.45E-03 | 18651 | 0.014 | 0.010 | 0.019 | 0.002 | 5.931 | 0.043 | 3.07E-09 | 18721 |
| 160 | VTA_R | -0.009 | -0.016 | -0.003 | 0.003 | -2.695 | -0.020 | 7.04E-03 | 18651 | 0.015 | 0.011 | 0.019 | 0.002 | 7.057 | 0.052 | 1.76E-12 | 18721 |
| 161 | SN_pc_L | -0.015 | -0.027 | -0.003 | 0.006 | -2.493 | -0.018 | 1.27E-02 | 18651 | 0.017 | 0.010 | 0.025 | 0.004 | 4.553 | 0.033 | 5.32E-06 | 18721 |
| 162 | SN_pc_R | -0.029 | -0.044 | -0.013 | 0.008 | -3.595 | -0.026 | 3.26E-04 | 18651 | 0.025 | 0.015 | 0.035 | 0.005 | 4.978 | 0.036 | 6.48E-07 | 18721 |
| 163 | SN_pr_L | -0.005 | -0.022 | 0.011 | 0.008 | -0.652 | -0.005 | 5.14E-01 | 18651 | 0.014 | 0.004 | 0.025 | 0.005 | 2.775 | 0.020 | 5.52E-03 | 18721 |
| 164 | SN_pr_R | -0.026 | -0.044 | -0.008 | 0.009 | -2.795 | -0.020 | 5.20E-03 | 18651 | 0.015 | 0.004 | 0.027 | 0.006 | 2.615 | 0.019 | 8.92E-03 | 18721 |
| 165 | Red_N_L | -0.021 | -0.041 | -0.001 | 0.010 | -2.031 | -0.015 | 4.23E-02 | 18651 | 0.042 | 0.029 | 0.055 | 0.006 | 6.525 | 0.048 | 6.97E-11 | 18721 |
| 166 | Red_N_R | -0.015 | -0.037 | 0.006 | 0.011 | -1.396 | -0.010 | 1.63E-01 | 18651 | 0.044 | 0.031 | 0.058 | 0.007 | 6.329 | 0.046 | 2.52E-10 | 18721 |
| 167 | LC_L | 0.004 | -0.004 | 0.011 | 0.004 | 1.038 | 0.008 | 2.99E-01 | 18651 | 0.007 | 0.002 | 0.011 | 0.002 | 2.788 | 0.020 | 5.31E-03 | 18721 |
| 168 | LC_R | 0.001 | -0.007 | 0.010 | 0.004 | 0.352 | 0.003 | 7.25E-01 | 18651 | 0.009 | 0.004 | 0.014 | 0.003 | 3.349 | 0.024 | 8.14E-04 | 18721 |
| 169 | Raphe_D | -0.050 | -0.079 | -0.021 | 0.015 | -3.380 | -0.025 | 7.25E-04 | 18651 | 0.087 | 0.069 | 0.105 | 0.009 | 9.351 | 0.068 | 9.67E-21 | 18721 |
| 170 | Raphe_M | -0.006 | -0.015 | 0.003 | 0.004 | -1.324 | -0.010 | 1.86E-01 | 18651 | 0.019 | 0.014 | 0.025 | 0.003 | 6.827 | 0.050 | 8.96E-12 | 18721 |

| | | Tea intake | | | | | | | | Water intake | | | | | | | |
|---|---|---|---|---|---|---|---|---|---|---|---|---|---|---|---|---|---|
| Number | AAL 3 region | beta | 95%CI_l | 95%CI_h | se | t | r | p | df.e | beta | 95%CI_l | 95%CI_h | se | t | r | p | df.e |
| 0 | Total brain | 64.706 | 11.326 | 118.087 | 27.234 | 2.376 | 0.017 | 1.75E-02 | 18672 | -160.661 | -231.380 | -89.943 | 36.079 | -4.453 | -0.033 | 8.52E-06 | 18637 |
| 1 | Precentral_L | 1.057 | -0.822 | 2.935 | 0.958 | 1.103 | 0.008 | 2.70E-01 | 18672 | -1.198 | -3.691 | 1.294 | 1.272 | -0.943 | -0.007 | 3.46E-01 | 18637 |
| 2 | Precentral_R | 1.623 | -0.099 | 3.345 | 0.879 | 1.848 | 0.014 | 6.47E-02 | 18672 | -2.650 | -4.934 | -0.367 | 1.165 | -2.275 | -0.017 | 2.29E-02 | 18637 |
| 3 | Frontal_Sup_2_L | 1.161 | -0.822 | 3.144 | 1.012 | 1.148 | 0.008 | 2.51E-01 | 18672 | -2.270 | -4.901 | 0.360 | 1.342 | -1.692 | -0.012 | 9.07E-02 | 18637 |
| 4 | Frontal_Sup_2_R | 2.501 | 0.315 | 4.687 | 1.115 | 2.243 | 0.016 | 2.49E-02 | 18672 | -1.645 | -4.546 | 1.255 | 1.480 | -1.112 | -0.008 | 2.66E-01 | 18637 |
| 5 | Frontal_Mid_2_L | 1.369 | -0.714 | 3.453 | 1.063 | 1.288 | 0.009 | 1.98E-01 | 18672 | -0.767 | -3.530 | 1.996 | 1.409 | -0.544 | -0.004 | 5.86E-01 | 18637 |
| 6 | Frontal_Mid_2_R | 1.265 | -1.092 | 3.622 | 1.202 | 1.052 | 0.008 | 2.93E-01 | 18672 | -1.688 | -4.812 | 1.436 | 1.594 | -1.059 | -0.008 | 2.90E-01 | 18637 |
| 7 | Frontal_Inf_Oper_L | 0.181 | -0.435 | 0.796 | 0.314 | 0.575 | 0.004 | 5.65E-01 | 18672 | -0.692 | -1.508 | 0.125 | 0.417 | -1.660 | -0.012 | 9.68E-02 | 18637 |
| 8 | Frontal_Inf_Oper_R | 0.288 | -0.484 | 1.059 | 0.394 | 0.730 | 0.005 | 4.65E-01 | 18672 | -1.215 | -2.238 | -0.192 | 0.522 | -2.327 | -0.017 | 2.00E-02 | 18637 |
| 9 | Frontal_Inf_Tri_L | 0.915 | -0.393 | 2.222 | 0.667 | 1.371 | 0.010 | 1.70E-01 | 18672 | -1.906 | -3.639 | -0.172 | 0.884 | -2.155 | -0.016 | 3.12E-02 | 18637 |
| 10 | Frontal_Inf_Tri_R | 0.827 | -0.199 | 1.853 | 0.524 | 1.580 | 0.012 | 1.14E-01 | 18672 | -1.205 | -2.566 | 0.156 | 0.694 | -1.736 | -0.013 | 8.26E-02 | 18637 |
| 11 | Frontal_Inf_Orb_2_L | -0.044 | -0.436 | 0.347 | 0.199 | -0.223 | -0.002 | 8.23E-01 | 18672 | -0.265 | -0.784 | 0.253 | 0.264 | -1.004 | -0.007 | 3.15E-01 | 18637 |
| 12 | Frontal_Inf_Orb_2_R | -0.040 | -0.449 | 0.369 | 0.209 | -0.191 | -0.001 | 8.49E-01 | 18672 | -0.600 | -1.142 | -0.058 | 0.276 | -2.170 | -0.016 | 3.00E-02 | 18637 |
| 13 | Rolandic_Oper_L | 0.359 | -0.232 | 0.951 | 0.302 | 1.190 | 0.009 | 2.34E-01 | 18672 | -1.191 | -1.975 | -0.408 | 0.400 | -2.980 | -0.022 | 2.89E-03 | 18637 |
| 14 | Rolandic_Oper_R | 1.460 | 0.615 | 2.305 | 0.431 | 3.386 | 0.025 | 7.10E-04 | 18672 | -1.433 | -2.554 | -0.312 | 0.572 | -2.505 | -0.018 | 1.23E-02 | 18637 |
| 15 | Supp_Motor_Area_L | 0.519 | -0.886 | 1.925 | 0.717 | 0.724 | 0.005 | 4.69E-01 | 18672 | 1.022 | -0.842 | 2.885 | 0.951 | 1.075 | 0.008 | 2.83E-01 | 18637 |
| 16 | Supp_Motor_Area_R | 0.524 | -1.028 | 2.077 | 0.792 | 0.662 | 0.005 | 5.08E-01 | 18672 | 1.440 | -0.619 | 3.499 | 1.050 | 1.371 | 0.010 | 1.71E-01 | 18637 |
| 17 | Olfactory_L | -0.060 | -0.269 | 0.149 | 0.107 | -0.560 | -0.004 | 5.75E-01 | 18672 | -0.226 | -0.503 | 0.051 | 0.141 | -1.598 | -0.012 | 1.10E-01 | 18637 |
| 18 | Olfactory_R | -0.075 | -0.284 | 0.134 | 0.107 | -0.701 | -0.005 | 4.83E-01 | 18672 | -0.304 | -0.581 | -0.026 | 0.142 | -2.145 | -0.016 | 3.19E-02 | 18637 |

| | | | | | | | | | | | | | | | | | |
|---|---|---|---|---|---|---|---|---|---|---|---|---|---|---|---|---|---|
| 19 | Frontal_Sup_Medial_L | 0.278 | -1.039 | 1.595 | 0.672 | 0.413 | 0.003 | 6.79E-01 | 18672 | -0.817 | -2.563 | 0.929 | 0.891 | -0.918 | -0.007 | 3.59E-01 | 18637 |
| 20 | Frontal_Sup_Medial_R | 0.311 | -0.648 | 1.270 | 0.489 | 0.636 | 0.005 | 5.25E-01 | 18672 | -1.338 | -2.610 | -0.067 | 0.649 | -2.064 | -0.015 | 3.91E-02 | 18637 |
| 21 | Frontal_Med_Orb_L | 0.374 | -0.118 | 0.866 | 0.251 | 1.490 | 0.011 | 1.36E-01 | 18672 | -0.588 | -1.240 | 0.064 | 0.333 | -1.769 | -0.013 | 7.70E-02 | 18637 |
| 22 | Frontal_Med_Orb_R | 0.555 | -0.050 | 1.159 | 0.308 | 1.798 | 0.013 | 7.21E-02 | 18672 | -0.929 | -1.729 | -0.128 | 0.408 | -2.273 | -0.017 | 2.30E-02 | 18637 |
| 23 | Rectus_L | 0.598 | 0.089 | 1.107 | 0.260 | 2.301 | 0.017 | 2.14E-02 | 18672 | -0.642 | -1.316 | 0.032 | 0.344 | -1.866 | -0.014 | 6.21E-02 | 18637 |
| 24 | Rectus_R | 0.488 | 0.017 | 0.959 | 0.240 | 2.029 | 0.015 | 4.24E-02 | 18672 | -0.593 | -1.217 | 0.031 | 0.318 | -1.862 | -0.014 | 6.25E-02 | 18637 |
| 25 | OFCmed_L | 0.167 | -0.128 | 0.462 | 0.151 | 1.110 | 0.008 | 2.67E-01 | 18672 | -0.389 | -0.780 | 0.002 | 0.200 | -1.949 | -0.014 | 5.13E-02 | 18637 |
| 26 | OFCmed_R | -0.011 | -0.355 | 0.333 | 0.175 | -0.065 | 0.000 | 9.49E-01 | 18672 | -0.450 | -0.906 | 0.006 | 0.233 | -1.935 | -0.014 | 5.29E-02 | 18637 |
| 27 | OFCant_L | 0.148 | -0.194 | 0.489 | 0.174 | 0.848 | 0.006 | 3.97E-01 | 18672 | -0.433 | -0.885 | 0.020 | 0.231 | -1.875 | -0.014 | 6.08E-02 | 18637 |
| 28 | OFCant_R | 0.150 | -0.301 | 0.601 | 0.230 | 0.652 | 0.005 | 5.14E-01 | 18672 | -1.048 | -1.646 | -0.450 | 0.305 | -3.436 | -0.025 | 5.92E-04 | 18637 |
| 29 | OFCpost_L | 0.030 | -0.398 | 0.457 | 0.218 | 0.135 | 0.001 | 8.92E-01 | 18672 | -0.525 | -1.092 | 0.041 | 0.289 | -1.817 | -0.013 | 6.93E-02 | 18637 |
| 30 | OFCpost_R | 0.090 | -0.305 | 0.485 | 0.201 | 0.446 | 0.003 | 6.56E-01 | 18672 | -0.866 | -1.390 | -0.342 | 0.267 | -3.240 | -0.024 | 1.20E-03 | 18637 |
| 31 | OFClat_L | -0.021 | -0.166 | 0.123 | 0.074 | -0.291 | -0.002 | 7.71E-01 | 18672 | -0.065 | -0.256 | 0.126 | 0.098 | -0.664 | -0.005 | 5.07E-01 | 18637 |
| 32 | OFClat_R | -0.002 | -0.124 | 0.119 | 0.062 | -0.039 | 0.000 | 9.69E-01 | 18672 | -0.334 | -0.496 | -0.173 | 0.082 | -4.056 | -0.030 | 5.02E-05 | 18637 |
| 33 | Insula_L | 0.368 | -0.954 | 1.690 | 0.674 | 0.545 | 0.004 | 5.85E-01 | 18672 | -2.928 | -4.679 | -1.177 | 0.893 | -3.277 | -0.024 | 1.05E-03 | 18637 |
| 34 | Insula_R | 0.730 | -0.558 | 2.018 | 0.657 | 1.111 | 0.008 | 2.67E-01 | 18672 | -3.521 | -5.227 | -1.815 | 0.870 | -4.046 | -0.030 | 5.24E-05 | 18637 |
| 37 | Cingulate_Mid_L | 1.664 | 0.548 | 2.781 | 0.570 | 2.921 | 0.021 | 3.49E-03 | 18672 | -1.340 | -2.820 | 0.140 | 0.755 | -1.775 | -0.013 | 7.60E-02 | 18637 |
| 38 | Cingulate_Mid_R | 1.740 | 0.549 | 2.932 | 0.608 | 2.863 | 0.021 | 4.20E-03 | 18672 | -1.464 | -3.043 | 0.115 | 0.805 | -1.818 | -0.013 | 6.91E-02 | 18637 |
| 39 | Cingulate_Post_L | 0.260 | -0.082 | 0.601 | 0.174 | 1.490 | 0.011 | 1.36E-01 | 18672 | -0.699 | -1.151 | -0.246 | 0.231 | -3.024 | -0.022 | 2.49E-03 | 18637 |
| 40 | Cingulate_Post_R | 0.133 | -0.056 | 0.323 | 0.097 | 1.380 | 0.010 | 1.68E-01 | 18672 | -0.235 | -0.486 | 0.016 | 0.128 | -1.833 | -0.013 | 6.68E-02 | 18637 |
| 41 | Hippocampus_L | -0.086 | -0.638 | 0.466 | 0.282 | -0.305 | -0.002 | 7.60E-01 | 18672 | -1.293 | -2.025 | -0.561 | 0.374 | -3.461 | -0.025 | 5.40E-04 | 18637 |
| 42 | Hippocampus_R | -0.311 | -0.884 | 0.261 | 0.292 | -1.065 | -0.008 | 2.87E-01 | 18672 | -0.712 | -1.472 | 0.047 | 0.388 | -1.838 | -0.013 | 6.60E-02 | 18637 |
| 43 | ParaHippocampal_L | 0.095 | -0.395 | 0.586 | 0.250 | 0.381 | 0.003 | 7.03E-01 | 18672 | -1.052 | -1.702 | -0.402 | 0.332 | -3.172 | -0.023 | 1.52E-03 | 18637 |
| 44 | ParaHippocampal_R | 0.142 | -0.477 | 0.760 | 0.316 | 0.449 | 0.003 | 6.54E-01 | 18672 | -1.392 | -2.213 | -0.572 | 0.419 | -3.327 | -0.024 | 8.80E-04 | 18637 |
| 45 | Amygdala_L | -0.016 | -0.206 | 0.175 | 0.097 | -0.164 | -0.001 | 8.70E-01 | 18672 | -0.556 | -0.808 | -0.303 | 0.129 | -4.313 | -0.032 | 1.62E-05 | 18637 |
| 46 | Amygdala_R | -0.023 | -0.218 | 0.173 | 0.100 | -0.226 | -0.002 | 8.21E-01 | 18672 | -0.634 | -0.892 | -0.375 | 0.132 | -4.806 | -0.035 | 1.55E-06 | 18637 |
| 47 | Calcarine_L | 0.370 | -1.449 | 2.189 | 0.928 | 0.399 | 0.003 | 6.90E-01 | 18672 | -0.325 | -2.736 | 2.086 | 1.230 | -0.264 | -0.002 | 7.92E-01 | 18637 |
| 48 | Calcarine_R | 0.894 | -0.626 | 2.414 | 0.776 | 1.153 | 0.008 | 2.49E-01 | 18672 | -0.626 | -2.642 | 1.390 | 1.028 | -0.608 | -0.004 | 5.43E-01 | 18637 |
| 49 | Cuneus_L | 0.349 | -0.792 | 1.489 | 0.582 | 0.599 | 0.004 | 5.49E-01 | 18672 | -0.665 | -2.178 | 0.849 | 0.772 | -0.861 | -0.006 | 3.89E-01 | 18637 |
| 50 | Cuneus_R | 0.442 | -0.431 | 1.314 | 0.445 | 0.992 | 0.007 | 3.21E-01 | 18672 | -1.257 | -2.416 | -0.099 | 0.591 | -2.128 | -0.016 | 3.34E-02 | 18637 |
| 51 | Lingual_L | 0.743 | -0.351 | 1.836 | 0.558 | 1.332 | 0.010 | 1.83E-01 | 18672 | -1.109 | -2.560 | 0.342 | 0.740 | -1.498 | -0.011 | 1.34E-01 | 18637 |
| 52 | Lingual_R | 1.086 | -0.060 | 2.232 | 0.585 | 1.858 | 0.014 | 6.32E-02 | 18672 | -1.866 | -3.385 | -0.347 | 0.775 | -2.408 | -0.018 | 1.61E-02 | 18637 |
| 53 | Occipital_Sup_L | 1.623 | 0.871 | 2.374 | 0.383 | 4.233 | 0.031 | 2.31E-05 | 18672 | -1.491 | -2.488 | -0.494 | 0.509 | -2.931 | -0.021 | 3.38E-03 | 18637 |
| 54 | Occipital_Sup_R | 1.008 | 0.193 | 1.823 | 0.416 | 2.425 | 0.018 | 1.53E-02 | 18672 | -0.929 | -2.011 | 0.152 | 0.552 | -1.685 | -0.012 | 9.21E-02 | 18637 |
| 55 | Occipital_Mid_L | 0.962 | -0.800 | 2.724 | 0.899 | 1.070 | 0.008 | 2.85E-01 | 18672 | -4.061 | -6.397 | -1.726 | 1.192 | -3.409 | -0.025 | 6.55E-04 | 18637 |
| 56 | Occipital_Mid_R | 1.508 | 0.282 | 2.734 | 0.625 | 2.411 | 0.018 | 1.59E-02 | 18672 | -1.392 | -3.019 | 0.235 | 0.830 | -1.677 | -0.012 | 9.36E-02 | 18637 |
| 57 | Occipital_Inf_L | 0.302 | -0.283 | 0.886 | 0.298 | 1.011 | 0.007 | 3.12E-01 | 18672 | -1.134 | -1.910 | -0.359 | 0.396 | -2.867 | -0.021 | 4.15E-03 | 18637 |
| 58 | Occipital_Inf_R | 0.542 | -0.133 | 1.218 | 0.345 | 1.573 | 0.012 | 1.16E-01 | 18672 | -1.015 | -1.911 | -0.119 | 0.457 | -2.219 | -0.016 | 2.65E-02 | 18637 |
| 59 | Fusiform_L | 1.379 | 0.099 | 2.659 | 0.653 | 2.111 | 0.015 | 3.48E-02 | 18672 | -2.638 | -4.334 | -0.942 | 0.865 | -3.049 | -0.022 | 2.30E-03 | 18637 |
| 60 | Fusiform_R | 1.245 | -0.163 | 2.653 | 0.718 | 1.734 | 0.013 | 8.30E-02 | 18672 | -2.328 | -4.194 | -0.463 | 0.952 | -2.446 | -0.018 | 1.44E-02 | 18637 |
| 61 | Postcentral_L | 2.865 | 0.938 | 4.791 | 0.983 | 2.915 | 0.021 | 3.56E-03 | 18672 | -3.135 | -5.688 | -0.581 | 1.303 | -2.406 | -0.018 | 1.61E-02 | 18637 |
| 62 | Postcentral_R | 2.213 | 0.238 | 4.188 | 1.008 | 2.196 | 0.016 | 2.81E-02 | 18672 | -2.163 | -4.781 | 0.456 | 1.336 | -1.619 | -0.012 | 1.05E-01 | 18637 |
| 63 | Parietal_Sup_L | 0.097 | -1.013 | 1.207 | 0.566 | 0.172 | 0.001 | 8.63E-01 | 18672 | -0.350 | -1.822 | 1.122 | 0.751 | -0.466 | -0.003 | 6.41E-01 | 18637 |
| 64 | Parietal_Sup_R | -0.025 | -1.010 | 0.960 | 0.502 | -0.050 | 0.000 | 9.60E-01 | 18672 | 0.075 | -1.231 | 1.381 | 0.666 | 0.112 | 0.001 | 9.10E-01 | 18637 |
| 65 | Parietal_Inf_L | 2.061 | 0.608 | 3.515 | 0.741 | 2.780 | 0.020 | 5.44E-03 | 18672 | -1.336 | -3.263 | 0.590 | 0.983 | -1.360 | -0.010 | 1.74E-01 | 18637 |
| 66 | Parietal_Inf_R | 0.744 | -0.198 | 1.685 | 0.480 | 1.548 | 0.011 | 1.22E-01 | 18672 | -1.142 | -2.390 | 0.105 | 0.637 | -1.795 | -0.013 | 7.27E-02 | 18637 |
| 67 | SupraMarginal_L | 1.112 | 0.260 | 1.963 | 0.434 | 2.560 | 0.019 | 1.05E-02 | 18672 | -1.063 | -2.193 | 0.066 | 0.576 | -1.845 | -0.014 | 6.50E-02 | 18637 |
| 68 | SupraMarginal_R | 1.245 | 0.040 | 2.450 | 0.615 | 2.026 | 0.015 | 4.28E-02 | 18672 | -3.827 | -5.424 | -2.231 | 0.814 | -4.700 | -0.034 | 2.62E-06 | 18637 |

| | | | | | | | | | | | | | | | | | |
|---|---|---|---|---|---|---|---|---|---|---|---|---|---|---|---|---|---|
| 69 | Angular_L | 0.695 | -0.142 | 1.531 | 0.427 | 1.628 | 0.012 | 1.04E-01 | 18672 | -0.213 | -1.322 | 0.896 | 0.566 | -0.376 | -0.003 | 7.07E-01 | 18637 |
| 70 | Angular_R | 0.925 | -0.196 | 2.047 | 0.572 | 1.617 | 0.012 | 1.06E-01 | 18672 | 0.013 | -1.475 | 1.501 | 0.759 | 0.017 | 0.000 | 9.86E-01 | 18637 |
| 71 | Precuneus_L | 0.784 | -1.255 | 2.823 | 1.040 | 0.754 | 0.006 | 4.51E-01 | 18672 | -1.049 | -3.751 | 1.653 | 1.378 | -0.761 | -0.006 | 4.47E-01 | 18637 |
| 72 | Precuneus_R | 0.905 | -0.912 | 2.721 | 0.927 | 0.976 | 0.007 | 3.29E-01 | 18672 | -2.415 | -4.821 | -0.009 | 1.228 | -1.967 | -0.014 | 4.92E-02 | 18637 |
| 73 | Paracentral_Lobule_L | -0.009 | -0.942 | 0.925 | 0.476 | -0.018 | 0.000 | 9.85E-01 | 18672 | 1.463 | 0.225 | 2.702 | 0.632 | 2.316 | 0.017 | 2.06E-02 | 18637 |
| 74 | Paracentral_Lobule_R | 0.056 | -0.612 | 0.725 | 0.341 | 0.165 | 0.001 | 8.69E-01 | 18672 | 0.363 | -0.523 | 1.250 | 0.452 | 0.803 | 0.006 | 4.22E-01 | 18637 |
| 75 | Caudate_L | -0.677 | -1.423 | 0.069 | 0.380 | -1.780 | -0.013 | 7.51E-02 | 18672 | 0.094 | -0.895 | 1.082 | 0.504 | 0.185 | 0.001 | 8.53E-01 | 18637 |
| 76 | Caudate_R | -1.022 | -1.866 | -0.178 | 0.430 | -2.374 | -0.017 | 1.76E-02 | 18672 | 0.450 | -0.669 | 1.568 | 0.571 | 0.788 | 0.006 | 4.31E-01 | 18637 |
| 77 | Putamen_L | -1.893 | -3.026 | -0.761 | 0.578 | -3.278 | -0.024 | 1.05E-03 | 18672 | -2.619 | -4.120 | -1.117 | 0.766 | -3.419 | -0.025 | 6.30E-04 | 18637 |
| 78 | Putamen_R | -1.829 | -2.982 | -0.676 | 0.588 | -3.108 | -0.023 | 1.89E-03 | 18672 | -2.335 | -3.865 | -0.806 | 0.780 | -2.992 | -0.022 | 2.77E-03 | 18637 |
| 79 | Pallidum_L | -0.152 | -0.253 | -0.051 | 0.051 | -2.961 | -0.022 | 3.07E-03 | 18672 | -0.067 | -0.201 | 0.066 | 0.068 | -0.989 | -0.007 | 3.23E-01 | 18637 |
| 80 | Pallidum_R | -0.215 | -0.326 | -0.105 | 0.056 | -3.822 | -0.028 | 1.33E-04 | 18672 | -0.078 | -0.224 | 0.069 | 0.075 | -1.042 | -0.008 | 2.97E-01 | 18637 |
| 83 | Heschl_L | 0.262 | 0.058 | 0.467 | 0.104 | 2.519 | 0.018 | 1.18E-02 | 18672 | -0.451 | -0.722 | -0.180 | 0.138 | -3.265 | -0.024 | 1.10E-03 | 18637 |
| 84 | Heschl_R | 0.438 | 0.210 | 0.666 | 0.116 | 3.769 | 0.028 | 1.65E-04 | 18672 | -0.618 | -0.920 | -0.315 | 0.154 | -4.005 | -0.029 | 6.22E-05 | 18637 |
| 85 | Temporal_Sup_L | 2.096 | 0.622 | 3.570 | 0.752 | 2.788 | 0.020 | 5.31E-03 | 18672 | -3.855 | -5.810 | -1.900 | 0.997 | -3.866 | -0.028 | 1.11E-04 | 18637 |
| 86 | Temporal_Sup_R | 1.571 | -0.060 | 3.202 | 0.832 | 1.887 | 0.014 | 5.91E-02 | 18672 | -5.537 | -7.698 | -3.376 | 1.103 | -5.022 | -0.037 | 5.16E-07 | 18637 |
| 87 | Temporal_Pole_Sup_L | 0.349 | -0.349 | 1.047 | 0.356 | 0.979 | 0.007 | 3.28E-01 | 18672 | -1.570 | -2.496 | -0.645 | 0.472 | -3.327 | -0.024 | 8.80E-04 | 18637 |
| 88 | Temporal_Pole_Sup_R | 0.439 | -0.297 | 1.174 | 0.375 | 1.169 | 0.009 | 2.42E-01 | 18672 | -1.399 | -2.373 | -0.425 | 0.497 | -2.814 | -0.021 | 4.89E-03 | 18637 |
| 89 | Temporal_Mid_L | 2.311 | -0.145 | 4.768 | 1.253 | 1.844 | 0.013 | 6.52E-02 | 18672 | -6.087 | -9.342 | -2.832 | 1.661 | -3.665 | -0.027 | 2.48E-04 | 18637 |
| 90 | Temporal_Mid_R | 2.304 | 0.122 | 4.485 | 1.113 | 2.070 | 0.015 | 3.85E-02 | 18672 | -6.033 | -8.922 | -3.144 | 1.474 | -4.093 | -0.030 | 4.27E-05 | 18637 |
| 91 | Temporal_Pole_Mid_L | 0.732 | 0.146 | 1.318 | 0.299 | 2.447 | 0.018 | 1.44E-02 | 18672 | -0.574 | -1.351 | 0.204 | 0.397 | -1.445 | -0.011 | 1.48E-01 | 18637 |
| 92 | Temporal_Pole_Mid_R | 1.252 | 0.540 | 1.965 | 0.364 | 3.444 | 0.025 | 5.74E-04 | 18672 | -1.441 | -2.386 | -0.495 | 0.482 | -2.987 | -0.022 | 2.82E-03 | 18637 |
| 93 | Temporal_Inf_L | 0.521 | -1.167 | 2.210 | 0.861 | 0.605 | 0.004 | 5.45E-01 | 18672 | -3.898 | -6.135 | -1.660 | 1.142 | -3.414 | -0.025 | 6.42E-04 | 18637 |
| 94 | Temporal_Inf_R | 1.377 | -0.464 | 3.218 | 0.939 | 1.466 | 0.011 | 1.43E-01 | 18672 | -4.749 | -7.189 | -2.309 | 1.245 | -3.816 | -0.028 | 1.36E-04 | 18637 |
| 95 | Cerebellum_Crus1_L | -0.325 | -2.330 | 1.679 | 1.023 | -0.318 | -0.002 | 7.50E-01 | 18672 | -1.095 | -3.753 | 1.564 | 1.356 | -0.807 | -0.006 | 4.20E-01 | 18637 |
| 96 | Cerebellum_Crus1_R | 0.014 | -2.030 | 2.059 | 1.043 | 0.014 | 0.000 | 9.89E-01 | 18672 | -1.602 | -4.315 | 1.110 | 1.384 | -1.158 | -0.008 | 2.47E-01 | 18637 |
| 97 | Cerebellum_Crus2_L | 0.845 | -0.746 | 2.435 | 0.811 | 1.041 | 0.008 | 2.98E-01 | 18672 | -3.376 | -5.483 | -1.270 | 1.075 | -3.142 | -0.023 | 1.68E-03 | 18637 |
| 98 | Cerebellum_Crus2_R | 0.709 | -0.762 | 2.180 | 0.751 | 0.945 | 0.007 | 3.45E-01 | 18672 | -3.887 | -5.837 | -1.938 | 0.995 | -3.909 | -0.029 | 9.31E-05 | 18637 |
| 99 | Cerebellum_3_L | -0.124 | -0.230 | -0.017 | 0.054 | -2.271 | -0.017 | 2.32E-02 | 18672 | 0.023 | -0.119 | 0.164 | 0.072 | 0.312 | 0.002 | 7.55E-01 | 18637 |
| 100 | Cerebellum_3_R | -0.067 | -0.211 | 0.077 | 0.073 | -0.915 | -0.007 | 3.60E-01 | 18672 | -0.019 | -0.210 | 0.172 | 0.098 | -0.195 | -0.001 | 8.45E-01 | 18637 |
| 101 | Cerebellum_4_5_L | 0.532 | -0.332 | 1.395 | 0.440 | 1.207 | 0.009 | 2.27E-01 | 18672 | -0.477 | -1.623 | 0.668 | 0.584 | -0.817 | -0.006 | 4.14E-01 | 18637 |
| 102 | Cerebellum_4_5_R | 0.745 | 0.041 | 1.448 | 0.359 | 2.076 | 0.015 | 3.80E-02 | 18672 | -0.434 | -1.367 | 0.500 | 0.476 | -0.911 | -0.007 | 3.62E-01 | 18637 |
| 103 | Cerebellum_6_L | 1.050 | -0.441 | 2.542 | 0.761 | 1.380 | 0.010 | 1.68E-01 | 18672 | -1.685 | -3.664 | 0.293 | 1.009 | -1.670 | -0.012 | 9.49E-02 | 18637 |
| 104 | Cerebellum_6_R | 0.988 | -0.583 | 2.560 | 0.802 | 1.233 | 0.009 | 2.18E-01 | 18672 | -1.695 | -3.781 | 0.390 | 1.064 | -1.593 | -0.012 | 1.11E-01 | 18637 |
| 105 | Cerebellum_7b_L | 0.238 | -0.248 | 0.725 | 0.248 | 0.960 | 0.007 | 3.37E-01 | 18672 | -1.300 | -1.944 | -0.656 | 0.328 | -3.959 | -0.029 | 7.57E-05 | 18637 |
| 106 | Cerebellum_7b_R | 0.379 | -0.103 | 0.861 | 0.246 | 1.542 | 0.011 | 1.23E-01 | 18672 | -1.213 | -1.851 | -0.574 | 0.326 | -3.723 | -0.027 | 1.97E-04 | 18637 |
| 107 | Cerebellum_8_L | 0.953 | -0.591 | 2.497 | 0.788 | 1.210 | 0.009 | 2.26E-01 | 18672 | -3.833 | -5.878 | -1.788 | 1.043 | -3.674 | -0.027 | 2.39E-04 | 18637 |
| 108 | Cerebellum_8_R | 0.657 | -1.353 | 2.666 | 1.025 | 0.641 | 0.005 | 5.22E-01 | 18672 | -5.704 | -8.366 | -3.041 | 1.359 | -4.198 | -0.031 | 2.70E-05 | 18637 |
| 109 | Cerebellum_9_L | -0.462 | -1.335 | 0.411 | 0.445 | -1.038 | -0.008 | 2.99E-01 | 18672 | -1.376 | -2.532 | -0.219 | 0.590 | -2.332 | -0.017 | 1.97E-02 | 18637 |
| 110 | Cerebellum_9_R | -0.676 | -1.534 | 0.182 | 0.438 | -1.545 | -0.011 | 1.22E-01 | 18672 | -1.262 | -2.399 | -0.124 | 0.580 | -2.174 | -0.016 | 2.97E-02 | 18637 |
| 111 | Cerebellum_10_L | -0.029 | -0.091 | 0.033 | 0.032 | -0.917 | -0.007 | 3.59E-01 | 18672 | -0.058 | -0.141 | 0.024 | 0.042 | -1.393 | -0.010 | 1.64E-01 | 18637 |
| 112 | Cerebellum_10_R | -0.050 | -0.115 | 0.014 | 0.033 | -1.527 | -0.011 | 1.27E-01 | 18672 | 0.025 | -0.060 | 0.111 | 0.044 | 0.576 | 0.004 | 5.65E-01 | 18637 |
| 113 | Vermis_1_2 | -0.068 | -0.108 | -0.028 | 0.021 | -3.319 | -0.024 | 9.07E-04 | 18672 | 0.010 | -0.044 | 0.063 | 0.027 | 0.361 | 0.003 | 7.18E-01 | 18637 |
| 114 | Vermis_3 | -0.162 | -0.322 | -0.002 | 0.082 | -1.989 | -0.015 | 4.68E-02 | 18672 | -0.071 | -0.283 | 0.142 | 0.108 | -0.653 | -0.005 | 5.14E-01 | 18637 |
| 115 | Vermis_4_5 | 0.015 | -0.455 | 0.485 | 0.240 | 0.061 | 0.000 | 9.51E-01 | 18672 | -0.023 | -0.647 | 0.602 | 0.319 | -0.072 | -0.001 | 9.43E-01 | 18637 |
| 116 | Vermis_6 | -0.117 | -0.379 | 0.145 | 0.134 | -0.875 | -0.006 | 3.82E-01 | 18672 | 0.058 | -0.290 | 0.405 | 0.177 | 0.325 | 0.002 | 7.45E-01 | 18637 |
| 117 | Vermis_7 | -0.151 | -0.299 | -0.003 | 0.076 | -1.998 | -0.015 | 4.57E-02 | 18672 | -0.018 | -0.214 | 0.179 | 0.100 | -0.178 | -0.001 | 8.59E-01 | 18637 |
| 118 | Vermis_8 | -0.239 | -0.482 | 0.004 | 0.124 | -1.926 | -0.014 | 5.42E-02 | 18672 | -0.282 | -0.605 | 0.041 | 0.165 | -1.714 | -0.013 | 8.66E-02 | 18637 |

| | | | | | | | | | | | | | | | | | |
|---|---|---|---|---|---|---|---|---|---|---|---|---|---|---|---|---|---|
| 119 | Vermis_9 | -0.178 | -0.375 | 0.019 | 0.101 | -1.766 | -0.013 | 7.73E-02 | 18672 | -0.207 | -0.468 | 0.055 | 0.133 | -1.551 | -0.011 | 1.21E-01 | 18637 |
| 120 | Vermis_10 | -0.036 | -0.091 | 0.019 | 0.028 | -1.292 | -0.009 | 1.96E-01 | 18672 | 0.020 | -0.053 | 0.093 | 0.037 | 0.534 | 0.004 | 5.93E-01 | 18637 |
| 121 | Thal_AV_L | 0.015 | -0.007 | 0.037 | 0.011 | 1.342 | 0.010 | 1.80E-01 | 18672 | -0.039 | -0.068 | -0.010 | 0.015 | -2.600 | -0.019 | 9.33E-03 | 18637 |
| 122 | Thal_AV_R | 0.025 | -0.006 | 0.057 | 0.016 | 1.561 | 0.011 | 1.19E-01 | 18672 | -0.058 | -0.100 | -0.017 | 0.021 | -2.736 | -0.020 | 6.23E-03 | 18637 |
| 123 | Thal_LP_L | 0.035 | 0.011 | 0.060 | 0.012 | 2.824 | 0.021 | 4.74E-03 | 18672 | -0.078 | -0.110 | -0.046 | 0.017 | -4.721 | -0.035 | 2.37E-06 | 18637 |
| 124 | Thal_LP_R | 0.055 | 0.023 | 0.087 | 0.016 | 3.406 | 0.025 | 6.62E-04 | 18672 | -0.130 | -0.172 | -0.088 | 0.022 | -6.059 | -0.044 | 1.39E-09 | 18637 |
| 125 | Thal_VA_L | -0.021 | -0.047 | 0.005 | 0.013 | -1.583 | -0.012 | 1.14E-01 | 18672 | 0.017 | -0.018 | 0.051 | 0.018 | 0.952 | 0.007 | 3.41E-01 | 18637 |
| 126 | Thal_VA_R | -0.022 | -0.054 | 0.010 | 0.016 | -1.349 | -0.010 | 1.77E-01 | 18672 | -0.008 | -0.050 | 0.035 | 0.022 | -0.348 | -0.003 | 7.28E-01 | 18637 |
| 127 | Thal_VL_L | 0.088 | 0.014 | 0.162 | 0.038 | 2.322 | 0.017 | 2.03E-02 | 18672 | -0.233 | -0.331 | -0.135 | 0.050 | -4.644 | -0.034 | 3.45E-06 | 18637 |
| 128 | Thal_VL_R | 0.156 | 0.058 | 0.255 | 0.050 | 3.114 | 0.023 | 1.85E-03 | 18672 | -0.405 | -0.536 | -0.275 | 0.067 | -6.091 | -0.045 | 1.14E-09 | 18637 |
| 129 | Thal_VPL_L | 0.198 | 0.110 | 0.286 | 0.045 | 4.406 | 0.032 | 1.06E-05 | 18672 | -0.372 | -0.489 | -0.256 | 0.060 | -6.245 | -0.046 | 4.33E-10 | 18637 |
| 130 | Thal_VPL_R | 0.227 | 0.134 | 0.319 | 0.047 | 4.790 | 0.035 | 1.68E-06 | 18672 | -0.452 | -0.575 | -0.329 | 0.063 | -7.204 | -0.053 | 6.06E-13 | 18637 |
| 131 | Thal_IL_L | 0.079 | 0.041 | 0.117 | 0.019 | 4.069 | 0.030 | 4.75E-05 | 18672 | -0.165 | -0.215 | -0.114 | 0.026 | -6.380 | -0.047 | 1.81E-10 | 18637 |
| 132 | Thal_IL_R | 0.104 | 0.061 | 0.147 | 0.022 | 4.716 | 0.034 | 2.43E-06 | 18672 | -0.214 | -0.272 | -0.157 | 0.029 | -7.321 | -0.054 | 2.56E-13 | 18637 |
| 133 | Thal_Re_L | 0.000 | 0.000 | 0.000 | 0.000 | -2.689 | -0.020 | 7.17E-03 | 18672 | 0.000 | 0.000 | 0.000 | 0.000 | -1.909 | -0.014 | 5.63E-02 | 18637 |
| 134 | Thal_Re_R | 0.000 | -0.001 | 0.001 | 0.001 | -0.212 | -0.002 | 8.32E-01 | 18672 | -0.004 | -0.005 | -0.002 | 0.001 | -4.313 | -0.032 | 1.62E-05 | 18637 |
| 135 | Thal_MDm_L | 0.643 | 0.360 | 0.925 | 0.144 | 4.457 | 0.033 | 8.37E-06 | 18672 | -1.211 | -1.585 | -0.836 | 0.191 | -6.332 | -0.046 | 2.47E-10 | 18637 |
| 136 | Thal_MDm_R | 0.796 | 0.467 | 1.124 | 0.168 | 4.747 | 0.035 | 2.08E-06 | 18672 | -1.390 | -1.825 | -0.955 | 0.222 | -6.258 | -0.046 | 3.99E-10 | 18637 |
| 137 | Thal_MDl_L | 0.119 | 0.070 | 0.168 | 0.025 | 4.768 | 0.035 | 1.88E-06 | 18672 | -0.241 | -0.306 | -0.177 | 0.033 | -7.292 | -0.053 | 3.18E-13 | 18637 |
| 138 | Thal_MDl_R | 0.149 | 0.093 | 0.205 | 0.028 | 5.255 | 0.038 | 1.49E-07 | 18672 | -0.269 | -0.342 | -0.195 | 0.038 | -7.144 | -0.052 | 9.38E-13 | 18637 |
| 139 | Thal_LGN_L | 0.005 | -0.003 | 0.012 | 0.004 | 1.225 | 0.009 | 2.21E-01 | 18672 | -0.013 | -0.023 | -0.003 | 0.005 | -2.656 | -0.019 | 7.92E-03 | 18637 |
| 140 | Thal_LGN_R | 0.002 | -0.013 | 0.018 | 0.008 | 0.306 | 0.002 | 7.60E-01 | 18672 | -0.061 | -0.082 | -0.040 | 0.011 | -5.699 | -0.042 | 1.23E-08 | 18637 |
| 141 | Thal_MGN_L | 0.017 | 0.003 | 0.030 | 0.007 | 2.449 | 0.018 | 1.43E-02 | 18672 | -0.050 | -0.067 | -0.032 | 0.009 | -5.537 | -0.041 | 3.11E-08 | 18637 |
| 142 | Thal_MGN_R | 0.017 | 0.003 | 0.032 | 0.007 | 2.328 | 0.017 | 1.99E-02 | 18672 | -0.061 | -0.080 | -0.042 | 0.010 | -6.250 | -0.046 | 4.18E-10 | 18637 |
| 143 | Thal_PuA_L | 0.024 | 0.008 | 0.039 | 0.008 | 2.982 | 0.022 | 2.87E-03 | 18672 | -0.059 | -0.080 | -0.038 | 0.011 | -5.582 | -0.041 | 2.42E-08 | 18637 |
| 144 | Thal_PuA_R | 0.053 | 0.020 | 0.085 | 0.017 | 3.171 | 0.023 | 1.52E-03 | 18672 | -0.143 | -0.186 | -0.100 | 0.022 | -6.497 | -0.048 | 8.39E-11 | 18637 |
| 145 | Thal_PuM_L | 0.627 | 0.316 | 0.938 | 0.159 | 3.951 | 0.029 | 7.82E-05 | 18672 | -1.294 | -1.706 | -0.881 | 0.210 | -6.151 | -0.045 | 7.85E-10 | 18637 |
| 146 | Thal_PuM_R | 0.863 | 0.475 | 1.250 | 0.198 | 4.360 | 0.032 | 1.31E-05 | 18672 | -1.687 | -2.201 | -1.174 | 0.262 | -6.438 | -0.047 | 1.24E-10 | 18637 |
| 147 | Thal_PuL_L | 0.136 | 0.080 | 0.193 | 0.029 | 4.714 | 0.034 | 2.45E-06 | 18672 | -0.256 | -0.331 | -0.180 | 0.038 | -6.660 | -0.049 | 2.81E-11 | 18637 |
| 148 | Thal_PuL_R | 0.225 | 0.135 | 0.315 | 0.046 | 4.888 | 0.036 | 1.03E-06 | 18672 | -0.416 | -0.536 | -0.296 | 0.061 | -6.814 | -0.050 | 9.78E-12 | 18637 |
| 149 | Thal_PuI_L | 0.019 | 0.004 | 0.035 | 0.008 | 2.426 | 0.018 | 1.53E-02 | 18672 | -0.032 | -0.053 | -0.012 | 0.010 | -3.108 | -0.023 | 1.89E-03 | 18637 |
| 150 | Thal_PuI_R | 0.025 | 0.005 | 0.046 | 0.010 | 2.443 | 0.018 | 1.46E-02 | 18672 | -0.063 | -0.090 | -0.036 | 0.014 | -4.571 | -0.033 | 4.89E-06 | 18637 |
| 151 | ACC_sub_L | 0.074 | -0.104 | 0.253 | 0.091 | 0.820 | 0.006 | 4.13E-01 | 18672 | -0.129 | -0.365 | 0.107 | 0.120 | -1.071 | -0.008 | 2.84E-01 | 18637 |
| 152 | ACC_sub_R | 0.101 | -0.065 | 0.268 | 0.085 | 1.194 | 0.009 | 2.33E-01 | 18672 | -0.092 | -0.312 | 0.129 | 0.113 | -0.813 | -0.006 | 4.16E-01 | 18637 |
| 153 | ACC_pre_L | 0.103 | -0.354 | 0.559 | 0.233 | 0.441 | 0.003 | 6.59E-01 | 18672 | -0.295 | -0.900 | 0.310 | 0.309 | -0.956 | -0.007 | 3.39E-01 | 18637 |
| 154 | ACC_pre_R | -0.092 | -0.531 | 0.348 | 0.224 | -0.410 | -0.003 | 6.82E-01 | 18672 | -0.662 | -1.244 | -0.079 | 0.297 | -2.225 | -0.016 | 2.61E-02 | 18637 |
| 155 | ACC_sup_L | 0.217 | -0.279 | 0.714 | 0.253 | 0.859 | 0.006 | 3.90E-01 | 18672 | -1.160 | -1.818 | -0.502 | 0.336 | -3.457 | -0.025 | 5.48E-04 | 18637 |
| 156 | ACC_sup_R | -0.153 | -0.666 | 0.361 | 0.262 | -0.582 | -0.004 | 5.60E-01 | 18672 | -0.654 | -1.336 | 0.028 | 0.348 | -1.880 | -0.014 | 6.01E-02 | 18637 |
| 157 | N_Acc_L | -0.188 | -0.338 | -0.039 | 0.076 | -2.469 | -0.018 | 1.35E-02 | 18672 | -0.313 | -0.511 | -0.115 | 0.101 | -3.099 | -0.023 | 1.95E-03 | 18637 |
| 158 | N_Acc_R | -0.184 | -0.313 | -0.054 | 0.066 | -2.785 | -0.020 | 5.36E-03 | 18672 | -0.248 | -0.420 | -0.077 | 0.087 | -2.840 | -0.021 | 4.51E-03 | 18637 |
| 159 | VTA_L | 0.007 | 0.002 | 0.012 | 0.003 | 2.686 | 0.020 | 7.23E-03 | 18672 | -0.011 | -0.018 | -0.004 | 0.004 | -3.082 | -0.023 | 2.06E-03 | 18637 |
| 160 | VTA_R | 0.008 | 0.003 | 0.012 | 0.002 | 3.202 | 0.023 | 1.37E-03 | 18672 | -0.010 | -0.016 | -0.004 | 0.003 | -3.104 | -0.023 | 1.91E-03 | 18637 |
| 161 | SN_pc_L | 0.014 | 0.005 | 0.022 | 0.004 | 3.150 | 0.023 | 1.64E-03 | 18672 | -0.014 | -0.026 | -0.003 | 0.006 | -2.521 | -0.018 | 1.17E-02 | 18637 |
| 162 | SN_pc_R | 0.021 | 0.009 | 0.032 | 0.006 | 3.641 | 0.027 | 2.72E-04 | 18672 | -0.021 | -0.036 | -0.007 | 0.007 | -2.844 | -0.021 | 4.46E-03 | 18637 |
| 163 | SN_pr_L | 0.010 | -0.002 | 0.021 | 0.006 | 1.658 | 0.012 | 9.73E-02 | 18672 | -0.018 | -0.033 | -0.003 | 0.008 | -2.346 | -0.017 | 1.90E-02 | 18637 |
| 164 | SN_pr_R | 0.021 | 0.008 | 0.034 | 0.007 | 3.169 | 0.023 | 1.53E-03 | 18672 | -0.016 | -0.033 | 0.001 | 0.009 | -1.836 | -0.013 | 6.64E-02 | 18637 |
| 165 | Red_N_L | 0.002 | -0.012 | 0.016 | 0.007 | 0.248 | 0.002 | 8.04E-01 | 18672 | -0.029 | -0.048 | -0.010 | 0.010 | -3.011 | -0.022 | 2.61E-03 | 18637 |
| 166 | Red_N_R | 0.006 | -0.010 | 0.021 | 0.008 | 0.748 | 0.005 | 4.54E-01 | 18672 | -0.028 | -0.048 | -0.007 | 0.010 | -2.653 | -0.019 | 7.99E-03 | 18637 |

| | | | | | | | | | | | | | | | | | |
|---|---|---|---|---|---|---|---|---|---|---|---|---|---|---|---|---|---|
| 167 | LC_L | -0.008 | -0.014 | -0.003 | 0.003 | -3.079 | -0.023 | 2.08E-03 | 18672 | 0.000 | -0.007 | 0.007 | 0.004 | -0.069 | -0.001 | 9.45E-01 | 18637 |
| 168 | LC_R | -0.009 | -0.015 | -0.003 | 0.003 | -3.009 | -0.022 | 2.63E-03 | 18672 | -0.001 | -0.009 | 0.007 | 0.004 | -0.281 | -0.002 | 7.79E-01 | 18637 |
| 169 | Raphe_D | -0.004 | -0.025 | 0.016 | 0.010 | -0.385 | -0.003 | 7.00E-01 | 18672 | -0.064 | -0.091 | -0.036 | 0.014 | -4.585 | -0.034 | 4.58E-06 | 18637 |
| 170 | Raphe_M | -0.002 | -0.009 | 0.004 | 0.003 | -0.720 | -0.005 | 4.71E-01 | 18672 | -0.006 | -0.014 | 0.002 | 0.004 | -1.371 | -0.010 | 1.70E-01 | 18637 |

| | | Cooked vegetable intake | | | | | | | | Processed meat intake | | | | | | | |
|---|---|---|---|---|---|---|---|---|---|---|---|---|---|---|---|---|---|
| Number | AAL 3 region | beta | 95%CI_l | 95%CI_h | se | t | r | p | df.e | beta | 95%CI_l | 95%CI_h | se | t | r | p | df.e |
| 0 | Total brain | -89.216 | -178.191 | -0.241 | 45.393 | -1.965 | -0.014 | 4.94E-02 | 18581 | -514.783 | -645.621 | -383.945 | 66.751 | -7.712 | -0.056 | 1.30E-14 | 18733 |
| 1 | Precentral_L | -1.155 | -4.285 | 1.976 | 1.597 | -0.723 | -0.005 | 4.70E-01 | 18581 | -10.080 | -14.690 | -5.469 | 2.352 | -4.285 | -0.031 | 1.83E-05 | 18733 |
| 2 | Precentral_R | -0.787 | -3.654 | 2.080 | 1.463 | -0.538 | -0.004 | 5.90E-01 | 18581 | -7.535 | -11.761 | -3.309 | 2.156 | -3.495 | -0.026 | 4.75E-04 | 18733 |
| 3 | Frontal_Sup_2_L | -0.811 | -4.116 | 2.494 | 1.686 | -0.481 | -0.004 | 6.30E-01 | 18581 | -10.827 | -15.693 | -5.962 | 2.482 | -4.362 | -0.032 | 1.30E-05 | 18733 |
| 4 | Frontal_Sup_2_R | -1.322 | -4.966 | 2.322 | 1.859 | -0.711 | -0.005 | 4.77E-01 | 18581 | -14.394 | -19.755 | -9.033 | 2.735 | -5.263 | -0.038 | 1.43E-07 | 18733 |
| 5 | Frontal_Mid_2_L | -1.964 | -5.435 | 1.508 | 1.771 | -1.109 | -0.008 | 2.68E-01 | 18581 | -9.067 | -14.179 | -3.956 | 2.608 | -3.477 | -0.025 | 5.08E-04 | 18733 |
| 6 | Frontal_Mid_2_R | -3.910 | -7.836 | 0.017 | 2.003 | -1.951 | -0.014 | 5.10E-02 | 18581 | -15.032 | -20.814 | -9.251 | 2.950 | -5.096 | -0.037 | 3.50E-07 | 18733 |
| 7 | Frontal_Inf_Oper_L | -0.382 | -1.407 | 0.644 | 0.523 | -0.730 | -0.005 | 4.65E-01 | 18581 | -4.191 | -5.701 | -2.681 | 0.770 | -5.440 | -0.040 | 5.39E-08 | 18733 |
| 8 | Frontal_Inf_Oper_R | -0.804 | -2.091 | 0.482 | 0.656 | -1.225 | -0.009 | 2.20E-01 | 18581 | -5.521 | -7.414 | -3.628 | 0.966 | -5.717 | -0.042 | 1.10E-08 | 18733 |
| 9 | Frontal_Inf_Tri_L | -1.278 | -3.456 | 0.901 | 1.111 | -1.150 | -0.008 | 2.50E-01 | 18581 | -9.302 | -12.508 | -6.095 | 1.636 | -5.686 | -0.042 | 1.32E-08 | 18733 |
| 10 | Frontal_Inf_Tri_R | 0.458 | -1.253 | 2.170 | 0.873 | 0.525 | 0.004 | 6.00E-01 | 18581 | -7.121 | -9.639 | -4.604 | 1.284 | -5.544 | -0.040 | 2.99E-08 | 18733 |
| 11 | Frontal_Inf_Orb_2_L | -0.318 | -0.970 | 0.333 | 0.332 | -0.957 | -0.007 | 3.39E-01 | 18581 | -2.428 | -3.387 | -1.469 | 0.489 | -4.964 | -0.036 | 6.97E-07 | 18733 |
| 12 | Frontal_Inf_Orb_2_R | 0.057 | -0.624 | 0.738 | 0.348 | 0.163 | 0.001 | 8.70E-01 | 18581 | -2.339 | -3.342 | -1.337 | 0.512 | -4.573 | -0.033 | 4.85E-06 | 18733 |
| 13 | Rolandic_Oper_L | -1.183 | -2.168 | -0.198 | 0.503 | -2.354 | -0.017 | 1.86E-02 | 18581 | -3.394 | -4.844 | -1.944 | 0.740 | -4.588 | -0.034 | 4.50E-06 | 18733 |
| 14 | Rolandic_Oper_R | -0.285 | -1.695 | 1.125 | 0.719 | -0.396 | -0.003 | 6.92E-01 | 18581 | -5.692 | -7.767 | -3.617 | 1.059 | -5.377 | -0.039 | 7.65E-08 | 18733 |
| 15 | Supp_Motor_Area_L | -0.498 | -2.838 | 1.841 | 1.194 | -0.417 | -0.003 | 6.76E-01 | 18581 | -7.131 | -10.579 | -3.683 | 1.759 | -4.054 | -0.030 | 5.06E-05 | 18733 |
| 16 | Supp_Motor_Area_R | 0.281 | -2.306 | 2.868 | 1.320 | 0.213 | 0.002 | 8.31E-01 | 18581 | -6.983 | -10.793 | -3.173 | 1.944 | -3.593 | -0.026 | 3.28E-04 | 18733 |
| 17 | Olfactory_L | -0.335 | -0.683 | 0.013 | 0.178 | -1.886 | -0.014 | 5.93E-02 | 18581 | -0.886 | -1.398 | -0.374 | 0.261 | -3.392 | -0.025 | 6.94E-04 | 18733 |
| 18 | Olfactory_R | -0.348 | -0.697 | 0.001 | 0.178 | -1.952 | -0.014 | 5.10E-02 | 18581 | -0.837 | -1.351 | -0.324 | 0.262 | -3.194 | -0.023 | 1.40E-03 | 18733 |
| 19 | Frontal_Sup_Medial_L | 0.131 | -2.063 | 2.325 | 1.119 | 0.117 | 0.001 | 9.07E-01 | 18581 | -10.324 | -13.552 | -7.096 | 1.647 | -6.269 | -0.046 | 3.71E-10 | 18733 |
| 20 | Frontal_Sup_Medial_R | -1.187 | -2.785 | 0.410 | 0.815 | -1.457 | -0.011 | 1.45E-01 | 18581 | -7.511 | -9.861 | -5.161 | 1.199 | -6.265 | -0.046 | 3.80E-10 | 18733 |
| 21 | Frontal_Med_Orb_L | -1.058 | -1.878 | -0.238 | 0.418 | -2.529 | -0.019 | 1.14E-02 | 18581 | -3.034 | -4.240 | -1.827 | 0.616 | -4.928 | -0.036 | 8.37E-07 | 18733 |
| 22 | Frontal_Med_Orb_R | -1.276 | -2.283 | -0.268 | 0.514 | -2.481 | -0.018 | 1.31E-02 | 18581 | -4.247 | -5.730 | -2.764 | 0.757 | -5.614 | -0.041 | 2.01E-08 | 18733 |
| 23 | Rectus_L | -1.068 | -1.917 | -0.219 | 0.433 | -2.466 | -0.018 | 1.37E-02 | 18581 | -3.028 | -4.276 | -1.779 | 0.637 | -4.754 | -0.035 | 2.01E-06 | 18733 |
| 24 | Rectus_R | -1.017 | -1.801 | -0.232 | 0.400 | -2.539 | -0.019 | 1.11E-02 | 18581 | -2.550 | -3.705 | -1.394 | 0.589 | -4.326 | -0.032 | 1.53E-05 | 18733 |
| 25 | OFCmed_L | -0.271 | -0.764 | 0.221 | 0.251 | -1.081 | -0.008 | 2.80E-01 | 18581 | -1.810 | -2.533 | -1.086 | 0.369 | -4.903 | -0.036 | 9.50E-07 | 18733 |
| 26 | OFCmed_R | -0.459 | -1.033 | 0.114 | 0.292 | -1.571 | -0.012 | 1.16E-01 | 18581 | -2.092 | -2.936 | -1.248 | 0.430 | -4.861 | -0.035 | 1.18E-06 | 18733 |
| 27 | OFCant_L | -0.439 | -1.008 | 0.130 | 0.290 | -1.513 | -0.011 | 1.30E-01 | 18581 | -1.548 | -2.385 | -0.710 | 0.427 | -3.622 | -0.026 | 2.93E-04 | 18733 |
| 28 | OFCant_R | -0.603 | -1.355 | 0.148 | 0.383 | -1.573 | -0.012 | 1.16E-01 | 18581 | -2.526 | -3.632 | -1.420 | 0.564 | -4.477 | -0.033 | 7.63E-06 | 18733 |
| 29 | OFCpost_L | -0.415 | -1.127 | 0.298 | 0.363 | -1.140 | -0.008 | 2.54E-01 | 18581 | -1.693 | -2.742 | -0.644 | 0.535 | -3.164 | -0.023 | 1.56E-03 | 18733 |
| 30 | OFCpost_R | -0.422 | -1.080 | 0.237 | 0.336 | -1.255 | -0.009 | 2.09E-01 | 18581 | -2.153 | -3.122 | -1.184 | 0.494 | -4.354 | -0.032 | 1.34E-05 | 18733 |
| 31 | OFClat_L | -0.018 | -0.258 | 0.223 | 0.123 | -0.144 | -0.001 | 8.86E-01 | 18581 | -0.834 | -1.187 | -0.480 | 0.180 | -4.620 | -0.034 | 3.86E-06 | 18733 |
| 32 | OFClat_R | -0.021 | -0.224 | 0.183 | 0.104 | -0.198 | -0.001 | 8.43E-01 | 18581 | -0.720 | -1.019 | -0.422 | 0.152 | -4.726 | -0.035 | 2.31E-06 | 18733 |
| 33 | Insula_L | -2.050 | -4.253 | 0.153 | 1.124 | -1.824 | -0.013 | 6.82E-02 | 18581 | -7.278 | -10.519 | -4.036 | 1.654 | -4.401 | -0.032 | 1.09E-05 | 18733 |
| 34 | Insula_R | -2.094 | -4.239 | 0.052 | 1.095 | -1.912 | -0.014 | 5.58E-02 | 18581 | -7.487 | -10.645 | -4.329 | 1.611 | -4.647 | -0.034 | 3.40E-06 | 18733 |
| 37 | Cingulate_Mid_L | -1.646 | -3.508 | 0.215 | 0.950 | -1.733 | -0.013 | 8.31E-02 | 18581 | -10.941 | -13.678 | -8.203 | 1.397 | -7.834 | -0.057 | 4.99E-15 | 18733 |
| 38 | Cingulate_Mid_R | -1.174 | -3.159 | 0.811 | 1.013 | -1.159 | -0.009 | 2.46E-01 | 18581 | -12.263 | -15.182 | -9.345 | 1.489 | -8.236 | -0.060 | 1.90E-16 | 18733 |
| 39 | Cingulate_Post_L | -0.493 | -1.062 | 0.076 | 0.290 | -1.697 | -0.012 | 8.96E-02 | 18581 | -1.175 | -2.013 | -0.338 | 0.427 | -2.750 | -0.020 | 5.96E-03 | 18733 |
| 40 | Cingulate_Post_R | -0.275 | -0.591 | 0.041 | 0.161 | -1.709 | -0.013 | 8.76E-02 | 18581 | -0.547 | -1.012 | -0.082 | 0.237 | -2.307 | -0.017 | 2.11E-02 | 18733 |
| 41 | Hippocampus_L | 0.335 | -0.585 | 1.254 | 0.469 | 0.713 | 0.005 | 4.76E-01 | 18581 | -2.125 | -3.479 | -0.770 | 0.691 | -3.074 | -0.022 | 2.12E-03 | 18733 |
| 42 | Hippocampus_R | 0.268 | -0.686 | 1.222 | 0.487 | 0.551 | 0.004 | 5.82E-01 | 18581 | -2.581 | -3.986 | -1.176 | 0.717 | -3.602 | -0.026 | 3.17E-04 | 18733 |
| 43 | ParaHippocampal_L | 0.730 | -0.087 | 1.547 | 0.417 | 1.752 | 0.013 | 7.98E-02 | 18581 | -2.381 | -3.585 | -1.178 | 0.614 | -3.878 | -0.028 | 1.06E-04 | 18733 |

| | | | | | | | | | | | | | | | | | |
|---|---|---|---|---|---|---|---|---|---|---|---|---|---|---|---|---|---|
| 44 | ParaHippocampal_R | 0.379 | -0.652 | 1.410 | 0.526 | 0.720 | 0.005 | 4.71E-01 | 18581 | -2.852 | -4.371 | -1.333 | 0.775 | -3.681 | -0.027 | 2.33E-04 | 18733 |
| 45 | Amygdala_L | -0.069 | -0.386 | 0.249 | 0.162 | -0.424 | -0.003 | 6.71E-01 | 18581 | -0.429 | -0.897 | 0.038 | 0.239 | -1.799 | -0.013 | 7.20E-02 | 18733 |
| 46 | Amygdala_R | -0.192 | -0.517 | 0.132 | 0.166 | -1.161 | -0.009 | 2.46E-01 | 18581 | -0.791 | -1.270 | -0.313 | 0.244 | -3.241 | -0.024 | 1.19E-03 | 18733 |
| 47 | Calcarine_L | -1.598 | -4.629 | 1.433 | 1.546 | -1.034 | -0.008 | 3.01E-01 | 18581 | -3.918 | -8.382 | 0.546 | 2.277 | -1.721 | -0.013 | 8.54E-02 | 18733 |
| 48 | Calcarine_R | -2.449 | -4.983 | 0.086 | 1.293 | -1.894 | -0.014 | 5.83E-02 | 18581 | -3.227 | -6.957 | 0.503 | 1.903 | -1.696 | -0.012 | 8.99E-02 | 18733 |
| 49 | Cuneus_L | -0.806 | -2.706 | 1.095 | 0.970 | -0.831 | -0.006 | 4.06E-01 | 18581 | -3.259 | -6.059 | -0.459 | 1.428 | -2.282 | -0.017 | 2.25E-02 | 18733 |
| 50 | Cuneus_R | -1.373 | -2.828 | 0.081 | 0.742 | -1.850 | -0.014 | 6.43E-02 | 18581 | -3.970 | -6.113 | -1.828 | 1.093 | -3.633 | -0.027 | 2.81E-04 | 18733 |
| 51 | Lingual_L | -0.898 | -2.721 | 0.926 | 0.930 | -0.965 | -0.007 | 3.35E-01 | 18581 | -6.014 | -8.697 | -3.331 | 1.369 | -4.393 | -0.032 | 1.12E-05 | 18733 |
| 52 | Lingual_R | -1.830 | -3.743 | 0.082 | 0.976 | -1.876 | -0.014 | 6.07E-02 | 18581 | -4.820 | -7.633 | -2.008 | 1.435 | -3.359 | -0.025 | 7.83E-04 | 18733 |
| 53 | Occipital_Sup_L | -0.537 | -1.791 | 0.716 | 0.640 | -0.840 | -0.006 | 4.01E-01 | 18581 | -4.084 | -5.928 | -2.240 | 0.941 | -4.341 | -0.032 | 1.43E-05 | 18733 |
| 54 | Occipital_Sup_R | -0.288 | -1.648 | 1.071 | 0.694 | -0.415 | -0.003 | 6.78E-01 | 18581 | -3.127 | -5.127 | -1.127 | 1.020 | -3.065 | -0.022 | 2.18E-03 | 18733 |
| 55 | Occipital_Mid_L | -2.908 | -5.846 | 0.030 | 1.499 | -1.940 | -0.014 | 5.24E-02 | 18581 | -7.213 | -11.538 | -2.888 | 2.207 | -3.269 | -0.024 | 1.08E-03 | 18733 |
| 56 | Occipital_Mid_R | -0.673 | -2.717 | 1.371 | 1.043 | -0.645 | -0.005 | 5.19E-01 | 18581 | -1.296 | -4.306 | 1.714 | 1.536 | -0.844 | -0.006 | 3.99E-01 | 18733 |
| 57 | Occipital_Inf_L | -0.814 | -1.789 | 0.161 | 0.498 | -1.636 | -0.012 | 1.02E-01 | 18581 | -0.716 | -2.153 | 0.720 | 0.733 | -0.978 | -0.007 | 3.28E-01 | 18733 |
| 58 | Occipital_Inf_R | -0.639 | -1.766 | 0.487 | 0.575 | -1.112 | -0.008 | 2.66E-01 | 18581 | 1.403 | -0.256 | 3.062 | 0.846 | 1.658 | 0.012 | 9.73E-02 | 18733 |
| 59 | Fusiform_L | 0.644 | -1.490 | 2.778 | 1.089 | 0.591 | 0.004 | 5.54E-01 | 18581 | -4.402 | -7.545 | -1.260 | 1.603 | -2.746 | -0.020 | 6.04E-03 | 18733 |
| 60 | Fusiform_R | 0.278 | -2.068 | 2.623 | 1.197 | 0.232 | 0.002 | 8.16E-01 | 18581 | -5.486 | -8.941 | -2.031 | 1.763 | -3.112 | -0.023 | 1.86E-03 | 18733 |
| 61 | Postcentral_L | 0.147 | -3.066 | 3.359 | 1.639 | 0.089 | 0.001 | 9.29E-01 | 18581 | -17.027 | -21.748 | -12.305 | 2.409 | -7.068 | -0.052 | 1.62E-12 | 18733 |
| 62 | Postcentral_R | -1.468 | -4.761 | 1.825 | 1.680 | -0.874 | -0.006 | 3.82E-01 | 18581 | -11.707 | -16.549 | -6.865 | 2.470 | -4.739 | -0.035 | 2.16E-06 | 18733 |
| 63 | Parietal_Sup_L | -1.213 | -3.063 | 0.638 | 0.944 | -1.285 | -0.009 | 1.99E-01 | 18581 | -4.934 | -7.656 | -2.211 | 1.389 | -3.552 | -0.026 | 3.83E-04 | 18733 |
| 64 | Parietal_Sup_R | -1.044 | -2.686 | 0.598 | 0.838 | -1.246 | -0.009 | 2.13E-01 | 18581 | -3.238 | -5.653 | -0.822 | 1.232 | -2.627 | -0.019 | 8.62E-03 | 18733 |
| 65 | Parietal_Inf_L | -1.714 | -4.136 | 0.709 | 1.236 | -1.386 | -0.010 | 1.66E-01 | 18581 | -12.303 | -15.865 | -8.741 | 1.817 | -6.769 | -0.049 | 1.33E-11 | 18733 |
| 66 | Parietal_Inf_R | -0.298 | -1.867 | 1.270 | 0.800 | -0.373 | -0.003 | 7.09E-01 | 18581 | -5.038 | -7.348 | -2.729 | 1.178 | -4.277 | -0.031 | 1.90E-05 | 18733 |
| 67 | SupraMarginal_L | -0.298 | -1.718 | 1.122 | 0.724 | -0.411 | -0.003 | 6.81E-01 | 18581 | -6.054 | -8.143 | -3.966 | 1.065 | -5.682 | -0.041 | 1.35E-08 | 18733 |
| 68 | SupraMarginal_R | -1.725 | -3.734 | 0.283 | 1.025 | -1.684 | -0.012 | 9.22E-02 | 18581 | -7.995 | -10.949 | -5.040 | 1.507 | -5.303 | -0.039 | 1.15E-07 | 18733 |
| 69 | Angular_L | -0.863 | -2.257 | 0.531 | 0.711 | -1.213 | -0.009 | 2.25E-01 | 18581 | -3.807 | -5.860 | -1.754 | 1.047 | -3.635 | -0.027 | 2.78E-04 | 18733 |
| 70 | Angular_R | -0.645 | -2.517 | 1.227 | 0.955 | -0.675 | -0.005 | 5.00E-01 | 18581 | -2.629 | -5.382 | 0.124 | 1.405 | -1.872 | -0.014 | 6.13E-02 | 18733 |
| 71 | Precuneus_L | -2.467 | -5.868 | 0.933 | 1.735 | -1.422 | -0.010 | 1.55E-01 | 18581 | -11.872 | -16.873 | -6.871 | 2.551 | -4.653 | -0.034 | 3.29E-06 | 18733 |
| 72 | Precuneus_R | -2.956 | -5.985 | 0.074 | 1.546 | -1.912 | -0.014 | 5.59E-02 | 18581 | -9.032 | -13.488 | -4.577 | 2.273 | -3.973 | -0.029 | 7.11E-05 | 18733 |
| 73 | Paracentral_Lobule_L | 0.439 | -1.117 | 1.995 | 0.794 | 0.553 | 0.004 | 5.80E-01 | 18581 | -4.030 | -6.321 | -1.740 | 1.169 | -3.449 | -0.025 | 5.65E-04 | 18733 |
| 74 | Paracentral_Lobule_R | 0.338 | -0.775 | 1.452 | 0.568 | 0.596 | 0.004 | 5.51E-01 | 18581 | -2.441 | -4.081 | -0.800 | 0.837 | -2.916 | -0.021 | 3.55E-03 | 18733 |
| 75 | Caudate_L | 0.297 | -0.946 | 1.541 | 0.634 | 0.469 | 0.003 | 6.39E-01 | 18581 | -3.160 | -4.990 | -1.331 | 0.934 | -3.385 | -0.025 | 7.13E-04 | 18733 |
| 76 | Caudate_R | -0.201 | -1.608 | 1.206 | 0.718 | -0.281 | -0.002 | 7.79E-01 | 18581 | -2.846 | -4.917 | -0.774 | 1.057 | -2.692 | -0.020 | 7.11E-03 | 18733 |
| 77 | Putamen_L | -0.931 | -2.817 | 0.956 | 0.963 | -0.967 | -0.007 | 3.34E-01 | 18581 | -4.377 | -7.156 | -1.599 | 1.418 | -3.088 | -0.023 | 2.02E-03 | 18733 |
| 78 | Putamen_R | -1.388 | -3.309 | 0.533 | 0.980 | -1.416 | -0.010 | 1.57E-01 | 18581 | -4.651 | -7.481 | -1.822 | 1.444 | -3.222 | -0.024 | 1.28E-03 | 18733 |
| 79 | Pallidum_L | 0.007 | -0.161 | 0.174 | 0.086 | 0.076 | 0.001 | 9.39E-01 | 18581 | -0.352 | -0.599 | -0.105 | 0.126 | -2.791 | -0.020 | 5.25E-03 | 18733 |
| 80 | Pallidum_R | 0.052 | -0.132 | 0.236 | 0.094 | 0.555 | 0.004 | 5.79E-01 | 18581 | -0.486 | -0.757 | -0.214 | 0.138 | -3.510 | -0.026 | 4.50E-04 | 18733 |
| 83 | Heschl_L | -0.343 | -0.683 | -0.002 | 0.174 | -1.973 | -0.014 | 4.85E-02 | 18581 | -1.263 | -1.763 | -0.762 | 0.255 | -4.943 | -0.036 | 7.75E-07 | 18733 |
| 84 | Heschl_R | -0.215 | -0.596 | 0.165 | 0.194 | -1.108 | -0.008 | 2.68E-01 | 18581 | -1.351 | -1.910 | -0.792 | 0.285 | -4.734 | -0.035 | 2.22E-06 | 18733 |
| 85 | Temporal_Sup_L | -2.907 | -5.365 | -0.450 | 1.254 | -2.319 | -0.017 | 2.04E-02 | 18581 | -9.823 | -13.437 | -6.208 | 1.844 | -5.327 | -0.039 | 1.01E-07 | 18733 |
| 86 | Temporal_Sup_R | -0.812 | -3.531 | 1.907 | 1.387 | -0.585 | -0.004 | 5.58E-01 | 18581 | -13.484 | -17.481 | -9.486 | 2.039 | -6.612 | -0.048 | 3.91E-11 | 18733 |
| 87 | Temporal_Pole_Sup_L | -0.977 | -2.141 | 0.186 | 0.594 | -1.646 | -0.012 | 9.98E-02 | 18581 | -1.726 | -3.440 | -0.013 | 0.874 | -1.975 | -0.014 | 4.83E-02 | 18733 |
| 88 | Temporal_Pole_Sup_R | -0.009 | -1.234 | 1.216 | 0.625 | -0.015 | 0.000 | 9.88E-01 | 18581 | -2.556 | -4.360 | -0.752 | 0.920 | -2.777 | -0.020 | 5.50E-03 | 18733 |
| 89 | Temporal_Mid_L | -2.296 | -6.391 | 1.799 | 2.089 | -1.099 | -0.008 | 2.72E-01 | 18581 | -15.292 | -21.318 | -9.266 | 3.074 | -4.974 | -0.036 | 6.62E-07 | 18733 |
| 90 | Temporal_Mid_R | -2.959 | -6.594 | 0.675 | 1.854 | -1.596 | -0.012 | 1.11E-01 | 18581 | -9.181 | -14.533 | -3.829 | 2.730 | -3.362 | -0.025 | 7.75E-04 | 18733 |
| 91 | Temporal_Pole_Mid_L | -0.024 | -1.002 | 0.954 | 0.499 | -0.049 | 0.000 | 9.61E-01 | 18581 | -0.530 | -1.971 | 0.910 | 0.735 | -0.721 | -0.005 | 4.71E-01 | 18733 |
| 92 | Temporal_Pole_Mid_R | -0.984 | -2.172 | 0.204 | 0.606 | -1.624 | -0.012 | 1.04E-01 | 18581 | -1.286 | -3.037 | 0.464 | 0.893 | -1.440 | -0.011 | 1.50E-01 | 18733 |
| 93 | Temporal_Inf_L | -1.387 | -4.203 | 1.428 | 1.436 | -0.966 | -0.007 | 3.34E-01 | 18581 | -4.308 | -8.453 | -0.163 | 2.115 | -2.037 | -0.015 | 4.16E-02 | 18733 |

| | | | | | | | | | | | | | | | | | |
|---|---|---|---|---|---|---|---|---|---|---|---|---|---|---|---|---|---|
| 94 | Temporal_Inf_R | -5.233 | -8.300 | -2.166 | 1.565 | -3.345 | -0.025 | 8.25E-04 | 18581 | -0.980 | -5.501 | 3.541 | 2.307 | -0.425 | -0.003 | 6.71E-01 | 18733 |
| 95 | Cerebellum_Crus1_L | -1.235 | -4.579 | 2.108 | 1.706 | -0.724 | -0.005 | 4.69E-01 | 18581 | -2.908 | -7.828 | 2.012 | 2.510 | -1.158 | -0.008 | 2.47E-01 | 18733 |
| 96 | Cerebellum_Crus1_R | 0.189 | -3.220 | 3.598 | 1.739 | 0.109 | 0.001 | 9.14E-01 | 18581 | -2.995 | -8.013 | 2.023 | 2.560 | -1.170 | -0.009 | 2.42E-01 | 18733 |
| 97 | Cerebellum_Crus2_L | -1.482 | -4.132 | 1.168 | 1.352 | -1.096 | -0.008 | 2.73E-01 | 18581 | -7.786 | -11.689 | -3.883 | 1.991 | -3.910 | -0.029 | 9.26E-05 | 18733 |
| 98 | Cerebellum_Crus2_R | -0.522 | -2.974 | 1.929 | 1.251 | -0.418 | -0.003 | 6.76E-01 | 18581 | -5.138 | -8.749 | -1.527 | 1.842 | -2.789 | -0.020 | 5.29E-03 | 18733 |
| 99 | Cerebellum_3_L | 0.065 | -0.113 | 0.243 | 0.091 | 0.714 | 0.005 | 4.75E-01 | 18581 | -0.056 | -0.318 | 0.206 | 0.134 | -0.417 | -0.003 | 6.77E-01 | 18733 |
| 100 | Cerebellum_3_R | 0.171 | -0.069 | 0.411 | 0.122 | 1.396 | 0.010 | 1.63E-01 | 18581 | -0.209 | -0.563 | 0.144 | 0.180 | -1.159 | -0.008 | 2.46E-01 | 18733 |
| 101 | Cerebellum_4_5_L | 0.392 | -1.047 | 1.831 | 0.734 | 0.534 | 0.004 | 5.93E-01 | 18581 | -1.618 | -3.737 | 0.501 | 1.081 | -1.497 | -0.011 | 1.35E-01 | 18733 |
| 102 | Cerebellum_4_5_R | 0.844 | -0.329 | 2.016 | 0.598 | 1.410 | 0.010 | 1.58E-01 | 18581 | -1.825 | -3.551 | -0.099 | 0.881 | -2.072 | -0.015 | 3.83E-02 | 18733 |
| 103 | Cerebellum_6_L | 0.046 | -2.442 | 2.534 | 1.269 | 0.036 | 0.000 | 9.71E-01 | 18581 | -3.619 | -7.281 | 0.042 | 1.868 | -1.937 | -0.014 | 5.27E-02 | 18733 |
| 104 | Cerebellum_6_R | 0.491 | -2.129 | 3.112 | 1.337 | 0.368 | 0.003 | 7.13E-01 | 18581 | -3.161 | -7.020 | 0.697 | 1.968 | -1.606 | -0.012 | 1.08E-01 | 18733 |
| 105 | Cerebellum_7b_L | -0.539 | -1.349 | 0.271 | 0.413 | -1.304 | -0.010 | 1.92E-01 | 18581 | -2.144 | -3.337 | -0.951 | 0.609 | -3.522 | -0.026 | 4.29E-04 | 18733 |
| 106 | Cerebellum_7b_R | -0.301 | -1.103 | 0.502 | 0.410 | -0.734 | -0.005 | 4.63E-01 | 18581 | -2.298 | -3.481 | -1.116 | 0.603 | -3.809 | -0.028 | 1.40E-04 | 18733 |
| 107 | Cerebellum_8_L | -1.526 | -4.097 | 1.045 | 1.312 | -1.163 | -0.009 | 2.45E-01 | 18581 | -1.553 | -5.341 | 2.236 | 1.933 | -0.803 | -0.006 | 4.22E-01 | 18733 |
| 108 | Cerebellum_8_R | -1.410 | -4.759 | 1.939 | 1.709 | -0.826 | -0.006 | 4.09E-01 | 18581 | -8.395 | -13.326 | -3.465 | 2.515 | -3.338 | -0.024 | 8.46E-04 | 18733 |
| 109 | Cerebellum_9_L | -0.563 | -2.015 | 0.890 | 0.741 | -0.759 | -0.006 | 4.48E-01 | 18581 | 1.890 | -0.252 | 4.032 | 1.093 | 1.730 | 0.013 | 8.37E-02 | 18733 |
| 110 | Cerebellum_9_R | -0.619 | -2.048 | 0.810 | 0.729 | -0.849 | -0.006 | 3.96E-01 | 18581 | 2.267 | 0.161 | 4.373 | 1.074 | 2.110 | 0.015 | 3.48E-02 | 18733 |
| 111 | Cerebellum_10_L | -0.047 | -0.151 | 0.056 | 0.053 | -0.900 | -0.007 | 3.68E-01 | 18581 | 0.294 | 0.142 | 0.446 | 0.078 | 3.786 | 0.028 | 1.53E-04 | 18733 |
| 112 | Cerebellum_10_R | -0.027 | -0.134 | 0.080 | 0.055 | -0.492 | -0.004 | 6.23E-01 | 18581 | 0.099 | -0.059 | 0.257 | 0.081 | 1.227 | 0.009 | 2.20E-01 | 18733 |
| 113 | Vermis_1_2 | 0.052 | -0.016 | 0.119 | 0.034 | 1.507 | 0.011 | 1.32E-01 | 18581 | -0.026 | -0.125 | 0.072 | 0.050 | -0.524 | -0.004 | 6.01E-01 | 18733 |
| 114 | Vermis_3 | 0.105 | -0.162 | 0.371 | 0.136 | 0.771 | 0.006 | 4.41E-01 | 18581 | -0.413 | -0.805 | -0.020 | 0.200 | -2.061 | -0.015 | 3.93E-02 | 18733 |
| 115 | Vermis_4_5 | 0.090 | -0.694 | 0.874 | 0.400 | 0.224 | 0.002 | 8.23E-01 | 18581 | -0.016 | -1.170 | 1.138 | 0.589 | -0.026 | 0.000 | 9.79E-01 | 18733 |
| 116 | Vermis_6 | -0.076 | -0.512 | 0.361 | 0.223 | -0.340 | -0.002 | 7.34E-01 | 18581 | 0.539 | -0.103 | 1.182 | 0.328 | 1.645 | 0.012 | 1.00E-01 | 18733 |
| 117 | Vermis_7 | -0.095 | -0.342 | 0.152 | 0.126 | -0.752 | -0.006 | 4.52E-01 | 18581 | 0.302 | -0.061 | 0.665 | 0.185 | 1.629 | 0.012 | 1.03E-01 | 18733 |
| 118 | Vermis_8 | -0.081 | -0.487 | 0.324 | 0.207 | -0.394 | -0.003 | 6.94E-01 | 18581 | 0.256 | -0.342 | 0.853 | 0.305 | 0.839 | 0.006 | 4.01E-01 | 18733 |
| 119 | Vermis_9 | -0.030 | -0.358 | 0.298 | 0.167 | -0.180 | -0.001 | 8.57E-01 | 18581 | 0.334 | -0.150 | 0.818 | 0.247 | 1.352 | 0.010 | 1.76E-01 | 18733 |
| 120 | Vermis_10 | 0.107 | 0.016 | 0.199 | 0.047 | 2.299 | 0.017 | 2.15E-02 | 18581 | 0.216 | 0.081 | 0.350 | 0.069 | 3.133 | 0.023 | 1.73E-03 | 18733 |
| 121 | Thal_AV_L | 0.010 | -0.027 | 0.047 | 0.019 | 0.544 | 0.004 | 5.87E-01 | 18581 | -0.085 | -0.139 | -0.031 | 0.028 | -3.067 | -0.022 | 2.16E-03 | 18733 |
| 122 | Thal_AV_R | 0.028 | -0.025 | 0.080 | 0.027 | 1.028 | 0.008 | 3.04E-01 | 18581 | -0.116 | -0.193 | -0.038 | 0.039 | -2.932 | -0.021 | 3.37E-03 | 18733 |
| 123 | Thal_LP_L | -0.004 | -0.044 | 0.037 | 0.021 | -0.180 | -0.001 | 8.57E-01 | 18581 | -0.129 | -0.189 | -0.069 | 0.031 | -4.208 | -0.031 | 2.58E-05 | 18733 |
| 124 | Thal_LP_R | 0.011 | -0.042 | 0.064 | 0.027 | 0.414 | 0.003 | 6.79E-01 | 18581 | -0.160 | -0.238 | -0.082 | 0.040 | -4.020 | -0.029 | 5.83E-05 | 18733 |
| 125 | Thal_VA_L | 0.016 | -0.027 | 0.060 | 0.022 | 0.729 | 0.005 | 4.66E-01 | 18581 | -0.044 | -0.108 | 0.019 | 0.033 | -1.365 | -0.010 | 1.72E-01 | 18733 |
| 126 | Thal_VA_R | 0.006 | -0.048 | 0.059 | 0.027 | 0.206 | 0.002 | 8.36E-01 | 18581 | -0.043 | -0.121 | 0.036 | 0.040 | -1.065 | -0.008 | 2.87E-01 | 18733 |
| 127 | Thal_VL_L | -0.068 | -0.191 | 0.056 | 0.063 | -1.075 | -0.008 | 2.82E-01 | 18581 | -0.151 | -0.332 | 0.031 | 0.093 | -1.622 | -0.012 | 1.05E-01 | 18733 |
| 128 | Thal_VL_R | -0.125 | -0.289 | 0.039 | 0.084 | -1.491 | -0.011 | 1.36E-01 | 18581 | -0.337 | -0.579 | -0.096 | 0.123 | -2.739 | -0.020 | 6.17E-03 | 18733 |
| 129 | Thal_VPL_L | -0.018 | -0.165 | 0.130 | 0.075 | -0.235 | -0.002 | 8.15E-01 | 18581 | -0.150 | -0.367 | 0.067 | 0.110 | -1.358 | -0.010 | 1.75E-01 | 18733 |
| 130 | Thal_VPL_R | -0.056 | -0.211 | 0.098 | 0.079 | -0.716 | -0.005 | 4.74E-01 | 18581 | -0.362 | -0.590 | -0.135 | 0.116 | -3.119 | -0.023 | 1.82E-03 | 18733 |
| 131 | Thal_IL_L | -0.053 | -0.117 | 0.010 | 0.032 | -1.644 | -0.012 | 1.00E-01 | 18581 | -0.231 | -0.324 | -0.137 | 0.048 | -4.828 | -0.035 | 1.39E-06 | 18733 |
| 132 | Thal_IL_R | -0.016 | -0.088 | 0.056 | 0.037 | -0.431 | -0.003 | 6.66E-01 | 18581 | -0.291 | -0.398 | -0.185 | 0.054 | -5.379 | -0.039 | 7.60E-08 | 18733 |
| 133 | Thal_Re_L | 0.000 | 0.000 | 0.000 | 0.000 | -0.111 | -0.001 | 9.12E-01 | 18581 | -0.001 | -0.001 | 0.000 | 0.000 | -2.950 | -0.022 | 3.18E-03 | 18733 |
| 134 | Thal_Re_R | 0.000 | -0.002 | 0.002 | 0.001 | 0.263 | 0.002 | 7.92E-01 | 18581 | -0.005 | -0.008 | -0.002 | 0.002 | -3.015 | -0.022 | 2.57E-03 | 18733 |
| 135 | Thal_MDm_L | -0.189 | -0.660 | 0.282 | 0.240 | -0.787 | -0.006 | 4.31E-01 | 18581 | -2.063 | -2.756 | -1.369 | 0.354 | -5.831 | -0.043 | 5.60E-09 | 18733 |
| 136 | Thal_MDm_R | 0.044 | -0.503 | 0.592 | 0.279 | 0.158 | 0.001 | 8.74E-01 | 18581 | -2.262 | -3.068 | -1.457 | 0.411 | -5.503 | -0.040 | 3.77E-08 | 18733 |
| 137 | Thal_MDl_L | -0.078 | -0.160 | 0.003 | 0.042 | -1.878 | -0.014 | 6.04E-02 | 18581 | -0.357 | -0.477 | -0.237 | 0.061 | -5.830 | -0.043 | 5.63E-09 | 18733 |
| 138 | Thal_MDl_R | -0.017 | -0.110 | 0.076 | 0.047 | -0.359 | -0.003 | 7.20E-01 | 18581 | -0.397 | -0.534 | -0.261 | 0.070 | -5.707 | -0.042 | 1.16E-08 | 18733 |
| 139 | Thal_LGN_L | 0.000 | -0.012 | 0.012 | 0.006 | 0.009 | 0.000 | 9.93E-01 | 18581 | -0.033 | -0.051 | -0.015 | 0.009 | -3.566 | -0.026 | 3.64E-04 | 18733 |
| 140 | Thal_LGN_R | 0.000 | -0.026 | 0.026 | 0.013 | 0.009 | 0.000 | 9.93E-01 | 18581 | -0.097 | -0.136 | -0.058 | 0.020 | -4.903 | -0.036 | 9.54E-07 | 18733 |
| 141 | Thal_MGN_L | -0.008 | -0.031 | 0.014 | 0.011 | -0.746 | -0.005 | 4.56E-01 | 18581 | -0.066 | -0.098 | -0.033 | 0.017 | -3.963 | -0.029 | 7.42E-05 | 18733 |

| | | | | | | | | | | | | | | | | | |
|---|---|---|---|---|---|---|---|---|---|---|---|---|---|---|---|---|---|
| 142 | Thal_MGN_R | 0.002 | -0.022 | 0.026 | 0.012 | 0.135 | 0.001 | 8.93E-01 | 18581 | -0.107 | -0.142 | -0.072 | 0.018 | -5.935 | -0.043 | 2.99E-09 | 18733 |
| 143 | Thal_PuA_L | -0.005 | -0.031 | 0.021 | 0.013 | -0.376 | -0.003 | 7.07E-01 | 18581 | -0.103 | -0.141 | -0.064 | 0.020 | -5.252 | -0.038 | 1.52E-07 | 18733 |
| 144 | Thal_PuA_R | 0.001 | -0.054 | 0.055 | 0.028 | 0.026 | 0.000 | 9.79E-01 | 18581 | -0.260 | -0.339 | -0.180 | 0.041 | -6.374 | -0.047 | 1.88E-10 | 18733 |
| 145 | Thal_PuM_L | -0.102 | -0.621 | 0.416 | 0.265 | -0.387 | -0.003 | 6.99E-01 | 18581 | -2.260 | -3.023 | -1.496 | 0.389 | -5.804 | -0.042 | 6.59E-09 | 18733 |
| 146 | Thal_PuM_R | -0.054 | -0.700 | 0.592 | 0.330 | -0.165 | -0.001 | 8.69E-01 | 18581 | -2.848 | -3.799 | -1.897 | 0.485 | -5.872 | -0.043 | 4.38E-09 | 18733 |
| 147 | Thal_PuL_L | -0.048 | -0.143 | 0.046 | 0.048 | -1.000 | -0.007 | 3.17E-01 | 18581 | -0.379 | -0.519 | -0.240 | 0.071 | -5.339 | -0.039 | 9.48E-08 | 18733 |
| 148 | Thal_PuL_R | -0.033 | -0.183 | 0.117 | 0.077 | -0.429 | -0.003 | 6.68E-01 | 18581 | -0.628 | -0.850 | -0.407 | 0.113 | -5.562 | -0.041 | 2.70E-08 | 18733 |
| 149 | Thal_PuI_L | 0.009 | -0.017 | 0.035 | 0.013 | 0.700 | 0.005 | 4.84E-01 | 18581 | -0.046 | -0.084 | -0.008 | 0.019 | -2.369 | -0.017 | 1.78E-02 | 18733 |
| 150 | Thal_PuI_R | 0.002 | -0.032 | 0.036 | 0.017 | 0.096 | 0.001 | 9.23E-01 | 18581 | -0.071 | -0.122 | -0.021 | 0.026 | -2.788 | -0.020 | 5.30E-03 | 18733 |
| 151 | ACC_sub_L | -0.290 | -0.587 | 0.007 | 0.151 | -1.915 | -0.014 | 5.54E-02 | 18581 | -0.605 | -1.042 | -0.168 | 0.223 | -2.715 | -0.020 | 6.63E-03 | 18733 |
| 152 | ACC_sub_R | -0.284 | -0.562 | -0.007 | 0.141 | -2.010 | -0.015 | 4.44E-02 | 18581 | -0.568 | -0.977 | -0.160 | 0.208 | -2.727 | -0.020 | 6.39E-03 | 18733 |
| 153 | ACC_pre_L | -0.940 | -1.700 | -0.179 | 0.388 | -2.421 | -0.018 | 1.55E-02 | 18581 | -2.752 | -3.872 | -1.632 | 0.571 | -4.816 | -0.035 | 1.47E-06 | 18733 |
| 154 | ACC_pre_R | -1.172 | -1.904 | -0.439 | 0.374 | -3.136 | -0.023 | 1.71E-03 | 18581 | -2.874 | -3.953 | -1.795 | 0.550 | -5.223 | -0.038 | 1.78E-07 | 18733 |
| 155 | ACC_sup_L | -1.516 | -2.343 | -0.690 | 0.422 | -3.596 | -0.026 | 3.24E-04 | 18581 | -3.556 | -4.773 | -2.340 | 0.621 | -5.729 | -0.042 | 1.03E-08 | 18733 |
| 156 | ACC_sup_R | -1.100 | -1.957 | -0.243 | 0.437 | -2.516 | -0.018 | 1.19E-02 | 18581 | -2.980 | -4.241 | -1.720 | 0.643 | -4.634 | -0.034 | 3.61E-06 | 18733 |
| 157 | N_Acc_L | -0.116 | -0.364 | 0.133 | 0.127 | -0.910 | -0.007 | 3.63E-01 | 18581 | -0.820 | -1.187 | -0.454 | 0.187 | -4.391 | -0.032 | 1.14E-05 | 18733 |
| 158 | N_Acc_R | -0.077 | -0.292 | 0.139 | 0.110 | -0.697 | -0.005 | 4.86E-01 | 18581 | -0.626 | -0.943 | -0.309 | 0.162 | -3.871 | -0.028 | 1.09E-04 | 18733 |
| 159 | VTA_L | 0.001 | -0.008 | 0.010 | 0.004 | 0.210 | 0.002 | 8.33E-01 | 18581 | -0.011 | -0.024 | 0.002 | 0.007 | -1.630 | -0.012 | 1.03E-01 | 18733 |
| 160 | VTA_R | 0.004 | -0.004 | 0.012 | 0.004 | 0.984 | 0.007 | 3.25E-01 | 18581 | -0.011 | -0.023 | 0.000 | 0.006 | -1.887 | -0.014 | 5.91E-02 | 18733 |
| 161 | SN_pc_L | -0.009 | -0.023 | 0.005 | 0.007 | -1.207 | -0.009 | 2.28E-01 | 18581 | -0.018 | -0.039 | 0.002 | 0.011 | -1.747 | -0.013 | 8.06E-02 | 18733 |
| 162 | SN_pc_R | -0.009 | -0.028 | 0.009 | 0.009 | -0.973 | -0.007 | 3.31E-01 | 18581 | -0.033 | -0.060 | -0.005 | 0.014 | -2.355 | -0.017 | 1.85E-02 | 18733 |
| 163 | SN_pr_L | -0.020 | -0.039 | -0.001 | 0.010 | -2.025 | -0.015 | 4.29E-02 | 18581 | -0.007 | -0.036 | 0.021 | 0.014 | -0.517 | -0.004 | 6.05E-01 | 18733 |
| 164 | SN_pr_R | -0.019 | -0.040 | 0.003 | 0.011 | -1.712 | -0.013 | 8.69E-02 | 18581 | -0.017 | -0.048 | 0.015 | 0.016 | -1.049 | -0.008 | 2.94E-01 | 18733 |
| 165 | Red_N_L | -0.002 | -0.025 | 0.022 | 0.012 | -0.136 | -0.001 | 8.92E-01 | 18581 | -0.069 | -0.104 | -0.034 | 0.018 | -3.900 | -0.028 | 9.65E-05 | 18733 |
| 166 | Red_N_R | -0.003 | -0.028 | 0.023 | 0.013 | -0.196 | -0.001 | 8.45E-01 | 18581 | -0.061 | -0.099 | -0.023 | 0.019 | -3.166 | -0.023 | 1.55E-03 | 18733 |
| 167 | LC_L | 0.011 | 0.002 | 0.020 | 0.004 | 2.427 | 0.018 | 1.52E-02 | 18581 | -0.001 | -0.014 | 0.012 | 0.007 | -0.159 | -0.001 | 8.73E-01 | 18733 |
| 168 | LC_R | 0.011 | 0.002 | 0.021 | 0.005 | 2.316 | 0.017 | 2.06E-02 | 18581 | 0.003 | -0.011 | 0.017 | 0.007 | 0.387 | 0.003 | 6.99E-01 | 18733 |
| 169 | Raphe_D | 0.007 | -0.027 | 0.041 | 0.017 | 0.418 | 0.003 | 6.76E-01 | 18581 | -0.145 | -0.196 | -0.095 | 0.026 | -5.677 | -0.041 | 1.39E-08 | 18733 |
| 170 | Raphe_M | 0.010 | 0.000 | 0.020 | 0.005 | 1.892 | 0.014 | 5.85E-02 | 18581 | -0.014 | -0.029 | 0.002 | 0.008 | -1.736 | -0.013 | 8.26E-02 | 18733 |

| | | Salad / raw vegetable intake | | | | | | | | Poultry intake | | | | | | | |
|---|---|---|---|---|---|---|---|---|---|---|---|---|---|---|---|---|---|
| Number | AAL 3 region | beta | 95%CI_l | 95%CI_h | se | t | r | p | df.e | beta | 95%CI_l | 95%CI_h | se | t | r | p | df.e |
| 0 | Total brain | -63.200 | -140.787 | 14.388 | 39.583 | -1.597 | -0.012 | 1.10E-01 | 18549 | -202.967 | -345.962 | -59.971 | 72.954 | -2.782 | -0.020 | 5.41E-03 | 18738 |
| 1 | Precentral_L | -2.369 | -5.098 | 0.360 | 1.392 | -1.702 | -0.012 | 8.88E-02 | 18549 | -4.794 | -9.827 | 0.239 | 2.568 | -1.867 | -0.014 | 6.19E-02 | 18738 |
| 2 | Precentral_R | -1.339 | -3.841 | 1.162 | 1.276 | -1.049 | -0.008 | 2.94E-01 | 18549 | -3.693 | -8.304 | 0.919 | 2.353 | -1.569 | -0.011 | 1.17E-01 | 18738 |
| 3 | Frontal_Sup_2_L | -5.290 | -8.173 | -2.407 | 1.471 | -3.597 | -0.026 | 3.23E-04 | 18549 | -5.368 | -10.680 | -0.055 | 2.710 | -1.980 | -0.014 | 4.77E-02 | 18738 |
| 4 | Frontal_Sup_2_R | -5.796 | -8.972 | -2.619 | 1.621 | -3.576 | -0.026 | 3.49E-04 | 18549 | -10.929 | -16.783 | -5.075 | 2.987 | -3.659 | -0.027 | 2.54E-04 | 18738 |
| 5 | Frontal_Mid_2_L | -4.977 | -8.003 | -1.951 | 1.544 | -3.224 | -0.024 | 1.27E-03 | 18549 | -11.076 | -16.654 | -5.499 | 2.846 | -3.892 | -0.028 | 9.96E-05 | 18738 |
| 6 | Frontal_Mid_2_R | -5.643 | -9.066 | -2.221 | 1.746 | -3.232 | -0.024 | 1.23E-03 | 18549 | -12.708 | -19.020 | -6.396 | 3.220 | -3.946 | -0.029 | 7.97E-05 | 18738 |
| 7 | Frontal_Inf_Oper_L | -0.122 | -1.016 | 0.773 | 0.456 | -0.267 | -0.002 | 7.90E-01 | 18549 | -1.831 | -3.480 | -0.182 | 0.841 | -2.177 | -0.016 | 2.95E-02 | 18738 |
| 8 | Frontal_Inf_Oper_R | -1.004 | -2.125 | 0.117 | 0.572 | -1.755 | -0.013 | 7.93E-02 | 18549 | -3.582 | -5.649 | -1.515 | 1.054 | -3.397 | -0.025 | 6.82E-04 | 18738 |
| 9 | Frontal_Inf_Tri_L | -1.590 | -3.490 | 0.311 | 0.970 | -1.639 | -0.012 | 1.01E-01 | 18549 | -5.218 | -8.720 | -1.717 | 1.786 | -2.921 | -0.021 | 3.49E-03 | 18738 |
| 10 | Frontal_Inf_Tri_R | -0.593 | -2.084 | 0.899 | 0.761 | -0.779 | -0.006 | 4.36E-01 | 18549 | -5.695 | -8.444 | -2.946 | 1.402 | -4.061 | -0.030 | 4.91E-05 | 18738 |
| 11 | Frontal_Inf_Orb_2_L | -0.233 | -0.801 | 0.336 | 0.290 | -0.802 | -0.006 | 4.23E-01 | 18549 | -1.412 | -2.459 | -0.366 | 0.534 | -2.645 | -0.019 | 8.17E-03 | 18738 |
| 12 | Frontal_Inf_Orb_2_R | -0.100 | -0.694 | 0.494 | 0.303 | -0.330 | -0.002 | 7.42E-01 | 18549 | -2.175 | -3.270 | -1.081 | 0.558 | -3.895 | -0.028 | 9.84E-05 | 18738 |
| 13 | Rolandic_Oper_L | -0.285 | -1.144 | 0.574 | 0.438 | -0.651 | -0.005 | 5.15E-01 | 18549 | -2.181 | -3.764 | -0.598 | 0.808 | -2.700 | -0.020 | 6.94E-03 | 18738 |
| 14 | Rolandic_Oper_R | -0.391 | -1.621 | 0.838 | 0.627 | -0.624 | -0.005 | 5.33E-01 | 18549 | -4.330 | -6.595 | -2.065 | 1.156 | -3.747 | -0.027 | 1.80E-04 | 18738 |
| 15 | Supp_Motor_Area_L | -2.880 | -4.919 | -0.841 | 1.040 | -2.769 | -0.020 | 5.64E-03 | 18549 | -3.726 | -7.489 | 0.038 | 1.920 | -1.940 | -0.014 | 5.24E-02 | 18738 |
| 16 | Supp_Motor_Area_R | -2.218 | -4.472 | 0.036 | 1.150 | -1.929 | -0.014 | 5.38E-02 | 18549 | -1.897 | -6.056 | 2.262 | 2.122 | -0.894 | -0.007 | 3.71E-01 | 18738 |

| | | | | | | | | | | | | | | | | | |
|---|---|---|---|---|---|---|---|---|---|---|---|---|---|---|---|---|---|
| 17 | Olfactory_L | -0.409 | -0.713 | -0.106 | 0.155 | -2.641 | -0.019 | 8.26E-03 | 18549 | -0.132 | -0.692 | 0.427 | 0.285 | -0.464 | -0.003 | 6.43E-01 | 18738 |
| 18 | Olfactory_R | -0.467 | -0.772 | -0.162 | 0.155 | -3.004 | -0.022 | 2.66E-03 | 18549 | -0.286 | -0.847 | 0.275 | 0.286 | -0.998 | -0.007 | 3.18E-01 | 18738 |
| 19 | Frontal_Sup_Medial_L | -2.252 | -4.166 | -0.338 | 0.977 | -2.306 | -0.017 | 2.11E-02 | 18549 | -8.542 | -12.067 | -5.018 | 1.798 | -4.750 | -0.035 | 2.05E-06 | 18738 |
| 20 | Frontal_Sup_Medial_R | -2.108 | -3.502 | -0.715 | 0.711 | -2.965 | -0.022 | 3.03E-03 | 18549 | -4.683 | -7.250 | -2.117 | 1.309 | -3.577 | -0.026 | 3.49E-04 | 18738 |
| 21 | Frontal_Med_Orb_L | -1.111 | -1.826 | -0.396 | 0.365 | -3.044 | -0.022 | 2.33E-03 | 18549 | -1.311 | -2.629 | 0.007 | 0.672 | -1.950 | -0.014 | 5.11E-02 | 18738 |
| 22 | Frontal_Med_Orb_R | -1.450 | -2.329 | -0.571 | 0.448 | -3.233 | -0.024 | 1.23E-03 | 18549 | -2.100 | -3.720 | -0.480 | 0.826 | -2.541 | -0.019 | 1.11E-02 | 18738 |
| 23 | Rectus_L | -0.990 | -1.730 | -0.249 | 0.378 | -2.621 | -0.019 | 8.78E-03 | 18549 | -1.420 | -2.784 | -0.057 | 0.696 | -2.042 | -0.015 | 4.12E-02 | 18738 |
| 24 | Rectus_R | -0.984 | -1.669 | -0.300 | 0.349 | -2.819 | -0.021 | 4.83E-03 | 18549 | -1.313 | -2.574 | -0.052 | 0.644 | -2.040 | -0.015 | 4.13E-02 | 18738 |
| 25 | OFCmed_L | -0.605 | -1.034 | -0.175 | 0.219 | -2.761 | -0.020 | 5.76E-03 | 18549 | -1.128 | -1.918 | -0.338 | 0.403 | -2.798 | -0.020 | 5.15E-03 | 18738 |
| 26 | OFCmed_R | -0.755 | -1.255 | -0.255 | 0.255 | -2.961 | -0.022 | 3.07E-03 | 18549 | -1.107 | -2.028 | -0.186 | 0.470 | -2.357 | -0.017 | 1.84E-02 | 18738 |
| 27 | OFCant_L | -0.735 | -1.232 | -0.239 | 0.253 | -2.904 | -0.021 | 3.68E-03 | 18549 | -0.539 | -1.453 | 0.376 | 0.466 | -1.155 | -0.008 | 2.48E-01 | 18738 |
| 28 | OFCant_R | -0.829 | -1.485 | -0.173 | 0.335 | -2.479 | -0.018 | 1.32E-02 | 18549 | -1.620 | -2.828 | -0.413 | 0.616 | -2.630 | -0.019 | 8.53E-03 | 18738 |
| 29 | OFCpost_L | -0.604 | -1.225 | 0.017 | 0.317 | -1.906 | -0.014 | 5.67E-02 | 18549 | -0.326 | -1.471 | 0.818 | 0.584 | -0.559 | -0.004 | 5.76E-01 | 18738 |
| 30 | OFCpost_R | -0.733 | -1.308 | -0.159 | 0.293 | -2.504 | -0.018 | 1.23E-02 | 18549 | -1.245 | -2.303 | -0.187 | 0.540 | -2.307 | -0.017 | 2.11E-02 | 18738 |
| 31 | OFClat_L | -0.079 | -0.289 | 0.131 | 0.107 | -0.738 | -0.005 | 4.61E-01 | 18549 | -0.689 | -1.075 | -0.303 | 0.197 | -3.501 | -0.026 | 4.65E-04 | 18738 |
| 32 | OFClat_R | -0.138 | -0.315 | 0.039 | 0.090 | -1.523 | -0.011 | 1.28E-01 | 18549 | -0.729 | -1.055 | -0.403 | 0.166 | -4.382 | -0.032 | 1.18E-05 | 18738 |
| 33 | Insula_L | -1.396 | -3.317 | 0.526 | 0.980 | -1.424 | -0.010 | 1.55E-01 | 18549 | -2.341 | -5.881 | 1.198 | 1.806 | -1.297 | -0.009 | 1.95E-01 | 18738 |
| 34 | Insula_R | -1.241 | -3.113 | 0.631 | 0.955 | -1.299 | -0.010 | 1.94E-01 | 18549 | -2.938 | -6.387 | 0.512 | 1.760 | -1.669 | -0.012 | 9.51E-02 | 18738 |
| 37 | Cingulate_Mid_L | -1.012 | -2.636 | 0.612 | 0.828 | -1.222 | -0.009 | 2.22E-01 | 18549 | -6.147 | -9.138 | -3.156 | 1.526 | -4.028 | -0.029 | 5.65E-05 | 18738 |
| 38 | Cingulate_Mid_R | -1.190 | -2.922 | 0.541 | 0.883 | -1.347 | -0.010 | 1.78E-01 | 18549 | -7.117 | -10.307 | -3.927 | 1.627 | -4.373 | -0.032 | 1.23E-05 | 18738 |
| 39 | Cingulate_Post_L | -0.125 | -0.622 | 0.371 | 0.253 | -0.496 | -0.004 | 6.20E-01 | 18549 | -0.961 | -1.875 | -0.047 | 0.466 | -2.061 | -0.015 | 3.94E-02 | 18738 |
| 40 | Cingulate_Post_R | -0.155 | -0.430 | 0.120 | 0.140 | -1.106 | -0.008 | 2.69E-01 | 18549 | -0.317 | -0.824 | 0.190 | 0.259 | -1.225 | -0.009 | 2.21E-01 | 18738 |
| 41 | Hippocampus_L | -0.314 | -1.116 | 0.489 | 0.409 | -0.766 | -0.006 | 4.44E-01 | 18549 | -1.542 | -3.021 | -0.064 | 0.754 | -2.045 | -0.015 | 4.09E-02 | 18738 |
| 42 | Hippocampus_R | -0.232 | -1.064 | 0.601 | 0.425 | -0.546 | -0.004 | 5.85E-01 | 18549 | -1.014 | -2.547 | 0.518 | 0.782 | -1.297 | -0.009 | 1.95E-01 | 18738 |
| 43 | ParaHippocampal_L | -0.014 | -0.726 | 0.698 | 0.363 | -0.039 | 0.000 | 9.69E-01 | 18549 | -1.320 | -2.634 | -0.006 | 0.670 | -1.969 | -0.014 | 4.90E-02 | 18738 |
| 44 | ParaHippocampal_R | -0.367 | -1.267 | 0.532 | 0.459 | -0.800 | -0.006 | 4.24E-01 | 18549 | -1.875 | -3.533 | -0.217 | 0.846 | -2.217 | -0.016 | 2.66E-02 | 18738 |
| 45 | Amygdala_L | -0.155 | -0.432 | 0.123 | 0.141 | -1.093 | -0.008 | 2.74E-01 | 18549 | -0.067 | -0.578 | 0.443 | 0.261 | -0.258 | -0.002 | 7.96E-01 | 18738 |
| 46 | Amygdala_R | -0.214 | -0.498 | 0.069 | 0.145 | -1.481 | -0.011 | 1.39E-01 | 18549 | -0.096 | -0.618 | 0.426 | 0.266 | -0.360 | -0.003 | 7.19E-01 | 18738 |
| 47 | Calcarine_L | 0.373 | -2.270 | 3.016 | 1.348 | 0.277 | 0.002 | 7.82E-01 | 18549 | -0.820 | -5.690 | 4.050 | 2.485 | -0.330 | -0.002 | 7.41E-01 | 18738 |
| 48 | Calcarine_R | 0.935 | -1.274 | 3.144 | 1.127 | 0.830 | 0.006 | 4.07E-01 | 18549 | -2.661 | -6.732 | 1.409 | 2.077 | -1.281 | -0.009 | 2.00E-01 | 18738 |
| 49 | Cuneus_L | -0.589 | -2.246 | 1.068 | 0.845 | -0.697 | -0.005 | 4.86E-01 | 18549 | 0.377 | -2.679 | 3.432 | 1.559 | 0.242 | 0.002 | 8.09E-01 | 18738 |
| 50 | Cuneus_R | 0.235 | -1.034 | 1.504 | 0.647 | 0.363 | 0.003 | 7.17E-01 | 18549 | -1.605 | -3.943 | 0.733 | 1.193 | -1.345 | -0.010 | 1.79E-01 | 18738 |
| 51 | Lingual_L | 0.428 | -1.162 | 2.018 | 0.811 | 0.527 | 0.004 | 5.98E-01 | 18549 | -2.167 | -5.095 | 0.761 | 1.494 | -1.450 | -0.011 | 1.47E-01 | 18738 |
| 52 | Lingual_R | -0.002 | -1.668 | 1.664 | 0.850 | -0.002 | 0.000 | 9.98E-01 | 18549 | -1.972 | -5.041 | 1.097 | 1.566 | -1.259 | -0.009 | 2.08E-01 | 18738 |
| 53 | Occipital_Sup_L | 0.065 | -1.029 | 1.158 | 0.558 | 0.116 | 0.001 | 9.08E-01 | 18549 | -2.468 | -4.481 | -0.455 | 1.027 | -2.403 | -0.018 | 1.63E-02 | 18738 |
| 54 | Occipital_Sup_R | 0.223 | -0.962 | 1.408 | 0.605 | 0.369 | 0.003 | 7.12E-01 | 18549 | -2.647 | -4.829 | -0.464 | 1.114 | -2.377 | -0.017 | 1.75E-02 | 18738 |
| 55 | Occipital_Mid_L | -0.276 | -2.839 | 2.286 | 1.307 | -0.211 | -0.002 | 8.33E-01 | 18549 | -1.343 | -6.064 | 3.378 | 2.409 | -0.558 | -0.004 | 5.77E-01 | 18738 |
| 56 | Occipital_Mid_R | -1.520 | -3.303 | 0.262 | 0.909 | -1.672 | -0.012 | 9.46E-02 | 18549 | 0.231 | -3.054 | 3.515 | 1.676 | 0.138 | 0.001 | 8.91E-01 | 18738 |
| 57 | Occipital_Inf_L | -0.024 | -0.875 | 0.827 | 0.434 | -0.055 | 0.000 | 9.56E-01 | 18549 | 0.966 | -0.601 | 2.533 | 0.799 | 1.208 | 0.009 | 2.27E-01 | 18738 |
| 58 | Occipital_Inf_R | -1.530 | -2.513 | -0.548 | 0.501 | -3.053 | -0.022 | 2.27E-03 | 18549 | 2.589 | 0.779 | 4.399 | 0.923 | 2.804 | 0.020 | 5.05E-03 | 18738 |
| 59 | Fusiform_L | 0.013 | -1.848 | 1.874 | 0.949 | 0.014 | 0.000 | 9.89E-01 | 18549 | 0.164 | -3.266 | 3.594 | 1.750 | 0.094 | 0.001 | 9.25E-01 | 18738 |
| 60 | Fusiform_R | 0.189 | -1.858 | 2.235 | 1.044 | 0.181 | 0.001 | 8.57E-01 | 18549 | -0.980 | -4.751 | 2.792 | 1.924 | -0.509 | -0.004 | 6.11E-01 | 18738 |
| 61 | Postcentral_L | -0.534 | -3.335 | 2.267 | 1.429 | -0.374 | -0.003 | 7.09E-01 | 18549 | -11.579 | -16.736 | -6.423 | 2.631 | -4.402 | -0.032 | 1.08E-05 | 18738 |
| 62 | Postcentral_R | 1.230 | -1.642 | 4.101 | 1.465 | 0.839 | 0.006 | 4.01E-01 | 18549 | -9.488 | -14.774 | -4.201 | 2.697 | -3.518 | -0.026 | 4.36E-04 | 18738 |
| 63 | Parietal_Sup_L | -0.549 | -2.163 | 1.064 | 0.823 | -0.667 | -0.005 | 5.05E-01 | 18549 | -3.459 | -6.431 | -0.487 | 1.516 | -2.282 | -0.017 | 2.25E-02 | 18738 |
| 64 | Parietal_Sup_R | 0.372 | -1.060 | 1.803 | 0.730 | 0.509 | 0.004 | 6.11E-01 | 18549 | -3.066 | -5.702 | -0.430 | 1.345 | -2.280 | -0.017 | 2.26E-02 | 18738 |
| 65 | Parietal_Inf_L | -1.130 | -3.243 | 0.983 | 1.078 | -1.048 | -0.008 | 2.95E-01 | 18549 | -3.846 | -7.737 | 0.045 | 1.985 | -1.937 | -0.014 | 5.27E-02 | 18738 |
| 66 | Parietal_Inf_R | -0.819 | -2.187 | 0.550 | 0.698 | -1.173 | -0.009 | 2.41E-01 | 18549 | -1.924 | -4.445 | 0.597 | 1.286 | -1.496 | -0.011 | 1.35E-01 | 18738 |

| | | | | | | | | | | | | | | | | | |
|---|---|---|---|---|---|---|---|---|---|---|---|---|---|---|---|---|---|
| 67 | SupraMarginal_L | -0.615 | -1.853 | 0.623 | 0.632 | -0.974 | -0.007 | 3.30E-01 | 18549 | -3.739 | -6.019 | -1.458 | 1.163 | -3.214 | -0.023 | 1.31E-03 | 18738 |
| 68 | SupraMarginal_R | -1.513 | -3.264 | 0.238 | 0.893 | -1.694 | -0.012 | 9.03E-02 | 18549 | -4.789 | -8.015 | -1.563 | 1.646 | -2.910 | -0.021 | 3.62E-03 | 18738 |
| 69 | Angular_L | -0.643 | -1.859 | 0.572 | 0.620 | -1.037 | -0.008 | 3.00E-01 | 18549 | 0.431 | -1.810 | 2.671 | 1.143 | 0.377 | 0.003 | 7.06E-01 | 18738 |
| 70 | Angular_R | -0.653 | -2.283 | 0.977 | 0.832 | -0.786 | -0.006 | 4.32E-01 | 18549 | -0.695 | -3.700 | 2.309 | 1.533 | -0.453 | -0.003 | 6.50E-01 | 18738 |
| 71 | Precuneus_L | -1.947 | -4.910 | 1.015 | 1.511 | -1.288 | -0.009 | 1.98E-01 | 18549 | -5.068 | -10.527 | 0.392 | 2.785 | -1.819 | -0.013 | 6.89E-02 | 18738 |
| 72 | Precuneus_R | -1.916 | -4.557 | 0.725 | 1.347 | -1.422 | -0.010 | 1.55E-01 | 18549 | -4.502 | -9.366 | 0.362 | 2.481 | -1.814 | -0.013 | 6.96E-02 | 18738 |
| 73 | Paracentral_Lobule_L | -0.509 | -1.865 | 0.847 | 0.692 | -0.736 | -0.005 | 4.62E-01 | 18549 | -1.105 | -3.605 | 1.396 | 1.276 | -0.866 | -0.006 | 3.87E-01 | 18738 |
| 74 | Paracentral_Lobule_R | 0.154 | -0.817 | 1.125 | 0.496 | 0.310 | 0.002 | 7.56E-01 | 18549 | -0.721 | -2.512 | 1.070 | 0.914 | -0.789 | -0.006 | 4.30E-01 | 18738 |
| 75 | Caudate_L | -1.126 | -2.211 | -0.042 | 0.553 | -2.035 | -0.015 | 4.18E-02 | 18549 | -2.689 | -4.686 | -0.692 | 1.019 | -2.639 | -0.019 | 8.31E-03 | 18738 |
| 76 | Caudate_R | -1.443 | -2.670 | -0.216 | 0.626 | -2.304 | -0.017 | 2.12E-02 | 18549 | -2.846 | -5.106 | -0.585 | 1.153 | -2.468 | -0.018 | 1.36E-02 | 18738 |
| 77 | Putamen_L | -0.027 | -1.674 | 1.621 | 0.840 | -0.032 | 0.000 | 9.75E-01 | 18549 | -0.646 | -3.679 | 2.388 | 1.548 | -0.417 | -0.003 | 6.77E-01 | 18738 |
| 78 | Putamen_R | -0.190 | -1.867 | 1.486 | 0.855 | -0.222 | -0.002 | 8.24E-01 | 18549 | -0.278 | -3.367 | 2.811 | 1.576 | -0.176 | -0.001 | 8.60E-01 | 18738 |
| 79 | Pallidum_L | 0.024 | -0.122 | 0.170 | 0.075 | 0.321 | 0.002 | 7.48E-01 | 18549 | -0.200 | -0.469 | 0.070 | 0.138 | -1.453 | -0.011 | 1.46E-01 | 18738 |
| 80 | Pallidum_R | 0.034 | -0.127 | 0.195 | 0.082 | 0.415 | 0.003 | 6.78E-01 | 18549 | -0.146 | -0.442 | 0.150 | 0.151 | -0.965 | -0.007 | 3.35E-01 | 18738 |
| 83 | Heschl_L | -0.117 | -0.413 | 0.180 | 0.151 | -0.770 | -0.006 | 4.41E-01 | 18549 | -0.600 | -1.147 | -0.053 | 0.279 | -2.151 | -0.016 | 3.15E-02 | 18738 |
| 84 | Heschl_R | -0.213 | -0.544 | 0.119 | 0.169 | -1.257 | -0.009 | 2.09E-01 | 18549 | -0.909 | -1.520 | -0.298 | 0.312 | -2.917 | -0.021 | 3.53E-03 | 18738 |
| 85 | Temporal_Sup_L | -1.295 | -3.438 | 0.847 | 1.093 | -1.185 | -0.009 | 2.36E-01 | 18549 | -6.253 | -10.199 | -2.306 | 2.013 | -3.106 | -0.023 | 1.90E-03 | 18738 |
| 86 | Temporal_Sup_R | -0.677 | -3.048 | 1.693 | 1.209 | -0.560 | -0.004 | 5.75E-01 | 18549 | -8.759 | -13.124 | -4.395 | 2.227 | -3.933 | -0.029 | 8.40E-05 | 18738 |
| 87 | Temporal_Pole_Sup_L | -1.333 | -2.347 | -0.319 | 0.517 | -2.577 | -0.019 | 9.98E-03 | 18549 | -0.858 | -2.729 | 1.012 | 0.954 | -0.900 | -0.007 | 3.68E-01 | 18738 |
| 88 | Temporal_Pole_Sup_R | -0.500 | -1.569 | 0.568 | 0.545 | -0.917 | -0.007 | 3.59E-01 | 18549 | -1.622 | -3.591 | 0.348 | 1.005 | -1.614 | -0.012 | 1.07E-01 | 18738 |
| 89 | Temporal_Mid_L | -1.954 | -5.527 | 1.618 | 1.823 | -1.072 | -0.008 | 2.84E-01 | 18549 | -5.161 | -11.742 | 1.419 | 3.357 | -1.537 | -0.011 | 1.24E-01 | 18738 |
| 90 | Temporal_Mid_R | -3.749 | -6.920 | -0.578 | 1.618 | -2.318 | -0.017 | 2.05E-02 | 18549 | -3.069 | -8.912 | 2.774 | 2.981 | -1.030 | -0.008 | 3.03E-01 | 18738 |
| 91 | Temporal_Pole_Mid_L | -0.173 | -1.026 | 0.679 | 0.435 | -0.399 | -0.003 | 6.90E-01 | 18549 | -0.657 | -2.229 | 0.914 | 0.802 | -0.820 | -0.006 | 4.12E-01 | 18738 |
| 92 | Temporal_Pole_Mid_R | -0.466 | -1.503 | 0.571 | 0.529 | -0.881 | -0.006 | 3.79E-01 | 18549 | -1.606 | -3.516 | 0.305 | 0.975 | -1.647 | -0.012 | 9.95E-02 | 18738 |
| 93 | Temporal_Inf_L | -1.032 | -3.487 | 1.424 | 1.253 | -0.823 | -0.006 | 4.10E-01 | 18549 | -0.710 | -5.233 | 3.813 | 2.308 | -0.308 | -0.002 | 7.58E-01 | 18738 |
| 94 | Temporal_Inf_R | -4.859 | -7.538 | -2.181 | 1.366 | -3.556 | -0.026 | 3.77E-04 | 18549 | 0.651 | -4.282 | 5.584 | 2.517 | 0.259 | 0.002 | 7.96E-01 | 18738 |
| 95 | Cerebellum_Crus1_L | 2.777 | -0.136 | 5.691 | 1.487 | 1.868 | 0.014 | 6.17E-02 | 18549 | 6.546 | 1.177 | 11.914 | 2.739 | 2.390 | 0.017 | 1.69E-02 | 18738 |
| 96 | Cerebellum_Crus1_R | 2.592 | -0.378 | 5.562 | 1.515 | 1.711 | 0.013 | 8.72E-02 | 18549 | 6.116 | 0.641 | 11.592 | 2.793 | 2.190 | 0.016 | 2.86E-02 | 18738 |
| 97 | Cerebellum_Crus2_L | 2.123 | -0.187 | 4.433 | 1.179 | 1.801 | 0.013 | 7.17E-02 | 18549 | 1.887 | -2.373 | 6.147 | 2.174 | 0.868 | 0.006 | 3.85E-01 | 18738 |
| 98 | Cerebellum_Crus2_R | 2.758 | 0.621 | 4.896 | 1.090 | 2.530 | 0.019 | 1.14E-02 | 18549 | 3.497 | -0.444 | 7.437 | 2.010 | 1.739 | 0.013 | 8.20E-02 | 18738 |
| 99 | Cerebellum_3_L | 0.101 | -0.055 | 0.256 | 0.079 | 1.269 | 0.009 | 2.04E-01 | 18549 | 0.479 | 0.194 | 0.765 | 0.146 | 3.287 | 0.024 | 1.01E-03 | 18738 |
| 100 | Cerebellum_3_R | 0.150 | -0.059 | 0.360 | 0.107 | 1.407 | 0.010 | 1.59E-01 | 18549 | 0.491 | 0.105 | 0.877 | 0.197 | 2.494 | 0.018 | 1.26E-02 | 18738 |
| 101 | Cerebellum_4_5_L | 1.456 | 0.202 | 2.711 | 0.640 | 2.276 | 0.017 | 2.28E-02 | 18549 | 1.828 | -0.484 | 4.140 | 1.180 | 1.550 | 0.011 | 1.21E-01 | 18738 |
| 102 | Cerebellum_4_5_R | 1.082 | 0.059 | 2.104 | 0.522 | 2.073 | 0.015 | 3.82E-02 | 18549 | 0.471 | -1.412 | 2.355 | 0.961 | 0.490 | 0.004 | 6.24E-01 | 18738 |
| 103 | Cerebellum_6_L | 2.685 | 0.515 | 4.855 | 1.107 | 2.426 | 0.018 | 1.53E-02 | 18549 | 4.033 | 0.037 | 8.028 | 2.038 | 1.978 | 0.014 | 4.79E-02 | 18738 |
| 104 | Cerebellum_6_R | 2.555 | 0.269 | 4.841 | 1.166 | 2.191 | 0.016 | 2.85E-02 | 18549 | 4.185 | -0.024 | 8.395 | 2.148 | 1.949 | 0.014 | 5.13E-02 | 18738 |
| 105 | Cerebellum_7b_L | 0.622 | -0.085 | 1.328 | 0.360 | 1.725 | 0.013 | 8.45E-02 | 18549 | 0.557 | -0.745 | 1.859 | 0.664 | 0.838 | 0.006 | 4.02E-01 | 18738 |
| 106 | Cerebellum_7b_R | 0.923 | 0.223 | 1.623 | 0.357 | 2.585 | 0.019 | 9.74E-03 | 18549 | 0.019 | -1.272 | 1.310 | 0.659 | 0.029 | 0.000 | 9.77E-01 | 18738 |
| 107 | Cerebellum_8_L | 1.695 | -0.547 | 3.937 | 1.144 | 1.482 | 0.011 | 1.38E-01 | 18549 | 4.832 | 0.699 | 8.964 | 2.108 | 2.291 | 0.017 | 2.19E-02 | 18738 |
| 108 | Cerebellum_8_R | 2.823 | -0.097 | 5.743 | 1.490 | 1.895 | 0.014 | 5.81E-02 | 18549 | 1.156 | -4.225 | 6.538 | 2.746 | 0.421 | 0.003 | 6.74E-01 | 18738 |
| 109 | Cerebellum_9_L | 1.494 | 0.225 | 2.762 | 0.647 | 2.308 | 0.017 | 2.10E-02 | 18549 | 5.111 | 2.775 | 7.447 | 1.192 | 4.288 | 0.031 | 1.81E-05 | 18738 |
| 110 | Cerebellum_9_R | 1.335 | 0.087 | 2.582 | 0.636 | 2.097 | 0.015 | 3.60E-02 | 18549 | 4.378 | 2.081 | 6.675 | 1.172 | 3.736 | 0.027 | 1.88E-04 | 18738 |
| 111 | Cerebellum_10_L | -0.001 | -0.091 | 0.088 | 0.046 | -0.032 | 0.000 | 9.75E-01 | 18549 | 0.357 | 0.191 | 0.523 | 0.085 | 4.222 | 0.031 | 2.43E-05 | 18738 |
| 112 | Cerebellum_10_R | 0.049 | -0.044 | 0.143 | 0.048 | 1.034 | 0.008 | 3.01E-01 | 18549 | 0.233 | 0.061 | 0.406 | 0.088 | 2.653 | 0.019 | 7.98E-03 | 18738 |
| 113 | Vermis_1_2 | 0.016 | -0.043 | 0.074 | 0.030 | 0.522 | 0.004 | 6.02E-01 | 18549 | 0.172 | 0.064 | 0.280 | 0.055 | 3.132 | 0.023 | 1.74E-03 | 18738 |
| 114 | Vermis_3 | 0.029 | -0.204 | 0.261 | 0.119 | 0.241 | 0.002 | 8.10E-01 | 18549 | 0.501 | 0.072 | 0.929 | 0.219 | 2.290 | 0.017 | 2.21E-02 | 18738 |
| 115 | Vermis_4_5 | 0.750 | 0.067 | 1.434 | 0.349 | 2.151 | 0.016 | 3.15E-02 | 18549 | 0.840 | -0.419 | 2.100 | 0.643 | 1.308 | 0.010 | 1.91E-01 | 18738 |
| 116 | Vermis_6 | 0.372 | -0.008 | 0.753 | 0.194 | 1.917 | 0.014 | 5.53E-02 | 18549 | 1.031 | 0.330 | 1.732 | 0.358 | 2.884 | 0.021 | 3.93E-03 | 18738 |

| | | | | | | | | | | | | | | | | | |
|---|---|---|---|---|---|---|---|---|---|---|---|---|---|---|---|---|---|
| 117 | Vermis_7 | 0.225 | 0.009 | 0.440 | 0.110 | 2.046 | 0.015 | 4.07E-02 | 18549 | 0.543 | 0.147 | 0.940 | 0.202 | 2.688 | 0.020 | 7.19E-03 | 18738 |
| 118 | Vermis_8 | 0.475 | 0.121 | 0.828 | 0.180 | 2.633 | 0.019 | 8.46E-03 | 18549 | 1.160 | 0.508 | 1.811 | 0.332 | 3.490 | 0.025 | 4.85E-04 | 18738 |
| 119 | Vermis_9 | 0.453 | 0.166 | 0.740 | 0.146 | 3.099 | 0.023 | 1.95E-03 | 18549 | 1.017 | 0.489 | 1.545 | 0.269 | 3.775 | 0.028 | 1.61E-04 | 18738 |
| 120 | Vermis_10 | 0.097 | 0.017 | 0.177 | 0.041 | 2.384 | 0.017 | 1.71E-02 | 18549 | 0.346 | 0.198 | 0.493 | 0.075 | 4.605 | 0.034 | 4.16E-06 | 18738 |
| 121 | Thal_AV_L | -0.023 | -0.055 | 0.009 | 0.016 | -1.421 | -0.010 | 1.55E-01 | 18549 | -0.050 | -0.109 | 0.009 | 0.030 | -1.646 | -0.012 | 9.97E-02 | 18738 |
| 122 | Thal_AV_R | -0.039 | -0.084 | 0.007 | 0.023 | -1.648 | -0.012 | 9.94E-02 | 18549 | -0.073 | -0.157 | 0.012 | 0.043 | -1.687 | -0.012 | 9.17E-02 | 18738 |
| 123 | Thal_LP_L | -0.015 | -0.051 | 0.020 | 0.018 | -0.841 | -0.006 | 4.01E-01 | 18549 | -0.097 | -0.163 | -0.032 | 0.033 | -2.914 | -0.021 | 3.57E-03 | 18738 |
| 124 | Thal_LP_R | -0.022 | -0.069 | 0.024 | 0.024 | -0.954 | -0.007 | 3.40E-01 | 18549 | -0.105 | -0.190 | -0.020 | 0.043 | -2.412 | -0.018 | 1.59E-02 | 18738 |
| 125 | Thal_VA_L | -0.030 | -0.068 | 0.008 | 0.019 | -1.553 | -0.011 | 1.21E-01 | 18549 | -0.015 | -0.084 | 0.055 | 0.036 | -0.411 | -0.003 | 6.81E-01 | 18738 |
| 126 | Thal_VA_R | -0.060 | -0.107 | -0.013 | 0.024 | -2.528 | -0.019 | 1.15E-02 | 18549 | -0.035 | -0.121 | 0.050 | 0.044 | -0.809 | -0.006 | 4.18E-01 | 18738 |
| 127 | Thal_VL_L | -0.150 | -0.258 | -0.042 | 0.055 | -2.734 | -0.020 | 6.27E-03 | 18549 | -0.139 | -0.337 | 0.060 | 0.101 | -1.371 | -0.010 | 1.70E-01 | 18738 |
| 128 | Thal_VL_R | -0.202 | -0.345 | -0.058 | 0.073 | -2.761 | -0.020 | 5.77E-03 | 18549 | -0.305 | -0.569 | -0.042 | 0.134 | -2.269 | -0.017 | 2.33E-02 | 18738 |
| 129 | Thal_VPL_L | 0.008 | -0.120 | 0.136 | 0.065 | 0.118 | 0.001 | 9.06E-01 | 18549 | -0.107 | -0.343 | 0.129 | 0.121 | -0.889 | -0.006 | 3.74E-01 | 18738 |
| 130 | Thal_VPL_R | 0.013 | -0.122 | 0.147 | 0.069 | 0.183 | 0.001 | 8.54E-01 | 18549 | -0.197 | -0.446 | 0.051 | 0.127 | -1.557 | -0.011 | 1.19E-01 | 18738 |
| 131 | Thal_IL_L | -0.023 | -0.078 | 0.033 | 0.028 | -0.810 | -0.006 | 4.18E-01 | 18549 | -0.167 | -0.270 | -0.065 | 0.052 | -3.207 | -0.023 | 1.34E-03 | 18738 |
| 132 | Thal_IL_R | -0.042 | -0.104 | 0.021 | 0.032 | -1.297 | -0.010 | 1.95E-01 | 18549 | -0.160 | -0.276 | -0.044 | 0.059 | -2.708 | -0.020 | 6.77E-03 | 18738 |
| 133 | Thal_Re_L | 0.000 | 0.000 | 0.000 | 0.000 | -0.074 | -0.001 | 9.41E-01 | 18549 | 0.000 | -0.001 | 0.000 | 0.000 | -0.564 | -0.004 | 5.72E-01 | 18738 |
| 134 | Thal_Re_R | -0.003 | -0.005 | -0.001 | 0.001 | -2.855 | -0.021 | 4.31E-03 | 18549 | -0.002 | -0.005 | 0.002 | 0.002 | -0.987 | -0.007 | 3.24E-01 | 18738 |
| 135 | Thal_MDm_L | -0.389 | -0.800 | 0.021 | 0.209 | -1.859 | -0.014 | 6.31E-02 | 18549 | -1.251 | -2.008 | -0.493 | 0.386 | -3.238 | -0.024 | 1.21E-03 | 18738 |
| 136 | Thal_MDm_R | -0.440 | -0.917 | 0.038 | 0.243 | -1.805 | -0.013 | 7.10E-02 | 18549 | -0.977 | -1.857 | -0.097 | 0.449 | -2.177 | -0.016 | 2.95E-02 | 18738 |
| 137 | Thal_MDl_L | -0.071 | -0.143 | 0.000 | 0.036 | -1.968 | -0.014 | 4.90E-02 | 18549 | -0.227 | -0.359 | -0.096 | 0.067 | -3.400 | -0.025 | 6.76E-04 | 18738 |
| 138 | Thal_MDl_R | -0.083 | -0.164 | -0.002 | 0.041 | -2.008 | -0.015 | 4.47E-02 | 18549 | -0.201 | -0.350 | -0.052 | 0.076 | -2.646 | -0.019 | 8.16E-03 | 18738 |
| 139 | Thal_LGN_L | -0.003 | -0.013 | 0.008 | 0.006 | -0.479 | -0.004 | 6.32E-01 | 18549 | -0.036 | -0.056 | -0.016 | 0.010 | -3.569 | -0.026 | 3.59E-04 | 18738 |
| 140 | Thal_LGN_R | -0.009 | -0.032 | 0.014 | 0.012 | -0.747 | -0.005 | 4.55E-01 | 18549 | -0.099 | -0.141 | -0.056 | 0.022 | -4.572 | -0.033 | 4.85E-06 | 18738 |
| 141 | Thal_MGN_L | -0.005 | -0.024 | 0.014 | 0.010 | -0.513 | -0.004 | 6.08E-01 | 18549 | -0.044 | -0.079 | -0.008 | 0.018 | -2.427 | -0.018 | 1.52E-02 | 18738 |
| 142 | Thal_MGN_R | 0.001 | -0.020 | 0.021 | 0.011 | 0.048 | 0.000 | 9.62E-01 | 18549 | -0.062 | -0.101 | -0.023 | 0.020 | -3.150 | -0.023 | 1.63E-03 | 18738 |
| 143 | Thal_PuA_L | 0.000 | -0.023 | 0.023 | 0.012 | -0.004 | 0.000 | 9.97E-01 | 18549 | -0.060 | -0.102 | -0.018 | 0.021 | -2.799 | -0.020 | 5.14E-03 | 18738 |
| 144 | Thal_PuA_R | 0.006 | -0.042 | 0.053 | 0.024 | 0.235 | 0.002 | 8.15E-01 | 18549 | -0.128 | -0.215 | -0.040 | 0.044 | -2.867 | -0.021 | 4.15E-03 | 18738 |
| 145 | Thal_PuM_L | 0.001 | -0.451 | 0.453 | 0.231 | 0.003 | 0.000 | 9.98E-01 | 18549 | -1.216 | -2.049 | -0.383 | 0.425 | -2.860 | -0.021 | 4.24E-03 | 18738 |
| 146 | Thal_PuM_R | 0.005 | -0.558 | 0.569 | 0.287 | 0.019 | 0.000 | 9.85E-01 | 18549 | -1.063 | -2.101 | -0.025 | 0.530 | -2.007 | -0.015 | 4.48E-02 | 18738 |
| 147 | Thal_PuL_L | -0.002 | -0.084 | 0.081 | 0.042 | -0.044 | 0.000 | 9.65E-01 | 18549 | -0.224 | -0.376 | -0.072 | 0.078 | -2.892 | -0.021 | 3.83E-03 | 18738 |
| 148 | Thal_PuL_R | -0.008 | -0.140 | 0.123 | 0.067 | -0.126 | -0.001 | 8.99E-01 | 18549 | -0.270 | -0.512 | -0.028 | 0.123 | -2.188 | -0.016 | 2.87E-02 | 18738 |
| 149 | Thal_PuI_L | 0.010 | -0.013 | 0.032 | 0.011 | 0.832 | 0.006 | 4.05E-01 | 18549 | -0.024 | -0.065 | 0.018 | 0.021 | -1.129 | -0.008 | 2.59E-01 | 18738 |
| 150 | Thal_PuI_R | 0.011 | -0.019 | 0.041 | 0.015 | 0.716 | 0.005 | 4.74E-01 | 18549 | -0.029 | -0.084 | 0.026 | 0.028 | -1.046 | -0.008 | 2.96E-01 | 18738 |
| 151 | ACC_sub_L | -0.446 | -0.705 | -0.187 | 0.132 | -3.379 | -0.025 | 7.30E-04 | 18549 | -0.463 | -0.940 | 0.014 | 0.243 | -1.902 | -0.014 | 5.72E-02 | 18738 |
| 152 | ACC_sub_R | -0.306 | -0.548 | -0.064 | 0.123 | -2.480 | -0.018 | 1.31E-02 | 18549 | -0.677 | -1.123 | -0.231 | 0.227 | -2.978 | -0.022 | 2.90E-03 | 18738 |
| 153 | ACC_pre_L | -1.371 | -2.034 | -0.707 | 0.339 | -4.048 | -0.030 | 5.18E-05 | 18549 | -1.808 | -3.030 | -0.585 | 0.624 | -2.898 | -0.021 | 3.76E-03 | 18738 |
| 154 | ACC_pre_R | -1.090 | -1.730 | -0.451 | 0.326 | -3.343 | -0.025 | 8.32E-04 | 18549 | -2.160 | -3.337 | -0.982 | 0.601 | -3.595 | -0.026 | 3.25E-04 | 18738 |
| 155 | ACC_sup_L | -1.344 | -2.066 | -0.623 | 0.368 | -3.653 | -0.027 | 2.60E-04 | 18549 | -2.549 | -3.877 | -1.220 | 0.678 | -3.760 | -0.027 | 1.70E-04 | 18738 |
| 156 | ACC_sup_R | -1.308 | -2.055 | -0.561 | 0.381 | -3.431 | -0.025 | 6.03E-04 | 18549 | -3.036 | -4.412 | -1.660 | 0.702 | -4.325 | -0.032 | 1.54E-05 | 18738 |
| 157 | N_Acc_L | -0.149 | -0.367 | 0.068 | 0.111 | -1.349 | -0.010 | 1.77E-01 | 18549 | -0.338 | -0.738 | 0.062 | 0.204 | -1.654 | -0.012 | 9.81E-02 | 18738 |
| 158 | N_Acc_R | -0.080 | -0.268 | 0.108 | 0.096 | -0.834 | -0.006 | 4.04E-01 | 18549 | -0.268 | -0.614 | 0.078 | 0.176 | -1.519 | -0.011 | 1.29E-01 | 18738 |
| 159 | VTA_L | 0.000 | -0.007 | 0.008 | 0.004 | 0.053 | 0.000 | 9.58E-01 | 18549 | -0.014 | -0.028 | 0.000 | 0.007 | -1.921 | -0.014 | 5.48E-02 | 18738 |
| 160 | VTA_R | 0.001 | -0.006 | 0.008 | 0.004 | 0.256 | 0.002 | 7.98E-01 | 18549 | -0.014 | -0.027 | -0.001 | 0.006 | -2.159 | -0.016 | 3.09E-02 | 18738 |
| 161 | SN_pc_L | 0.001 | -0.011 | 0.013 | 0.006 | 0.191 | 0.001 | 8.49E-01 | 18549 | -0.022 | -0.044 | 0.001 | 0.012 | -1.887 | -0.014 | 5.91E-02 | 18738 |
| 162 | SN_pc_R | -0.005 | -0.021 | 0.011 | 0.008 | -0.619 | -0.005 | 5.36E-01 | 18549 | -0.032 | -0.061 | -0.002 | 0.015 | -2.101 | -0.015 | 3.57E-02 | 18738 |
| 163 | SN_pr_L | -0.005 | -0.022 | 0.012 | 0.009 | -0.573 | -0.004 | 5.67E-01 | 18549 | -0.039 | -0.070 | -0.008 | 0.016 | -2.490 | -0.018 | 1.28E-02 | 18738 |
| 164 | SN_pr_R | -0.009 | -0.027 | 0.010 | 0.010 | -0.903 | -0.007 | 3.66E-01 | 18549 | -0.027 | -0.061 | 0.008 | 0.018 | -1.531 | -0.011 | 1.26E-01 | 18738 |

| | | | | | | | | | | | | | | | | | |
|---|---|---|---|---|---|---|---|---|---|---|---|---|---|---|---|---|---|
| 165 | Red_N_L | 0.000 | -0.021 | 0.020 | 0.011 | -0.039 | 0.000 | 9.69E-01 | 18549 | -0.067 | -0.105 | -0.030 | 0.019 | -3.490 | -0.025 | 4.85E-04 | 18738 |
| 166 | Red_N_R | -0.002 | -0.024 | 0.021 | 0.011 | -0.147 | -0.001 | 8.83E-01 | 18549 | -0.056 | -0.098 | -0.015 | 0.021 | -2.681 | -0.020 | 7.36E-03 | 18738 |
| 167 | LC_L | 0.001 | -0.007 | 0.008 | 0.004 | 0.179 | 0.001 | 8.58E-01 | 18549 | 0.008 | -0.006 | 0.022 | 0.007 | 1.112 | 0.008 | 2.66E-01 | 18738 |
| 168 | LC_R | -0.001 | -0.010 | 0.007 | 0.004 | -0.267 | -0.002 | 7.90E-01 | 18549 | 0.012 | -0.004 | 0.027 | 0.008 | 1.505 | 0.011 | 1.32E-01 | 18738 |
| 169 | Raphe_D | -0.006 | -0.036 | 0.024 | 0.015 | -0.400 | -0.003 | 6.89E-01 | 18549 | -0.068 | -0.123 | -0.013 | 0.028 | -2.438 | -0.018 | 1.48E-02 | 18738 |
| 170 | Raphe_M | -0.003 | -0.012 | 0.006 | 0.005 | -0.638 | -0.005 | 5.23E-01 | 18549 | -0.006 | -0.022 | 0.011 | 0.009 | -0.675 | -0.005 | 5.00E-01 | 18738 |

| | | Fresh fruit intake | | | | | | | | Beef intake | | | | | | | |
|---|---|---|---|---|---|---|---|---|---|---|---|---|---|---|---|---|---|
| Number | AAL 3 region | beta | 95%CI_l | 95%CI_h | se | t | r | p | df.e | beta | 95%CI_l | 95%CI_h | se | t | r | p | df.e |
| 0 | Total brain | 139.732 | 44.360 | 235.105 | 48.657 | 2.872 | 0.021 | 4.09E-03 | 18641 | -419.674 | -584.949 | -254.399 | 84.320 | -4.977 | -0.036 | 6.51E-07 | 18719 |
| 1 | Precentral_L | -3.823 | -7.180 | -0.465 | 1.713 | -2.231 | -0.016 | 2.57E-02 | 18641 | -8.725 | -14.547 | -2.903 | 2.970 | -2.938 | -0.021 | 3.31E-03 | 18719 |
| 2 | Precentral_R | 1.443 | -1.636 | 4.522 | 1.571 | 0.919 | 0.007 | 3.58E-01 | 18641 | -7.983 | -13.318 | -2.648 | 2.722 | -2.933 | -0.021 | 3.36E-03 | 18719 |
| 3 | Frontal_Sup_2_L | -0.038 | -3.583 | 3.507 | 1.809 | -0.021 | 0.000 | 9.83E-01 | 18641 | -8.582 | -14.730 | -2.435 | 3.136 | -2.736 | -0.020 | 6.22E-03 | 18719 |
| 4 | Frontal_Sup_2_R | -0.413 | -4.321 | 3.496 | 1.994 | -0.207 | -0.002 | 8.36E-01 | 18641 | -11.283 | -18.058 | -4.509 | 3.456 | -3.265 | -0.024 | 1.10E-03 | 18719 |
| 5 | Frontal_Mid_2_L | -2.022 | -5.746 | 1.702 | 1.900 | -1.064 | -0.008 | 2.87E-01 | 18641 | -10.304 | -16.756 | -3.852 | 3.292 | -3.130 | -0.023 | 1.75E-03 | 18719 |
| 6 | Frontal_Mid_2_R | 1.174 | -3.037 | 5.386 | 2.149 | 0.546 | 0.004 | 5.85E-01 | 18641 | -17.523 | -24.823 | -10.223 | 3.724 | -4.705 | -0.034 | 2.56E-06 | 18719 |
| 7 | Frontal_Inf_Oper_L | -0.230 | -1.330 | 0.870 | 0.561 | -0.409 | -0.003 | 6.82E-01 | 18641 | -3.074 | -4.981 | -1.166 | 0.973 | -3.158 | -0.023 | 1.59E-03 | 18719 |
| 8 | Frontal_Inf_Oper_R | 0.152 | -1.227 | 1.530 | 0.703 | 0.216 | 0.002 | 8.29E-01 | 18641 | -3.424 | -5.815 | -1.033 | 1.220 | -2.807 | -0.021 | 5.01E-03 | 18719 |
| 9 | Frontal_Inf_Tri_L | -0.592 | -2.929 | 1.744 | 1.192 | -0.497 | -0.004 | 6.19E-01 | 18641 | -7.582 | -11.632 | -3.532 | 2.066 | -3.670 | -0.027 | 2.44E-04 | 18719 |
| 10 | Frontal_Inf_Tri_R | -0.483 | -2.318 | 1.352 | 0.936 | -0.516 | -0.004 | 6.06E-01 | 18641 | -5.903 | -9.083 | -2.723 | 1.622 | -3.639 | -0.027 | 2.75E-04 | 18719 |
| 11 | Frontal_Inf_Orb_2_L | -0.298 | -0.996 | 0.400 | 0.356 | -0.838 | -0.006 | 4.02E-01 | 18641 | -1.110 | -2.321 | 0.101 | 0.618 | -1.797 | -0.013 | 7.24E-02 | 18719 |
| 12 | Frontal_Inf_Orb_2_R | 0.215 | -0.515 | 0.945 | 0.373 | 0.577 | 0.004 | 5.64E-01 | 18641 | -2.040 | -3.306 | -0.774 | 0.646 | -3.158 | -0.023 | 1.59E-03 | 18719 |
| 13 | Rolandic_Oper_L | 0.379 | -0.678 | 1.435 | 0.539 | 0.702 | 0.005 | 4.83E-01 | 18641 | -2.454 | -4.286 | -0.622 | 0.935 | -2.625 | -0.019 | 8.67E-03 | 18719 |
| 14 | Rolandic_Oper_R | 1.690 | 0.179 | 3.202 | 0.771 | 2.192 | 0.016 | 2.84E-02 | 18641 | -3.417 | -6.038 | -0.795 | 1.337 | -2.555 | -0.019 | 1.06E-02 | 18719 |
| 15 | Supp_Motor_Area_L | 2.194 | -0.319 | 4.707 | 1.282 | 1.711 | 0.013 | 8.71E-02 | 18641 | -5.935 | -10.289 | -1.580 | 2.221 | -2.672 | -0.020 | 7.56E-03 | 18719 |
| 16 | Supp_Motor_Area_R | 2.159 | -0.618 | 4.935 | 1.417 | 1.524 | 0.011 | 1.27E-01 | 18641 | -7.490 | -12.301 | -2.678 | 2.455 | -3.051 | -0.022 | 2.28E-03 | 18719 |
| 17 | Olfactory_L | 0.469 | 0.096 | 0.842 | 0.190 | 2.465 | 0.018 | 1.37E-02 | 18641 | -0.256 | -0.903 | 0.391 | 0.330 | -0.776 | -0.006 | 4.38E-01 | 18719 |
| 18 | Olfactory_R | 0.334 | -0.040 | 0.709 | 0.191 | 1.752 | 0.013 | 7.99E-02 | 18641 | -0.519 | -1.168 | 0.130 | 0.331 | -1.568 | -0.011 | 1.17E-01 | 18719 |
| 19 | Frontal_Sup_Medial_L | 3.526 | 1.173 | 5.880 | 1.201 | 2.937 | 0.022 | 3.32E-03 | 18641 | -10.375 | -14.453 | -6.296 | 2.081 | -4.986 | -0.036 | 6.22E-07 | 18719 |
| 20 | Frontal_Sup_Medial_R | 1.339 | -0.374 | 3.052 | 0.874 | 1.532 | 0.011 | 1.25E-01 | 18641 | -6.470 | -9.439 | -3.501 | 1.515 | -4.272 | -0.031 | 1.95E-05 | 18719 |
| 21 | Frontal_Med_Orb_L | 1.019 | 0.140 | 1.899 | 0.449 | 2.272 | 0.017 | 2.31E-02 | 18641 | -3.098 | -4.623 | -1.574 | 0.778 | -3.984 | -0.029 | 6.79E-05 | 18719 |
| 22 | Frontal_Med_Orb_R | 1.017 | -0.063 | 2.098 | 0.551 | 1.845 | 0.014 | 6.50E-02 | 18641 | -3.251 | -5.125 | -1.378 | 0.956 | -3.401 | -0.025 | 6.72E-04 | 18719 |
| 23 | Rectus_L | 1.035 | 0.125 | 1.945 | 0.464 | 2.230 | 0.016 | 2.58E-02 | 18641 | -2.540 | -4.117 | -0.962 | 0.805 | -3.156 | -0.023 | 1.60E-03 | 18719 |
| 24 | Rectus_R | 0.842 | 0.000 | 1.684 | 0.429 | 1.961 | 0.014 | 4.99E-02 | 18641 | -1.583 | -3.043 | -0.124 | 0.745 | -2.127 | -0.016 | 3.35E-02 | 18719 |
| 25 | OFCmed_L | 0.115 | -0.412 | 0.643 | 0.269 | 0.428 | 0.003 | 6.68E-01 | 18641 | -1.355 | -2.269 | -0.440 | 0.467 | -2.904 | -0.021 | 3.69E-03 | 18719 |
| 26 | OFCmed_R | -0.130 | -0.745 | 0.485 | 0.314 | -0.414 | -0.003 | 6.79E-01 | 18641 | -1.832 | -2.898 | -0.767 | 0.544 | -3.371 | -0.025 | 7.50E-04 | 18719 |
| 27 | OFCant_L | 0.009 | -0.601 | 0.619 | 0.311 | 0.029 | 0.000 | 9.77E-01 | 18641 | -0.993 | -2.050 | 0.065 | 0.539 | -1.840 | -0.013 | 6.57E-02 | 18719 |
| 28 | OFCant_R | -0.327 | -1.133 | 0.478 | 0.411 | -0.796 | -0.006 | 4.26E-01 | 18641 | -3.238 | -4.634 | -1.841 | 0.712 | -4.544 | -0.033 | 5.55E-06 | 18719 |
| 29 | OFCpost_L | 0.179 | -0.585 | 0.943 | 0.390 | 0.459 | 0.003 | 6.46E-01 | 18641 | -1.227 | -2.551 | 0.097 | 0.676 | -1.816 | -0.013 | 6.94E-02 | 18719 |
| 30 | OFCpost_R | 0.106 | -0.599 | 0.811 | 0.360 | 0.295 | 0.002 | 7.68E-01 | 18641 | -2.322 | -3.545 | -1.098 | 0.624 | -3.720 | -0.027 | 2.00E-04 | 18719 |
| 31 | OFClat_L | -0.148 | -0.405 | 0.110 | 0.131 | -1.125 | -0.008 | 2.61E-01 | 18641 | -0.606 | -1.053 | -0.159 | 0.228 | -2.660 | -0.019 | 7.83E-03 | 18719 |
| 32 | OFClat_R | 0.026 | -0.192 | 0.243 | 0.111 | 0.232 | 0.002 | 8.16E-01 | 18641 | -0.519 | -0.897 | -0.142 | 0.192 | -2.699 | -0.020 | 6.96E-03 | 18719 |
| 33 | Insula_L | 2.225 | -0.135 | 4.586 | 1.204 | 1.848 | 0.014 | 6.46E-02 | 18641 | -6.349 | -10.444 | -2.255 | 2.089 | -3.040 | -0.022 | 2.37E-03 | 18719 |
| 34 | Insula_R | 3.090 | 0.790 | 5.390 | 1.174 | 2.633 | 0.019 | 8.47E-03 | 18641 | -6.613 | -10.602 | -2.623 | 2.035 | -3.249 | -0.024 | 1.16E-03 | 18719 |
| 37 | Cingulate_Mid_L | 3.413 | 1.416 | 5.410 | 1.019 | 3.350 | 0.025 | 8.10E-04 | 18641 | -10.985 | -14.443 | -7.527 | 1.764 | -6.226 | -0.045 | 4.88E-10 | 18719 |
| 38 | Cingulate_Mid_R | 4.224 | 2.095 | 6.353 | 1.086 | 3.889 | 0.028 | 1.01E-04 | 18641 | -10.766 | -14.453 | -7.078 | 1.881 | -5.723 | -0.042 | 1.06E-08 | 18719 |
| 39 | Cingulate_Post_L | -0.397 | -1.007 | 0.213 | 0.311 | -1.275 | -0.009 | 2.02E-01 | 18641 | -2.075 | -3.132 | -1.017 | 0.539 | -3.847 | -0.028 | 1.20E-04 | 18719 |
| 40 | Cingulate_Post_R | -0.241 | -0.580 | 0.097 | 0.173 | -1.397 | -0.010 | 1.62E-01 | 18641 | -0.901 | -1.488 | -0.315 | 0.299 | -3.012 | -0.022 | 2.60E-03 | 18719 |
| 41 | Hippocampus_L | 1.026 | 0.039 | 2.013 | 0.504 | 2.037 | 0.015 | 4.17E-02 | 18641 | -1.993 | -3.703 | -0.283 | 0.872 | -2.284 | -0.017 | 2.24E-02 | 18719 |

| | | | | | | | | | | | | | | | | | |
|---|---|---|---|---|---|---|---|---|---|---|---|---|---|---|---|---|---|
| 42 | Hippocampus_R | 1.755 | 0.731 | 2.778 | 0.522 | 3.361 | 0.025 | 7.78E-04 | 18641 | -1.842 | -3.615 | -0.069 | 0.905 | -2.036 | -0.015 | 4.18E-02 | 18719 |
| 43 | ParaHippocampal_L | 1.623 | 0.746 | 2.500 | 0.447 | 3.628 | 0.027 | 2.86E-04 | 18641 | -1.794 | -3.313 | -0.275 | 0.775 | -2.314 | -0.017 | 2.07E-02 | 18719 |
| 44 | ParaHippocampal_R | 2.267 | 1.161 | 3.374 | 0.565 | 4.017 | 0.029 | 5.93E-05 | 18641 | -2.850 | -4.766 | -0.933 | 0.978 | -2.914 | -0.021 | 3.57E-03 | 18719 |
| 45 | Amygdala_L | 0.320 | -0.021 | 0.661 | 0.174 | 1.841 | 0.013 | 6.56E-02 | 18641 | 0.125 | -0.466 | 0.715 | 0.301 | 0.414 | 0.003 | 6.79E-01 | 18719 |
| 46 | Amygdala_R | 0.365 | 0.017 | 0.714 | 0.178 | 2.055 | 0.015 | 3.99E-02 | 18641 | -0.477 | -1.081 | 0.127 | 0.308 | -1.547 | -0.011 | 1.22E-01 | 18719 |
| 47 | Calcarine_L | -0.371 | -3.625 | 2.883 | 1.660 | -0.223 | -0.002 | 8.23E-01 | 18641 | -1.193 | -6.829 | 4.443 | 2.875 | -0.415 | -0.003 | 6.78E-01 | 18719 |
| 48 | Calcarine_R | 1.445 | -1.273 | 4.163 | 1.387 | 1.042 | 0.008 | 2.97E-01 | 18641 | -3.581 | -8.291 | 1.128 | 2.403 | -1.490 | -0.011 | 1.36E-01 | 18719 |
| 49 | Cuneus_L | 0.620 | -1.420 | 2.660 | 1.041 | 0.595 | 0.004 | 5.52E-01 | 18641 | -2.848 | -6.383 | 0.686 | 1.803 | -1.580 | -0.012 | 1.14E-01 | 18719 |
| 50 | Cuneus_R | 1.088 | -0.472 | 2.649 | 0.796 | 1.368 | 0.010 | 1.71E-01 | 18641 | -4.467 | -7.171 | -1.763 | 1.380 | -3.238 | -0.024 | 1.21E-03 | 18719 |
| 51 | Lingual_L | 1.607 | -0.350 | 3.563 | 0.998 | 1.610 | 0.012 | 1.07E-01 | 18641 | -2.968 | -6.356 | 0.421 | 1.729 | -1.717 | -0.013 | 8.60E-02 | 18719 |
| 52 | Lingual_R | 1.075 | -0.976 | 3.126 | 1.046 | 1.028 | 0.008 | 3.04E-01 | 18641 | -3.918 | -7.468 | -0.368 | 1.811 | -2.163 | -0.016 | 3.05E-02 | 18719 |
| 53 | Occipital_Sup_L | 0.929 | -0.415 | 2.274 | 0.686 | 1.355 | 0.010 | 1.75E-01 | 18641 | -3.649 | -5.978 | -1.320 | 1.188 | -3.071 | -0.022 | 2.13E-03 | 18719 |
| 54 | Occipital_Sup_R | 0.545 | -0.913 | 2.002 | 0.744 | 0.732 | 0.005 | 4.64E-01 | 18641 | -5.572 | -8.096 | -3.048 | 1.288 | -4.327 | -0.032 | 1.52E-05 | 18719 |
| 55 | Occipital_Mid_L | -0.458 | -3.608 | 2.692 | 1.607 | -0.285 | -0.002 | 7.76E-01 | 18641 | -8.472 | -13.932 | -3.013 | 2.785 | -3.042 | -0.022 | 2.36E-03 | 18719 |
| 56 | Occipital_Mid_R | -1.924 | -4.117 | 0.270 | 1.119 | -1.719 | -0.013 | 8.56E-02 | 18641 | -3.647 | -7.445 | 0.152 | 1.938 | -1.882 | -0.014 | 5.99E-02 | 18719 |
| 57 | Occipital_Inf_L | -0.370 | -1.417 | 0.676 | 0.534 | -0.693 | -0.005 | 4.88E-01 | 18641 | -0.634 | -2.446 | 1.179 | 0.925 | -0.685 | -0.005 | 4.93E-01 | 18719 |
| 58 | Occipital_Inf_R | -1.006 | -2.216 | 0.203 | 0.617 | -1.631 | -0.012 | 1.03E-01 | 18641 | 1.356 | -0.738 | 3.450 | 1.068 | 1.269 | 0.009 | 2.05E-01 | 18719 |
| 59 | Fusiform_L | 4.198 | 1.910 | 6.486 | 1.167 | 3.596 | 0.026 | 3.24E-04 | 18641 | -3.664 | -7.630 | 0.303 | 2.024 | -1.810 | -0.013 | 7.03E-02 | 18719 |
| 60 | Fusiform_R | 6.857 | 4.342 | 9.372 | 1.283 | 5.345 | 0.039 | 9.16E-08 | 18641 | -2.356 | -6.714 | 2.002 | 2.223 | -1.060 | -0.008 | 2.89E-01 | 18719 |
| 61 | Postcentral_L | 4.607 | 1.162 | 8.052 | 1.758 | 2.621 | 0.019 | 8.78E-03 | 18641 | -15.235 | -21.201 | -9.270 | 3.044 | -5.006 | -0.037 | 5.62E-07 | 18719 |
| 62 | Postcentral_R | 6.366 | 2.835 | 9.897 | 1.801 | 3.534 | 0.026 | 4.10E-04 | 18641 | -12.053 | -18.170 | -5.936 | 3.121 | -3.862 | -0.028 | 1.13E-04 | 18719 |
| 63 | Parietal_Sup_L | 0.871 | -1.113 | 2.856 | 1.012 | 0.861 | 0.006 | 3.89E-01 | 18641 | -3.306 | -6.745 | 0.132 | 1.754 | -1.885 | -0.014 | 5.95E-02 | 18719 |
| 64 | Parietal_Sup_R | 0.864 | -0.897 | 2.626 | 0.898 | 0.962 | 0.007 | 3.36E-01 | 18641 | -5.024 | -8.074 | -1.974 | 1.556 | -3.229 | -0.024 | 1.25E-03 | 18719 |
| 65 | Parietal_Inf_L | 3.636 | 1.037 | 6.235 | 1.326 | 2.742 | 0.020 | 6.12E-03 | 18641 | -10.661 | -15.162 | -6.161 | 2.296 | -4.644 | -0.034 | 3.45E-06 | 18719 |
| 66 | Parietal_Inf_R | 1.631 | -0.053 | 3.314 | 0.859 | 1.898 | 0.014 | 5.76E-02 | 18641 | -4.385 | -7.300 | -1.469 | 1.487 | -2.948 | -0.022 | 3.20E-03 | 18719 |
| 67 | SupraMarginal_L | 0.831 | -0.692 | 2.355 | 0.777 | 1.070 | 0.008 | 2.85E-01 | 18641 | -4.785 | -7.422 | -2.147 | 1.346 | -3.556 | -0.026 | 3.78E-04 | 18719 |
| 68 | SupraMarginal_R | 2.836 | 0.681 | 4.991 | 1.100 | 2.579 | 0.019 | 9.90E-03 | 18641 | -6.710 | -10.442 | -2.978 | 1.904 | -3.524 | -0.026 | 4.26E-04 | 18719 |
| 69 | Angular_L | 1.270 | -0.226 | 2.767 | 0.764 | 1.664 | 0.012 | 9.62E-02 | 18641 | -4.491 | -7.082 | -1.899 | 1.322 | -3.396 | -0.025 | 6.86E-04 | 18719 |
| 70 | Angular_R | -0.050 | -2.056 | 1.956 | 1.024 | -0.049 | 0.000 | 9.61E-01 | 18641 | -7.591 | -11.064 | -4.117 | 1.772 | -4.283 | -0.031 | 1.85E-05 | 18719 |
| 71 | Precuneus_L | 2.728 | -0.917 | 6.373 | 1.860 | 1.467 | 0.011 | 1.42E-01 | 18641 | -10.847 | -17.164 | -4.531 | 3.222 | -3.366 | -0.025 | 7.63E-04 | 18719 |
| 72 | Precuneus_R | 1.789 | -1.459 | 5.036 | 1.657 | 1.080 | 0.008 | 2.80E-01 | 18641 | -12.415 | -18.040 | -6.790 | 2.870 | -4.326 | -0.032 | 1.52E-05 | 18719 |
| 73 | Paracentral_Lobule_L | 0.436 | -1.234 | 2.105 | 0.852 | 0.512 | 0.004 | 6.09E-01 | 18641 | -2.253 | -5.146 | 0.640 | 1.476 | -1.526 | -0.011 | 1.27E-01 | 18719 |
| 74 | Paracentral_Lobule_R | 1.190 | -0.006 | 2.386 | 0.610 | 1.951 | 0.014 | 5.11E-02 | 18641 | -1.909 | -3.981 | 0.162 | 1.057 | -1.806 | -0.013 | 7.09E-02 | 18719 |
| 75 | Caudate_L | 0.119 | -1.214 | 1.451 | 0.680 | 0.175 | 0.001 | 8.61E-01 | 18641 | -2.870 | -5.181 | -0.558 | 1.179 | -2.434 | -0.018 | 1.50E-02 | 18719 |
| 76 | Caudate_R | -0.121 | -1.629 | 1.387 | 0.769 | -0.158 | -0.001 | 8.75E-01 | 18641 | -2.989 | -5.605 | -0.373 | 1.335 | -2.239 | -0.016 | 2.51E-02 | 18719 |
| 77 | Putamen_L | 0.753 | -1.270 | 2.775 | 1.032 | 0.730 | 0.005 | 4.66E-01 | 18641 | -2.446 | -5.955 | 1.063 | 1.790 | -1.366 | -0.010 | 1.72E-01 | 18719 |
| 78 | Putamen_R | 1.330 | -0.729 | 3.389 | 1.050 | 1.266 | 0.009 | 2.06E-01 | 18641 | -3.215 | -6.788 | 0.359 | 1.823 | -1.763 | -0.013 | 7.79E-02 | 18719 |
| 79 | Pallidum_L | 0.079 | -0.101 | 0.259 | 0.092 | 0.861 | 0.006 | 3.89E-01 | 18641 | -0.242 | -0.554 | 0.069 | 0.159 | -1.524 | -0.011 | 1.28E-01 | 18719 |
| 80 | Pallidum_R | 0.144 | -0.054 | 0.341 | 0.101 | 1.425 | 0.010 | 1.54E-01 | 18641 | -0.301 | -0.643 | 0.042 | 0.175 | -1.722 | -0.013 | 8.50E-02 | 18719 |
| 83 | Heschl_L | 0.017 | -0.348 | 0.381 | 0.186 | 0.089 | 0.001 | 9.29E-01 | 18641 | -1.097 | -1.730 | -0.465 | 0.323 | -3.399 | -0.025 | 6.77E-04 | 18719 |
| 84 | Heschl_R | 0.310 | -0.098 | 0.717 | 0.208 | 1.489 | 0.011 | 1.37E-01 | 18641 | -0.900 | -1.607 | -0.194 | 0.361 | -2.497 | -0.018 | 1.25E-02 | 18719 |
| 85 | Temporal_Sup_L | 1.345 | -1.288 | 3.979 | 1.344 | 1.001 | 0.007 | 3.17E-01 | 18641 | -9.516 | -14.081 | -4.951 | 2.329 | -4.086 | -0.030 | 4.41E-05 | 18719 |
| 86 | Temporal_Sup_R | 3.759 | 0.844 | 6.673 | 1.487 | 2.528 | 0.019 | 1.15E-02 | 18641 | -10.225 | -15.275 | -5.176 | 2.576 | -3.969 | -0.029 | 7.24E-05 | 18719 |
| 87 | Temporal_Pole_Sup_L | 0.841 | -0.406 | 2.089 | 0.636 | 1.322 | 0.010 | 1.86E-01 | 18641 | -2.115 | -4.278 | 0.048 | 1.103 | -1.916 | -0.014 | 5.53E-02 | 18719 |
| 88 | Temporal_Pole_Sup_R | 2.300 | 0.986 | 3.614 | 0.670 | 3.432 | 0.025 | 6.01E-04 | 18641 | -3.489 | -5.766 | -1.212 | 1.161 | -3.004 | -0.022 | 2.67E-03 | 18719 |
| 89 | Temporal_Mid_L | 4.755 | 0.360 | 9.150 | 2.242 | 2.121 | 0.016 | 3.40E-02 | 18641 | -16.233 | -23.840 | -8.625 | 3.881 | -4.182 | -0.031 | 2.90E-05 | 18719 |
| 90 | Temporal_Mid_R | 2.245 | -1.658 | 6.148 | 1.991 | 1.128 | 0.008 | 2.59E-01 | 18641 | -10.046 | -16.800 | -3.293 | 3.445 | -2.916 | -0.021 | 3.55E-03 | 18719 |
| 91 | Temporal_Pole_Mid_L | 0.924 | -0.126 | 1.973 | 0.535 | 1.725 | 0.013 | 8.46E-02 | 18641 | -1.159 | -2.977 | 0.658 | 0.927 | -1.250 | -0.009 | 2.11E-01 | 18719 |

| | | | | | | | | | | | | | | | | | |
|---|---|---|---|---|---|---|---|---|---|---|---|---|---|---|---|---|---|
| 92 | Temporal_Pole_Mid_R | 1.516 | 0.241 | 2.791 | 0.650 | 2.331 | 0.017 | 1.98E-02 | 18641 | -2.724 | -4.932 | -0.515 | 1.127 | -2.417 | -0.018 | 1.57E-02 | 18719 |
| 93 | Temporal_Inf_L | 1.688 | -1.331 | 4.707 | 1.540 | 1.096 | 0.008 | 2.73E-01 | 18641 | -2.569 | -7.797 | 2.658 | 2.667 | -0.963 | -0.007 | 3.35E-01 | 18719 |
| 94 | Temporal_Inf_R | 0.968 | -2.325 | 4.261 | 1.680 | 0.576 | 0.004 | 5.64E-01 | 18641 | -5.416 | -11.115 | 0.283 | 2.908 | -1.863 | -0.014 | 6.25E-02 | 18719 |
| 95 | Cerebellum_Crus1_L | 3.628 | 0.043 | 7.213 | 1.829 | 1.984 | 0.015 | 4.73E-02 | 18641 | 2.062 | -4.150 | 8.274 | 3.169 | 0.651 | 0.005 | 5.15E-01 | 18719 |
| 96 | Cerebellum_Crus1_R | 4.581 | 0.924 | 8.239 | 1.866 | 2.455 | 0.018 | 1.41E-02 | 18641 | 3.317 | -3.018 | 9.652 | 3.232 | 1.026 | 0.008 | 3.05E-01 | 18719 |
| 97 | Cerebellum_Crus2_L | 2.931 | 0.087 | 5.775 | 1.451 | 2.020 | 0.015 | 4.34E-02 | 18641 | -3.115 | -8.042 | 1.812 | 2.514 | -1.239 | -0.009 | 2.15E-01 | 18719 |
| 98 | Cerebellum_Crus2_R | 2.476 | -0.155 | 5.107 | 1.342 | 1.845 | 0.014 | 6.51E-02 | 18641 | -0.276 | -4.833 | 4.282 | 2.325 | -0.119 | -0.001 | 9.06E-01 | 18719 |
| 99 | Cerebellum_3_L | 0.310 | 0.120 | 0.501 | 0.097 | 3.187 | 0.023 | 1.44E-03 | 18641 | 0.221 | -0.110 | 0.552 | 0.169 | 1.309 | 0.010 | 1.91E-01 | 18719 |
| 100 | Cerebellum_3_R | 0.477 | 0.220 | 0.735 | 0.131 | 3.631 | 0.027 | 2.83E-04 | 18641 | 0.311 | -0.135 | 0.758 | 0.228 | 1.367 | 0.010 | 1.72E-01 | 18719 |
| 101 | Cerebellum_4_5_L | 1.894 | 0.350 | 3.438 | 0.788 | 2.404 | 0.018 | 1.62E-02 | 18641 | 1.620 | -1.056 | 4.295 | 1.365 | 1.186 | 0.009 | 2.35E-01 | 18719 |
| 102 | Cerebellum_4_5_R | 2.215 | 0.956 | 3.473 | 0.642 | 3.450 | 0.025 | 5.62E-04 | 18641 | 0.536 | -1.644 | 2.716 | 1.112 | 0.482 | 0.004 | 6.30E-01 | 18719 |
| 103 | Cerebellum_6_L | 3.420 | 0.752 | 6.088 | 1.361 | 2.513 | 0.018 | 1.20E-02 | 18641 | 2.726 | -1.898 | 7.349 | 2.359 | 1.155 | 0.008 | 2.48E-01 | 18719 |
| 104 | Cerebellum_6_R | 4.080 | 1.269 | 6.892 | 1.434 | 2.845 | 0.021 | 4.45E-03 | 18641 | 4.218 | -0.653 | 9.090 | 2.485 | 1.697 | 0.012 | 8.96E-02 | 18719 |
| 105 | Cerebellum_7b_L | 1.093 | 0.224 | 1.963 | 0.444 | 2.465 | 0.018 | 1.37E-02 | 18641 | -0.463 | -1.969 | 1.042 | 0.768 | -0.603 | -0.004 | 5.46E-01 | 18719 |
| 106 | Cerebellum_7b_R | 1.164 | 0.302 | 2.025 | 0.440 | 2.647 | 0.019 | 8.14E-03 | 18641 | -0.668 | -2.160 | 0.825 | 0.761 | -0.877 | -0.006 | 3.81E-01 | 18719 |
| 107 | Cerebellum_8_L | 3.271 | 0.511 | 6.031 | 1.408 | 2.323 | 0.017 | 2.02E-02 | 18641 | 4.525 | -0.257 | 9.306 | 2.440 | 1.855 | 0.014 | 6.37E-02 | 18719 |
| 108 | Cerebellum_8_R | 4.644 | 1.053 | 8.235 | 1.832 | 2.535 | 0.019 | 1.13E-02 | 18641 | -0.846 | -7.070 | 5.378 | 3.175 | -0.266 | -0.002 | 7.90E-01 | 18719 |
| 109 | Cerebellum_9_L | 0.445 | -1.115 | 2.005 | 0.796 | 0.560 | 0.004 | 5.76E-01 | 18641 | 3.799 | 1.095 | 6.504 | 1.380 | 2.754 | 0.020 | 5.90E-03 | 18719 |
| 110 | Cerebellum_9_R | -0.110 | -1.643 | 1.424 | 0.783 | -0.140 | -0.001 | 8.89E-01 | 18641 | 3.470 | 0.811 | 6.128 | 1.356 | 2.558 | 0.019 | 1.05E-02 | 18719 |
| 111 | Cerebellum_10_L | -0.091 | -0.201 | 0.020 | 0.057 | -1.602 | -0.012 | 1.09E-01 | 18641 | 0.386 | 0.195 | 0.578 | 0.098 | 3.947 | 0.029 | 7.94E-05 | 18719 |
| 112 | Cerebellum_10_R | -0.092 | -0.207 | 0.023 | 0.059 | -1.570 | -0.011 | 1.17E-01 | 18641 | 0.227 | 0.027 | 0.426 | 0.102 | 2.226 | 0.016 | 2.60E-02 | 18719 |
| 113 | Vermis_1_2 | 0.122 | 0.050 | 0.194 | 0.037 | 3.312 | 0.024 | 9.28E-04 | 18641 | 0.036 | -0.089 | 0.160 | 0.064 | 0.558 | 0.004 | 5.77E-01 | 18719 |
| 114 | Vermis_3 | 0.471 | 0.185 | 0.757 | 0.146 | 3.227 | 0.024 | 1.25E-03 | 18641 | 0.011 | -0.485 | 0.507 | 0.253 | 0.043 | 0.000 | 9.65E-01 | 18719 |
| 115 | Vermis_4_5 | 0.776 | -0.065 | 1.618 | 0.429 | 1.809 | 0.013 | 7.04E-02 | 18641 | 0.393 | -1.064 | 1.851 | 0.744 | 0.529 | 0.004 | 5.97E-01 | 18719 |
| 116 | Vermis_6 | 0.477 | 0.009 | 0.945 | 0.239 | 1.996 | 0.015 | 4.59E-02 | 18641 | 1.174 | 0.363 | 1.986 | 0.414 | 2.837 | 0.021 | 4.55E-03 | 18719 |
| 117 | Vermis_7 | 0.217 | -0.047 | 0.482 | 0.135 | 1.611 | 0.012 | 1.07E-01 | 18641 | 0.622 | 0.163 | 1.080 | 0.234 | 2.659 | 0.019 | 7.85E-03 | 18719 |
| 118 | Vermis_8 | 0.096 | -0.339 | 0.530 | 0.222 | 0.431 | 0.003 | 6.66E-01 | 18641 | 0.924 | 0.171 | 1.678 | 0.385 | 2.404 | 0.018 | 1.62E-02 | 18719 |
| 119 | Vermis_9 | 0.227 | -0.125 | 0.580 | 0.180 | 1.263 | 0.009 | 2.06E-01 | 18641 | 0.796 | 0.185 | 1.407 | 0.312 | 2.553 | 0.019 | 1.07E-02 | 18719 |
| 120 | Vermis_10 | 0.075 | -0.024 | 0.173 | 0.050 | 1.487 | 0.011 | 1.37E-01 | 18641 | 0.270 | 0.099 | 0.440 | 0.087 | 3.104 | 0.023 | 1.91E-03 | 18719 |
| 121 | Thal_AV_L | -0.018 | -0.058 | 0.021 | 0.020 | -0.905 | -0.007 | 3.65E-01 | 18641 | -0.117 | -0.185 | -0.049 | 0.035 | -3.365 | -0.025 | 7.67E-04 | 18719 |
| 122 | Thal_AV_R | 0.001 | -0.056 | 0.057 | 0.029 | 0.024 | 0.000 | 9.81E-01 | 18641 | -0.187 | -0.284 | -0.089 | 0.050 | -3.753 | -0.027 | 1.75E-04 | 18719 |
| 123 | Thal_LP_L | -0.003 | -0.047 | 0.040 | 0.022 | -0.157 | -0.001 | 8.75E-01 | 18641 | -0.183 | -0.259 | -0.107 | 0.039 | -4.742 | -0.035 | 2.14E-06 | 18719 |
| 124 | Thal_LP_R | -0.008 | -0.065 | 0.049 | 0.029 | -0.273 | -0.002 | 7.85E-01 | 18641 | -0.280 | -0.379 | -0.182 | 0.050 | -5.580 | -0.041 | 2.44E-08 | 18719 |
| 125 | Thal_VA_L | -0.034 | -0.080 | 0.013 | 0.024 | -1.427 | -0.010 | 1.53E-01 | 18641 | -0.036 | -0.117 | 0.045 | 0.041 | -0.872 | -0.006 | 3.83E-01 | 18719 |
| 126 | Thal_VA_R | -0.015 | -0.073 | 0.042 | 0.029 | -0.529 | -0.004 | 5.96E-01 | 18641 | -0.062 | -0.161 | 0.037 | 0.051 | -1.220 | -0.009 | 2.22E-01 | 18719 |
| 127 | Thal_VL_L | -0.116 | -0.248 | 0.017 | 0.068 | -1.715 | -0.013 | 8.63E-02 | 18641 | -0.386 | -0.616 | -0.157 | 0.117 | -3.301 | -0.024 | 9.66E-04 | 18719 |
| 128 | Thal_VL_R | -0.097 | -0.273 | 0.079 | 0.090 | -1.078 | -0.008 | 2.81E-01 | 18641 | -0.754 | -1.059 | -0.449 | 0.156 | -4.850 | -0.035 | 1.25E-06 | 18719 |
| 129 | Thal_VPL_L | -0.165 | -0.323 | -0.007 | 0.080 | -2.052 | -0.015 | 4.01E-02 | 18641 | -0.434 | -0.708 | -0.161 | 0.139 | -3.115 | -0.023 | 1.84E-03 | 18719 |
| 130 | Thal_VPL_R | -0.121 | -0.287 | 0.045 | 0.085 | -1.429 | -0.010 | 1.53E-01 | 18641 | -0.546 | -0.833 | -0.259 | 0.147 | -3.725 | -0.027 | 1.96E-04 | 18719 |
| 131 | Thal_IL_L | -0.029 | -0.097 | 0.039 | 0.035 | -0.835 | -0.006 | 4.04E-01 | 18641 | -0.330 | -0.448 | -0.211 | 0.060 | -5.468 | -0.040 | 4.62E-08 | 18719 |
| 132 | Thal_IL_R | 0.006 | -0.072 | 0.083 | 0.039 | 0.145 | 0.001 | 8.85E-01 | 18641 | -0.348 | -0.482 | -0.214 | 0.068 | -5.084 | -0.037 | 3.74E-07 | 18719 |
| 133 | Thal_Re_L | 0.000 | 0.000 | 0.000 | 0.000 | 0.534 | 0.004 | 5.93E-01 | 18641 | 0.000 | -0.001 | 0.000 | 0.000 | -1.687 | -0.012 | 9.17E-02 | 18719 |
| 134 | Thal_Re_R | 0.001 | -0.001 | 0.004 | 0.001 | 1.059 | 0.008 | 2.90E-01 | 18641 | -0.006 | -0.010 | -0.002 | 0.002 | -2.798 | -0.020 | 5.14E-03 | 18719 |
| 135 | Thal_MDm_L | -0.128 | -0.634 | 0.377 | 0.258 | -0.498 | -0.004 | 6.18E-01 | 18641 | -2.505 | -3.380 | -1.629 | 0.447 | -5.608 | -0.041 | 2.07E-08 | 18719 |
| 136 | Thal_MDm_R | 0.067 | -0.520 | 0.655 | 0.300 | 0.224 | 0.002 | 8.23E-01 | 18641 | -2.564 | -3.581 | -1.546 | 0.519 | -4.939 | -0.036 | 7.93E-07 | 18719 |
| 137 | Thal_MDl_L | -0.025 | -0.112 | 0.063 | 0.045 | -0.552 | -0.004 | 5.81E-01 | 18641 | -0.466 | -0.618 | -0.315 | 0.077 | -6.032 | -0.044 | 1.65E-09 | 18719 |
| 138 | Thal_MDl_R | 0.012 | -0.088 | 0.111 | 0.051 | 0.229 | 0.002 | 8.19E-01 | 18641 | -0.490 | -0.662 | -0.318 | 0.088 | -5.577 | -0.041 | 2.48E-08 | 18719 |
| 139 | Thal_LGN_L | 0.012 | -0.002 | 0.025 | 0.007 | 1.704 | 0.012 | 8.85E-02 | 18641 | -0.048 | -0.071 | -0.025 | 0.012 | -4.079 | -0.030 | 4.55E-05 | 18719 |

| | | | | | | | | | | | | | | | | | |
|---|---|---|---|---|---|---|---|---|---|---|---|---|---|---|---|---|---|
| 140 | Thal_LGN_R | 0.002 | -0.027 | 0.030 | 0.014 | 0.121 | 0.001 | 9.04E-01 | 18641 | -0.143 | -0.192 | -0.094 | 0.025 | -5.720 | -0.042 | 1.08E-08 | 18719 |
| 141 | Thal_MGN_L | 0.001 | -0.023 | 0.024 | 0.012 | 0.065 | 0.000 | 9.48E-01 | 18641 | -0.095 | -0.136 | -0.054 | 0.021 | -4.520 | -0.033 | 6.23E-06 | 18719 |
| 142 | Thal_MGN_R | 0.009 | -0.017 | 0.035 | 0.013 | 0.673 | 0.005 | 5.01E-01 | 18641 | -0.104 | -0.149 | -0.059 | 0.023 | -4.572 | -0.033 | 4.87E-06 | 18719 |
| 143 | Thal_PuA_L | 0.002 | -0.026 | 0.030 | 0.014 | 0.160 | 0.001 | 8.73E-01 | 18641 | -0.127 | -0.175 | -0.078 | 0.025 | -5.132 | -0.037 | 2.89E-07 | 18719 |
| 144 | Thal_PuA_R | 0.026 | -0.032 | 0.084 | 0.030 | 0.870 | 0.006 | 3.84E-01 | 18641 | -0.233 | -0.334 | -0.132 | 0.051 | -4.533 | -0.033 | 5.85E-06 | 18719 |
| 145 | Thal_PuM_L | -0.116 | -0.673 | 0.440 | 0.284 | -0.409 | -0.003 | 6.82E-01 | 18641 | -2.618 | -3.581 | -1.654 | 0.492 | -5.324 | -0.039 | 1.03E-07 | 18719 |
| 146 | Thal_PuM_R | 0.037 | -0.656 | 0.730 | 0.354 | 0.104 | 0.001 | 9.17E-01 | 18641 | -2.776 | -3.977 | -1.575 | 0.613 | -4.530 | -0.033 | 5.94E-06 | 18719 |
| 147 | Thal_PuL_L | -0.035 | -0.136 | 0.067 | 0.052 | -0.675 | -0.005 | 5.00E-01 | 18641 | -0.489 | -0.665 | -0.314 | 0.090 | -5.457 | -0.040 | 4.90E-08 | 18719 |
| 148 | Thal_PuL_R | -0.013 | -0.174 | 0.149 | 0.082 | -0.153 | -0.001 | 8.78E-01 | 18641 | -0.675 | -0.955 | -0.396 | 0.143 | -4.733 | -0.035 | 2.23E-06 | 18719 |
| 149 | Thal_PuI_L | -0.019 | -0.046 | 0.009 | 0.014 | -1.341 | -0.010 | 1.80E-01 | 18641 | -0.074 | -0.121 | -0.026 | 0.024 | -3.012 | -0.022 | 2.60E-03 | 18719 |
| 150 | Thal_PuI_R | -0.023 | -0.060 | 0.013 | 0.019 | -1.248 | -0.009 | 2.12E-01 | 18641 | -0.112 | -0.175 | -0.049 | 0.032 | -3.465 | -0.025 | 5.32E-04 | 18719 |
| 151 | ACC_sub_L | 0.132 | -0.186 | 0.450 | 0.162 | 0.814 | 0.006 | 4.16E-01 | 18641 | -0.195 | -0.747 | 0.356 | 0.281 | -0.694 | -0.005 | 4.88E-01 | 18719 |
| 152 | ACC_sub_R | 0.041 | -0.256 | 0.339 | 0.152 | 0.273 | 0.002 | 7.85E-01 | 18641 | -0.121 | -0.637 | 0.394 | 0.263 | -0.461 | -0.003 | 6.45E-01 | 18719 |
| 153 | ACC_pre_L | 1.128 | 0.312 | 1.943 | 0.416 | 2.710 | 0.020 | 6.73E-03 | 18641 | -2.289 | -3.704 | -0.875 | 0.721 | -3.173 | -0.023 | 1.51E-03 | 18719 |
| 154 | ACC_pre_R | 1.113 | 0.327 | 1.899 | 0.401 | 2.776 | 0.020 | 5.50E-03 | 18641 | -1.919 | -3.281 | -0.557 | 0.695 | -2.761 | -0.020 | 5.76E-03 | 18719 |
| 155 | ACC_sup_L | 0.734 | -0.152 | 1.621 | 0.452 | 1.623 | 0.012 | 1.05E-01 | 18641 | -2.532 | -4.069 | -0.995 | 0.784 | -3.229 | -0.024 | 1.24E-03 | 18719 |
| 156 | ACC_sup_R | 1.117 | 0.199 | 2.036 | 0.469 | 2.385 | 0.017 | 1.71E-02 | 18641 | -2.237 | -3.829 | -0.645 | 0.812 | -2.754 | -0.020 | 5.89E-03 | 18719 |
| 157 | N_Acc_L | 0.279 | 0.012 | 0.545 | 0.136 | 2.048 | 0.015 | 4.06E-02 | 18641 | -0.340 | -0.803 | 0.122 | 0.236 | -1.443 | -0.011 | 1.49E-01 | 18719 |
| 158 | N_Acc_R | 0.153 | -0.077 | 0.384 | 0.118 | 1.303 | 0.010 | 1.93E-01 | 18641 | -0.318 | -0.718 | 0.082 | 0.204 | -1.556 | -0.011 | 1.20E-01 | 18719 |
| 159 | VTA_L | 0.008 | -0.001 | 0.018 | 0.005 | 1.743 | 0.013 | 8.14E-02 | 18641 | -0.010 | -0.026 | 0.007 | 0.008 | -1.148 | -0.008 | 2.51E-01 | 18719 |
| 160 | VTA_R | 0.010 | 0.002 | 0.019 | 0.004 | 2.323 | 0.017 | 2.02E-02 | 18641 | -0.016 | -0.031 | -0.002 | 0.007 | -2.187 | -0.016 | 2.87E-02 | 18719 |
| 161 | SN_pc_L | 0.019 | 0.003 | 0.034 | 0.008 | 2.408 | 0.018 | 1.60E-02 | 18641 | -0.033 | -0.060 | -0.007 | 0.013 | -2.511 | -0.018 | 1.20E-02 | 18719 |
| 162 | SN_pc_R | 0.024 | 0.004 | 0.043 | 0.010 | 2.336 | 0.017 | 1.95E-02 | 18641 | -0.049 | -0.083 | -0.014 | 0.018 | -2.773 | -0.020 | 5.57E-03 | 18719 |
| 163 | SN_pr_L | 0.006 | -0.015 | 0.026 | 0.010 | 0.554 | 0.004 | 5.80E-01 | 18641 | -0.036 | -0.071 | -0.001 | 0.018 | -1.988 | -0.015 | 4.69E-02 | 18719 |
| 164 | SN_pr_R | 0.008 | -0.015 | 0.031 | 0.012 | 0.654 | 0.005 | 5.13E-01 | 18641 | -0.035 | -0.075 | 0.005 | 0.020 | -1.716 | -0.013 | 8.63E-02 | 18719 |
| 165 | Red_N_L | 0.031 | 0.006 | 0.056 | 0.013 | 2.395 | 0.018 | 1.66E-02 | 18641 | -0.065 | -0.109 | -0.021 | 0.022 | -2.916 | -0.021 | 3.54E-03 | 18719 |
| 166 | Red_N_R | 0.044 | 0.016 | 0.071 | 0.014 | 3.103 | 0.023 | 1.92E-03 | 18641 | -0.062 | -0.109 | -0.014 | 0.024 | -2.529 | -0.018 | 1.14E-02 | 18719 |
| 167 | LC_L | 0.012 | 0.003 | 0.022 | 0.005 | 2.533 | 0.019 | 1.13E-02 | 18641 | 0.004 | -0.013 | 0.020 | 0.008 | 0.425 | 0.003 | 6.71E-01 | 18719 |
| 168 | LC_R | 0.009 | -0.001 | 0.019 | 0.005 | 1.718 | 0.013 | 8.58E-02 | 18641 | 0.001 | -0.017 | 0.019 | 0.009 | 0.115 | 0.001 | 9.09E-01 | 18719 |
| 169 | Raphe_D | 0.041 | 0.005 | 0.078 | 0.019 | 2.216 | 0.016 | 2.67E-02 | 18641 | -0.138 | -0.202 | -0.075 | 0.032 | -4.276 | -0.031 | 1.91E-05 | 18719 |
| 170 | Raphe_M | 0.014 | 0.002 | 0.025 | 0.006 | 2.379 | 0.017 | 1.74E-02 | 18641 | -0.009 | -0.028 | 0.010 | 0.010 | -0.904 | -0.007 | 3.66E-01 | 18719 |

| | | Dried fruit intake | | | | | | | | Lamb/mutton intake | | | | | | | |
|---|---|---|---|---|---|---|---|---|---|---|---|---|---|---|---|---|---|
| **Number** | **AAL 3 region** | **beta** | **95%CI_l** | **95%CI_h** | **se** | **t** | **r** | **p** | **df.e** | **beta** | **95%CI_l** | **95%CI_h** | **se** | **t** | **r** | **p** | **df.e** |
| 0 | Total brain | 350.276 | 239.002 | 461.550 | 56.770 | 6.170 | 0.045 | 6.97E-10 | 18518 | -620.066 | -830.228 | -409.904 | 107.221 | -5.783 | -0.042 | 7.45E-09 | 18709 |
| 1 | Precentral_L | 4.505 | 0.585 | 8.424 | 2.000 | 2.253 | 0.017 | 2.43E-02 | 18518 | -12.945 | -20.349 | -5.541 | 3.777 | -3.427 | -0.025 | 6.12E-04 | 18709 |
| 2 | Precentral_R | 6.695 | 3.108 | 10.281 | 1.830 | 3.659 | 0.027 | 2.54E-04 | 18518 | -14.863 | -21.645 | -8.082 | 3.460 | -4.296 | -0.031 | 1.75E-05 | 18709 |
| 3 | Frontal_Sup_2_L | 10.064 | 5.932 | 14.196 | 2.108 | 4.774 | 0.035 | 1.82E-06 | 18518 | -13.057 | -20.872 | -5.243 | 3.987 | -3.275 | -0.024 | 1.06E-03 | 18709 |
| 4 | Frontal_Sup_2_R | 9.883 | 5.325 | 14.441 | 2.326 | 4.250 | 0.031 | 2.15E-05 | 18518 | -19.080 | -27.690 | -10.469 | 4.393 | -4.343 | -0.032 | 1.41E-05 | 18709 |
| 5 | Frontal_Mid_2_L | 9.767 | 5.425 | 14.110 | 2.215 | 4.409 | 0.032 | 1.05E-05 | 18518 | -14.133 | -22.340 | -5.927 | 4.187 | -3.376 | -0.025 | 7.38E-04 | 18709 |
| 6 | Frontal_Mid_2_R | 10.934 | 6.020 | 15.847 | 2.507 | 4.362 | 0.032 | 1.30E-05 | 18518 | -20.010 | -29.296 | -10.724 | 4.737 | -4.224 | -0.031 | 2.41E-05 | 18709 |
| 7 | Frontal_Inf_Oper_L | 2.732 | 1.449 | 4.015 | 0.655 | 4.173 | 0.031 | 3.02E-05 | 18518 | -3.864 | -6.290 | -1.439 | 1.237 | -3.123 | -0.023 | 1.79E-03 | 18709 |
| 8 | Frontal_Inf_Oper_R | 2.618 | 1.008 | 4.227 | 0.821 | 3.188 | 0.023 | 1.44E-03 | 18518 | -4.825 | -7.865 | -1.784 | 1.551 | -3.110 | -0.023 | 1.87E-03 | 18709 |
| 9 | Frontal_Inf_Tri_L | 6.402 | 3.676 | 9.128 | 1.391 | 4.603 | 0.034 | 4.19E-06 | 18518 | -7.824 | -12.975 | -2.673 | 2.628 | -2.977 | -0.022 | 2.91E-03 | 18709 |
| 10 | Frontal_Inf_Tri_R | 4.173 | 2.032 | 6.315 | 1.092 | 3.820 | 0.028 | 1.34E-04 | 18518 | -6.512 | -10.557 | -2.468 | 2.063 | -3.156 | -0.023 | 1.60E-03 | 18709 |
| 11 | Frontal_Inf_Orb_2_L | 2.576 | 1.761 | 3.391 | 0.416 | 6.194 | 0.045 | 5.98E-10 | 18518 | -0.837 | -2.378 | 0.703 | 0.786 | -1.066 | -0.008 | 2.87E-01 | 18709 |
| 12 | Frontal_Inf_Orb_2_R | 2.519 | 1.666 | 3.371 | 0.435 | 5.792 | 0.043 | 7.08E-09 | 18518 | -3.035 | -4.645 | -1.426 | 0.821 | -3.696 | -0.027 | 2.19E-04 | 18709 |
| 13 | Rolandic_Oper_L | 1.566 | 0.332 | 2.800 | 0.629 | 2.488 | 0.018 | 1.29E-02 | 18518 | -2.760 | -5.089 | -0.430 | 1.189 | -2.322 | -0.017 | 2.03E-02 | 18709 |
| 14 | Rolandic_Oper_R | 2.786 | 1.021 | 4.550 | 0.900 | 3.094 | 0.023 | 1.98E-03 | 18518 | -6.406 | -9.739 | -3.073 | 1.700 | -3.767 | -0.028 | 1.66E-04 | 18709 |

| | | | | | | | | | | | | | | | | | |
|---|---|---|---|---|---|---|---|---|---|---|---|---|---|---|---|---|---|
| 15 | Supp_Motor_Area_L | 3.359 | 0.431 | 6.287 | 1.494 | 2.249 | 0.017 | 2.45E-02 | 18518 | -9.605 | -15.139 | -4.071 | 2.823 | -3.402 | -0.025 | 6.71E-04 | 18709 |
| 16 | Supp_Motor_Area_R | 4.661 | 1.422 | 7.900 | 1.653 | 2.821 | 0.021 | 4.80E-03 | 18518 | -7.793 | -13.911 | -1.675 | 3.121 | -2.497 | -0.018 | 1.25E-02 | 18709 |
| 17 | Olfactory_L | 0.328 | -0.108 | 0.764 | 0.222 | 1.473 | 0.011 | 1.41E-01 | 18518 | -0.660 | -1.484 | 0.163 | 0.420 | -1.573 | -0.011 | 1.16E-01 | 18709 |
| 18 | Olfactory_R | 0.342 | -0.096 | 0.779 | 0.223 | 1.532 | 0.011 | 1.26E-01 | 18518 | -0.985 | -1.810 | -0.160 | 0.421 | -2.341 | -0.017 | 1.93E-02 | 18709 |
| 19 | Frontal_Sup_Medial_L | 6.920 | 4.175 | 9.666 | 1.401 | 4.941 | 0.036 | 7.84E-07 | 18518 | -13.549 | -18.735 | -8.363 | 2.646 | -5.121 | -0.037 | 3.07E-07 | 18709 |
| 20 | Frontal_Sup_Medial_R | 5.652 | 3.654 | 7.650 | 1.019 | 5.545 | 0.041 | 2.98E-08 | 18518 | -8.431 | -12.206 | -4.655 | 1.926 | -4.377 | -0.032 | 1.21E-05 | 18709 |
| 21 | Frontal_Med_Orb_L | 2.192 | 1.164 | 3.219 | 0.524 | 4.182 | 0.031 | 2.90E-05 | 18518 | -2.196 | -4.134 | -0.257 | 0.989 | -2.220 | -0.016 | 2.64E-02 | 18709 |
| 22 | Frontal_Med_Orb_R | 2.788 | 1.526 | 4.051 | 0.644 | 4.330 | 0.032 | 1.50E-05 | 18518 | -3.401 | -5.784 | -1.019 | 1.215 | -2.798 | -0.020 | 5.14E-03 | 18709 |
| 23 | Rectus_L | 2.473 | 1.410 | 3.535 | 0.542 | 4.561 | 0.033 | 5.13E-06 | 18518 | -2.990 | -4.997 | -0.984 | 1.024 | -2.922 | -0.021 | 3.49E-03 | 18709 |
| 24 | Rectus_R | 2.427 | 1.444 | 3.409 | 0.501 | 4.840 | 0.036 | 1.31E-06 | 18518 | -2.762 | -4.617 | -0.906 | 0.947 | -2.918 | -0.021 | 3.53E-03 | 18709 |
| 25 | OFCmed_L | 1.520 | 0.905 | 2.136 | 0.314 | 4.845 | 0.036 | 1.27E-06 | 18518 | -2.021 | -3.184 | -0.859 | 0.593 | -3.408 | -0.025 | 6.56E-04 | 18709 |
| 26 | OFCmed_R | 1.515 | 0.798 | 2.232 | 0.366 | 4.140 | 0.030 | 3.49E-05 | 18518 | -2.368 | -3.723 | -1.013 | 0.691 | -3.426 | -0.025 | 6.13E-04 | 18709 |
| 27 | OFCant_L | 1.263 | 0.551 | 1.975 | 0.363 | 3.477 | 0.026 | 5.09E-04 | 18518 | -1.837 | -3.183 | -0.492 | 0.686 | -2.678 | -0.020 | 7.42E-03 | 18709 |
| 28 | OFCant_R | 2.005 | 1.065 | 2.945 | 0.480 | 4.181 | 0.031 | 2.92E-05 | 18518 | -3.594 | -5.370 | -1.818 | 0.906 | -3.966 | -0.029 | 7.33E-05 | 18709 |
| 29 | OFCpost_L | 1.220 | 0.329 | 2.110 | 0.454 | 2.684 | 0.020 | 7.28E-03 | 18518 | -2.385 | -4.069 | -0.701 | 0.859 | -2.775 | -0.020 | 5.52E-03 | 18709 |
| 30 | OFCpost_R | 1.945 | 1.122 | 2.769 | 0.420 | 4.630 | 0.034 | 3.67E-06 | 18518 | -3.570 | -5.126 | -2.015 | 0.794 | -4.499 | -0.033 | 6.88E-06 | 18709 |
| 31 | OFClat_L | 0.777 | 0.477 | 1.078 | 0.153 | 5.067 | 0.037 | 4.09E-07 | 18518 | -0.688 | -1.256 | -0.120 | 0.290 | -2.373 | -0.017 | 1.76E-02 | 18709 |
| 32 | OFClat_R | 0.831 | 0.577 | 1.085 | 0.129 | 6.417 | 0.047 | 1.42E-10 | 18518 | -1.045 | -1.525 | -0.566 | 0.245 | -4.272 | -0.031 | 1.95E-05 | 18709 |
| 33 | Insula_L | 3.848 | 1.091 | 6.606 | 1.407 | 2.735 | 0.020 | 6.24E-03 | 18518 | -8.711 | -13.917 | -3.504 | 2.656 | -3.279 | -0.024 | 1.04E-03 | 18709 |
| 34 | Insula_R | 4.462 | 1.776 | 7.149 | 1.371 | 3.256 | 0.024 | 1.13E-03 | 18518 | -9.514 | -14.585 | -4.443 | 2.587 | -3.678 | -0.027 | 2.36E-04 | 18709 |
| 37 | Cingulate_Mid_L | 5.337 | 3.005 | 7.670 | 1.190 | 4.485 | 0.033 | 7.33E-06 | 18518 | -14.496 | -18.894 | -10.098 | 2.244 | -6.461 | -0.047 | 1.07E-10 | 18709 |
| 38 | Cingulate_Mid_R | 5.041 | 2.554 | 7.528 | 1.269 | 3.973 | 0.029 | 7.13E-05 | 18518 | -14.809 | -19.499 | -10.119 | 2.393 | -6.189 | -0.045 | 6.17E-10 | 18709 |
| 39 | Cingulate_Post_L | 0.382 | -0.330 | 1.093 | 0.363 | 1.052 | 0.008 | 2.93E-01 | 18518 | -3.633 | -4.977 | -2.289 | 0.686 | -5.300 | -0.039 | 1.17E-07 | 18709 |
| 40 | Cingulate_Post_R | -0.017 | -0.412 | 0.378 | 0.202 | -0.085 | -0.001 | 9.32E-01 | 18518 | -1.756 | -2.501 | -1.011 | 0.380 | -4.619 | -0.034 | 3.88E-06 | 18709 |
| 41 | Hippocampus_L | 1.912 | 0.761 | 3.062 | 0.587 | 3.255 | 0.024 | 1.13E-03 | 18518 | -0.566 | -2.740 | 1.609 | 1.110 | -0.510 | -0.004 | 6.10E-01 | 18709 |
| 42 | Hippocampus_R | 2.033 | 0.838 | 3.227 | 0.609 | 3.336 | 0.025 | 8.51E-04 | 18518 | 0.629 | -1.626 | 2.885 | 1.151 | 0.547 | 0.004 | 5.85E-01 | 18709 |
| 43 | ParaHippocampal_L | 2.707 | 1.684 | 3.731 | 0.522 | 5.185 | 0.038 | 2.18E-07 | 18518 | -0.900 | -2.832 | 1.033 | 0.986 | -0.913 | -0.007 | 3.61E-01 | 18709 |
| 44 | ParaHippocampal_R | 3.697 | 2.406 | 4.989 | 0.659 | 5.613 | 0.041 | 2.02E-08 | 18518 | -0.067 | -2.505 | 2.371 | 1.244 | -0.054 | 0.000 | 9.57E-01 | 18709 |
| 45 | Amygdala_L | 0.536 | 0.138 | 0.933 | 0.203 | 2.642 | 0.019 | 8.24E-03 | 18518 | -0.050 | -0.801 | 0.701 | 0.383 | -0.131 | -0.001 | 8.96E-01 | 18709 |
| 46 | Amygdala_R | 0.550 | 0.143 | 0.957 | 0.208 | 2.648 | 0.019 | 8.11E-03 | 18518 | -0.266 | -1.034 | 0.502 | 0.392 | -0.679 | -0.005 | 4.97E-01 | 18709 |
| 47 | Calcarine_L | -1.251 | -5.042 | 2.540 | 1.934 | -0.647 | -0.005 | 5.18E-01 | 18518 | -2.041 | -9.208 | 5.126 | 3.656 | -0.558 | -0.004 | 5.77E-01 | 18709 |
| 48 | Calcarine_R | -0.368 | -3.535 | 2.799 | 1.616 | -0.228 | -0.002 | 8.20E-01 | 18518 | -2.800 | -8.791 | 3.191 | 3.057 | -0.916 | -0.007 | 3.60E-01 | 18709 |
| 49 | Cuneus_L | -0.278 | -2.659 | 2.103 | 1.215 | -0.229 | -0.002 | 8.19E-01 | 18518 | -2.630 | -7.125 | 1.865 | 2.293 | -1.147 | -0.008 | 2.51E-01 | 18709 |
| 50 | Cuneus_R | 0.945 | -0.877 | 2.766 | 0.929 | 1.017 | 0.007 | 3.09E-01 | 18518 | -6.725 | -10.162 | -3.287 | 1.754 | -3.835 | -0.028 | 1.26E-04 | 18709 |
| 51 | Lingual_L | 1.456 | -0.823 | 3.736 | 1.163 | 1.252 | 0.009 | 2.11E-01 | 18518 | -6.321 | -10.629 | -2.014 | 2.198 | -2.877 | -0.021 | 4.03E-03 | 18709 |
| 52 | Lingual_R | 2.437 | 0.048 | 4.826 | 1.219 | 1.999 | 0.015 | 4.56E-02 | 18518 | -4.150 | -8.665 | 0.366 | 2.304 | -1.801 | -0.013 | 7.17E-02 | 18709 |
| 53 | Occipital_Sup_L | 1.361 | -0.206 | 2.929 | 0.800 | 1.702 | 0.013 | 8.87E-02 | 18518 | -4.339 | -7.301 | -1.378 | 1.511 | -2.872 | -0.021 | 4.09E-03 | 18709 |
| 54 | Occipital_Sup_R | 2.290 | 0.590 | 3.990 | 0.867 | 2.640 | 0.019 | 8.29E-03 | 18518 | -6.352 | -9.561 | -3.143 | 1.637 | -3.880 | -0.028 | 1.05E-04 | 18709 |
| 55 | Occipital_Mid_L | 2.933 | -0.746 | 6.612 | 1.877 | 1.563 | 0.011 | 1.18E-01 | 18518 | -11.493 | -18.435 | -4.551 | 3.542 | -3.245 | -0.024 | 1.18E-03 | 18709 |
| 56 | Occipital_Mid_R | 0.620 | -1.938 | 3.179 | 1.305 | 0.475 | 0.003 | 6.35E-01 | 18518 | -6.243 | -11.074 | -1.412 | 2.465 | -2.533 | -0.019 | 1.13E-02 | 18709 |
| 57 | Occipital_Inf_L | -0.498 | -1.719 | 0.723 | 0.623 | -0.799 | -0.006 | 4.24E-01 | 18518 | -1.508 | -3.814 | 0.797 | 1.176 | -1.282 | -0.009 | 2.00E-01 | 18709 |
| 58 | Occipital_Inf_R | -2.251 | -3.661 | -0.841 | 0.719 | -3.130 | -0.023 | 1.75E-03 | 18518 | -2.173 | -4.837 | 0.490 | 1.359 | -1.600 | -0.012 | 1.10E-01 | 18709 |
| 59 | Fusiform_L | 5.771 | 3.101 | 8.440 | 1.362 | 4.237 | 0.031 | 2.28E-05 | 18518 | -3.337 | -8.382 | 1.708 | 2.574 | -1.296 | -0.009 | 1.95E-01 | 18709 |
| 60 | Fusiform_R | 7.246 | 4.312 | 10.181 | 1.497 | 4.840 | 0.036 | 1.31E-06 | 18518 | -2.660 | -8.205 | 2.884 | 2.829 | -0.941 | -0.007 | 3.47E-01 | 18709 |
| 61 | Postcentral_L | 11.133 | 7.115 | 15.151 | 2.050 | 5.431 | 0.040 | 5.67E-08 | 18518 | -18.515 | -26.102 | -10.927 | 3.871 | -4.783 | -0.035 | 1.74E-06 | 18709 |
| 62 | Postcentral_R | 10.827 | 6.709 | 14.946 | 2.101 | 5.153 | 0.038 | 2.59E-07 | 18518 | -15.139 | -22.918 | -7.359 | 3.969 | -3.814 | -0.028 | 1.37E-04 | 18709 |
| 63 | Parietal_Sup_L | 1.137 | -1.178 | 3.451 | 1.181 | 0.963 | 0.007 | 3.36E-01 | 18518 | -7.028 | -11.400 | -2.656 | 2.231 | -3.151 | -0.023 | 1.63E-03 | 18709 |
| 64 | Parietal_Sup_R | 0.061 | -1.994 | 2.115 | 1.048 | 0.058 | 0.000 | 9.54E-01 | 18518 | -7.866 | -11.743 | -3.989 | 1.978 | -3.977 | -0.029 | 7.01E-05 | 18709 |

| | | | | | | | | | | | | | | | | | |
|---|---|---|---|---|---|---|---|---|---|---|---|---|---|---|---|---|---|
| 65 | Parietal_Inf_L | 3.370 | 0.340 | 6.399 | 1.546 | 2.180 | 0.016 | 2.93E-02 | 18518 | -16.800 | -22.522 | -11.078 | 2.919 | -5.755 | -0.042 | 8.82E-09 | 18709 |
| 66 | Parietal_Inf_R | 0.706 | -1.257 | 2.669 | 1.002 | 0.705 | 0.005 | 4.81E-01 | 18518 | -8.265 | -11.971 | -4.559 | 1.891 | -4.371 | -0.032 | 1.24E-05 | 18709 |
| 67 | SupraMarginal_L | 1.759 | -0.018 | 3.535 | 0.906 | 1.941 | 0.014 | 5.23E-02 | 18518 | -4.725 | -8.081 | -1.369 | 1.712 | -2.760 | -0.020 | 5.79E-03 | 18709 |
| 68 | SupraMarginal_R | 5.107 | 2.595 | 7.619 | 1.281 | 3.985 | 0.029 | 6.77E-05 | 18518 | -9.559 | -14.303 | -4.815 | 2.420 | -3.950 | -0.029 | 7.86E-05 | 18709 |
| 69 | Angular_L | 1.772 | 0.027 | 3.517 | 0.890 | 1.990 | 0.015 | 4.66E-02 | 18518 | -6.915 | -10.209 | -3.620 | 1.681 | -4.114 | -0.030 | 3.91E-05 | 18709 |
| 70 | Angular_R | -0.130 | -2.470 | 2.211 | 1.194 | -0.108 | -0.001 | 9.14E-01 | 18518 | -6.853 | -11.271 | -2.434 | 2.254 | -3.040 | -0.022 | 2.37E-03 | 18709 |
| 71 | Precuneus_L | 3.386 | -0.870 | 7.641 | 2.171 | 1.559 | 0.011 | 1.19E-01 | 18518 | -16.237 | -24.265 | -8.209 | 4.096 | -3.964 | -0.029 | 7.39E-05 | 18709 |
| 72 | Precuneus_R | 3.014 | -0.777 | 6.804 | 1.934 | 1.558 | 0.011 | 1.19E-01 | 18518 | -16.454 | -23.605 | -9.304 | 3.648 | -4.511 | -0.033 | 6.50E-06 | 18709 |
| 73 | Paracentral_Lobule_L | 2.249 | 0.303 | 4.195 | 0.993 | 2.265 | 0.017 | 2.35E-02 | 18518 | -4.256 | -7.935 | -0.577 | 1.877 | -2.268 | -0.017 | 2.34E-02 | 18709 |
| 74 | Paracentral_Lobule_R | 2.471 | 1.077 | 3.865 | 0.711 | 3.475 | 0.026 | 5.13E-04 | 18518 | -1.737 | -4.372 | 0.898 | 1.344 | -1.292 | -0.009 | 1.96E-01 | 18709 |
| 75 | Caudate_L | 2.567 | 1.013 | 4.121 | 0.793 | 3.237 | 0.024 | 1.21E-03 | 18518 | -2.076 | -5.015 | 0.862 | 1.499 | -1.385 | -0.010 | 1.66E-01 | 18709 |
| 76 | Caudate_R | 2.909 | 1.150 | 4.668 | 0.898 | 3.241 | 0.024 | 1.19E-03 | 18518 | -2.045 | -5.370 | 1.280 | 1.696 | -1.205 | -0.009 | 2.28E-01 | 18709 |
| 77 | Putamen_L | 3.285 | 0.922 | 5.647 | 1.205 | 2.725 | 0.020 | 6.43E-03 | 18518 | -7.330 | -11.790 | -2.870 | 2.275 | -3.221 | -0.024 | 1.28E-03 | 18709 |
| 78 | Putamen_R | 2.975 | 0.570 | 5.381 | 1.227 | 2.425 | 0.018 | 1.53E-02 | 18518 | -7.788 | -12.330 | -3.246 | 2.317 | -3.361 | -0.025 | 7.78E-04 | 18709 |
| 79 | Pallidum_L | 0.236 | 0.026 | 0.446 | 0.107 | 2.207 | 0.016 | 2.73E-02 | 18518 | -0.612 | -1.009 | -0.216 | 0.202 | -3.027 | -0.022 | 2.47E-03 | 18709 |
| 80 | Pallidum_R | 0.247 | 0.017 | 0.477 | 0.118 | 2.101 | 0.015 | 3.56E-02 | 18518 | -0.702 | -1.138 | -0.267 | 0.222 | -3.162 | -0.023 | 1.57E-03 | 18709 |
| 83 | Heschl_L | 0.287 | -0.140 | 0.713 | 0.217 | 1.318 | 0.010 | 1.88E-01 | 18518 | -1.387 | -2.191 | -0.583 | 0.410 | -3.380 | -0.025 | 7.26E-04 | 18709 |
| 84 | Heschl_R | 0.458 | -0.018 | 0.934 | 0.243 | 1.885 | 0.014 | 5.94E-02 | 18518 | -1.725 | -2.623 | -0.827 | 0.458 | -3.764 | -0.028 | 1.68E-04 | 18709 |
| 85 | Temporal_Sup_L | 5.066 | 1.994 | 8.139 | 1.568 | 3.232 | 0.024 | 1.23E-03 | 18518 | -8.821 | -14.626 | -3.015 | 2.962 | -2.978 | -0.022 | 2.90E-03 | 18709 |
| 86 | Temporal_Sup_R | 9.073 | 5.673 | 12.474 | 1.735 | 5.230 | 0.038 | 1.71E-07 | 18518 | -15.772 | -22.191 | -9.353 | 3.275 | -4.816 | -0.035 | 1.48E-06 | 18709 |
| 87 | Temporal_Pole_Sup_L | 2.248 | 0.790 | 3.707 | 0.744 | 3.022 | 0.022 | 2.52E-03 | 18518 | -3.921 | -6.672 | -1.171 | 1.403 | -2.795 | -0.020 | 5.20E-03 | 18709 |
| 88 | Temporal_Pole_Sup_R | 3.418 | 1.883 | 4.953 | 0.783 | 4.366 | 0.032 | 1.27E-05 | 18518 | -4.887 | -7.782 | -1.992 | 1.477 | -3.309 | -0.024 | 9.38E-04 | 18709 |
| 89 | Temporal_Mid_L | 12.123 | 7.003 | 17.244 | 2.612 | 4.641 | 0.034 | 3.49E-06 | 18518 | -17.731 | -27.404 | -8.057 | 4.935 | -3.593 | -0.026 | 3.28E-04 | 18709 |
| 90 | Temporal_Mid_R | 8.683 | 4.134 | 13.233 | 2.321 | 3.741 | 0.027 | 1.84E-04 | 18518 | -17.648 | -26.231 | -9.065 | 4.379 | -4.030 | -0.029 | 5.59E-05 | 18709 |
| 91 | Temporal_Pole_Mid_L | 1.923 | 0.698 | 3.149 | 0.625 | 3.077 | 0.023 | 2.10E-03 | 18518 | -1.593 | -3.906 | 0.719 | 1.180 | -1.351 | -0.010 | 1.77E-01 | 18709 |
| 92 | Temporal_Pole_Mid_R | 3.059 | 1.571 | 4.548 | 0.759 | 4.029 | 0.030 | 5.63E-05 | 18518 | -2.291 | -5.100 | 0.518 | 1.433 | -1.599 | -0.012 | 1.10E-01 | 18709 |
| 93 | Temporal_Inf_L | 3.646 | 0.124 | 7.167 | 1.797 | 2.029 | 0.015 | 4.24E-02 | 18518 | -6.857 | -13.504 | -0.209 | 3.392 | -2.022 | -0.015 | 4.32E-02 | 18709 |
| 94 | Temporal_Inf_R | 3.311 | -0.531 | 7.154 | 1.960 | 1.689 | 0.012 | 9.12E-02 | 18518 | -7.074 | -14.323 | 0.174 | 3.698 | -1.913 | -0.014 | 5.58E-02 | 18709 |
| 95 | Cerebellum_Crus1_L | 3.275 | -0.907 | 7.456 | 2.133 | 1.535 | 0.011 | 1.25E-01 | 18518 | 5.248 | -2.650 | 13.146 | 4.029 | 1.302 | 0.010 | 1.93E-01 | 18709 |
| 96 | Cerebellum_Crus1_R | 4.651 | 0.387 | 8.915 | 2.175 | 2.138 | 0.016 | 3.25E-02 | 18518 | 3.034 | -5.024 | 11.092 | 4.111 | 0.738 | 0.005 | 4.61E-01 | 18709 |
| 97 | Cerebellum_Crus2_L | 7.523 | 4.210 | 10.835 | 1.690 | 4.451 | 0.033 | 8.59E-06 | 18518 | -4.791 | -11.059 | 1.478 | 3.198 | -1.498 | -0.011 | 1.34E-01 | 18709 |
| 98 | Cerebellum_Crus2_R | 6.764 | 3.699 | 9.829 | 1.564 | 4.326 | 0.032 | 1.53E-05 | 18518 | -2.516 | -8.315 | 3.283 | 2.959 | -0.850 | -0.006 | 3.95E-01 | 18709 |
| 99 | Cerebellum_3_L | 0.159 | -0.064 | 0.381 | 0.114 | 1.396 | 0.010 | 1.63E-01 | 18518 | -0.002 | -0.423 | 0.418 | 0.215 | -0.011 | 0.000 | 9.91E-01 | 18709 |
| 100 | Cerebellum_3_R | 0.280 | -0.021 | 0.580 | 0.153 | 1.822 | 0.013 | 6.85E-02 | 18518 | -0.568 | -1.136 | -0.001 | 0.290 | -1.963 | -0.014 | 4.97E-02 | 18709 |
| 101 | Cerebellum_4_5_L | 1.726 | -0.075 | 3.527 | 0.919 | 1.878 | 0.014 | 6.04E-02 | 18518 | -0.827 | -4.229 | 2.575 | 1.736 | -0.477 | -0.003 | 6.34E-01 | 18709 |
| 102 | Cerebellum_4_5_R | 1.917 | 0.449 | 3.385 | 0.749 | 2.560 | 0.019 | 1.05E-02 | 18518 | -2.323 | -5.095 | 0.449 | 1.414 | -1.643 | -0.012 | 1.00E-01 | 18709 |
| 103 | Cerebellum_6_L | 3.519 | 0.406 | 6.633 | 1.589 | 2.215 | 0.016 | 2.67E-02 | 18518 | 0.803 | -5.076 | 6.682 | 2.999 | 0.268 | 0.002 | 7.89E-01 | 18709 |
| 104 | Cerebellum_6_R | 4.427 | 1.149 | 7.705 | 1.672 | 2.647 | 0.019 | 8.12E-03 | 18518 | 0.432 | -5.763 | 6.627 | 3.161 | 0.137 | 0.001 | 8.91E-01 | 18709 |
| 105 | Cerebellum_7b_L | 2.624 | 1.611 | 3.637 | 0.517 | 5.078 | 0.037 | 3.86E-07 | 18518 | -1.325 | -3.241 | 0.591 | 0.977 | -1.355 | -0.010 | 1.75E-01 | 18709 |
| 106 | Cerebellum_7b_R | 2.988 | 1.985 | 3.991 | 0.512 | 5.837 | 0.043 | 5.40E-09 | 18518 | -1.212 | -3.111 | 0.687 | 0.969 | -1.251 | -0.009 | 2.11E-01 | 18709 |
| 107 | Cerebellum_8_L | 4.079 | 0.862 | 7.295 | 1.641 | 2.485 | 0.018 | 1.30E-02 | 18518 | 0.343 | -5.739 | 6.424 | 3.103 | 0.110 | 0.001 | 9.12E-01 | 18709 |
| 108 | Cerebellum_8_R | 9.658 | 5.470 | 13.846 | 2.136 | 4.521 | 0.033 | 6.20E-06 | 18518 | -5.682 | -13.598 | 2.234 | 4.039 | -1.407 | -0.010 | 1.59E-01 | 18709 |
| 109 | Cerebellum_9_L | -1.571 | -3.391 | 0.249 | 0.928 | -1.692 | -0.012 | 9.06E-02 | 18518 | -0.050 | -3.489 | 3.389 | 1.754 | -0.028 | 0.000 | 9.77E-01 | 18709 |
| 110 | Cerebellum_9_R | -1.266 | -3.056 | 0.523 | 0.913 | -1.387 | -0.010 | 1.65E-01 | 18518 | -0.864 | -4.244 | 2.516 | 1.725 | -0.501 | -0.004 | 6.16E-01 | 18709 |
| 111 | Cerebellum_10_L | -0.200 | -0.329 | -0.071 | 0.066 | -3.040 | -0.022 | 2.37E-03 | 18518 | 0.024 | -0.220 | 0.268 | 0.124 | 0.193 | 0.001 | 8.47E-01 | 18709 |
| 112 | Cerebellum_10_R | -0.043 | -0.177 | 0.092 | 0.069 | -0.624 | -0.005 | 5.33E-01 | 18518 | -0.029 | -0.283 | 0.225 | 0.130 | -0.222 | -0.002 | 8.25E-01 | 18709 |
| 113 | Vermis_1_2 | 0.074 | -0.010 | 0.158 | 0.043 | 1.717 | 0.013 | 8.60E-02 | 18518 | -0.145 | -0.304 | 0.013 | 0.081 | -1.797 | -0.013 | 7.24E-02 | 18709 |
| 114 | Vermis_3 | 0.439 | 0.105 | 0.772 | 0.170 | 2.575 | 0.019 | 1.00E-02 | 18518 | -0.792 | -1.422 | -0.161 | 0.322 | -2.462 | -0.018 | 1.38E-02 | 18709 |

| | | | | | | | | | | | | | | | | | |
|---|---|---|---|---|---|---|---|---|---|---|---|---|---|---|---|---|---|
| 115 | Vermis_4_5 | 0.545 | -0.437 | 1.526 | 0.501 | 1.088 | 0.008 | 2.77E-01 | 18518 | -0.614 | -2.467 | 1.239 | 0.946 | -0.650 | -0.005 | 5.16E-01 | 18709 |
| 116 | Vermis_6 | 0.002 | -0.544 | 0.549 | 0.279 | 0.009 | 0.000 | 9.93E-01 | 18518 | 0.416 | -0.616 | 1.447 | 0.526 | 0.790 | 0.006 | 4.29E-01 | 18709 |
| 117 | Vermis_7 | 0.045 | -0.264 | 0.354 | 0.158 | 0.288 | 0.002 | 7.74E-01 | 18518 | 0.231 | -0.352 | 0.814 | 0.297 | 0.777 | 0.006 | 4.37E-01 | 18709 |
| 118 | Vermis_8 | -0.256 | -0.764 | 0.251 | 0.259 | -0.991 | -0.007 | 3.22E-01 | 18518 | 0.390 | -0.568 | 1.349 | 0.489 | 0.798 | 0.006 | 4.25E-01 | 18709 |
| 119 | Vermis_9 | -0.318 | -0.729 | 0.093 | 0.210 | -1.516 | -0.011 | 1.29E-01 | 18518 | 0.370 | -0.407 | 1.147 | 0.396 | 0.934 | 0.007 | 3.50E-01 | 18709 |
| 120 | Vermis_10 | -0.097 | -0.211 | 0.018 | 0.058 | -1.651 | -0.012 | 9.87E-02 | 18518 | 0.085 | -0.132 | 0.302 | 0.111 | 0.770 | 0.006 | 4.41E-01 | 18709 |
| 121 | Thal_AV_L | 0.113 | 0.067 | 0.159 | 0.023 | 4.809 | 0.035 | 1.53E-06 | 18518 | -0.160 | -0.247 | -0.073 | 0.044 | -3.618 | -0.026 | 2.98E-04 | 18709 |
| 122 | Thal_AV_R | 0.187 | 0.121 | 0.253 | 0.034 | 5.567 | 0.041 | 2.63E-08 | 18518 | -0.239 | -0.363 | -0.115 | 0.063 | -3.780 | -0.028 | 1.57E-04 | 18709 |
| 123 | Thal_LP_L | 0.102 | 0.051 | 0.153 | 0.026 | 3.918 | 0.029 | 8.97E-05 | 18518 | -0.229 | -0.325 | -0.133 | 0.049 | -4.662 | -0.034 | 3.16E-06 | 18709 |
| 124 | Thal_LP_R | 0.153 | 0.086 | 0.219 | 0.034 | 4.506 | 0.033 | 6.64E-06 | 18518 | -0.325 | -0.450 | -0.200 | 0.064 | -5.084 | -0.037 | 3.74E-07 | 18709 |
| 125 | Thal_VA_L | 0.065 | 0.010 | 0.119 | 0.028 | 2.331 | 0.017 | 1.98E-02 | 18518 | -0.074 | -0.176 | 0.029 | 0.052 | -1.413 | -0.010 | 1.58E-01 | 18709 |
| 126 | Thal_VA_R | 0.134 | 0.067 | 0.201 | 0.034 | 3.935 | 0.029 | 8.33E-05 | 18518 | -0.100 | -0.226 | 0.026 | 0.064 | -1.560 | -0.011 | 1.19E-01 | 18709 |
| 127 | Thal_VL_L | 0.292 | 0.137 | 0.446 | 0.079 | 3.700 | 0.027 | 2.16E-04 | 18518 | -0.627 | -0.919 | -0.335 | 0.149 | -4.212 | -0.031 | 2.54E-05 | 18709 |
| 128 | Thal_VL_R | 0.621 | 0.416 | 0.826 | 0.105 | 5.934 | 0.044 | 3.02E-09 | 18518 | -0.994 | -1.382 | -0.607 | 0.198 | -5.028 | -0.037 | 5.01E-07 | 18709 |
| 129 | Thal_VPL_L | 0.191 | 0.007 | 0.376 | 0.094 | 2.038 | 0.015 | 4.16E-02 | 18518 | -0.701 | -1.049 | -0.353 | 0.177 | -3.953 | -0.029 | 7.76E-05 | 18709 |
| 130 | Thal_VPL_R | 0.259 | 0.066 | 0.453 | 0.099 | 2.627 | 0.019 | 8.63E-03 | 18518 | -0.979 | -1.344 | -0.613 | 0.186 | -5.250 | -0.038 | 1.54E-07 | 18709 |
| 131 | Thal_IL_L | 0.187 | 0.107 | 0.266 | 0.041 | 4.593 | 0.034 | 4.39E-06 | 18518 | -0.489 | -0.639 | -0.339 | 0.077 | -6.380 | -0.047 | 1.82E-10 | 18709 |
| 132 | Thal_IL_R | 0.261 | 0.171 | 0.352 | 0.046 | 5.679 | 0.042 | 1.38E-08 | 18518 | -0.523 | -0.694 | -0.353 | 0.087 | -6.017 | -0.044 | 1.81E-09 | 18709 |
| 133 | Thal_Re_L | 0.001 | 0.000 | 0.001 | 0.000 | 3.698 | 0.027 | 2.18E-04 | 18518 | 0.000 | -0.001 | 0.000 | 0.000 | -1.408 | -0.010 | 1.59E-01 | 18709 |
| 134 | Thal_Re_R | 0.008 | 0.005 | 0.011 | 0.001 | 5.937 | 0.044 | 2.96E-09 | 18518 | -0.007 | -0.012 | -0.002 | 0.003 | -2.655 | -0.019 | 7.93E-03 | 18709 |
| 135 | Thal_MDm_L | 1.778 | 1.188 | 2.368 | 0.301 | 5.910 | 0.043 | 3.49E-09 | 18518 | -3.527 | -4.641 | -2.414 | 0.568 | -6.211 | -0.045 | 5.39E-10 | 18709 |
| 136 | Thal_MDm_R | 2.185 | 1.500 | 2.870 | 0.350 | 6.250 | 0.046 | 4.18E-10 | 18518 | -3.741 | -5.035 | -2.447 | 0.660 | -5.668 | -0.041 | 1.47E-08 | 18709 |
| 137 | Thal_MDl_L | 0.310 | 0.208 | 0.412 | 0.052 | 5.958 | 0.044 | 2.60E-09 | 18518 | -0.660 | -0.852 | -0.467 | 0.098 | -6.710 | -0.049 | 2.00E-11 | 18709 |
| 138 | Thal_MDl_R | 0.402 | 0.286 | 0.517 | 0.059 | 6.791 | 0.050 | 1.14E-11 | 18518 | -0.672 | -0.891 | -0.453 | 0.112 | -6.012 | -0.044 | 1.87E-09 | 18709 |
| 139 | Thal_LGN_L | 0.017 | 0.002 | 0.033 | 0.008 | 2.161 | 0.016 | 3.07E-02 | 18518 | -0.069 | -0.098 | -0.040 | 0.015 | -4.613 | -0.034 | 3.99E-06 | 18709 |
| 140 | Thal_LGN_R | 0.054 | 0.021 | 0.087 | 0.017 | 3.234 | 0.024 | 1.22E-03 | 18518 | -0.176 | -0.238 | -0.113 | 0.032 | -5.527 | -0.040 | 3.29E-08 | 18709 |
| 141 | Thal_MGN_L | 0.034 | 0.006 | 0.061 | 0.014 | 2.383 | 0.018 | 1.72E-02 | 18518 | -0.173 | -0.225 | -0.121 | 0.027 | -6.488 | -0.047 | 8.91E-11 | 18709 |
| 142 | Thal_MGN_R | 0.048 | 0.018 | 0.078 | 0.015 | 3.118 | 0.023 | 1.82E-03 | 18518 | -0.187 | -0.243 | -0.130 | 0.029 | -6.453 | -0.047 | 1.13E-10 | 18709 |
| 143 | Thal_PuA_L | 0.048 | 0.015 | 0.080 | 0.017 | 2.868 | 0.021 | 4.13E-03 | 18518 | -0.218 | -0.280 | -0.157 | 0.031 | -6.944 | -0.051 | 3.94E-12 | 18709 |
| 144 | Thal_PuA_R | 0.122 | 0.054 | 0.189 | 0.035 | 3.508 | 0.026 | 4.52E-04 | 18518 | -0.460 | -0.589 | -0.332 | 0.065 | -7.043 | -0.051 | 1.95E-12 | 18709 |
| 145 | Thal_PuM_L | 1.326 | 0.677 | 1.976 | 0.331 | 4.002 | 0.029 | 6.30E-05 | 18518 | -4.171 | -5.396 | -2.946 | 0.625 | -6.674 | -0.049 | 2.56E-11 | 18709 |
| 146 | Thal_PuM_R | 1.701 | 0.892 | 2.510 | 0.413 | 4.122 | 0.030 | 3.78E-05 | 18518 | -5.101 | -6.628 | -3.575 | 0.779 | -6.550 | -0.048 | 5.91E-11 | 18709 |
| 147 | Thal_PuL_L | 0.251 | 0.132 | 0.369 | 0.060 | 4.147 | 0.030 | 3.38E-05 | 18518 | -0.717 | -0.941 | -0.494 | 0.114 | -6.291 | -0.046 | 3.22E-10 | 18709 |
| 148 | Thal_PuL_R | 0.395 | 0.207 | 0.584 | 0.096 | 4.115 | 0.030 | 3.89E-05 | 18518 | -1.112 | -1.468 | -0.757 | 0.181 | -6.134 | -0.045 | 8.76E-10 | 18709 |
| 149 | Thal_PuI_L | 0.029 | -0.004 | 0.061 | 0.016 | 1.746 | 0.013 | 8.08E-02 | 18518 | -0.098 | -0.159 | -0.037 | 0.031 | -3.156 | -0.023 | 1.60E-03 | 18709 |
| 150 | Thal_PuI_R | 0.045 | 0.003 | 0.088 | 0.022 | 2.079 | 0.015 | 3.76E-02 | 18518 | -0.192 | -0.272 | -0.111 | 0.041 | -4.665 | -0.034 | 3.11E-06 | 18709 |
| 151 | ACC_sub_L | -0.159 | -0.531 | 0.213 | 0.190 | -0.838 | -0.006 | 4.02E-01 | 18518 | -0.381 | -1.083 | 0.321 | 0.358 | -1.064 | -0.008 | 2.87E-01 | 18709 |
| 152 | ACC_sub_R | -0.314 | -0.662 | 0.033 | 0.177 | -1.774 | -0.013 | 7.60E-02 | 18518 | -0.315 | -0.971 | 0.341 | 0.335 | -0.942 | -0.007 | 3.46E-01 | 18709 |
| 153 | ACC_pre_L | 1.477 | 0.523 | 2.430 | 0.486 | 3.036 | 0.022 | 2.40E-03 | 18518 | -2.884 | -4.682 | -1.085 | 0.917 | -3.143 | -0.023 | 1.67E-03 | 18709 |
| 154 | ACC_pre_R | 1.472 | 0.554 | 2.390 | 0.468 | 3.142 | 0.023 | 1.68E-03 | 18518 | -3.614 | -5.346 | -1.882 | 0.884 | -4.090 | -0.030 | 4.33E-05 | 18709 |
| 155 | ACC_sup_L | 0.900 | -0.136 | 1.936 | 0.529 | 1.703 | 0.013 | 8.86E-02 | 18518 | -3.628 | -5.583 | -1.672 | 0.998 | -3.636 | -0.027 | 2.77E-04 | 18709 |
| 156 | ACC_sup_R | 0.684 | -0.388 | 1.756 | 0.547 | 1.251 | 0.009 | 2.11E-01 | 18518 | -3.110 | -5.135 | -1.084 | 1.033 | -3.010 | -0.022 | 2.62E-03 | 18709 |
| 157 | N_Acc_L | 0.448 | 0.136 | 0.759 | 0.159 | 2.819 | 0.021 | 4.82E-03 | 18518 | -0.409 | -0.997 | 0.180 | 0.300 | -1.362 | -0.010 | 1.73E-01 | 18709 |
| 158 | N_Acc_R | 0.451 | 0.182 | 0.720 | 0.137 | 3.284 | 0.024 | 1.02E-03 | 18518 | -0.477 | -0.986 | 0.032 | 0.260 | -1.838 | -0.013 | 6.61E-02 | 18709 |
| 159 | VTA_L | 0.017 | 0.006 | 0.027 | 0.006 | 2.952 | 0.022 | 3.16E-03 | 18518 | 0.002 | -0.018 | 0.023 | 0.011 | 0.212 | 0.002 | 8.32E-01 | 18709 |
| 160 | VTA_R | 0.021 | 0.011 | 0.031 | 0.005 | 4.135 | 0.030 | 3.57E-05 | 18518 | -0.014 | -0.032 | 0.005 | 0.010 | -1.426 | -0.010 | 1.54E-01 | 18709 |
| 161 | SN_pc_L | 0.007 | -0.011 | 0.024 | 0.009 | 0.767 | 0.006 | 4.43E-01 | 18518 | -0.040 | -0.073 | -0.006 | 0.017 | -2.340 | -0.017 | 1.93E-02 | 18709 |
| 162 | SN_pc_R | 0.020 | -0.004 | 0.043 | 0.012 | 1.657 | 0.012 | 9.76E-02 | 18518 | -0.072 | -0.115 | -0.028 | 0.022 | -3.217 | -0.024 | 1.30E-03 | 18709 |

| | | | | | | | | | | | | | | | | | |
|---|---|---|---|---|---|---|---|---|---|---|---|---|---|---|---|---|---|
| 163 | SN_pr_L | 0.013 | -0.011 | 0.037 | 0.012 | 1.086 | 0.008 | 2.78E-01 | 18518 | -0.042 | -0.087 | 0.004 | 0.023 | -1.805 | -0.013 | 7.10E-02 | 18709 |
| 164 | SN_pr_R | 0.022 | -0.005 | 0.049 | 0.014 | 1.604 | 0.012 | 1.09E-01 | 18518 | -0.036 | -0.087 | 0.014 | 0.026 | -1.406 | -0.010 | 1.60E-01 | 18709 |
| 165 | Red_N_L | 0.037 | 0.008 | 0.067 | 0.015 | 2.472 | 0.018 | 1.34E-02 | 18518 | -0.082 | -0.138 | -0.027 | 0.028 | -2.895 | -0.021 | 3.80E-03 | 18709 |
| 166 | Red_N_R | 0.065 | 0.032 | 0.097 | 0.016 | 3.940 | 0.029 | 8.17E-05 | 18518 | -0.101 | -0.162 | -0.040 | 0.031 | -3.269 | -0.024 | 1.08E-03 | 18709 |
| 167 | LC_L | 0.005 | -0.006 | 0.016 | 0.006 | 0.852 | 0.006 | 3.94E-01 | 18518 | -0.010 | -0.031 | 0.011 | 0.011 | -0.966 | -0.007 | 3.34E-01 | 18709 |
| 168 | LC_R | 0.003 | -0.009 | 0.015 | 0.006 | 0.503 | 0.004 | 6.15E-01 | 18518 | -0.014 | -0.037 | 0.009 | 0.012 | -1.218 | -0.009 | 2.23E-01 | 18709 |
| 169 | Raphe_D | 0.116 | 0.074 | 0.159 | 0.022 | 5.350 | 0.039 | 8.88E-08 | 18518 | -0.186 | -0.267 | -0.105 | 0.041 | -4.518 | -0.033 | 6.29E-06 | 18709 |
| 170 | Raphe_M | 0.019 | 0.006 | 0.032 | 0.007 | 2.840 | 0.021 | 4.51E-03 | 18518 | -0.014 | -0.038 | 0.011 | 0.013 | -1.086 | -0.008 | 2.77E-01 | 18709 |

| | | Bread intake | | | | | | | | Pork intake | | | | | | | |
|---|---|---|---|---|---|---|---|---|---|---|---|---|---|---|---|---|---|
| **Number** | **AAL 3 region** | **beta** | **95%CI_l** | **95%CI_h** | **se** | **t** | **r** | **p** | **df.e** | **beta** | **95%CI_l** | **95%CI_h** | **se** | **t** | **r** | **p** | **df.e** |
| 0 | Total brain | -4.371 | -23.834 | 15.091 | 9.929 | -0.440 | -0.003 | 6.60E-01 | 18607 | -591.260 | -785.720 | -396.801 | 99.209 | -5.960 | -0.044 | 2.57E-09 | 18697 |
| 1 | Precentral_L | -0.300 | -0.986 | 0.385 | 0.350 | -0.859 | -0.006 | 3.90E-01 | 18607 | -9.043 | -15.896 | -2.189 | 3.497 | -2.586 | -0.019 | 9.71E-03 | 18697 |
| 2 | Precentral_R | 0.351 | -0.277 | 0.979 | 0.320 | 1.094 | 0.008 | 2.74E-01 | 18607 | -12.589 | -18.867 | -6.312 | 3.203 | -3.931 | -0.029 | 8.49E-05 | 18697 |
| 3 | Frontal_Sup_2_L | 0.764 | 0.040 | 1.487 | 0.369 | 2.070 | 0.015 | 3.85E-02 | 18607 | -16.730 | -23.963 | -9.496 | 3.690 | -4.533 | -0.033 | 5.84E-06 | 18697 |
| 4 | Frontal_Sup_2_R | 0.857 | 0.059 | 1.654 | 0.407 | 2.105 | 0.015 | 3.53E-02 | 18607 | -17.157 | -25.129 | -9.184 | 4.067 | -4.218 | -0.031 | 2.48E-05 | 18697 |
| 5 | Frontal_Mid_2_L | 0.331 | -0.428 | 1.091 | 0.387 | 0.856 | 0.006 | 3.92E-01 | 18607 | -8.993 | -16.589 | -1.398 | 3.875 | -2.321 | -0.017 | 2.03E-02 | 18697 |
| 6 | Frontal_Mid_2_R | 0.570 | -0.289 | 1.429 | 0.438 | 1.300 | 0.010 | 1.94E-01 | 18607 | -19.092 | -27.685 | -10.498 | 4.384 | -4.355 | -0.032 | 1.34E-05 | 18697 |
| 7 | Frontal_Inf_Oper_L | -0.113 | -0.337 | 0.111 | 0.114 | -0.989 | -0.007 | 3.23E-01 | 18607 | -3.606 | -5.851 | -1.361 | 1.145 | -3.149 | -0.023 | 1.64E-03 | 18697 |
| 8 | Frontal_Inf_Oper_R | 0.173 | -0.108 | 0.455 | 0.144 | 1.206 | 0.009 | 2.28E-01 | 18607 | -5.893 | -8.708 | -3.078 | 1.436 | -4.104 | -0.030 | 4.08E-05 | 18697 |
| 9 | Frontal_Inf_Tri_L | 0.109 | -0.368 | 0.586 | 0.243 | 0.449 | 0.003 | 6.53E-01 | 18607 | -11.163 | -15.929 | -6.396 | 2.432 | -4.591 | -0.034 | 4.45E-06 | 18697 |
| 10 | Frontal_Inf_Tri_R | 0.030 | -0.344 | 0.404 | 0.191 | 0.157 | 0.001 | 8.75E-01 | 18607 | -10.087 | -13.830 | -6.345 | 1.909 | -5.283 | -0.039 | 1.28E-07 | 18697 |
| 11 | Frontal_Inf_Orb_2_L | 0.146 | 0.004 | 0.289 | 0.073 | 2.015 | 0.015 | 4.39E-02 | 18607 | -2.389 | -3.814 | -0.963 | 0.727 | -3.285 | -0.024 | 1.02E-03 | 18697 |
| 12 | Frontal_Inf_Orb_2_R | 0.117 | -0.032 | 0.266 | 0.076 | 1.541 | 0.011 | 1.23E-01 | 18607 | -2.638 | -4.128 | -1.148 | 0.760 | -3.470 | -0.025 | 5.22E-04 | 18697 |
| 13 | Rolandic_Oper_L | 0.040 | -0.175 | 0.256 | 0.110 | 0.368 | 0.003 | 7.13E-01 | 18607 | -4.448 | -6.603 | -2.292 | 1.100 | -4.043 | -0.030 | 5.29E-05 | 18697 |
| 14 | Rolandic_Oper_R | 0.064 | -0.245 | 0.372 | 0.157 | 0.404 | 0.003 | 6.86E-01 | 18607 | -8.810 | -11.894 | -5.725 | 1.574 | -5.598 | -0.041 | 2.19E-08 | 18697 |
| 15 | Supp_Motor_Area_L | -0.083 | -0.596 | 0.430 | 0.262 | -0.318 | -0.002 | 7.51E-01 | 18607 | -6.434 | -11.559 | -1.309 | 2.615 | -2.461 | -0.018 | 1.39E-02 | 18697 |
| 16 | Supp_Motor_Area_R | -0.161 | -0.727 | 0.405 | 0.289 | -0.557 | -0.004 | 5.78E-01 | 18607 | -2.097 | -7.761 | 3.566 | 2.890 | -0.726 | -0.005 | 4.68E-01 | 18697 |
| 17 | Olfactory_L | -0.045 | -0.121 | 0.032 | 0.039 | -1.148 | -0.008 | 2.51E-01 | 18607 | -1.229 | -1.991 | -0.468 | 0.389 | -3.163 | -0.023 | 1.56E-03 | 18697 |
| 18 | Olfactory_R | -0.024 | -0.100 | 0.053 | 0.039 | -0.612 | -0.004 | 5.41E-01 | 18607 | -1.304 | -2.067 | -0.540 | 0.390 | -3.347 | -0.024 | 8.19E-04 | 18697 |
| 19 | Frontal_Sup_Medial_L | 0.279 | -0.202 | 0.759 | 0.245 | 1.137 | 0.008 | 2.55E-01 | 18607 | -10.905 | -15.709 | -6.102 | 2.451 | -4.450 | -0.033 | 8.64E-06 | 18697 |
| 20 | Frontal_Sup_Medial_R | 0.294 | -0.056 | 0.643 | 0.178 | 1.647 | 0.012 | 9.97E-02 | 18607 | -7.069 | -10.565 | -3.573 | 1.784 | -3.963 | -0.029 | 7.43E-05 | 18697 |
| 21 | Frontal_Med_Orb_L | 0.170 | -0.010 | 0.349 | 0.092 | 1.853 | 0.014 | 6.40E-02 | 18607 | -3.928 | -5.721 | -2.134 | 0.915 | -4.293 | -0.031 | 1.78E-05 | 18697 |
| 22 | Frontal_Med_Orb_R | 0.219 | -0.001 | 0.440 | 0.113 | 1.949 | 0.014 | 5.14E-02 | 18607 | -4.649 | -6.853 | -2.444 | 1.125 | -4.133 | -0.030 | 3.59E-05 | 18697 |
| 23 | Rectus_L | 0.135 | -0.051 | 0.320 | 0.095 | 1.420 | 0.010 | 1.56E-01 | 18607 | -3.343 | -5.200 | -1.487 | 0.947 | -3.530 | -0.026 | 4.17E-04 | 18697 |
| 24 | Rectus_R | 0.044 | -0.128 | 0.216 | 0.088 | 0.505 | 0.004 | 6.14E-01 | 18607 | -2.930 | -4.648 | -1.213 | 0.876 | -3.344 | -0.024 | 8.28E-04 | 18697 |
| 25 | OFCmed_L | 0.018 | -0.090 | 0.125 | 0.055 | 0.324 | 0.002 | 7.46E-01 | 18607 | -1.764 | -2.841 | -0.688 | 0.549 | -3.213 | -0.023 | 1.32E-03 | 18697 |
| 26 | OFCmed_R | 0.010 | -0.116 | 0.135 | 0.064 | 0.155 | 0.001 | 8.77E-01 | 18607 | -2.038 | -3.292 | -0.783 | 0.640 | -3.184 | -0.023 | 1.45E-03 | 18697 |
| 27 | OFCant_L | 0.014 | -0.111 | 0.138 | 0.063 | 0.214 | 0.002 | 8.30E-01 | 18607 | -1.833 | -3.078 | -0.588 | 0.635 | -2.886 | -0.021 | 3.90E-03 | 18697 |
| 28 | OFCant_R | 0.061 | -0.103 | 0.226 | 0.084 | 0.731 | 0.005 | 4.65E-01 | 18607 | -3.573 | -5.217 | -1.928 | 0.839 | -4.259 | -0.031 | 2.06E-05 | 18697 |
| 29 | OFCpost_L | 0.028 | -0.128 | 0.184 | 0.080 | 0.353 | 0.003 | 7.24E-01 | 18607 | -3.156 | -4.714 | -1.597 | 0.795 | -3.970 | -0.029 | 7.22E-05 | 18697 |
| 30 | OFCpost_R | 0.101 | -0.043 | 0.245 | 0.074 | 1.373 | 0.010 | 1.70E-01 | 18607 | -2.962 | -4.402 | -1.521 | 0.735 | -4.030 | -0.029 | 5.60E-05 | 18697 |
| 31 | OFClat_L | 0.019 | -0.034 | 0.071 | 0.027 | 0.703 | 0.005 | 4.82E-01 | 18607 | -0.908 | -1.434 | -0.382 | 0.268 | -3.384 | -0.025 | 7.17E-04 | 18697 |
| 32 | OFClat_R | 0.014 | -0.031 | 0.058 | 0.023 | 0.596 | 0.004 | 5.51E-01 | 18607 | -0.827 | -1.271 | -0.383 | 0.227 | -3.650 | -0.027 | 2.63E-04 | 18697 |
| 33 | Insula_L | 0.020 | -0.462 | 0.502 | 0.246 | 0.081 | 0.001 | 9.35E-01 | 18607 | -13.326 | -18.143 | -8.508 | 2.458 | -5.422 | -0.040 | 5.96E-08 | 18697 |
| 34 | Insula_R | 0.092 | -0.377 | 0.562 | 0.239 | 0.385 | 0.003 | 7.00E-01 | 18607 | -12.344 | -17.037 | -7.651 | 2.394 | -5.155 | -0.038 | 2.56E-07 | 18697 |
| 37 | Cingulate_Mid_L | 0.298 | -0.110 | 0.705 | 0.208 | 1.433 | 0.011 | 1.52E-01 | 18607 | -12.976 | -17.046 | -8.906 | 2.076 | -6.249 | -0.046 | 4.21E-10 | 18697 |
| 38 | Cingulate_Mid_R | 0.165 | -0.269 | 0.599 | 0.221 | 0.746 | 0.005 | 4.56E-01 | 18607 | -13.839 | -18.179 | -9.499 | 2.214 | -6.250 | -0.046 | 4.19E-10 | 18697 |
| 39 | Cingulate_Post_L | 0.005 | -0.119 | 0.130 | 0.063 | 0.082 | 0.001 | 9.35E-01 | 18607 | -2.562 | -3.806 | -1.317 | 0.635 | -4.036 | -0.030 | 5.47E-05 | 18697 |

| | | | | | | | | | | | | | | | | | |
|---|---|---|---|---|---|---|---|---|---|---|---|---|---|---|---|---|---|
| 40 | Cingulate_Post_R | -0.019 | -0.088 | 0.050 | 0.035 | -0.538 | -0.004 | 5.91E-01 | 18607 | -1.143 | -1.834 | -0.453 | 0.352 | -3.247 | -0.024 | 1.17E-03 | 18697 |
| 41 | Hippocampus_L | -0.004 | -0.205 | 0.197 | 0.103 | -0.040 | 0.000 | 9.68E-01 | 18607 | -1.845 | -3.857 | 0.167 | 1.027 | -1.797 | -0.013 | 7.23E-02 | 18697 |
| 42 | Hippocampus_R | -0.031 | -0.240 | 0.177 | 0.106 | -0.295 | -0.002 | 7.68E-01 | 18607 | -0.829 | -2.914 | 1.257 | 1.064 | -0.779 | -0.006 | 4.36E-01 | 18697 |
| 43 | ParaHippocampal_L | 0.059 | -0.119 | 0.238 | 0.091 | 0.650 | 0.005 | 5.16E-01 | 18607 | -2.164 | -3.951 | -0.376 | 0.912 | -2.373 | -0.017 | 1.77E-02 | 18697 |
| 44 | ParaHippocampal_R | 0.110 | -0.116 | 0.335 | 0.115 | 0.954 | 0.007 | 3.40E-01 | 18607 | -1.910 | -4.165 | 0.346 | 1.151 | -1.660 | -0.012 | 9.70E-02 | 18697 |
| 45 | Amygdala_L | 0.013 | -0.057 | 0.082 | 0.035 | 0.364 | 0.003 | 7.16E-01 | 18607 | -0.544 | -1.238 | 0.151 | 0.354 | -1.534 | -0.011 | 1.25E-01 | 18697 |
| 46 | Amygdala_R | 0.020 | -0.051 | 0.091 | 0.036 | 0.562 | 0.004 | 5.74E-01 | 18607 | -0.741 | -1.452 | -0.030 | 0.363 | -2.043 | -0.015 | 4.10E-02 | 18697 |
| 47 | Calcarine_L | -1.484 | -2.147 | -0.820 | 0.338 | -4.384 | -0.032 | 1.17E-05 | 18607 | -2.444 | -9.080 | 4.192 | 3.386 | -0.722 | -0.005 | 4.70E-01 | 18697 |
| 48 | Calcarine_R | -1.232 | -1.786 | -0.677 | 0.283 | -4.354 | -0.032 | 1.34E-05 | 18607 | -2.213 | -7.760 | 3.334 | 2.830 | -0.782 | -0.006 | 4.34E-01 | 18697 |
| 49 | Cuneus_L | -0.689 | -1.105 | -0.273 | 0.212 | -3.248 | -0.024 | 1.17E-03 | 18607 | -4.724 | -8.884 | -0.563 | 2.123 | -2.225 | -0.016 | 2.61E-02 | 18697 |
| 50 | Cuneus_R | -0.501 | -0.819 | -0.183 | 0.162 | -3.084 | -0.023 | 2.05E-03 | 18607 | -3.936 | -7.119 | -0.752 | 1.624 | -2.423 | -0.018 | 1.54E-02 | 18697 |
| 51 | Lingual_L | -0.571 | -0.970 | -0.173 | 0.203 | -2.808 | -0.021 | 4.99E-03 | 18607 | -5.330 | -9.320 | -1.341 | 2.035 | -2.619 | -0.019 | 8.83E-03 | 18697 |
| 52 | Lingual_R | -0.567 | -0.985 | -0.150 | 0.213 | -2.662 | -0.020 | 7.77E-03 | 18607 | -4.823 | -9.004 | -0.641 | 2.133 | -2.261 | -0.017 | 2.38E-02 | 18697 |
| 53 | Occipital_Sup_L | -0.119 | -0.394 | 0.155 | 0.140 | -0.854 | -0.006 | 3.93E-01 | 18607 | -5.139 | -7.881 | -2.397 | 1.399 | -3.673 | -0.027 | 2.40E-04 | 18697 |
| 54 | Occipital_Sup_R | -0.251 | -0.548 | 0.047 | 0.152 | -1.653 | -0.012 | 9.84E-02 | 18607 | -5.224 | -8.196 | -2.251 | 1.517 | -3.445 | -0.025 | 5.73E-04 | 18697 |
| 55 | Occipital_Mid_L | -0.744 | -1.386 | -0.101 | 0.328 | -2.268 | -0.017 | 2.33E-02 | 18607 | -9.736 | -16.163 | -3.309 | 3.279 | -2.969 | -0.022 | 2.99E-03 | 18697 |
| 56 | Occipital_Mid_R | -0.363 | -0.811 | 0.084 | 0.228 | -1.592 | -0.012 | 1.11E-01 | 18607 | -6.586 | -11.059 | -2.112 | 2.282 | -2.886 | -0.021 | 3.91E-03 | 18697 |
| 57 | Occipital_Inf_L | -0.221 | -0.435 | -0.008 | 0.109 | -2.033 | -0.015 | 4.21E-02 | 18607 | -0.909 | -3.044 | 1.225 | 1.089 | -0.835 | -0.006 | 4.04E-01 | 18697 |
| 58 | Occipital_Inf_R | -0.255 | -0.501 | -0.008 | 0.126 | -2.025 | -0.015 | 4.29E-02 | 18607 | 0.768 | -1.697 | 3.233 | 1.258 | 0.611 | 0.004 | 5.41E-01 | 18697 |
| 59 | Fusiform_L | 0.170 | -0.297 | 0.636 | 0.238 | 0.713 | 0.005 | 4.76E-01 | 18607 | -3.691 | -8.360 | 0.978 | 2.382 | -1.550 | -0.011 | 1.21E-01 | 18697 |
| 60 | Fusiform_R | 0.239 | -0.274 | 0.753 | 0.262 | 0.914 | 0.007 | 3.61E-01 | 18607 | -4.982 | -10.110 | 0.146 | 2.616 | -1.904 | -0.014 | 5.69E-02 | 18697 |
| 61 | Postcentral_L | 0.440 | -0.263 | 1.143 | 0.358 | 1.227 | 0.009 | 2.20E-01 | 18607 | -15.447 | -22.466 | -8.427 | 3.581 | -4.313 | -0.032 | 1.62E-05 | 18697 |
| 62 | Postcentral_R | 0.814 | 0.094 | 1.535 | 0.368 | 2.215 | 0.016 | 2.68E-02 | 18607 | -14.531 | -21.731 | -7.331 | 3.673 | -3.956 | -0.029 | 7.66E-05 | 18697 |
| 63 | Parietal_Sup_L | -0.444 | -0.848 | -0.039 | 0.206 | -2.149 | -0.016 | 3.17E-02 | 18607 | -5.746 | -9.793 | -1.698 | 2.065 | -2.783 | -0.020 | 5.40E-03 | 18697 |
| 64 | Parietal_Sup_R | -0.427 | -0.786 | -0.067 | 0.183 | -2.328 | -0.017 | 1.99E-02 | 18607 | -5.443 | -9.032 | -1.854 | 1.831 | -2.972 | -0.022 | 2.96E-03 | 18697 |
| 65 | Parietal_Inf_L | -0.280 | -0.810 | 0.250 | 0.270 | -1.037 | -0.008 | 3.00E-01 | 18607 | -14.018 | -19.314 | -8.722 | 2.702 | -5.188 | -0.038 | 2.14E-07 | 18697 |
| 66 | Parietal_Inf_R | 0.012 | -0.331 | 0.355 | 0.175 | 0.068 | 0.001 | 9.45E-01 | 18607 | -7.966 | -11.399 | -4.534 | 1.751 | -4.550 | -0.033 | 5.41E-06 | 18697 |
| 67 | SupraMarginal_L | 0.171 | -0.139 | 0.482 | 0.158 | 1.081 | 0.008 | 2.79E-01 | 18607 | -7.364 | -10.467 | -4.260 | 1.583 | -4.650 | -0.034 | 3.34E-06 | 18697 |
| 68 | SupraMarginal_R | 0.249 | -0.190 | 0.688 | 0.224 | 1.112 | 0.008 | 2.66E-01 | 18607 | -10.059 | -14.452 | -5.665 | 2.241 | -4.488 | -0.033 | 7.24E-06 | 18697 |
| 69 | Angular_L | -0.257 | -0.562 | 0.048 | 0.156 | -1.654 | -0.012 | 9.82E-02 | 18607 | -5.645 | -8.694 | -2.596 | 1.556 | -3.629 | -0.027 | 2.86E-04 | 18697 |
| 70 | Angular_R | -0.229 | -0.638 | 0.180 | 0.209 | -1.098 | -0.008 | 2.72E-01 | 18607 | -8.723 | -12.813 | -4.634 | 2.086 | -4.181 | -0.031 | 2.92E-05 | 18697 |
| 71 | Precuneus_L | -0.480 | -1.224 | 0.264 | 0.380 | -1.264 | -0.009 | 2.06E-01 | 18607 | -13.102 | -20.537 | -5.668 | 3.793 | -3.455 | -0.025 | 5.52E-04 | 18697 |
| 72 | Precuneus_R | -0.630 | -1.292 | 0.033 | 0.338 | -1.863 | -0.014 | 6.24E-02 | 18607 | -8.930 | -15.553 | -2.308 | 3.379 | -2.643 | -0.019 | 8.22E-03 | 18697 |
| 73 | Paracentral_Lobule_L | -0.010 | -0.351 | 0.330 | 0.174 | -0.059 | 0.000 | 9.53E-01 | 18607 | -2.331 | -5.736 | 1.075 | 1.737 | -1.341 | -0.010 | 1.80E-01 | 18697 |
| 74 | Paracentral_Lobule_R | 0.011 | -0.232 | 0.255 | 0.124 | 0.092 | 0.001 | 9.27E-01 | 18607 | -1.870 | -4.310 | 0.569 | 1.245 | -1.503 | -0.011 | 1.33E-01 | 18697 |
| 75 | Caudate_L | 0.076 | -0.196 | 0.348 | 0.139 | 0.549 | 0.004 | 5.83E-01 | 18607 | -3.992 | -6.710 | -1.274 | 1.387 | -2.879 | -0.021 | 4.00E-03 | 18697 |
| 76 | Caudate_R | 0.217 | -0.091 | 0.525 | 0.157 | 1.383 | 0.010 | 1.67E-01 | 18607 | -3.921 | -6.999 | -0.843 | 1.570 | -2.497 | -0.018 | 1.25E-02 | 18697 |
| 77 | Putamen_L | 0.157 | -0.256 | 0.569 | 0.210 | 0.746 | 0.005 | 4.56E-01 | 18607 | -6.595 | -10.723 | -2.466 | 2.106 | -3.131 | -0.023 | 1.75E-03 | 18697 |
| 78 | Putamen_R | 0.283 | -0.138 | 0.703 | 0.214 | 1.319 | 0.010 | 1.87E-01 | 18607 | -7.216 | -11.420 | -3.011 | 2.145 | -3.364 | -0.025 | 7.70E-04 | 18697 |
| 79 | Pallidum_L | -0.003 | -0.039 | 0.034 | 0.019 | -0.136 | -0.001 | 8.92E-01 | 18607 | -0.416 | -0.783 | -0.049 | 0.187 | -2.220 | -0.016 | 2.64E-02 | 18697 |
| 80 | Pallidum_R | 0.013 | -0.028 | 0.053 | 0.021 | 0.618 | 0.005 | 5.37E-01 | 18607 | -0.638 | -1.041 | -0.235 | 0.206 | -3.104 | -0.023 | 1.91E-03 | 18697 |
| 83 | Heschl_L | -0.001 | -0.075 | 0.074 | 0.038 | -0.022 | 0.000 | 9.82E-01 | 18607 | -2.050 | -2.795 | -1.306 | 0.380 | -5.400 | -0.039 | 6.75E-08 | 18697 |
| 84 | Heschl_R | 0.046 | -0.038 | 0.129 | 0.042 | 1.076 | 0.008 | 2.82E-01 | 18607 | -2.298 | -3.130 | -1.467 | 0.424 | -5.417 | -0.040 | 6.12E-08 | 18697 |
| 85 | Temporal_Sup_L | 0.374 | -0.164 | 0.911 | 0.274 | 1.363 | 0.010 | 1.73E-01 | 18607 | -15.028 | -20.398 | -9.657 | 2.740 | -5.484 | -0.040 | 4.20E-08 | 18697 |
| 86 | Temporal_Sup_R | 0.149 | -0.446 | 0.743 | 0.303 | 0.491 | 0.004 | 6.24E-01 | 18607 | -18.266 | -24.205 | -12.326 | 3.030 | -6.028 | -0.044 | 1.70E-09 | 18697 |
| 87 | Temporal_Pole_Sup_L | 0.142 | -0.112 | 0.396 | 0.130 | 1.094 | 0.008 | 2.74E-01 | 18607 | -3.776 | -6.320 | -1.231 | 1.298 | -2.908 | -0.021 | 3.64E-03 | 18697 |
| 88 | Temporal_Pole_Sup_R | 0.242 | -0.025 | 0.510 | 0.137 | 1.774 | 0.013 | 7.61E-02 | 18607 | -3.673 | -6.352 | -0.994 | 1.367 | -2.687 | -0.020 | 7.22E-03 | 18697 |
| 89 | Temporal_Mid_L | -0.596 | -1.492 | 0.299 | 0.457 | -1.305 | -0.010 | 1.92E-01 | 18607 | -22.603 | -31.555 | -13.651 | 4.567 | -4.949 | -0.036 | 7.53E-07 | 18697 |

| | | | | | | | | | | | | | | | | | |
|---|---|---|---|---|---|---|---|---|---|---|---|---|---|---|---|---|---|
| 90 | Temporal_Mid_R | -0.375 | -1.170 | 0.421 | 0.406 | -0.923 | -0.007 | 3.56E-01 | 18607 | -17.089 | -25.035 | -9.143 | 4.054 | -4.216 | -0.031 | 2.50E-05 | 18697 |
| 91 | Temporal_Pole_Mid_L | -0.012 | -0.225 | 0.202 | 0.109 | -0.107 | -0.001 | 9.15E-01 | 18607 | -1.540 | -3.680 | 0.599 | 1.091 | -1.411 | -0.010 | 1.58E-01 | 18697 |
| 92 | Temporal_Pole_Mid_R | 0.139 | -0.121 | 0.399 | 0.133 | 1.048 | 0.008 | 2.95E-01 | 18607 | -2.407 | -5.007 | 0.192 | 1.326 | -1.815 | -0.013 | 6.95E-02 | 18697 |
| 93 | Temporal_Inf_L | -0.261 | -0.877 | 0.356 | 0.314 | -0.829 | -0.006 | 4.07E-01 | 18607 | -5.672 | -11.823 | 0.480 | 3.138 | -1.807 | -0.013 | 7.07E-02 | 18697 |
| 94 | Temporal_Inf_R | -0.098 | -0.770 | 0.573 | 0.343 | -0.287 | -0.002 | 7.74E-01 | 18607 | -5.007 | -11.719 | 1.705 | 3.425 | -1.462 | -0.011 | 1.44E-01 | 18697 |
| 95 | Cerebellum_Crus1_L | -0.645 | -1.376 | 0.085 | 0.373 | -1.731 | -0.013 | 8.35E-02 | 18607 | 3.672 | -3.643 | 10.987 | 3.732 | 0.984 | 0.007 | 3.25E-01 | 18697 |
| 96 | Cerebellum_Crus1_R | -0.255 | -1.000 | 0.490 | 0.380 | -0.671 | -0.005 | 5.02E-01 | 18607 | -2.225 | -9.686 | 5.236 | 3.806 | -0.585 | -0.004 | 5.59E-01 | 18697 |
| 97 | Cerebellum_Crus2_L | -0.374 | -0.954 | 0.206 | 0.296 | -1.264 | -0.009 | 2.06E-01 | 18607 | -3.713 | -9.515 | 2.090 | 2.960 | -1.254 | -0.009 | 2.10E-01 | 18697 |
| 98 | Cerebellum_Crus2_R | -0.284 | -0.821 | 0.252 | 0.274 | -1.039 | -0.008 | 2.99E-01 | 18607 | -2.294 | -7.662 | 3.073 | 2.738 | -0.838 | -0.006 | 4.02E-01 | 18697 |
| 99 | Cerebellum_3_L | -0.039 | -0.078 | 0.000 | 0.020 | -1.980 | -0.015 | 4.77E-02 | 18607 | 0.031 | -0.359 | 0.420 | 0.199 | 0.155 | 0.001 | 8.77E-01 | 18697 |
| 100 | Cerebellum_3_R | -0.063 | -0.116 | -0.010 | 0.027 | -2.350 | -0.017 | 1.88E-02 | 18607 | -0.442 | -0.968 | 0.084 | 0.268 | -1.648 | -0.012 | 9.93E-02 | 18697 |
| 101 | Cerebellum_4_5_L | -0.094 | -0.409 | 0.221 | 0.161 | -0.584 | -0.004 | 5.59E-01 | 18607 | 0.340 | -2.810 | 3.491 | 1.607 | 0.212 | 0.002 | 8.32E-01 | 18697 |
| 102 | Cerebellum_4_5_R | -0.098 | -0.355 | 0.158 | 0.131 | -0.752 | -0.006 | 4.52E-01 | 18607 | -1.561 | -4.127 | 1.005 | 1.309 | -1.192 | -0.009 | 2.33E-01 | 18697 |
| 103 | Cerebellum_6_L | -0.229 | -0.773 | 0.315 | 0.278 | -0.824 | -0.006 | 4.10E-01 | 18607 | 0.390 | -5.054 | 5.835 | 2.778 | 0.140 | 0.001 | 8.88E-01 | 18697 |
| 104 | Cerebellum_6_R | -0.392 | -0.965 | 0.182 | 0.293 | -1.339 | -0.010 | 1.81E-01 | 18607 | -0.206 | -5.942 | 5.529 | 2.926 | -0.071 | -0.001 | 9.44E-01 | 18697 |
| 105 | Cerebellum_7b_L | 0.021 | -0.156 | 0.199 | 0.090 | 0.236 | 0.002 | 8.13E-01 | 18607 | -0.617 | -2.390 | 1.157 | 0.905 | -0.682 | -0.005 | 4.95E-01 | 18697 |
| 106 | Cerebellum_7b_R | -0.046 | -0.222 | 0.129 | 0.090 | -0.517 | -0.004 | 6.06E-01 | 18607 | -1.132 | -2.890 | 0.626 | 0.897 | -1.262 | -0.009 | 2.07E-01 | 18697 |
| 107 | Cerebellum_8_L | 0.021 | -0.541 | 0.584 | 0.287 | 0.074 | 0.001 | 9.41E-01 | 18607 | 5.376 | -0.253 | 11.005 | 2.872 | 1.872 | 0.014 | 6.12E-02 | 18697 |
| 108 | Cerebellum_8_R | 0.116 | -0.617 | 0.848 | 0.374 | 0.310 | 0.002 | 7.57E-01 | 18607 | 0.404 | -6.923 | 7.731 | 3.738 | 0.108 | 0.001 | 9.14E-01 | 18697 |
| 109 | Cerebellum_9_L | -0.152 | -0.470 | 0.166 | 0.162 | -0.938 | -0.007 | 3.48E-01 | 18607 | 4.069 | 0.885 | 7.252 | 1.624 | 2.505 | 0.018 | 1.23E-02 | 18697 |
| 110 | Cerebellum_9_R | -0.206 | -0.519 | 0.106 | 0.160 | -1.293 | -0.009 | 1.96E-01 | 18607 | 4.478 | 1.349 | 7.607 | 1.596 | 2.805 | 0.021 | 5.03E-03 | 18697 |
| 111 | Cerebellum_10_L | 0.002 | -0.020 | 0.025 | 0.012 | 0.190 | 0.001 | 8.50E-01 | 18607 | 0.238 | 0.012 | 0.464 | 0.115 | 2.066 | 0.015 | 3.89E-02 | 18697 |
| 112 | Cerebellum_10_R | 0.006 | -0.018 | 0.029 | 0.012 | 0.490 | 0.004 | 6.24E-01 | 18607 | 0.059 | -0.176 | 0.294 | 0.120 | 0.495 | 0.004 | 6.20E-01 | 18697 |
| 113 | Vermis_1_2 | -0.016 | -0.031 | -0.001 | 0.007 | -2.130 | -0.016 | 3.32E-02 | 18607 | -0.096 | -0.243 | 0.051 | 0.075 | -1.279 | -0.009 | 2.01E-01 | 18697 |
| 114 | Vermis_3 | -0.045 | -0.103 | 0.014 | 0.030 | -1.497 | -0.011 | 1.34E-01 | 18607 | -0.532 | -1.116 | 0.052 | 0.298 | -1.786 | -0.013 | 7.41E-02 | 18697 |
| 115 | Vermis_4_5 | -0.018 | -0.190 | 0.153 | 0.088 | -0.209 | -0.002 | 8.34E-01 | 18607 | -0.148 | -1.863 | 1.568 | 0.875 | -0.169 | -0.001 | 8.66E-01 | 18697 |
| 116 | Vermis_6 | -0.046 | -0.142 | 0.049 | 0.049 | -0.946 | -0.007 | 3.44E-01 | 18607 | 0.707 | -0.248 | 1.662 | 0.487 | 1.450 | 0.011 | 1.47E-01 | 18697 |
| 117 | Vermis_7 | 0.029 | -0.025 | 0.083 | 0.028 | 1.042 | 0.008 | 2.97E-01 | 18607 | 0.475 | -0.065 | 1.014 | 0.275 | 1.723 | 0.013 | 8.48E-02 | 18697 |
| 118 | Vermis_8 | 0.002 | -0.087 | 0.090 | 0.045 | 0.038 | 0.000 | 9.69E-01 | 18607 | 1.336 | 0.449 | 2.223 | 0.453 | 2.952 | 0.022 | 3.16E-03 | 18697 |
| 119 | Vermis_9 | -0.053 | -0.125 | 0.019 | 0.037 | -1.451 | -0.011 | 1.47E-01 | 18607 | 0.870 | 0.151 | 1.589 | 0.367 | 2.372 | 0.017 | 1.77E-02 | 18697 |
| 120 | Vermis_10 | -0.019 | -0.039 | 0.001 | 0.010 | -1.861 | -0.014 | 6.28E-02 | 18607 | 0.317 | 0.117 | 0.518 | 0.102 | 3.103 | 0.023 | 1.92E-03 | 18697 |
| 121 | Thal_AV_L | 0.008 | 0.000 | 0.016 | 0.004 | 2.054 | 0.015 | 4.00E-02 | 18607 | -0.103 | -0.183 | -0.022 | 0.041 | -2.507 | -0.018 | 1.22E-02 | 18697 |
| 122 | Thal_AV_R | 0.011 | 0.000 | 0.023 | 0.006 | 1.897 | 0.014 | 5.79E-02 | 18607 | -0.174 | -0.289 | -0.060 | 0.059 | -2.978 | -0.022 | 2.90E-03 | 18697 |
| 123 | Thal_LP_L | 0.009 | 0.000 | 0.018 | 0.005 | 2.012 | 0.015 | 4.42E-02 | 18607 | -0.189 | -0.278 | -0.100 | 0.045 | -4.162 | -0.030 | 3.16E-05 | 18697 |
| 124 | Thal_LP_R | 0.012 | 0.000 | 0.023 | 0.006 | 1.991 | 0.015 | 4.65E-02 | 18607 | -0.244 | -0.360 | -0.128 | 0.059 | -4.121 | -0.030 | 3.79E-05 | 18697 |
| 125 | Thal_VA_L | -0.002 | -0.011 | 0.007 | 0.005 | -0.418 | -0.003 | 6.76E-01 | 18607 | -0.023 | -0.118 | 0.071 | 0.048 | -0.484 | -0.004 | 6.28E-01 | 18697 |
| 126 | Thal_VA_R | 0.005 | -0.007 | 0.016 | 0.006 | 0.783 | 0.006 | 4.34E-01 | 18607 | -0.059 | -0.175 | 0.058 | 0.059 | -0.985 | -0.007 | 3.25E-01 | 18697 |
| 127 | Thal_VL_L | 0.043 | 0.016 | 0.070 | 0.014 | 3.131 | 0.023 | 1.75E-03 | 18607 | -0.423 | -0.693 | -0.153 | 0.138 | -3.067 | -0.022 | 2.16E-03 | 18697 |
| 128 | Thal_VL_R | 0.056 | 0.021 | 0.092 | 0.018 | 3.087 | 0.023 | 2.02E-03 | 18607 | -0.620 | -0.979 | -0.262 | 0.183 | -3.389 | -0.025 | 7.03E-04 | 18697 |
| 129 | Thal_VPL_L | 0.007 | -0.025 | 0.039 | 0.016 | 0.411 | 0.003 | 6.81E-01 | 18607 | -0.673 | -0.995 | -0.351 | 0.164 | -4.100 | -0.030 | 4.14E-05 | 18697 |
| 130 | Thal_VPL_R | -0.001 | -0.034 | 0.033 | 0.017 | -0.034 | 0.000 | 9.73E-01 | 18607 | -0.729 | -1.067 | -0.390 | 0.173 | -4.222 | -0.031 | 2.44E-05 | 18697 |
| 131 | Thal_IL_L | 0.008 | -0.006 | 0.022 | 0.007 | 1.069 | 0.008 | 2.85E-01 | 18607 | -0.351 | -0.490 | -0.211 | 0.071 | -4.937 | -0.036 | 7.98E-07 | 18697 |
| 132 | Thal_IL_R | 0.013 | -0.003 | 0.029 | 0.008 | 1.593 | 0.012 | 1.11E-01 | 18607 | -0.421 | -0.579 | -0.263 | 0.081 | -5.231 | -0.038 | 1.70E-07 | 18697 |
| 133 | Thal_Re_L | 0.000 | 0.000 | 0.000 | 0.000 | -0.063 | 0.000 | 9.50E-01 | 18607 | 0.000 | -0.001 | 0.000 | 0.000 | -1.237 | -0.009 | 2.16E-01 | 18697 |
| 134 | Thal_Re_R | 0.000 | 0.000 | 0.001 | 0.000 | 1.109 | 0.008 | 2.67E-01 | 18607 | -0.006 | -0.011 | -0.001 | 0.002 | -2.462 | -0.018 | 1.38E-02 | 18697 |
| 135 | Thal_MDm_L | 0.097 | -0.006 | 0.200 | 0.053 | 1.852 | 0.014 | 6.41E-02 | 18607 | -3.011 | -4.041 | -1.980 | 0.526 | -5.727 | -0.042 | 1.04E-08 | 18697 |
| 136 | Thal_MDm_R | 0.104 | -0.016 | 0.223 | 0.061 | 1.699 | 0.012 | 8.94E-02 | 18607 | -3.364 | -4.562 | -2.167 | 0.611 | -5.508 | -0.040 | 3.68E-08 | 18697 |
| 137 | Thal_MDl_L | 0.026 | 0.009 | 0.044 | 0.009 | 2.903 | 0.021 | 3.70E-03 | 18607 | -0.534 | -0.712 | -0.355 | 0.091 | -5.862 | -0.043 | 4.65E-09 | 18697 |

| | | | | | | | | | | | | | | | | | |
|---|---|---|---|---|---|---|---|---|---|---|---|---|---|---|---|---|---|
| 138 | Thal_MDl_R | 0.022 | 0.002 | 0.043 | 0.010 | 2.176 | 0.016 | 2.96E-02 | 18607 | -0.584 | -0.787 | -0.382 | 0.103 | -5.649 | -0.041 | 1.63E-08 | 18697 |
| 139 | Thal_LGN_L | 0.002 | -0.001 | 0.004 | 0.001 | 1.250 | 0.009 | 2.11E-01 | 18607 | -0.043 | -0.070 | -0.016 | 0.014 | -3.130 | -0.023 | 1.75E-03 | 18697 |
| 140 | Thal_LGN_R | 0.002 | -0.004 | 0.008 | 0.003 | 0.684 | 0.005 | 4.94E-01 | 18607 | -0.119 | -0.176 | -0.061 | 0.029 | -4.033 | -0.029 | 5.52E-05 | 18697 |
| 141 | Thal_MGN_L | -0.001 | -0.006 | 0.003 | 0.002 | -0.602 | -0.004 | 5.47E-01 | 18607 | -0.111 | -0.159 | -0.063 | 0.025 | -4.501 | -0.033 | 6.79E-06 | 18697 |
| 142 | Thal_MGN_R | -0.002 | -0.007 | 0.004 | 0.003 | -0.600 | -0.004 | 5.48E-01 | 18607 | -0.134 | -0.187 | -0.082 | 0.027 | -5.010 | -0.037 | 5.50E-07 | 18697 |
| 143 | Thal_PuA_L | -0.002 | -0.008 | 0.003 | 0.003 | -0.856 | -0.006 | 3.92E-01 | 18607 | -0.148 | -0.206 | -0.091 | 0.029 | -5.094 | -0.037 | 3.54E-07 | 18697 |
| 144 | Thal_PuA_R | -0.003 | -0.015 | 0.009 | 0.006 | -0.500 | -0.004 | 6.17E-01 | 18607 | -0.332 | -0.451 | -0.214 | 0.061 | -5.486 | -0.040 | 4.16E-08 | 18697 |
| 145 | Thal_PuM_L | -0.003 | -0.116 | 0.110 | 0.058 | -0.052 | 0.000 | 9.59E-01 | 18607 | -3.384 | -4.519 | -2.250 | 0.579 | -5.846 | -0.043 | 5.11E-09 | 18697 |
| 146 | Thal_PuM_R | 0.001 | -0.140 | 0.142 | 0.072 | 0.012 | 0.000 | 9.90E-01 | 18607 | -4.120 | -5.534 | -2.707 | 0.721 | -5.713 | -0.042 | 1.13E-08 | 18697 |
| 147 | Thal_PuL_L | 0.004 | -0.017 | 0.025 | 0.011 | 0.375 | 0.003 | 7.08E-01 | 18607 | -0.610 | -0.817 | -0.403 | 0.106 | -5.777 | -0.042 | 7.73E-09 | 18697 |
| 148 | Thal_PuL_R | 0.005 | -0.028 | 0.038 | 0.017 | 0.300 | 0.002 | 7.64E-01 | 18607 | -0.942 | -1.271 | -0.613 | 0.168 | -5.611 | -0.041 | 2.04E-08 | 18697 |
| 149 | Thal_PuI_L | -0.001 | -0.007 | 0.004 | 0.003 | -0.446 | -0.003 | 6.55E-01 | 18607 | -0.125 | -0.181 | -0.068 | 0.029 | -4.342 | -0.032 | 1.42E-05 | 18697 |
| 150 | Thal_PuI_R | -0.004 | -0.012 | 0.003 | 0.004 | -1.073 | -0.008 | 2.83E-01 | 18607 | -0.121 | -0.195 | -0.046 | 0.038 | -3.175 | -0.023 | 1.50E-03 | 18697 |
| 151 | ACC_sub_L | 0.008 | -0.056 | 0.073 | 0.033 | 0.256 | 0.002 | 7.98E-01 | 18607 | -1.131 | -1.781 | -0.482 | 0.331 | -3.415 | -0.025 | 6.40E-04 | 18697 |
| 152 | ACC_sub_R | -0.008 | -0.069 | 0.052 | 0.031 | -0.266 | -0.002 | 7.90E-01 | 18607 | -0.749 | -1.356 | -0.142 | 0.310 | -2.418 | -0.018 | 1.56E-02 | 18697 |
| 153 | ACC_pre_L | 0.120 | -0.046 | 0.287 | 0.085 | 1.415 | 0.010 | 1.57E-01 | 18607 | -3.238 | -4.902 | -1.573 | 0.849 | -3.813 | -0.028 | 1.38E-04 | 18697 |
| 154 | ACC_pre_R | -0.038 | -0.198 | 0.122 | 0.082 | -0.467 | -0.003 | 6.41E-01 | 18607 | -3.459 | -5.063 | -1.856 | 0.818 | -4.229 | -0.031 | 2.36E-05 | 18697 |
| 155 | ACC_sup_L | 0.140 | -0.041 | 0.321 | 0.092 | 1.516 | 0.011 | 1.29E-01 | 18607 | -4.327 | -6.136 | -2.519 | 0.923 | -4.691 | -0.034 | 2.74E-06 | 18697 |
| 156 | ACC_sup_R | 0.030 | -0.157 | 0.218 | 0.096 | 0.316 | 0.002 | 7.52E-01 | 18607 | -4.159 | -6.033 | -2.285 | 0.956 | -4.349 | -0.032 | 1.37E-05 | 18697 |
| 157 | N_Acc_L | 0.001 | -0.053 | 0.056 | 0.028 | 0.054 | 0.000 | 9.57E-01 | 18607 | -0.879 | -1.424 | -0.335 | 0.278 | -3.168 | -0.023 | 1.54E-03 | 18697 |
| 158 | N_Acc_R | 0.025 | -0.022 | 0.072 | 0.024 | 1.039 | 0.008 | 2.99E-01 | 18607 | -0.795 | -1.266 | -0.324 | 0.240 | -3.309 | -0.024 | 9.38E-04 | 18697 |
| 159 | VTA_L | 0.001 | -0.001 | 0.003 | 0.001 | 0.962 | 0.007 | 3.36E-01 | 18607 | -0.007 | -0.026 | 0.012 | 0.010 | -0.695 | -0.005 | 4.87E-01 | 18697 |
| 160 | VTA_R | 0.001 | -0.001 | 0.003 | 0.001 | 1.232 | 0.009 | 2.18E-01 | 18607 | -0.012 | -0.029 | 0.005 | 0.009 | -1.377 | -0.010 | 1.69E-01 | 18697 |
| 161 | SN_pc_L | 0.000 | -0.003 | 0.003 | 0.002 | 0.170 | 0.001 | 8.65E-01 | 18607 | -0.033 | -0.064 | -0.002 | 0.016 | -2.094 | -0.015 | 3.63E-02 | 18697 |
| 162 | SN_pc_R | 0.001 | -0.003 | 0.005 | 0.002 | 0.315 | 0.002 | 7.53E-01 | 18607 | -0.051 | -0.091 | -0.010 | 0.021 | -2.462 | -0.018 | 1.38E-02 | 18697 |
| 163 | SN_pr_L | 0.001 | -0.003 | 0.005 | 0.002 | 0.439 | 0.003 | 6.61E-01 | 18607 | -0.036 | -0.077 | 0.006 | 0.021 | -1.677 | -0.012 | 9.35E-02 | 18697 |
| 164 | SN_pr_R | 0.000 | -0.004 | 0.005 | 0.002 | 0.146 | 0.001 | 8.84E-01 | 18607 | -0.031 | -0.078 | 0.016 | 0.024 | -1.289 | -0.009 | 1.97E-01 | 18697 |
| 165 | Red_N_L | 0.002 | -0.004 | 0.007 | 0.003 | 0.617 | 0.005 | 5.37E-01 | 18607 | -0.082 | -0.134 | -0.031 | 0.026 | -3.119 | -0.023 | 1.82E-03 | 18697 |
| 166 | Red_N_R | 0.002 | -0.003 | 0.008 | 0.003 | 0.782 | 0.006 | 4.34E-01 | 18607 | -0.072 | -0.128 | -0.016 | 0.029 | -2.507 | -0.018 | 1.22E-02 | 18697 |
| 167 | LC_L | -0.001 | -0.003 | 0.001 | 0.001 | -0.748 | -0.005 | 4.54E-01 | 18607 | -0.011 | -0.030 | 0.008 | 0.010 | -1.123 | -0.008 | 2.62E-01 | 18697 |
| 168 | LC_R | 0.000 | -0.002 | 0.002 | 0.001 | -0.371 | -0.003 | 7.11E-01 | 18607 | -0.011 | -0.032 | 0.010 | 0.011 | -1.012 | -0.007 | 3.11E-01 | 18697 |
| 169 | Raphe_D | 0.001 | -0.006 | 0.009 | 0.004 | 0.320 | 0.002 | 7.49E-01 | 18607 | -0.123 | -0.198 | -0.049 | 0.038 | -3.238 | -0.024 | 1.21E-03 | 18697 |
| 170 | Raphe_M | 0.001 | -0.001 | 0.003 | 0.001 | 0.720 | 0.005 | 4.72E-01 | 18607 | -0.017 | -0.040 | 0.005 | 0.012 | -1.503 | -0.011 | 1.33E-01 | 18697 |

| | | Oily fish intake | | | | | | | | Cheese intake | | | | | | | |
|---|---|---|---|---|---|---|---|---|---|---|---|---|---|---|---|---|---|
| **Number** | **AAL 3 region** | **beta** | **95%CI_l** | **95%CI_h** | **se** | **t** | **r** | **p** | **df.e** | **beta** | **95%CI_l** | **95%CI_h** | **se** | **t** | **r** | **p** | **df.e** |
| 0 | Total brain | -3.612 | -150.408 | 143.185 | 74.893 | -0.048 | 0.000 | 9.62E-01 | 18725 | -49.240 | -173.822 | 75.342 | 63.559 | -0.775 | -0.006 | 4.39E-01 | 18410 |
| 1 | Precentral_L | -1.180 | -6.346 | 3.985 | 2.635 | -0.448 | -0.003 | 6.54E-01 | 18725 | -5.890 | -10.269 | -1.512 | 2.234 | -2.637 | -0.019 | 8.37E-03 | 18410 |
| 2 | Precentral_R | -0.714 | -5.447 | 4.019 | 2.415 | -0.296 | -0.002 | 7.67E-01 | 18725 | -4.187 | -8.199 | -0.175 | 2.047 | -2.046 | -0.015 | 4.08E-02 | 18410 |
| 3 | Frontal_Sup_2_L | 3.072 | -2.380 | 8.524 | 2.782 | 1.105 | 0.008 | 2.69E-01 | 18725 | 2.129 | -2.497 | 6.755 | 2.360 | 0.902 | 0.007 | 3.67E-01 | 18410 |
| 4 | Frontal_Sup_2_R | -0.167 | -6.177 | 5.843 | 3.066 | -0.055 | 0.000 | 9.56E-01 | 18725 | 1.036 | -4.068 | 6.140 | 2.604 | 0.398 | 0.003 | 6.91E-01 | 18410 |
| 5 | Frontal_Mid_2_L | -2.088 | -7.814 | 3.639 | 2.922 | -0.715 | -0.005 | 4.75E-01 | 18725 | 2.408 | -2.454 | 7.270 | 2.481 | 0.971 | 0.007 | 3.32E-01 | 18410 |
| 6 | Frontal_Mid_2_R | -4.439 | -10.919 | 2.042 | 3.306 | -1.343 | -0.010 | 1.79E-01 | 18725 | 2.333 | -3.165 | 7.830 | 2.805 | 0.832 | 0.006 | 4.06E-01 | 18410 |
| 7 | Frontal_Inf_Oper_L | -0.618 | -2.310 | 1.074 | 0.863 | -0.715 | -0.005 | 4.74E-01 | 18725 | 1.067 | -0.369 | 2.503 | 0.733 | 1.457 | 0.011 | 1.45E-01 | 18410 |
| 8 | Frontal_Inf_Oper_R | -0.950 | -3.072 | 1.171 | 1.082 | -0.878 | -0.006 | 3.80E-01 | 18725 | 0.474 | -1.327 | 2.276 | 0.919 | 0.516 | 0.004 | 6.06E-01 | 18410 |
| 9 | Frontal_Inf_Tri_L | -1.081 | -4.675 | 2.512 | 1.833 | -0.590 | -0.004 | 5.55E-01 | 18725 | 4.286 | 1.236 | 7.335 | 1.556 | 2.755 | 0.020 | 5.88E-03 | 18410 |
| 10 | Frontal_Inf_Tri_R | 0.166 | -2.656 | 2.988 | 1.440 | 0.115 | 0.001 | 9.08E-01 | 18725 | 2.038 | -0.360 | 4.435 | 1.223 | 1.666 | 0.012 | 9.57E-02 | 18410 |
| 11 | Frontal_Inf_Orb_2_L | 0.271 | -0.803 | 1.345 | 0.548 | 0.494 | 0.004 | 6.21E-01 | 18725 | 1.867 | 0.954 | 2.779 | 0.466 | 4.010 | 0.030 | 6.10E-05 | 18410 |
| 12 | Frontal_Inf_Orb_2_R | -0.017 | -1.140 | 1.107 | 0.573 | -0.029 | 0.000 | 9.77E-01 | 18725 | 2.326 | 1.372 | 3.280 | 0.487 | 4.780 | 0.035 | 1.76E-06 | 18410 |

| | | | | | | | | | | | | | | | | | |
|---|---|---|---|---|---|---|---|---|---|---|---|---|---|---|---|---|---|
| 13 | Rolandic_Oper_L | 1.660 | 0.036 | 3.285 | 0.829 | 2.003 | 0.015 | 4.52E-02 | 18725 | 0.382 | -0.998 | 1.762 | 0.704 | 0.543 | 0.004 | 5.87E-01 | 18410 |
| 14 | Rolandic_Oper_R | 2.288 | -0.038 | 4.613 | 1.186 | 1.928 | 0.014 | 5.38E-02 | 18725 | -1.090 | -3.066 | 0.885 | 1.008 | -1.082 | -0.008 | 2.79E-01 | 18410 |
| 15 | Supp_Motor_Area_L | -1.890 | -5.751 | 1.972 | 1.970 | -0.959 | -0.007 | 3.37E-01 | 18725 | -4.312 | -7.589 | -1.036 | 1.672 | -2.579 | -0.019 | 9.90E-03 | 18410 |
| 16 | Supp_Motor_Area_R | -1.633 | -5.901 | 2.635 | 2.178 | -0.750 | -0.005 | 4.53E-01 | 18725 | -3.661 | -7.284 | -0.038 | 1.848 | -1.981 | -0.015 | 4.76E-02 | 18410 |
| 17 | Olfactory_L | 0.343 | -0.231 | 0.917 | 0.293 | 1.171 | 0.009 | 2.42E-01 | 18725 | -0.045 | -0.533 | 0.442 | 0.249 | -0.182 | -0.001 | 8.56E-01 | 18410 |
| 18 | Olfactory_R | 0.003 | -0.572 | 0.579 | 0.294 | 0.011 | 0.000 | 9.91E-01 | 18725 | -0.091 | -0.579 | 0.398 | 0.249 | -0.364 | -0.003 | 7.16E-01 | 18410 |
| 19 | Frontal_Sup_Medial_L | -2.178 | -5.797 | 1.442 | 1.847 | -1.179 | -0.009 | 2.38E-01 | 18725 | 2.776 | -0.298 | 5.850 | 1.568 | 1.770 | 0.013 | 7.67E-02 | 18410 |
| 20 | Frontal_Sup_Medial_R | -0.979 | -3.614 | 1.656 | 1.344 | -0.728 | -0.005 | 4.66E-01 | 18725 | 0.683 | -1.553 | 2.919 | 1.141 | 0.599 | 0.004 | 5.49E-01 | 18410 |
| 21 | Frontal_Med_Orb_L | -0.377 | -1.730 | 0.976 | 0.690 | -0.546 | -0.004 | 5.85E-01 | 18725 | 1.013 | -0.135 | 2.162 | 0.586 | 1.729 | 0.013 | 8.38E-02 | 18410 |
| 22 | Frontal_Med_Orb_R | 0.673 | -0.989 | 2.336 | 0.848 | 0.794 | 0.006 | 4.27E-01 | 18725 | 1.358 | -0.054 | 2.769 | 0.720 | 1.885 | 0.014 | 5.94E-02 | 18410 |
| 23 | Rectus_L | 0.330 | -1.070 | 1.730 | 0.714 | 0.462 | 0.003 | 6.44E-01 | 18725 | 0.656 | -0.532 | 1.845 | 0.606 | 1.082 | 0.008 | 2.79E-01 | 18410 |
| 24 | Rectus_R | 0.942 | -0.353 | 2.237 | 0.661 | 1.426 | 0.010 | 1.54E-01 | 18725 | 0.640 | -0.460 | 1.739 | 0.561 | 1.140 | 0.008 | 2.54E-01 | 18410 |
| 25 | OFCmed_L | 0.076 | -0.735 | 0.888 | 0.414 | 0.185 | 0.001 | 8.54E-01 | 18725 | 0.062 | -0.627 | 0.752 | 0.352 | 0.178 | 0.001 | 8.59E-01 | 18410 |
| 26 | OFCmed_R | 0.509 | -0.436 | 1.454 | 0.482 | 1.055 | 0.008 | 2.91E-01 | 18725 | 0.492 | -0.311 | 1.295 | 0.410 | 1.201 | 0.009 | 2.30E-01 | 18410 |
| 27 | OFCant_L | -0.321 | -1.259 | 0.617 | 0.479 | -0.671 | -0.005 | 5.02E-01 | 18725 | -0.552 | -1.349 | 0.245 | 0.407 | -1.357 | -0.010 | 1.75E-01 | 18410 |
| 28 | OFCant_R | -0.653 | -1.893 | 0.586 | 0.632 | -1.033 | -0.008 | 3.01E-01 | 18725 | 0.817 | -0.236 | 1.870 | 0.537 | 1.521 | 0.011 | 1.28E-01 | 18410 |
| 29 | OFCpost_L | 0.447 | -0.728 | 1.621 | 0.599 | 0.745 | 0.005 | 4.56E-01 | 18725 | 0.244 | -0.754 | 1.242 | 0.509 | 0.480 | 0.004 | 6.31E-01 | 18410 |
| 30 | OFCpost_R | -0.422 | -1.507 | 0.664 | 0.554 | -0.761 | -0.006 | 4.47E-01 | 18725 | 0.594 | -0.327 | 1.515 | 0.470 | 1.264 | 0.009 | 2.06E-01 | 18410 |
| 31 | OFClat_L | 0.004 | -0.392 | 0.401 | 0.202 | 0.022 | 0.000 | 9.83E-01 | 18725 | 0.517 | 0.180 | 0.853 | 0.172 | 3.009 | 0.022 | 2.63E-03 | 18410 |
| 32 | OFClat_R | -0.254 | -0.589 | 0.081 | 0.171 | -1.486 | -0.011 | 1.37E-01 | 18725 | 0.777 | 0.492 | 1.061 | 0.145 | 5.358 | 0.039 | 8.52E-08 | 18410 |
| 33 | Insula_L | 0.752 | -2.880 | 4.384 | 1.853 | 0.406 | 0.003 | 6.85E-01 | 18725 | 2.078 | -1.006 | 5.161 | 1.573 | 1.321 | 0.010 | 1.87E-01 | 18410 |
| 34 | Insula_R | 0.501 | -3.038 | 4.040 | 1.806 | 0.277 | 0.002 | 7.81E-01 | 18725 | 0.935 | -2.069 | 3.939 | 1.533 | 0.610 | 0.004 | 5.42E-01 | 18410 |
| 37 | Cingulate_Mid_L | -2.244 | -5.314 | 0.826 | 1.566 | -1.433 | -0.010 | 1.52E-01 | 18725 | -0.526 | -3.134 | 2.081 | 1.330 | -0.396 | -0.003 | 6.92E-01 | 18410 |
| 38 | Cingulate_Mid_R | -1.738 | -5.013 | 1.536 | 1.671 | -1.041 | -0.008 | 2.98E-01 | 18725 | -1.617 | -4.397 | 1.162 | 1.418 | -1.140 | -0.008 | 2.54E-01 | 18410 |
| 39 | Cingulate_Post_L | -0.823 | -1.761 | 0.115 | 0.479 | -1.719 | -0.013 | 8.56E-02 | 18725 | -1.021 | -1.817 | -0.225 | 0.406 | -2.514 | -0.019 | 1.20E-02 | 18410 |
| 40 | Cingulate_Post_R | -0.356 | -0.876 | 0.164 | 0.265 | -1.341 | -0.010 | 1.80E-01 | 18725 | -0.774 | -1.216 | -0.333 | 0.225 | -3.439 | -0.025 | 5.86E-04 | 18410 |
| 41 | Hippocampus_L | 1.549 | 0.032 | 3.066 | 0.774 | 2.001 | 0.015 | 4.54E-02 | 18725 | -0.154 | -1.444 | 1.135 | 0.658 | -0.235 | -0.002 | 8.15E-01 | 18410 |
| 42 | Hippocampus_R | 1.892 | 0.319 | 3.464 | 0.802 | 2.357 | 0.017 | 1.84E-02 | 18725 | 0.163 | -1.175 | 1.501 | 0.682 | 0.239 | 0.002 | 8.11E-01 | 18410 |
| 43 | ParaHippocampal_L | 0.521 | -0.827 | 1.869 | 0.688 | 0.758 | 0.006 | 4.49E-01 | 18725 | 0.241 | -0.904 | 1.387 | 0.584 | 0.413 | 0.003 | 6.79E-01 | 18410 |
| 44 | ParaHippocampal_R | 1.074 | -0.627 | 2.775 | 0.868 | 1.238 | 0.009 | 2.16E-01 | 18725 | 0.792 | -0.654 | 2.237 | 0.737 | 1.074 | 0.008 | 2.83E-01 | 18410 |
| 45 | Amygdala_L | -0.396 | -0.920 | 0.128 | 0.267 | -1.480 | -0.011 | 1.39E-01 | 18725 | 0.252 | -0.192 | 0.697 | 0.227 | 1.112 | 0.008 | 2.66E-01 | 18410 |
| 46 | Amygdala_R | -0.202 | -0.738 | 0.334 | 0.273 | -0.739 | -0.005 | 4.60E-01 | 18725 | 0.194 | -0.261 | 0.649 | 0.232 | 0.835 | 0.006 | 4.04E-01 | 18410 |
| 47 | Calcarine_L | 2.203 | -2.796 | 7.202 | 2.550 | 0.864 | 0.006 | 3.88E-01 | 18725 | -12.034 | -16.272 | -7.796 | 2.162 | -5.565 | -0.041 | 2.65E-08 | 18410 |
| 48 | Calcarine_R | 1.692 | -2.485 | 5.869 | 2.131 | 0.794 | 0.006 | 4.27E-01 | 18725 | -9.885 | -13.427 | -6.344 | 1.807 | -5.471 | -0.040 | 4.54E-08 | 18410 |
| 49 | Cuneus_L | 1.368 | -1.768 | 4.504 | 1.600 | 0.855 | 0.006 | 3.92E-01 | 18725 | -5.914 | -8.574 | -3.253 | 1.357 | -4.357 | -0.032 | 1.33E-05 | 18410 |
| 50 | Cuneus_R | 0.212 | -2.187 | 2.612 | 1.224 | 0.174 | 0.001 | 8.62E-01 | 18725 | -5.287 | -7.323 | -3.251 | 1.039 | -5.090 | -0.037 | 3.61E-07 | 18410 |
| 51 | Lingual_L | 0.703 | -2.303 | 3.709 | 1.534 | 0.459 | 0.003 | 6.47E-01 | 18725 | -4.511 | -7.065 | -1.957 | 1.303 | -3.463 | -0.026 | 5.36E-04 | 18410 |
| 52 | Lingual_R | 0.667 | -2.483 | 3.817 | 1.607 | 0.415 | 0.003 | 6.78E-01 | 18725 | -3.761 | -6.437 | -1.085 | 1.365 | -2.755 | -0.020 | 5.87E-03 | 18410 |
| 53 | Occipital_Sup_L | 0.660 | -1.407 | 2.727 | 1.054 | 0.626 | 0.005 | 5.31E-01 | 18725 | -1.880 | -3.636 | -0.124 | 0.896 | -2.099 | -0.015 | 3.58E-02 | 18410 |
| 54 | Occipital_Sup_R | -0.228 | -2.469 | 2.013 | 1.143 | -0.200 | -0.001 | 8.42E-01 | 18725 | -1.819 | -3.722 | 0.085 | 0.971 | -1.873 | -0.014 | 6.11E-02 | 18410 |
| 55 | Occipital_Mid_L | 0.888 | -3.957 | 5.733 | 2.472 | 0.359 | 0.003 | 7.19E-01 | 18725 | -5.753 | -9.865 | -1.642 | 2.098 | -2.743 | -0.020 | 6.10E-03 | 18410 |
| 56 | Occipital_Mid_R | -2.427 | -5.797 | 0.944 | 1.720 | -1.411 | -0.010 | 1.58E-01 | 18725 | -3.686 | -6.547 | -0.825 | 1.460 | -2.525 | -0.019 | 1.16E-02 | 18410 |
| 57 | Occipital_Inf_L | 0.795 | -0.813 | 2.403 | 0.820 | 0.969 | 0.007 | 3.32E-01 | 18725 | -3.000 | -4.365 | -1.635 | 0.696 | -4.307 | -0.032 | 1.66E-05 | 18410 |
| 58 | Occipital_Inf_R | -3.225 | -5.082 | -1.367 | 0.947 | -3.403 | -0.025 | 6.67E-04 | 18725 | -4.719 | -6.293 | -3.144 | 0.803 | -5.873 | -0.043 | 4.35E-09 | 18410 |
| 59 | Fusiform_L | 1.160 | -2.360 | 4.679 | 1.796 | 0.646 | 0.005 | 5.18E-01 | 18725 | -0.883 | -3.871 | 2.105 | 1.524 | -0.579 | -0.004 | 5.63E-01 | 18410 |
| 60 | Fusiform_R | 2.524 | -1.347 | 6.394 | 1.974 | 1.278 | 0.009 | 2.01E-01 | 18725 | 0.389 | -2.899 | 3.677 | 1.677 | 0.232 | 0.002 | 8.17E-01 | 18410 |
| 61 | Postcentral_L | 1.889 | -3.406 | 7.184 | 2.702 | 0.699 | 0.005 | 4.84E-01 | 18725 | 3.805 | -0.687 | 8.298 | 2.292 | 1.660 | 0.012 | 9.69E-02 | 18410 |
| 62 | Postcentral_R | 4.110 | -1.317 | 9.536 | 2.769 | 1.484 | 0.011 | 1.38E-01 | 18725 | 2.459 | -2.150 | 7.067 | 2.351 | 1.046 | 0.008 | 2.96E-01 | 18410 |

| | | | | | | | | | | | | | | | | | |
|---|---|---|---|---|---|---|---|---|---|---|---|---|---|---|---|---|---|
| 63 | Parietal_Sup_L | -3.187 | -6.237 | -0.137 | 1.556 | -2.048 | -0.015 | 4.06E-02 | 18725 | -3.600 | -6.191 | -1.010 | 1.322 | -2.724 | -0.020 | 6.45E-03 | 18410 |
| 64 | Parietal_Sup_R | -1.807 | -4.513 | 0.899 | 1.380 | -1.309 | -0.010 | 1.91E-01 | 18725 | -4.784 | -7.079 | -2.488 | 1.171 | -4.084 | -0.030 | 4.44E-05 | 18410 |
| 65 | Parietal_Inf_L | -1.990 | -5.985 | 2.004 | 2.038 | -0.977 | -0.007 | 3.29E-01 | 18725 | -0.522 | -3.912 | 2.869 | 1.730 | -0.302 | -0.002 | 7.63E-01 | 18410 |
| 66 | Parietal_Inf_R | -1.324 | -3.912 | 1.263 | 1.320 | -1.003 | -0.007 | 3.16E-01 | 18725 | -1.669 | -3.862 | 0.525 | 1.119 | -1.491 | -0.011 | 1.36E-01 | 18410 |
| 67 | SupraMarginal_L | 0.420 | -1.921 | 2.761 | 1.194 | 0.351 | 0.003 | 7.25E-01 | 18725 | 1.479 | -0.508 | 3.467 | 1.014 | 1.459 | 0.011 | 1.45E-01 | 18410 |
| 68 | SupraMarginal_R | -0.608 | -3.920 | 2.704 | 1.690 | -0.360 | -0.003 | 7.19E-01 | 18725 | 2.360 | -0.452 | 5.172 | 1.435 | 1.645 | 0.012 | 1.00E-01 | 18410 |
| 69 | Angular_L | 1.143 | -1.155 | 3.440 | 1.172 | 0.975 | 0.007 | 3.30E-01 | 18725 | -1.337 | -3.287 | 0.613 | 0.995 | -1.344 | -0.010 | 1.79E-01 | 18410 |
| 70 | Angular_R | 0.754 | -2.329 | 3.838 | 1.573 | 0.480 | 0.004 | 6.32E-01 | 18725 | -2.280 | -4.900 | 0.340 | 1.337 | -1.706 | -0.013 | 8.81E-02 | 18410 |
| 71 | Precuneus_L | -2.128 | -7.731 | 3.475 | 2.858 | -0.745 | -0.005 | 4.57E-01 | 18725 | -6.370 | -11.124 | -1.615 | 2.426 | -2.626 | -0.019 | 8.65E-03 | 18410 |
| 72 | Precuneus_R | -3.935 | -8.926 | 1.057 | 2.547 | -1.545 | -0.011 | 1.22E-01 | 18725 | -8.197 | -12.435 | -3.960 | 2.162 | -3.792 | -0.028 | 1.50E-04 | 18410 |
| 73 | Paracentral_Lobule_L | -0.455 | -3.021 | 2.110 | 1.309 | -0.348 | -0.003 | 7.28E-01 | 18725 | -4.074 | -6.250 | -1.897 | 1.110 | -3.669 | -0.027 | 2.44E-04 | 18410 |
| 74 | Paracentral_Lobule_R | -1.051 | -2.888 | 0.786 | 0.937 | -1.121 | -0.008 | 2.62E-01 | 18725 | -1.856 | -3.415 | -0.296 | 0.796 | -2.332 | -0.017 | 1.97E-02 | 18410 |
| 75 | Caudate_L | -0.480 | -2.529 | 1.568 | 1.045 | -0.460 | -0.003 | 6.46E-01 | 18725 | 0.264 | -1.474 | 2.003 | 0.887 | 0.298 | 0.002 | 7.66E-01 | 18410 |
| 76 | Caudate_R | -0.698 | -3.017 | 1.621 | 1.183 | -0.590 | -0.004 | 5.55E-01 | 18725 | 0.146 | -1.820 | 2.111 | 1.003 | 0.145 | 0.001 | 8.85E-01 | 18410 |
| 77 | Putamen_L | -2.881 | -5.993 | 0.231 | 1.588 | -1.815 | -0.013 | 6.96E-02 | 18725 | 0.342 | -2.298 | 2.982 | 1.347 | 0.254 | 0.002 | 8.00E-01 | 18410 |
| 78 | Putamen_R | -3.318 | -6.488 | -0.149 | 1.617 | -2.052 | -0.015 | 4.02E-02 | 18725 | -0.059 | -2.748 | 2.631 | 1.372 | -0.043 | 0.000 | 9.66E-01 | 18410 |
| 79 | Pallidum_L | -0.255 | -0.531 | 0.022 | 0.141 | -1.806 | -0.013 | 7.10E-02 | 18725 | 0.068 | -0.167 | 0.303 | 0.120 | 0.565 | 0.004 | 5.72E-01 | 18410 |
| 80 | Pallidum_R | -0.259 | -0.562 | 0.045 | 0.155 | -1.671 | -0.012 | 9.48E-02 | 18725 | -0.038 | -0.295 | 0.220 | 0.132 | -0.286 | -0.002 | 7.75E-01 | 18410 |
| 83 | Heschl_L | 0.318 | -0.243 | 0.879 | 0.286 | 1.111 | 0.008 | 2.67E-01 | 18725 | -0.085 | -0.561 | 0.391 | 0.243 | -0.350 | -0.003 | 7.26E-01 | 18410 |
| 84 | Heschl_R | 0.350 | -0.277 | 0.977 | 0.320 | 1.095 | 0.008 | 2.73E-01 | 18725 | -0.022 | -0.554 | 0.510 | 0.272 | -0.081 | -0.001 | 9.36E-01 | 18410 |
| 85 | Temporal_Sup_L | 2.299 | -1.753 | 6.350 | 2.067 | 1.112 | 0.008 | 2.66E-01 | 18725 | 2.484 | -0.955 | 5.924 | 1.755 | 1.416 | 0.010 | 1.57E-01 | 18410 |
| 86 | Temporal_Sup_R | 3.586 | -0.896 | 8.069 | 2.287 | 1.568 | 0.011 | 1.17E-01 | 18725 | 2.380 | -1.427 | 6.186 | 1.942 | 1.225 | 0.009 | 2.20E-01 | 18410 |
| 87 | Temporal_Pole_Sup_L | -0.392 | -2.311 | 1.527 | 0.979 | -0.400 | -0.003 | 6.89E-01 | 18725 | 0.537 | -1.092 | 2.165 | 0.831 | 0.646 | 0.005 | 5.18E-01 | 18410 |
| 88 | Temporal_Pole_Sup_R | 0.803 | -1.218 | 2.825 | 1.031 | 0.779 | 0.006 | 4.36E-01 | 18725 | 1.405 | -0.311 | 3.122 | 0.876 | 1.605 | 0.012 | 1.09E-01 | 18410 |
| 89 | Temporal_Mid_L | 4.999 | -1.755 | 11.753 | 3.446 | 1.451 | 0.011 | 1.47E-01 | 18725 | 3.251 | -2.481 | 8.984 | 2.925 | 1.112 | 0.008 | 2.66E-01 | 18410 |
| 90 | Temporal_Mid_R | -0.373 | -6.370 | 5.624 | 3.059 | -0.122 | -0.001 | 9.03E-01 | 18725 | -1.169 | -6.261 | 3.924 | 2.598 | -0.450 | -0.003 | 6.53E-01 | 18410 |
| 91 | Temporal_Pole_Mid_L | 0.805 | -0.808 | 2.417 | 0.823 | 0.978 | 0.007 | 3.28E-01 | 18725 | 0.413 | -0.957 | 1.782 | 0.699 | 0.591 | 0.004 | 5.55E-01 | 18410 |
| 92 | Temporal_Pole_Mid_R | 2.464 | 0.503 | 4.424 | 1.000 | 2.463 | 0.018 | 1.38E-02 | 18725 | 1.793 | 0.129 | 3.457 | 0.849 | 2.112 | 0.016 | 3.47E-02 | 18410 |
| 93 | Temporal_Inf_L | 0.965 | -3.677 | 5.608 | 2.368 | 0.407 | 0.003 | 6.84E-01 | 18725 | -1.524 | -5.466 | 2.419 | 2.011 | -0.758 | -0.006 | 4.49E-01 | 18410 |
| 94 | Temporal_Inf_R | 2.653 | -2.411 | 7.716 | 2.583 | 1.027 | 0.008 | 3.04E-01 | 18725 | -1.078 | -5.372 | 3.217 | 2.191 | -0.492 | -0.004 | 6.23E-01 | 18410 |
| 95 | Cerebellum_Crus1_L | 4.553 | -0.957 | 10.064 | 2.811 | 1.620 | 0.012 | 1.05E-01 | 18725 | 0.451 | -4.216 | 5.118 | 2.381 | 0.189 | 0.001 | 8.50E-01 | 18410 |
| 96 | Cerebellum_Crus1_R | 0.255 | -5.364 | 5.875 | 2.867 | 0.089 | 0.001 | 9.29E-01 | 18725 | -0.414 | -5.175 | 4.347 | 2.429 | -0.170 | -0.001 | 8.65E-01 | 18410 |
| 97 | Cerebellum_Crus2_L | -0.762 | -5.135 | 3.610 | 2.231 | -0.342 | -0.002 | 7.33E-01 | 18725 | 4.712 | 1.006 | 8.419 | 1.891 | 2.492 | 0.018 | 1.27E-02 | 18410 |
| 98 | Cerebellum_Crus2_R | 1.780 | -2.264 | 5.824 | 2.063 | 0.863 | 0.006 | 3.88E-01 | 18725 | 3.268 | -0.160 | 6.696 | 1.749 | 1.868 | 0.014 | 6.17E-02 | 18410 |
| 99 | Cerebellum_3_L | 0.107 | -0.187 | 0.400 | 0.150 | 0.714 | 0.005 | 4.75E-01 | 18725 | -0.341 | -0.590 | -0.093 | 0.127 | -2.689 | -0.020 | 7.17E-03 | 18410 |
| 100 | Cerebellum_3_R | -0.034 | -0.430 | 0.362 | 0.202 | -0.167 | -0.001 | 8.68E-01 | 18725 | -0.560 | -0.896 | -0.224 | 0.171 | -3.270 | -0.024 | 1.08E-03 | 18410 |
| 101 | Cerebellum_4_5_L | 0.127 | -2.246 | 2.500 | 1.211 | 0.105 | 0.001 | 9.17E-01 | 18725 | -1.777 | -3.789 | 0.236 | 1.027 | -1.730 | -0.013 | 8.36E-02 | 18410 |
| 102 | Cerebellum_4_5_R | -0.057 | -1.991 | 1.876 | 0.986 | -0.058 | 0.000 | 9.54E-01 | 18725 | -1.521 | -3.161 | 0.119 | 0.837 | -1.818 | -0.013 | 6.90E-02 | 18410 |
| 103 | Cerebellum_6_L | 1.698 | -2.403 | 5.799 | 2.092 | 0.812 | 0.006 | 4.17E-01 | 18725 | -2.276 | -5.752 | 1.201 | 1.774 | -1.283 | -0.009 | 1.99E-01 | 18410 |
| 104 | Cerebellum_6_R | 0.224 | -4.097 | 4.545 | 2.205 | 0.102 | 0.001 | 9.19E-01 | 18725 | -2.053 | -5.718 | 1.611 | 1.869 | -1.098 | -0.008 | 2.72E-01 | 18410 |
| 105 | Cerebellum_7b_L | -0.135 | -1.471 | 1.202 | 0.682 | -0.197 | -0.001 | 8.44E-01 | 18725 | 2.013 | 0.881 | 3.146 | 0.578 | 3.484 | 0.026 | 4.95E-04 | 18410 |
| 106 | Cerebellum_7b_R | 0.573 | -0.752 | 1.897 | 0.676 | 0.848 | 0.006 | 3.97E-01 | 18725 | 2.223 | 1.100 | 3.347 | 0.573 | 3.880 | 0.029 | 1.05E-04 | 18410 |
| 107 | Cerebellum_8_L | -1.926 | -6.168 | 2.317 | 2.165 | -0.890 | -0.007 | 3.74E-01 | 18725 | 2.434 | -1.160 | 6.028 | 1.834 | 1.328 | 0.010 | 1.84E-01 | 18410 |
| 108 | Cerebellum_8_R | -1.540 | -7.062 | 3.982 | 2.817 | -0.547 | -0.004 | 5.85E-01 | 18725 | 6.831 | 2.151 | 11.511 | 2.388 | 2.861 | 0.021 | 4.23E-03 | 18410 |
| 109 | Cerebellum_9_L | -1.924 | -4.322 | 0.475 | 1.224 | -1.572 | -0.011 | 1.16E-01 | 18725 | 0.108 | -1.927 | 2.143 | 1.038 | 0.104 | 0.001 | 9.17E-01 | 18410 |
| 110 | Cerebellum_9_R | -3.134 | -5.491 | -0.776 | 1.203 | -2.605 | -0.019 | 9.19E-03 | 18725 | 0.366 | -1.633 | 2.365 | 1.020 | 0.359 | 0.003 | 7.20E-01 | 18410 |
| 111 | Cerebellum_10_L | -0.097 | -0.267 | 0.073 | 0.087 | -1.120 | -0.008 | 2.63E-01 | 18725 | -0.160 | -0.305 | -0.016 | 0.074 | -2.175 | -0.016 | 2.96E-02 | 18410 |
| 112 | Cerebellum_10_R | 0.019 | -0.158 | 0.196 | 0.090 | 0.214 | 0.002 | 8.31E-01 | 18725 | 0.000 | -0.150 | 0.150 | 0.077 | 0.004 | 0.000 | 9.97E-01 | 18410 |

| | | | | | | | | | | | | | | | | | |
|---|---|---|---|---|---|---|---|---|---|---|---|---|---|---|---|---|---|
| 113 | Vermis_1_2 | 0.058 | -0.053 | 0.168 | 0.056 | 1.025 | 0.007 | 3.05E-01 | 18725 | -0.156 | -0.250 | -0.062 | 0.048 | -3.260 | -0.024 | 1.12E-03 | 18410 |
| 114 | Vermis_3 | 0.252 | -0.188 | 0.692 | 0.224 | 1.122 | 0.008 | 2.62E-01 | 18725 | -0.144 | -0.517 | 0.228 | 0.190 | -0.759 | -0.006 | 4.48E-01 | 18410 |
| 115 | Vermis_4_5 | -0.372 | -1.665 | 0.920 | 0.659 | -0.565 | -0.004 | 5.72E-01 | 18725 | 0.110 | -0.987 | 1.206 | 0.559 | 0.196 | 0.001 | 8.45E-01 | 18410 |
| 116 | Vermis_6 | -0.425 | -1.144 | 0.295 | 0.367 | -1.157 | -0.008 | 2.47E-01 | 18725 | -0.556 | -1.167 | 0.054 | 0.311 | -1.787 | -0.013 | 7.40E-02 | 18410 |
| 117 | Vermis_7 | -0.174 | -0.581 | 0.233 | 0.208 | -0.839 | -0.006 | 4.02E-01 | 18725 | -0.327 | -0.671 | 0.018 | 0.176 | -1.856 | -0.014 | 6.35E-02 | 18410 |
| 118 | Vermis_8 | -0.096 | -0.764 | 0.573 | 0.341 | -0.280 | -0.002 | 7.79E-01 | 18725 | -0.180 | -0.747 | 0.387 | 0.289 | -0.623 | -0.005 | 5.34E-01 | 18410 |
| 119 | Vermis_9 | -0.527 | -1.068 | 0.015 | 0.276 | -1.904 | -0.014 | 5.69E-02 | 18725 | 0.112 | -0.347 | 0.571 | 0.234 | 0.478 | 0.004 | 6.33E-01 | 18410 |
| 120 | Vermis_10 | -0.112 | -0.263 | 0.039 | 0.077 | -1.452 | -0.011 | 1.46E-01 | 18725 | -0.103 | -0.231 | 0.025 | 0.065 | -1.576 | -0.012 | 1.15E-01 | 18410 |
| 121 | Thal_AV_L | 0.032 | -0.029 | 0.092 | 0.031 | 1.026 | 0.008 | 3.05E-01 | 18725 | 0.088 | 0.037 | 0.140 | 0.026 | 3.368 | 0.025 | 7.59E-04 | 18410 |
| 122 | Thal_AV_R | 0.108 | 0.022 | 0.195 | 0.044 | 2.453 | 0.018 | 1.42E-02 | 18725 | 0.120 | 0.046 | 0.193 | 0.037 | 3.199 | 0.024 | 1.38E-03 | 18410 |
| 123 | Thal_LP_L | -0.012 | -0.079 | 0.055 | 0.034 | -0.360 | -0.003 | 7.19E-01 | 18725 | 0.087 | 0.030 | 0.144 | 0.029 | 3.006 | 0.022 | 2.65E-03 | 18410 |
| 124 | Thal_LP_R | 0.040 | -0.048 | 0.127 | 0.045 | 0.886 | 0.006 | 3.76E-01 | 18725 | 0.110 | 0.035 | 0.184 | 0.038 | 2.896 | 0.021 | 3.78E-03 | 18410 |
| 125 | Thal_VA_L | 0.037 | -0.034 | 0.109 | 0.036 | 1.024 | 0.007 | 3.06E-01 | 18725 | 0.039 | -0.022 | 0.100 | 0.031 | 1.255 | 0.009 | 2.10E-01 | 18410 |
| 126 | Thal_VA_R | 0.097 | 0.010 | 0.185 | 0.045 | 2.173 | 0.016 | 2.98E-02 | 18725 | 0.060 | -0.015 | 0.134 | 0.038 | 1.570 | 0.012 | 1.16E-01 | 18410 |
| 127 | Thal_VL_L | -0.091 | -0.295 | 0.113 | 0.104 | -0.875 | -0.006 | 3.82E-01 | 18725 | 0.324 | 0.151 | 0.497 | 0.088 | 3.677 | 0.027 | 2.37E-04 | 18410 |
| 128 | Thal_VL_R | 0.114 | -0.157 | 0.384 | 0.138 | 0.823 | 0.006 | 4.11E-01 | 18725 | 0.520 | 0.290 | 0.750 | 0.117 | 4.438 | 0.033 | 9.15E-06 | 18410 |
| 129 | Thal_VPL_L | -0.078 | -0.321 | 0.164 | 0.124 | -0.632 | -0.005 | 5.27E-01 | 18725 | 0.278 | 0.072 | 0.484 | 0.105 | 2.647 | 0.020 | 8.12E-03 | 18410 |
| 130 | Thal_VPL_R | -0.176 | -0.431 | 0.078 | 0.130 | -1.357 | -0.010 | 1.75E-01 | 18725 | 0.252 | 0.035 | 0.468 | 0.111 | 2.277 | 0.017 | 2.28E-02 | 18410 |
| 131 | Thal_IL_L | -0.017 | -0.121 | 0.088 | 0.054 | -0.309 | -0.002 | 7.58E-01 | 18725 | 0.110 | 0.021 | 0.199 | 0.045 | 2.413 | 0.018 | 1.58E-02 | 18410 |
| 132 | Thal_IL_R | -0.005 | -0.124 | 0.114 | 0.061 | -0.080 | -0.001 | 9.36E-01 | 18725 | 0.126 | 0.024 | 0.227 | 0.052 | 2.434 | 0.018 | 1.49E-02 | 18410 |
| 133 | Thal_Re_L | 0.000 | 0.000 | 0.001 | 0.000 | 0.901 | 0.007 | 3.68E-01 | 18725 | 0.000 | 0.000 | 0.001 | 0.000 | 0.637 | 0.005 | 5.24E-01 | 18410 |
| 134 | Thal_Re_R | 0.002 | -0.001 | 0.006 | 0.002 | 1.296 | 0.009 | 1.95E-01 | 18725 | 0.004 | 0.001 | 0.007 | 0.002 | 2.374 | 0.017 | 1.76E-02 | 18410 |
| 135 | Thal_MDm_L | 0.203 | -0.575 | 0.980 | 0.396 | 0.511 | 0.004 | 6.09E-01 | 18725 | 1.155 | 0.495 | 1.815 | 0.337 | 3.431 | 0.025 | 6.04E-04 | 18410 |
| 136 | Thal_MDm_R | 0.135 | -0.768 | 1.038 | 0.461 | 0.292 | 0.002 | 7.70E-01 | 18725 | 1.400 | 0.633 | 2.167 | 0.391 | 3.579 | 0.026 | 3.46E-04 | 18410 |
| 137 | Thal_MDl_L | -0.026 | -0.160 | 0.109 | 0.069 | -0.375 | -0.003 | 7.07E-01 | 18725 | 0.233 | 0.118 | 0.347 | 0.058 | 3.989 | 0.029 | 6.67E-05 | 18410 |
| 138 | Thal_MDl_R | 0.015 | -0.138 | 0.168 | 0.078 | 0.193 | 0.001 | 8.47E-01 | 18725 | 0.252 | 0.122 | 0.382 | 0.066 | 3.806 | 0.028 | 1.42E-04 | 18410 |
| 139 | Thal_LGN_L | -0.002 | -0.022 | 0.018 | 0.010 | -0.192 | -0.001 | 8.48E-01 | 18725 | -0.002 | -0.019 | 0.016 | 0.009 | -0.182 | -0.001 | 8.56E-01 | 18410 |
| 140 | Thal_LGN_R | -0.024 | -0.068 | 0.019 | 0.022 | -1.093 | -0.008 | 2.74E-01 | 18725 | 0.020 | -0.017 | 0.057 | 0.019 | 1.061 | 0.008 | 2.89E-01 | 18410 |
| 141 | Thal_MGN_L | -0.019 | -0.055 | 0.018 | 0.019 | -1.011 | -0.007 | 3.12E-01 | 18725 | -0.018 | -0.049 | 0.013 | 0.016 | -1.147 | -0.008 | 2.51E-01 | 18410 |
| 142 | Thal_MGN_R | -0.026 | -0.065 | 0.014 | 0.020 | -1.263 | -0.009 | 2.07E-01 | 18725 | 0.000 | -0.034 | 0.033 | 0.017 | -0.010 | 0.000 | 9.92E-01 | 18410 |
| 143 | Thal_PuA_L | 0.001 | -0.042 | 0.044 | 0.022 | 0.037 | 0.000 | 9.70E-01 | 18725 | -0.015 | -0.052 | 0.021 | 0.019 | -0.811 | -0.006 | 4.17E-01 | 18410 |
| 144 | Thal_PuA_R | -0.047 | -0.137 | 0.042 | 0.046 | -1.034 | -0.008 | 3.01E-01 | 18725 | -0.003 | -0.079 | 0.073 | 0.039 | -0.084 | -0.001 | 9.33E-01 | 18410 |
| 145 | Thal_PuM_L | 0.280 | -0.575 | 1.135 | 0.436 | 0.643 | 0.005 | 5.21E-01 | 18725 | 0.385 | -0.342 | 1.111 | 0.371 | 1.037 | 0.008 | 3.00E-01 | 18410 |
| 146 | Thal_PuM_R | -0.189 | -1.254 | 0.877 | 0.544 | -0.347 | -0.003 | 7.28E-01 | 18725 | 0.356 | -0.549 | 1.262 | 0.462 | 0.771 | 0.006 | 4.41E-01 | 18410 |
| 147 | Thal_PuL_L | 0.004 | -0.152 | 0.160 | 0.080 | 0.052 | 0.000 | 9.58E-01 | 18725 | 0.130 | -0.003 | 0.263 | 0.068 | 1.922 | 0.014 | 5.46E-02 | 18410 |
| 148 | Thal_PuL_R | -0.090 | -0.338 | 0.158 | 0.127 | -0.712 | -0.005 | 4.76E-01 | 18725 | 0.154 | -0.057 | 0.365 | 0.108 | 1.428 | 0.011 | 1.53E-01 | 18410 |
| 149 | Thal_PuI_L | 0.007 | -0.036 | 0.049 | 0.022 | 0.317 | 0.002 | 7.51E-01 | 18725 | 0.036 | 0.000 | 0.073 | 0.018 | 1.984 | 0.015 | 4.73E-02 | 18410 |
| 150 | Thal_PuI_R | 0.002 | -0.055 | 0.058 | 0.029 | 0.056 | 0.000 | 9.56E-01 | 18725 | 0.039 | -0.009 | 0.087 | 0.024 | 1.593 | 0.012 | 1.11E-01 | 18410 |
| 151 | ACC_sub_L | -0.183 | -0.672 | 0.306 | 0.250 | -0.733 | -0.005 | 4.64E-01 | 18725 | -0.442 | -0.857 | -0.026 | 0.212 | -2.085 | -0.015 | 3.71E-02 | 18410 |
| 152 | ACC_sub_R | -0.257 | -0.714 | 0.200 | 0.233 | -1.101 | -0.008 | 2.71E-01 | 18725 | -0.284 | -0.673 | 0.104 | 0.198 | -1.436 | -0.011 | 1.51E-01 | 18410 |
| 153 | ACC_pre_L | -0.799 | -2.054 | 0.455 | 0.640 | -1.249 | -0.009 | 2.12E-01 | 18725 | -0.179 | -1.245 | 0.886 | 0.544 | -0.330 | -0.002 | 7.41E-01 | 18410 |
| 154 | ACC_pre_R | -0.377 | -1.586 | 0.832 | 0.617 | -0.611 | -0.004 | 5.41E-01 | 18725 | -0.130 | -1.156 | 0.896 | 0.524 | -0.248 | -0.002 | 8.04E-01 | 18410 |
| 155 | ACC_sup_L | -1.134 | -2.498 | 0.229 | 0.696 | -1.631 | -0.012 | 1.03E-01 | 18725 | 0.064 | -1.095 | 1.222 | 0.591 | 0.108 | 0.001 | 9.14E-01 | 18410 |
| 156 | ACC_sup_R | -0.522 | -1.935 | 0.890 | 0.721 | -0.725 | -0.005 | 4.69E-01 | 18725 | -0.491 | -1.690 | 0.709 | 0.612 | -0.802 | -0.006 | 4.23E-01 | 18410 |
| 157 | N_Acc_L | 0.281 | -0.129 | 0.692 | 0.209 | 1.343 | 0.010 | 1.79E-01 | 18725 | 0.248 | -0.100 | 0.597 | 0.178 | 1.395 | 0.010 | 1.63E-01 | 18410 |
| 158 | N_Acc_R | 0.011 | -0.344 | 0.366 | 0.181 | 0.063 | 0.000 | 9.50E-01 | 18725 | 0.187 | -0.114 | 0.489 | 0.154 | 1.218 | 0.009 | 2.23E-01 | 18410 |
| 159 | VTA_L | 0.006 | -0.009 | 0.020 | 0.007 | 0.783 | 0.006 | 4.33E-01 | 18725 | 0.001 | -0.011 | 0.013 | 0.006 | 0.133 | 0.001 | 8.94E-01 | 18410 |
| 160 | VTA_R | 0.000 | -0.013 | 0.013 | 0.007 | 0.004 | 0.000 | 9.97E-01 | 18725 | 0.004 | -0.007 | 0.015 | 0.006 | 0.730 | 0.005 | 4.66E-01 | 18410 |

| | | | | | | | | | | | | | | | | | |
|---|---|---|---|---|---|---|---|---|---|---|---|---|---|---|---|---|---|
| 161 | SN_pc_L | -0.005 | -0.028 | 0.019 | 0.012 | -0.386 | -0.003 | 6.99E-01 | 18725 | -0.001 | -0.020 | 0.019 | 0.010 | -0.056 | 0.000 | 9.56E-01 | 18410 |
| 162 | SN_pc_R | -0.024 | -0.054 | 0.007 | 0.016 | -1.536 | -0.011 | 1.25E-01 | 18725 | 0.001 | -0.025 | 0.027 | 0.013 | 0.080 | 0.001 | 9.36E-01 | 18410 |
| 163 | SN_pr_L | -0.016 | -0.048 | 0.015 | 0.016 | -1.024 | -0.007 | 3.06E-01 | 18725 | 0.021 | -0.006 | 0.047 | 0.014 | 1.514 | 0.011 | 1.30E-01 | 18410 |
| 164 | SN_pr_R | -0.022 | -0.057 | 0.014 | 0.018 | -1.200 | -0.009 | 2.30E-01 | 18725 | 0.005 | -0.025 | 0.035 | 0.015 | 0.348 | 0.003 | 7.28E-01 | 18410 |
| 165 | Red_N_L | 0.007 | -0.032 | 0.046 | 0.020 | 0.343 | 0.003 | 7.32E-01 | 18725 | -0.006 | -0.039 | 0.027 | 0.017 | -0.372 | -0.003 | 7.10E-01 | 18410 |
| 166 | Red_N_R | -0.010 | -0.052 | 0.032 | 0.022 | -0.455 | -0.003 | 6.49E-01 | 18725 | 0.008 | -0.028 | 0.044 | 0.018 | 0.451 | 0.003 | 6.52E-01 | 18410 |
| 167 | LC_L | 0.015 | 0.000 | 0.029 | 0.007 | 1.982 | 0.014 | 4.75E-02 | 18725 | -0.016 | -0.029 | -0.004 | 0.006 | -2.624 | -0.019 | 8.69E-03 | 18410 |
| 168 | LC_R | 0.006 | -0.010 | 0.021 | 0.008 | 0.700 | 0.005 | 4.84E-01 | 18725 | -0.023 | -0.036 | -0.010 | 0.007 | -3.360 | -0.025 | 7.81E-04 | 18410 |
| 169 | Raphe_D | 0.022 | -0.035 | 0.078 | 0.029 | 0.751 | 0.005 | 4.53E-01 | 18725 | 0.021 | -0.027 | 0.068 | 0.024 | 0.843 | 0.006 | 3.99E-01 | 18410 |
| 170 | Raphe_M | 0.020 | 0.003 | 0.037 | 0.009 | 2.295 | 0.017 | 2.18E-02 | 18725 | -0.015 | -0.030 | -0.001 | 0.007 | -2.079 | -0.015 | 3.77E-02 | 18410 |

**Non-oily fish intake**

| Number | AAL 3 region | beta | 95%CI_l | 95%CI_h | se | t | r | p | df.e |
|---|---|---|---|---|---|---|---|---|---|
| 0 | Total brain | 34.897 | -137.429 | 207.224 | 87.918 | 0.397 | 0.003 | 6.91E-01 | 18700 |
| 1 | Precentral_L | -2.070 | -8.137 | 3.997 | 3.095 | -0.669 | -0.005 | 5.04E-01 | 18700 |
| 2 | Precentral_R | -2.073 | -7.632 | 3.486 | 2.836 | -0.731 | -0.005 | 4.65E-01 | 18700 |
| 3 | Frontal_Sup_2_L | -5.168 | -11.573 | 1.238 | 3.268 | -1.581 | -0.012 | 1.14E-01 | 18700 |
| 4 | Frontal_Sup_2_R | -5.242 | -12.302 | 1.818 | 3.602 | -1.455 | -0.011 | 1.46E-01 | 18700 |
| 5 | Frontal_Mid_2_L | -0.330 | -7.059 | 6.400 | 3.433 | -0.096 | -0.001 | 9.24E-01 | 18700 |
| 6 | Frontal_Mid_2_R | -2.241 | -9.853 | 5.372 | 3.884 | -0.577 | -0.004 | 5.64E-01 | 18700 |
| 7 | Frontal_Inf_Oper_L | -2.555 | -4.542 | -0.567 | 1.014 | -2.520 | -0.018 | 1.18E-02 | 18700 |
| 8 | Frontal_Inf_Oper_R | -2.782 | -5.273 | -0.290 | 1.271 | -2.189 | -0.016 | 2.86E-02 | 18700 |
| 9 | Frontal_Inf_Tri_L | -0.143 | -4.365 | 4.079 | 2.154 | -0.066 | 0.000 | 9.47E-01 | 18700 |
| 10 | Frontal_Inf_Tri_R | -1.354 | -4.669 | 1.961 | 1.691 | -0.801 | -0.006 | 4.23E-01 | 18700 |
| 11 | Frontal_Inf_Orb_2_L | 0.062 | -1.200 | 1.324 | 0.644 | 0.097 | 0.001 | 9.23E-01 | 18700 |
| 12 | Frontal_Inf_Orb_2_R | -0.250 | -1.570 | 1.070 | 0.673 | -0.372 | -0.003 | 7.10E-01 | 18700 |
| 13 | Rolandic_Oper_L | 0.620 | -1.289 | 2.529 | 0.974 | 0.636 | 0.005 | 5.25E-01 | 18700 |
| 14 | Rolandic_Oper_R | 1.492 | -1.239 | 4.223 | 1.393 | 1.071 | 0.008 | 2.84E-01 | 18700 |
| 15 | Supp_Motor_Area_L | -2.978 | -7.513 | 1.558 | 2.314 | -1.287 | -0.009 | 1.98E-01 | 18700 |
| 16 | Supp_Motor_Area_R | -0.405 | -5.419 | 4.609 | 2.558 | -0.158 | -0.001 | 8.74E-01 | 18700 |
| 17 | Olfactory_L | 0.679 | 0.005 | 1.354 | 0.344 | 1.974 | 0.014 | 4.84E-02 | 18700 |
| 18 | Olfactory_R | 0.403 | -0.273 | 1.080 | 0.345 | 1.169 | 0.009 | 2.42E-01 | 18700 |
| 19 | Frontal_Sup_Medial_L | -4.328 | -8.581 | -0.076 | 2.170 | -1.995 | -0.015 | 4.61E-02 | 18700 |
| 20 | Frontal_Sup_Medial_R | -1.746 | -4.842 | 1.349 | 1.579 | -1.106 | -0.008 | 2.69E-01 | 18700 |
| 21 | Frontal_Med_Orb_L | -0.550 | -2.138 | 1.039 | 0.810 | -0.678 | -0.005 | 4.98E-01 | 18700 |
| 22 | Frontal_Med_Orb_R | -0.891 | -2.842 | 1.061 | 0.996 | -0.894 | -0.007 | 3.71E-01 | 18700 |
| 23 | Rectus_L | 0.424 | -1.220 | 2.068 | 0.839 | 0.506 | 0.004 | 6.13E-01 | 18700 |
| 24 | Rectus_R | 0.431 | -1.089 | 1.952 | 0.776 | 0.556 | 0.004 | 5.78E-01 | 18700 |
| 25 | OFCmed_L | -0.545 | -1.498 | 0.408 | 0.486 | -1.120 | -0.008 | 2.63E-01 | 18700 |
| 26 | OFCmed_R | -0.503 | -1.614 | 0.607 | 0.567 | -0.888 | -0.006 | 3.74E-01 | 18700 |
| 27 | OFCant_L | -0.634 | -1.736 | 0.468 | 0.562 | -1.127 | -0.008 | 2.60E-01 | 18700 |
| 28 | OFCant_R | -1.427 | -2.883 | 0.028 | 0.743 | -1.922 | -0.014 | 5.46E-02 | 18700 |
| 29 | OFCpost_L | -0.093 | -1.474 | 1.287 | 0.704 | -0.132 | -0.001 | 8.95E-01 | 18700 |
| 30 | OFCpost_R | -0.922 | -2.198 | 0.354 | 0.651 | -1.417 | -0.010 | 1.57E-01 | 18700 |
| 31 | OFClat_L | -0.112 | -0.578 | 0.354 | 0.238 | -0.471 | -0.003 | 6.37E-01 | 18700 |
| 32 | OFClat_R | -0.351 | -0.744 | 0.043 | 0.201 | -1.748 | -0.013 | 8.05E-02 | 18700 |
| 33 | Insula_L | 0.423 | -3.845 | 4.692 | 2.178 | 0.194 | 0.001 | 8.46E-01 | 18700 |
| 34 | Insula_R | -0.135 | -4.293 | 4.023 | 2.121 | -0.064 | 0.000 | 9.49E-01 | 18700 |
| 37 | Cingulate_Mid_L | -1.251 | -4.858 | 2.356 | 1.840 | -0.680 | -0.005 | 4.97E-01 | 18700 |

| | | | | | | | | | |
|---|---|---|---|---|---|---|---|---|---|
| 38 | Cingulate_Mid_R | -1.554 | -5.401 | 2.292 | 1.962 | -0.792 | -0.006 | 4.28E-01 | 18700 |
| 39 | Cingulate_Post_L | -0.280 | -1.383 | 0.822 | 0.562 | -0.498 | -0.004 | 6.18E-01 | 18700 |
| 40 | Cingulate_Post_R | 0.010 | -0.601 | 0.622 | 0.312 | 0.032 | 0.000 | 9.74E-01 | 18700 |
| 41 | Hippocampus_L | 0.550 | -1.233 | 2.332 | 0.909 | 0.604 | 0.004 | 5.46E-01 | 18700 |
| 42 | Hippocampus_R | 0.769 | -1.078 | 2.617 | 0.943 | 0.816 | 0.006 | 4.14E-01 | 18700 |
| 43 | ParaHippocampal_L | 0.587 | -0.996 | 2.170 | 0.808 | 0.727 | 0.005 | 4.67E-01 | 18700 |
| 44 | ParaHippocampal_R | -0.002 | -1.998 | 1.994 | 1.018 | -0.002 | 0.000 | 9.99E-01 | 18700 |
| 45 | Amygdala_L | -0.061 | -0.677 | 0.554 | 0.314 | -0.196 | -0.001 | 8.45E-01 | 18700 |
| 46 | Amygdala_R | -0.372 | -1.001 | 0.257 | 0.321 | -1.158 | -0.008 | 2.47E-01 | 18700 |
| 47 | Calcarine_L | 7.637 | 1.765 | 13.510 | 2.996 | 2.549 | 0.019 | 1.08E-02 | 18700 |
| 48 | Calcarine_R | 5.131 | 0.224 | 10.039 | 2.504 | 2.049 | 0.015 | 4.04E-02 | 18700 |
| 49 | Cuneus_L | 4.228 | 0.546 | 7.911 | 1.879 | 2.251 | 0.016 | 2.44E-02 | 18700 |
| 50 | Cuneus_R | 1.223 | -1.594 | 4.040 | 1.437 | 0.851 | 0.006 | 3.95E-01 | 18700 |
| 51 | Lingual_L | 5.347 | 1.817 | 8.876 | 1.801 | 2.969 | 0.022 | 2.99E-03 | 18700 |
| 52 | Lingual_R | 3.518 | -0.182 | 7.217 | 1.887 | 1.864 | 0.014 | 6.24E-02 | 18700 |
| 53 | Occipital_Sup_L | 0.411 | -2.017 | 2.838 | 1.238 | 0.332 | 0.002 | 7.40E-01 | 18700 |
| 54 | Occipital_Sup_R | -2.525 | -5.157 | 0.107 | 1.343 | -1.880 | -0.014 | 6.01E-02 | 18700 |
| 55 | Occipital_Mid_L | 3.212 | -2.476 | 8.901 | 2.902 | 1.107 | 0.008 | 2.68E-01 | 18700 |
| 56 | Occipital_Mid_R | -1.886 | -5.848 | 2.075 | 2.021 | -0.933 | -0.007 | 3.51E-01 | 18700 |
| 57 | Occipital_Inf_L | 2.871 | 0.982 | 4.759 | 0.963 | 2.980 | 0.022 | 2.89E-03 | 18700 |
| 58 | Occipital_Inf_R | 0.115 | -2.067 | 2.296 | 1.113 | 0.103 | 0.001 | 9.18E-01 | 18700 |
| 59 | Fusiform_L | 3.493 | -0.640 | 7.627 | 2.109 | 1.656 | 0.012 | 9.76E-02 | 18700 |
| 60 | Fusiform_R | 4.680 | 0.138 | 9.221 | 2.317 | 2.020 | 0.015 | 4.34E-02 | 18700 |
| 61 | Postcentral_L | -3.461 | -9.680 | 2.759 | 3.173 | -1.091 | -0.008 | 2.75E-01 | 18700 |
| 62 | Postcentral_R | -2.707 | -9.083 | 3.669 | 3.253 | -0.832 | -0.006 | 4.05E-01 | 18700 |
| 63 | Parietal_Sup_L | -3.760 | -7.344 | -0.177 | 1.828 | -2.057 | -0.015 | 3.97E-02 | 18700 |
| 64 | Parietal_Sup_R | -1.475 | -4.655 | 1.705 | 1.622 | -0.909 | -0.007 | 3.63E-01 | 18700 |
| 65 | Parietal_Inf_L | -1.948 | -6.639 | 2.744 | 2.393 | -0.814 | -0.006 | 4.16E-01 | 18700 |
| 66 | Parietal_Inf_R | -1.381 | -4.419 | 1.656 | 1.550 | -0.891 | -0.007 | 3.73E-01 | 18700 |
| 67 | SupraMarginal_L | -0.977 | -3.727 | 1.772 | 1.403 | -0.697 | -0.005 | 4.86E-01 | 18700 |
| 68 | SupraMarginal_R | -3.230 | -7.121 | 0.660 | 1.985 | -1.628 | -0.012 | 1.04E-01 | 18700 |
| 69 | Angular_L | 1.736 | -0.964 | 4.436 | 1.377 | 1.261 | 0.009 | 2.07E-01 | 18700 |
| 70 | Angular_R | 4.021 | 0.402 | 7.641 | 1.847 | 2.178 | 0.016 | 2.94E-02 | 18700 |
| 71 | Precuneus_L | -1.682 | -8.265 | 4.901 | 3.358 | -0.501 | -0.004 | 6.17E-01 | 18700 |
| 72 | Precuneus_R | -0.988 | -6.851 | 4.875 | 2.991 | -0.330 | -0.002 | 7.41E-01 | 18700 |
| 73 | Paracentral_Lobule_L | -1.362 | -4.374 | 1.651 | 1.537 | -0.886 | -0.006 | 3.76E-01 | 18700 |
| 74 | Paracentral_Lobule_R | -2.659 | -4.817 | -0.501 | 1.101 | -2.415 | -0.018 | 1.57E-02 | 18700 |
| 75 | Caudate_L | 0.935 | -1.474 | 3.343 | 1.229 | 0.761 | 0.006 | 4.47E-01 | 18700 |
| 76 | Caudate_R | 1.324 | -1.403 | 4.050 | 1.391 | 0.952 | 0.007 | 3.41E-01 | 18700 |
| 77 | Putamen_L | -1.385 | -5.042 | 2.271 | 1.866 | -0.743 | -0.005 | 4.58E-01 | 18700 |
| 78 | Putamen_R | -1.762 | -5.486 | 1.962 | 1.900 | -0.928 | -0.007 | 3.54E-01 | 18700 |
| 79 | Pallidum_L | -0.220 | -0.545 | 0.105 | 0.166 | -1.327 | -0.010 | 1.84E-01 | 18700 |
| 80 | Pallidum_R | -0.231 | -0.587 | 0.126 | 0.182 | -1.267 | -0.009 | 2.05E-01 | 18700 |
| 83 | Heschl_L | 0.106 | -0.553 | 0.766 | 0.336 | 0.316 | 0.002 | 7.52E-01 | 18700 |
| 84 | Heschl_R | 0.455 | -0.281 | 1.192 | 0.376 | 1.212 | 0.009 | 2.26E-01 | 18700 |
| 85 | Temporal_Sup_L | 0.104 | -4.656 | 4.864 | 2.428 | 0.043 | 0.000 | 9.66E-01 | 18700 |
| 86 | Temporal_Sup_R | 2.271 | -2.994 | 7.536 | 2.686 | 0.845 | 0.006 | 3.98E-01 | 18700 |
| 87 | Temporal_Pole_Sup_L | -0.499 | -2.753 | 1.755 | 1.150 | -0.434 | -0.003 | 6.64E-01 | 18700 |

| | | | | | | | | | |
|---|---|---|---|---|---|---|---|---|---|
| 88 | Temporal_Pole_Sup_R | -0.221 | -2.594 | 2.153 | 1.211 | -0.182 | -0.001 | 8.56E-01 | 18700 |
| 89 | Temporal_Mid_L | -0.770 | -8.700 | 7.159 | 4.045 | -0.190 | -0.001 | 8.49E-01 | 18700 |
| 90 | Temporal_Mid_R | -2.141 | -9.178 | 4.896 | 3.590 | -0.596 | -0.004 | 5.51E-01 | 18700 |
| 91 | Temporal_Pole_Mid_L | 0.304 | -1.590 | 2.198 | 0.966 | 0.314 | 0.002 | 7.53E-01 | 18700 |
| 92 | Temporal_Pole_Mid_R | -0.711 | -3.013 | 1.592 | 1.175 | -0.605 | -0.004 | 5.45E-01 | 18700 |
| 93 | Temporal_Inf_L | 1.893 | -3.557 | 7.342 | 2.780 | 0.681 | 0.005 | 4.96E-01 | 18700 |
| 94 | Temporal_Inf_R | 1.395 | -4.544 | 7.335 | 3.030 | 0.460 | 0.003 | 6.45E-01 | 18700 |
| 95 | Cerebellum_Crus1_L | 12.269 | 5.802 | 18.737 | 3.299 | 3.719 | 0.027 | 2.01E-04 | 18700 |
| 96 | Cerebellum_Crus1_R | 8.806 | 2.210 | 15.401 | 3.365 | 2.617 | 0.019 | 8.88E-03 | 18700 |
| 97 | Cerebellum_Crus2_L | 2.164 | -2.968 | 7.296 | 2.618 | 0.827 | 0.006 | 4.08E-01 | 18700 |
| 98 | Cerebellum_Crus2_R | 2.449 | -2.298 | 7.195 | 2.422 | 1.011 | 0.007 | 3.12E-01 | 18700 |
| 99 | Cerebellum_3_L | 0.360 | 0.015 | 0.704 | 0.176 | 2.046 | 0.015 | 4.08E-02 | 18700 |
| 100 | Cerebellum_3_R | 0.352 | -0.113 | 0.817 | 0.237 | 1.484 | 0.011 | 1.38E-01 | 18700 |
| 101 | Cerebellum_4_5_L | 2.984 | 0.198 | 5.770 | 1.421 | 2.099 | 0.015 | 3.58E-02 | 18700 |
| 102 | Cerebellum_4_5_R | 1.739 | -0.531 | 4.010 | 1.158 | 1.502 | 0.011 | 1.33E-01 | 18700 |
| 103 | Cerebellum_6_L | 6.843 | 2.029 | 11.656 | 2.456 | 2.786 | 0.020 | 5.33E-03 | 18700 |
| 104 | Cerebellum_6_R | 6.267 | 1.196 | 11.338 | 2.587 | 2.422 | 0.018 | 1.54E-02 | 18700 |
| 105 | Cerebellum_7b_L | 0.739 | -0.830 | 2.308 | 0.800 | 0.923 | 0.007 | 3.56E-01 | 18700 |
| 106 | Cerebellum_7b_R | 0.779 | -0.775 | 2.334 | 0.793 | 0.982 | 0.007 | 3.26E-01 | 18700 |
| 107 | Cerebellum_8_L | 3.703 | -1.275 | 8.682 | 2.540 | 1.458 | 0.011 | 1.45E-01 | 18700 |
| 108 | Cerebellum_8_R | 4.055 | -2.424 | 10.534 | 3.305 | 1.227 | 0.009 | 2.20E-01 | 18700 |
| 109 | Cerebellum_9_L | 1.434 | -1.383 | 4.251 | 1.437 | 0.998 | 0.007 | 3.18E-01 | 18700 |
| 110 | Cerebellum_9_R | 0.830 | -1.940 | 3.599 | 1.413 | 0.587 | 0.004 | 5.57E-01 | 18700 |
| 111 | Cerebellum_10_L | 0.167 | -0.033 | 0.367 | 0.102 | 1.637 | 0.012 | 1.02E-01 | 18700 |
| 112 | Cerebellum_10_R | 0.199 | -0.009 | 0.407 | 0.106 | 1.872 | 0.014 | 6.12E-02 | 18700 |
| 113 | Vermis_1_2 | 0.084 | -0.046 | 0.214 | 0.066 | 1.265 | 0.009 | 2.06E-01 | 18700 |
| 114 | Vermis_3 | 0.274 | -0.242 | 0.791 | 0.264 | 1.041 | 0.008 | 2.98E-01 | 18700 |
| 115 | Vermis_4_5 | 0.761 | -0.757 | 2.279 | 0.775 | 0.982 | 0.007 | 3.26E-01 | 18700 |
| 116 | Vermis_6 | 0.731 | -0.114 | 1.576 | 0.431 | 1.695 | 0.012 | 9.01E-02 | 18700 |
| 117 | Vermis_7 | 0.487 | 0.009 | 0.965 | 0.244 | 1.997 | 0.015 | 4.58E-02 | 18700 |
| 118 | Vermis_8 | 0.831 | 0.046 | 1.616 | 0.400 | 2.076 | 0.015 | 3.79E-02 | 18700 |
| 119 | Vermis_9 | 0.144 | -0.492 | 0.780 | 0.325 | 0.444 | 0.003 | 6.57E-01 | 18700 |
| 120 | Vermis_10 | 0.056 | -0.122 | 0.233 | 0.091 | 0.615 | 0.004 | 5.38E-01 | 18700 |
| 121 | Thal_AV_L | -0.011 | -0.082 | 0.060 | 0.036 | -0.304 | -0.002 | 7.61E-01 | 18700 |
| 122 | Thal_AV_R | -0.021 | -0.123 | 0.080 | 0.052 | -0.410 | -0.003 | 6.82E-01 | 18700 |
| 123 | Thal_LP_L | -0.049 | -0.128 | 0.030 | 0.040 | -1.212 | -0.009 | 2.25E-01 | 18700 |
| 124 | Thal_LP_R | -0.045 | -0.148 | 0.057 | 0.052 | -0.866 | -0.006 | 3.86E-01 | 18700 |
| 125 | Thal_VA_L | 0.025 | -0.059 | 0.109 | 0.043 | 0.589 | 0.004 | 5.56E-01 | 18700 |
| 126 | Thal_VA_R | 0.010 | -0.093 | 0.113 | 0.053 | 0.188 | 0.001 | 8.50E-01 | 18700 |
| 127 | Thal_VL_L | -0.161 | -0.401 | 0.078 | 0.122 | -1.322 | -0.010 | 1.86E-01 | 18700 |
| 128 | Thal_VL_R | -0.112 | -0.430 | 0.206 | 0.162 | -0.689 | -0.005 | 4.91E-01 | 18700 |
| 129 | Thal_VPL_L | -0.180 | -0.465 | 0.105 | 0.145 | -1.237 | -0.009 | 2.16E-01 | 18700 |
| 130 | Thal_VPL_R | -0.143 | -0.443 | 0.156 | 0.153 | -0.936 | -0.007 | 3.49E-01 | 18700 |
| 131 | Thal_IL_L | -0.046 | -0.169 | 0.078 | 0.063 | -0.725 | -0.005 | 4.68E-01 | 18700 |
| 132 | Thal_IL_R | -0.077 | -0.217 | 0.063 | 0.071 | -1.083 | -0.008 | 2.79E-01 | 18700 |
| 133 | Thal_Re_L | 0.000 | 0.000 | 0.001 | 0.000 | 0.232 | 0.002 | 8.17E-01 | 18700 |
| 134 | Thal_Re_R | -0.003 | -0.007 | 0.002 | 0.002 | -1.238 | -0.009 | 2.16E-01 | 18700 |
| 135 | Thal_MDm_L | -0.520 | -1.434 | 0.393 | 0.466 | -1.117 | -0.008 | 2.64E-01 | 18700 |

| | | | | | | | | | |
|---|---|---|---|---|---|---|---|---|---|
| 136 | Thal_MDm_R | -0.749 | -1.810 | 0.312 | 0.541 | -1.384 | -0.010 | 1.67E-01 | 18700 |
| 137 | Thal_MDl_L | -0.091 | -0.249 | 0.067 | 0.081 | -1.129 | -0.008 | 2.59E-01 | 18700 |
| 138 | Thal_MDl_R | -0.126 | -0.305 | 0.054 | 0.092 | -1.370 | -0.010 | 1.71E-01 | 18700 |
| 139 | Thal_LGN_L | -0.036 | -0.060 | -0.012 | 0.012 | -2.908 | -0.021 | 3.64E-03 | 18700 |
| 140 | Thal_LGN_R | -0.070 | -0.121 | -0.019 | 0.026 | -2.691 | -0.020 | 7.14E-03 | 18700 |
| 141 | Thal_MGN_L | -0.034 | -0.077 | 0.008 | 0.022 | -1.576 | -0.012 | 1.15E-01 | 18700 |
| 142 | Thal_MGN_R | -0.031 | -0.077 | 0.016 | 0.024 | -1.300 | -0.010 | 1.94E-01 | 18700 |
| 143 | Thal_PuA_L | -0.021 | -0.072 | 0.029 | 0.026 | -0.824 | -0.006 | 4.10E-01 | 18700 |
| 144 | Thal_PuA_R | -0.066 | -0.171 | 0.039 | 0.054 | -1.231 | -0.009 | 2.18E-01 | 18700 |
| 145 | Thal_PuM_L | -0.345 | -1.350 | 0.660 | 0.513 | -0.674 | -0.005 | 5.01E-01 | 18700 |
| 146 | Thal_PuM_R | -0.721 | -1.973 | 0.531 | 0.639 | -1.128 | -0.008 | 2.59E-01 | 18700 |
| 147 | Thal_PuL_L | -0.068 | -0.252 | 0.115 | 0.094 | -0.730 | -0.005 | 4.65E-01 | 18700 |
| 148 | Thal_PuL_R | -0.161 | -0.453 | 0.131 | 0.149 | -1.082 | -0.008 | 2.79E-01 | 18700 |
| 149 | Thal_PuI_L | -0.013 | -0.062 | 0.037 | 0.025 | -0.493 | -0.004 | 6.22E-01 | 18700 |
| 150 | Thal_PuI_R | -0.022 | -0.088 | 0.044 | 0.034 | -0.648 | -0.005 | 5.17E-01 | 18700 |
| 151 | ACC_sub_L | -0.047 | -0.622 | 0.528 | 0.293 | -0.159 | -0.001 | 8.74E-01 | 18700 |
| 152 | ACC_sub_R | 0.008 | -0.530 | 0.545 | 0.274 | 0.029 | 0.000 | 9.77E-01 | 18700 |
| 153 | ACC_pre_L | -0.116 | -1.590 | 1.358 | 0.752 | -0.154 | -0.001 | 8.77E-01 | 18700 |
| 154 | ACC_pre_R | -0.069 | -1.489 | 1.351 | 0.724 | -0.095 | -0.001 | 9.24E-01 | 18700 |
| 155 | ACC_sup_L | -1.368 | -2.970 | 0.234 | 0.817 | -1.674 | -0.012 | 9.42E-02 | 18700 |
| 156 | ACC_sup_R | -1.104 | -2.763 | 0.556 | 0.847 | -1.303 | -0.010 | 1.92E-01 | 18700 |
| 157 | N_Acc_L | 0.295 | -0.187 | 0.777 | 0.246 | 1.201 | 0.009 | 2.30E-01 | 18700 |
| 158 | N_Acc_R | 0.188 | -0.229 | 0.605 | 0.213 | 0.883 | 0.006 | 3.77E-01 | 18700 |
| 159 | VTA_L | 0.002 | -0.014 | 0.019 | 0.009 | 0.285 | 0.002 | 7.75E-01 | 18700 |
| 160 | VTA_R | -0.003 | -0.018 | 0.012 | 0.008 | -0.370 | -0.003 | 7.11E-01 | 18700 |
| 161 | SN_pc_L | 0.001 | -0.026 | 0.028 | 0.014 | 0.066 | 0.000 | 9.48E-01 | 18700 |
| 162 | SN_pc_R | -0.013 | -0.049 | 0.023 | 0.018 | -0.722 | -0.005 | 4.71E-01 | 18700 |
| 163 | SN_pr_L | -0.028 | -0.065 | 0.009 | 0.019 | -1.477 | -0.011 | 1.40E-01 | 18700 |
| 164 | SN_pr_R | -0.026 | -0.068 | 0.015 | 0.021 | -1.243 | -0.009 | 2.14E-01 | 18700 |
| 165 | Red_N_L | -0.007 | -0.052 | 0.039 | 0.023 | -0.280 | -0.002 | 7.79E-01 | 18700 |
| 166 | Red_N_R | -0.032 | -0.081 | 0.018 | 0.025 | -1.247 | -0.009 | 2.12E-01 | 18700 |
| 167 | LC_L | 0.015 | -0.002 | 0.032 | 0.009 | 1.731 | 0.013 | 8.34E-02 | 18700 |
| 168 | LC_R | 0.014 | -0.005 | 0.033 | 0.009 | 1.484 | 0.011 | 1.38E-01 | 18700 |
| 169 | Raphe_D | 0.012 | -0.054 | 0.078 | 0.034 | 0.361 | 0.003 | 7.18E-01 | 18700 |
| 170 | Raphe_M | 0.019 | -0.001 | 0.039 | 0.010 | 1.862 | 0.014 | 6.26E-02 | 18700 |

#### Table S3: Associations between grey matter volumes and diet phenotypes (baseline).

| Number | AAL 3 region | beta | 95%CI_l | 95%CI_h | se | t | r | p | df.e | beta | 95%CI_l | 95%CI_h | se | t | r | p | df.e |
|---|---|---|---|---|---|---|---|---|---|---|---|---|---|---|---|---|---|
| | | Coffee intake | | | | | | | | Cereal intake | | | | | | | |
| 0 | Total brain | -308.583 | -377.049 | -240.118 | 34.930 | -8.834 | -0.064 | 1.09E-18 | 18834 | 270.579 | 221.143 | 320.016 | 25.221 | 10.728 | 0.078 | 8.97E-27 | 18852 |
| 1 | Precentral_L | -6.832 | -9.243 | -4.421 | 1.230 | -5.554 | -0.040 | 2.83E-08 | 18834 | 4.739 | 2.996 | 6.481 | 0.889 | 5.331 | 0.039 | 9.88E-08 | 18852 |
| 2 | Precentral_R | -6.251 | -8.460 | -4.042 | 1.127 | -5.546 | -0.040 | 2.96E-08 | 18834 | 5.073 | 3.477 | 6.669 | 0.814 | 6.231 | 0.045 | 4.72E-10 | 18852 |
| 3 | Frontal_Sup_2_L | -9.211 | -11.754 | -6.669 | 1.297 | -7.101 | -0.052 | 1.29E-12 | 18834 | 6.777 | 4.939 | 8.616 | 0.938 | 7.226 | 0.053 | 5.16E-13 | 18852 |
| 4 | Frontal_Sup_2_R | -10.428 | -13.231 | -7.626 | 1.430 | -7.293 | -0.053 | 3.15E-13 | 18834 | 8.147 | 6.122 | 10.173 | 1.033 | 7.885 | 0.057 | 3.32E-15 | 18852 |
| 5 | Frontal_Mid_2_L | -8.593 | -11.264 | -5.921 | 1.363 | -6.304 | -0.046 | 2.96E-10 | 18834 | 5.395 | 3.463 | 7.327 | 0.986 | 5.473 | 0.040 | 4.47E-08 | 18852 |
| 6 | Frontal_Mid_2_R | -10.513 | -13.536 | -7.490 | 1.542 | -6.817 | -0.050 | 9.61E-12 | 18834 | 8.012 | 5.828 | 10.196 | 1.114 | 7.190 | 0.052 | 6.74E-13 | 18852 |
| 7 | Frontal_Inf_Oper_L | -1.972 | -2.762 | -1.182 | 0.403 | -4.893 | -0.036 | 9.99E-07 | 18834 | 1.672 | 1.101 | 2.243 | 0.291 | 5.742 | 0.042 | 9.48E-09 | 18852 |
| 8 | Frontal_Inf_Oper_R | -2.830 | -3.821 | -1.840 | 0.505 | -5.600 | -0.041 | 2.18E-08 | 18834 | 1.999 | 1.283 | 2.715 | 0.365 | 5.475 | 0.040 | 4.44E-08 | 18852 |
| 9 | Frontal_Inf_Tri_L | -4.246 | -5.923 | -2.569 | 0.855 | -4.963 | -0.036 | 6.99E-07 | 18834 | 3.774 | 2.562 | 4.985 | 0.618 | 6.105 | 0.044 | 1.05E-09 | 18852 |
| 10 | Frontal_Inf_Tri_R | -3.699 | -5.015 | -2.382 | 0.672 | -5.505 | -0.040 | 3.73E-08 | 18834 | 3.086 | 2.135 | 4.038 | 0.485 | 6.359 | 0.046 | 2.08E-10 | 18852 |
| 11 | Frontal_Inf_Orb_2_L | -1.062 | -1.563 | -0.560 | 0.256 | -4.150 | -0.030 | 3.33E-05 | 18834 | 1.161 | 0.798 | 1.523 | 0.185 | 6.281 | 0.046 | 3.44E-10 | 18852 |
| 12 | Frontal_Inf_Orb_2_R | -1.516 | -2.040 | -0.992 | 0.267 | -5.670 | -0.041 | 1.45E-08 | 18834 | 1.365 | 0.987 | 1.744 | 0.193 | 7.068 | 0.051 | 1.62E-12 | 18852 |
| 13 | Rolandic_Oper_L | -1.732 | -2.490 | -0.974 | 0.387 | -4.476 | -0.033 | 7.65E-06 | 18834 | 1.625 | 1.077 | 2.173 | 0.280 | 5.810 | 0.042 | 6.35E-09 | 18852 |
| 14 | Rolandic_Oper_R | -3.131 | -4.216 | -2.046 | 0.553 | -5.658 | -0.041 | 1.56E-08 | 18834 | 2.623 | 1.839 | 3.407 | 0.400 | 6.558 | 0.048 | 5.60E-11 | 18852 |
| 15 | Supp_Motor_Area_L | -4.703 | -6.507 | -2.898 | 0.920 | -5.109 | -0.037 | 3.27E-07 | 18834 | 2.588 | 1.283 | 3.892 | 0.665 | 3.889 | 0.028 | 1.01E-04 | 18852 |
| 16 | Supp_Motor_Area_R | -5.213 | -7.206 | -3.220 | 1.017 | -5.127 | -0.037 | 2.97E-07 | 18834 | 3.375 | 1.934 | 4.816 | 0.735 | 4.591 | 0.033 | 4.43E-06 | 18852 |
| 17 | Olfactory_L | -0.456 | -0.724 | -0.188 | 0.137 | -3.335 | -0.024 | 8.55E-04 | 18834 | 0.556 | 0.362 | 0.750 | 0.099 | 5.629 | 0.041 | 1.84E-08 | 18852 |
| 18 | Olfactory_R | -0.507 | -0.775 | -0.238 | 0.137 | -3.692 | -0.027 | 2.23E-04 | 18834 | 0.471 | 0.277 | 0.665 | 0.099 | 4.753 | 0.035 | 2.02E-06 | 18852 |
| 19 | Frontal_Sup_Medial_L | -5.950 | -7.640 | -4.261 | 0.862 | -6.903 | -0.050 | 5.26E-12 | 18834 | 5.446 | 4.227 | 6.666 | 0.622 | 8.753 | 0.064 | 2.26E-18 | 18852 |
| 20 | Frontal_Sup_Medial_R | -4.447 | -5.676 | -3.218 | 0.627 | -7.092 | -0.052 | 1.36E-12 | 18834 | 3.398 | 2.510 | 4.287 | 0.453 | 7.499 | 0.055 | 6.69E-14 | 18852 |
| 21 | Frontal_Med_Orb_L | -1.865 | -2.497 | -1.234 | 0.322 | -5.791 | -0.042 | 7.12E-09 | 18834 | 1.952 | 1.496 | 2.408 | 0.233 | 8.393 | 0.061 | 5.09E-17 | 18852 |
| 22 | Frontal_Med_Orb_R | -2.219 | -2.995 | -1.443 | 0.396 | -5.606 | -0.041 | 2.10E-08 | 18834 | 2.393 | 1.833 | 2.954 | 0.286 | 8.374 | 0.061 | 5.94E-17 | 18852 |
| 23 | Rectus_L | -1.631 | -2.285 | -0.978 | 0.334 | -4.891 | -0.036 | 1.01E-06 | 18834 | 1.860 | 1.388 | 2.333 | 0.241 | 7.724 | 0.056 | 1.18E-14 | 18852 |
| 24 | Rectus_R | -1.505 | -2.110 | -0.900 | 0.309 | -4.876 | -0.036 | 1.09E-06 | 18834 | 1.706 | 1.269 | 2.142 | 0.223 | 7.655 | 0.056 | 2.03E-14 | 18852 |
| 25 | OFCmed_L | -0.994 | -1.373 | -0.615 | 0.193 | -5.139 | -0.037 | 2.79E-07 | 18834 | 0.844 | 0.570 | 1.117 | 0.140 | 6.039 | 0.044 | 1.58E-09 | 18852 |
| 26 | OFCmed_R | -1.060 | -1.502 | -0.618 | 0.225 | -4.705 | -0.034 | 2.56E-06 | 18834 | 0.876 | 0.557 | 1.195 | 0.163 | 5.383 | 0.039 | 7.42E-08 | 18852 |
| 27 | OFCant_L | -0.985 | -1.423 | -0.546 | 0.224 | -4.404 | -0.032 | 1.07E-05 | 18834 | 0.696 | 0.379 | 1.013 | 0.162 | 4.309 | 0.031 | 1.65E-05 | 18852 |
| 28 | OFCant_R | -1.213 | -1.792 | -0.634 | 0.295 | -4.109 | -0.030 | 4.00E-05 | 18834 | 1.470 | 1.052 | 1.888 | 0.213 | 6.896 | 0.050 | 5.54E-12 | 18852 |
| 29 | OFCpost_L | -1.162 | -1.710 | -0.613 | 0.280 | -4.151 | -0.030 | 3.33E-05 | 18834 | 0.834 | 0.438 | 1.231 | 0.202 | 4.127 | 0.030 | 3.69E-05 | 18852 |
| 30 | OFCpost_R | -1.090 | -1.597 | -0.583 | 0.259 | -4.213 | -0.031 | 2.53E-05 | 18834 | 1.189 | 0.823 | 1.556 | 0.187 | 6.368 | 0.046 | 1.96E-10 | 18852 |
| 31 | OFClat_L | -0.378 | -0.563 | -0.193 | 0.094 | -4.004 | -0.029 | 6.26E-05 | 18834 | 0.377 | 0.243 | 0.511 | 0.068 | 5.525 | 0.040 | 3.34E-08 | 18852 |
| 32 | OFClat_R | -0.393 | -0.550 | -0.237 | 0.080 | -4.932 | -0.036 | 8.21E-07 | 18834 | 0.485 | 0.372 | 0.598 | 0.058 | 8.425 | 0.061 | 3.85E-17 | 18852 |
| 33 | Insula_L | -6.880 | -8.574 | -5.187 | 0.864 | -7.963 | -0.058 | 1.77E-15 | 18834 | 4.306 | 3.080 | 5.531 | 0.625 | 6.887 | 0.050 | 5.87E-12 | 18852 |
| 34 | Insula_R | -6.721 | -8.372 | -5.071 | 0.842 | -7.982 | -0.058 | 1.53E-15 | 18834 | 4.792 | 3.599 | 5.986 | 0.609 | 7.869 | 0.057 | 3.75E-15 | 18852 |
| 37 | Cingulate_Mid_L | -5.663 | -7.097 | -4.229 | 0.732 | -7.742 | -0.056 | 1.03E-14 | 18834 | 5.001 | 3.965 | 6.037 | 0.528 | 9.463 | 0.069 | 3.34E-21 | 18852 |
| 38 | Cingulate_Mid_R | -5.685 | -7.213 | -4.157 | 0.780 | -7.290 | -0.053 | 3.21E-13 | 18834 | 5.304 | 4.200 | 6.408 | 0.563 | 9.416 | 0.068 | 5.21E-21 | 18852 |
| 39 | Cingulate_Post_L | -0.587 | -1.025 | -0.149 | 0.224 | -2.625 | -0.019 | 8.67E-03 | 18834 | 0.663 | 0.347 | 0.980 | 0.161 | 4.106 | 0.030 | 4.04E-05 | 18852 |
| 40 | Cingulate_Post_R | -0.187 | -0.430 | 0.056 | 0.124 | -1.510 | -0.011 | 1.31E-01 | 18834 | 0.217 | 0.042 | 0.393 | 0.090 | 2.424 | 0.018 | 1.54E-02 | 18852 |
| 41 | Hippocampus_L | -0.710 | -1.419 | -0.001 | 0.362 | -1.963 | -0.014 | 4.96E-02 | 18834 | 1.518 | 1.006 | 2.030 | 0.261 | 5.812 | 0.042 | 6.26E-09 | 18852 |
| 42 | Hippocampus_R | -0.627 | -1.363 | 0.108 | 0.375 | -1.672 | -0.012 | 9.44E-02 | 18834 | 1.131 | 0.600 | 1.663 | 0.271 | 4.175 | 0.030 | 2.99E-05 | 18852 |
| 43 | ParaHippocampal_L | -0.932 | -1.561 | -0.302 | 0.321 | -2.899 | -0.021 | 3.74E-03 | 18834 | 1.475 | 1.020 | 1.930 | 0.232 | 6.360 | 0.046 | 2.07E-10 | 18852 |
| 44 | ParaHippocampal_R | -1.168 | -1.963 | -0.373 | 0.406 | -2.879 | -0.021 | 3.99E-03 | 18834 | 1.910 | 1.336 | 2.484 | 0.293 | 6.523 | 0.047 | 7.04E-11 | 18852 |
| 45 | Amygdala_L | -0.656 | -0.900 | -0.411 | 0.125 | -5.255 | -0.038 | 1.49E-07 | 18834 | 0.482 | 0.305 | 0.658 | 0.090 | 5.341 | 0.039 | 9.35E-08 | 18852 |
| 46 | Amygdala_R | -0.774 | -1.024 | -0.524 | 0.128 | -6.062 | -0.044 | 1.37E-09 | 18834 | 0.521 | 0.340 | 0.702 | 0.092 | 5.645 | 0.041 | 1.67E-08 | 18852 |
| 47 | Calcarine_L | -5.548 | -7.880 | -3.215 | 1.190 | -4.662 | -0.034 | 3.15E-06 | 18834 | 0.774 | -0.914 | 2.461 | 0.861 | 0.899 | 0.007 | 3.69E-01 | 18852 |

| | | | | | | | | | | | | | | | | | |
|---|---|---|---|---|---|---|---|---|---|---|---|---|---|---|---|---|---|
| 48 | Calcarine_R | -4.380 | -6.329 | -2.431 | 0.994 | -4.405 | -0.032 | 1.06E-05 | 18834 | 1.132 | -0.278 | 2.541 | 0.719 | 1.574 | 0.011 | 1.16E-01 | 18852 |
| 49 | Cuneus_L | -3.430 | -4.895 | -1.965 | 0.747 | -4.590 | -0.033 | 4.47E-06 | 18834 | 0.860 | -0.199 | 1.919 | 0.540 | 1.591 | 0.012 | 1.12E-01 | 18852 |
| 50 | Cuneus_R | -2.807 | -3.927 | -1.686 | 0.572 | -4.911 | -0.036 | 9.16E-07 | 18834 | 1.291 | 0.481 | 2.101 | 0.413 | 3.124 | 0.023 | 1.79E-03 | 18852 |
| 51 | Lingual_L | -2.995 | -4.399 | -1.590 | 0.717 | -4.179 | -0.030 | 2.94E-05 | 18834 | 0.843 | -0.172 | 1.859 | 0.518 | 1.628 | 0.012 | 1.04E-01 | 18852 |
| 52 | Lingual_R | -2.330 | -3.801 | -0.859 | 0.751 | -3.104 | -0.023 | 1.91E-03 | 18834 | 1.580 | 0.517 | 2.643 | 0.542 | 2.913 | 0.021 | 3.58E-03 | 18852 |
| 53 | Occipital_Sup_L | -2.293 | -3.258 | -1.328 | 0.492 | -4.656 | -0.034 | 3.24E-06 | 18834 | 1.395 | 0.697 | 2.092 | 0.356 | 3.918 | 0.029 | 8.96E-05 | 18852 |
| 54 | Occipital_Sup_R | -2.548 | -3.595 | -1.502 | 0.534 | -4.773 | -0.035 | 1.83E-06 | 18834 | 1.786 | 1.030 | 2.543 | 0.386 | 4.629 | 0.034 | 3.70E-06 | 18852 |
| 55 | Occipital_Mid_L | -3.547 | -5.810 | -1.285 | 1.154 | -3.073 | -0.022 | 2.12E-03 | 18834 | 1.127 | -0.509 | 2.762 | 0.835 | 1.350 | 0.010 | 1.77E-01 | 18852 |
| 56 | Occipital_Mid_R | -2.448 | -4.023 | -0.873 | 0.804 | -3.046 | -0.022 | 2.32E-03 | 18834 | 0.966 | -0.173 | 2.105 | 0.581 | 1.663 | 0.012 | 9.63E-02 | 18852 |
| 57 | Occipital_Inf_L | -1.021 | -1.772 | -0.271 | 0.383 | -2.668 | -0.019 | 7.64E-03 | 18834 | -0.173 | -0.716 | 0.369 | 0.277 | -0.626 | -0.005 | 5.31E-01 | 18852 |
| 58 | Occipital_Inf_R | -0.510 | -1.378 | 0.358 | 0.443 | -1.152 | -0.008 | 2.49E-01 | 18834 | -0.013 | -0.640 | 0.614 | 0.320 | -0.041 | 0.000 | 9.67E-01 | 18852 |
| 59 | Fusiform_L | -2.089 | -3.733 | -0.445 | 0.839 | -2.490 | -0.018 | 1.28E-02 | 18834 | 3.192 | 2.005 | 4.379 | 0.606 | 5.271 | 0.038 | 1.37E-07 | 18852 |
| 60 | Fusiform_R | -3.112 | -4.919 | -1.305 | 0.922 | -3.376 | -0.025 | 7.37E-04 | 18834 | 3.605 | 2.300 | 4.909 | 0.665 | 5.417 | 0.039 | 6.12E-08 | 18852 |
| 61 | Postcentral_L | -7.503 | -9.975 | -5.032 | 1.261 | -5.951 | -0.043 | 2.72E-09 | 18834 | 7.016 | 5.230 | 8.801 | 0.911 | 7.702 | 0.056 | 1.40E-14 | 18852 |
| 62 | Postcentral_R | -5.354 | -7.888 | -2.821 | 1.293 | -4.142 | -0.030 | 3.46E-05 | 18834 | 6.145 | 4.316 | 7.974 | 0.933 | 6.584 | 0.048 | 4.69E-11 | 18852 |
| 63 | Parietal_Sup_L | -3.289 | -4.714 | -1.865 | 0.727 | -4.525 | -0.033 | 6.08E-06 | 18834 | 0.631 | -0.398 | 1.661 | 0.525 | 1.202 | 0.009 | 2.29E-01 | 18852 |
| 64 | Parietal_Sup_R | -2.405 | -3.669 | -1.141 | 0.645 | -3.730 | -0.027 | 1.92E-04 | 18834 | 0.832 | -0.081 | 1.746 | 0.466 | 1.786 | 0.013 | 7.41E-02 | 18852 |
| 65 | Parietal_Inf_L | -6.005 | -7.868 | -4.141 | 0.951 | -6.316 | -0.046 | 2.74E-10 | 18834 | 4.355 | 3.008 | 5.703 | 0.687 | 6.337 | 0.046 | 2.39E-10 | 18852 |
| 66 | Parietal_Inf_R | -2.630 | -3.839 | -1.422 | 0.617 | -4.265 | -0.031 | 2.01E-05 | 18834 | 2.973 | 2.100 | 3.846 | 0.446 | 6.673 | 0.049 | 2.57E-11 | 18852 |
| 67 | SupraMarginal_L | -2.260 | -3.353 | -1.166 | 0.558 | -4.050 | -0.030 | 5.13E-05 | 18834 | 2.557 | 1.767 | 3.346 | 0.403 | 6.347 | 0.046 | 2.24E-10 | 18852 |
| 68 | SupraMarginal_R | -3.075 | -4.623 | -1.527 | 0.790 | -3.893 | -0.028 | 9.92E-05 | 18834 | 5.033 | 3.917 | 6.149 | 0.570 | 8.836 | 0.064 | 1.08E-18 | 18852 |
| 69 | Angular_L | -2.669 | -3.743 | -1.595 | 0.548 | -4.870 | -0.035 | 1.12E-06 | 18834 | 1.064 | 0.287 | 1.841 | 0.396 | 2.685 | 0.020 | 7.25E-03 | 18852 |
| 70 | Angular_R | -2.699 | -4.138 | -1.259 | 0.734 | -3.674 | -0.027 | 2.39E-04 | 18834 | 1.805 | 0.764 | 2.845 | 0.531 | 3.400 | 0.025 | 6.76E-04 | 18852 |
| 71 | Precuneus_L | -5.981 | -8.600 | -3.362 | 1.336 | -4.476 | -0.033 | 7.66E-06 | 18834 | 2.733 | 0.840 | 4.626 | 0.966 | 2.829 | 0.021 | 4.67E-03 | 18852 |
| 72 | Precuneus_R | -5.339 | -7.671 | -3.007 | 1.190 | -4.488 | -0.033 | 7.24E-06 | 18834 | 2.572 | 0.886 | 4.258 | 0.860 | 2.991 | 0.022 | 2.79E-03 | 18852 |
| 73 | Paracentral_Lobule_L | -2.217 | -3.416 | -1.018 | 0.612 | -3.625 | -0.026 | 2.90E-04 | 18834 | 0.514 | -0.352 | 1.381 | 0.442 | 1.163 | 0.008 | 2.45E-01 | 18852 |
| 74 | Paracentral_Lobule_R | -1.357 | -2.215 | -0.498 | 0.438 | -3.096 | -0.023 | 1.96E-03 | 18834 | 0.642 | 0.021 | 1.263 | 0.317 | 2.027 | 0.015 | 4.27E-02 | 18852 |
| 75 | Caudate_L | -1.177 | -2.134 | -0.220 | 0.488 | -2.410 | -0.018 | 1.59E-02 | 18834 | -0.270 | -0.963 | 0.422 | 0.353 | -0.766 | -0.006 | 4.44E-01 | 18852 |
| 76 | Caudate_R | -1.791 | -2.875 | -0.706 | 0.553 | -3.235 | -0.024 | 1.22E-03 | 18834 | -0.297 | -1.081 | 0.488 | 0.400 | -0.742 | -0.005 | 4.58E-01 | 18852 |
| 77 | Putamen_L | -4.237 | -5.690 | -2.784 | 0.741 | -5.715 | -0.042 | 1.11E-08 | 18834 | 3.374 | 2.324 | 4.424 | 0.536 | 6.298 | 0.046 | 3.09E-10 | 18852 |
| 78 | Putamen_R | -4.869 | -6.348 | -3.389 | 0.755 | -6.451 | -0.047 | 1.14E-10 | 18834 | 3.382 | 2.312 | 4.452 | 0.546 | 6.196 | 0.045 | 5.90E-10 | 18852 |
| 79 | Pallidum_L | -0.127 | -0.256 | 0.003 | 0.066 | -1.921 | -0.014 | 5.47E-02 | 18834 | 0.137 | 0.044 | 0.231 | 0.048 | 2.881 | 0.021 | 3.97E-03 | 18852 |
| 80 | Pallidum_R | -0.227 | -0.369 | -0.085 | 0.072 | -3.131 | -0.023 | 1.75E-03 | 18834 | 0.140 | 0.037 | 0.243 | 0.052 | 2.677 | 0.019 | 7.44E-03 | 18852 |
| 83 | Heschl_L | -0.842 | -1.104 | -0.580 | 0.134 | -6.301 | -0.046 | 3.03E-10 | 18834 | 0.541 | 0.352 | 0.731 | 0.097 | 5.604 | 0.041 | 2.13E-08 | 18852 |
| 84 | Heschl_R | -0.779 | -1.071 | -0.486 | 0.149 | -5.216 | -0.038 | 1.85E-07 | 18834 | 0.647 | 0.435 | 0.858 | 0.108 | 5.994 | 0.044 | 2.09E-09 | 18852 |
| 85 | Temporal_Sup_L | -6.502 | -8.391 | -4.613 | 0.964 | -6.745 | -0.049 | 1.57E-11 | 18834 | 4.234 | 2.868 | 5.601 | 0.697 | 6.074 | 0.044 | 1.27E-09 | 18852 |
| 86 | Temporal_Sup_R | -5.729 | -7.822 | -3.636 | 1.068 | -5.366 | -0.039 | 8.16E-08 | 18834 | 6.288 | 4.777 | 7.799 | 0.771 | 8.157 | 0.059 | 3.65E-16 | 18852 |
| 87 | Temporal_Pole_Sup_L | -2.412 | -3.308 | -1.516 | 0.457 | -5.277 | -0.038 | 1.33E-07 | 18834 | 1.995 | 1.348 | 2.643 | 0.330 | 6.042 | 0.044 | 1.55E-09 | 18852 |
| 88 | Temporal_Pole_Sup_R | -2.774 | -3.717 | -1.831 | 0.481 | -5.765 | -0.042 | 8.31E-09 | 18834 | 2.058 | 1.377 | 2.740 | 0.348 | 5.922 | 0.043 | 3.23E-09 | 18852 |
| 89 | Temporal_Mid_L | -6.771 | -9.925 | -3.617 | 1.609 | -4.208 | -0.031 | 2.59E-05 | 18834 | 7.475 | 5.197 | 9.753 | 1.162 | 6.432 | 0.047 | 1.29E-10 | 18852 |
| 90 | Temporal_Mid_R | -6.978 | -9.777 | -4.179 | 1.428 | -4.886 | -0.036 | 1.04E-06 | 18834 | 6.090 | 4.067 | 8.112 | 1.032 | 5.902 | 0.043 | 3.65E-09 | 18852 |
| 91 | Temporal_Pole_Mid_L | -1.058 | -1.812 | -0.305 | 0.385 | -2.752 | -0.020 | 5.93E-03 | 18834 | 1.261 | 0.716 | 1.805 | 0.278 | 4.540 | 0.033 | 5.67E-06 | 18852 |
| 92 | Temporal_Pole_Mid_R | -1.950 | -2.866 | -1.035 | 0.467 | -4.178 | -0.030 | 2.96E-05 | 18834 | 2.115 | 1.455 | 2.776 | 0.337 | 6.277 | 0.046 | 3.53E-10 | 18852 |
| 93 | Temporal_Inf_L | -2.577 | -4.745 | -0.409 | 1.106 | -2.330 | -0.017 | 1.98E-02 | 18834 | 2.246 | 0.679 | 3.812 | 0.799 | 2.809 | 0.020 | 4.97E-03 | 18852 |
| 94 | Temporal_Inf_R | -4.778 | -7.142 | -2.414 | 1.206 | -3.962 | -0.029 | 7.46E-05 | 18834 | 2.253 | 0.545 | 3.961 | 0.871 | 2.585 | 0.019 | 9.74E-03 | 18852 |
| 95 | Cerebellum_Crus1_L | -2.446 | -5.022 | 0.130 | 1.314 | -1.861 | -0.014 | 6.27E-02 | 18834 | 3.411 | 1.549 | 5.272 | 0.949 | 3.592 | 0.026 | 3.29E-04 | 18852 |
| 96 | Cerebellum_Crus1_R | -3.231 | -5.858 | -0.605 | 1.340 | -2.412 | -0.018 | 1.59E-02 | 18834 | 3.886 | 1.988 | 5.783 | 0.968 | 4.014 | 0.029 | 5.98E-05 | 18852 |
| 97 | Cerebellum_Crus2_L | -2.250 | -4.292 | -0.207 | 1.042 | -2.158 | -0.016 | 3.09E-02 | 18834 | 4.682 | 3.207 | 6.157 | 0.752 | 6.223 | 0.045 | 4.98E-10 | 18852 |
| 98 | Cerebellum_Crus2_R | -1.529 | -3.418 | 0.360 | 0.964 | -1.586 | -0.012 | 1.13E-01 | 18834 | 4.545 | 3.181 | 5.908 | 0.696 | 6.534 | 0.048 | 6.58E-11 | 18852 |

| | | | | | | | | | | | | | | | | | |
|---|---|---|---|---|---|---|---|---|---|---|---|---|---|---|---|---|---|
| 99 | Cerebellum_3_L | 0.092 | -0.046 | 0.229 | 0.070 | 1.309 | 0.010 | 1.91E-01 | 18834 | 0.202 | 0.103 | 0.301 | 0.050 | 3.998 | 0.029 | 6.42E-05 | 18852 |
| 100 | Cerebellum_3_R | -0.083 | -0.268 | 0.102 | 0.094 | -0.880 | -0.006 | 3.79E-01 | 18834 | 0.271 | 0.137 | 0.405 | 0.068 | 3.977 | 0.029 | 7.00E-05 | 18852 |
| 101 | Cerebellum_4_5_L | 0.628 | -0.481 | 1.737 | 0.566 | 1.110 | 0.008 | 2.67E-01 | 18834 | 1.050 | 0.249 | 1.851 | 0.409 | 2.570 | 0.019 | 1.02E-02 | 18852 |
| 102 | Cerebellum_4_5_R | -0.162 | -1.065 | 0.742 | 0.461 | -0.351 | -0.003 | 7.25E-01 | 18834 | 1.268 | 0.615 | 1.920 | 0.333 | 3.809 | 0.028 | 1.40E-04 | 18852 |
| 103 | Cerebellum_6_L | -0.678 | -2.596 | 1.240 | 0.979 | -0.693 | -0.005 | 4.88E-01 | 18834 | 2.402 | 1.016 | 3.787 | 0.707 | 3.397 | 0.025 | 6.81E-04 | 18852 |
| 104 | Cerebellum_6_R | -1.189 | -3.209 | 0.832 | 1.031 | -1.153 | -0.008 | 2.49E-01 | 18834 | 3.343 | 1.884 | 4.802 | 0.744 | 4.492 | 0.033 | 7.11E-06 | 18852 |
| 105 | Cerebellum_7b_L | -0.152 | -0.776 | 0.473 | 0.319 | -0.475 | -0.003 | 6.34E-01 | 18834 | 1.476 | 1.025 | 1.927 | 0.230 | 6.413 | 0.047 | 1.46E-10 | 18852 |
| 106 | Cerebellum_7b_R | -0.415 | -1.034 | 0.204 | 0.316 | -1.315 | -0.010 | 1.88E-01 | 18834 | 1.777 | 1.330 | 2.224 | 0.228 | 7.801 | 0.057 | 6.47E-15 | 18852 |
| 107 | Cerebellum_8_L | 1.237 | -0.747 | 3.221 | 1.012 | 1.222 | 0.009 | 2.22E-01 | 18834 | 3.627 | 2.194 | 5.060 | 0.731 | 4.961 | 0.036 | 7.08E-07 | 18852 |
| 108 | Cerebellum_8_R | 0.094 | -2.487 | 2.676 | 1.317 | 0.072 | 0.001 | 9.43E-01 | 18834 | 6.495 | 4.631 | 8.359 | 0.951 | 6.830 | 0.050 | 8.75E-12 | 18852 |
| 109 | Cerebellum_9_L | 1.835 | 0.715 | 2.956 | 0.572 | 3.210 | 0.023 | 1.33E-03 | 18834 | 0.879 | 0.069 | 1.690 | 0.413 | 2.126 | 0.015 | 3.35E-02 | 18852 |
| 110 | Cerebellum_9_R | 1.846 | 0.744 | 2.947 | 0.562 | 3.284 | 0.024 | 1.02E-03 | 18834 | 0.863 | 0.066 | 1.659 | 0.406 | 2.123 | 0.015 | 3.38E-02 | 18852 |
| 111 | Cerebellum_10_L | 0.038 | -0.042 | 0.117 | 0.041 | 0.935 | 0.007 | 3.50E-01 | 18834 | -0.012 | -0.069 | 0.046 | 0.029 | -0.399 | -0.003 | 6.90E-01 | 18852 |
| 112 | Cerebellum_10_R | 0.043 | -0.040 | 0.125 | 0.042 | 1.011 | 0.007 | 3.12E-01 | 18834 | 0.013 | -0.047 | 0.073 | 0.030 | 0.430 | 0.003 | 6.67E-01 | 18852 |
| 113 | Vermis_1_2 | -0.014 | -0.066 | 0.038 | 0.026 | -0.528 | -0.004 | 5.98E-01 | 18834 | 0.087 | 0.050 | 0.124 | 0.019 | 4.562 | 0.033 | 5.09E-06 | 18852 |
| 114 | Vermis_3 | 0.076 | -0.130 | 0.281 | 0.105 | 0.720 | 0.005 | 4.71E-01 | 18834 | 0.364 | 0.216 | 0.513 | 0.076 | 4.811 | 0.035 | 1.51E-06 | 18852 |
| 115 | Vermis_4_5 | 0.743 | 0.139 | 1.347 | 0.308 | 2.412 | 0.018 | 1.59E-02 | 18834 | 0.418 | -0.018 | 0.855 | 0.223 | 1.878 | 0.014 | 6.03E-02 | 18852 |
| 116 | Vermis_6 | 0.434 | 0.098 | 0.770 | 0.172 | 2.530 | 0.018 | 1.14E-02 | 18834 | 0.066 | -0.177 | 0.309 | 0.124 | 0.532 | 0.004 | 5.94E-01 | 18852 |
| 117 | Vermis_7 | 0.218 | 0.027 | 0.408 | 0.097 | 2.243 | 0.016 | 2.49E-02 | 18834 | 0.109 | -0.028 | 0.247 | 0.070 | 1.559 | 0.011 | 1.19E-01 | 18852 |
| 118 | Vermis_8 | 0.503 | 0.190 | 0.815 | 0.159 | 3.155 | 0.023 | 1.61E-03 | 18834 | 0.135 | -0.091 | 0.361 | 0.115 | 1.172 | 0.009 | 2.41E-01 | 18852 |
| 119 | Vermis_9 | 0.413 | 0.160 | 0.666 | 0.129 | 3.195 | 0.023 | 1.40E-03 | 18834 | 0.266 | 0.083 | 0.450 | 0.093 | 2.852 | 0.021 | 4.35E-03 | 18852 |
| 120 | Vermis_10 | 0.061 | -0.010 | 0.131 | 0.036 | 1.683 | 0.012 | 9.24E-02 | 18834 | 0.026 | -0.025 | 0.077 | 0.026 | 0.987 | 0.007 | 3.24E-01 | 18852 |
| 121 | Thal_AV_L | -0.055 | -0.083 | -0.027 | 0.014 | -3.818 | -0.028 | 1.35E-04 | 18834 | 0.032 | 0.011 | 0.052 | 0.010 | 3.046 | 0.022 | 2.32E-03 | 18852 |
| 122 | Thal_AV_R | -0.077 | -0.117 | -0.037 | 0.021 | -3.746 | -0.027 | 1.80E-04 | 18834 | 0.045 | 0.016 | 0.074 | 0.015 | 3.003 | 0.022 | 2.68E-03 | 18852 |
| 123 | Thal_LP_L | -0.080 | -0.111 | -0.049 | 0.016 | -5.008 | -0.036 | 5.56E-07 | 18834 | 0.074 | 0.051 | 0.096 | 0.012 | 6.385 | 0.046 | 1.75E-10 | 18852 |
| 124 | Thal_LP_R | -0.135 | -0.175 | -0.094 | 0.021 | -6.482 | -0.047 | 9.28E-11 | 18834 | 0.107 | 0.078 | 0.137 | 0.015 | 7.148 | 0.052 | 9.15E-13 | 18852 |
| 125 | Thal_VA_L | 0.001 | -0.032 | 0.035 | 0.017 | 0.085 | 0.001 | 9.32E-01 | 18834 | -0.026 | -0.051 | -0.002 | 0.012 | -2.152 | -0.016 | 3.14E-02 | 18852 |
| 126 | Thal_VA_R | 0.004 | -0.037 | 0.045 | 0.021 | 0.178 | 0.001 | 8.59E-01 | 18834 | -0.021 | -0.050 | 0.009 | 0.015 | -1.364 | -0.010 | 1.73E-01 | 18852 |
| 127 | Thal_VL_L | -0.111 | -0.206 | -0.015 | 0.049 | -2.278 | -0.017 | 2.27E-02 | 18834 | 0.111 | 0.042 | 0.180 | 0.035 | 3.169 | 0.023 | 1.53E-03 | 18852 |
| 128 | Thal_VL_R | -0.267 | -0.394 | -0.141 | 0.064 | -4.153 | -0.030 | 3.29E-05 | 18834 | 0.232 | 0.140 | 0.323 | 0.047 | 4.979 | 0.036 | 6.44E-07 | 18852 |
| 129 | Thal_VPL_L | -0.269 | -0.382 | -0.156 | 0.058 | -4.660 | -0.034 | 3.19E-06 | 18834 | 0.271 | 0.189 | 0.352 | 0.042 | 6.489 | 0.047 | 8.88E-11 | 18852 |
| 130 | Thal_VPL_R | -0.392 | -0.511 | -0.273 | 0.061 | -6.462 | -0.047 | 1.06E-10 | 18834 | 0.290 | 0.204 | 0.376 | 0.044 | 6.609 | 0.048 | 3.96E-11 | 18852 |
| 131 | Thal_IL_L | -0.195 | -0.244 | -0.147 | 0.025 | -7.830 | -0.057 | 5.15E-15 | 18834 | 0.146 | 0.111 | 0.182 | 0.018 | 8.109 | 0.059 | 5.40E-16 | 18852 |
| 132 | Thal_IL_R | -0.262 | -0.317 | -0.206 | 0.028 | -9.253 | -0.067 | 2.42E-20 | 18834 | 0.168 | 0.128 | 0.208 | 0.020 | 8.217 | 0.060 | 2.22E-16 | 18852 |
| 133 | Thal_Re_L | 0.000 | 0.000 | 0.000 | 0.000 | -0.216 | -0.002 | 8.29E-01 | 18834 | 0.000 | 0.000 | 0.000 | 0.000 | 2.924 | 0.021 | 3.46E-03 | 18852 |
| 134 | Thal_Re_R | -0.003 | -0.005 | -0.001 | 0.001 | -3.688 | -0.027 | 2.26E-04 | 18834 | 0.003 | 0.002 | 0.004 | 0.001 | 4.756 | 0.035 | 1.99E-06 | 18852 |
| 135 | Thal_MDm_L | -1.774 | -2.136 | -1.412 | 0.185 | -9.605 | -0.070 | 8.52E-22 | 18834 | 1.162 | 0.901 | 1.424 | 0.134 | 8.704 | 0.063 | 3.46E-18 | 18852 |
| 136 | Thal_MDm_R | -2.133 | -2.554 | -1.713 | 0.215 | -9.943 | -0.072 | 3.09E-23 | 18834 | 1.285 | 0.981 | 1.590 | 0.155 | 8.282 | 0.060 | 1.29E-16 | 18852 |
| 137 | Thal_MDl_L | -0.285 | -0.348 | -0.222 | 0.032 | -8.911 | -0.065 | 5.50E-19 | 18834 | 0.213 | 0.168 | 0.258 | 0.023 | 9.210 | 0.067 | 3.60E-20 | 18852 |
| 138 | Thal_MDl_R | -0.347 | -0.418 | -0.275 | 0.036 | -9.540 | -0.069 | 1.60E-21 | 18834 | 0.230 | 0.178 | 0.281 | 0.026 | 8.749 | 0.064 | 2.32E-18 | 18852 |
| 139 | Thal_LGN_L | -0.012 | -0.022 | -0.002 | 0.005 | -2.469 | -0.018 | 1.36E-02 | 18834 | 0.019 | 0.012 | 0.026 | 0.004 | 5.440 | 0.040 | 5.38E-08 | 18852 |
| 140 | Thal_LGN_R | -0.038 | -0.058 | -0.017 | 0.010 | -3.635 | -0.026 | 2.79E-04 | 18834 | 0.057 | 0.042 | 0.071 | 0.007 | 7.604 | 0.055 | 3.00E-14 | 18852 |
| 141 | Thal_MGN_L | -0.047 | -0.064 | -0.030 | 0.009 | -5.384 | -0.039 | 7.39E-08 | 18834 | 0.055 | 0.042 | 0.067 | 0.006 | 8.697 | 0.063 | 3.68E-18 | 18852 |
| 142 | Thal_MGN_R | -0.061 | -0.080 | -0.043 | 0.009 | -6.487 | -0.047 | 8.97E-11 | 18834 | 0.056 | 0.042 | 0.069 | 0.007 | 8.184 | 0.060 | 2.92E-16 | 18852 |
| 143 | Thal_PuA_L | -0.060 | -0.080 | -0.039 | 0.010 | -5.810 | -0.042 | 6.35E-09 | 18834 | 0.064 | 0.049 | 0.078 | 0.007 | 8.596 | 0.062 | 8.92E-18 | 18852 |
| 144 | Thal_PuA_R | -0.150 | -0.191 | -0.108 | 0.021 | -7.029 | -0.051 | 2.15E-12 | 18834 | 0.129 | 0.099 | 0.159 | 0.015 | 8.382 | 0.061 | 5.55E-17 | 18852 |
| 145 | Thal_PuM_L | -1.577 | -1.976 | -1.178 | 0.204 | -7.749 | -0.056 | 9.70E-15 | 18834 | 1.211 | 0.923 | 1.499 | 0.147 | 8.232 | 0.060 | 1.96E-16 | 18852 |
| 146 | Thal_PuM_R | -2.114 | -2.611 | -1.618 | 0.253 | -8.343 | -0.061 | 7.77E-17 | 18834 | 1.431 | 1.072 | 1.790 | 0.183 | 7.808 | 0.057 | 6.13E-15 | 18852 |
| 147 | Thal_PuL_L | -0.308 | -0.381 | -0.235 | 0.037 | -8.295 | -0.060 | 1.16E-16 | 18834 | 0.226 | 0.173 | 0.278 | 0.027 | 8.412 | 0.061 | 4.33E-17 | 18852 |

| | | | | | | | | | | | | | | | | | |
|---|---|---|---|---|---|---|---|---|---|---|---|---|---|---|---|---|---|
| 148 | Thal_PuL_R | -0.527 | -0.643 | -0.411 | 0.059 | -8.932 | -0.065 | 4.56E-19 | 18834 | 0.342 | 0.259 | 0.426 | 0.043 | 8.020 | 0.058 | 1.12E-15 | 18852 |
| 149 | Thal_Pul_L | -0.035 | -0.055 | -0.015 | 0.010 | -3.477 | -0.025 | 5.09E-04 | 18834 | 0.032 | 0.018 | 0.046 | 0.007 | 4.403 | 0.032 | 1.07E-05 | 18852 |
| 150 | Thal_Pul_R | -0.053 | -0.079 | -0.027 | 0.013 | -3.974 | -0.029 | 7.08E-05 | 18834 | 0.042 | 0.023 | 0.061 | 0.010 | 4.363 | 0.032 | 1.29E-05 | 18852 |
| 151 | ACC_sub_L | -0.334 | -0.562 | -0.105 | 0.117 | -2.863 | -0.021 | 4.20E-03 | 18834 | 0.305 | 0.140 | 0.471 | 0.084 | 3.624 | 0.026 | 2.91E-04 | 18852 |
| 152 | ACC_sub_R | -0.351 | -0.564 | -0.137 | 0.109 | -3.218 | -0.023 | 1.29E-03 | 18834 | 0.224 | 0.070 | 0.379 | 0.079 | 2.846 | 0.021 | 4.43E-03 | 18852 |
| 153 | ACC_pre_L | -1.548 | -2.134 | -0.963 | 0.299 | -5.182 | -0.038 | 2.22E-07 | 18834 | 1.572 | 1.148 | 1.995 | 0.216 | 7.280 | 0.053 | 3.46E-13 | 18852 |
| 154 | ACC_pre_R | -1.413 | -1.977 | -0.849 | 0.288 | -4.909 | -0.036 | 9.25E-07 | 18834 | 1.296 | 0.889 | 1.704 | 0.208 | 6.232 | 0.045 | 4.69E-10 | 18852 |
| 155 | ACC_sup_L | -1.919 | -2.555 | -1.283 | 0.324 | -5.915 | -0.043 | 3.37E-09 | 18834 | 1.494 | 1.034 | 1.954 | 0.235 | 6.367 | 0.046 | 1.98E-10 | 18852 |
| 156 | ACC_sup_R | -1.032 | -1.691 | -0.372 | 0.336 | -3.067 | -0.022 | 2.16E-03 | 18834 | 0.865 | 0.388 | 1.342 | 0.243 | 3.557 | 0.026 | 3.76E-04 | 18852 |
| 157 | N_Acc_L | -0.335 | -0.527 | -0.144 | 0.098 | -3.428 | -0.025 | 6.09E-04 | 18834 | 0.388 | 0.250 | 0.527 | 0.071 | 5.496 | 0.040 | 3.94E-08 | 18852 |
| 158 | N_Acc_R | -0.333 | -0.499 | -0.167 | 0.085 | -3.938 | -0.029 | 8.25E-05 | 18834 | 0.325 | 0.205 | 0.445 | 0.061 | 5.317 | 0.039 | 1.07E-07 | 18852 |
| 159 | VTA_L | -0.014 | -0.020 | -0.007 | 0.003 | -3.962 | -0.029 | 7.45E-05 | 18834 | 0.014 | 0.009 | 0.018 | 0.002 | 5.485 | 0.040 | 4.20E-08 | 18852 |
| 160 | VTA_R | -0.013 | -0.019 | -0.007 | 0.003 | -4.101 | -0.030 | 4.12E-05 | 18834 | 0.014 | 0.010 | 0.018 | 0.002 | 6.308 | 0.046 | 2.89E-10 | 18852 |
| 161 | SN_pc_L | -0.022 | -0.033 | -0.011 | 0.006 | -3.953 | -0.029 | 7.73E-05 | 18834 | 0.019 | 0.011 | 0.026 | 0.004 | 4.648 | 0.034 | 3.38E-06 | 18852 |
| 162 | SN_pc_R | -0.035 | -0.049 | -0.021 | 0.007 | -4.836 | -0.035 | 1.33E-06 | 18834 | 0.028 | 0.017 | 0.038 | 0.005 | 5.259 | 0.038 | 1.46E-07 | 18852 |
| 163 | SN_pr_L | -0.014 | -0.028 | 0.001 | 0.008 | -1.814 | -0.013 | 6.97E-02 | 18834 | 0.016 | 0.005 | 0.026 | 0.005 | 2.922 | 0.021 | 3.48E-03 | 18852 |
| 164 | SN_pr_R | -0.029 | -0.045 | -0.012 | 0.008 | -3.436 | -0.025 | 5.91E-04 | 18834 | 0.019 | 0.007 | 0.031 | 0.006 | 3.127 | 0.023 | 1.77E-03 | 18852 |
| 165 | Red_N_L | -0.030 | -0.049 | -0.012 | 0.009 | -3.294 | -0.024 | 9.89E-04 | 18834 | 0.037 | 0.024 | 0.050 | 0.007 | 5.533 | 0.040 | 3.19E-08 | 18852 |
| 166 | Red_N_R | -0.035 | -0.055 | -0.015 | 0.010 | -3.490 | -0.025 | 4.85E-04 | 18834 | 0.040 | 0.025 | 0.054 | 0.007 | 5.463 | 0.040 | 4.74E-08 | 18852 |
| 167 | LC_L | -0.002 | -0.009 | 0.004 | 0.003 | -0.720 | -0.005 | 4.71E-01 | 18834 | 0.003 | -0.002 | 0.008 | 0.002 | 1.301 | 0.009 | 1.93E-01 | 18852 |
| 168 | LC_R | -0.002 | -0.009 | 0.005 | 0.004 | -0.547 | -0.004 | 5.84E-01 | 18834 | 0.003 | -0.002 | 0.009 | 0.003 | 1.166 | 0.008 | 2.44E-01 | 18852 |
| 169 | Raphe_D | -0.068 | -0.094 | -0.041 | 0.013 | -5.040 | -0.037 | 4.70E-07 | 18834 | 0.080 | 0.061 | 0.099 | 0.010 | 8.234 | 0.060 | 1.93E-16 | 18852 |
| 170 | Raphe_M | -0.010 | -0.018 | -0.002 | 0.004 | -2.361 | -0.017 | 1.82E-02 | 18834 | 0.013 | 0.008 | 0.019 | 0.003 | 4.545 | 0.033 | 5.53E-06 | 18852 |

| | | Tea intake | | | | | | | | Water intake | | | | | | | |
|---|---|---|---|---|---|---|---|---|---|---|---|---|---|---|---|---|---|
| **Number** | **AAL 3 region** | **beta** | **95%CI_l** | **95%CI_h** | **se** | **t** | **r** | **p** | **df.e** | **beta** | **95%CI_l** | **95%CI_h** | **se** | **t** | **r** | **p** | **df.e** |
| 0 | Total brain | 15.805 | -35.995 | 67.604 | 26.427 | 0.598 | 0.004 | 5.50E-01 | 18775 | -117.015 | -184.464 | -49.565 | 34.411 | -3.400 | -0.025 | 6.74E-04 | 18750 |
| 1 | Precentral_L | 0.233 | -1.589 | 2.054 | 0.929 | 0.250 | 0.002 | 8.02E-01 | 18775 | -1.425 | -3.797 | 0.948 | 1.210 | -1.177 | -0.009 | 2.39E-01 | 18750 |
| 2 | Precentral_R | 1.040 | -0.630 | 2.709 | 0.852 | 1.221 | 0.009 | 2.22E-01 | 18775 | -2.556 | -4.730 | -0.382 | 1.109 | -2.305 | -0.017 | 2.12E-02 | 18750 |
| 3 | Frontal_Sup_2_L | 0.733 | -1.190 | 2.656 | 0.981 | 0.747 | 0.005 | 4.55E-01 | 18775 | -2.967 | -5.474 | -0.461 | 1.279 | -2.320 | -0.017 | 2.03E-02 | 18750 |
| 4 | Frontal_Sup_2_R | 1.629 | -0.492 | 3.749 | 1.082 | 1.505 | 0.011 | 1.32E-01 | 18775 | -2.388 | -5.151 | 0.375 | 1.410 | -1.694 | -0.012 | 9.03E-02 | 18750 |
| 5 | Frontal_Mid_2_L | -0.059 | -2.079 | 1.961 | 1.031 | -0.057 | 0.000 | 9.55E-01 | 18775 | -0.584 | -3.217 | 2.050 | 1.344 | -0.434 | -0.003 | 6.64E-01 | 18750 |
| 6 | Frontal_Mid_2_R | 0.682 | -1.604 | 2.968 | 1.166 | 0.585 | 0.004 | 5.59E-01 | 18775 | -1.831 | -4.810 | 1.149 | 1.520 | -1.204 | -0.009 | 2.28E-01 | 18750 |
| 7 | Frontal_Inf_Oper_L | 0.024 | -0.572 | 0.621 | 0.304 | 0.080 | 0.001 | 9.36E-01 | 18775 | -1.009 | -1.786 | -0.233 | 0.396 | -2.547 | -0.019 | 1.09E-02 | 18750 |
| 8 | Frontal_Inf_Oper_R | -0.184 | -0.933 | 0.564 | 0.382 | -0.483 | -0.004 | 6.29E-01 | 18775 | -1.334 | -2.309 | -0.359 | 0.498 | -2.682 | -0.020 | 7.33E-03 | 18750 |
| 9 | Frontal_Inf_Tri_L | 0.083 | -1.185 | 1.351 | 0.647 | 0.129 | 0.001 | 8.98E-01 | 18775 | -1.812 | -3.464 | -0.161 | 0.842 | -2.151 | -0.016 | 3.15E-02 | 18750 |
| 10 | Frontal_Inf_Tri_R | 0.152 | -0.844 | 1.147 | 0.508 | 0.298 | 0.002 | 7.65E-01 | 18775 | -1.357 | -2.654 | -0.061 | 0.661 | -2.052 | -0.015 | 4.01E-02 | 18750 |
| 11 | Frontal_Inf_Orb_2_L | -0.153 | -0.532 | 0.226 | 0.193 | -0.790 | -0.006 | 4.30E-01 | 18775 | -0.253 | -0.747 | 0.240 | 0.252 | -1.005 | -0.007 | 3.15E-01 | 18750 |
| 12 | Frontal_Inf_Orb_2_R | -0.099 | -0.496 | 0.297 | 0.202 | -0.492 | -0.004 | 6.23E-01 | 18775 | -0.414 | -0.930 | 0.103 | 0.263 | -1.571 | -0.011 | 1.16E-01 | 18750 |
| 13 | Rolandic_Oper_L | -0.126 | -0.699 | 0.448 | 0.293 | -0.429 | -0.003 | 6.68E-01 | 18775 | -0.607 | -1.353 | 0.140 | 0.381 | -1.593 | -0.012 | 1.11E-01 | 18750 |
| 14 | Rolandic_Oper_R | 1.061 | 0.241 | 1.881 | 0.418 | 2.535 | 0.018 | 1.13E-02 | 18775 | -1.129 | -2.197 | -0.061 | 0.545 | -2.073 | -0.015 | 3.82E-02 | 18750 |
| 15 | Supp_Motor_Area_L | -0.116 | -1.478 | 1.247 | 0.695 | -0.167 | -0.001 | 8.68E-01 | 18775 | -0.501 | -2.277 | 1.275 | 0.906 | -0.553 | -0.004 | 5.80E-01 | 18750 |
| 16 | Supp_Motor_Area_R | -0.337 | -1.843 | 1.168 | 0.768 | -0.439 | -0.003 | 6.61E-01 | 18775 | -0.027 | -1.990 | 1.936 | 1.002 | -0.027 | 0.000 | 9.79E-01 | 18750 |
| 17 | Olfactory_L | -0.095 | -0.297 | 0.108 | 0.103 | -0.917 | -0.007 | 3.59E-01 | 18775 | -0.273 | -0.537 | -0.009 | 0.135 | -2.030 | -0.015 | 4.24E-02 | 18750 |
| 18 | Olfactory_R | -0.136 | -0.339 | 0.067 | 0.104 | -1.312 | -0.010 | 1.90E-01 | 18775 | -0.367 | -0.631 | -0.102 | 0.135 | -2.717 | -0.020 | 6.59E-03 | 18750 |
| 19 | Frontal_Sup_Medial_L | -0.854 | -2.132 | 0.424 | 0.652 | -1.309 | -0.010 | 1.90E-01 | 18775 | -1.698 | -3.362 | -0.034 | 0.849 | -2.000 | -0.015 | 4.56E-02 | 18750 |
| 20 | Frontal_Sup_Medial_R | -0.529 | -1.459 | 0.401 | 0.475 | -1.115 | -0.008 | 2.65E-01 | 18775 | -1.305 | -2.516 | -0.094 | 0.618 | -2.112 | -0.015 | 3.47E-02 | 18750 |
| 21 | Frontal_Med_Orb_L | -0.093 | -0.570 | 0.385 | 0.244 | -0.381 | -0.003 | 7.03E-01 | 18775 | -0.434 | -1.055 | 0.188 | 0.317 | -1.367 | -0.010 | 1.72E-01 | 18750 |
| 22 | Frontal_Med_Orb_R | 0.092 | -0.495 | 0.679 | 0.299 | 0.308 | 0.002 | 7.58E-01 | 18775 | -0.747 | -1.511 | 0.017 | 0.390 | -1.916 | -0.014 | 5.54E-02 | 18750 |
| 23 | Rectus_L | 0.110 | -0.384 | 0.605 | 0.252 | 0.438 | 0.003 | 6.61E-01 | 18775 | -0.353 | -0.997 | 0.290 | 0.328 | -1.075 | -0.008 | 2.82E-01 | 18750 |

| | | | | | | | | | | | | | | | | | |
|---|---|---|---|---|---|---|---|---|---|---|---|---|---|---|---|---|---|
| 24 | Rectus_R | -0.041 | -0.498 | 0.416 | 0.233 | -0.177 | -0.001 | 8.60E-01 | 18775 | -0.461 | -1.056 | 0.135 | 0.304 | -1.516 | -0.011 | 1.29E-01 | 18750 |
| 25 | OFCmed_L | -0.039 | -0.326 | 0.247 | 0.146 | -0.269 | -0.002 | 7.88E-01 | 18775 | -0.323 | -0.696 | 0.051 | 0.190 | -1.694 | -0.012 | 9.02E-02 | 18750 |
| 26 | OFCmed_R | -0.159 | -0.492 | 0.175 | 0.170 | -0.933 | -0.007 | 3.51E-01 | 18775 | -0.581 | -1.015 | -0.146 | 0.222 | -2.617 | -0.019 | 8.87E-03 | 18750 |
| 27 | OFCant_L | 0.017 | -0.314 | 0.348 | 0.169 | 0.101 | 0.001 | 9.20E-01 | 18775 | -0.589 | -1.020 | -0.158 | 0.220 | -2.679 | -0.020 | 7.40E-03 | 18750 |
| 28 | OFCant_R | 0.138 | -0.299 | 0.575 | 0.223 | 0.619 | 0.005 | 5.36E-01 | 18775 | -1.077 | -1.647 | -0.508 | 0.291 | -3.709 | -0.027 | 2.09E-04 | 18750 |
| 29 | OFCpost_L | -0.015 | -0.429 | 0.399 | 0.211 | -0.070 | -0.001 | 9.44E-01 | 18775 | -0.686 | -1.226 | -0.146 | 0.275 | -2.490 | -0.018 | 1.28E-02 | 18750 |
| 30 | OFCpost_R | 0.039 | -0.345 | 0.422 | 0.196 | 0.198 | 0.001 | 8.43E-01 | 18775 | -0.699 | -1.198 | -0.199 | 0.255 | -2.743 | -0.020 | 6.09E-03 | 18750 |
| 31 | OFClat_L | -0.049 | -0.188 | 0.091 | 0.071 | -0.681 | -0.005 | 4.96E-01 | 18775 | -0.174 | -0.357 | 0.008 | 0.093 | -1.875 | -0.014 | 6.08E-02 | 18750 |
| 32 | OFClat_R | -0.053 | -0.171 | 0.066 | 0.060 | -0.872 | -0.006 | 3.83E-01 | 18775 | -0.254 | -0.408 | -0.100 | 0.078 | -3.236 | -0.024 | 1.22E-03 | 18750 |
| 33 | Insula_L | 0.128 | -1.155 | 1.410 | 0.654 | 0.195 | 0.001 | 8.45E-01 | 18775 | -2.109 | -3.779 | -0.440 | 0.852 | -2.477 | -0.018 | 1.33E-02 | 18750 |
| 34 | Insula_R | 0.283 | -0.966 | 1.533 | 0.638 | 0.444 | 0.003 | 6.57E-01 | 18775 | -2.700 | -4.326 | -1.073 | 0.830 | -3.253 | -0.024 | 1.14E-03 | 18750 |
| 37 | Cingulate_Mid_L | 0.612 | -0.473 | 1.697 | 0.554 | 1.105 | 0.008 | 2.69E-01 | 18775 | -1.431 | -2.843 | -0.019 | 0.720 | -1.986 | -0.015 | 4.70E-02 | 18750 |
| 38 | Cingulate_Mid_R | 0.488 | -0.669 | 1.644 | 0.590 | 0.826 | 0.006 | 4.09E-01 | 18775 | -1.285 | -2.790 | 0.220 | 0.768 | -1.673 | -0.012 | 9.43E-02 | 18750 |
| 39 | Cingulate_Post_L | 0.176 | -0.155 | 0.507 | 0.169 | 1.041 | 0.008 | 2.98E-01 | 18775 | -0.573 | -1.004 | -0.142 | 0.220 | -2.605 | -0.019 | 9.20E-03 | 18750 |
| 40 | Cingulate_Post_R | 0.091 | -0.093 | 0.274 | 0.094 | 0.966 | 0.007 | 3.34E-01 | 18775 | -0.265 | -0.504 | -0.026 | 0.122 | -2.169 | -0.016 | 3.01E-02 | 18750 |
| 41 | Hippocampus_L | 0.008 | -0.527 | 0.544 | 0.273 | 0.029 | 0.000 | 9.76E-01 | 18775 | -0.756 | -1.452 | -0.060 | 0.355 | -2.129 | -0.016 | 3.32E-02 | 18750 |
| 42 | Hippocampus_R | -0.141 | -0.697 | 0.414 | 0.283 | -0.498 | -0.004 | 6.19E-01 | 18775 | -0.400 | -1.123 | 0.323 | 0.369 | -1.085 | -0.008 | 2.78E-01 | 18750 |
| 43 | ParaHippocampal_L | 0.267 | -0.209 | 0.742 | 0.243 | 1.099 | 0.008 | 2.72E-01 | 18775 | -0.612 | -1.230 | 0.007 | 0.315 | -1.939 | -0.014 | 5.25E-02 | 18750 |
| 44 | ParaHippocampal_R | 0.147 | -0.454 | 0.747 | 0.306 | 0.479 | 0.003 | 6.32E-01 | 18775 | -0.417 | -1.199 | 0.364 | 0.399 | -1.047 | -0.008 | 2.95E-01 | 18750 |
| 45 | Amygdala_L | 0.093 | -0.091 | 0.278 | 0.094 | 0.990 | 0.007 | 3.22E-01 | 18775 | -0.370 | -0.610 | -0.130 | 0.123 | -3.019 | -0.022 | 2.54E-03 | 18750 |
| 46 | Amygdala_R | 0.087 | -0.102 | 0.276 | 0.097 | 0.904 | 0.007 | 3.66E-01 | 18775 | -0.361 | -0.607 | -0.115 | 0.126 | -2.878 | -0.021 | 4.00E-03 | 18750 |
| 47 | Calcarine_L | 0.271 | -1.493 | 2.034 | 0.900 | 0.301 | 0.002 | 7.63E-01 | 18775 | -0.268 | -2.563 | 2.028 | 1.171 | -0.229 | -0.002 | 8.19E-01 | 18750 |
| 48 | Calcarine_R | 0.532 | -0.941 | 2.006 | 0.752 | 0.708 | 0.005 | 4.79E-01 | 18775 | 0.087 | -1.832 | 2.005 | 0.979 | 0.088 | 0.001 | 9.30E-01 | 18750 |
| 49 | Cuneus_L | 0.007 | -1.100 | 1.114 | 0.565 | 0.012 | 0.000 | 9.91E-01 | 18775 | 0.549 | -0.892 | 1.991 | 0.736 | 0.747 | 0.005 | 4.55E-01 | 18750 |
| 50 | Cuneus_R | 0.301 | -0.546 | 1.147 | 0.432 | 0.696 | 0.005 | 4.86E-01 | 18775 | -1.190 | -2.292 | -0.087 | 0.563 | -2.115 | -0.015 | 3.45E-02 | 18750 |
| 51 | Lingual_L | 0.483 | -0.578 | 1.544 | 0.541 | 0.893 | 0.007 | 3.72E-01 | 18775 | -0.661 | -2.045 | 0.722 | 0.706 | -0.937 | -0.007 | 3.49E-01 | 18750 |
| 52 | Lingual_R | 0.537 | -0.575 | 1.649 | 0.567 | 0.947 | 0.007 | 3.44E-01 | 18775 | -1.064 | -2.511 | 0.384 | 0.738 | -1.441 | -0.011 | 1.50E-01 | 18750 |
| 53 | Occipital_Sup_L | 1.238 | 0.509 | 1.967 | 0.372 | 3.328 | 0.024 | 8.76E-04 | 18775 | -0.746 | -1.697 | 0.205 | 0.485 | -1.538 | -0.011 | 1.24E-01 | 18750 |
| 54 | Occipital_Sup_R | 0.886 | 0.096 | 1.677 | 0.403 | 2.199 | 0.016 | 2.79E-02 | 18775 | -1.468 | -2.497 | -0.438 | 0.525 | -2.794 | -0.020 | 5.20E-03 | 18750 |
| 55 | Occipital_Mid_L | 0.284 | -1.425 | 1.994 | 0.872 | 0.326 | 0.002 | 7.44E-01 | 18775 | -3.767 | -5.991 | -1.543 | 1.135 | -3.320 | -0.024 | 9.01E-04 | 18750 |
| 56 | Occipital_Mid_R | 0.798 | -0.392 | 1.987 | 0.607 | 1.315 | 0.010 | 1.89E-01 | 18775 | -1.958 | -3.507 | -0.410 | 0.790 | -2.478 | -0.018 | 1.32E-02 | 18750 |
| 57 | Occipital_Inf_L | -0.095 | -0.662 | 0.472 | 0.289 | -0.329 | -0.002 | 7.42E-01 | 18775 | -1.252 | -1.990 | -0.514 | 0.377 | -3.324 | -0.024 | 8.89E-04 | 18750 |
| 58 | Occipital_Inf_R | 0.370 | -0.286 | 1.025 | 0.334 | 1.105 | 0.008 | 2.69E-01 | 18775 | -1.452 | -2.306 | -0.599 | 0.436 | -3.334 | -0.024 | 8.56E-04 | 18750 |
| 59 | Fusiform_L | 0.617 | -0.625 | 1.858 | 0.633 | 0.973 | 0.007 | 3.30E-01 | 18775 | -1.745 | -3.363 | -0.127 | 0.825 | -2.114 | -0.015 | 3.45E-02 | 18750 |
| 60 | Fusiform_R | 0.033 | -1.332 | 1.397 | 0.696 | 0.047 | 0.000 | 9.63E-01 | 18775 | -1.120 | -2.897 | 0.657 | 0.907 | -1.235 | -0.009 | 2.17E-01 | 18750 |
| 61 | Postcentral_L | 1.303 | -0.566 | 3.171 | 0.953 | 1.366 | 0.010 | 1.72E-01 | 18775 | -0.896 | -3.330 | 1.537 | 1.242 | -0.722 | -0.005 | 4.70E-01 | 18750 |
| 62 | Postcentral_R | 0.815 | -1.100 | 2.730 | 0.977 | 0.834 | 0.006 | 4.04E-01 | 18775 | -0.096 | -2.589 | 2.398 | 1.272 | -0.075 | -0.001 | 9.40E-01 | 18750 |
| 63 | Parietal_Sup_L | -0.844 | -1.921 | 0.232 | 0.549 | -1.537 | -0.011 | 1.24E-01 | 18775 | -0.175 | -1.578 | 1.227 | 0.715 | -0.245 | -0.002 | 8.06E-01 | 18750 |
| 64 | Parietal_Sup_R | -0.937 | -1.893 | 0.018 | 0.487 | -1.923 | -0.014 | 5.45E-02 | 18775 | 0.123 | -1.121 | 1.367 | 0.635 | 0.194 | 0.001 | 8.47E-01 | 18750 |
| 65 | Parietal_Inf_L | 0.438 | -0.971 | 1.847 | 0.719 | 0.609 | 0.004 | 5.42E-01 | 18775 | -0.619 | -2.455 | 1.217 | 0.937 | -0.661 | -0.005 | 5.08E-01 | 18750 |
| 66 | Parietal_Inf_R | 0.167 | -0.747 | 1.081 | 0.466 | 0.359 | 0.003 | 7.20E-01 | 18775 | -0.750 | -1.940 | 0.440 | 0.607 | -1.235 | -0.009 | 2.17E-01 | 18750 |
| 67 | SupraMarginal_L | 0.366 | -0.461 | 1.192 | 0.422 | 0.868 | 0.006 | 3.86E-01 | 18775 | -0.669 | -1.745 | 0.406 | 0.549 | -1.220 | -0.009 | 2.23E-01 | 18750 |
| 68 | SupraMarginal_R | 0.439 | -0.731 | 1.608 | 0.597 | 0.735 | 0.005 | 4.62E-01 | 18775 | -1.470 | -2.993 | 0.053 | 0.777 | -1.892 | -0.014 | 5.85E-02 | 18750 |
| 69 | Angular_L | -0.178 | -0.990 | 0.634 | 0.414 | -0.430 | -0.003 | 6.67E-01 | 18775 | 0.228 | -0.829 | 1.286 | 0.540 | 0.423 | 0.003 | 6.72E-01 | 18750 |
| 70 | Angular_R | -0.269 | -1.357 | 0.820 | 0.555 | -0.484 | -0.004 | 6.28E-01 | 18775 | -0.483 | -1.899 | 0.934 | 0.723 | -0.668 | -0.005 | 5.04E-01 | 18750 |
| 71 | Precuneus_L | -0.730 | -2.709 | 1.250 | 1.010 | -0.722 | -0.005 | 4.70E-01 | 18775 | -1.073 | -3.650 | 1.505 | 1.315 | -0.816 | -0.006 | 4.15E-01 | 18750 |
| 72 | Precuneus_R | -0.301 | -2.064 | 1.462 | 0.899 | -0.335 | -0.002 | 7.38E-01 | 18775 | -2.743 | -5.038 | -0.448 | 1.171 | -2.342 | -0.017 | 1.92E-02 | 18750 |
| 73 | Paracentral_Lobule_L | -0.493 | -1.399 | 0.412 | 0.462 | -1.068 | -0.008 | 2.85E-01 | 18775 | 0.542 | -0.638 | 1.721 | 0.602 | 0.900 | 0.007 | 3.68E-01 | 18750 |
| 74 | Paracentral_Lobule_R | -0.366 | -1.015 | 0.283 | 0.331 | -1.105 | -0.008 | 2.69E-01 | 18775 | 0.194 | -0.651 | 1.040 | 0.431 | 0.451 | 0.003 | 6.52E-01 | 18750 |

| | | | | | | | | | | | | | | | | | |
|---|---|---|---|---|---|---|---|---|---|---|---|---|---|---|---|---|---|
| 75 | Caudate_L | 0.091 | -0.633 | 0.815 | 0.370 | 0.246 | 0.002 | 8.05E-01 | 18775 | 0.629 | -0.314 | 1.571 | 0.481 | 1.308 | 0.010 | 1.91E-01 | 18750 |
| 76 | Caudate_R | -0.316 | -1.136 | 0.504 | 0.418 | -0.755 | -0.006 | 4.51E-01 | 18775 | 0.800 | -0.267 | 1.868 | 0.545 | 1.469 | 0.011 | 1.42E-01 | 18750 |
| 77 | Putamen_L | -1.112 | -2.210 | -0.014 | 0.560 | -1.985 | -0.014 | 4.72E-02 | 18775 | -1.173 | -2.603 | 0.258 | 0.730 | -1.607 | -0.012 | 1.08E-01 | 18750 |
| 78 | Putamen_R | -1.208 | -2.326 | -0.089 | 0.571 | -2.116 | -0.015 | 3.44E-02 | 18775 | -0.871 | -2.328 | 0.586 | 0.744 | -1.171 | -0.009 | 2.42E-01 | 18750 |
| 79 | Pallidum_L | -0.130 | -0.228 | -0.033 | 0.050 | -2.615 | -0.019 | 8.92E-03 | 18775 | -0.063 | -0.190 | 0.064 | 0.065 | -0.970 | -0.007 | 3.32E-01 | 18750 |
| 80 | Pallidum_R | -0.157 | -0.264 | -0.050 | 0.055 | -2.874 | -0.021 | 4.05E-03 | 18775 | -0.074 | -0.214 | 0.065 | 0.071 | -1.046 | -0.008 | 2.96E-01 | 18750 |
| 83 | Heschl_L | 0.118 | -0.080 | 0.316 | 0.101 | 1.166 | 0.009 | 2.44E-01 | 18775 | -0.286 | -0.544 | -0.029 | 0.132 | -2.177 | -0.016 | 2.95E-02 | 18750 |
| 84 | Heschl_R | 0.388 | 0.166 | 0.609 | 0.113 | 3.436 | 0.025 | 5.91E-04 | 18775 | -0.435 | -0.723 | -0.147 | 0.147 | -2.963 | -0.022 | 3.05E-03 | 18750 |
| 85 | Temporal_Sup_L | 1.408 | -0.021 | 2.837 | 0.729 | 1.931 | 0.014 | 5.35E-02 | 18775 | -2.660 | -4.521 | -0.799 | 0.949 | -2.801 | -0.020 | 5.09E-03 | 18750 |
| 86 | Temporal_Sup_R | 1.214 | -0.368 | 2.796 | 0.807 | 1.504 | 0.011 | 1.33E-01 | 18775 | -4.677 | -6.736 | -2.618 | 1.050 | -4.453 | -0.033 | 8.52E-06 | 18750 |
| 87 | Temporal_Pole_Sup_L | 0.244 | -0.434 | 0.921 | 0.346 | 0.705 | 0.005 | 4.81E-01 | 18775 | -1.355 | -2.236 | -0.474 | 0.449 | -3.015 | -0.022 | 2.58E-03 | 18750 |
| 88 | Temporal_Pole_Sup_R | 0.685 | -0.028 | 1.398 | 0.364 | 1.883 | 0.014 | 5.97E-02 | 18775 | -1.500 | -2.427 | -0.573 | 0.473 | -3.171 | -0.023 | 1.52E-03 | 18750 |
| 89 | Temporal_Mid_L | 0.718 | -1.666 | 3.101 | 1.216 | 0.590 | 0.004 | 5.55E-01 | 18775 | -5.447 | -8.549 | -2.344 | 1.583 | -3.441 | -0.025 | 5.81E-04 | 18750 |
| 90 | Temporal_Mid_R | 0.282 | -1.834 | 2.397 | 1.079 | 0.261 | 0.002 | 7.94E-01 | 18775 | -4.736 | -7.491 | -1.982 | 1.405 | -3.370 | -0.025 | 7.52E-04 | 18750 |
| 91 | Temporal_Pole_Mid_L | 0.558 | -0.012 | 1.127 | 0.290 | 1.920 | 0.014 | 5.48E-02 | 18775 | -0.451 | -1.191 | 0.290 | 0.378 | -1.194 | -0.009 | 2.33E-01 | 18750 |
| 92 | Temporal_Pole_Mid_R | 1.011 | 0.320 | 1.702 | 0.353 | 2.869 | 0.021 | 4.13E-03 | 18775 | -1.306 | -2.206 | -0.407 | 0.459 | -2.846 | -0.021 | 4.43E-03 | 18750 |
| 93 | Temporal_Inf_L | -0.572 | -2.210 | 1.066 | 0.836 | -0.684 | -0.005 | 4.94E-01 | 18775 | -3.034 | -5.166 | -0.901 | 1.088 | -2.788 | -0.020 | 5.31E-03 | 18750 |
| 94 | Temporal_Inf_R | -0.157 | -1.942 | 1.629 | 0.911 | -0.172 | -0.001 | 8.63E-01 | 18775 | -2.699 | -5.025 | -0.373 | 1.187 | -2.275 | -0.017 | 2.29E-02 | 18750 |
| 95 | Cerebellum_Crus1_L | -0.508 | -2.455 | 1.438 | 0.993 | -0.512 | -0.004 | 6.09E-01 | 18775 | 0.233 | -2.302 | 2.768 | 1.293 | 0.180 | 0.001 | 8.57E-01 | 18750 |
| 96 | Cerebellum_Crus1_R | -0.428 | -2.412 | 1.556 | 1.012 | -0.423 | -0.003 | 6.72E-01 | 18775 | -0.808 | -3.393 | 1.776 | 1.318 | -0.613 | -0.004 | 5.40E-01 | 18750 |
| 97 | Cerebellum_Crus2_L | 0.667 | -0.875 | 2.209 | 0.787 | 0.848 | 0.006 | 3.97E-01 | 18775 | -1.152 | -3.162 | 0.857 | 1.025 | -1.124 | -0.008 | 2.61E-01 | 18750 |
| 98 | Cerebellum_Crus2_R | 0.173 | -1.253 | 1.599 | 0.727 | 0.238 | 0.002 | 8.12E-01 | 18775 | -2.060 | -3.918 | -0.203 | 0.948 | -2.174 | -0.016 | 2.97E-02 | 18750 |
| 99 | Cerebellum_3_L | -0.114 | -0.217 | -0.010 | 0.053 | -2.159 | -0.016 | 3.09E-02 | 18775 | 0.000 | -0.135 | 0.135 | 0.069 | 0.000 | 0.000 | 1.00E+00 | 18750 |
| 100 | Cerebellum_3_R | -0.064 | -0.203 | 0.076 | 0.071 | -0.895 | -0.007 | 3.71E-01 | 18775 | -0.029 | -0.211 | 0.153 | 0.093 | -0.307 | -0.002 | 7.59E-01 | 18750 |
| 101 | Cerebellum_4_5_L | 0.339 | -0.498 | 1.177 | 0.427 | 0.795 | 0.006 | 4.27E-01 | 18775 | -0.102 | -1.192 | 0.989 | 0.556 | -0.183 | -0.001 | 8.55E-01 | 18750 |
| 102 | Cerebellum_4_5_R | 0.211 | -0.471 | 0.893 | 0.348 | 0.606 | 0.004 | 5.45E-01 | 18775 | 0.201 | -0.688 | 1.090 | 0.453 | 0.443 | 0.003 | 6.58E-01 | 18750 |
| 103 | Cerebellum_6_L | 0.385 | -1.063 | 1.834 | 0.739 | 0.522 | 0.004 | 6.02E-01 | 18775 | -0.369 | -2.255 | 1.517 | 0.962 | -0.383 | -0.003 | 7.02E-01 | 18750 |
| 104 | Cerebellum_6_R | 0.094 | -1.431 | 1.620 | 0.778 | 0.121 | 0.001 | 9.03E-01 | 18775 | -0.556 | -2.543 | 1.431 | 1.014 | -0.548 | -0.004 | 5.83E-01 | 18750 |
| 105 | Cerebellum_7b_L | 0.025 | -0.447 | 0.497 | 0.241 | 0.104 | 0.001 | 9.17E-01 | 18775 | -0.576 | -1.190 | 0.039 | 0.313 | -1.837 | -0.013 | 6.63E-02 | 18750 |
| 106 | Cerebellum_7b_R | 0.192 | -0.275 | 0.659 | 0.238 | 0.805 | 0.006 | 4.21E-01 | 18775 | -0.546 | -1.155 | 0.062 | 0.311 | -1.759 | -0.013 | 7.86E-02 | 18750 |
| 107 | Cerebellum_8_L | -0.167 | -1.665 | 1.331 | 0.764 | -0.218 | -0.002 | 8.28E-01 | 18775 | -1.709 | -3.661 | 0.242 | 0.996 | -1.717 | -0.013 | 8.61E-02 | 18750 |
| 108 | Cerebellum_8_R | -0.641 | -2.590 | 1.308 | 0.994 | -0.645 | -0.005 | 5.19E-01 | 18775 | -3.083 | -5.623 | -0.544 | 1.296 | -2.380 | -0.017 | 1.73E-02 | 18750 |
| 109 | Cerebellum_9_L | -0.172 | -1.019 | 0.675 | 0.432 | -0.398 | -0.003 | 6.91E-01 | 18775 | -0.841 | -1.944 | 0.262 | 0.563 | -1.494 | -0.011 | 1.35E-01 | 18750 |
| 110 | Cerebellum_9_R | -0.341 | -1.174 | 0.491 | 0.425 | -0.804 | -0.006 | 4.21E-01 | 18775 | -1.087 | -2.172 | -0.002 | 0.553 | -1.964 | -0.014 | 4.95E-02 | 18750 |
| 111 | Cerebellum_10_L | -0.005 | -0.065 | 0.055 | 0.031 | -0.152 | -0.001 | 8.79E-01 | 18775 | 0.009 | -0.070 | 0.087 | 0.040 | 0.217 | 0.002 | 8.28E-01 | 18750 |
| 112 | Cerebellum_10_R | -0.059 | -0.122 | 0.003 | 0.032 | -1.860 | -0.014 | 6.29E-02 | 18775 | 0.051 | -0.031 | 0.132 | 0.041 | 1.223 | 0.009 | 2.21E-01 | 18750 |
| 113 | Vermis_1_2 | -0.062 | -0.101 | -0.023 | 0.020 | -3.102 | -0.023 | 1.93E-03 | 18775 | -0.010 | -0.061 | 0.041 | 0.026 | -0.384 | -0.003 | 7.01E-01 | 18750 |
| 114 | Vermis_3 | -0.196 | -0.351 | -0.041 | 0.079 | -2.475 | -0.018 | 1.33E-02 | 18775 | -0.076 | -0.278 | 0.126 | 0.103 | -0.739 | -0.005 | 4.60E-01 | 18750 |
| 115 | Vermis_4_5 | -0.195 | -0.651 | 0.261 | 0.233 | -0.838 | -0.006 | 4.02E-01 | 18775 | 0.107 | -0.487 | 0.701 | 0.303 | 0.352 | 0.003 | 7.24E-01 | 18750 |
| 116 | Vermis_6 | -0.175 | -0.429 | 0.079 | 0.130 | -1.348 | -0.010 | 1.78E-01 | 18775 | 0.106 | -0.225 | 0.437 | 0.169 | 0.628 | 0.005 | 5.30E-01 | 18750 |
| 117 | Vermis_7 | -0.119 | -0.263 | 0.025 | 0.073 | -1.623 | -0.012 | 1.05E-01 | 18775 | 0.053 | -0.134 | 0.241 | 0.096 | 0.560 | 0.004 | 5.75E-01 | 18750 |
| 118 | Vermis_8 | -0.236 | -0.472 | 0.000 | 0.120 | -1.960 | -0.014 | 5.00E-02 | 18775 | -0.118 | -0.426 | 0.189 | 0.157 | -0.754 | -0.006 | 4.51E-01 | 18750 |
| 119 | Vermis_9 | -0.051 | -0.242 | 0.141 | 0.098 | -0.519 | -0.004 | 6.04E-01 | 18775 | -0.135 | -0.385 | 0.114 | 0.127 | -1.062 | -0.008 | 2.88E-01 | 18750 |
| 120 | Vermis_10 | -0.016 | -0.069 | 0.037 | 0.027 | -0.585 | -0.004 | 5.59E-01 | 18775 | -0.015 | -0.085 | 0.054 | 0.035 | -0.432 | -0.003 | 6.66E-01 | 18750 |
| 121 | Thal_AV_L | 0.014 | -0.007 | 0.036 | 0.011 | 1.317 | 0.010 | 1.88E-01 | 18775 | -0.021 | -0.049 | 0.007 | 0.014 | -1.487 | -0.011 | 1.37E-01 | 18750 |
| 122 | Thal_AV_R | 0.021 | -0.009 | 0.052 | 0.016 | 1.354 | 0.010 | 1.76E-01 | 18775 | -0.030 | -0.070 | 0.010 | 0.020 | -1.475 | -0.011 | 1.40E-01 | 18750 |
| 123 | Thal_LP_L | 0.019 | -0.004 | 0.043 | 0.012 | 1.589 | 0.012 | 1.12E-01 | 18775 | -0.038 | -0.069 | -0.008 | 0.016 | -2.442 | -0.018 | 1.46E-02 | 18750 |
| 124 | Thal_LP_R | 0.050 | 0.019 | 0.081 | 0.016 | 3.166 | 0.023 | 1.55E-03 | 18775 | -0.068 | -0.108 | -0.028 | 0.020 | -3.327 | -0.024 | 8.81E-04 | 18750 |
| 125 | Thal_VA_L | -0.013 | -0.038 | 0.013 | 0.013 | -0.985 | -0.007 | 3.24E-01 | 18775 | 0.020 | -0.013 | 0.053 | 0.017 | 1.176 | 0.009 | 2.40E-01 | 18750 |

| | | | | | | | | | | | | | | | | | |
|---|---|---|---|---|---|---|---|---|---|---|---|---|---|---|---|---|---|
| 126 | Thal_VA_R | -0.022 | -0.053 | 0.009 | 0.016 | -1.372 | -0.010 | 1.70E-01 | 18775 | -0.005 | -0.045 | 0.035 | 0.021 | -0.243 | -0.002 | 8.08E-01 | 18750 |
| 127 | Thal_VL_L | 0.070 | -0.002 | 0.142 | 0.037 | 1.902 | 0.014 | 5.72E-02 | 18775 | -0.180 | -0.274 | -0.087 | 0.048 | -3.781 | -0.028 | 1.57E-04 | 18750 |
| 128 | Thal_VL_R | 0.118 | 0.022 | 0.213 | 0.049 | 2.416 | 0.018 | 1.57E-02 | 18775 | -0.275 | -0.399 | -0.151 | 0.063 | -4.348 | -0.032 | 1.38E-05 | 18750 |
| 129 | Thal_VPL_L | 0.157 | 0.072 | 0.243 | 0.044 | 3.605 | 0.026 | 3.13E-04 | 18775 | -0.251 | -0.363 | -0.140 | 0.057 | -4.427 | -0.032 | 9.62E-06 | 18750 |
| 130 | Thal_VPL_R | 0.194 | 0.104 | 0.283 | 0.046 | 4.223 | 0.031 | 2.43E-05 | 18775 | -0.252 | -0.369 | -0.135 | 0.060 | -4.225 | -0.031 | 2.41E-05 | 18750 |
| 131 | Thal_IL_L | 0.067 | 0.030 | 0.104 | 0.019 | 3.548 | 0.026 | 3.89E-04 | 18775 | -0.100 | -0.148 | -0.051 | 0.025 | -4.053 | -0.030 | 5.08E-05 | 18750 |
| 132 | Thal_IL_R | 0.090 | 0.048 | 0.132 | 0.021 | 4.191 | 0.031 | 2.80E-05 | 18775 | -0.123 | -0.178 | -0.069 | 0.028 | -4.424 | -0.032 | 9.73E-06 | 18750 |
| 133 | Thal_Re_L | 0.000 | 0.000 | 0.000 | 0.000 | -2.609 | -0.019 | 9.08E-03 | 18775 | 0.000 | 0.000 | 0.000 | 0.000 | -0.083 | -0.001 | 9.34E-01 | 18750 |
| 134 | Thal_Re_R | 0.000 | -0.002 | 0.001 | 0.001 | -0.697 | -0.005 | 4.86E-01 | 18775 | -0.002 | -0.004 | -0.001 | 0.001 | -2.571 | -0.019 | 1.01E-02 | 18750 |
| 135 | Thal_MDm_L | 0.534 | 0.260 | 0.808 | 0.140 | 3.817 | 0.028 | 1.36E-04 | 18775 | -0.761 | -1.118 | -0.404 | 0.182 | -4.180 | -0.031 | 2.93E-05 | 18750 |
| 136 | Thal_MDm_R | 0.692 | 0.374 | 1.011 | 0.162 | 4.262 | 0.031 | 2.03E-05 | 18775 | -0.867 | -1.281 | -0.452 | 0.211 | -4.098 | -0.030 | 4.18E-05 | 18750 |
| 137 | Thal_MDl_L | 0.095 | 0.048 | 0.143 | 0.024 | 3.946 | 0.029 | 7.96E-05 | 18775 | -0.148 | -0.210 | -0.086 | 0.032 | -4.702 | -0.034 | 2.60E-06 | 18750 |
| 138 | Thal_MDl_R | 0.123 | 0.069 | 0.177 | 0.028 | 4.486 | 0.033 | 7.30E-06 | 18775 | -0.167 | -0.237 | -0.097 | 0.036 | -4.669 | -0.034 | 3.05E-06 | 18750 |
| 139 | Thal_LGN_L | -0.003 | -0.011 | 0.004 | 0.004 | -0.952 | -0.007 | 3.41E-01 | 18775 | -0.011 | -0.020 | -0.002 | 0.005 | -2.307 | -0.017 | 2.11E-02 | 18750 |
| 140 | Thal_LGN_R | -0.008 | -0.024 | 0.007 | 0.008 | -1.058 | -0.008 | 2.90E-01 | 18775 | -0.048 | -0.067 | -0.028 | 0.010 | -4.665 | -0.034 | 3.11E-06 | 18750 |
| 141 | Thal_MGN_L | 0.006 | -0.007 | 0.019 | 0.007 | 0.952 | 0.007 | 3.41E-01 | 18775 | -0.037 | -0.053 | -0.020 | 0.009 | -4.284 | -0.031 | 1.84E-05 | 18750 |
| 142 | Thal_MGN_R | 0.010 | -0.004 | 0.024 | 0.007 | 1.419 | 0.010 | 1.56E-01 | 18775 | -0.044 | -0.062 | -0.026 | 0.009 | -4.750 | -0.035 | 2.05E-06 | 18750 |
| 143 | Thal_PuA_L | 0.011 | -0.004 | 0.027 | 0.008 | 1.469 | 0.011 | 1.42E-01 | 18775 | -0.043 | -0.063 | -0.023 | 0.010 | -4.255 | -0.031 | 2.10E-05 | 18750 |
| 144 | Thal_PuA_R | 0.039 | 0.007 | 0.070 | 0.016 | 2.408 | 0.018 | 1.60E-02 | 18775 | -0.101 | -0.142 | -0.060 | 0.021 | -4.810 | -0.035 | 1.52E-06 | 18750 |
| 145 | Thal_PuM_L | 0.452 | 0.150 | 0.754 | 0.154 | 2.936 | 0.021 | 3.33E-03 | 18775 | -0.863 | -1.256 | -0.470 | 0.200 | -4.304 | -0.031 | 1.69E-05 | 18750 |
| 146 | Thal_PuM_R | 0.724 | 0.348 | 1.100 | 0.192 | 3.772 | 0.028 | 1.62E-04 | 18775 | -1.055 | -1.545 | -0.566 | 0.250 | -4.224 | -0.031 | 2.41E-05 | 18750 |
| 147 | Thal_PuL_L | 0.104 | 0.049 | 0.159 | 0.028 | 3.702 | 0.027 | 2.15E-04 | 18775 | -0.154 | -0.226 | -0.083 | 0.037 | -4.218 | -0.031 | 2.48E-05 | 18750 |
| 148 | Thal_PuL_R | 0.190 | 0.103 | 0.278 | 0.045 | 4.264 | 0.031 | 2.02E-05 | 18775 | -0.236 | -0.350 | -0.122 | 0.058 | -4.062 | -0.030 | 4.88E-05 | 18750 |
| 149 | Thal_PuI_L | 0.011 | -0.004 | 0.026 | 0.008 | 1.404 | 0.010 | 1.60E-01 | 18775 | -0.019 | -0.039 | 0.000 | 0.010 | -1.930 | -0.014 | 5.36E-02 | 18750 |
| 150 | Thal_PuI_R | 0.018 | -0.002 | 0.037 | 0.010 | 1.742 | 0.013 | 8.15E-02 | 18775 | -0.030 | -0.055 | -0.004 | 0.013 | -2.250 | -0.016 | 2.45E-02 | 18750 |
| 151 | ACC_sub_L | 0.036 | -0.136 | 0.209 | 0.088 | 0.412 | 0.003 | 6.80E-01 | 18775 | -0.211 | -0.435 | 0.014 | 0.115 | -1.835 | -0.013 | 6.65E-02 | 18750 |
| 152 | ACC_sub_R | 0.052 | -0.110 | 0.213 | 0.082 | 0.626 | 0.005 | 5.31E-01 | 18775 | -0.205 | -0.415 | 0.005 | 0.107 | -1.910 | -0.014 | 5.61E-02 | 18750 |
| 153 | ACC_pre_L | -0.142 | -0.585 | 0.301 | 0.226 | -0.630 | -0.005 | 5.29E-01 | 18775 | -0.646 | -1.222 | -0.070 | 0.294 | -2.197 | -0.016 | 2.80E-02 | 18750 |
| 154 | ACC_pre_R | -0.282 | -0.709 | 0.144 | 0.217 | -1.298 | -0.009 | 1.94E-01 | 18775 | -0.713 | -1.268 | -0.158 | 0.283 | -2.519 | -0.018 | 1.18E-02 | 18750 |
| 155 | ACC_sup_L | 0.182 | -0.299 | 0.663 | 0.245 | 0.743 | 0.005 | 4.58E-01 | 18775 | -0.943 | -1.569 | -0.316 | 0.319 | -2.951 | -0.022 | 3.17E-03 | 18750 |
| 156 | ACC_sup_R | 0.018 | -0.480 | 0.516 | 0.254 | 0.070 | 0.001 | 9.44E-01 | 18775 | -0.708 | -1.356 | -0.059 | 0.331 | -2.138 | -0.016 | 3.25E-02 | 18750 |
| 157 | N_Acc_L | -0.128 | -0.272 | 0.017 | 0.074 | -1.727 | -0.013 | 8.43E-02 | 18775 | -0.089 | -0.277 | 0.100 | 0.096 | -0.920 | -0.007 | 3.57E-01 | 18750 |
| 158 | N_Acc_R | -0.160 | -0.285 | -0.034 | 0.064 | -2.499 | -0.018 | 1.25E-02 | 18775 | -0.105 | -0.268 | 0.058 | 0.083 | -1.258 | -0.009 | 2.08E-01 | 18750 |
| 159 | VTA_L | 0.008 | 0.003 | 0.013 | 0.003 | 2.930 | 0.021 | 3.39E-03 | 18775 | -0.009 | -0.016 | -0.002 | 0.003 | -2.694 | -0.020 | 7.07E-03 | 18750 |
| 160 | VTA_R | 0.009 | 0.004 | 0.013 | 0.002 | 3.712 | 0.027 | 2.06E-04 | 18775 | -0.007 | -0.013 | -0.001 | 0.003 | -2.425 | -0.018 | 1.53E-02 | 18750 |
| 161 | SN_pc_L | 0.015 | 0.006 | 0.023 | 0.004 | 3.483 | 0.025 | 4.96E-04 | 18775 | -0.014 | -0.025 | -0.004 | 0.005 | -2.668 | -0.019 | 7.64E-03 | 18750 |
| 162 | SN_pc_R | 0.020 | 0.009 | 0.031 | 0.005 | 3.658 | 0.027 | 2.55E-04 | 18775 | -0.017 | -0.031 | -0.003 | 0.007 | -2.376 | -0.017 | 1.75E-02 | 18750 |
| 163 | SN_pr_L | 0.010 | -0.002 | 0.021 | 0.006 | 1.689 | 0.012 | 9.12E-02 | 18775 | -0.019 | -0.034 | -0.005 | 0.007 | -2.630 | -0.019 | 8.56E-03 | 18750 |
| 164 | SN_pr_R | 0.017 | 0.004 | 0.029 | 0.006 | 2.625 | 0.019 | 8.68E-03 | 18775 | -0.019 | -0.035 | -0.003 | 0.008 | -2.314 | -0.017 | 2.07E-02 | 18750 |
| 165 | Red_N_L | 0.001 | -0.013 | 0.014 | 0.007 | 0.085 | 0.001 | 9.33E-01 | 18775 | -0.029 | -0.047 | -0.012 | 0.009 | -3.234 | -0.024 | 1.22E-03 | 18750 |
| 166 | Red_N_R | 0.002 | -0.013 | 0.017 | 0.008 | 0.292 | 0.002 | 7.71E-01 | 18775 | -0.022 | -0.041 | -0.002 | 0.010 | -2.180 | -0.016 | 2.93E-02 | 18750 |
| 167 | LC_L | -0.006 | -0.011 | -0.001 | 0.003 | -2.265 | -0.017 | 2.35E-02 | 18775 | -0.004 | -0.011 | 0.002 | 0.003 | -1.274 | -0.009 | 2.03E-01 | 18750 |
| 168 | LC_R | -0.006 | -0.011 | 0.000 | 0.003 | -2.068 | -0.015 | 3.87E-02 | 18775 | -0.006 | -0.013 | 0.002 | 0.004 | -1.491 | -0.011 | 1.36E-01 | 18750 |
| 169 | Raphe_D | -0.005 | -0.025 | 0.015 | 0.010 | -0.512 | -0.004 | 6.08E-01 | 18775 | -0.046 | -0.072 | -0.020 | 0.013 | -3.474 | -0.025 | 5.15E-04 | 18750 |
| 170 | Raphe_M | -0.003 | -0.009 | 0.003 | 0.003 | -0.959 | -0.007 | 3.37E-01 | 18775 | -0.007 | -0.015 | 0.001 | 0.004 | -1.677 | -0.012 | 9.36E-02 | 18750 |

| | | Cooked vegetable intake | | | | | | | | Processed meat intake | | | | | | | |
|---|---|---|---|---|---|---|---|---|---|---|---|---|---|---|---|---|---|
| **Number** | **AAL 3 region** | **beta** | **95%CI_l** | **95%CI_h** | **se** | **t** | **r** | **p** | **df.e** | **beta** | **95%CI_l** | **95%CI_h** | **se** | **t** | **r** | **p** | **df.e** |
| 0 | Total brain | -153.752 | -253.273 | -54.231 | 50.774 | -3.028 | -0.022 | 2.46E-03 | 18661 | -319.575 | -450.441 | -188.710 | 66.765 | -4.787 | -0.035 | 1.71E-06 | 18865 |
| 1 | Precentral_L | -2.700 | -6.202 | 0.801 | 1.786 | -1.511 | -0.011 | 1.31E-01 | 18661 | -5.118 | -9.722 | -0.515 | 2.349 | -2.179 | -0.016 | 2.93E-02 | 18865 |

| | | | | | | | | | | | | | | | | | |
|---|---|---|---|---|---|---|---|---|---|---|---|---|---|---|---|---|---|
| 2 | Precentral_R | -1.535 | -4.741 | 1.672 | 1.636 | -0.938 | -0.007 | 3.48E-01 | 18661 | -4.252 | -8.470 | -0.034 | 2.152 | -1.976 | -0.014 | 4.82E-02 | 18865 |
| 3 | Frontal_Sup_2_L | -4.404 | -8.101 | -0.708 | 1.886 | -2.335 | -0.017 | 1.95E-02 | 18661 | -3.736 | -8.598 | 1.125 | 2.480 | -1.507 | -0.011 | 1.32E-01 | 18865 |
| 4 | Frontal_Sup_2_R | -3.610 | -7.685 | 0.464 | 2.079 | -1.737 | -0.013 | 8.24E-02 | 18661 | -6.481 | -11.839 | -1.123 | 2.734 | -2.371 | -0.017 | 1.78E-02 | 18865 |
| 5 | Frontal_Mid_2_L | -2.407 | -6.289 | 1.475 | 1.980 | -1.215 | -0.009 | 2.24E-01 | 18661 | -1.255 | -6.361 | 3.851 | 2.605 | -0.482 | -0.004 | 6.30E-01 | 18865 |
| 6 | Frontal_Mid_2_R | -3.511 | -7.900 | 0.878 | 2.239 | -1.568 | -0.011 | 1.17E-01 | 18661 | -6.255 | -12.033 | -0.477 | 2.948 | -2.122 | -0.015 | 3.38E-02 | 18865 |
| 7 | Frontal_Inf_Oper_L | -0.522 | -1.668 | 0.624 | 0.585 | -0.893 | -0.007 | 3.72E-01 | 18661 | -1.703 | -3.212 | -0.195 | 0.770 | -2.213 | -0.016 | 2.69E-02 | 18865 |
| 8 | Frontal_Inf_Oper_R | -1.182 | -2.620 | 0.256 | 0.734 | -1.611 | -0.012 | 1.07E-01 | 18661 | -1.787 | -3.679 | 0.105 | 0.965 | -1.851 | -0.013 | 6.41E-02 | 18865 |
| 9 | Frontal_Inf_Tri_L | -2.319 | -4.750 | 0.113 | 1.241 | -1.869 | -0.014 | 6.16E-02 | 18661 | -3.220 | -6.422 | -0.017 | 1.634 | -1.971 | -0.014 | 4.88E-02 | 18865 |
| 10 | Frontal_Inf_Tri_R | -0.378 | -2.288 | 1.533 | 0.975 | -0.388 | -0.003 | 6.98E-01 | 18661 | -3.003 | -5.519 | -0.488 | 1.283 | -2.340 | -0.017 | 1.93E-02 | 18865 |
| 11 | Frontal_Inf_Orb_2_L | -1.114 | -1.842 | -0.386 | 0.371 | -3.000 | -0.022 | 2.71E-03 | 18661 | -0.384 | -1.342 | 0.574 | 0.489 | -0.785 | -0.006 | 4.32E-01 | 18865 |
| 12 | Frontal_Inf_Orb_2_R | -0.922 | -1.682 | -0.161 | 0.388 | -2.374 | -0.017 | 1.76E-02 | 18661 | -0.878 | -1.879 | 0.123 | 0.511 | -1.719 | -0.013 | 8.56E-02 | 18865 |
| 13 | Rolandic_Oper_L | -1.863 | -2.963 | -0.762 | 0.562 | -3.317 | -0.024 | 9.10E-04 | 18661 | -1.402 | -2.851 | 0.046 | 0.739 | -1.897 | -0.014 | 5.78E-02 | 18865 |
| 14 | Rolandic_Oper_R | -2.665 | -4.240 | -1.090 | 0.803 | -3.317 | -0.024 | 9.10E-04 | 18661 | -2.385 | -4.458 | -0.312 | 1.058 | -2.256 | -0.016 | 2.41E-02 | 18865 |
| 15 | Supp_Motor_Area_L | -1.961 | -4.579 | 0.656 | 1.336 | -1.469 | -0.011 | 1.42E-01 | 18661 | -3.457 | -6.901 | -0.012 | 1.757 | -1.967 | -0.014 | 4.92E-02 | 18865 |
| 16 | Supp_Motor_Area_R | -1.820 | -4.713 | 1.074 | 1.476 | -1.233 | -0.009 | 2.18E-01 | 18661 | -3.688 | -7.494 | 0.118 | 1.942 | -1.900 | -0.014 | 5.75E-02 | 18865 |
| 17 | Olfactory_L | -0.539 | -0.928 | -0.149 | 0.199 | -2.711 | -0.020 | 6.71E-03 | 18661 | -0.195 | -0.707 | 0.317 | 0.261 | -0.745 | -0.005 | 4.56E-01 | 18865 |
| 18 | Olfactory_R | -0.457 | -0.847 | -0.066 | 0.199 | -2.291 | -0.017 | 2.20E-02 | 18661 | -0.174 | -0.687 | 0.340 | 0.262 | -0.664 | -0.005 | 5.07E-01 | 18865 |
| 19 | Frontal_Sup_Medial_L | -2.405 | -4.859 | 0.050 | 1.252 | -1.920 | -0.014 | 5.48E-02 | 18661 | -4.668 | -7.896 | -1.439 | 1.647 | -2.834 | -0.021 | 4.60E-03 | 18865 |
| 20 | Frontal_Sup_Medial_R | -2.380 | -4.167 | -0.593 | 0.912 | -2.611 | -0.019 | 9.04E-03 | 18661 | -3.279 | -5.628 | -0.930 | 1.198 | -2.736 | -0.020 | 6.22E-03 | 18865 |
| 21 | Frontal_Med_Orb_L | -1.455 | -2.373 | -0.538 | 0.468 | -3.109 | -0.023 | 1.88E-03 | 18661 | -0.893 | -2.099 | 0.313 | 0.615 | -1.451 | -0.011 | 1.47E-01 | 18865 |
| 22 | Frontal_Med_Orb_R | -1.615 | -2.743 | -0.487 | 0.575 | -2.807 | -0.021 | 5.01E-03 | 18661 | -1.365 | -2.847 | 0.117 | 0.756 | -1.805 | -0.013 | 7.10E-02 | 18865 |
| 23 | Rectus_L | -1.573 | -2.523 | -0.624 | 0.484 | -3.249 | -0.024 | 1.16E-03 | 18661 | -1.391 | -2.639 | -0.143 | 0.637 | -2.184 | -0.016 | 2.90E-02 | 18865 |
| 24 | Rectus_R | -1.297 | -2.175 | -0.419 | 0.448 | -2.896 | -0.021 | 3.79E-03 | 18661 | -0.971 | -2.126 | 0.184 | 0.589 | -1.648 | -0.012 | 9.94E-02 | 18865 |
| 25 | OFCmed_L | -1.115 | -1.665 | -0.565 | 0.281 | -3.971 | -0.029 | 7.19E-05 | 18661 | -0.965 | -1.688 | -0.241 | 0.369 | -2.613 | -0.019 | 8.98E-03 | 18865 |
| 26 | OFCmed_R | -0.706 | -1.347 | -0.065 | 0.327 | -2.158 | -0.016 | 3.09E-02 | 18661 | -0.612 | -1.455 | 0.231 | 0.430 | -1.422 | -0.010 | 1.55E-01 | 18865 |
| 27 | OFCant_L | -1.033 | -1.669 | -0.396 | 0.325 | -3.181 | -0.023 | 1.47E-03 | 18661 | -0.668 | -1.505 | 0.169 | 0.427 | -1.565 | -0.011 | 1.18E-01 | 18865 |
| 28 | OFCant_R | -0.955 | -1.796 | -0.114 | 0.429 | -2.227 | -0.016 | 2.60E-02 | 18661 | -0.890 | -1.995 | 0.214 | 0.564 | -1.580 | -0.012 | 1.14E-01 | 18865 |
| 29 | OFCpost_L | -1.413 | -2.209 | -0.616 | 0.406 | -3.476 | -0.025 | 5.09E-04 | 18661 | -0.320 | -1.368 | 0.727 | 0.534 | -0.600 | -0.004 | 5.49E-01 | 18865 |
| 30 | OFCpost_R | -1.332 | -2.069 | -0.596 | 0.376 | -3.546 | -0.026 | 3.93E-04 | 18661 | -0.910 | -1.878 | 0.058 | 0.494 | -1.843 | -0.013 | 6.53E-02 | 18865 |
| 31 | OFClat_L | -0.291 | -0.560 | -0.023 | 0.137 | -2.126 | -0.016 | 3.35E-02 | 18661 | -0.376 | -0.729 | -0.022 | 0.180 | -2.082 | -0.015 | 3.73E-02 | 18865 |
| 32 | OFClat_R | -0.409 | -0.636 | -0.182 | 0.116 | -3.532 | -0.026 | 4.13E-04 | 18661 | -0.330 | -0.628 | -0.031 | 0.152 | -2.165 | -0.016 | 3.04E-02 | 18865 |
| 33 | Insula_L | -3.156 | -5.618 | -0.694 | 1.256 | -2.513 | -0.018 | 1.20E-02 | 18661 | -2.830 | -6.070 | 0.409 | 1.653 | -1.713 | -0.012 | 8.68E-02 | 18865 |
| 34 | Insula_R | -3.595 | -5.996 | -1.195 | 1.225 | -2.936 | -0.021 | 3.33E-03 | 18661 | -3.176 | -6.333 | -0.019 | 1.611 | -1.972 | -0.014 | 4.86E-02 | 18865 |
| 37 | Cingulate_Mid_L | -2.952 | -5.035 | -0.868 | 1.063 | -2.777 | -0.020 | 5.49E-03 | 18661 | -7.593 | -10.333 | -4.854 | 1.398 | -5.433 | -0.040 | 5.62E-08 | 18865 |
| 38 | Cingulate_Mid_R | -2.251 | -4.470 | -0.032 | 1.132 | -1.988 | -0.015 | 4.68E-02 | 18661 | -7.364 | -10.284 | -4.444 | 1.490 | -4.943 | -0.036 | 7.77E-07 | 18865 |
| 39 | Cingulate_Post_L | -0.776 | -1.411 | -0.140 | 0.324 | -2.393 | -0.018 | 1.67E-02 | 18661 | -1.577 | -2.412 | -0.741 | 0.426 | -3.697 | -0.027 | 2.19E-04 | 18865 |
| 40 | Cingulate_Post_R | -0.435 | -0.788 | -0.082 | 0.180 | -2.418 | -0.018 | 1.56E-02 | 18661 | -0.670 | -1.134 | -0.206 | 0.237 | -2.831 | -0.021 | 4.64E-03 | 18865 |
| 41 | Hippocampus_L | -0.457 | -1.485 | 0.571 | 0.524 | -0.871 | -0.006 | 3.84E-01 | 18661 | -0.702 | -2.054 | 0.651 | 0.690 | -1.016 | -0.007 | 3.09E-01 | 18865 |
| 42 | Hippocampus_R | -0.479 | -1.546 | 0.588 | 0.544 | -0.880 | -0.006 | 3.79E-01 | 18661 | -0.618 | -2.021 | 0.786 | 0.716 | -0.863 | -0.006 | 3.88E-01 | 18865 |
| 43 | ParaHippocampal_L | -0.421 | -1.335 | 0.492 | 0.466 | -0.904 | -0.007 | 3.66E-01 | 18661 | -1.877 | -3.079 | -0.675 | 0.613 | -3.062 | -0.022 | 2.20E-03 | 18865 |
| 44 | ParaHippocampal_R | -0.195 | -1.350 | 0.959 | 0.589 | -0.331 | -0.002 | 7.40E-01 | 18661 | -2.353 | -3.870 | -0.835 | 0.774 | -3.039 | -0.022 | 2.37E-03 | 18865 |
| 45 | Amygdala_L | -0.395 | -0.750 | -0.040 | 0.181 | -2.182 | -0.016 | 2.91E-02 | 18661 | 0.031 | -0.436 | 0.498 | 0.238 | 0.130 | 0.001 | 8.97E-01 | 18865 |
| 46 | Amygdala_R | -0.368 | -0.732 | -0.005 | 0.185 | -1.987 | -0.015 | 4.70E-02 | 18661 | -0.280 | -0.758 | 0.198 | 0.244 | -1.149 | -0.008 | 2.51E-01 | 18865 |
| 47 | Calcarine_L | -1.349 | -4.735 | 2.038 | 1.728 | -0.781 | -0.006 | 4.35E-01 | 18661 | -4.546 | -9.001 | -0.091 | 2.273 | -2.000 | -0.015 | 4.55E-02 | 18865 |
| 48 | Calcarine_R | -1.656 | -4.486 | 1.174 | 1.444 | -1.147 | -0.008 | 2.51E-01 | 18661 | -3.055 | -6.778 | 0.668 | 1.899 | -1.608 | -0.012 | 1.08E-01 | 18865 |
| 49 | Cuneus_L | -1.171 | -3.298 | 0.956 | 1.085 | -1.079 | -0.008 | 2.80E-01 | 18661 | -4.169 | -6.965 | -1.372 | 1.427 | -2.921 | -0.021 | 3.49E-03 | 18865 |
| 50 | Cuneus_R | -1.138 | -2.765 | 0.489 | 0.830 | -1.371 | -0.010 | 1.71E-01 | 18661 | -3.535 | -5.673 | -1.396 | 1.091 | -3.240 | -0.024 | 1.20E-03 | 18865 |
| 51 | Lingual_L | -1.680 | -3.719 | 0.360 | 1.041 | -1.614 | -0.012 | 1.07E-01 | 18661 | -4.272 | -6.953 | -1.591 | 1.368 | -3.123 | -0.023 | 1.79E-03 | 18865 |
| 52 | Lingual_R | -2.461 | -4.598 | -0.324 | 1.090 | -2.257 | -0.017 | 2.40E-02 | 18661 | -4.078 | -6.887 | -1.270 | 1.433 | -2.846 | -0.021 | 4.43E-03 | 18865 |

| | | | | | | | | | | | | | | | | | |
|---|---|---|---|---|---|---|---|---|---|---|---|---|---|---|---|---|---|
| 53 | Occipital_Sup_L | -1.087 | -2.489 | 0.315 | 0.715 | -1.520 | -0.011 | 1.29E-01 | 18661 | -4.631 | -6.473 | -2.789 | 0.940 | -4.927 | -0.036 | 8.43E-07 | 18865 |
| 54 | Occipital_Sup_R | -1.225 | -2.746 | 0.295 | 0.776 | -1.580 | -0.012 | 1.14E-01 | 18661 | -2.541 | -4.538 | -0.543 | 1.019 | -2.493 | -0.018 | 1.27E-02 | 18865 |
| 55 | Occipital_Mid_L | -2.551 | -5.833 | 0.730 | 1.674 | -1.524 | -0.011 | 1.28E-01 | 18661 | -5.382 | -9.701 | -1.063 | 2.204 | -2.443 | -0.018 | 1.46E-02 | 18865 |
| 56 | Occipital_Mid_R | -0.842 | -3.129 | 1.444 | 1.166 | -0.722 | -0.005 | 4.70E-01 | 18661 | -1.040 | -4.047 | 1.967 | 1.534 | -0.678 | -0.005 | 4.98E-01 | 18865 |
| 57 | Occipital_Inf_L | -1.295 | -2.384 | -0.206 | 0.556 | -2.330 | -0.017 | 1.98E-02 | 18661 | -0.378 | -1.811 | 1.054 | 0.731 | -0.518 | -0.004 | 6.05E-01 | 18865 |
| 58 | Occipital_Inf_R | -1.783 | -3.044 | -0.523 | 0.643 | -2.773 | -0.020 | 5.57E-03 | 18661 | 1.773 | 0.117 | 3.429 | 0.845 | 2.098 | 0.015 | 3.59E-02 | 18865 |
| 59 | Fusiform_L | -0.172 | -2.559 | 2.215 | 1.218 | -0.142 | -0.001 | 8.87E-01 | 18661 | -5.953 | -9.091 | -2.816 | 1.601 | -3.720 | -0.027 | 2.00E-04 | 18865 |
| 60 | Fusiform_R | -1.848 | -4.470 | 0.774 | 1.338 | -1.381 | -0.010 | 1.67E-01 | 18661 | -5.680 | -9.127 | -2.232 | 1.759 | -3.229 | -0.024 | 1.24E-03 | 18865 |
| 61 | Postcentral_L | -3.303 | -6.893 | 0.288 | 1.832 | -1.803 | -0.013 | 7.14E-02 | 18661 | -9.732 | -14.453 | -5.010 | 2.409 | -4.040 | -0.029 | 5.37E-05 | 18865 |
| 62 | Postcentral_R | -3.795 | -7.474 | -0.117 | 1.877 | -2.022 | -0.015 | 4.32E-02 | 18661 | -9.485 | -14.320 | -4.650 | 2.467 | -3.845 | -0.028 | 1.21E-04 | 18865 |
| 63 | Parietal_Sup_L | -1.715 | -3.782 | 0.352 | 1.054 | -1.627 | -0.012 | 1.04E-01 | 18661 | -4.104 | -6.824 | -1.384 | 1.388 | -2.957 | -0.022 | 3.11E-03 | 18865 |
| 64 | Parietal_Sup_R | -0.587 | -2.421 | 1.247 | 0.936 | -0.627 | -0.005 | 5.31E-01 | 18661 | -3.090 | -5.502 | -0.678 | 1.231 | -2.511 | -0.018 | 1.20E-02 | 18865 |
| 65 | Parietal_Inf_L | -1.848 | -4.557 | 0.860 | 1.382 | -1.338 | -0.010 | 1.81E-01 | 18661 | -9.842 | -13.400 | -6.283 | 1.816 | -5.421 | -0.039 | 6.00E-08 | 18865 |
| 66 | Parietal_Inf_R | -0.340 | -2.094 | 1.414 | 0.895 | -0.380 | -0.003 | 7.04E-01 | 18661 | -3.445 | -5.753 | -1.137 | 1.178 | -2.925 | -0.021 | 3.44E-03 | 18865 |
| 67 | SupraMarginal_L | -1.897 | -3.483 | -0.310 | 0.810 | -2.343 | -0.017 | 1.91E-02 | 18661 | -3.611 | -5.698 | -1.524 | 1.065 | -3.392 | -0.025 | 6.96E-04 | 18865 |
| 68 | SupraMarginal_R | -2.467 | -4.715 | -0.220 | 1.147 | -2.152 | -0.016 | 3.14E-02 | 18661 | -4.494 | -7.449 | -1.539 | 1.508 | -2.981 | -0.022 | 2.87E-03 | 18865 |
| 69 | Angular_L | -0.772 | -2.332 | 0.788 | 0.796 | -0.970 | -0.007 | 3.32E-01 | 18661 | -3.676 | -5.727 | -1.626 | 1.046 | -3.514 | -0.026 | 4.42E-04 | 18865 |
| 70 | Angular_R | -0.719 | -2.809 | 1.371 | 1.066 | -0.674 | -0.005 | 5.00E-01 | 18661 | -2.675 | -5.423 | 0.073 | 1.402 | -1.908 | -0.014 | 5.64E-02 | 18865 |
| 71 | Precuneus_L | -1.131 | -4.933 | 2.671 | 1.940 | -0.583 | -0.004 | 5.60E-01 | 18661 | -14.675 | -19.672 | -9.679 | 2.549 | -5.757 | -0.042 | 8.69E-09 | 18865 |
| 72 | Precuneus_R | -1.824 | -5.211 | 1.562 | 1.728 | -1.056 | -0.008 | 2.91E-01 | 18661 | -9.365 | -13.816 | -4.914 | 2.271 | -4.124 | -0.030 | 3.74E-05 | 18865 |
| 73 | Paracentral_Lobule_L | -0.494 | -2.232 | 1.245 | 0.887 | -0.557 | -0.004 | 5.78E-01 | 18661 | -2.833 | -5.121 | -0.544 | 1.168 | -2.426 | -0.018 | 1.53E-02 | 18865 |
| 74 | Paracentral_Lobule_R | -0.898 | -2.143 | 0.347 | 0.635 | -1.413 | -0.010 | 1.58E-01 | 18661 | -2.092 | -3.731 | -0.453 | 0.836 | -2.501 | -0.018 | 1.24E-02 | 18865 |
| 75 | Caudate_L | -0.240 | -1.629 | 1.148 | 0.708 | -0.339 | -0.002 | 7.34E-01 | 18661 | -1.232 | -3.060 | 0.595 | 0.932 | -1.322 | -0.010 | 1.86E-01 | 18865 |
| 76 | Caudate_R | -0.285 | -1.859 | 1.288 | 0.803 | -0.355 | -0.003 | 7.22E-01 | 18661 | -0.642 | -2.712 | 1.429 | 1.056 | -0.607 | -0.004 | 5.44E-01 | 18865 |
| 77 | Putamen_L | -0.393 | -2.503 | 1.717 | 1.077 | -0.365 | -0.003 | 7.15E-01 | 18661 | -2.112 | -4.888 | 0.663 | 1.416 | -1.492 | -0.011 | 1.36E-01 | 18865 |
| 78 | Putamen_R | -0.960 | -3.110 | 1.189 | 1.097 | -0.876 | -0.006 | 3.81E-01 | 18661 | -2.469 | -5.296 | 0.358 | 1.442 | -1.712 | -0.012 | 8.70E-02 | 18865 |
| 79 | Pallidum_L | -0.006 | -0.194 | 0.181 | 0.096 | -0.067 | 0.000 | 9.47E-01 | 18661 | -0.145 | -0.392 | 0.101 | 0.126 | -1.155 | -0.008 | 2.48E-01 | 18865 |
| 80 | Pallidum_R | -0.074 | -0.280 | 0.132 | 0.105 | -0.705 | -0.005 | 4.81E-01 | 18661 | -0.210 | -0.481 | 0.061 | 0.138 | -1.521 | -0.011 | 1.28E-01 | 18865 |
| 83 | Heschl_L | -0.420 | -0.801 | -0.040 | 0.194 | -2.168 | -0.016 | 3.02E-02 | 18661 | -0.686 | -1.186 | -0.185 | 0.255 | -2.686 | -0.020 | 7.24E-03 | 18865 |
| 84 | Heschl_R | -0.633 | -1.058 | -0.208 | 0.217 | -2.918 | -0.021 | 3.53E-03 | 18661 | -0.632 | -1.191 | -0.073 | 0.285 | -2.217 | -0.016 | 2.66E-02 | 18865 |
| 85 | Temporal_Sup_L | -4.936 | -7.680 | -2.192 | 1.400 | -3.525 | -0.026 | 4.24E-04 | 18661 | -4.515 | -8.124 | -0.905 | 1.841 | -2.452 | -0.018 | 1.42E-02 | 18865 |
| 86 | Temporal_Sup_R | -4.756 | -7.794 | -1.718 | 1.550 | -3.068 | -0.022 | 2.16E-03 | 18661 | -4.548 | -8.544 | -0.551 | 2.039 | -2.231 | -0.016 | 2.57E-02 | 18865 |
| 87 | Temporal_Pole_Sup_L | -1.003 | -2.305 | 0.298 | 0.664 | -1.511 | -0.011 | 1.31E-01 | 18661 | -0.663 | -2.374 | 1.048 | 0.873 | -0.759 | -0.006 | 4.48E-01 | 18865 |
| 88 | Temporal_Pole_Sup_R | -0.552 | -1.921 | 0.817 | 0.699 | -0.790 | -0.006 | 4.29E-01 | 18661 | -1.320 | -3.121 | 0.480 | 0.919 | -1.437 | -0.010 | 1.51E-01 | 18865 |
| 89 | Temporal_Mid_L | -6.018 | -10.598 | -1.438 | 2.336 | -2.576 | -0.019 | 1.00E-02 | 18661 | -8.039 | -14.062 | -2.017 | 3.072 | -2.617 | -0.019 | 8.89E-03 | 18865 |
| 90 | Temporal_Mid_R | -3.380 | -7.446 | 0.686 | 2.074 | -1.629 | -0.012 | 1.03E-01 | 18661 | -5.772 | -11.118 | -0.425 | 2.728 | -2.116 | -0.015 | 3.44E-02 | 18865 |
| 91 | Temporal_Pole_Mid_L | 0.076 | -1.019 | 1.170 | 0.558 | 0.136 | 0.001 | 8.92E-01 | 18661 | -1.365 | -2.803 | 0.073 | 0.734 | -1.860 | -0.014 | 6.28E-02 | 18865 |
| 92 | Temporal_Pole_Mid_R | -0.923 | -2.251 | 0.405 | 0.678 | -1.363 | -0.010 | 1.73E-01 | 18661 | -0.798 | -2.544 | 0.949 | 0.891 | -0.895 | -0.007 | 3.71E-01 | 18865 |
| 93 | Temporal_Inf_L | -4.402 | -7.551 | -1.254 | 1.606 | -2.741 | -0.020 | 6.14E-03 | 18661 | -2.730 | -6.870 | 1.410 | 2.112 | -1.293 | -0.009 | 1.96E-01 | 18865 |
| 94 | Temporal_Inf_R | -5.271 | -8.703 | -1.840 | 1.750 | -3.011 | -0.022 | 2.60E-03 | 18661 | -1.449 | -5.962 | 3.064 | 2.302 | -0.629 | -0.005 | 5.29E-01 | 18865 |
| 95 | Cerebellum_Crus1_L | -2.161 | -5.901 | 1.580 | 1.908 | -1.132 | -0.008 | 2.58E-01 | 18661 | -5.847 | -10.762 | -0.933 | 2.507 | -2.332 | -0.017 | 1.97E-02 | 18865 |
| 96 | Cerebellum_Crus1_R | 0.428 | -3.384 | 4.239 | 1.945 | 0.220 | 0.002 | 8.26E-01 | 18661 | -4.199 | -9.211 | 0.813 | 2.557 | -1.642 | -0.012 | 1.01E-01 | 18865 |
| 97 | Cerebellum_Crus2_L | -2.334 | -5.298 | 0.631 | 1.512 | -1.543 | -0.011 | 1.23E-01 | 18661 | -8.275 | -12.171 | -4.378 | 1.988 | -4.162 | -0.030 | 3.16E-05 | 18865 |
| 98 | Cerebellum_Crus2_R | -1.070 | -3.808 | 1.668 | 1.397 | -0.766 | -0.006 | 4.44E-01 | 18661 | -4.665 | -8.269 | -1.062 | 1.838 | -2.538 | -0.018 | 1.12E-02 | 18865 |
| 99 | Cerebellum_3_L | 0.265 | 0.066 | 0.464 | 0.101 | 2.609 | 0.019 | 9.08E-03 | 18661 | -0.125 | -0.386 | 0.137 | 0.133 | -0.936 | -0.007 | 3.49E-01 | 18865 |
| 100 | Cerebellum_3_R | 0.255 | -0.013 | 0.523 | 0.137 | 1.862 | 0.014 | 6.26E-02 | 18661 | -0.214 | -0.567 | 0.139 | 0.180 | -1.189 | -0.009 | 2.34E-01 | 18865 |
| 101 | Cerebellum_4_5_L | 1.827 | 0.219 | 3.435 | 0.820 | 2.227 | 0.016 | 2.59E-02 | 18661 | -1.287 | -3.403 | 0.829 | 1.079 | -1.192 | -0.009 | 2.33E-01 | 18865 |
| 102 | Cerebellum_4_5_R | 1.003 | -0.308 | 2.313 | 0.669 | 1.500 | 0.011 | 1.34E-01 | 18661 | -1.419 | -3.142 | 0.305 | 0.879 | -1.613 | -0.012 | 1.07E-01 | 18865 |
| 103 | Cerebellum_6_L | 1.862 | -0.921 | 4.644 | 1.420 | 1.311 | 0.010 | 1.90E-01 | 18661 | -4.510 | -8.170 | -0.851 | 1.867 | -2.416 | -0.018 | 1.57E-02 | 18865 |

| | | | | | | | | | | | | | | | | | |
|---|---|---|---|---|---|---|---|---|---|---|---|---|---|---|---|---|---|
| 104 | Cerebellum_6_R | 1.855 | -1.075 | 4.785 | 1.495 | 1.241 | 0.009 | 2.15E-01 | 18661 | -3.666 | -7.521 | 0.188 | 1.966 | -1.864 | -0.014 | 6.23E-02 | 18865 |
| 105 | Cerebellum_7b_L | -0.643 | -1.549 | 0.262 | 0.462 | -1.393 | -0.010 | 1.64E-01 | 18661 | -1.680 | -2.871 | -0.488 | 0.608 | -2.763 | -0.020 | 5.74E-03 | 18865 |
| 106 | Cerebellum_7b_R | -0.597 | -1.494 | 0.300 | 0.458 | -1.305 | -0.010 | 1.92E-01 | 18661 | -1.548 | -2.729 | -0.367 | 0.602 | -2.570 | -0.019 | 1.02E-02 | 18865 |
| 107 | Cerebellum_8_L | -2.105 | -4.981 | 0.770 | 1.467 | -1.435 | -0.011 | 1.51E-01 | 18661 | -1.370 | -5.156 | 2.417 | 1.932 | -0.709 | -0.005 | 4.78E-01 | 18865 |
| 108 | Cerebellum_8_R | -2.355 | -6.098 | 1.389 | 1.910 | -1.233 | -0.009 | 2.18E-01 | 18661 | -5.425 | -10.352 | -0.498 | 2.513 | -2.158 | -0.016 | 3.09E-02 | 18865 |
| 109 | Cerebellum_9_L | -0.427 | -2.054 | 1.200 | 0.830 | -0.515 | -0.004 | 6.07E-01 | 18661 | 1.841 | -0.299 | 3.981 | 1.092 | 1.686 | 0.012 | 9.18E-02 | 18865 |
| 110 | Cerebellum_9_R | -0.547 | -2.146 | 1.053 | 0.816 | -0.670 | -0.005 | 5.03E-01 | 18661 | 1.779 | -0.324 | 3.882 | 1.073 | 1.658 | 0.012 | 9.74E-02 | 18865 |
| 111 | Cerebellum_10_L | 0.050 | -0.065 | 0.166 | 0.059 | 0.851 | 0.006 | 3.95E-01 | 18661 | 0.244 | 0.092 | 0.395 | 0.077 | 3.150 | 0.023 | 1.64E-03 | 18865 |
| 112 | Cerebellum_10_R | 0.057 | -0.063 | 0.178 | 0.061 | 0.939 | 0.007 | 3.48E-01 | 18661 | 0.121 | -0.037 | 0.279 | 0.081 | 1.499 | 0.011 | 1.34E-01 | 18865 |
| 113 | Vermis_1_2 | 0.097 | 0.022 | 0.172 | 0.038 | 2.530 | 0.019 | 1.14E-02 | 18661 | -0.035 | -0.134 | 0.063 | 0.050 | -0.702 | -0.005 | 4.83E-01 | 18865 |
| 114 | Vermis_3 | 0.202 | -0.096 | 0.500 | 0.152 | 1.327 | 0.010 | 1.84E-01 | 18661 | -0.355 | -0.747 | 0.037 | 0.200 | -1.776 | -0.013 | 7.58E-02 | 18865 |
| 115 | Vermis_4_5 | -0.211 | -1.087 | 0.664 | 0.447 | -0.473 | -0.003 | 6.36E-01 | 18661 | -0.605 | -1.757 | 0.547 | 0.588 | -1.029 | -0.007 | 3.04E-01 | 18865 |
| 116 | Vermis_6 | 0.256 | -0.231 | 0.744 | 0.249 | 1.031 | 0.008 | 3.02E-01 | 18661 | -0.349 | -0.990 | 0.293 | 0.327 | -1.065 | -0.008 | 2.87E-01 | 18865 |
| 117 | Vermis_7 | 0.184 | -0.091 | 0.460 | 0.141 | 1.311 | 0.010 | 1.90E-01 | 18661 | -0.013 | -0.376 | 0.350 | 0.185 | -0.071 | -0.001 | 9.43E-01 | 18865 |
| 118 | Vermis_8 | 0.438 | -0.015 | 0.891 | 0.231 | 1.894 | 0.014 | 5.82E-02 | 18661 | 0.107 | -0.489 | 0.704 | 0.304 | 0.353 | 0.003 | 7.24E-01 | 18865 |
| 119 | Vermis_9 | 0.141 | -0.227 | 0.508 | 0.188 | 0.750 | 0.005 | 4.54E-01 | 18661 | 0.234 | -0.250 | 0.717 | 0.247 | 0.947 | 0.007 | 3.44E-01 | 18865 |
| 120 | Vermis_10 | 0.082 | -0.021 | 0.184 | 0.052 | 1.562 | 0.011 | 1.18E-01 | 18661 | 0.191 | 0.056 | 0.325 | 0.069 | 2.774 | 0.020 | 5.54E-03 | 18865 |
| 121 | Thal_AV_L | -0.022 | -0.063 | 0.019 | 0.021 | -1.040 | -0.008 | 2.98E-01 | 18661 | -0.075 | -0.129 | -0.021 | 0.028 | -2.720 | -0.020 | 6.53E-03 | 18865 |
| 122 | Thal_AV_R | 0.009 | -0.050 | 0.068 | 0.030 | 0.299 | 0.002 | 7.65E-01 | 18661 | -0.096 | -0.173 | -0.019 | 0.039 | -2.432 | -0.018 | 1.50E-02 | 18865 |
| 123 | Thal_LP_L | -0.019 | -0.065 | 0.026 | 0.023 | -0.840 | -0.006 | 4.01E-01 | 18661 | -0.097 | -0.156 | -0.037 | 0.030 | -3.165 | -0.023 | 1.55E-03 | 18865 |
| 124 | Thal_LP_R | 0.005 | -0.055 | 0.064 | 0.030 | 0.154 | 0.001 | 8.78E-01 | 18661 | -0.105 | -0.183 | -0.027 | 0.040 | -2.651 | -0.019 | 8.04E-03 | 18865 |
| 125 | Thal_VA_L | -0.018 | -0.067 | 0.030 | 0.025 | -0.734 | -0.005 | 4.63E-01 | 18661 | -0.045 | -0.109 | 0.019 | 0.033 | -1.390 | -0.010 | 1.64E-01 | 18865 |
| 126 | Thal_VA_R | -0.007 | -0.067 | 0.052 | 0.030 | -0.237 | -0.002 | 8.13E-01 | 18661 | -0.043 | -0.121 | 0.035 | 0.040 | -1.079 | -0.008 | 2.81E-01 | 18865 |
| 127 | Thal_VL_L | 0.016 | -0.122 | 0.154 | 0.070 | 0.230 | 0.002 | 8.18E-01 | 18661 | -0.154 | -0.335 | 0.028 | 0.093 | -1.659 | -0.012 | 9.71E-02 | 18865 |
| 128 | Thal_VL_R | 0.044 | -0.139 | 0.227 | 0.093 | 0.469 | 0.003 | 6.39E-01 | 18661 | -0.282 | -0.523 | -0.042 | 0.123 | -2.298 | -0.017 | 2.16E-02 | 18865 |
| 129 | Thal_VPL_L | 0.131 | -0.033 | 0.296 | 0.084 | 1.567 | 0.011 | 1.17E-01 | 18661 | -0.066 | -0.282 | 0.150 | 0.110 | -0.596 | -0.004 | 5.51E-01 | 18865 |
| 130 | Thal_VPL_R | 0.071 | -0.102 | 0.243 | 0.088 | 0.800 | 0.006 | 4.24E-01 | 18661 | -0.135 | -0.362 | 0.092 | 0.116 | -1.165 | -0.008 | 2.44E-01 | 18865 |
| 131 | Thal_IL_L | -0.040 | -0.112 | 0.031 | 0.036 | -1.115 | -0.008 | 2.65E-01 | 18661 | -0.102 | -0.195 | -0.008 | 0.048 | -2.134 | -0.016 | 3.28E-02 | 18865 |
| 132 | Thal_IL_R | -0.039 | -0.120 | 0.041 | 0.041 | -0.958 | -0.007 | 3.38E-01 | 18661 | -0.130 | -0.236 | -0.024 | 0.054 | -2.410 | -0.018 | 1.60E-02 | 18865 |
| 133 | Thal_Re_L | 0.000 | -0.001 | 0.000 | 0.000 | -1.258 | -0.009 | 2.08E-01 | 18661 | 0.000 | 0.000 | 0.000 | 0.000 | -0.235 | -0.002 | 8.14E-01 | 18865 |
| 134 | Thal_Re_R | 0.000 | -0.003 | 0.002 | 0.001 | -0.333 | -0.002 | 7.39E-01 | 18661 | -0.002 | -0.005 | 0.001 | 0.002 | -1.267 | -0.009 | 2.05E-01 | 18865 |
| 135 | Thal_MDm_L | -0.339 | -0.865 | 0.188 | 0.269 | -1.260 | -0.009 | 2.08E-01 | 18661 | -0.936 | -1.628 | -0.243 | 0.353 | -2.649 | -0.019 | 8.07E-03 | 18865 |
| 136 | Thal_MDm_R | -0.212 | -0.824 | 0.400 | 0.312 | -0.680 | -0.005 | 4.97E-01 | 18661 | -1.005 | -1.810 | -0.201 | 0.410 | -2.449 | -0.018 | 1.43E-02 | 18865 |
| 137 | Thal_MDl_L | -0.072 | -0.163 | 0.019 | 0.046 | -1.551 | -0.011 | 1.21E-01 | 18661 | -0.185 | -0.305 | -0.065 | 0.061 | -3.024 | -0.022 | 2.49E-03 | 18865 |
| 138 | Thal_MDl_R | -0.030 | -0.133 | 0.074 | 0.053 | -0.565 | -0.004 | 5.72E-01 | 18661 | -0.196 | -0.332 | -0.060 | 0.069 | -2.820 | -0.021 | 4.81E-03 | 18865 |
| 139 | Thal_LGN_L | -0.001 | -0.015 | 0.013 | 0.007 | -0.164 | -0.001 | 8.70E-01 | 18661 | -0.025 | -0.043 | -0.007 | 0.009 | -2.696 | -0.020 | 7.02E-03 | 18865 |
| 140 | Thal_LGN_R | -0.008 | -0.037 | 0.022 | 0.015 | -0.503 | -0.004 | 6.15E-01 | 18661 | -0.062 | -0.101 | -0.023 | 0.020 | -3.132 | -0.023 | 1.74E-03 | 18865 |
| 141 | Thal_MGN_L | -0.017 | -0.042 | 0.007 | 0.013 | -1.379 | -0.010 | 1.68E-01 | 18661 | -0.038 | -0.071 | -0.006 | 0.017 | -2.299 | -0.017 | 2.15E-02 | 18865 |
| 142 | Thal_MGN_R | -0.020 | -0.047 | 0.007 | 0.014 | -1.442 | -0.011 | 1.49E-01 | 18661 | -0.063 | -0.098 | -0.027 | 0.018 | -3.473 | -0.025 | 5.16E-04 | 18865 |
| 143 | Thal_PuA_L | -0.016 | -0.046 | 0.013 | 0.015 | -1.107 | -0.008 | 2.68E-01 | 18661 | -0.065 | -0.104 | -0.027 | 0.020 | -3.345 | -0.024 | 8.26E-04 | 18865 |
| 144 | Thal_PuA_R | -0.039 | -0.100 | 0.022 | 0.031 | -1.262 | -0.009 | 2.07E-01 | 18661 | -0.143 | -0.222 | -0.063 | 0.041 | -3.504 | -0.026 | 4.59E-04 | 18865 |
| 145 | Thal_PuM_L | -0.298 | -0.878 | 0.282 | 0.296 | -1.007 | -0.007 | 3.14E-01 | 18661 | -1.301 | -2.063 | -0.539 | 0.389 | -3.346 | -0.024 | 8.21E-04 | 18865 |
| 146 | Thal_PuM_R | -0.311 | -1.033 | 0.411 | 0.368 | -0.844 | -0.006 | 3.99E-01 | 18661 | -1.469 | -2.419 | -0.520 | 0.485 | -3.033 | -0.022 | 2.43E-03 | 18865 |
| 147 | Thal_PuL_L | -0.050 | -0.156 | 0.056 | 0.054 | -0.930 | -0.007 | 3.52E-01 | 18661 | -0.216 | -0.355 | -0.077 | 0.071 | -3.051 | -0.022 | 2.29E-03 | 18865 |
| 148 | Thal_PuL_R | -0.069 | -0.237 | 0.100 | 0.086 | -0.799 | -0.006 | 4.25E-01 | 18661 | -0.306 | -0.527 | -0.085 | 0.113 | -2.710 | -0.020 | 6.74E-03 | 18865 |
| 149 | Thal_PuI_L | 0.024 | -0.005 | 0.052 | 0.015 | 1.604 | 0.012 | 1.09E-01 | 18661 | -0.023 | -0.061 | 0.014 | 0.019 | -1.212 | -0.009 | 2.26E-01 | 18865 |
| 150 | Thal_PuI_R | 0.018 | -0.021 | 0.056 | 0.019 | 0.902 | 0.007 | 3.67E-01 | 18661 | -0.036 | -0.086 | 0.014 | 0.026 | -1.418 | -0.010 | 1.56E-01 | 18865 |
| 151 | ACC_sub_L | -0.524 | -0.856 | -0.193 | 0.169 | -3.099 | -0.023 | 1.95E-03 | 18661 | -0.055 | -0.491 | 0.381 | 0.223 | -0.247 | -0.002 | 8.05E-01 | 18865 |
| 152 | ACC_sub_R | -0.537 | -0.847 | -0.228 | 0.158 | -3.401 | -0.025 | 6.72E-04 | 18661 | 0.015 | -0.393 | 0.422 | 0.208 | 0.070 | 0.001 | 9.44E-01 | 18865 |

| | | | | | | | | | | | | | | | | | |
|---|---|---|---|---|---|---|---|---|---|---|---|---|---|---|---|---|---|
| 153 | ACC_pre_L | -0.976 | -1.827 | -0.125 | 0.434 | -2.249 | -0.016 | 2.45E-02 | 18661 | -0.965 | -2.084 | 0.153 | 0.571 | -1.692 | -0.012 | 9.07E-02 | 18865 |
| 154 | ACC_pre_R | -0.955 | -1.774 | -0.136 | 0.418 | -2.287 | -0.017 | 2.22E-02 | 18661 | -0.979 | -2.056 | 0.099 | 0.550 | -1.780 | -0.013 | 7.50E-02 | 18865 |
| 155 | ACC_sup_L | -1.331 | -2.256 | -0.407 | 0.472 | -2.823 | -0.021 | 4.76E-03 | 18661 | -1.422 | -2.638 | -0.206 | 0.620 | -2.293 | -0.017 | 2.19E-02 | 18865 |
| 156 | ACC_sup_R | -0.733 | -1.691 | 0.224 | 0.488 | -1.502 | -0.011 | 1.33E-01 | 18661 | -1.062 | -2.321 | 0.197 | 0.642 | -1.653 | -0.012 | 9.83E-02 | 18865 |
| 157 | N_Acc_L | -0.262 | -0.540 | 0.017 | 0.142 | -1.842 | -0.013 | 6.55E-02 | 18661 | -0.316 | -0.682 | 0.050 | 0.187 | -1.694 | -0.012 | 9.04E-02 | 18865 |
| 158 | N_Acc_R | -0.141 | -0.382 | 0.100 | 0.123 | -1.150 | -0.008 | 2.50E-01 | 18661 | -0.219 | -0.536 | 0.097 | 0.162 | -1.357 | -0.010 | 1.75E-01 | 18865 |
| 159 | VTA_L | 0.012 | 0.002 | 0.021 | 0.005 | 2.318 | 0.017 | 2.04E-02 | 18661 | -0.004 | -0.016 | 0.009 | 0.007 | -0.538 | -0.004 | 5.91E-01 | 18865 |
| 160 | VTA_R | 0.012 | 0.003 | 0.021 | 0.004 | 2.610 | 0.019 | 9.05E-03 | 18661 | -0.003 | -0.014 | 0.009 | 0.006 | -0.439 | -0.003 | 6.60E-01 | 18865 |
| 161 | SN_pc_L | -0.005 | -0.020 | 0.011 | 0.008 | -0.581 | -0.004 | 5.62E-01 | 18661 | -0.008 | -0.028 | 0.013 | 0.011 | -0.741 | -0.005 | 4.59E-01 | 18865 |
| 162 | SN_pc_R | -0.006 | -0.027 | 0.014 | 0.011 | -0.612 | -0.004 | 5.40E-01 | 18661 | -0.018 | -0.045 | 0.010 | 0.014 | -1.270 | -0.009 | 2.04E-01 | 18865 |
| 163 | SN_pr_L | -0.011 | -0.033 | 0.010 | 0.011 | -1.042 | -0.008 | 2.97E-01 | 18661 | -0.008 | -0.036 | 0.020 | 0.014 | -0.561 | -0.004 | 5.75E-01 | 18865 |
| 164 | SN_pr_R | -0.012 | -0.036 | 0.012 | 0.012 | -1.002 | -0.007 | 3.16E-01 | 18661 | -0.006 | -0.037 | 0.026 | 0.016 | -0.361 | -0.003 | 7.18E-01 | 18865 |
| 165 | Red_N_L | 0.013 | -0.014 | 0.039 | 0.013 | 0.943 | 0.007 | 3.46E-01 | 18661 | -0.027 | -0.062 | 0.007 | 0.018 | -1.553 | -0.011 | 1.20E-01 | 18865 |
| 166 | Red_N_R | 0.013 | -0.016 | 0.041 | 0.015 | 0.874 | 0.006 | 3.82E-01 | 18661 | -0.019 | -0.057 | 0.019 | 0.019 | -0.989 | -0.007 | 3.23E-01 | 18865 |
| 167 | LC_L | 0.016 | 0.006 | 0.025 | 0.005 | 3.117 | 0.023 | 1.83E-03 | 18661 | 0.003 | -0.010 | 0.015 | 0.007 | 0.381 | 0.003 | 7.03E-01 | 18865 |
| 168 | LC_R | 0.007 | -0.003 | 0.018 | 0.005 | 1.347 | 0.010 | 1.78E-01 | 18661 | 0.005 | -0.009 | 0.019 | 0.007 | 0.716 | 0.005 | 4.74E-01 | 18865 |
| 169 | Raphe_D | 0.012 | -0.026 | 0.050 | 0.019 | 0.622 | 0.005 | 5.34E-01 | 18661 | -0.059 | -0.109 | -0.009 | 0.026 | -2.295 | -0.017 | 2.17E-02 | 18865 |
| 170 | Raphe_M | 0.018 | 0.006 | 0.029 | 0.006 | 3.026 | 0.022 | 2.48E-03 | 18661 | -0.003 | -0.018 | 0.013 | 0.008 | -0.336 | -0.002 | 7.37E-01 | 18865 |

| | | Salad / raw vegetable intake | | | | | | | | Poultry intake | | | | | | | |
|---|---|---|---|---|---|---|---|---|---|---|---|---|---|---|---|---|---|
| Number | AAL 3 region | beta | 95%CI_l | 95%CI_h | se | t | r | p | df.e | beta | 95%CI_l | 95%CI_h | se | t | r | p | df.e |
| 0 | Total brain | -68.496 | -147.064 | 10.071 | 40.084 | -1.709 | -0.012 | 8.75E-02 | 18701 | -148.310 | -295.115 | -1.505 | 74.897 | -1.980 | -0.014 | 4.77E-02 | 18863 |
| 1 | Precentral_L | -1.398 | -4.157 | 1.362 | 1.408 | -0.993 | -0.007 | 3.21E-01 | 18701 | -3.477 | -8.637 | 1.682 | 2.632 | -1.321 | -0.010 | 1.87E-01 | 18863 |
| 2 | Precentral_R | -0.212 | -2.741 | 2.318 | 1.290 | -0.164 | -0.001 | 8.70E-01 | 18701 | -2.975 | -7.703 | 1.754 | 2.412 | -1.233 | -0.009 | 2.18E-01 | 18863 |
| 3 | Frontal_Sup_2_L | -3.487 | -6.403 | -0.571 | 1.488 | -2.344 | -0.017 | 1.91E-02 | 18701 | -3.175 | -8.626 | 2.275 | 2.781 | -1.142 | -0.008 | 2.53E-01 | 18863 |
| 4 | Frontal_Sup_2_R | -2.337 | -5.551 | 0.878 | 1.640 | -1.425 | -0.010 | 1.54E-01 | 18701 | -6.316 | -12.323 | -0.310 | 3.065 | -2.061 | -0.015 | 3.93E-02 | 18863 |
| 5 | Frontal_Mid_2_L | -1.851 | -4.914 | 1.212 | 1.563 | -1.185 | -0.009 | 2.36E-01 | 18701 | -4.395 | -10.120 | 1.329 | 2.920 | -1.505 | -0.011 | 1.32E-01 | 18863 |
| 6 | Frontal_Mid_2_R | -2.177 | -5.641 | 1.287 | 1.767 | -1.232 | -0.009 | 2.18E-01 | 18701 | -9.997 | -16.474 | -3.521 | 3.304 | -3.026 | -0.022 | 2.48E-03 | 18863 |
| 7 | Frontal_Inf_Oper_L | -0.200 | -1.105 | 0.705 | 0.462 | -0.434 | -0.003 | 6.64E-01 | 18701 | -1.180 | -2.871 | 0.511 | 0.863 | -1.368 | -0.010 | 1.71E-01 | 18863 |
| 8 | Frontal_Inf_Oper_R | -0.900 | -2.034 | 0.235 | 0.579 | -1.555 | -0.011 | 1.20E-01 | 18701 | -2.092 | -4.213 | 0.029 | 1.082 | -1.933 | -0.014 | 5.33E-02 | 18863 |
| 9 | Frontal_Inf_Tri_L | -1.225 | -3.146 | 0.695 | 0.980 | -1.251 | -0.009 | 2.11E-01 | 18701 | -3.536 | -7.126 | 0.053 | 1.831 | -1.931 | -0.014 | 5.35E-02 | 18863 |
| 10 | Frontal_Inf_Tri_R | 0.528 | -0.981 | 2.036 | 0.769 | 0.686 | 0.005 | 4.93E-01 | 18701 | -3.345 | -6.164 | -0.525 | 1.438 | -2.325 | -0.017 | 2.01E-02 | 18863 |
| 11 | Frontal_Inf_Orb_2_L | -0.477 | -1.052 | 0.097 | 0.293 | -1.628 | -0.012 | 1.04E-01 | 18701 | -0.872 | -1.946 | 0.201 | 0.548 | -1.592 | -0.012 | 1.11E-01 | 18863 |
| 12 | Frontal_Inf_Orb_2_R | -0.137 | -0.738 | 0.464 | 0.307 | -0.446 | -0.003 | 6.55E-01 | 18701 | -1.172 | -2.295 | -0.050 | 0.573 | -2.047 | -0.015 | 4.06E-02 | 18863 |
| 13 | Rolandic_Oper_L | -0.712 | -1.581 | 0.157 | 0.443 | -1.605 | -0.012 | 1.08E-01 | 18701 | -1.528 | -3.152 | 0.096 | 0.828 | -1.844 | -0.013 | 6.52E-02 | 18863 |
| 14 | Rolandic_Oper_R | -0.382 | -1.627 | 0.862 | 0.635 | -0.602 | -0.004 | 5.47E-01 | 18701 | -2.987 | -5.311 | -0.664 | 1.185 | -2.520 | -0.018 | 1.17E-02 | 18863 |
| 15 | Supp_Motor_Area_L | -1.381 | -3.448 | 0.686 | 1.055 | -1.310 | -0.010 | 1.90E-01 | 18701 | -3.304 | -7.165 | 0.557 | 1.970 | -1.677 | -0.012 | 9.35E-02 | 18863 |
| 16 | Supp_Motor_Area_R | -0.314 | -2.596 | 1.968 | 1.164 | -0.270 | -0.002 | 7.87E-01 | 18701 | -1.632 | -5.898 | 2.633 | 2.176 | -0.750 | -0.005 | 4.53E-01 | 18863 |
| 17 | Olfactory_L | -0.339 | -0.646 | -0.032 | 0.157 | -2.165 | -0.016 | 3.04E-02 | 18701 | 0.185 | -0.389 | 0.759 | 0.293 | 0.633 | 0.005 | 5.27E-01 | 18863 |
| 18 | Olfactory_R | -0.383 | -0.692 | -0.075 | 0.157 | -2.440 | -0.018 | 1.47E-02 | 18701 | -0.008 | -0.584 | 0.567 | 0.294 | -0.028 | 0.000 | 9.78E-01 | 18863 |
| 19 | Frontal_Sup_Medial_L | -2.508 | -4.445 | -0.571 | 0.988 | -2.538 | -0.019 | 1.11E-02 | 18701 | -6.506 | -10.125 | -2.887 | 1.846 | -3.524 | -0.026 | 4.26E-04 | 18863 |
| 20 | Frontal_Sup_Medial_R | -1.418 | -2.828 | -0.008 | 0.719 | -1.972 | -0.014 | 4.87E-02 | 18701 | -3.760 | -6.394 | -1.127 | 1.343 | -2.799 | -0.020 | 5.13E-03 | 18863 |
| 21 | Frontal_Med_Orb_L | -1.253 | -1.976 | -0.529 | 0.369 | -3.393 | -0.025 | 6.92E-04 | 18701 | -0.752 | -2.104 | 0.601 | 0.690 | -1.089 | -0.008 | 2.76E-01 | 18863 |
| 22 | Frontal_Med_Orb_R | -1.519 | -2.408 | -0.630 | 0.454 | -3.348 | -0.024 | 8.15E-04 | 18701 | -1.929 | -3.591 | -0.268 | 0.848 | -2.276 | -0.017 | 2.29E-02 | 18863 |
| 23 | Rectus_L | -0.826 | -1.576 | -0.077 | 0.382 | -2.161 | -0.016 | 3.07E-02 | 18701 | -1.402 | -2.802 | -0.003 | 0.714 | -1.964 | -0.014 | 4.96E-02 | 18863 |
| 24 | Rectus_R | -0.622 | -1.315 | 0.071 | 0.354 | -1.758 | -0.013 | 7.87E-02 | 18701 | -1.526 | -2.821 | -0.232 | 0.661 | -2.311 | -0.017 | 2.09E-02 | 18863 |
| 25 | OFCmed_L | -0.531 | -0.966 | -0.097 | 0.222 | -2.397 | -0.018 | 1.66E-02 | 18701 | -1.055 | -1.866 | -0.244 | 0.414 | -2.549 | -0.019 | 1.08E-02 | 18863 |
| 26 | OFCmed_R | -0.787 | -1.293 | -0.281 | 0.258 | -3.048 | -0.022 | 2.31E-03 | 18701 | -0.870 | -1.815 | 0.076 | 0.482 | -1.803 | -0.013 | 7.14E-02 | 18863 |
| 27 | OFCant_L | -0.717 | -1.219 | -0.215 | 0.256 | -2.798 | -0.020 | 5.15E-03 | 18701 | -0.342 | -1.280 | 0.596 | 0.478 | -0.715 | -0.005 | 4.75E-01 | 18863 |
| 28 | OFCant_R | -0.897 | -1.560 | -0.234 | 0.338 | -2.651 | -0.019 | 8.04E-03 | 18701 | -1.074 | -2.312 | 0.165 | 0.632 | -1.700 | -0.012 | 8.92E-02 | 18863 |

| | | | | | | | | | | | | | | | | | |
|---|---|---|---|---|---|---|---|---|---|---|---|---|---|---|---|---|---|
| 29 | OFCpost_L | -0.812 | -1.440 | -0.183 | 0.321 | -2.531 | -0.019 | 1.14E-02 | 18701 | -0.057 | -1.231 | 1.117 | 0.599 | -0.096 | -0.001 | 9.24E-01 | 18863 |
| 30 | OFCpost_R | -0.884 | -1.465 | -0.304 | 0.296 | -2.985 | -0.022 | 2.84E-03 | 18701 | -0.645 | -1.730 | 0.440 | 0.554 | -1.165 | -0.008 | 2.44E-01 | 18863 |
| 31 | OFClat_L | -0.154 | -0.367 | 0.058 | 0.108 | -1.427 | -0.010 | 1.54E-01 | 18701 | -0.545 | -0.941 | -0.149 | 0.202 | -2.697 | -0.020 | 7.00E-03 | 18863 |
| 32 | OFClat_R | -0.110 | -0.289 | 0.069 | 0.091 | -1.202 | -0.009 | 2.29E-01 | 18701 | -0.511 | -0.845 | -0.176 | 0.171 | -2.991 | -0.022 | 2.79E-03 | 18863 |
| 33 | Insula_L | -1.714 | -3.658 | 0.230 | 0.992 | -1.728 | -0.013 | 8.40E-02 | 18701 | -2.125 | -5.757 | 1.506 | 1.853 | -1.147 | -0.008 | 2.51E-01 | 18863 |
| 34 | Insula_R | -1.254 | -3.149 | 0.641 | 0.967 | -1.297 | -0.009 | 1.95E-01 | 18701 | -2.963 | -6.502 | 0.576 | 1.805 | -1.641 | -0.012 | 1.01E-01 | 18863 |
| 37 | Cingulate_Mid_L | 0.790 | -0.856 | 2.435 | 0.839 | 0.941 | 0.007 | 3.47E-01 | 18701 | -4.683 | -7.755 | -1.611 | 1.567 | -2.988 | -0.022 | 2.81E-03 | 18863 |
| 38 | Cingulate_Mid_R | 0.269 | -1.484 | 2.022 | 0.894 | 0.301 | 0.002 | 7.63E-01 | 18701 | -5.824 | -9.098 | -2.550 | 1.671 | -3.486 | -0.025 | 4.91E-04 | 18863 |
| 39 | Cingulate_Post_L | -0.394 | -0.896 | 0.108 | 0.256 | -1.539 | -0.011 | 1.24E-01 | 18701 | -0.906 | -1.843 | 0.031 | 0.478 | -1.894 | -0.014 | 5.82E-02 | 18863 |
| 40 | Cingulate_Post_R | -0.256 | -0.535 | 0.022 | 0.142 | -1.805 | -0.013 | 7.10E-02 | 18701 | -0.298 | -0.818 | 0.222 | 0.265 | -1.124 | -0.008 | 2.61E-01 | 18863 |
| 41 | Hippocampus_L | 0.320 | -0.492 | 1.131 | 0.414 | 0.772 | 0.006 | 4.40E-01 | 18701 | -1.671 | -3.187 | -0.155 | 0.774 | -2.160 | -0.016 | 3.08E-02 | 18863 |
| 42 | Hippocampus_R | 0.443 | -0.399 | 1.285 | 0.430 | 1.031 | 0.008 | 3.03E-01 | 18701 | -0.835 | -2.407 | 0.738 | 0.802 | -1.040 | -0.008 | 2.98E-01 | 18863 |
| 43 | ParaHippocampal_L | 0.258 | -0.462 | 0.979 | 0.368 | 0.703 | 0.005 | 4.82E-01 | 18701 | -1.649 | -2.996 | -0.302 | 0.687 | -2.399 | -0.017 | 1.64E-02 | 18863 |
| 44 | ParaHippocampal_R | 0.521 | -0.389 | 1.432 | 0.464 | 1.122 | 0.008 | 2.62E-01 | 18701 | -1.919 | -3.620 | -0.218 | 0.868 | -2.211 | -0.016 | 2.71E-02 | 18863 |
| 45 | Amygdala_L | -0.127 | -0.407 | 0.154 | 0.143 | -0.886 | -0.006 | 3.76E-01 | 18701 | -0.216 | -0.739 | 0.308 | 0.267 | -0.807 | -0.006 | 4.20E-01 | 18863 |
| 46 | Amygdala_R | -0.192 | -0.479 | 0.095 | 0.146 | -1.313 | -0.010 | 1.89E-01 | 18701 | -0.195 | -0.731 | 0.340 | 0.273 | -0.714 | -0.005 | 4.75E-01 | 18863 |
| 47 | Calcarine_L | -0.508 | -3.181 | 2.164 | 1.363 | -0.373 | -0.003 | 7.09E-01 | 18701 | 3.198 | -1.797 | 8.192 | 2.548 | 1.255 | 0.009 | 2.10E-01 | 18863 |
| 48 | Calcarine_R | -0.456 | -2.689 | 1.777 | 1.139 | -0.400 | -0.003 | 6.89E-01 | 18701 | 0.322 | -3.852 | 4.495 | 2.129 | 0.151 | 0.001 | 8.80E-01 | 18863 |
| 49 | Cuneus_L | 0.003 | -1.676 | 1.682 | 0.856 | 0.004 | 0.000 | 9.97E-01 | 18701 | 2.164 | -0.972 | 5.300 | 1.600 | 1.353 | 0.010 | 1.76E-01 | 18863 |
| 50 | Cuneus_R | -0.450 | -1.734 | 0.834 | 0.655 | -0.687 | -0.005 | 4.92E-01 | 18701 | -0.716 | -3.114 | 1.683 | 1.224 | -0.585 | -0.004 | 5.59E-01 | 18863 |
| 51 | Lingual_L | 0.509 | -1.101 | 2.119 | 0.821 | 0.619 | 0.005 | 5.36E-01 | 18701 | 0.494 | -2.513 | 3.501 | 1.534 | 0.322 | 0.002 | 7.47E-01 | 18863 |
| 52 | Lingual_R | -0.214 | -1.899 | 1.471 | 0.860 | -0.249 | -0.002 | 8.03E-01 | 18701 | -0.037 | -3.185 | 3.112 | 1.606 | -0.023 | 0.000 | 9.82E-01 | 18863 |
| 53 | Occipital_Sup_L | 0.251 | -0.856 | 1.357 | 0.564 | 0.444 | 0.003 | 6.57E-01 | 18701 | -0.644 | -2.712 | 1.423 | 1.055 | -0.611 | -0.004 | 5.41E-01 | 18863 |
| 54 | Occipital_Sup_R | -0.232 | -1.431 | 0.968 | 0.612 | -0.379 | -0.003 | 7.05E-01 | 18701 | -0.964 | -3.205 | 1.276 | 1.143 | -0.844 | -0.006 | 3.99E-01 | 18863 |
| 55 | Occipital_Mid_L | -0.758 | -3.349 | 1.833 | 1.322 | -0.574 | -0.004 | 5.66E-01 | 18701 | 0.741 | -4.101 | 5.583 | 2.470 | 0.300 | 0.002 | 7.64E-01 | 18863 |
| 56 | Occipital_Mid_R | -1.931 | -3.735 | -0.128 | 0.920 | -2.099 | -0.015 | 3.58E-02 | 18701 | 1.394 | -1.976 | 4.765 | 1.720 | 0.811 | 0.006 | 4.17E-01 | 18863 |
| 57 | Occipital_Inf_L | -1.084 | -1.943 | -0.225 | 0.438 | -2.472 | -0.018 | 1.34E-02 | 18701 | 1.373 | -0.233 | 2.980 | 0.820 | 1.676 | 0.012 | 9.38E-02 | 18863 |
| 58 | Occipital_Inf_R | -1.734 | -2.728 | -0.740 | 0.507 | -3.419 | -0.025 | 6.31E-04 | 18701 | 1.817 | -0.040 | 3.674 | 0.947 | 1.918 | 0.014 | 5.51E-02 | 18863 |
| 59 | Fusiform_L | -0.098 | -1.982 | 1.786 | 0.961 | -0.102 | -0.001 | 9.19E-01 | 18701 | -0.632 | -4.151 | 2.886 | 1.795 | -0.352 | -0.003 | 7.25E-01 | 18863 |
| 60 | Fusiform_R | -0.054 | -2.122 | 2.015 | 1.055 | -0.051 | 0.000 | 9.59E-01 | 18701 | -1.312 | -5.179 | 2.554 | 1.973 | -0.665 | -0.005 | 5.06E-01 | 18863 |
| 61 | Postcentral_L | 1.510 | -1.324 | 4.343 | 1.445 | 1.044 | 0.008 | 2.96E-01 | 18701 | -9.734 | -15.027 | -4.441 | 2.700 | -3.605 | -0.026 | 3.13E-04 | 18863 |
| 62 | Postcentral_R | 3.036 | 0.134 | 5.939 | 1.481 | 2.050 | 0.015 | 4.04E-02 | 18701 | -7.986 | -13.407 | -2.564 | 2.766 | -2.887 | -0.021 | 3.89E-03 | 18863 |
| 63 | Parietal_Sup_L | -0.467 | -2.100 | 1.165 | 0.833 | -0.561 | -0.004 | 5.75E-01 | 18701 | -2.713 | -5.762 | 0.337 | 1.556 | -1.744 | -0.013 | 8.12E-02 | 18863 |
| 64 | Parietal_Sup_R | 0.172 | -1.275 | 1.620 | 0.738 | 0.233 | 0.002 | 8.16E-01 | 18701 | -1.105 | -3.809 | 1.599 | 1.380 | -0.801 | -0.006 | 4.23E-01 | 18863 |
| 65 | Parietal_Inf_L | -1.631 | -3.768 | 0.506 | 1.090 | -1.496 | -0.011 | 1.35E-01 | 18701 | -3.189 | -7.182 | 0.804 | 2.037 | -1.565 | -0.011 | 1.17E-01 | 18863 |
| 66 | Parietal_Inf_R | -0.177 | -1.562 | 1.207 | 0.706 | -0.251 | -0.002 | 8.02E-01 | 18701 | -2.139 | -4.727 | 0.450 | 1.321 | -1.620 | -0.012 | 1.05E-01 | 18863 |
| 67 | SupraMarginal_L | -0.891 | -2.143 | 0.361 | 0.639 | -1.395 | -0.010 | 1.63E-01 | 18701 | -4.162 | -6.501 | -1.822 | 1.194 | -3.487 | -0.025 | 4.90E-04 | 18863 |
| 68 | SupraMarginal_R | -0.327 | -2.100 | 1.447 | 0.905 | -0.361 | -0.003 | 7.18E-01 | 18701 | -5.384 | -8.697 | -2.072 | 1.690 | -3.186 | -0.023 | 1.44E-03 | 18863 |
| 69 | Angular_L | -0.646 | -1.876 | 0.585 | 0.628 | -1.028 | -0.008 | 3.04E-01 | 18701 | -0.459 | -2.758 | 1.841 | 1.173 | -0.391 | -0.003 | 6.96E-01 | 18863 |
| 70 | Angular_R | -0.206 | -1.855 | 1.443 | 0.841 | -0.244 | -0.002 | 8.07E-01 | 18701 | 0.030 | -3.051 | 3.112 | 1.572 | 0.019 | 0.000 | 9.85E-01 | 18863 |
| 71 | Precuneus_L | -0.684 | -3.685 | 2.318 | 1.531 | -0.447 | -0.003 | 6.55E-01 | 18701 | -3.888 | -9.492 | 1.716 | 2.859 | -1.360 | -0.010 | 1.74E-01 | 18863 |
| 72 | Precuneus_R | -1.653 | -4.326 | 1.021 | 1.364 | -1.212 | -0.009 | 2.26E-01 | 18701 | -2.945 | -7.937 | 2.046 | 2.547 | -1.157 | -0.008 | 2.47E-01 | 18863 |
| 73 | Paracentral_Lobule_L | 0.557 | -0.816 | 1.930 | 0.700 | 0.795 | 0.006 | 4.27E-01 | 18701 | -2.086 | -4.652 | 0.479 | 1.309 | -1.594 | -0.012 | 1.11E-01 | 18863 |
| 74 | Paracentral_Lobule_R | 0.249 | -0.735 | 1.233 | 0.502 | 0.496 | 0.004 | 6.20E-01 | 18701 | -0.706 | -2.543 | 1.131 | 0.937 | -0.753 | -0.005 | 4.51E-01 | 18863 |
| 75 | Caudate_L | -0.167 | -1.265 | 0.930 | 0.560 | -0.299 | -0.002 | 7.65E-01 | 18701 | -1.661 | -3.710 | 0.388 | 1.045 | -1.589 | -0.012 | 1.12E-01 | 18863 |
| 76 | Caudate_R | -0.529 | -1.772 | 0.713 | 0.634 | -0.835 | -0.006 | 4.04E-01 | 18701 | -1.454 | -3.776 | 0.868 | 1.185 | -1.228 | -0.009 | 2.20E-01 | 18863 |
| 77 | Putamen_L | -1.129 | -2.794 | 0.535 | 0.849 | -1.330 | -0.010 | 1.84E-01 | 18701 | 0.542 | -2.570 | 3.654 | 1.588 | 0.341 | 0.002 | 7.33E-01 | 18863 |
| 78 | Putamen_R | -1.251 | -2.947 | 0.445 | 0.865 | -1.446 | -0.011 | 1.48E-01 | 18701 | -0.232 | -3.402 | 2.939 | 1.617 | -0.143 | -0.001 | 8.86E-01 | 18863 |
| 79 | Pallidum_L | 0.050 | -0.098 | 0.198 | 0.076 | 0.658 | 0.005 | 5.10E-01 | 18701 | -0.073 | -0.350 | 0.204 | 0.141 | -0.517 | -0.004 | 6.05E-01 | 18863 |

| | | | | | | | | | | | | | | | | | |
|---|---|---|---|---|---|---|---|---|---|---|---|---|---|---|---|---|---|
| 80 | Pallidum_R | -0.001 | -0.164 | 0.161 | 0.083 | -0.018 | 0.000 | 9.86E-01 | 18701 | -0.072 | -0.376 | 0.232 | 0.155 | -0.465 | -0.003 | 6.42E-01 | 18863 |
| 83 | Heschl_L | -0.313 | -0.613 | -0.012 | 0.153 | -2.041 | -0.015 | 4.12E-02 | 18701 | -0.388 | -0.949 | 0.173 | 0.286 | -1.357 | -0.010 | 1.75E-01 | 18863 |
| 84 | Heschl_R | -0.112 | -0.447 | 0.224 | 0.171 | -0.652 | -0.005 | 5.15E-01 | 18701 | -0.484 | -1.111 | 0.142 | 0.320 | -1.515 | -0.011 | 1.30E-01 | 18863 |
| 85 | Temporal_Sup_L | -2.957 | -5.123 | -0.790 | 1.105 | -2.675 | -0.020 | 7.48E-03 | 18701 | -5.829 | -9.875 | -1.782 | 2.065 | -2.823 | -0.021 | 4.76E-03 | 18863 |
| 86 | Temporal_Sup_R | -1.863 | -4.262 | 0.535 | 1.224 | -1.523 | -0.011 | 1.28E-01 | 18701 | -6.876 | -11.356 | -2.395 | 2.286 | -3.008 | -0.022 | 2.63E-03 | 18863 |
| 87 | Temporal_Pole_Sup_L | -1.280 | -2.306 | -0.253 | 0.524 | -2.444 | -0.018 | 1.45E-02 | 18701 | -1.308 | -3.227 | 0.610 | 0.979 | -1.337 | -0.010 | 1.81E-01 | 18863 |
| 88 | Temporal_Pole_Sup_R | -1.106 | -2.186 | -0.026 | 0.551 | -2.008 | -0.015 | 4.46E-02 | 18701 | -1.510 | -3.529 | 0.510 | 1.030 | -1.465 | -0.011 | 1.43E-01 | 18863 |
| 89 | Temporal_Mid_L | -2.179 | -5.791 | 1.434 | 1.843 | -1.182 | -0.009 | 2.37E-01 | 18701 | -6.195 | -12.944 | 0.555 | 3.444 | -1.799 | -0.013 | 7.21E-02 | 18863 |
| 90 | Temporal_Mid_R | -2.706 | -5.915 | 0.502 | 1.637 | -1.653 | -0.012 | 9.83E-02 | 18701 | -3.720 | -9.714 | 2.274 | 3.058 | -1.216 | -0.009 | 2.24E-01 | 18863 |
| 91 | Temporal_Pole_Mid_L | -0.185 | -1.048 | 0.678 | 0.440 | -0.421 | -0.003 | 6.74E-01 | 18701 | -1.599 | -3.212 | 0.014 | 0.823 | -1.944 | -0.014 | 5.20E-02 | 18863 |
| 92 | Temporal_Pole_Mid_R | -0.533 | -1.580 | 0.514 | 0.534 | -0.998 | -0.007 | 3.19E-01 | 18701 | -1.545 | -3.503 | 0.414 | 0.999 | -1.546 | -0.011 | 1.22E-01 | 18863 |
| 93 | Temporal_Inf_L | -0.860 | -3.343 | 1.624 | 1.267 | -0.679 | -0.005 | 4.97E-01 | 18701 | -2.116 | -6.757 | 2.525 | 2.368 | -0.894 | -0.007 | 3.71E-01 | 18863 |
| 94 | Temporal_Inf_R | -3.086 | -5.795 | -0.377 | 1.382 | -2.233 | -0.016 | 2.55E-02 | 18701 | -0.655 | -5.715 | 4.405 | 2.582 | -0.254 | -0.002 | 8.00E-01 | 18863 |
| 95 | Cerebellum_Crus1_L | -1.475 | -4.426 | 1.476 | 1.505 | -0.980 | -0.007 | 3.27E-01 | 18701 | 5.928 | 0.416 | 11.440 | 2.812 | 2.108 | 0.015 | 3.50E-02 | 18863 |
| 96 | Cerebellum_Crus1_R | -0.529 | -3.538 | 2.479 | 1.535 | -0.345 | -0.003 | 7.30E-01 | 18701 | 6.256 | 0.637 | 11.875 | 2.867 | 2.182 | 0.016 | 2.91E-02 | 18863 |
| 97 | Cerebellum_Crus2_L | -1.499 | -3.840 | 0.842 | 1.194 | -1.255 | -0.009 | 2.09E-01 | 18701 | 0.039 | -4.333 | 4.411 | 2.231 | 0.017 | 0.000 | 9.86E-01 | 18863 |
| 98 | Cerebellum_Crus2_R | 0.185 | -1.978 | 2.348 | 1.104 | 0.168 | 0.001 | 8.67E-01 | 18701 | 0.947 | -3.094 | 4.988 | 2.062 | 0.459 | 0.003 | 6.46E-01 | 18863 |
| 99 | Cerebellum_3_L | 0.082 | -0.075 | 0.239 | 0.080 | 1.020 | 0.007 | 3.08E-01 | 18701 | 0.568 | 0.275 | 0.861 | 0.150 | 3.797 | 0.028 | 1.47E-04 | 18863 |
| 100 | Cerebellum_3_R | 0.046 | -0.166 | 0.257 | 0.108 | 0.423 | 0.003 | 6.73E-01 | 18701 | 0.536 | 0.141 | 0.932 | 0.202 | 2.657 | 0.019 | 7.88E-03 | 18863 |
| 101 | Cerebellum_4_5_L | 0.623 | -0.646 | 1.892 | 0.647 | 0.962 | 0.007 | 3.36E-01 | 18701 | 2.660 | 0.288 | 5.032 | 1.210 | 2.198 | 0.016 | 2.80E-02 | 18863 |
| 102 | Cerebellum_4_5_R | 0.371 | -0.663 | 1.405 | 0.528 | 0.703 | 0.005 | 4.82E-01 | 18701 | 0.933 | -0.999 | 2.866 | 0.986 | 0.947 | 0.007 | 3.44E-01 | 18863 |
| 103 | Cerebellum_6_L | 0.720 | -1.474 | 2.915 | 1.120 | 0.643 | 0.005 | 5.20E-01 | 18701 | 2.891 | -1.212 | 6.994 | 2.093 | 1.381 | 0.010 | 1.67E-01 | 18863 |
| 104 | Cerebellum_6_R | 0.813 | -1.499 | 3.125 | 1.179 | 0.689 | 0.005 | 4.91E-01 | 18701 | 4.090 | -0.231 | 8.412 | 2.205 | 1.855 | 0.014 | 6.36E-02 | 18863 |
| 105 | Cerebellum_7b_L | -0.385 | -1.100 | 0.331 | 0.365 | -1.054 | -0.008 | 2.92E-01 | 18701 | 0.084 | -1.252 | 1.421 | 0.682 | 0.124 | 0.001 | 9.01E-01 | 18863 |
| 106 | Cerebellum_7b_R | -0.012 | -0.721 | 0.697 | 0.362 | -0.034 | 0.000 | 9.73E-01 | 18701 | -0.737 | -2.061 | 0.587 | 0.676 | -1.091 | -0.008 | 2.75E-01 | 18863 |
| 107 | Cerebellum_8_L | -0.973 | -3.244 | 1.298 | 1.159 | -0.840 | -0.006 | 4.01E-01 | 18701 | 2.254 | -1.990 | 6.499 | 2.166 | 1.041 | 0.008 | 2.98E-01 | 18863 |
| 108 | Cerebellum_8_R | -2.084 | -5.040 | 0.872 | 1.508 | -1.382 | -0.010 | 1.67E-01 | 18701 | -2.102 | -7.627 | 3.422 | 2.819 | -0.746 | -0.005 | 4.56E-01 | 18863 |
| 109 | Cerebellum_9_L | -0.458 | -1.742 | 0.825 | 0.655 | -0.700 | -0.005 | 4.84E-01 | 18701 | 3.481 | 1.082 | 5.879 | 1.224 | 2.845 | 0.021 | 4.45E-03 | 18863 |
| 110 | Cerebellum_9_R | -0.500 | -1.762 | 0.762 | 0.644 | -0.777 | -0.006 | 4.37E-01 | 18701 | 3.265 | 0.907 | 5.623 | 1.203 | 2.714 | 0.020 | 6.65E-03 | 18863 |
| 111 | Cerebellum_10_L | -0.129 | -0.219 | -0.038 | 0.046 | -2.773 | -0.020 | 5.56E-03 | 18701 | 0.222 | 0.052 | 0.392 | 0.087 | 2.564 | 0.019 | 1.04E-02 | 18863 |
| 112 | Cerebellum_10_R | -0.044 | -0.139 | 0.050 | 0.048 | -0.915 | -0.007 | 3.60E-01 | 18701 | 0.204 | 0.027 | 0.381 | 0.090 | 2.264 | 0.016 | 2.36E-02 | 18863 |
| 113 | Vermis_1_2 | 0.038 | -0.021 | 0.098 | 0.030 | 1.275 | 0.009 | 2.02E-01 | 18701 | 0.193 | 0.082 | 0.303 | 0.056 | 3.423 | 0.025 | 6.20E-04 | 18863 |
| 114 | Vermis_3 | 0.073 | -0.162 | 0.308 | 0.120 | 0.608 | 0.004 | 5.43E-01 | 18701 | 0.585 | 0.145 | 1.024 | 0.224 | 2.608 | 0.019 | 9.12E-03 | 18863 |
| 115 | Vermis_4_5 | 0.481 | -0.211 | 1.172 | 0.353 | 1.363 | 0.010 | 1.73E-01 | 18701 | 1.070 | -0.222 | 2.362 | 0.659 | 1.624 | 0.012 | 1.04E-01 | 18863 |
| 116 | Vermis_6 | 0.214 | -0.171 | 0.599 | 0.196 | 1.089 | 0.008 | 2.76E-01 | 18701 | 1.118 | 0.398 | 1.837 | 0.367 | 3.046 | 0.022 | 2.33E-03 | 18863 |
| 117 | Vermis_7 | 0.065 | -0.153 | 0.283 | 0.111 | 0.585 | 0.004 | 5.59E-01 | 18701 | 0.615 | 0.208 | 1.022 | 0.208 | 2.964 | 0.022 | 3.04E-03 | 18863 |
| 118 | Vermis_8 | 0.254 | -0.103 | 0.612 | 0.182 | 1.393 | 0.010 | 1.64E-01 | 18701 | 1.391 | 0.723 | 2.059 | 0.341 | 4.079 | 0.030 | 4.54E-05 | 18863 |
| 119 | Vermis_9 | 0.162 | -0.129 | 0.452 | 0.148 | 1.092 | 0.008 | 2.75E-01 | 18701 | 0.928 | 0.386 | 1.470 | 0.277 | 3.354 | 0.024 | 7.97E-04 | 18863 |
| 120 | Vermis_10 | 0.043 | -0.038 | 0.124 | 0.041 | 1.050 | 0.008 | 2.94E-01 | 18701 | 0.351 | 0.200 | 0.502 | 0.077 | 4.555 | 0.033 | 5.28E-06 | 18863 |
| 121 | Thal_AV_L | 0.000 | -0.032 | 0.033 | 0.016 | 0.022 | 0.000 | 9.82E-01 | 18701 | -0.050 | -0.110 | 0.011 | 0.031 | -1.613 | -0.012 | 1.07E-01 | 18863 |
| 122 | Thal_AV_R | 0.027 | -0.019 | 0.073 | 0.024 | 1.133 | 0.008 | 2.57E-01 | 18701 | -0.082 | -0.168 | 0.004 | 0.044 | -1.863 | -0.014 | 6.25E-02 | 18863 |
| 123 | Thal_LP_L | -0.013 | -0.049 | 0.023 | 0.018 | -0.695 | -0.005 | 4.87E-01 | 18701 | -0.129 | -0.196 | -0.062 | 0.034 | -3.764 | -0.027 | 1.68E-04 | 18863 |
| 124 | Thal_LP_R | 0.004 | -0.043 | 0.050 | 0.024 | 0.156 | 0.001 | 8.76E-01 | 18701 | -0.129 | -0.216 | -0.042 | 0.045 | -2.894 | -0.021 | 3.80E-03 | 18863 |
| 125 | Thal_VA_L | -0.001 | -0.039 | 0.037 | 0.020 | -0.041 | 0.000 | 9.67E-01 | 18701 | -0.020 | -0.091 | 0.051 | 0.036 | -0.548 | -0.004 | 5.83E-01 | 18863 |
| 126 | Thal_VA_R | -0.005 | -0.052 | 0.042 | 0.024 | -0.219 | -0.002 | 8.27E-01 | 18701 | -0.036 | -0.124 | 0.052 | 0.045 | -0.802 | -0.006 | 4.22E-01 | 18863 |
| 127 | Thal_VL_L | -0.081 | -0.190 | 0.028 | 0.056 | -1.453 | -0.011 | 1.46E-01 | 18701 | -0.272 | -0.476 | -0.069 | 0.104 | -2.620 | -0.019 | 8.81E-03 | 18863 |
| 128 | Thal_VL_R | -0.050 | -0.195 | 0.094 | 0.074 | -0.685 | -0.005 | 4.94E-01 | 18701 | -0.469 | -0.739 | -0.199 | 0.138 | -3.403 | -0.025 | 6.69E-04 | 18863 |
| 129 | Thal_VPL_L | -0.072 | -0.202 | 0.057 | 0.066 | -1.091 | -0.008 | 2.75E-01 | 18701 | -0.217 | -0.459 | 0.025 | 0.124 | -1.755 | -0.013 | 7.92E-02 | 18863 |
| 130 | Thal_VPL_R | -0.045 | -0.181 | 0.091 | 0.070 | -0.649 | -0.005 | 5.16E-01 | 18701 | -0.335 | -0.589 | -0.080 | 0.130 | -2.576 | -0.019 | 1.00E-02 | 18863 |

| | | | | | | | | | | | | | | | | | |
|---|---|---|---|---|---|---|---|---|---|---|---|---|---|---|---|---|---|
| 131 | Thal_IL_L | -0.016 | -0.072 | 0.040 | 0.029 | -0.549 | -0.004 | 5.83E-01 | 18701 | -0.167 | -0.272 | -0.062 | 0.053 | -3.119 | -0.023 | 1.82E-03 | 18863 |
| 132 | Thal_IL_R | -0.001 | -0.064 | 0.063 | 0.032 | -0.017 | 0.000 | 9.86E-01 | 18701 | -0.180 | -0.299 | -0.061 | 0.061 | -2.974 | -0.022 | 2.95E-03 | 18863 |
| 133 | Thal_Re_L | 0.000 | 0.000 | 0.000 | 0.000 | 1.001 | 0.007 | 3.17E-01 | 18701 | 0.000 | -0.001 | 0.000 | 0.000 | -1.139 | -0.008 | 2.55E-01 | 18863 |
| 134 | Thal_Re_R | 0.000 | -0.002 | 0.002 | 0.001 | 0.390 | 0.003 | 6.97E-01 | 18701 | -0.003 | -0.007 | 0.000 | 0.002 | -1.861 | -0.014 | 6.28E-02 | 18863 |
| 135 | Thal_MDm_L | -0.137 | -0.553 | 0.278 | 0.212 | -0.647 | -0.005 | 5.18E-01 | 18701 | -1.353 | -2.129 | -0.577 | 0.396 | -3.417 | -0.025 | 6.35E-04 | 18863 |
| 136 | Thal_MDm_R | -0.011 | -0.493 | 0.472 | 0.246 | -0.043 | 0.000 | 9.66E-01 | 18701 | -1.150 | -2.052 | -0.248 | 0.460 | -2.498 | -0.018 | 1.25E-02 | 18863 |
| 137 | Thal_MDl_L | -0.033 | -0.104 | 0.039 | 0.037 | -0.887 | -0.006 | 3.75E-01 | 18701 | -0.278 | -0.412 | -0.144 | 0.069 | -4.054 | -0.030 | 5.05E-05 | 18863 |
| 138 | Thal_MDl_R | -0.003 | -0.085 | 0.078 | 0.042 | -0.084 | -0.001 | 9.33E-01 | 18701 | -0.241 | -0.394 | -0.088 | 0.078 | -3.095 | -0.023 | 1.97E-03 | 18863 |
| 139 | Thal_LGN_L | 0.000 | -0.011 | 0.010 | 0.006 | -0.082 | -0.001 | 9.35E-01 | 18701 | -0.034 | -0.054 | -0.013 | 0.010 | -3.252 | -0.024 | 1.15E-03 | 18863 |
| 140 | Thal_LGN_R | -0.007 | -0.030 | 0.016 | 0.012 | -0.599 | -0.004 | 5.49E-01 | 18701 | -0.090 | -0.133 | -0.047 | 0.022 | -4.068 | -0.030 | 4.76E-05 | 18863 |
| 141 | Thal_MGN_L | -0.021 | -0.041 | -0.002 | 0.010 | -2.154 | -0.016 | 3.12E-02 | 18701 | -0.049 | -0.086 | -0.013 | 0.019 | -2.654 | -0.019 | 7.97E-03 | 18863 |
| 142 | Thal_MGN_R | -0.011 | -0.032 | 0.010 | 0.011 | -1.046 | -0.008 | 2.95E-01 | 18701 | -0.066 | -0.106 | -0.026 | 0.020 | -3.271 | -0.024 | 1.07E-03 | 18863 |
| 143 | Thal_PuA_L | -0.015 | -0.038 | 0.008 | 0.012 | -1.243 | -0.009 | 2.14E-01 | 18701 | -0.064 | -0.107 | -0.021 | 0.022 | -2.895 | -0.021 | 3.80E-03 | 18863 |
| 144 | Thal_PuA_R | -0.019 | -0.066 | 0.029 | 0.024 | -0.764 | -0.006 | 4.45E-01 | 18701 | -0.139 | -0.229 | -0.050 | 0.046 | -3.054 | -0.022 | 2.26E-03 | 18863 |
| 145 | Thal_PuM_L | -0.115 | -0.573 | 0.342 | 0.233 | -0.494 | -0.004 | 6.21E-01 | 18701 | -1.290 | -2.144 | -0.435 | 0.436 | -2.959 | -0.022 | 3.09E-03 | 18863 |
| 146 | Thal_PuM_R | -0.083 | -0.653 | 0.487 | 0.291 | -0.285 | -0.002 | 7.76E-01 | 18701 | -1.362 | -2.427 | -0.297 | 0.543 | -2.508 | -0.018 | 1.22E-02 | 18863 |
| 147 | Thal_PuL_L | -0.020 | -0.104 | 0.063 | 0.043 | -0.472 | -0.003 | 6.37E-01 | 18701 | -0.244 | -0.400 | -0.088 | 0.080 | -3.070 | -0.022 | 2.14E-03 | 18863 |
| 148 | Thal_PuL_R | -0.020 | -0.153 | 0.113 | 0.068 | -0.299 | -0.002 | 7.65E-01 | 18701 | -0.333 | -0.581 | -0.085 | 0.127 | -2.633 | -0.019 | 8.46E-03 | 18863 |
| 149 | Thal_PuI_L | -0.002 | -0.025 | 0.020 | 0.012 | -0.209 | -0.002 | 8.34E-01 | 18701 | -0.054 | -0.097 | -0.012 | 0.022 | -2.515 | -0.018 | 1.19E-02 | 18863 |
| 150 | Thal_PuI_R | 0.003 | -0.027 | 0.033 | 0.015 | 0.182 | 0.001 | 8.55E-01 | 18701 | -0.059 | -0.116 | -0.003 | 0.029 | -2.078 | -0.015 | 3.78E-02 | 18863 |
| 151 | ACC_sub_L | -0.436 | -0.697 | -0.174 | 0.133 | -3.265 | -0.024 | 1.10E-03 | 18701 | -0.275 | -0.764 | 0.214 | 0.250 | -1.103 | -0.008 | 2.70E-01 | 18863 |
| 152 | ACC_sub_R | -0.337 | -0.581 | -0.093 | 0.125 | -2.704 | -0.020 | 6.87E-03 | 18701 | -0.386 | -0.843 | 0.071 | 0.233 | -1.654 | -0.012 | 9.80E-02 | 18863 |
| 153 | ACC_pre_L | -0.986 | -1.657 | -0.315 | 0.343 | -2.879 | -0.021 | 4.00E-03 | 18701 | -0.866 | -2.120 | 0.388 | 0.640 | -1.353 | -0.010 | 1.76E-01 | 18863 |
| 154 | ACC_pre_R | -0.814 | -1.460 | -0.168 | 0.330 | -2.468 | -0.018 | 1.36E-02 | 18701 | -1.058 | -2.266 | 0.150 | 0.616 | -1.716 | -0.012 | 8.61E-02 | 18863 |
| 155 | ACC_sup_L | -0.382 | -1.112 | 0.348 | 0.372 | -1.026 | -0.008 | 3.05E-01 | 18701 | -1.739 | -3.102 | -0.377 | 0.695 | -2.502 | -0.018 | 1.24E-02 | 18863 |
| 156 | ACC_sup_R | -0.776 | -1.532 | -0.021 | 0.385 | -2.015 | -0.015 | 4.39E-02 | 18701 | -1.746 | -3.158 | -0.335 | 0.720 | -2.426 | -0.018 | 1.53E-02 | 18863 |
| 157 | N_Acc_L | -0.171 | -0.391 | 0.048 | 0.112 | -1.531 | -0.011 | 1.26E-01 | 18701 | -0.124 | -0.534 | 0.287 | 0.209 | -0.591 | -0.004 | 5.55E-01 | 18863 |
| 158 | N_Acc_R | -0.070 | -0.260 | 0.120 | 0.097 | -0.720 | -0.005 | 4.72E-01 | 18701 | -0.161 | -0.516 | 0.194 | 0.181 | -0.888 | -0.006 | 3.75E-01 | 18863 |
| 159 | VTA_L | 0.003 | -0.005 | 0.011 | 0.004 | 0.795 | 0.006 | 4.27E-01 | 18701 | -0.014 | -0.028 | 0.000 | 0.007 | -1.899 | -0.014 | 5.76E-02 | 18863 |
| 160 | VTA_R | 0.001 | -0.006 | 0.008 | 0.004 | 0.379 | 0.003 | 7.05E-01 | 18701 | -0.014 | -0.027 | -0.001 | 0.007 | -2.156 | -0.016 | 3.11E-02 | 18863 |
| 161 | SN_pc_L | -0.002 | -0.014 | 0.010 | 0.006 | -0.311 | -0.002 | 7.56E-01 | 18701 | -0.031 | -0.054 | -0.008 | 0.012 | -2.597 | -0.019 | 9.42E-03 | 18863 |
| 162 | SN_pc_R | -0.004 | -0.021 | 0.012 | 0.008 | -0.514 | -0.004 | 6.07E-01 | 18701 | -0.041 | -0.071 | -0.010 | 0.016 | -2.618 | -0.019 | 8.86E-03 | 18863 |
| 163 | SN_pr_L | -0.006 | -0.023 | 0.011 | 0.009 | -0.703 | -0.005 | 4.82E-01 | 18701 | -0.063 | -0.094 | -0.031 | 0.016 | -3.907 | -0.028 | 9.38E-05 | 18863 |
| 164 | SN_pr_R | -0.010 | -0.029 | 0.009 | 0.010 | -1.047 | -0.008 | 2.95E-01 | 18701 | -0.043 | -0.078 | -0.008 | 0.018 | -2.390 | -0.017 | 1.69E-02 | 18863 |
| 165 | Red_N_L | 0.013 | -0.008 | 0.034 | 0.011 | 1.249 | 0.009 | 2.12E-01 | 18701 | -0.064 | -0.103 | -0.025 | 0.020 | -3.225 | -0.023 | 1.26E-03 | 18863 |
| 166 | Red_N_R | 0.010 | -0.013 | 0.033 | 0.012 | 0.877 | 0.006 | 3.81E-01 | 18701 | -0.046 | -0.088 | -0.004 | 0.022 | -2.131 | -0.016 | 3.31E-02 | 18863 |
| 167 | LC_L | 0.005 | -0.003 | 0.013 | 0.004 | 1.288 | 0.009 | 1.98E-01 | 18701 | 0.018 | 0.003 | 0.032 | 0.007 | 2.409 | 0.018 | 1.60E-02 | 18863 |
| 168 | LC_R | 0.002 | -0.006 | 0.011 | 0.004 | 0.539 | 0.004 | 5.90E-01 | 18701 | 0.020 | 0.005 | 0.036 | 0.008 | 2.523 | 0.018 | 1.16E-02 | 18863 |
| 169 | Raphe_D | 0.007 | -0.023 | 0.037 | 0.015 | 0.439 | 0.003 | 6.61E-01 | 18701 | -0.052 | -0.109 | 0.004 | 0.029 | -1.823 | -0.013 | 6.84E-02 | 18863 |
| 170 | Raphe_M | 0.004 | -0.005 | 0.013 | 0.005 | 0.822 | 0.006 | 4.11E-01 | 18701 | 0.005 | -0.012 | 0.022 | 0.009 | 0.529 | 0.004 | 5.97E-01 | 18863 |

| | | Fresh fruit intake | | | | | | | | Beef intake | | | | | | | |
|---|---|---|---|---|---|---|---|---|---|---|---|---|---|---|---|---|---|
| **Number** | **AAL 3 region** | **beta** | **95%CI_l** | **95%CI_h** | **se** | **t** | **r** | **p** | **df.e** | **beta** | **95%CI_l** | **95%CI_h** | **se** | **t** | **r** | **p** | **df.e** |
| 0 | Total brain | 42.218 | -56.901 | 141.337 | 50.569 | 0.835 | 0.006 | 4.04E-01 | 18761 | -407.572 | -568.427 | -246.716 | 82.065 | -4.966 | -0.036 | 6.88E-07 | 18856 |
| 1 | Precentral_L | -2.745 | -6.232 | 0.743 | 1.779 | -1.543 | -0.011 | 1.23E-01 | 18761 | -6.181 | -11.839 | -0.522 | 2.887 | -2.141 | -0.016 | 3.23E-02 | 18856 |
| 2 | Precentral_R | 1.270 | -1.926 | 4.465 | 1.630 | 0.779 | 0.006 | 4.36E-01 | 18761 | -7.285 | -12.467 | -2.102 | 2.644 | -2.755 | -0.020 | 5.88E-03 | 18856 |
| 3 | Frontal_Sup_2_L | -0.491 | -4.173 | 3.191 | 1.878 | -0.261 | -0.002 | 7.94E-01 | 18761 | -11.394 | -17.368 | -5.419 | 3.048 | -3.738 | -0.027 | 1.86E-04 | 18856 |
| 4 | Frontal_Sup_2_R | -1.654 | -5.713 | 2.405 | 2.071 | -0.799 | -0.006 | 4.25E-01 | 18761 | -11.441 | -18.026 | -4.855 | 3.360 | -3.405 | -0.025 | 6.63E-04 | 18856 |
| 5 | Frontal_Mid_2_L | -2.649 | -6.516 | 1.217 | 1.973 | -1.343 | -0.010 | 1.79E-01 | 18761 | -9.554 | -15.830 | -3.278 | 3.202 | -2.984 | -0.022 | 2.85E-03 | 18856 |
| 6 | Frontal_Mid_2_R | -2.000 | -6.374 | 2.374 | 2.231 | -0.896 | -0.007 | 3.70E-01 | 18761 | -15.496 | -22.595 | -8.396 | 3.622 | -4.278 | -0.031 | 1.89E-05 | 18856 |

| | | | | | | | | | | | | | | | | | |
|---|---|---|---|---|---|---|---|---|---|---|---|---|---|---|---|---|---|
| 7 | Frontal_Inf_Oper_L | -0.304 | -1.446 | 0.838 | 0.583 | -0.521 | -0.004 | 6.02E-01 | 18761 | -3.815 | -5.668 | -1.963 | 0.945 | -4.036 | -0.029 | 5.45E-05 | 18856 |
| 8 | Frontal_Inf_Oper_R | 0.026 | -1.406 | 1.459 | 0.731 | 0.036 | 0.000 | 9.71E-01 | 18761 | -4.641 | -6.966 | -2.316 | 1.186 | -3.913 | -0.028 | 9.16E-05 | 18856 |
| 9 | Frontal_Inf_Tri_L | -1.259 | -3.682 | 1.165 | 1.236 | -1.018 | -0.007 | 3.09E-01 | 18761 | -7.721 | -11.657 | -3.786 | 2.008 | -3.846 | -0.028 | 1.21E-04 | 18856 |
| 10 | Frontal_Inf_Tri_R | -0.679 | -2.583 | 1.226 | 0.972 | -0.698 | -0.005 | 4.85E-01 | 18761 | -6.761 | -9.852 | -3.670 | 1.577 | -4.287 | -0.031 | 1.82E-05 | 18856 |
| 11 | Frontal_Inf_Orb_2_L | -0.578 | -1.303 | 0.146 | 0.370 | -1.565 | -0.011 | 1.18E-01 | 18761 | -1.697 | -2.875 | -0.520 | 0.601 | -2.826 | -0.021 | 4.71E-03 | 18856 |
| 12 | Frontal_Inf_Orb_2_R | 0.058 | -0.700 | 0.816 | 0.387 | 0.150 | 0.001 | 8.81E-01 | 18761 | -3.131 | -4.362 | -1.901 | 0.628 | -4.990 | -0.036 | 6.11E-07 | 18856 |
| 13 | Rolandic_Oper_L | -1.010 | -2.107 | 0.087 | 0.560 | -1.805 | -0.013 | 7.10E-02 | 18761 | -1.776 | -3.556 | 0.004 | 0.908 | -1.956 | -0.014 | 5.05E-02 | 18856 |
| 14 | Rolandic_Oper_R | 0.875 | -0.694 | 2.444 | 0.800 | 1.093 | 0.008 | 2.74E-01 | 18761 | -4.872 | -7.419 | -2.326 | 1.299 | -3.750 | -0.027 | 1.77E-04 | 18856 |
| 15 | Supp_Motor_Area_L | 2.361 | -0.248 | 4.971 | 1.331 | 1.774 | 0.013 | 7.61E-02 | 18761 | -4.031 | -8.265 | 0.203 | 2.160 | -1.866 | -0.014 | 6.21E-02 | 18856 |
| 16 | Supp_Motor_Area_R | 2.291 | -0.591 | 5.173 | 1.470 | 1.558 | 0.011 | 1.19E-01 | 18761 | -4.625 | -9.303 | 0.053 | 2.387 | -1.938 | -0.014 | 5.27E-02 | 18856 |
| 17 | Olfactory_L | 0.016 | -0.372 | 0.403 | 0.198 | 0.079 | 0.001 | 9.37E-01 | 18761 | -0.438 | -1.067 | 0.191 | 0.321 | -1.364 | -0.010 | 1.73E-01 | 18856 |
| 18 | Olfactory_R | -0.095 | -0.484 | 0.293 | 0.198 | -0.481 | -0.004 | 6.31E-01 | 18761 | -0.609 | -1.240 | 0.022 | 0.322 | -1.893 | -0.014 | 5.84E-02 | 18856 |
| 19 | Frontal_Sup_Medial_L | 2.054 | -0.392 | 4.499 | 1.248 | 1.646 | 0.012 | 9.97E-02 | 18761 | -10.061 | -14.028 | -6.094 | 2.024 | -4.971 | -0.036 | 6.71E-07 | 18856 |
| 20 | Frontal_Sup_Medial_R | 0.700 | -1.080 | 2.480 | 0.908 | 0.771 | 0.006 | 4.41E-01 | 18761 | -7.573 | -10.459 | -4.687 | 1.472 | -5.144 | -0.037 | 2.72E-07 | 18856 |
| 21 | Frontal_Med_Orb_L | 0.403 | -0.510 | 1.316 | 0.466 | 0.865 | 0.006 | 3.87E-01 | 18761 | -3.296 | -4.778 | -1.814 | 0.756 | -4.359 | -0.032 | 1.32E-05 | 18856 |
| 22 | Frontal_Med_Orb_R | 0.604 | -0.517 | 1.726 | 0.572 | 1.056 | 0.008 | 2.91E-01 | 18761 | -3.424 | -5.245 | -1.602 | 0.929 | -3.684 | -0.027 | 2.30E-04 | 18856 |
| 23 | Rectus_L | 0.407 | -0.538 | 1.351 | 0.482 | 0.844 | 0.006 | 3.98E-01 | 18761 | -2.600 | -4.135 | -1.066 | 0.783 | -3.322 | -0.024 | 8.96E-04 | 18856 |
| 24 | Rectus_R | 0.314 | -0.559 | 1.188 | 0.446 | 0.705 | 0.005 | 4.81E-01 | 18761 | -2.286 | -3.705 | -0.866 | 0.724 | -3.156 | -0.023 | 1.60E-03 | 18856 |
| 25 | OFCmed_L | -0.100 | -0.647 | 0.448 | 0.279 | -0.356 | -0.003 | 7.21E-01 | 18761 | -1.606 | -2.496 | -0.717 | 0.454 | -3.540 | -0.026 | 4.01E-04 | 18856 |
| 26 | OFCmed_R | -0.397 | -1.035 | 0.241 | 0.325 | -1.219 | -0.009 | 2.23E-01 | 18761 | -2.051 | -3.087 | -1.015 | 0.529 | -3.880 | -0.028 | 1.05E-04 | 18856 |
| 27 | OFCant_L | -0.407 | -1.040 | 0.226 | 0.323 | -1.261 | -0.009 | 2.07E-01 | 18761 | -1.402 | -2.431 | -0.374 | 0.525 | -2.673 | -0.019 | 7.53E-03 | 18856 |
| 28 | OFCant_R | -1.019 | -1.855 | -0.183 | 0.426 | -2.390 | -0.017 | 1.69E-02 | 18761 | -3.000 | -4.358 | -1.642 | 0.693 | -4.331 | -0.032 | 1.49E-05 | 18856 |
| 29 | OFCpost_L | -0.367 | -1.159 | 0.425 | 0.404 | -0.909 | -0.007 | 3.63E-01 | 18761 | -1.756 | -3.043 | -0.469 | 0.657 | -2.674 | -0.019 | 7.51E-03 | 18856 |
| 30 | OFCpost_R | -0.441 | -1.174 | 0.291 | 0.374 | -1.181 | -0.009 | 2.38E-01 | 18761 | -2.763 | -3.952 | -1.573 | 0.607 | -4.552 | -0.033 | 5.35E-06 | 18856 |
| 31 | OFClat_L | -0.331 | -0.598 | -0.063 | 0.136 | -2.423 | -0.018 | 1.54E-02 | 18761 | -0.881 | -1.315 | -0.446 | 0.222 | -3.975 | -0.029 | 7.07E-05 | 18856 |
| 32 | OFClat_R | -0.099 | -0.325 | 0.127 | 0.115 | -0.858 | -0.006 | 3.91E-01 | 18761 | -0.952 | -1.319 | -0.585 | 0.187 | -5.088 | -0.037 | 3.66E-07 | 18856 |
| 33 | Insula_L | -0.923 | -3.374 | 1.527 | 1.250 | -0.739 | -0.005 | 4.60E-01 | 18761 | -6.409 | -10.389 | -2.429 | 2.031 | -3.156 | -0.023 | 1.60E-03 | 18856 |
| 34 | Insula_R | -0.018 | -2.407 | 2.371 | 1.219 | -0.015 | 0.000 | 9.88E-01 | 18761 | -8.656 | -12.534 | -4.778 | 1.978 | -4.375 | -0.032 | 1.22E-05 | 18856 |
| 37 | Cingulate_Mid_L | 2.808 | 0.734 | 4.883 | 1.058 | 2.653 | 0.019 | 7.98E-03 | 18761 | -9.609 | -12.976 | -6.242 | 1.718 | -5.594 | -0.041 | 2.25E-08 | 18856 |
| 38 | Cingulate_Mid_R | 2.694 | 0.483 | 4.904 | 1.128 | 2.388 | 0.017 | 1.69E-02 | 18761 | -9.352 | -12.941 | -5.763 | 1.831 | -5.107 | -0.037 | 3.30E-07 | 18856 |
| 39 | Cingulate_Post_L | -0.228 | -0.861 | 0.406 | 0.323 | -0.704 | -0.005 | 4.81E-01 | 18761 | -2.095 | -3.122 | -1.067 | 0.524 | -3.995 | -0.029 | 6.48E-05 | 18856 |
| 40 | Cingulate_Post_R | -0.253 | -0.604 | 0.098 | 0.179 | -1.412 | -0.010 | 1.58E-01 | 18761 | -0.866 | -1.436 | -0.296 | 0.291 | -2.979 | -0.022 | 2.90E-03 | 18856 |
| 41 | Hippocampus_L | 0.457 | -0.567 | 1.481 | 0.522 | 0.875 | 0.006 | 3.82E-01 | 18761 | -2.394 | -4.057 | -0.731 | 0.848 | -2.822 | -0.021 | 4.77E-03 | 18856 |
| 42 | Hippocampus_R | 0.948 | -0.114 | 2.011 | 0.542 | 1.749 | 0.013 | 8.03E-02 | 18761 | -2.584 | -4.309 | -0.859 | 0.880 | -2.937 | -0.021 | 3.32E-03 | 18856 |
| 43 | ParaHippocampal_L | 0.964 | 0.054 | 1.874 | 0.464 | 2.077 | 0.015 | 3.78E-02 | 18761 | -1.806 | -3.284 | -0.329 | 0.754 | -2.396 | -0.017 | 1.66E-02 | 18856 |
| 44 | ParaHippocampal_R | 0.723 | -0.426 | 1.871 | 0.586 | 1.234 | 0.009 | 2.17E-01 | 18761 | -3.132 | -4.997 | -1.267 | 0.952 | -3.291 | -0.024 | 1.00E-03 | 18856 |
| 45 | Amygdala_L | 0.157 | -0.196 | 0.511 | 0.180 | 0.873 | 0.006 | 3.83E-01 | 18761 | -0.299 | -0.873 | 0.275 | 0.293 | -1.020 | -0.007 | 3.08E-01 | 18856 |
| 46 | Amygdala_R | 0.124 | -0.238 | 0.485 | 0.184 | 0.671 | 0.005 | 5.02E-01 | 18761 | -0.993 | -1.580 | -0.406 | 0.300 | -3.314 | -0.024 | 9.20E-04 | 18856 |
| 47 | Calcarine_L | -0.393 | -3.768 | 2.982 | 1.722 | -0.228 | -0.002 | 8.19E-01 | 18761 | 2.151 | -3.326 | 7.629 | 2.795 | 0.770 | 0.006 | 4.41E-01 | 18856 |
| 48 | Calcarine_R | 0.464 | -2.355 | 3.283 | 1.438 | 0.322 | 0.002 | 7.47E-01 | 18761 | 0.459 | -4.117 | 5.035 | 2.335 | 0.197 | 0.001 | 8.44E-01 | 18856 |
| 49 | Cuneus_L | 2.166 | 0.047 | 4.285 | 1.081 | 2.003 | 0.015 | 4.51E-02 | 18761 | -1.425 | -4.864 | 2.014 | 1.754 | -0.812 | -0.006 | 4.17E-01 | 18856 |
| 50 | Cuneus_R | 0.248 | -1.372 | 1.867 | 0.826 | 0.300 | 0.002 | 7.64E-01 | 18761 | -2.176 | -4.806 | 0.453 | 1.342 | -1.622 | -0.012 | 1.05E-01 | 18856 |
| 51 | Lingual_L | 0.223 | -1.808 | 2.255 | 1.036 | 0.216 | 0.002 | 8.29E-01 | 18761 | -1.359 | -4.657 | 1.939 | 1.682 | -0.808 | -0.006 | 4.19E-01 | 18856 |
| 52 | Lingual_R | -0.044 | -2.171 | 2.082 | 1.085 | -0.041 | 0.000 | 9.67E-01 | 18761 | -1.615 | -5.068 | 1.838 | 1.761 | -0.917 | -0.007 | 3.59E-01 | 18856 |
| 53 | Occipital_Sup_L | 1.100 | -0.296 | 2.496 | 0.712 | 1.544 | 0.011 | 1.23E-01 | 18761 | -4.431 | -6.698 | -2.165 | 1.156 | -3.833 | -0.028 | 1.27E-04 | 18856 |
| 54 | Occipital_Sup_R | 0.128 | -1.385 | 1.642 | 0.772 | 0.166 | 0.001 | 8.68E-01 | 18761 | -5.038 | -7.494 | -2.583 | 1.253 | -4.022 | -0.029 | 5.79E-05 | 18856 |
| 55 | Occipital_Mid_L | -1.382 | -4.651 | 1.888 | 1.668 | -0.828 | -0.006 | 4.08E-01 | 18761 | -7.707 | -13.016 | -2.398 | 2.709 | -2.845 | -0.021 | 4.44E-03 | 18856 |
| 56 | Occipital_Mid_R | -1.104 | -3.381 | 1.173 | 1.162 | -0.950 | -0.007 | 3.42E-01 | 18761 | -4.212 | -7.908 | -0.515 | 1.886 | -2.233 | -0.016 | 2.55E-02 | 18856 |
| 57 | Occipital_Inf_L | -1.338 | -2.423 | -0.253 | 0.554 | -2.417 | -0.018 | 1.57E-02 | 18761 | 0.191 | -1.571 | 1.953 | 0.899 | 0.212 | 0.002 | 8.32E-01 | 18856 |

| | | | | | | | | | | | | | | | | | |
|---|---|---|---|---|---|---|---|---|---|---|---|---|---|---|---|---|---|
| 58 | Occipital_Inf_R | -1.723 | -2.977 | -0.468 | 0.640 | -2.692 | -0.020 | 7.11E-03 | 18761 | 1.607 | -0.429 | 3.643 | 1.039 | 1.547 | 0.011 | 1.22E-01 | 18856 |
| 59 | Fusiform_L | 2.234 | -0.142 | 4.610 | 1.212 | 1.843 | 0.013 | 6.54E-02 | 18761 | -0.237 | -4.095 | 3.622 | 1.969 | -0.120 | -0.001 | 9.04E-01 | 18856 |
| 60 | Fusiform_R | 3.952 | 1.342 | 6.562 | 1.331 | 2.968 | 0.022 | 3.00E-03 | 18761 | -1.846 | -6.086 | 2.394 | 2.163 | -0.853 | -0.006 | 3.93E-01 | 18856 |
| 61 | Postcentral_L | 7.208 | 3.633 | 10.783 | 1.824 | 3.952 | 0.029 | 7.79E-05 | 18761 | -9.644 | -15.448 | -3.840 | 2.961 | -3.257 | -0.024 | 1.13E-03 | 18856 |
| 62 | Postcentral_R | 8.965 | 5.304 | 12.625 | 1.867 | 4.800 | 0.035 | 1.60E-06 | 18761 | -6.775 | -12.720 | -0.831 | 3.033 | -2.234 | -0.016 | 2.55E-02 | 18856 |
| 63 | Parietal_Sup_L | -0.076 | -2.136 | 1.985 | 1.051 | -0.072 | -0.001 | 9.43E-01 | 18761 | -4.883 | -8.227 | -1.539 | 1.706 | -2.862 | -0.021 | 4.21E-03 | 18856 |
| 64 | Parietal_Sup_R | 0.368 | -1.460 | 2.196 | 0.933 | 0.394 | 0.003 | 6.93E-01 | 18761 | -4.280 | -7.245 | -1.315 | 1.513 | -2.829 | -0.021 | 4.67E-03 | 18856 |
| 65 | Parietal_Inf_L | 3.275 | 0.577 | 5.972 | 1.376 | 2.379 | 0.017 | 1.74E-02 | 18761 | -7.799 | -12.176 | -3.422 | 2.233 | -3.493 | -0.025 | 4.80E-04 | 18856 |
| 66 | Parietal_Inf_R | 2.516 | 0.767 | 4.264 | 0.892 | 2.820 | 0.021 | 4.80E-03 | 18761 | -3.930 | -6.768 | -1.092 | 1.448 | -2.714 | -0.020 | 6.65E-03 | 18856 |
| 67 | SupraMarginal_L | 1.141 | -0.440 | 2.721 | 0.806 | 1.414 | 0.010 | 1.57E-01 | 18761 | -5.167 | -7.733 | -2.602 | 1.309 | -3.948 | -0.029 | 7.91E-05 | 18856 |
| 68 | SupraMarginal_R | 4.377 | 2.140 | 6.614 | 1.141 | 3.835 | 0.028 | 1.26E-04 | 18761 | -8.565 | -12.196 | -4.934 | 1.853 | -4.623 | -0.034 | 3.81E-06 | 18856 |
| 69 | Angular_L | 0.781 | -0.772 | 2.335 | 0.793 | 0.986 | 0.007 | 3.24E-01 | 18761 | -2.671 | -5.192 | -0.150 | 1.286 | -2.077 | -0.015 | 3.78E-02 | 18856 |
| 70 | Angular_R | 0.269 | -1.813 | 2.351 | 1.062 | 0.253 | 0.002 | 8.00E-01 | 18761 | -5.009 | -8.387 | -1.630 | 1.724 | -2.906 | -0.021 | 3.66E-03 | 18856 |
| 71 | Precuneus_L | 3.631 | -0.155 | 7.416 | 1.931 | 1.880 | 0.014 | 6.01E-02 | 18761 | -8.376 | -14.520 | -2.231 | 3.135 | -2.672 | -0.019 | 7.55E-03 | 18856 |
| 72 | Precuneus_R | 0.911 | -2.462 | 4.283 | 1.721 | 0.529 | 0.004 | 5.97E-01 | 18761 | -7.606 | -13.078 | -2.134 | 2.792 | -2.724 | -0.020 | 6.45E-03 | 18856 |
| 73 | Paracentral_Lobule_L | 1.791 | 0.058 | 3.525 | 0.884 | 2.025 | 0.015 | 4.29E-02 | 18761 | -1.759 | -4.573 | 1.055 | 1.435 | -1.225 | -0.009 | 2.20E-01 | 18856 |
| 74 | Paracentral_Lobule_R | 0.922 | -0.320 | 2.163 | 0.633 | 1.455 | 0.011 | 1.46E-01 | 18761 | -1.284 | -3.299 | 0.731 | 1.028 | -1.249 | -0.009 | 2.12E-01 | 18856 |
| 75 | Caudate_L | -0.096 | -1.480 | 1.288 | 0.706 | -0.136 | -0.001 | 8.92E-01 | 18761 | -4.080 | -6.327 | -1.833 | 1.146 | -3.559 | -0.026 | 3.73E-04 | 18856 |
| 76 | Caudate_R | -0.779 | -2.347 | 0.789 | 0.800 | -0.974 | -0.007 | 3.30E-01 | 18761 | -4.098 | -6.644 | -1.552 | 1.299 | -3.155 | -0.023 | 1.61E-03 | 18856 |
| 77 | Putamen_L | -1.411 | -3.512 | 0.689 | 1.072 | -1.317 | -0.010 | 1.88E-01 | 18761 | -5.773 | -9.185 | -2.362 | 1.740 | -3.317 | -0.024 | 9.12E-04 | 18856 |
| 78 | Putamen_R | -0.937 | -3.078 | 1.203 | 1.092 | -0.858 | -0.006 | 3.91E-01 | 18761 | -6.129 | -9.603 | -2.654 | 1.773 | -3.457 | -0.025 | 5.47E-04 | 18856 |
| 79 | Pallidum_L | -0.060 | -0.247 | 0.127 | 0.095 | -0.627 | -0.005 | 5.31E-01 | 18761 | -0.573 | -0.876 | -0.269 | 0.155 | -3.701 | -0.027 | 2.15E-04 | 18856 |
| 80 | Pallidum_R | -0.042 | -0.248 | 0.163 | 0.105 | -0.406 | -0.003 | 6.85E-01 | 18761 | -0.662 | -0.995 | -0.329 | 0.170 | -3.900 | -0.028 | 9.65E-05 | 18856 |
| 83 | Heschl_L | -0.337 | -0.716 | 0.042 | 0.193 | -1.743 | -0.013 | 8.14E-02 | 18761 | -0.989 | -1.603 | -0.374 | 0.314 | -3.151 | -0.023 | 1.63E-03 | 18856 |
| 84 | Heschl_R | 0.138 | -0.286 | 0.561 | 0.216 | 0.636 | 0.005 | 5.24E-01 | 18761 | -1.525 | -2.212 | -0.838 | 0.350 | -4.353 | -0.032 | 1.35E-05 | 18856 |
| 85 | Temporal_Sup_L | -0.925 | -3.658 | 1.808 | 1.395 | -0.663 | -0.005 | 5.07E-01 | 18761 | -9.173 | -13.610 | -4.737 | 2.263 | -4.053 | -0.030 | 5.08E-05 | 18856 |
| 86 | Temporal_Sup_R | 2.609 | -0.417 | 5.634 | 1.544 | 1.690 | 0.012 | 9.10E-02 | 18761 | -12.809 | -17.719 | -7.898 | 2.505 | -5.113 | -0.037 | 3.21E-07 | 18856 |
| 87 | Temporal_Pole_Sup_L | -0.421 | -1.716 | 0.875 | 0.661 | -0.637 | -0.005 | 5.24E-01 | 18761 | -2.765 | -4.868 | -0.662 | 1.073 | -2.577 | -0.019 | 9.97E-03 | 18856 |
| 88 | Temporal_Pole_Sup_R | 0.269 | -1.094 | 1.632 | 0.695 | 0.386 | 0.003 | 6.99E-01 | 18761 | -4.218 | -6.431 | -2.004 | 1.129 | -3.735 | -0.027 | 1.88E-04 | 18856 |
| 89 | Temporal_Mid_L | 1.329 | -3.228 | 5.886 | 2.325 | 0.572 | 0.004 | 5.68E-01 | 18761 | -16.029 | -23.430 | -8.629 | 3.776 | -4.246 | -0.031 | 2.19E-05 | 18856 |
| 90 | Temporal_Mid_R | -0.150 | -4.196 | 3.896 | 2.064 | -0.073 | -0.001 | 9.42E-01 | 18761 | -7.848 | -14.420 | -1.275 | 3.353 | -2.340 | -0.017 | 1.93E-02 | 18856 |
| 91 | Temporal_Pole_Mid_L | 0.226 | -0.863 | 1.315 | 0.556 | 0.406 | 0.003 | 6.84E-01 | 18761 | -0.400 | -2.169 | 1.368 | 0.902 | -0.444 | -0.003 | 6.57E-01 | 18856 |
| 92 | Temporal_Pole_Mid_R | -0.037 | -1.358 | 1.284 | 0.674 | -0.054 | 0.000 | 9.57E-01 | 18761 | -2.436 | -4.584 | -0.289 | 1.096 | -2.223 | -0.016 | 2.62E-02 | 18856 |
| 93 | Temporal_Inf_L | -0.697 | -3.828 | 2.434 | 1.597 | -0.436 | -0.003 | 6.63E-01 | 18761 | -3.309 | -8.398 | 1.781 | 2.597 | -1.274 | -0.009 | 2.03E-01 | 18856 |
| 94 | Temporal_Inf_R | -1.095 | -4.509 | 2.319 | 1.742 | -0.629 | -0.005 | 5.29E-01 | 18761 | -3.706 | -9.254 | 1.843 | 2.831 | -1.309 | -0.010 | 1.91E-01 | 18856 |
| 95 | Cerebellum_Crus1_L | 1.378 | -2.346 | 5.102 | 1.900 | 0.725 | 0.005 | 4.68E-01 | 18761 | 1.785 | -4.259 | 7.828 | 3.083 | 0.579 | 0.004 | 5.63E-01 | 18856 |
| 96 | Cerebellum_Crus1_R | 0.290 | -3.507 | 4.086 | 1.937 | 0.150 | 0.001 | 8.81E-01 | 18761 | 0.817 | -5.345 | 6.979 | 3.144 | 0.260 | 0.002 | 7.95E-01 | 18856 |
| 97 | Cerebellum_Crus2_L | -0.452 | -3.404 | 2.500 | 1.506 | -0.300 | -0.002 | 7.64E-01 | 18761 | -4.195 | -8.988 | 0.598 | 2.445 | -1.715 | -0.012 | 8.63E-02 | 18856 |
| 98 | Cerebellum_Crus2_R | -0.528 | -3.254 | 2.199 | 1.391 | -0.379 | -0.003 | 7.04E-01 | 18761 | -2.572 | -7.003 | 1.859 | 2.261 | -1.138 | -0.008 | 2.55E-01 | 18856 |
| 99 | Cerebellum_3_L | 0.088 | -0.110 | 0.286 | 0.101 | 0.872 | 0.006 | 3.83E-01 | 18761 | 0.621 | 0.299 | 0.942 | 0.164 | 3.785 | 0.028 | 1.54E-04 | 18856 |
| 100 | Cerebellum_3_R | 0.224 | -0.043 | 0.491 | 0.136 | 1.644 | 0.012 | 1.00E-01 | 18761 | 0.635 | 0.201 | 1.069 | 0.221 | 2.869 | 0.021 | 4.12E-03 | 18856 |
| 101 | Cerebellum_4_5_L | 0.730 | -0.872 | 2.333 | 0.817 | 0.894 | 0.007 | 3.72E-01 | 18761 | 2.897 | 0.297 | 5.498 | 1.327 | 2.184 | 0.016 | 2.90E-02 | 18856 |
| 102 | Cerebellum_4_5_R | 1.591 | 0.286 | 2.896 | 0.666 | 2.390 | 0.017 | 1.69E-02 | 18761 | 1.665 | -0.455 | 3.784 | 1.081 | 1.539 | 0.011 | 1.24E-01 | 18856 |
| 103 | Cerebellum_6_L | 2.308 | -0.463 | 5.078 | 1.414 | 1.632 | 0.012 | 1.03E-01 | 18761 | 1.877 | -2.623 | 6.376 | 2.296 | 0.817 | 0.006 | 4.14E-01 | 18856 |
| 104 | Cerebellum_6_R | 2.702 | -0.216 | 5.621 | 1.489 | 1.815 | 0.013 | 6.96E-02 | 18761 | 2.432 | -2.307 | 7.171 | 2.418 | 1.006 | 0.007 | 3.15E-01 | 18856 |
| 105 | Cerebellum_7b_L | -0.032 | -0.934 | 0.870 | 0.460 | -0.069 | -0.001 | 9.45E-01 | 18761 | -1.139 | -2.604 | 0.326 | 0.748 | -1.524 | -0.011 | 1.28E-01 | 18856 |
| 106 | Cerebellum_7b_R | 0.010 | -0.884 | 0.904 | 0.456 | 0.022 | 0.000 | 9.83E-01 | 18761 | -1.634 | -3.086 | -0.182 | 0.741 | -2.206 | -0.016 | 2.74E-02 | 18856 |
| 107 | Cerebellum_8_L | -0.104 | -2.972 | 2.764 | 1.463 | -0.071 | -0.001 | 9.43E-01 | 18761 | 0.948 | -3.706 | 5.602 | 2.374 | 0.399 | 0.003 | 6.90E-01 | 18856 |
| 108 | Cerebellum_8_R | -0.657 | -4.389 | 3.075 | 1.904 | -0.345 | -0.003 | 7.30E-01 | 18761 | -5.159 | -11.217 | 0.898 | 3.090 | -1.670 | -0.012 | 9.50E-02 | 18856 |

| | | | | | | | | | | | | | | | | | |
|---|---|---|---|---|---|---|---|---|---|---|---|---|---|---|---|---|---|
| 109 | Cerebellum_9_L | -1.258 | -2.879 | 0.364 | 0.827 | -1.521 | -0.011 | 1.28E-01 | 18761 | 2.222 | -0.408 | 4.852 | 1.342 | 1.656 | 0.012 | 9.77E-02 | 18856 |
| 110 | Cerebellum_9_R | -1.656 | -3.250 | -0.061 | 0.813 | -2.035 | -0.015 | 4.18E-02 | 18761 | 2.330 | -0.256 | 4.915 | 1.319 | 1.766 | 0.013 | 7.74E-02 | 18856 |
| 111 | Cerebellum_10_L | -0.037 | -0.152 | 0.078 | 0.059 | -0.632 | -0.005 | 5.27E-01 | 18761 | 0.236 | 0.050 | 0.423 | 0.095 | 2.484 | 0.018 | 1.30E-02 | 18856 |
| 112 | Cerebellum_10_R | -0.006 | -0.126 | 0.113 | 0.061 | -0.102 | -0.001 | 9.19E-01 | 18761 | 0.238 | 0.044 | 0.432 | 0.099 | 2.405 | 0.018 | 1.62E-02 | 18856 |
| 113 | Vermis_1_2 | 0.004 | -0.071 | 0.079 | 0.038 | 0.108 | 0.001 | 9.14E-01 | 18761 | 0.154 | 0.033 | 0.275 | 0.062 | 2.494 | 0.018 | 1.27E-02 | 18856 |
| 114 | Vermis_3 | -0.005 | -0.302 | 0.292 | 0.151 | -0.034 | 0.000 | 9.73E-01 | 18761 | 0.410 | -0.072 | 0.892 | 0.246 | 1.668 | 0.012 | 9.53E-02 | 18856 |
| 115 | Vermis_4_5 | -0.055 | -0.928 | 0.817 | 0.445 | -0.124 | -0.001 | 9.01E-01 | 18761 | 0.905 | -0.511 | 2.322 | 0.723 | 1.253 | 0.009 | 2.10E-01 | 18856 |
| 116 | Vermis_6 | 0.081 | -0.404 | 0.567 | 0.248 | 0.329 | 0.002 | 7.42E-01 | 18761 | 0.855 | 0.066 | 1.644 | 0.402 | 2.125 | 0.015 | 3.36E-02 | 18856 |
| 117 | Vermis_7 | -0.043 | -0.317 | 0.232 | 0.140 | -0.303 | -0.002 | 7.62E-01 | 18761 | 0.223 | -0.223 | 0.669 | 0.228 | 0.978 | 0.007 | 3.28E-01 | 18856 |
| 118 | Vermis_8 | -0.078 | -0.529 | 0.374 | 0.230 | -0.337 | -0.002 | 7.36E-01 | 18761 | 0.535 | -0.198 | 1.268 | 0.374 | 1.430 | 0.010 | 1.53E-01 | 18856 |
| 119 | Vermis_9 | -0.070 | -0.437 | 0.296 | 0.187 | -0.376 | -0.003 | 7.07E-01 | 18761 | 0.568 | -0.027 | 1.162 | 0.303 | 1.872 | 0.014 | 6.12E-02 | 18856 |
| 120 | Vermis_10 | -0.057 | -0.159 | 0.045 | 0.052 | -1.101 | -0.008 | 2.71E-01 | 18761 | 0.313 | 0.147 | 0.478 | 0.084 | 3.704 | 0.027 | 2.13E-04 | 18856 |
| 121 | Thal_AV_L | -0.001 | -0.041 | 0.040 | 0.021 | -0.025 | 0.000 | 9.80E-01 | 18761 | -0.112 | -0.178 | -0.046 | 0.034 | -3.315 | -0.024 | 9.18E-04 | 18856 |
| 122 | Thal_AV_R | -0.029 | -0.087 | 0.029 | 0.030 | -0.974 | -0.007 | 3.30E-01 | 18761 | -0.141 | -0.236 | -0.047 | 0.048 | -2.923 | -0.021 | 3.47E-03 | 18856 |
| 123 | Thal_LP_L | 0.000 | -0.046 | 0.045 | 0.023 | -0.016 | 0.000 | 9.87E-01 | 18761 | -0.161 | -0.235 | -0.088 | 0.037 | -4.307 | -0.031 | 1.67E-05 | 18856 |
| 124 | Thal_LP_R | -0.032 | -0.091 | 0.027 | 0.030 | -1.058 | -0.008 | 2.90E-01 | 18761 | -0.211 | -0.306 | -0.115 | 0.049 | -4.314 | -0.031 | 1.61E-05 | 18856 |
| 125 | Thal_VA_L | -0.007 | -0.055 | 0.041 | 0.025 | -0.285 | -0.002 | 7.76E-01 | 18761 | -0.057 | -0.136 | 0.021 | 0.040 | -1.434 | -0.010 | 1.52E-01 | 18856 |
| 126 | Thal_VA_R | -0.020 | -0.079 | 0.040 | 0.030 | -0.651 | -0.005 | 5.15E-01 | 18761 | -0.050 | -0.146 | 0.047 | 0.049 | -1.010 | -0.007 | 3.13E-01 | 18856 |
| 127 | Thal_VL_L | -0.060 | -0.197 | 0.077 | 0.070 | -0.855 | -0.006 | 3.92E-01 | 18761 | -0.411 | -0.634 | -0.188 | 0.114 | -3.611 | -0.026 | 3.06E-04 | 18856 |
| 128 | Thal_VL_R | -0.137 | -0.320 | 0.045 | 0.093 | -1.477 | -0.011 | 1.40E-01 | 18761 | -0.619 | -0.915 | -0.323 | 0.151 | -4.097 | -0.030 | 4.21E-05 | 18856 |
| 129 | Thal_VPL_L | -0.175 | -0.339 | -0.012 | 0.083 | -2.099 | -0.015 | 3.58E-02 | 18761 | -0.391 | -0.656 | -0.125 | 0.135 | -2.884 | -0.021 | 3.93E-03 | 18856 |
| 130 | Thal_VPL_R | -0.135 | -0.307 | 0.037 | 0.088 | -1.541 | -0.011 | 1.23E-01 | 18761 | -0.563 | -0.842 | -0.284 | 0.142 | -3.954 | -0.029 | 7.73E-05 | 18856 |
| 131 | Thal_IL_L | -0.040 | -0.110 | 0.031 | 0.036 | -1.100 | -0.008 | 2.72E-01 | 18761 | -0.334 | -0.449 | -0.219 | 0.059 | -5.694 | -0.041 | 1.26E-08 | 18856 |
| 132 | Thal_IL_R | -0.034 | -0.114 | 0.047 | 0.041 | -0.824 | -0.006 | 4.10E-01 | 18761 | -0.364 | -0.494 | -0.233 | 0.066 | -5.468 | -0.040 | 4.60E-08 | 18856 |
| 133 | Thal_Re_L | 0.000 | 0.000 | 0.000 | 0.000 | 0.014 | 0.000 | 9.89E-01 | 18761 | 0.000 | -0.001 | 0.000 | 0.000 | -0.777 | -0.006 | 4.37E-01 | 18856 |
| 134 | Thal_Re_R | -0.001 | -0.003 | 0.002 | 0.001 | -0.514 | -0.004 | 6.07E-01 | 18761 | -0.004 | -0.008 | 0.000 | 0.002 | -2.191 | -0.016 | 2.85E-02 | 18856 |
| 135 | Thal_MDm_L | -0.410 | -0.934 | 0.114 | 0.267 | -1.533 | -0.011 | 1.25E-01 | 18761 | -2.618 | -3.469 | -1.768 | 0.434 | -6.035 | -0.044 | 1.62E-09 | 18856 |
| 136 | Thal_MDm_R | -0.413 | -1.022 | 0.196 | 0.311 | -1.329 | -0.010 | 1.84E-01 | 18761 | -2.698 | -3.687 | -1.710 | 0.504 | -5.350 | -0.039 | 8.88E-08 | 18856 |
| 137 | Thal_MDl_L | -0.042 | -0.133 | 0.049 | 0.046 | -0.905 | -0.007 | 3.65E-01 | 18761 | -0.468 | -0.616 | -0.321 | 0.075 | -6.235 | -0.045 | 4.61E-10 | 18856 |
| 138 | Thal_MDl_R | -0.051 | -0.154 | 0.052 | 0.053 | -0.971 | -0.007 | 3.31E-01 | 18761 | -0.484 | -0.652 | -0.317 | 0.085 | -5.672 | -0.041 | 1.43E-08 | 18856 |
| 139 | Thal_LGN_L | 0.003 | -0.010 | 0.017 | 0.007 | 0.473 | 0.003 | 6.36E-01 | 18761 | -0.043 | -0.066 | -0.021 | 0.011 | -3.808 | -0.028 | 1.40E-04 | 18856 |
| 140 | Thal_LGN_R | -0.013 | -0.043 | 0.016 | 0.015 | -0.900 | -0.007 | 3.68E-01 | 18761 | -0.123 | -0.171 | -0.075 | 0.024 | -5.069 | -0.037 | 4.05E-07 | 18856 |
| 141 | Thal_MGN_L | -0.017 | -0.041 | 0.008 | 0.013 | -1.329 | -0.010 | 1.84E-01 | 18761 | -0.098 | -0.138 | -0.059 | 0.020 | -4.833 | -0.035 | 1.35E-06 | 18856 |
| 142 | Thal_MGN_R | -0.013 | -0.040 | 0.014 | 0.014 | -0.953 | -0.007 | 3.41E-01 | 18761 | -0.121 | -0.165 | -0.078 | 0.022 | -5.476 | -0.040 | 4.40E-08 | 18856 |
| 143 | Thal_PuA_L | -0.019 | -0.048 | 0.010 | 0.015 | -1.277 | -0.009 | 2.02E-01 | 18761 | -0.127 | -0.174 | -0.080 | 0.024 | -5.267 | -0.038 | 1.41E-07 | 18856 |
| 144 | Thal_PuA_R | -0.020 | -0.080 | 0.040 | 0.031 | -0.649 | -0.005 | 5.16E-01 | 18761 | -0.274 | -0.372 | -0.176 | 0.050 | -5.472 | -0.040 | 4.52E-08 | 18856 |
| 145 | Thal_PuM_L | -0.496 | -1.073 | 0.081 | 0.294 | -1.685 | -0.012 | 9.19E-02 | 18761 | -2.579 | -3.515 | -1.642 | 0.478 | -5.396 | -0.039 | 6.90E-08 | 18856 |
| 146 | Thal_PuM_R | -0.314 | -1.034 | 0.405 | 0.367 | -0.857 | -0.006 | 3.92E-01 | 18761 | -3.056 | -4.224 | -1.889 | 0.595 | -5.133 | -0.037 | 2.88E-07 | 18856 |
| 147 | Thal_PuL_L | -0.076 | -0.181 | 0.029 | 0.054 | -1.412 | -0.010 | 1.58E-01 | 18761 | -0.470 | -0.641 | -0.299 | 0.087 | -5.387 | -0.039 | 7.24E-08 | 18856 |
| 148 | Thal_PuL_R | -0.084 | -0.251 | 0.083 | 0.085 | -0.983 | -0.007 | 3.26E-01 | 18761 | -0.708 | -0.980 | -0.436 | 0.139 | -5.106 | -0.037 | 3.33E-07 | 18856 |
| 149 | Thal_PuI_L | -0.023 | -0.052 | 0.006 | 0.015 | -1.577 | -0.012 | 1.15E-01 | 18761 | -0.063 | -0.110 | -0.017 | 0.024 | -2.656 | -0.019 | 7.92E-03 | 18856 |
| 150 | Thal_PuI_R | -0.020 | -0.058 | 0.018 | 0.019 | -1.034 | -0.008 | 3.01E-01 | 18761 | -0.094 | -0.155 | -0.032 | 0.031 | -2.990 | -0.022 | 2.80E-03 | 18856 |
| 151 | ACC_sub_L | -0.057 | -0.387 | 0.274 | 0.169 | -0.336 | -0.002 | 7.37E-01 | 18761 | -0.351 | -0.887 | 0.186 | 0.274 | -1.282 | -0.009 | 2.00E-01 | 18856 |
| 152 | ACC_sub_R | -0.159 | -0.467 | 0.149 | 0.157 | -1.011 | -0.007 | 3.12E-01 | 18761 | -0.253 | -0.754 | 0.248 | 0.256 | -0.991 | -0.007 | 3.22E-01 | 18856 |
| 153 | ACC_pre_L | 0.052 | -0.795 | 0.898 | 0.432 | 0.119 | 0.001 | 9.05E-01 | 18761 | -2.618 | -3.992 | -1.244 | 0.701 | -3.734 | -0.027 | 1.89E-04 | 18856 |
| 154 | ACC_pre_R | 0.190 | -0.625 | 1.005 | 0.416 | 0.457 | 0.003 | 6.48E-01 | 18761 | -2.197 | -3.521 | -0.873 | 0.676 | -3.252 | -0.024 | 1.15E-03 | 18856 |
| 155 | ACC_sup_L | 0.097 | -0.823 | 1.018 | 0.469 | 0.208 | 0.002 | 8.36E-01 | 18761 | -2.133 | -3.627 | -0.638 | 0.762 | -2.797 | -0.020 | 5.16E-03 | 18856 |
| 156 | ACC_sup_R | 0.255 | -0.697 | 1.207 | 0.486 | 0.525 | 0.004 | 6.00E-01 | 18761 | -1.823 | -3.371 | -0.275 | 0.790 | -2.309 | -0.017 | 2.10E-02 | 18856 |
| 157 | N_Acc_L | -0.080 | -0.357 | 0.197 | 0.141 | -0.568 | -0.004 | 5.70E-01 | 18761 | -0.724 | -1.174 | -0.274 | 0.230 | -3.154 | -0.023 | 1.62E-03 | 18856 |

| | | | | | | | | | | | | | | | | | |
|---|---|---|---|---|---|---|---|---|---|---|---|---|---|---|---|---|---|
| 158 | N_Acc_R | -0.151 | -0.390 | 0.089 | 0.122 | -1.234 | -0.009 | 2.17E-01 | 18761 | -0.581 | -0.970 | -0.192 | 0.199 | -2.927 | -0.021 | 3.43E-03 | 18856 |
| 159 | VTA_L | 0.004 | -0.006 | 0.013 | 0.005 | 0.720 | 0.005 | 4.72E-01 | 18761 | 0.003 | -0.013 | 0.019 | 0.008 | 0.350 | 0.003 | 7.26E-01 | 18856 |
| 160 | VTA_R | 0.004 | -0.005 | 0.012 | 0.004 | 0.787 | 0.006 | 4.32E-01 | 18761 | 0.004 | -0.011 | 0.018 | 0.007 | 0.497 | 0.004 | 6.19E-01 | 18856 |
| 161 | SN_pc_L | 0.009 | -0.006 | 0.025 | 0.008 | 1.151 | 0.008 | 2.50E-01 | 18761 | -0.026 | -0.051 | -0.001 | 0.013 | -2.004 | -0.015 | 4.51E-02 | 18856 |
| 162 | SN_pc_R | 0.001 | -0.020 | 0.022 | 0.010 | 0.098 | 0.001 | 9.22E-01 | 18761 | -0.017 | -0.051 | 0.016 | 0.017 | -1.016 | -0.007 | 3.10E-01 | 18856 |
| 163 | SN_pr_L | -0.007 | -0.028 | 0.014 | 0.011 | -0.645 | -0.005 | 5.19E-01 | 18761 | -0.034 | -0.068 | 0.001 | 0.018 | -1.924 | -0.014 | 5.44E-02 | 18856 |
| 164 | SN_pr_R | -0.017 | -0.041 | 0.007 | 0.012 | -1.370 | -0.010 | 1.71E-01 | 18761 | -0.011 | -0.050 | 0.027 | 0.020 | -0.579 | -0.004 | 5.63E-01 | 18856 |
| 165 | Red_N_L | -0.002 | -0.028 | 0.025 | 0.013 | -0.119 | -0.001 | 9.05E-01 | 18761 | -0.035 | -0.077 | 0.008 | 0.022 | -1.597 | -0.012 | 1.10E-01 | 18856 |
| 166 | Red_N_R | -0.001 | -0.029 | 0.028 | 0.015 | -0.050 | 0.000 | 9.60E-01 | 18761 | -0.021 | -0.067 | 0.025 | 0.024 | -0.886 | -0.006 | 3.76E-01 | 18856 |
| 167 | LC_L | 0.002 | -0.008 | 0.011 | 0.005 | 0.322 | 0.002 | 7.47E-01 | 18761 | 0.013 | -0.003 | 0.029 | 0.008 | 1.573 | 0.011 | 1.16E-01 | 18856 |
| 168 | LC_R | -0.005 | -0.016 | 0.006 | 0.005 | -0.882 | -0.006 | 3.78E-01 | 18761 | 0.011 | -0.007 | 0.028 | 0.009 | 1.194 | 0.009 | 2.32E-01 | 18856 |
| 169 | Raphe_D | -0.010 | -0.048 | 0.027 | 0.019 | -0.541 | -0.004 | 5.88E-01 | 18761 | -0.082 | -0.144 | -0.021 | 0.031 | -2.620 | -0.019 | 8.79E-03 | 18856 |
| 170 | Raphe_M | 0.003 | -0.009 | 0.015 | 0.006 | 0.506 | 0.004 | 6.13E-01 | 18761 | 0.004 | -0.015 | 0.022 | 0.010 | 0.369 | 0.003 | 7.12E-01 | 18856 |

| | | Dried fruit intake | | | | | | | | Lamb/mutton intake | | | | | | | |
|---|---|---|---|---|---|---|---|---|---|---|---|---|---|---|---|---|---|
| Number | AAL 3 region | beta | 95%CI_l | 95%CI_h | se | t | r | p | df.e | beta | 95%CI_l | 95%CI_h | se | t | r | p | df.e |
| 0 | Total brain | 276.978 | 165.541 | 388.414 | 56.853 | 4.872 | 0.036 | 1.11E-06 | 18645 | -549.726 | -748.243 | -351.210 | 101.279 | -5.428 | -0.040 | 5.78E-08 | 18828 |
| 1 | Precentral_L | 1.972 | -1.952 | 5.896 | 2.002 | 0.985 | 0.007 | 3.25E-01 | 18645 | -8.809 | -15.794 | -1.825 | 3.563 | -2.472 | -0.018 | 1.34E-02 | 18828 |
| 2 | Precentral_R | 5.141 | 1.550 | 8.732 | 1.832 | 2.806 | 0.021 | 5.02E-03 | 18645 | -9.676 | -16.071 | -3.280 | 3.263 | -2.965 | -0.022 | 3.03E-03 | 18828 |
| 3 | Frontal_Sup_2_L | 7.493 | 3.356 | 11.630 | 2.111 | 3.550 | 0.026 | 3.86E-04 | 18645 | -12.695 | -20.070 | -5.319 | 3.763 | -3.374 | -0.025 | 7.43E-04 | 18828 |
| 4 | Frontal_Sup_2_R | 8.129 | 3.567 | 12.690 | 2.327 | 3.493 | 0.026 | 4.79E-04 | 18645 | -13.297 | -21.427 | -5.166 | 4.148 | -3.206 | -0.023 | 1.35E-03 | 18828 |
| 5 | Frontal_Mid_2_L | 7.795 | 3.449 | 12.141 | 2.217 | 3.516 | 0.026 | 4.39E-04 | 18645 | -11.307 | -19.051 | -3.562 | 3.951 | -2.862 | -0.021 | 4.22E-03 | 18828 |
| 6 | Frontal_Mid_2_R | 9.854 | 4.938 | 14.771 | 2.508 | 3.929 | 0.029 | 8.57E-05 | 18645 | -17.799 | -26.561 | -9.037 | 4.470 | -3.982 | -0.029 | 6.87E-05 | 18828 |
| 7 | Frontal_Inf_Oper_L | 2.707 | 1.423 | 3.991 | 0.655 | 4.133 | 0.030 | 3.59E-05 | 18645 | -3.811 | -6.099 | -1.524 | 1.167 | -3.266 | -0.024 | 1.09E-03 | 18828 |
| 8 | Frontal_Inf_Oper_R | 2.556 | 0.946 | 4.166 | 0.821 | 3.112 | 0.023 | 1.86E-03 | 18645 | -5.472 | -8.342 | -2.602 | 1.464 | -3.738 | -0.027 | 1.86E-04 | 18828 |
| 9 | Frontal_Inf_Tri_L | 5.978 | 3.253 | 8.703 | 1.390 | 4.300 | 0.031 | 1.71E-05 | 18645 | -4.433 | -9.292 | 0.426 | 2.479 | -1.788 | -0.013 | 7.37E-02 | 18828 |
| 10 | Frontal_Inf_Tri_R | 3.724 | 1.585 | 5.863 | 1.091 | 3.413 | 0.025 | 6.45E-04 | 18645 | -5.815 | -9.632 | -1.998 | 1.947 | -2.986 | -0.022 | 2.83E-03 | 18828 |
| 11 | Frontal_Inf_Orb_2_L | 1.440 | 0.625 | 2.254 | 0.416 | 3.463 | 0.025 | 5.36E-04 | 18645 | -0.610 | -2.063 | 0.844 | 0.741 | -0.822 | -0.006 | 4.11E-01 | 18828 |
| 12 | Frontal_Inf_Orb_2_R | 1.613 | 0.762 | 2.465 | 0.434 | 3.713 | 0.027 | 2.05E-04 | 18645 | -1.833 | -3.352 | -0.314 | 0.775 | -2.365 | -0.017 | 1.81E-02 | 18828 |
| 13 | Rolandic_Oper_L | 0.654 | -0.579 | 1.886 | 0.629 | 1.039 | 0.008 | 2.99E-01 | 18645 | -2.290 | -4.487 | -0.093 | 1.121 | -2.043 | -0.015 | 4.11E-02 | 18828 |
| 14 | Rolandic_Oper_R | 1.311 | -0.453 | 3.075 | 0.900 | 1.457 | 0.011 | 1.45E-01 | 18645 | -4.906 | -8.050 | -1.762 | 1.604 | -3.059 | -0.022 | 2.23E-03 | 18828 |
| 15 | Supp_Motor_Area_L | 0.640 | -2.293 | 3.572 | 1.496 | 0.428 | 0.003 | 6.69E-01 | 18645 | -6.394 | -11.619 | -1.169 | 2.666 | -2.399 | -0.017 | 1.65E-02 | 18828 |
| 16 | Supp_Motor_Area_R | 1.488 | -1.750 | 4.726 | 1.652 | 0.901 | 0.007 | 3.68E-01 | 18645 | -6.634 | -12.408 | -0.860 | 2.946 | -2.252 | -0.016 | 2.43E-02 | 18828 |
| 17 | Olfactory_L | 0.407 | -0.028 | 0.843 | 0.222 | 1.833 | 0.013 | 6.68E-02 | 18645 | -0.194 | -0.971 | 0.582 | 0.396 | -0.490 | -0.004 | 6.24E-01 | 18828 |
| 18 | Olfactory_R | 0.387 | -0.050 | 0.824 | 0.223 | 1.737 | 0.013 | 8.24E-02 | 18645 | -0.711 | -1.489 | 0.068 | 0.397 | -1.789 | -0.013 | 7.37E-02 | 18828 |
| 19 | Frontal_Sup_Medial_L | 6.944 | 4.197 | 9.691 | 1.402 | 4.955 | 0.036 | 7.31E-07 | 18645 | -11.561 | -16.458 | -6.665 | 2.498 | -4.628 | -0.034 | 3.71E-06 | 18828 |
| 20 | Frontal_Sup_Medial_R | 4.755 | 2.756 | 6.754 | 1.020 | 4.662 | 0.034 | 3.16E-06 | 18645 | -6.422 | -9.986 | -2.859 | 1.818 | -3.533 | -0.026 | 4.12E-04 | 18828 |
| 21 | Frontal_Med_Orb_L | 2.098 | 1.071 | 3.125 | 0.524 | 4.005 | 0.029 | 6.23E-05 | 18645 | -3.104 | -4.935 | -1.274 | 0.934 | -3.325 | -0.024 | 8.87E-04 | 18828 |
| 22 | Frontal_Med_Orb_R | 2.579 | 1.317 | 3.842 | 0.644 | 4.005 | 0.029 | 6.22E-05 | 18645 | -3.369 | -5.618 | -1.120 | 1.147 | -2.937 | -0.021 | 3.32E-03 | 18828 |
| 23 | Rectus_L | 1.925 | 0.862 | 2.988 | 0.542 | 3.549 | 0.026 | 3.88E-04 | 18645 | -2.459 | -4.353 | -0.564 | 0.967 | -2.544 | -0.019 | 1.10E-02 | 18828 |
| 24 | Rectus_R | 2.197 | 1.214 | 3.181 | 0.502 | 4.379 | 0.032 | 1.20E-05 | 18645 | -2.185 | -3.937 | -0.432 | 0.894 | -2.443 | -0.018 | 1.46E-02 | 18828 |
| 25 | OFCmed_L | 0.989 | 0.373 | 1.605 | 0.314 | 3.146 | 0.023 | 1.66E-03 | 18645 | -1.344 | -2.442 | -0.246 | 0.560 | -2.400 | -0.017 | 1.64E-02 | 18828 |
| 26 | OFCmed_R | 1.265 | 0.548 | 1.982 | 0.366 | 3.456 | 0.025 | 5.49E-04 | 18645 | -1.533 | -2.813 | -0.254 | 0.653 | -2.349 | -0.017 | 1.88E-02 | 18828 |
| 27 | OFCant_L | 0.647 | -0.066 | 1.360 | 0.364 | 1.779 | 0.013 | 7.52E-02 | 18645 | -1.728 | -2.998 | -0.459 | 0.648 | -2.669 | -0.019 | 7.61E-03 | 18828 |
| 28 | OFCant_R | 1.370 | 0.431 | 2.310 | 0.479 | 2.858 | 0.021 | 4.26E-03 | 18645 | -2.884 | -4.560 | -1.208 | 0.855 | -3.373 | -0.025 | 7.46E-04 | 18828 |
| 29 | OFCpost_L | 0.658 | -0.233 | 1.549 | 0.455 | 1.447 | 0.011 | 1.48E-01 | 18645 | -1.521 | -3.109 | 0.068 | 0.811 | -1.876 | -0.014 | 6.06E-02 | 18828 |
| 30 | OFCpost_R | 1.075 | 0.251 | 1.899 | 0.421 | 2.557 | 0.019 | 1.06E-02 | 18645 | -2.130 | -3.598 | -0.662 | 0.749 | -2.844 | -0.021 | 4.46E-03 | 18828 |
| 31 | OFClat_L | 0.680 | 0.379 | 0.980 | 0.153 | 4.433 | 0.032 | 9.34E-06 | 18645 | -0.398 | -0.935 | 0.138 | 0.274 | -1.456 | -0.011 | 1.46E-01 | 18828 |
| 32 | OFClat_R | 0.715 | 0.461 | 0.969 | 0.129 | 5.524 | 0.040 | 3.37E-08 | 18645 | -0.775 | -1.227 | -0.322 | 0.231 | -3.354 | -0.024 | 7.99E-04 | 18828 |
| 33 | Insula_L | 1.823 | -0.936 | 4.581 | 1.407 | 1.295 | 0.009 | 1.95E-01 | 18645 | -6.386 | -11.299 | -1.472 | 2.507 | -2.547 | -0.019 | 1.09E-02 | 18828 |

| | | | | | | | | | | | | | | | | | |
|---|---|---|---|---|---|---|---|---|---|---|---|---|---|---|---|---|---|
| 34 | Insula_R | 1.377 | -1.313 | 4.067 | 1.372 | 1.004 | 0.007 | 3.16E-01 | 18645 | -7.398 | -12.187 | -2.609 | 2.443 | -3.028 | -0.022 | 2.47E-03 | 18828 |
| 37 | Cingulate_Mid_L | 5.237 | 2.904 | 7.570 | 1.190 | 4.400 | 0.032 | 1.09E-05 | 18645 | -13.122 | -17.276 | -8.968 | 2.119 | -6.192 | -0.045 | 6.08E-10 | 18828 |
| 38 | Cingulate_Mid_R | 3.343 | 0.855 | 5.831 | 1.269 | 2.634 | 0.019 | 8.45E-03 | 18645 | -12.023 | -16.452 | -7.594 | 2.260 | -5.321 | -0.039 | 1.04E-07 | 18828 |
| 39 | Cingulate_Post_L | -0.109 | -0.821 | 0.603 | 0.363 | -0.299 | -0.002 | 7.65E-01 | 18645 | -2.606 | -3.875 | -1.338 | 0.647 | -4.028 | -0.029 | 5.64E-05 | 18828 |
| 40 | Cingulate_Post_R | -0.330 | -0.725 | 0.066 | 0.202 | -1.635 | -0.012 | 1.02E-01 | 18645 | -0.965 | -1.669 | -0.261 | 0.359 | -2.687 | -0.020 | 7.22E-03 | 18828 |
| 41 | Hippocampus_L | 1.713 | 0.563 | 2.864 | 0.587 | 2.919 | 0.021 | 3.52E-03 | 18645 | -0.979 | -3.031 | 1.074 | 1.047 | -0.935 | -0.007 | 3.50E-01 | 18828 |
| 42 | Hippocampus_R | 2.161 | 0.967 | 3.355 | 0.609 | 3.547 | 0.026 | 3.90E-04 | 18645 | -0.691 | -2.820 | 1.437 | 1.086 | -0.637 | -0.005 | 5.24E-01 | 18828 |
| 43 | ParaHippocampal_L | 2.322 | 1.300 | 3.343 | 0.521 | 4.455 | 0.033 | 8.43E-06 | 18645 | -1.584 | -3.407 | 0.240 | 0.930 | -1.702 | -0.012 | 8.87E-02 | 18828 |
| 44 | ParaHippocampal_R | 2.537 | 1.245 | 3.829 | 0.659 | 3.849 | 0.028 | 1.19E-04 | 18645 | -1.260 | -3.561 | 1.042 | 1.174 | -1.073 | -0.008 | 2.83E-01 | 18828 |
| 45 | Amygdala_L | 0.182 | -0.216 | 0.580 | 0.203 | 0.897 | 0.007 | 3.69E-01 | 18645 | -0.151 | -0.860 | 0.557 | 0.362 | -0.419 | -0.003 | 6.76E-01 | 18828 |
| 46 | Amygdala_R | 0.241 | -0.166 | 0.648 | 0.208 | 1.160 | 0.008 | 2.46E-01 | 18645 | -0.508 | -1.233 | 0.216 | 0.370 | -1.375 | -0.010 | 1.69E-01 | 18828 |
| 47 | Calcarine_L | 1.003 | -2.788 | 4.794 | 1.934 | 0.518 | 0.004 | 6.04E-01 | 18645 | -1.398 | -8.158 | 5.363 | 3.449 | -0.405 | -0.003 | 6.85E-01 | 18828 |
| 48 | Calcarine_R | 0.105 | -3.065 | 3.275 | 1.617 | 0.065 | 0.000 | 9.48E-01 | 18645 | -1.649 | -7.297 | 3.999 | 2.882 | -0.572 | -0.004 | 5.67E-01 | 18828 |
| 49 | Cuneus_L | 0.791 | -1.590 | 3.171 | 1.215 | 0.651 | 0.005 | 5.15E-01 | 18645 | -2.188 | -6.430 | 2.055 | 2.164 | -1.011 | -0.007 | 3.12E-01 | 18828 |
| 50 | Cuneus_R | -0.040 | -1.861 | 1.782 | 0.929 | -0.043 | 0.000 | 9.66E-01 | 18645 | -4.964 | -8.208 | -1.719 | 1.655 | -2.999 | -0.022 | 2.72E-03 | 18828 |
| 51 | Lingual_L | 2.700 | 0.417 | 4.982 | 1.165 | 2.318 | 0.017 | 2.04E-02 | 18645 | -4.775 | -8.846 | -0.704 | 2.077 | -2.299 | -0.017 | 2.15E-02 | 18828 |
| 52 | Lingual_R | 1.797 | -0.593 | 4.187 | 1.220 | 1.473 | 0.011 | 1.41E-01 | 18645 | -2.604 | -6.867 | 1.658 | 2.175 | -1.198 | -0.009 | 2.31E-01 | 18828 |
| 53 | Occipital_Sup_L | 1.726 | 0.158 | 3.295 | 0.800 | 2.157 | 0.016 | 3.10E-02 | 18645 | -4.608 | -7.406 | -1.810 | 1.427 | -3.228 | -0.024 | 1.25E-03 | 18828 |
| 54 | Occipital_Sup_R | 0.738 | -0.964 | 2.439 | 0.868 | 0.850 | 0.006 | 3.95E-01 | 18645 | -4.721 | -7.752 | -1.690 | 1.546 | -3.053 | -0.022 | 2.27E-03 | 18828 |
| 55 | Occipital_Mid_L | -1.550 | -5.228 | 2.128 | 1.876 | -0.826 | -0.006 | 4.09E-01 | 18645 | -9.889 | -16.443 | -3.336 | 3.343 | -2.958 | -0.022 | 3.10E-03 | 18828 |
| 56 | Occipital_Mid_R | 0.196 | -2.365 | 2.757 | 1.307 | 0.150 | 0.001 | 8.81E-01 | 18645 | -3.520 | -8.083 | 1.042 | 2.328 | -1.512 | -0.011 | 1.30E-01 | 18828 |
| 57 | Occipital_Inf_L | -1.227 | -2.446 | -0.008 | 0.622 | -1.973 | -0.014 | 4.85E-02 | 18645 | -2.495 | -4.669 | -0.322 | 1.109 | -2.250 | -0.016 | 2.45E-02 | 18828 |
| 58 | Occipital_Inf_R | -2.853 | -4.263 | -1.444 | 0.719 | -3.968 | -0.029 | 7.29E-05 | 18645 | -0.650 | -3.162 | 1.861 | 1.281 | -0.507 | -0.004 | 6.12E-01 | 18828 |
| 59 | Fusiform_L | 4.382 | 1.712 | 7.053 | 1.362 | 3.217 | 0.024 | 1.30E-03 | 18645 | -6.404 | -11.166 | -1.642 | 2.429 | -2.636 | -0.019 | 8.39E-03 | 18828 |
| 60 | Fusiform_R | 5.001 | 2.066 | 7.936 | 1.497 | 3.340 | 0.024 | 8.39E-04 | 18645 | -4.069 | -9.299 | 1.160 | 2.668 | -1.525 | -0.011 | 1.27E-01 | 18828 |
| 61 | Postcentral_L | 9.490 | 5.476 | 13.505 | 2.048 | 4.634 | 0.034 | 3.62E-06 | 18645 | -14.397 | -21.558 | -7.236 | 3.653 | -3.941 | -0.029 | 8.15E-05 | 18828 |
| 62 | Postcentral_R | 9.560 | 5.445 | 13.675 | 2.099 | 4.554 | 0.033 | 5.31E-06 | 18645 | -11.606 | -18.942 | -4.270 | 3.743 | -3.101 | -0.023 | 1.93E-03 | 18828 |
| 63 | Parietal_Sup_L | 0.344 | -1.971 | 2.658 | 1.181 | 0.291 | 0.002 | 7.71E-01 | 18645 | -3.801 | -7.927 | 0.325 | 2.105 | -1.806 | -0.013 | 7.10E-02 | 18828 |
| 64 | Parietal_Sup_R | 0.553 | -1.501 | 2.606 | 1.048 | 0.527 | 0.004 | 5.98E-01 | 18645 | -4.487 | -8.145 | -0.829 | 1.866 | -2.404 | -0.018 | 1.62E-02 | 18828 |
| 65 | Parietal_Inf_L | 3.410 | 0.379 | 6.441 | 1.546 | 2.205 | 0.016 | 2.74E-02 | 18645 | -13.144 | -18.545 | -7.744 | 2.755 | -4.771 | -0.035 | 1.85E-06 | 18828 |
| 66 | Parietal_Inf_R | 1.279 | -0.687 | 3.245 | 1.003 | 1.275 | 0.009 | 2.02E-01 | 18645 | -6.196 | -9.699 | -2.693 | 1.787 | -3.467 | -0.025 | 5.27E-04 | 18828 |
| 67 | SupraMarginal_L | 1.643 | -0.133 | 3.419 | 0.906 | 1.813 | 0.013 | 6.98E-02 | 18645 | -5.852 | -9.019 | -2.686 | 1.615 | -3.622 | -0.026 | 2.93E-04 | 18828 |
| 68 | SupraMarginal_R | 3.738 | 1.225 | 6.251 | 1.282 | 2.916 | 0.021 | 3.55E-03 | 18645 | -11.121 | -15.604 | -6.639 | 2.287 | -4.863 | -0.035 | 1.16E-06 | 18828 |
| 69 | Angular_L | 2.150 | 0.405 | 3.896 | 0.891 | 2.414 | 0.018 | 1.58E-02 | 18645 | -6.692 | -9.803 | -3.582 | 1.587 | -4.217 | -0.031 | 2.49E-05 | 18828 |
| 70 | Angular_R | 1.747 | -0.594 | 4.088 | 1.194 | 1.463 | 0.011 | 1.44E-01 | 18645 | -5.294 | -9.463 | -1.125 | 2.127 | -2.489 | -0.018 | 1.28E-02 | 18828 |
| 71 | Precuneus_L | 3.143 | -1.115 | 7.401 | 2.172 | 1.447 | 0.011 | 1.48E-01 | 18645 | -9.885 | -17.469 | -2.300 | 3.870 | -2.554 | -0.019 | 1.06E-02 | 18828 |
| 72 | Precuneus_R | 1.126 | -2.665 | 4.917 | 1.934 | 0.582 | 0.004 | 5.60E-01 | 18645 | -11.043 | -17.796 | -4.290 | 3.445 | -3.205 | -0.023 | 1.35E-03 | 18828 |
| 73 | Paracentral_Lobule_L | -0.039 | -1.986 | 1.909 | 0.994 | -0.039 | 0.000 | 9.69E-01 | 18645 | -2.103 | -5.576 | 1.369 | 1.772 | -1.187 | -0.009 | 2.35E-01 | 18828 |
| 74 | Paracentral_Lobule_R | 0.584 | -0.811 | 1.980 | 0.712 | 0.821 | 0.006 | 4.12E-01 | 18645 | -1.043 | -3.530 | 1.445 | 1.269 | -0.822 | -0.006 | 4.11E-01 | 18828 |
| 75 | Caudate_L | 1.739 | 0.186 | 3.292 | 0.792 | 2.195 | 0.016 | 2.82E-02 | 18645 | -3.630 | -6.404 | -0.856 | 1.415 | -2.565 | -0.019 | 1.03E-02 | 18828 |
| 76 | Caudate_R | 2.283 | 0.522 | 4.044 | 0.898 | 2.542 | 0.019 | 1.10E-02 | 18645 | -4.863 | -8.005 | -1.721 | 1.603 | -3.033 | -0.022 | 2.42E-03 | 18828 |
| 77 | Putamen_L | 2.897 | 0.536 | 5.259 | 1.205 | 2.405 | 0.018 | 1.62E-02 | 18645 | -7.688 | -11.899 | -3.477 | 2.149 | -3.578 | -0.026 | 3.47E-04 | 18828 |
| 78 | Putamen_R | 2.477 | 0.072 | 4.883 | 1.227 | 2.019 | 0.015 | 4.35E-02 | 18645 | -7.993 | -12.284 | -3.703 | 2.189 | -3.652 | -0.027 | 2.61E-04 | 18828 |
| 79 | Pallidum_L | 0.137 | -0.074 | 0.347 | 0.107 | 1.274 | 0.009 | 2.03E-01 | 18645 | -0.490 | -0.865 | -0.116 | 0.191 | -2.567 | -0.019 | 1.03E-02 | 18828 |
| 80 | Pallidum_R | 0.116 | -0.115 | 0.346 | 0.118 | 0.983 | 0.007 | 3.26E-01 | 18645 | -0.677 | -1.088 | -0.266 | 0.210 | -3.231 | -0.024 | 1.24E-03 | 18828 |
| 83 | Heschl_L | 0.030 | -0.396 | 0.456 | 0.217 | 0.138 | 0.001 | 8.90E-01 | 18645 | -0.864 | -1.623 | -0.105 | 0.387 | -2.232 | -0.016 | 2.56E-02 | 18828 |
| 84 | Heschl_R | 0.186 | -0.290 | 0.662 | 0.243 | 0.766 | 0.006 | 4.44E-01 | 18645 | -1.369 | -2.217 | -0.521 | 0.433 | -3.164 | -0.023 | 1.56E-03 | 18828 |
| 85 | Temporal_Sup_L | 2.001 | -1.073 | 5.074 | 1.568 | 1.276 | 0.009 | 2.02E-01 | 18645 | -7.622 | -13.099 | -2.144 | 2.795 | -2.727 | -0.020 | 6.39E-03 | 18828 |
| 86 | Temporal_Sup_R | 4.799 | 1.398 | 8.200 | 1.735 | 2.766 | 0.020 | 5.69E-03 | 18645 | -13.935 | -19.997 | -7.873 | 3.093 | -4.506 | -0.033 | 6.65E-06 | 18828 |

| | | | | | | | | | | | | | | | | | |
|---|---|---|---|---|---|---|---|---|---|---|---|---|---|---|---|---|---|
| 87 | Temporal_Pole_Sup_L | 1.162 | -0.294 | 2.619 | 0.743 | 1.565 | 0.011 | 1.18E-01 | 18645 | -2.657 | -5.252 | -0.062 | 1.324 | -2.007 | -0.015 | 4.48E-02 | 18828 |
| 88 | Temporal_Pole_Sup_R | 1.961 | 0.428 | 3.495 | 0.783 | 2.507 | 0.018 | 1.22E-02 | 18645 | -4.371 | -7.102 | -1.640 | 1.393 | -3.138 | -0.023 | 1.71E-03 | 18828 |
| 89 | Temporal_Mid_L | 7.888 | 2.761 | 13.014 | 2.615 | 3.016 | 0.022 | 2.57E-03 | 18645 | -14.104 | -23.239 | -4.968 | 4.661 | -3.026 | -0.022 | 2.48E-03 | 18828 |
| 90 | Temporal_Mid_R | 3.822 | -0.729 | 8.373 | 2.322 | 1.646 | 0.012 | 9.98E-02 | 18645 | -15.293 | -23.400 | -7.187 | 4.136 | -3.698 | -0.027 | 2.18E-04 | 18828 |
| 91 | Temporal_Pole_Mid_L | 1.273 | 0.050 | 2.497 | 0.624 | 2.040 | 0.015 | 4.14E-02 | 18645 | -1.503 | -3.685 | 0.679 | 1.113 | -1.350 | -0.010 | 1.77E-01 | 18828 |
| 92 | Temporal_Pole_Mid_R | 2.081 | 0.595 | 3.568 | 0.758 | 2.744 | 0.020 | 6.07E-03 | 18645 | -2.673 | -5.321 | -0.024 | 1.351 | -1.978 | -0.014 | 4.79E-02 | 18828 |
| 93 | Temporal_Inf_L | 1.276 | -2.247 | 4.798 | 1.797 | 0.710 | 0.005 | 4.78E-01 | 18645 | -6.727 | -13.008 | -0.446 | 3.204 | -2.099 | -0.015 | 3.58E-02 | 18828 |
| 94 | Temporal_Inf_R | 0.036 | -3.808 | 3.880 | 1.961 | 0.018 | 0.000 | 9.85E-01 | 18645 | -7.495 | -14.335 | -0.654 | 3.490 | -2.147 | -0.016 | 3.18E-02 | 18828 |
| 95 | Cerebellum_Crus1_L | 5.318 | 1.130 | 9.506 | 2.137 | 2.489 | 0.018 | 1.28E-02 | 18645 | -1.521 | -8.980 | 5.939 | 3.806 | -0.400 | -0.003 | 6.89E-01 | 18828 |
| 96 | Cerebellum_Crus1_R | 6.703 | 2.436 | 10.971 | 2.177 | 3.079 | 0.023 | 2.08E-03 | 18645 | -1.875 | -9.480 | 5.730 | 3.880 | -0.483 | -0.004 | 6.29E-01 | 18828 |
| 97 | Cerebellum_Crus2_L | 7.351 | 4.033 | 10.670 | 1.693 | 4.343 | 0.032 | 1.42E-05 | 18645 | -6.735 | -12.653 | -0.818 | 3.019 | -2.231 | -0.016 | 2.57E-02 | 18828 |
| 98 | Cerebellum_Crus2_R | 6.999 | 3.932 | 10.067 | 1.565 | 4.473 | 0.033 | 7.78E-06 | 18645 | -4.485 | -9.955 | 0.985 | 2.791 | -1.607 | -0.012 | 1.08E-01 | 18828 |
| 99 | Cerebellum_3_L | 0.167 | -0.056 | 0.389 | 0.114 | 1.467 | 0.011 | 1.42E-01 | 18645 | 0.166 | -0.231 | 0.563 | 0.202 | 0.821 | 0.006 | 4.12E-01 | 18828 |
| 100 | Cerebellum_3_R | 0.301 | 0.000 | 0.601 | 0.153 | 1.963 | 0.014 | 4.97E-02 | 18645 | -0.088 | -0.624 | 0.448 | 0.273 | -0.322 | -0.002 | 7.48E-01 | 18828 |
| 101 | Cerebellum_4_5_L | 2.727 | 0.926 | 4.529 | 0.919 | 2.967 | 0.022 | 3.01E-03 | 18645 | -0.448 | -3.659 | 2.762 | 1.638 | -0.274 | -0.002 | 7.84E-01 | 18828 |
| 102 | Cerebellum_4_5_R | 2.262 | 0.794 | 3.729 | 0.749 | 3.020 | 0.022 | 2.53E-03 | 18645 | -1.056 | -3.672 | 1.560 | 1.335 | -0.791 | -0.006 | 4.29E-01 | 18828 |
| 103 | Cerebellum_6_L | 4.996 | 1.877 | 8.115 | 1.591 | 3.140 | 0.023 | 1.69E-03 | 18645 | -5.118 | -10.672 | 0.436 | 2.834 | -1.806 | -0.013 | 7.09E-02 | 18828 |
| 104 | Cerebellum_6_R | 5.902 | 2.618 | 9.186 | 1.676 | 3.522 | 0.026 | 4.29E-04 | 18645 | -2.743 | -8.593 | 3.107 | 2.985 | -0.919 | -0.007 | 3.58E-01 | 18828 |
| 105 | Cerebellum_7b_L | 2.614 | 1.600 | 3.629 | 0.518 | 5.051 | 0.037 | 4.43E-07 | 18645 | -1.705 | -3.514 | 0.104 | 0.923 | -1.847 | -0.013 | 6.47E-02 | 18828 |
| 106 | Cerebellum_7b_R | 3.147 | 2.142 | 4.151 | 0.513 | 6.137 | 0.045 | 8.57E-10 | 18645 | -1.968 | -3.760 | -0.176 | 0.914 | -2.153 | -0.016 | 3.14E-02 | 18828 |
| 107 | Cerebellum_8_L | 4.952 | 1.730 | 8.174 | 1.644 | 3.013 | 0.022 | 2.59E-03 | 18645 | -2.212 | -7.958 | 3.534 | 2.931 | -0.755 | -0.005 | 4.51E-01 | 18828 |
| 108 | Cerebellum_8_R | 10.552 | 6.358 | 14.746 | 2.140 | 4.932 | 0.036 | 8.23E-07 | 18645 | -8.507 | -15.984 | -1.030 | 3.815 | -2.230 | -0.016 | 2.58E-02 | 18828 |
| 109 | Cerebellum_9_L | -1.590 | -3.411 | 0.232 | 0.929 | -1.711 | -0.013 | 8.72E-02 | 18645 | -1.650 | -4.897 | 1.596 | 1.656 | -0.997 | -0.007 | 3.19E-01 | 18828 |
| 110 | Cerebellum_9_R | -1.430 | -3.222 | 0.361 | 0.914 | -1.565 | -0.011 | 1.18E-01 | 18645 | -1.686 | -4.878 | 1.506 | 1.628 | -1.035 | -0.008 | 3.01E-01 | 18828 |
| 111 | Cerebellum_10_L | -0.177 | -0.306 | -0.047 | 0.066 | -2.680 | -0.020 | 7.37E-03 | 18645 | -0.044 | -0.274 | 0.186 | 0.117 | -0.374 | -0.003 | 7.08E-01 | 18828 |
| 112 | Cerebellum_10_R | -0.023 | -0.158 | 0.111 | 0.069 | -0.341 | -0.002 | 7.33E-01 | 18645 | -0.067 | -0.307 | 0.172 | 0.122 | -0.550 | -0.004 | 5.82E-01 | 18828 |
| 113 | Vermis_1_2 | 0.033 | -0.051 | 0.117 | 0.043 | 0.760 | 0.006 | 4.47E-01 | 18645 | -0.019 | -0.169 | 0.130 | 0.076 | -0.255 | -0.002 | 7.99E-01 | 18828 |
| 114 | Vermis_3 | 0.373 | 0.040 | 0.707 | 0.170 | 2.193 | 0.016 | 2.84E-02 | 18645 | -0.428 | -1.023 | 0.167 | 0.304 | -1.409 | -0.010 | 1.59E-01 | 18828 |
| 115 | Vermis_4_5 | 1.039 | 0.059 | 2.019 | 0.500 | 2.078 | 0.015 | 3.77E-02 | 18645 | -0.284 | -2.033 | 1.465 | 0.892 | -0.318 | -0.002 | 7.50E-01 | 18828 |
| 116 | Vermis_6 | 0.098 | -0.448 | 0.644 | 0.279 | 0.352 | 0.003 | 7.25E-01 | 18645 | -0.258 | -1.232 | 0.716 | 0.497 | -0.520 | -0.004 | 6.03E-01 | 18828 |
| 117 | Vermis_7 | 0.134 | -0.175 | 0.443 | 0.158 | 0.851 | 0.006 | 3.95E-01 | 18645 | -0.133 | -0.684 | 0.417 | 0.281 | -0.474 | -0.003 | 6.35E-01 | 18828 |
| 118 | Vermis_8 | -0.078 | -0.586 | 0.430 | 0.259 | -0.303 | -0.002 | 7.62E-01 | 18645 | -0.603 | -1.508 | 0.303 | 0.462 | -1.305 | -0.010 | 1.92E-01 | 18828 |
| 119 | Vermis_9 | -0.285 | -0.697 | 0.127 | 0.210 | -1.355 | -0.010 | 1.76E-01 | 18645 | -0.047 | -0.781 | 0.687 | 0.374 | -0.125 | -0.001 | 9.01E-01 | 18828 |
| 120 | Vermis_10 | -0.124 | -0.239 | -0.009 | 0.059 | -2.114 | -0.015 | 3.45E-02 | 18645 | 0.126 | -0.079 | 0.330 | 0.104 | 1.204 | 0.009 | 2.29E-01 | 18828 |
| 121 | Thal_AV_L | 0.097 | 0.051 | 0.143 | 0.023 | 4.162 | 0.030 | 3.17E-05 | 18645 | -0.137 | -0.218 | -0.055 | 0.042 | -3.272 | -0.024 | 1.07E-03 | 18828 |
| 122 | Thal_AV_R | 0.156 | 0.090 | 0.221 | 0.033 | 4.675 | 0.034 | 2.96E-06 | 18645 | -0.169 | -0.286 | -0.052 | 0.060 | -2.829 | -0.021 | 4.68E-03 | 18828 |
| 123 | Thal_LP_L | 0.112 | 0.061 | 0.163 | 0.026 | 4.332 | 0.032 | 1.49E-05 | 18645 | -0.184 | -0.275 | -0.093 | 0.046 | -3.977 | -0.029 | 7.00E-05 | 18828 |
| 124 | Thal_LP_R | 0.154 | 0.088 | 0.220 | 0.034 | 4.574 | 0.033 | 4.83E-06 | 18645 | -0.247 | -0.365 | -0.129 | 0.060 | -4.099 | -0.030 | 4.17E-05 | 18828 |
| 125 | Thal_VA_L | 0.062 | 0.008 | 0.116 | 0.028 | 2.234 | 0.016 | 2.55E-02 | 18645 | -0.068 | -0.164 | 0.029 | 0.049 | -1.370 | -0.010 | 1.71E-01 | 18828 |
| 126 | Thal_VA_R | 0.108 | 0.041 | 0.174 | 0.034 | 3.170 | 0.023 | 1.53E-03 | 18645 | -0.088 | -0.207 | 0.031 | 0.061 | -1.456 | -0.011 | 1.45E-01 | 18828 |
| 127 | Thal_VL_L | 0.279 | 0.124 | 0.433 | 0.079 | 3.537 | 0.026 | 4.06E-04 | 18645 | -0.577 | -0.853 | -0.302 | 0.141 | -4.108 | -0.030 | 4.01E-05 | 18828 |
| 128 | Thal_VL_R | 0.563 | 0.358 | 0.767 | 0.104 | 5.388 | 0.039 | 7.20E-08 | 18645 | -0.794 | -1.159 | -0.428 | 0.186 | -4.257 | -0.031 | 2.08E-05 | 18828 |
| 129 | Thal_VPL_L | 0.187 | 0.003 | 0.371 | 0.094 | 1.996 | 0.015 | 4.59E-02 | 18645 | -0.493 | -0.820 | -0.165 | 0.167 | -2.949 | -0.021 | 3.20E-03 | 18828 |
| 130 | Thal_VPL_R | 0.367 | 0.173 | 0.560 | 0.099 | 3.718 | 0.027 | 2.01E-04 | 18645 | -0.699 | -1.043 | -0.355 | 0.176 | -3.978 | -0.029 | 6.97E-05 | 18828 |
| 131 | Thal_IL_L | 0.196 | 0.117 | 0.276 | 0.041 | 4.841 | 0.035 | 1.30E-06 | 18645 | -0.379 | -0.520 | -0.237 | 0.072 | -5.235 | -0.038 | 1.67E-07 | 18828 |
| 132 | Thal_IL_R | 0.272 | 0.182 | 0.362 | 0.046 | 5.929 | 0.043 | 3.10E-09 | 18645 | -0.404 | -0.565 | -0.244 | 0.082 | -4.928 | -0.036 | 8.39E-07 | 18828 |
| 133 | Thal_Re_L | 0.001 | 0.000 | 0.001 | 0.000 | 3.361 | 0.025 | 7.78E-04 | 18645 | 0.000 | -0.001 | 0.000 | 0.000 | -0.680 | -0.005 | 4.96E-01 | 18828 |
| 134 | Thal_Re_R | 0.007 | 0.004 | 0.009 | 0.001 | 4.852 | 0.036 | 1.23E-06 | 18645 | -0.006 | -0.011 | -0.001 | 0.002 | -2.347 | -0.017 | 1.89E-02 | 18828 |
| 135 | Thal_MDm_L | 1.722 | 1.134 | 2.310 | 0.300 | 5.744 | 0.042 | 9.39E-09 | 18645 | -2.876 | -3.926 | -1.827 | 0.536 | -5.371 | -0.039 | 7.93E-08 | 18828 |

| | | | | | | | | | | | | | | | | | |
|---|---|---|---|---|---|---|---|---|---|---|---|---|---|---|---|---|---|
| 136 | Thal_MDm_R | 2.090 | 1.408 | 2.772 | 0.348 | 6.004 | 0.044 | 1.96E-09 | 18645 | -3.086 | -4.306 | -1.866 | 0.623 | -4.957 | -0.036 | 7.21E-07 | 18828 |
| 137 | Thal_MDl_L | 0.315 | 0.213 | 0.417 | 0.052 | 6.058 | 0.044 | 1.40E-09 | 18645 | -0.533 | -0.715 | -0.351 | 0.093 | -5.750 | -0.042 | 9.06E-09 | 18828 |
| 138 | Thal_MDl_R | 0.375 | 0.260 | 0.491 | 0.059 | 6.362 | 0.047 | 2.04E-10 | 18645 | -0.546 | -0.753 | -0.340 | 0.105 | -5.182 | -0.038 | 2.21E-07 | 18828 |
| 139 | Thal_LGN_L | 0.028 | 0.012 | 0.043 | 0.008 | 3.523 | 0.026 | 4.27E-04 | 18645 | -0.049 | -0.076 | -0.021 | 0.014 | -3.471 | -0.025 | 5.19E-04 | 18828 |
| 140 | Thal_LGN_R | 0.083 | 0.050 | 0.116 | 0.017 | 4.931 | 0.036 | 8.27E-07 | 18645 | -0.150 | -0.209 | -0.091 | 0.030 | -5.010 | -0.036 | 5.50E-07 | 18828 |
| 141 | Thal_MGN_L | 0.046 | 0.018 | 0.073 | 0.014 | 3.233 | 0.024 | 1.23E-03 | 18645 | -0.130 | -0.179 | -0.081 | 0.025 | -5.178 | -0.038 | 2.27E-07 | 18828 |
| 142 | Thal_MGN_R | 0.067 | 0.037 | 0.097 | 0.015 | 4.374 | 0.032 | 1.22E-05 | 18645 | -0.145 | -0.198 | -0.091 | 0.027 | -5.293 | -0.039 | 1.22E-07 | 18828 |
| 143 | Thal_PuA_L | 0.064 | 0.031 | 0.097 | 0.017 | 3.836 | 0.028 | 1.25E-04 | 18645 | -0.163 | -0.221 | -0.105 | 0.030 | -5.492 | -0.040 | 4.04E-08 | 18828 |
| 144 | Thal_PuA_R | 0.166 | 0.098 | 0.234 | 0.035 | 4.801 | 0.035 | 1.59E-06 | 18645 | -0.340 | -0.461 | -0.219 | 0.062 | -5.513 | -0.040 | 3.57E-08 | 18828 |
| 145 | Thal_PuM_L | 1.561 | 0.913 | 2.210 | 0.331 | 4.721 | 0.035 | 2.36E-06 | 18645 | -3.015 | -4.171 | -1.860 | 0.589 | -5.115 | -0.037 | 3.16E-07 | 18828 |
| 146 | Thal_PuM_R | 2.021 | 1.214 | 2.829 | 0.412 | 4.908 | 0.036 | 9.29E-07 | 18645 | -3.602 | -5.042 | -2.162 | 0.735 | -4.903 | -0.036 | 9.52E-07 | 18828 |
| 147 | Thal_PuL_L | 0.281 | 0.163 | 0.400 | 0.060 | 4.664 | 0.034 | 3.12E-06 | 18645 | -0.522 | -0.733 | -0.311 | 0.108 | -4.853 | -0.035 | 1.22E-06 | 18828 |
| 148 | Thal_PuL_R | 0.466 | 0.278 | 0.654 | 0.096 | 4.856 | 0.036 | 1.20E-06 | 18645 | -0.827 | -1.162 | -0.491 | 0.171 | -4.832 | -0.035 | 1.36E-06 | 18828 |
| 149 | Thal_PuI_L | 0.028 | -0.005 | 0.060 | 0.016 | 1.675 | 0.012 | 9.39E-02 | 18645 | -0.063 | -0.121 | -0.006 | 0.029 | -2.161 | -0.016 | 3.07E-02 | 18828 |
| 150 | Thal_PuI_R | 0.071 | 0.029 | 0.113 | 0.022 | 3.277 | 0.024 | 1.05E-03 | 18645 | -0.127 | -0.203 | -0.051 | 0.039 | -3.289 | -0.024 | 1.01E-03 | 18828 |
| 151 | ACC_sub_L | -0.072 | -0.444 | 0.299 | 0.190 | -0.381 | -0.003 | 7.03E-01 | 18645 | -0.708 | -1.370 | -0.046 | 0.338 | -2.097 | -0.015 | 3.60E-02 | 18828 |
| 152 | ACC_sub_R | -0.094 | -0.441 | 0.254 | 0.177 | -0.529 | -0.004 | 5.97E-01 | 18645 | -0.587 | -1.205 | 0.031 | 0.315 | -1.862 | -0.014 | 6.26E-02 | 18828 |
| 153 | ACC_pre_L | 0.944 | -0.008 | 1.897 | 0.486 | 1.943 | 0.014 | 5.20E-02 | 18645 | -3.453 | -5.149 | -1.756 | 0.865 | -3.990 | -0.029 | 6.63E-05 | 18828 |
| 154 | ACC_pre_R | 0.847 | -0.070 | 1.764 | 0.468 | 1.811 | 0.013 | 7.02E-02 | 18645 | -3.558 | -5.191 | -1.924 | 0.833 | -4.270 | -0.031 | 1.97E-05 | 18828 |
| 155 | ACC_sup_L | 0.688 | -0.348 | 1.724 | 0.529 | 1.302 | 0.010 | 1.93E-01 | 18645 | -3.154 | -4.997 | -1.311 | 0.940 | -3.354 | -0.024 | 7.99E-04 | 18828 |
| 156 | ACC_sup_R | 0.304 | -0.768 | 1.376 | 0.547 | 0.556 | 0.004 | 5.78E-01 | 18645 | -2.766 | -4.676 | -0.857 | 0.974 | -2.839 | -0.021 | 4.52E-03 | 18828 |
| 157 | N_Acc_L | 0.517 | 0.205 | 0.828 | 0.159 | 3.253 | 0.024 | 1.15E-03 | 18645 | -0.442 | -0.997 | 0.113 | 0.283 | -1.560 | -0.011 | 1.19E-01 | 18828 |
| 158 | N_Acc_R | 0.315 | 0.046 | 0.585 | 0.138 | 2.292 | 0.017 | 2.19E-02 | 18645 | -0.510 | -0.990 | -0.030 | 0.245 | -2.083 | -0.015 | 3.73E-02 | 18828 |
| 159 | VTA_L | 0.014 | 0.003 | 0.025 | 0.006 | 2.480 | 0.018 | 1.32E-02 | 18645 | 0.003 | -0.017 | 0.023 | 0.010 | 0.302 | 0.002 | 7.63E-01 | 18828 |
| 160 | VTA_R | 0.015 | 0.005 | 0.025 | 0.005 | 2.929 | 0.021 | 3.40E-03 | 18645 | -0.009 | -0.026 | 0.009 | 0.009 | -0.972 | -0.007 | 3.31E-01 | 18828 |
| 161 | SN_pc_L | 0.009 | -0.009 | 0.027 | 0.009 | 0.999 | 0.007 | 3.18E-01 | 18645 | -0.044 | -0.075 | -0.013 | 0.016 | -2.748 | -0.020 | 6.01E-03 | 18828 |
| 162 | SN_pc_R | 0.017 | -0.006 | 0.040 | 0.012 | 1.470 | 0.011 | 1.42E-01 | 18645 | -0.065 | -0.106 | -0.024 | 0.021 | -3.091 | -0.023 | 2.00E-03 | 18828 |
| 163 | SN_pr_L | 0.008 | -0.016 | 0.032 | 0.012 | 0.622 | 0.005 | 5.34E-01 | 18645 | -0.039 | -0.082 | 0.003 | 0.022 | -1.817 | -0.013 | 6.92E-02 | 18828 |
| 164 | SN_pr_R | 0.013 | -0.014 | 0.040 | 0.014 | 0.949 | 0.007 | 3.42E-01 | 18645 | -0.037 | -0.085 | 0.011 | 0.024 | -1.516 | -0.011 | 1.29E-01 | 18828 |
| 165 | Red_N_L | 0.044 | 0.015 | 0.074 | 0.015 | 2.939 | 0.022 | 3.30E-03 | 18645 | -0.067 | -0.119 | -0.014 | 0.027 | -2.492 | -0.018 | 1.27E-02 | 18828 |
| 166 | Red_N_R | 0.047 | 0.015 | 0.079 | 0.016 | 2.856 | 0.021 | 4.29E-03 | 18645 | -0.065 | -0.122 | -0.008 | 0.029 | -2.237 | -0.016 | 2.53E-02 | 18828 |
| 167 | LC_L | 0.003 | -0.008 | 0.014 | 0.006 | 0.489 | 0.004 | 6.25E-01 | 18645 | 0.006 | -0.014 | 0.026 | 0.010 | 0.609 | 0.004 | 5.42E-01 | 18828 |
| 168 | LC_R | -0.002 | -0.015 | 0.010 | 0.006 | -0.406 | -0.003 | 6.85E-01 | 18645 | 0.001 | -0.020 | 0.023 | 0.011 | 0.113 | 0.001 | 9.10E-01 | 18828 |
| 169 | Raphe_D | 0.096 | 0.053 | 0.139 | 0.022 | 4.413 | 0.032 | 1.02E-05 | 18645 | -0.100 | -0.176 | -0.023 | 0.039 | -2.564 | -0.019 | 1.04E-02 | 18828 |
| 170 | Raphe_M | 0.014 | 0.001 | 0.027 | 0.007 | 2.180 | 0.016 | 2.93E-02 | 18645 | 0.002 | -0.021 | 0.025 | 0.012 | 0.167 | 0.001 | 8.68E-01 | 18828 |

| | | Bread intake | | | | | | | | Pork intake | | | | | | | |
|---|---|---|---|---|---|---|---|---|---|---|---|---|---|---|---|---|---|
| Number | AAL 3 region | beta | 95%CI_l | 95%CI_h | se | t | r | p | df.e | beta | 95%CI_l | 95%CI_h | se | t | r | p | df.e |
| 0 | Total brain | 8.626 | -8.644 | 25.895 | 8.811 | 0.979 | 0.007 | 3.28E-01 | 18599 | -469.130 | -663.863 | -274.398 | 99.349 | -4.722 | -0.034 | 2.35E-06 | 18827 |
| 1 | Precentral_L | -0.062 | -0.669 | 0.546 | 0.310 | -0.200 | -0.001 | 8.42E-01 | 18599 | -6.083 | -12.930 | 0.764 | 3.493 | -1.741 | -0.013 | 8.16E-02 | 18827 |
| 2 | Precentral_R | 0.311 | -0.246 | 0.867 | 0.284 | 1.094 | 0.008 | 2.74E-01 | 18599 | -9.029 | -15.302 | -2.757 | 3.200 | -2.822 | -0.021 | 4.78E-03 | 18827 |
| 3 | Frontal_Sup_2_L | 0.954 | 0.312 | 1.596 | 0.327 | 2.914 | 0.021 | 3.58E-03 | 18599 | -10.291 | -17.524 | -3.057 | 3.690 | -2.789 | -0.020 | 5.30E-03 | 18827 |
| 4 | Frontal_Sup_2_R | 0.805 | 0.097 | 1.512 | 0.361 | 2.230 | 0.016 | 2.58E-02 | 18599 | -13.739 | -21.713 | -5.765 | 4.068 | -3.377 | -0.025 | 7.34E-04 | 18827 |
| 5 | Frontal_Mid_2_L | 0.163 | -0.511 | 0.836 | 0.344 | 0.474 | 0.003 | 6.36E-01 | 18599 | -9.292 | -16.887 | -1.697 | 3.875 | -2.398 | -0.017 | 1.65E-02 | 18827 |
| 6 | Frontal_Mid_2_R | 0.845 | 0.083 | 1.607 | 0.389 | 2.175 | 0.016 | 2.97E-02 | 18599 | -14.527 | -23.120 | -5.934 | 4.384 | -3.314 | -0.024 | 9.23E-04 | 18827 |
| 7 | Frontal_Inf_Oper_L | 0.063 | -0.136 | 0.261 | 0.101 | 0.616 | 0.005 | 5.38E-01 | 18599 | -2.387 | -4.631 | -0.142 | 1.145 | -2.084 | -0.015 | 3.71E-02 | 18827 |
| 8 | Frontal_Inf_Oper_R | 0.251 | 0.002 | 0.501 | 0.127 | 1.972 | 0.014 | 4.86E-02 | 18599 | -4.495 | -7.309 | -1.681 | 1.436 | -3.130 | -0.023 | 1.75E-03 | 18827 |
| 9 | Frontal_Inf_Tri_L | 0.242 | -0.181 | 0.664 | 0.216 | 1.120 | 0.008 | 2.63E-01 | 18599 | -8.551 | -13.314 | -3.787 | 2.430 | -3.519 | -0.026 | 4.35E-04 | 18827 |
| 10 | Frontal_Inf_Tri_R | 0.179 | -0.153 | 0.511 | 0.169 | 1.058 | 0.008 | 2.90E-01 | 18599 | -5.945 | -9.687 | -2.203 | 1.909 | -3.114 | -0.023 | 1.85E-03 | 18827 |
| 11 | Frontal_Inf_Orb_2_L | 0.145 | 0.019 | 0.271 | 0.064 | 2.252 | 0.017 | 2.43E-02 | 18599 | -2.020 | -3.445 | -0.595 | 0.727 | -2.778 | -0.020 | 5.47E-03 | 18827 |

| | | | | | | | | | | | | | | | | | |
|---|---|---|---|---|---|---|---|---|---|---|---|---|---|---|---|---|---|
| 12 | Frontal_Inf_Orb_2_R | 0.109 | -0.023 | 0.241 | 0.067 | 1.614 | 0.012 | 1.07E-01 | 18599 | -2.141 | -3.631 | -0.652 | 0.760 | -2.817 | -0.021 | 4.85E-03 | 18827 |
| 13 | Rolandic_Oper_L | 0.082 | -0.109 | 0.273 | 0.098 | 0.840 | 0.006 | 4.01E-01 | 18599 | -2.620 | -4.776 | -0.465 | 1.100 | -2.383 | -0.017 | 1.72E-02 | 18827 |
| 14 | Rolandic_Oper_R | 0.139 | -0.135 | 0.413 | 0.140 | 0.995 | 0.007 | 3.20E-01 | 18599 | -6.203 | -9.287 | -3.118 | 1.574 | -3.941 | -0.029 | 8.13E-05 | 18827 |
| 15 | Supp_Motor_Area_L | 0.139 | -0.316 | 0.594 | 0.232 | 0.599 | 0.004 | 5.49E-01 | 18599 | -2.585 | -7.710 | 2.539 | 2.615 | -0.989 | -0.007 | 3.23E-01 | 18827 |
| 16 | Supp_Motor_Area_R | 0.127 | -0.375 | 0.629 | 0.256 | 0.497 | 0.004 | 6.19E-01 | 18599 | -2.650 | -8.313 | 3.013 | 2.889 | -0.917 | -0.007 | 3.59E-01 | 18827 |
| 17 | Olfactory_L | 0.032 | -0.036 | 0.099 | 0.034 | 0.921 | 0.007 | 3.57E-01 | 18599 | -0.978 | -1.740 | -0.216 | 0.389 | -2.517 | -0.018 | 1.18E-02 | 18827 |
| 18 | Olfactory_R | 0.041 | -0.027 | 0.109 | 0.035 | 1.191 | 0.009 | 2.34E-01 | 18599 | -1.358 | -2.122 | -0.595 | 0.390 | -3.486 | -0.025 | 4.91E-04 | 18827 |
| 19 | Frontal_Sup_Medial_L | 0.524 | 0.098 | 0.950 | 0.217 | 2.411 | 0.018 | 1.59E-02 | 18599 | -6.959 | -11.763 | -2.156 | 2.451 | -2.840 | -0.021 | 4.52E-03 | 18827 |
| 20 | Frontal_Sup_Medial_R | 0.203 | -0.106 | 0.513 | 0.158 | 1.287 | 0.009 | 1.98E-01 | 18599 | -6.337 | -9.831 | -2.843 | 1.783 | -3.555 | -0.026 | 3.80E-04 | 18827 |
| 21 | Frontal_Med_Orb_L | 0.188 | 0.029 | 0.348 | 0.081 | 2.322 | 0.017 | 2.02E-02 | 18599 | -3.184 | -4.978 | -1.390 | 0.915 | -3.478 | -0.025 | 5.06E-04 | 18827 |
| 22 | Frontal_Med_Orb_R | 0.089 | -0.107 | 0.284 | 0.100 | 0.891 | 0.007 | 3.73E-01 | 18599 | -3.257 | -5.462 | -1.052 | 1.125 | -2.895 | -0.021 | 3.79E-03 | 18827 |
| 23 | Rectus_L | 0.157 | -0.007 | 0.322 | 0.084 | 1.873 | 0.014 | 6.11E-02 | 18599 | -2.981 | -4.838 | -1.123 | 0.948 | -3.145 | -0.023 | 1.66E-03 | 18827 |
| 24 | Rectus_R | 0.115 | -0.037 | 0.268 | 0.078 | 1.486 | 0.011 | 1.37E-01 | 18599 | -2.134 | -3.853 | -0.415 | 0.877 | -2.434 | -0.018 | 1.50E-02 | 18827 |
| 25 | OFCmed_L | -0.002 | -0.097 | 0.094 | 0.049 | -0.033 | 0.000 | 9.74E-01 | 18599 | -1.505 | -2.582 | -0.428 | 0.549 | -2.740 | -0.020 | 6.15E-03 | 18827 |
| 26 | OFCmed_R | 0.037 | -0.075 | 0.148 | 0.057 | 0.646 | 0.005 | 5.18E-01 | 18599 | -0.858 | -2.113 | 0.397 | 0.640 | -1.340 | -0.010 | 1.80E-01 | 18827 |
| 27 | OFCant_L | -0.031 | -0.141 | 0.080 | 0.056 | -0.544 | -0.004 | 5.86E-01 | 18599 | -1.567 | -2.811 | -0.322 | 0.635 | -2.468 | -0.018 | 1.36E-02 | 18827 |
| 28 | OFCant_R | 0.080 | -0.066 | 0.225 | 0.074 | 1.071 | 0.008 | 2.84E-01 | 18599 | -2.210 | -3.854 | -0.566 | 0.839 | -2.635 | -0.019 | 8.42E-03 | 18827 |
| 29 | OFCpost_L | 0.170 | 0.032 | 0.308 | 0.070 | 2.411 | 0.018 | 1.59E-02 | 18599 | -2.455 | -4.012 | -0.897 | 0.795 | -3.089 | -0.023 | 2.01E-03 | 18827 |
| 30 | OFCpost_R | 0.158 | 0.030 | 0.286 | 0.065 | 2.424 | 0.018 | 1.53E-02 | 18599 | -2.843 | -4.282 | -1.404 | 0.734 | -3.871 | -0.028 | 1.09E-04 | 18827 |
| 31 | OFClat_L | 0.022 | -0.025 | 0.068 | 0.024 | 0.911 | 0.007 | 3.62E-01 | 18599 | -0.632 | -1.157 | -0.106 | 0.268 | -2.355 | -0.017 | 1.85E-02 | 18827 |
| 32 | OFClat_R | 0.017 | -0.023 | 0.056 | 0.020 | 0.828 | 0.006 | 4.08E-01 | 18599 | -0.644 | -1.088 | -0.200 | 0.227 | -2.842 | -0.021 | 4.49E-03 | 18827 |
| 33 | Insula_L | 0.440 | 0.012 | 0.867 | 0.218 | 2.015 | 0.015 | 4.39E-02 | 18599 | -11.925 | -16.742 | -7.108 | 2.457 | -4.853 | -0.035 | 1.23E-06 | 18827 |
| 34 | Insula_R | 0.518 | 0.102 | 0.935 | 0.212 | 2.440 | 0.018 | 1.47E-02 | 18599 | -10.730 | -15.427 | -6.033 | 2.396 | -4.478 | -0.033 | 7.58E-06 | 18827 |
| 37 | Cingulate_Mid_L | 0.385 | 0.024 | 0.747 | 0.184 | 2.090 | 0.015 | 3.66E-02 | 18599 | -10.043 | -14.117 | -5.968 | 2.079 | -4.831 | -0.035 | 1.37E-06 | 18827 |
| 38 | Cingulate_Mid_R | 0.421 | 0.036 | 0.806 | 0.196 | 2.145 | 0.016 | 3.19E-02 | 18599 | -10.113 | -14.456 | -5.770 | 2.216 | -4.564 | -0.033 | 5.04E-06 | 18827 |
| 39 | Cingulate_Post_L | 0.068 | -0.042 | 0.179 | 0.056 | 1.215 | 0.009 | 2.24E-01 | 18599 | -2.355 | -3.599 | -1.111 | 0.635 | -3.711 | -0.027 | 2.07E-04 | 18827 |
| 40 | Cingulate_Post_R | 0.012 | -0.049 | 0.073 | 0.031 | 0.387 | 0.003 | 6.99E-01 | 18599 | -0.758 | -1.448 | -0.068 | 0.352 | -2.154 | -0.016 | 3.13E-02 | 18827 |
| 41 | Hippocampus_L | 0.182 | 0.004 | 0.361 | 0.091 | 1.999 | 0.015 | 4.56E-02 | 18599 | -2.717 | -4.729 | -0.705 | 1.027 | -2.647 | -0.019 | 8.13E-03 | 18827 |
| 42 | Hippocampus_R | 0.113 | -0.072 | 0.299 | 0.094 | 1.200 | 0.009 | 2.30E-01 | 18599 | -3.011 | -5.099 | -0.923 | 1.065 | -2.827 | -0.021 | 4.71E-03 | 18827 |
| 43 | ParaHippocampal_L | 0.172 | 0.014 | 0.331 | 0.081 | 2.129 | 0.016 | 3.33E-02 | 18599 | -2.761 | -4.549 | -0.973 | 0.912 | -3.027 | -0.022 | 2.47E-03 | 18827 |
| 44 | ParaHippocampal_R | 0.267 | 0.067 | 0.467 | 0.102 | 2.614 | 0.019 | 8.97E-03 | 18599 | -4.210 | -6.467 | -1.953 | 1.151 | -3.657 | -0.027 | 2.56E-04 | 18827 |
| 45 | Amygdala_L | 0.089 | 0.028 | 0.151 | 0.031 | 2.848 | 0.021 | 4.41E-03 | 18599 | -0.611 | -1.305 | 0.084 | 0.355 | -1.722 | -0.013 | 8.50E-02 | 18827 |
| 46 | Amygdala_R | 0.090 | 0.027 | 0.153 | 0.032 | 2.785 | 0.020 | 5.37E-03 | 18599 | -1.013 | -1.723 | -0.302 | 0.363 | -2.792 | -0.020 | 5.25E-03 | 18827 |
| 47 | Calcarine_L | -0.899 | -1.487 | -0.312 | 0.300 | -2.999 | -0.022 | 2.71E-03 | 18599 | -1.698 | -8.326 | 4.931 | 3.382 | -0.502 | -0.004 | 6.16E-01 | 18827 |
| 48 | Calcarine_R | -0.844 | -1.335 | -0.353 | 0.251 | -3.368 | -0.025 | 7.60E-04 | 18599 | -1.720 | -7.260 | 3.820 | 2.826 | -0.609 | -0.004 | 5.43E-01 | 18827 |
| 49 | Cuneus_L | -0.593 | -0.962 | -0.223 | 0.188 | -3.145 | -0.023 | 1.67E-03 | 18599 | -1.518 | -5.677 | 2.641 | 2.122 | -0.715 | -0.005 | 4.74E-01 | 18827 |
| 50 | Cuneus_R | -0.352 | -0.634 | -0.069 | 0.144 | -2.443 | -0.018 | 1.46E-02 | 18599 | -1.862 | -5.043 | 1.319 | 1.623 | -1.147 | -0.008 | 2.51E-01 | 18827 |
| 51 | Lingual_L | -0.309 | -0.663 | 0.045 | 0.181 | -1.709 | -0.013 | 8.74E-02 | 18599 | -3.272 | -7.264 | 0.719 | 2.037 | -1.607 | -0.012 | 1.08E-01 | 18827 |
| 52 | Lingual_R | -0.238 | -0.609 | 0.132 | 0.189 | -1.260 | -0.009 | 2.08E-01 | 18599 | -3.606 | -7.785 | 0.574 | 2.132 | -1.691 | -0.012 | 9.09E-02 | 18827 |
| 53 | Occipital_Sup_L | -0.109 | -0.352 | 0.134 | 0.124 | -0.880 | -0.006 | 3.79E-01 | 18599 | -3.181 | -5.925 | -0.438 | 1.399 | -2.273 | -0.017 | 2.30E-02 | 18827 |
| 54 | Occipital_Sup_R | 0.060 | -0.204 | 0.324 | 0.135 | 0.444 | 0.003 | 6.57E-01 | 18599 | -3.185 | -6.157 | -0.212 | 1.516 | -2.100 | -0.015 | 3.57E-02 | 18827 |
| 55 | Occipital_Mid_L | -0.175 | -0.746 | 0.395 | 0.291 | -0.603 | -0.004 | 5.47E-01 | 18599 | -6.196 | -12.621 | 0.228 | 3.278 | -1.891 | -0.014 | 5.87E-02 | 18827 |
| 56 | Occipital_Mid_R | 0.144 | -0.253 | 0.541 | 0.202 | 0.711 | 0.005 | 4.77E-01 | 18599 | -4.238 | -8.713 | 0.236 | 2.283 | -1.857 | -0.014 | 6.34E-02 | 18827 |
| 57 | Occipital_Inf_L | -0.003 | -0.193 | 0.186 | 0.097 | -0.036 | 0.000 | 9.71E-01 | 18599 | -1.980 | -4.112 | 0.152 | 1.088 | -1.820 | -0.013 | 6.88E-02 | 18827 |
| 58 | Occipital_Inf_R | 0.155 | -0.064 | 0.374 | 0.112 | 1.386 | 0.010 | 1.66E-01 | 18599 | 0.925 | -1.539 | 3.389 | 1.257 | 0.736 | 0.005 | 4.62E-01 | 18827 |
| 59 | Fusiform_L | -0.088 | -0.503 | 0.327 | 0.212 | -0.416 | -0.003 | 6.77E-01 | 18599 | -4.348 | -9.018 | 0.322 | 2.382 | -1.825 | -0.013 | 6.80E-02 | 18827 |
| 60 | Fusiform_R | 0.246 | -0.209 | 0.701 | 0.232 | 1.059 | 0.008 | 2.90E-01 | 18599 | -5.571 | -10.701 | -0.441 | 2.617 | -2.129 | -0.016 | 3.33E-02 | 18827 |
| 61 | Postcentral_L | 0.336 | -0.287 | 0.960 | 0.318 | 1.058 | 0.008 | 2.90E-01 | 18599 | -7.385 | -14.407 | -0.364 | 3.582 | -2.062 | -0.015 | 3.93E-02 | 18827 |
| 62 | Postcentral_R | 0.378 | -0.261 | 1.017 | 0.326 | 1.160 | 0.009 | 2.46E-01 | 18599 | -8.005 | -15.202 | -0.807 | 3.672 | -2.180 | -0.016 | 2.93E-02 | 18827 |

| | | | | | | | | | | | | | | | | | |
|---|---|---|---|---|---|---|---|---|---|---|---|---|---|---|---|---|---|
| 63 | Parietal_Sup_L | -0.165 | -0.524 | 0.195 | 0.183 | -0.897 | -0.007 | 3.69E-01 | 18599 | -3.420 | -7.467 | 0.627 | 2.065 | -1.656 | -0.012 | 9.77E-02 | 18827 |
| 64 | Parietal_Sup_R | 0.072 | -0.247 | 0.391 | 0.163 | 0.443 | 0.003 | 6.58E-01 | 18599 | -5.405 | -8.992 | -1.818 | 1.830 | -2.953 | -0.022 | 3.15E-03 | 18827 |
| 65 | Parietal_Inf_L | -0.130 | -0.601 | 0.340 | 0.240 | -0.543 | -0.004 | 5.87E-01 | 18599 | -8.564 | -13.861 | -3.266 | 2.703 | -3.168 | -0.023 | 1.53E-03 | 18827 |
| 66 | Parietal_Inf_R | -0.021 | -0.326 | 0.284 | 0.156 | -0.133 | -0.001 | 8.94E-01 | 18599 | -4.549 | -7.985 | -1.114 | 1.753 | -2.595 | -0.019 | 9.46E-03 | 18827 |
| 67 | SupraMarginal_L | 0.220 | -0.056 | 0.495 | 0.141 | 1.562 | 0.011 | 1.18E-01 | 18599 | -5.344 | -8.448 | -2.239 | 1.584 | -3.374 | -0.025 | 7.43E-04 | 18827 |
| 68 | SupraMarginal_R | 0.223 | -0.167 | 0.613 | 0.199 | 1.120 | 0.008 | 2.63E-01 | 18599 | -7.119 | -11.517 | -2.721 | 2.244 | -3.173 | -0.023 | 1.51E-03 | 18827 |
| 69 | Angular_L | -0.074 | -0.345 | 0.197 | 0.138 | -0.538 | -0.004 | 5.91E-01 | 18599 | -4.615 | -7.665 | -1.565 | 1.556 | -2.966 | -0.022 | 3.02E-03 | 18827 |
| 70 | Angular_R | -0.303 | -0.666 | 0.060 | 0.185 | -1.638 | -0.012 | 1.01E-01 | 18599 | -5.445 | -9.534 | -1.356 | 2.086 | -2.610 | -0.019 | 9.05E-03 | 18827 |
| 71 | Precuneus_L | -0.213 | -0.873 | 0.447 | 0.337 | -0.634 | -0.005 | 5.26E-01 | 18599 | -8.747 | -16.186 | -1.308 | 3.795 | -2.305 | -0.017 | 2.12E-02 | 18827 |
| 72 | Precuneus_R | -0.267 | -0.854 | 0.321 | 0.300 | -0.890 | -0.007 | 3.74E-01 | 18599 | -6.891 | -13.517 | -0.266 | 3.380 | -2.039 | -0.015 | 4.15E-02 | 18827 |
| 73 | Paracentral_Lobule_L | -0.075 | -0.377 | 0.227 | 0.154 | -0.489 | -0.004 | 6.25E-01 | 18599 | -1.249 | -4.655 | 2.157 | 1.738 | -0.719 | -0.005 | 4.72E-01 | 18827 |
| 74 | Paracentral_Lobule_R | 0.010 | -0.206 | 0.226 | 0.110 | 0.091 | 0.001 | 9.28E-01 | 18599 | -0.151 | -2.592 | 2.289 | 1.245 | -0.122 | -0.001 | 9.03E-01 | 18827 |
| 75 | Caudate_L | 0.369 | 0.128 | 0.610 | 0.123 | 3.002 | 0.022 | 2.68E-03 | 18599 | -4.013 | -6.733 | -1.293 | 1.388 | -2.892 | -0.021 | 3.84E-03 | 18827 |
| 76 | Caudate_R | 0.477 | 0.204 | 0.750 | 0.139 | 3.425 | 0.025 | 6.15E-04 | 18599 | -4.904 | -7.986 | -1.822 | 1.572 | -3.119 | -0.023 | 1.82E-03 | 18827 |
| 77 | Putamen_L | 0.439 | 0.073 | 0.805 | 0.187 | 2.353 | 0.017 | 1.86E-02 | 18599 | -5.546 | -9.675 | -1.416 | 2.107 | -2.632 | -0.019 | 8.49E-03 | 18827 |
| 78 | Putamen_R | 0.391 | 0.019 | 0.764 | 0.190 | 2.058 | 0.015 | 3.96E-02 | 18599 | -6.454 | -10.659 | -2.248 | 2.146 | -3.008 | -0.022 | 2.63E-03 | 18827 |
| 79 | Pallidum_L | 0.020 | -0.012 | 0.053 | 0.017 | 1.211 | 0.009 | 2.26E-01 | 18599 | -0.427 | -0.794 | -0.060 | 0.187 | -2.280 | -0.017 | 2.26E-02 | 18827 |
| 80 | Pallidum_R | 0.029 | -0.007 | 0.065 | 0.018 | 1.602 | 0.012 | 1.09E-01 | 18599 | -0.667 | -1.070 | -0.264 | 0.206 | -3.246 | -0.024 | 1.17E-03 | 18827 |
| 83 | Heschl_L | 0.031 | -0.035 | 0.097 | 0.034 | 0.929 | 0.007 | 3.53E-01 | 18599 | -1.264 | -2.008 | -0.520 | 0.380 | -3.329 | -0.024 | 8.75E-04 | 18827 |
| 84 | Heschl_R | 0.051 | -0.023 | 0.125 | 0.038 | 1.346 | 0.010 | 1.78E-01 | 18599 | -1.907 | -2.738 | -1.075 | 0.424 | -4.494 | -0.033 | 7.03E-06 | 18827 |
| 85 | Temporal_Sup_L | 0.778 | 0.301 | 1.255 | 0.243 | 3.199 | 0.023 | 1.38E-03 | 18599 | -11.692 | -17.063 | -6.322 | 2.740 | -4.268 | -0.031 | 1.99E-05 | 18827 |
| 86 | Temporal_Sup_R | 0.340 | -0.187 | 0.867 | 0.269 | 1.264 | 0.009 | 2.06E-01 | 18599 | -14.763 | -20.709 | -8.818 | 3.033 | -4.867 | -0.035 | 1.14E-06 | 18827 |
| 87 | Temporal_Pole_Sup_L | 0.317 | 0.092 | 0.542 | 0.115 | 2.757 | 0.020 | 5.84E-03 | 18599 | -4.277 | -6.822 | -1.732 | 1.298 | -3.294 | -0.024 | 9.88E-04 | 18827 |
| 88 | Temporal_Pole_Sup_R | 0.446 | 0.208 | 0.683 | 0.121 | 3.682 | 0.027 | 2.32E-04 | 18599 | -4.708 | -7.385 | -2.030 | 1.366 | -3.446 | -0.025 | 5.70E-04 | 18827 |
| 89 | Temporal_Mid_L | -0.067 | -0.861 | 0.728 | 0.405 | -0.164 | -0.001 | 8.69E-01 | 18599 | -21.080 | -30.035 | -12.126 | 4.568 | -4.614 | -0.034 | 3.97E-06 | 18827 |
| 90 | Temporal_Mid_R | 0.176 | -0.530 | 0.882 | 0.360 | 0.488 | 0.004 | 6.26E-01 | 18599 | -13.089 | -21.039 | -5.139 | 4.056 | -3.227 | -0.024 | 1.25E-03 | 18827 |
| 91 | Temporal_Pole_Mid_L | 0.051 | -0.138 | 0.241 | 0.097 | 0.532 | 0.004 | 5.95E-01 | 18599 | -2.698 | -4.838 | -0.558 | 1.092 | -2.472 | -0.018 | 1.35E-02 | 18827 |
| 92 | Temporal_Pole_Mid_R | 0.314 | 0.084 | 0.545 | 0.117 | 2.677 | 0.020 | 7.43E-03 | 18599 | -3.404 | -6.001 | -0.806 | 1.325 | -2.568 | -0.019 | 1.02E-02 | 18827 |
| 93 | Temporal_Inf_L | -0.288 | -0.834 | 0.258 | 0.279 | -1.034 | -0.008 | 3.01E-01 | 18599 | -7.058 | -13.215 | -0.902 | 3.141 | -2.247 | -0.016 | 2.46E-02 | 18827 |
| 94 | Temporal_Inf_R | 0.177 | -0.419 | 0.773 | 0.304 | 0.582 | 0.004 | 5.61E-01 | 18599 | -4.664 | -11.379 | 2.052 | 3.426 | -1.361 | -0.010 | 1.73E-01 | 18827 |
| 95 | Cerebellum_Crus1_L | -0.621 | -1.270 | 0.028 | 0.331 | -1.875 | -0.014 | 6.08E-02 | 18599 | 0.968 | -6.348 | 8.284 | 3.733 | 0.259 | 0.002 | 7.95E-01 | 18827 |
| 96 | Cerebellum_Crus1_R | -0.338 | -1.000 | 0.323 | 0.337 | -1.002 | -0.007 | 3.17E-01 | 18599 | -0.063 | -7.522 | 7.395 | 3.805 | -0.017 | 0.000 | 9.87E-01 | 18827 |
| 97 | Cerebellum_Crus2_L | -0.292 | -0.807 | 0.222 | 0.262 | -1.115 | -0.008 | 2.65E-01 | 18599 | -6.323 | -12.122 | -0.524 | 2.959 | -2.137 | -0.016 | 3.26E-02 | 18827 |
| 98 | Cerebellum_Crus2_R | -0.165 | -0.640 | 0.310 | 0.242 | -0.680 | -0.005 | 4.96E-01 | 18599 | -3.073 | -8.435 | 2.288 | 2.735 | -1.124 | -0.008 | 2.61E-01 | 18827 |
| 99 | Cerebellum_3_L | 0.001 | -0.034 | 0.035 | 0.018 | 0.045 | 0.000 | 9.64E-01 | 18599 | 0.290 | -0.099 | 0.679 | 0.199 | 1.460 | 0.011 | 1.44E-01 | 18827 |
| 100 | Cerebellum_3_R | -0.014 | -0.060 | 0.033 | 0.024 | -0.571 | -0.004 | 5.68E-01 | 18599 | -0.001 | -0.526 | 0.525 | 0.268 | -0.003 | 0.000 | 9.98E-01 | 18827 |
| 101 | Cerebellum_4_5_L | -0.003 | -0.282 | 0.276 | 0.142 | -0.019 | 0.000 | 9.85E-01 | 18599 | 2.793 | -0.357 | 5.942 | 1.607 | 1.738 | 0.013 | 8.22E-02 | 18827 |
| 102 | Cerebellum_4_5_R | 0.058 | -0.169 | 0.285 | 0.116 | 0.500 | 0.004 | 6.17E-01 | 18599 | 0.685 | -1.881 | 3.250 | 1.309 | 0.523 | 0.004 | 6.01E-01 | 18827 |
| 103 | Cerebellum_6_L | -0.440 | -0.923 | 0.043 | 0.246 | -1.785 | -0.013 | 7.42E-02 | 18599 | 0.043 | -5.404 | 5.490 | 2.779 | 0.015 | 0.000 | 9.88E-01 | 18827 |
| 104 | Cerebellum_6_R | -0.344 | -0.853 | 0.164 | 0.259 | -1.328 | -0.010 | 1.84E-01 | 18599 | 1.621 | -4.116 | 7.358 | 2.927 | 0.554 | 0.004 | 5.80E-01 | 18827 |
| 105 | Cerebellum_7b_L | -0.049 | -0.207 | 0.108 | 0.080 | -0.617 | -0.005 | 5.37E-01 | 18599 | -1.184 | -2.957 | 0.590 | 0.905 | -1.308 | -0.010 | 1.91E-01 | 18827 |
| 106 | Cerebellum_7b_R | -0.046 | -0.202 | 0.109 | 0.079 | -0.580 | -0.004 | 5.62E-01 | 18599 | -1.722 | -3.479 | 0.034 | 0.896 | -1.922 | -0.014 | 5.47E-02 | 18827 |
| 107 | Cerebellum_8_L | -0.144 | -0.643 | 0.355 | 0.255 | -0.565 | -0.004 | 5.72E-01 | 18599 | 1.425 | -4.210 | 7.060 | 2.875 | 0.496 | 0.004 | 6.20E-01 | 18827 |
| 108 | Cerebellum_8_R | 0.035 | -0.615 | 0.684 | 0.331 | 0.105 | 0.001 | 9.16E-01 | 18599 | -2.333 | -9.666 | 5.000 | 3.741 | -0.624 | -0.005 | 5.33E-01 | 18827 |
| 109 | Cerebellum_9_L | -0.116 | -0.398 | 0.166 | 0.144 | -0.807 | -0.006 | 4.20E-01 | 18599 | 0.988 | -2.196 | 4.173 | 1.625 | 0.608 | 0.004 | 5.43E-01 | 18827 |
| 110 | Cerebellum_9_R | -0.090 | -0.367 | 0.187 | 0.141 | -0.638 | -0.005 | 5.24E-01 | 18599 | 1.824 | -1.307 | 4.954 | 1.597 | 1.142 | 0.008 | 2.54E-01 | 18827 |
| 111 | Cerebellum_10_L | 0.009 | -0.011 | 0.029 | 0.010 | 0.899 | 0.007 | 3.68E-01 | 18599 | 0.178 | -0.048 | 0.404 | 0.115 | 1.545 | 0.011 | 1.22E-01 | 18827 |
| 112 | Cerebellum_10_R | 0.017 | -0.004 | 0.037 | 0.011 | 1.558 | 0.011 | 1.19E-01 | 18599 | 0.162 | -0.073 | 0.396 | 0.120 | 1.349 | 0.010 | 1.77E-01 | 18827 |
| 113 | Vermis_1_2 | 0.002 | -0.011 | 0.015 | 0.007 | 0.358 | 0.003 | 7.21E-01 | 18599 | -0.020 | -0.167 | 0.126 | 0.075 | -0.270 | -0.002 | 7.87E-01 | 18827 |

| | | | | | | | | | | | | | | | | | |
|---|---|---|---|---|---|---|---|---|---|---|---|---|---|---|---|---|---|
| 114 | Vermis_3 | 0.020 | -0.032 | 0.072 | 0.026 | 0.758 | 0.006 | 4.49E-01 | 18599 | -0.100 | -0.684 | 0.483 | 0.298 | -0.336 | -0.002 | 7.37E-01 | 18827 |
| 115 | Vermis_4_5 | 0.090 | -0.062 | 0.242 | 0.078 | 1.164 | 0.009 | 2.44E-01 | 18599 | 1.682 | -0.032 | 3.397 | 0.875 | 1.923 | 0.014 | 5.45E-02 | 18827 |
| 116 | Vermis_6 | -0.035 | -0.119 | 0.050 | 0.043 | -0.805 | -0.006 | 4.21E-01 | 18599 | 0.709 | -0.246 | 1.664 | 0.487 | 1.455 | 0.011 | 1.46E-01 | 18827 |
| 117 | Vermis_7 | -0.003 | -0.051 | 0.045 | 0.024 | -0.125 | -0.001 | 9.01E-01 | 18599 | 0.303 | -0.237 | 0.843 | 0.275 | 1.101 | 0.008 | 2.71E-01 | 18827 |
| 118 | Vermis_8 | -0.040 | -0.119 | 0.039 | 0.040 | -0.999 | -0.007 | 3.18E-01 | 18599 | 0.301 | -0.586 | 1.189 | 0.453 | 0.665 | 0.005 | 5.06E-01 | 18827 |
| 119 | Vermis_9 | -0.030 | -0.094 | 0.034 | 0.033 | -0.928 | -0.007 | 3.53E-01 | 18599 | 0.327 | -0.393 | 1.047 | 0.367 | 0.890 | 0.006 | 3.73E-01 | 18827 |
| 120 | Vermis_10 | -0.001 | -0.019 | 0.016 | 0.009 | -0.151 | -0.001 | 8.80E-01 | 18599 | 0.149 | -0.052 | 0.349 | 0.102 | 1.454 | 0.011 | 1.46E-01 | 18827 |
| 121 | Thal_AV_L | 0.010 | 0.003 | 0.017 | 0.004 | 2.709 | 0.020 | 6.76E-03 | 18599 | -0.100 | -0.180 | -0.019 | 0.041 | -2.432 | -0.018 | 1.50E-02 | 18827 |
| 122 | Thal_AV_R | 0.010 | 0.000 | 0.020 | 0.005 | 1.916 | 0.014 | 5.53E-02 | 18599 | -0.176 | -0.290 | -0.061 | 0.058 | -3.007 | -0.022 | 2.64E-03 | 18827 |
| 123 | Thal_LP_L | 0.012 | 0.004 | 0.020 | 0.004 | 3.073 | 0.023 | 2.12E-03 | 18599 | -0.163 | -0.252 | -0.074 | 0.045 | -3.585 | -0.026 | 3.38E-04 | 18827 |
| 124 | Thal_LP_R | 0.008 | -0.002 | 0.018 | 0.005 | 1.503 | 0.011 | 1.33E-01 | 18599 | -0.253 | -0.369 | -0.137 | 0.059 | -4.277 | -0.031 | 1.90E-05 | 18827 |
| 125 | Thal_VA_L | 0.005 | -0.004 | 0.013 | 0.004 | 1.092 | 0.008 | 2.75E-01 | 18599 | -0.051 | -0.146 | 0.044 | 0.048 | -1.059 | -0.008 | 2.90E-01 | 18827 |
| 126 | Thal_VA_R | 0.007 | -0.003 | 0.017 | 0.005 | 1.302 | 0.010 | 1.93E-01 | 18599 | -0.101 | -0.218 | 0.015 | 0.059 | -1.708 | -0.012 | 8.77E-02 | 18827 |
| 127 | Thal_VL_L | 0.055 | 0.031 | 0.079 | 0.012 | 4.506 | 0.033 | 6.64E-06 | 18599 | -0.392 | -0.663 | -0.122 | 0.138 | -2.847 | -0.021 | 4.41E-03 | 18827 |
| 128 | Thal_VL_R | 0.050 | 0.018 | 0.082 | 0.016 | 3.086 | 0.023 | 2.03E-03 | 18599 | -0.664 | -1.022 | -0.305 | 0.183 | -3.630 | -0.026 | 2.84E-04 | 18827 |
| 129 | Thal_VPL_L | 0.024 | -0.005 | 0.052 | 0.015 | 1.627 | 0.012 | 1.04E-01 | 18599 | -0.576 | -0.897 | -0.255 | 0.164 | -3.514 | -0.026 | 4.42E-04 | 18827 |
| 130 | Thal_VPL_R | 0.008 | -0.022 | 0.038 | 0.015 | 0.519 | 0.004 | 6.04E-01 | 18599 | -0.644 | -0.981 | -0.306 | 0.172 | -3.734 | -0.027 | 1.89E-04 | 18827 |
| 131 | Thal_IL_L | 0.017 | 0.004 | 0.029 | 0.006 | 2.665 | 0.020 | 7.71E-03 | 18599 | -0.387 | -0.526 | -0.248 | 0.071 | -5.455 | -0.040 | 4.95E-08 | 18827 |
| 132 | Thal_IL_R | 0.013 | -0.001 | 0.027 | 0.007 | 1.880 | 0.014 | 6.01E-02 | 18599 | -0.412 | -0.570 | -0.254 | 0.080 | -5.121 | -0.037 | 3.06E-07 | 18827 |
| 133 | Thal_Re_L | 0.000 | 0.000 | 0.000 | 0.000 | 0.609 | 0.004 | 5.42E-01 | 18599 | 0.000 | -0.001 | 0.000 | 0.000 | -0.984 | -0.007 | 3.25E-01 | 18827 |
| 134 | Thal_Re_R | 0.000 | 0.000 | 0.001 | 0.000 | 1.399 | 0.010 | 1.62E-01 | 18599 | -0.006 | -0.011 | -0.001 | 0.002 | -2.555 | -0.019 | 1.06E-02 | 18827 |
| 135 | Thal_MDm_L | 0.119 | 0.027 | 0.210 | 0.047 | 2.547 | 0.019 | 1.09E-02 | 18599 | -2.787 | -3.817 | -1.757 | 0.525 | -5.305 | -0.039 | 1.14E-07 | 18827 |
| 136 | Thal_MDm_R | 0.097 | -0.009 | 0.203 | 0.054 | 1.799 | 0.013 | 7.20E-02 | 18599 | -3.040 | -4.237 | -1.843 | 0.611 | -4.979 | -0.036 | 6.44E-07 | 18827 |
| 137 | Thal_MDl_L | 0.030 | 0.014 | 0.045 | 0.008 | 3.665 | 0.027 | 2.48E-04 | 18599 | -0.489 | -0.667 | -0.311 | 0.091 | -5.376 | -0.039 | 7.70E-08 | 18827 |
| 138 | Thal_MDl_R | 0.020 | 0.002 | 0.038 | 0.009 | 2.173 | 0.016 | 2.98E-02 | 18599 | -0.527 | -0.730 | -0.325 | 0.103 | -5.100 | -0.037 | 3.43E-07 | 18827 |
| 139 | Thal_LGN_L | 0.002 | 0.000 | 0.005 | 0.001 | 1.805 | 0.013 | 7.11E-02 | 18599 | -0.051 | -0.078 | -0.024 | 0.014 | -3.716 | -0.027 | 2.03E-04 | 18827 |
| 140 | Thal_LGN_R | 0.005 | 0.000 | 0.010 | 0.003 | 1.958 | 0.014 | 5.02E-02 | 18599 | -0.135 | -0.192 | -0.077 | 0.029 | -4.587 | -0.033 | 4.53E-06 | 18827 |
| 141 | Thal_MGN_L | 0.001 | -0.003 | 0.005 | 0.002 | 0.506 | 0.004 | 6.13E-01 | 18599 | -0.113 | -0.161 | -0.064 | 0.025 | -4.569 | -0.033 | 4.94E-06 | 18827 |
| 142 | Thal_MGN_R | 0.001 | -0.004 | 0.006 | 0.002 | 0.394 | 0.003 | 6.94E-01 | 18599 | -0.139 | -0.191 | -0.086 | 0.027 | -5.174 | -0.038 | 2.31E-07 | 18827 |
| 143 | Thal_PuA_L | 0.002 | -0.003 | 0.007 | 0.003 | 0.620 | 0.005 | 5.36E-01 | 18599 | -0.155 | -0.212 | -0.098 | 0.029 | -5.322 | -0.039 | 1.04E-07 | 18827 |
| 144 | Thal_PuA_R | 0.003 | -0.008 | 0.013 | 0.005 | 0.515 | 0.004 | 6.07E-01 | 18599 | -0.332 | -0.451 | -0.214 | 0.060 | -5.490 | -0.040 | 4.06E-08 | 18827 |
| 145 | Thal_PuM_L | 0.051 | -0.049 | 0.152 | 0.051 | 1.000 | 0.007 | 3.17E-01 | 18599 | -2.956 | -4.090 | -1.823 | 0.578 | -5.112 | -0.037 | 3.22E-07 | 18827 |
| 146 | Thal_PuM_R | 0.041 | -0.084 | 0.167 | 0.064 | 0.649 | 0.005 | 5.16E-01 | 18599 | -3.761 | -5.173 | -2.348 | 0.721 | -5.219 | -0.038 | 1.82E-07 | 18827 |
| 147 | Thal_PuL_L | 0.013 | -0.005 | 0.031 | 0.009 | 1.402 | 0.010 | 1.61E-01 | 18599 | -0.538 | -0.744 | -0.331 | 0.106 | -5.095 | -0.037 | 3.52E-07 | 18827 |
| 148 | Thal_PuL_R | 0.010 | -0.019 | 0.039 | 0.015 | 0.675 | 0.005 | 4.99E-01 | 18599 | -0.841 | -1.170 | -0.512 | 0.168 | -5.013 | -0.037 | 5.41E-07 | 18827 |
| 149 | Thal_PuI_L | 0.001 | -0.004 | 0.006 | 0.003 | 0.579 | 0.004 | 5.63E-01 | 18599 | -0.080 | -0.137 | -0.024 | 0.029 | -2.795 | -0.020 | 5.19E-03 | 18827 |
| 150 | Thal_PuI_R | -0.002 | -0.008 | 0.005 | 0.003 | -0.525 | -0.004 | 6.00E-01 | 18599 | -0.109 | -0.183 | -0.034 | 0.038 | -2.858 | -0.021 | 4.27E-03 | 18827 |
| 151 | ACC_sub_L | -0.006 | -0.064 | 0.052 | 0.029 | -0.203 | -0.001 | 8.39E-01 | 18599 | -0.955 | -1.604 | -0.306 | 0.331 | -2.883 | -0.021 | 3.94E-03 | 18827 |
| 152 | ACC_sub_R | -0.003 | -0.056 | 0.051 | 0.027 | -0.095 | -0.001 | 9.24E-01 | 18599 | -0.750 | -1.356 | -0.144 | 0.309 | -2.425 | -0.018 | 1.53E-02 | 18827 |
| 153 | ACC_pre_L | 0.069 | -0.079 | 0.216 | 0.075 | 0.914 | 0.007 | 3.61E-01 | 18599 | -2.727 | -4.391 | -1.064 | 0.849 | -3.214 | -0.023 | 1.31E-03 | 18827 |
| 154 | ACC_pre_R | 0.026 | -0.116 | 0.168 | 0.072 | 0.363 | 0.003 | 7.16E-01 | 18599 | -2.851 | -4.453 | -1.249 | 0.817 | -3.488 | -0.025 | 4.88E-04 | 18827 |
| 155 | ACC_sup_L | 0.171 | 0.011 | 0.331 | 0.082 | 2.088 | 0.015 | 3.68E-02 | 18599 | -2.757 | -4.564 | -0.949 | 0.922 | -2.989 | -0.022 | 2.80E-03 | 18827 |
| 156 | ACC_sup_R | 0.065 | -0.101 | 0.231 | 0.085 | 0.767 | 0.006 | 4.43E-01 | 18599 | -3.022 | -4.895 | -1.149 | 0.956 | -3.163 | -0.023 | 1.57E-03 | 18827 |
| 157 | N_Acc_L | 0.060 | 0.012 | 0.108 | 0.025 | 2.430 | 0.018 | 1.51E-02 | 18599 | -0.797 | -1.342 | -0.253 | 0.278 | -2.869 | -0.021 | 4.12E-03 | 18827 |
| 158 | N_Acc_R | 0.052 | 0.010 | 0.094 | 0.021 | 2.437 | 0.018 | 1.48E-02 | 18599 | -0.863 | -1.334 | -0.392 | 0.240 | -3.592 | -0.026 | 3.28E-04 | 18827 |
| 159 | VTA_L | 0.001 | -0.001 | 0.003 | 0.001 | 1.138 | 0.008 | 2.55E-01 | 18599 | -0.001 | -0.020 | 0.018 | 0.010 | -0.084 | -0.001 | 9.33E-01 | 18827 |
| 160 | VTA_R | 0.001 | 0.000 | 0.003 | 0.001 | 1.331 | 0.010 | 1.83E-01 | 18599 | -0.006 | -0.023 | 0.011 | 0.009 | -0.709 | -0.005 | 4.78E-01 | 18827 |
| 161 | SN_pc_L | 0.001 | -0.001 | 0.004 | 0.001 | 1.072 | 0.008 | 2.84E-01 | 18599 | -0.028 | -0.059 | 0.002 | 0.016 | -1.817 | -0.013 | 6.92E-02 | 18827 |
| 162 | SN_pc_R | 0.004 | 0.000 | 0.007 | 0.002 | 2.108 | 0.015 | 3.51E-02 | 18599 | -0.053 | -0.093 | -0.013 | 0.021 | -2.580 | -0.019 | 9.89E-03 | 18827 |

| | | | | | | | | | | | | | | | | | |
|---|---|---|---|---|---|---|---|---|---|---|---|---|---|---|---|---|---|
| 163 | SN_pr_L | 0.003 | 0.000 | 0.007 | 0.002 | 1.830 | 0.013 | 6.72E-02 | 18599 | -0.038 | -0.080 | 0.004 | 0.021 | -1.791 | -0.013 | 7.33E-02 | 18827 |
| 164 | SN_pr_R | 0.006 | 0.002 | 0.010 | 0.002 | 2.676 | 0.020 | 7.45E-03 | 18599 | -0.051 | -0.098 | -0.004 | 0.024 | -2.117 | -0.015 | 3.43E-02 | 18827 |
| 165 | Red_N_L | 0.003 | -0.001 | 0.008 | 0.002 | 1.417 | 0.010 | 1.56E-01 | 18599 | -0.072 | -0.124 | -0.020 | 0.026 | -2.739 | -0.020 | 6.16E-03 | 18827 |
| 166 | Red_N_R | 0.005 | 0.000 | 0.010 | 0.003 | 1.861 | 0.014 | 6.27E-02 | 18599 | -0.050 | -0.106 | 0.006 | 0.029 | -1.759 | -0.013 | 7.86E-02 | 18827 |
| 167 | LC_L | 0.001 | -0.001 | 0.003 | 0.001 | 1.204 | 0.009 | 2.29E-01 | 18599 | -0.001 | -0.020 | 0.019 | 0.010 | -0.071 | -0.001 | 9.43E-01 | 18827 |
| 168 | LC_R | 0.001 | -0.001 | 0.003 | 0.001 | 1.205 | 0.009 | 2.28E-01 | 18599 | 0.000 | -0.021 | 0.021 | 0.011 | 0.007 | 0.000 | 9.95E-01 | 18827 |
| 169 | Raphe_D | 0.003 | -0.004 | 0.009 | 0.003 | 0.805 | 0.006 | 4.21E-01 | 18599 | -0.093 | -0.167 | -0.018 | 0.038 | -2.436 | -0.018 | 1.49E-02 | 18827 |
| 170 | Raphe_M | 0.002 | 0.000 | 0.004 | 0.001 | 1.682 | 0.012 | 9.25E-02 | 18599 | -0.004 | -0.026 | 0.019 | 0.012 | -0.321 | -0.002 | 7.48E-01 | 18827 |

| | | Oily fish intake | | | | | | | | Cheese intake | | | | | | | |
|---|---|---|---|---|---|---|---|---|---|---|---|---|---|---|---|---|---|
| **Number** | **AAL 3 region** | **beta** | **95%CI_l** | **95%CI_h** | **se** | **t** | **r** | **p** | **df.e** | **beta** | **95%CI_l** | **95%CI_h** | **se** | **t** | **r** | **p** | **df.e** |
| 0 | Total brain | -155.729 | -305.500 | -5.958 | 76.410 | -2.038 | -0.015 | 4.16E-02 | 18842 | 0.163 | -126.741 | 127.068 | 64.744 | 0.003 | 0.000 | 9.98E-01 | 18590 |
| 1 | Precentral_L | -2.718 | -7.982 | 2.547 | 2.686 | -1.012 | -0.007 | 3.12E-01 | 18842 | -2.900 | -7.360 | 1.559 | 2.275 | -1.275 | -0.009 | 2.02E-01 | 18590 |
| 2 | Precentral_R | -2.806 | -7.632 | 2.020 | 2.462 | -1.140 | -0.008 | 2.54E-01 | 18842 | -1.818 | -5.902 | 2.266 | 2.083 | -0.873 | -0.006 | 3.83E-01 | 18590 |
| 3 | Frontal_Sup_2_L | -2.305 | -7.868 | 3.257 | 2.838 | -0.812 | -0.006 | 4.17E-01 | 18842 | 2.868 | -1.839 | 7.576 | 2.401 | 1.194 | 0.009 | 2.32E-01 | 18590 |
| 4 | Frontal_Sup_2_R | -4.411 | -10.541 | 1.718 | 3.127 | -1.411 | -0.010 | 1.58E-01 | 18842 | 1.344 | -3.847 | 6.536 | 2.649 | 0.508 | 0.004 | 6.12E-01 | 18590 |
| 5 | Frontal_Mid_2_L | -3.993 | -9.833 | 1.846 | 2.979 | -1.340 | -0.010 | 1.80E-01 | 18842 | 3.172 | -1.778 | 8.122 | 2.526 | 1.256 | 0.009 | 2.09E-01 | 18590 |
| 6 | Frontal_Mid_2_R | -7.229 | -13.839 | -0.618 | 3.372 | -2.143 | -0.016 | 3.21E-02 | 18842 | 1.171 | -4.424 | 6.766 | 2.855 | 0.410 | 0.003 | 6.82E-01 | 18590 |
| 7 | Frontal_Inf_Oper_L | -1.849 | -3.574 | -0.124 | 0.880 | -2.101 | -0.015 | 3.57E-02 | 18842 | 1.616 | 0.155 | 3.077 | 0.746 | 2.168 | 0.016 | 3.02E-02 | 18590 |
| 8 | Frontal_Inf_Oper_R | -1.727 | -3.891 | 0.437 | 1.104 | -1.565 | -0.011 | 1.18E-01 | 18842 | 1.875 | 0.042 | 3.708 | 0.935 | 2.005 | 0.015 | 4.50E-02 | 18590 |
| 9 | Frontal_Inf_Tri_L | -2.995 | -6.657 | 0.668 | 1.869 | -1.603 | -0.012 | 1.09E-01 | 18842 | 5.918 | 2.817 | 9.020 | 1.582 | 3.740 | 0.027 | 1.84E-04 | 18590 |
| 10 | Frontal_Inf_Tri_R | -0.071 | -2.949 | 2.807 | 1.468 | -0.048 | 0.000 | 9.62E-01 | 18842 | 4.435 | 1.995 | 6.874 | 1.245 | 3.563 | 0.026 | 3.67E-04 | 18590 |
| 11 | Frontal_Inf_Orb_2_L | -0.698 | -1.793 | 0.398 | 0.559 | -1.248 | -0.009 | 2.12E-01 | 18842 | 2.281 | 1.353 | 3.210 | 0.474 | 4.815 | 0.035 | 1.48E-06 | 18590 |
| 12 | Frontal_Inf_Orb_2_R | -0.452 | -1.597 | 0.694 | 0.584 | -0.773 | -0.006 | 4.40E-01 | 18842 | 2.837 | 1.866 | 3.807 | 0.495 | 5.731 | 0.042 | 1.02E-08 | 18590 |
| 13 | Rolandic_Oper_L | -1.048 | -2.705 | 0.609 | 0.845 | -1.240 | -0.009 | 2.15E-01 | 18842 | 1.322 | -0.083 | 2.726 | 0.717 | 1.844 | 0.014 | 6.52E-02 | 18590 |
| 14 | Rolandic_Oper_R | 0.166 | -2.204 | 2.537 | 1.209 | 0.137 | 0.001 | 8.91E-01 | 18842 | 0.050 | -1.960 | 2.060 | 1.025 | 0.049 | 0.000 | 9.61E-01 | 18590 |
| 15 | Supp_Motor_Area_L | -1.420 | -5.361 | 2.521 | 2.011 | -0.706 | -0.005 | 4.80E-01 | 18842 | -4.014 | -7.347 | -0.682 | 1.700 | -2.361 | -0.017 | 1.82E-02 | 18590 |
| 16 | Supp_Motor_Area_R | 0.168 | -4.186 | 4.522 | 2.221 | 0.076 | 0.001 | 9.40E-01 | 18842 | -2.903 | -6.585 | 0.779 | 1.879 | -1.545 | -0.011 | 1.22E-01 | 18590 |
| 17 | Olfactory_L | -0.039 | -0.624 | 0.547 | 0.299 | -0.130 | -0.001 | 8.97E-01 | 18842 | 0.392 | -0.105 | 0.888 | 0.253 | 1.545 | 0.011 | 1.22E-01 | 18590 |
| 18 | Olfactory_R | -0.127 | -0.714 | 0.460 | 0.300 | -0.424 | -0.003 | 6.71E-01 | 18842 | 0.469 | -0.028 | 0.967 | 0.254 | 1.849 | 0.014 | 6.45E-02 | 18590 |
| 19 | Frontal_Sup_Medial_L | -2.253 | -5.947 | 1.442 | 1.885 | -1.195 | -0.009 | 2.32E-01 | 18842 | 3.529 | 0.400 | 6.657 | 1.596 | 2.211 | 0.016 | 2.71E-02 | 18590 |
| 20 | Frontal_Sup_Medial_R | -1.629 | -4.317 | 1.059 | 1.371 | -1.188 | -0.009 | 2.35E-01 | 18842 | 0.625 | -1.652 | 2.902 | 1.162 | 0.538 | 0.004 | 5.91E-01 | 18590 |
| 21 | Frontal_Med_Orb_L | -1.003 | -2.383 | 0.377 | 0.704 | -1.425 | -0.010 | 1.54E-01 | 18842 | 1.207 | 0.038 | 2.376 | 0.596 | 2.024 | 0.015 | 4.30E-02 | 18590 |
| 22 | Frontal_Med_Orb_R | -0.561 | -2.257 | 1.135 | 0.865 | -0.649 | -0.005 | 5.17E-01 | 18842 | 1.644 | 0.208 | 3.081 | 0.733 | 2.243 | 0.016 | 2.49E-02 | 18590 |
| 23 | Rectus_L | -1.195 | -2.624 | 0.233 | 0.729 | -1.641 | -0.012 | 1.01E-01 | 18842 | 1.404 | 0.193 | 2.614 | 0.618 | 2.273 | 0.017 | 2.31E-02 | 18590 |
| 24 | Rectus_R | -0.371 | -1.692 | 0.951 | 0.674 | -0.550 | -0.004 | 5.83E-01 | 18842 | 1.461 | 0.342 | 2.581 | 0.571 | 2.558 | 0.019 | 1.05E-02 | 18590 |
| 25 | OFCmed_L | -0.693 | -1.521 | 0.135 | 0.422 | -1.640 | -0.012 | 1.01E-01 | 18842 | 0.138 | -0.564 | 0.840 | 0.358 | 0.384 | 0.003 | 7.01E-01 | 18590 |
| 26 | OFCmed_R | 0.006 | -0.959 | 0.970 | 0.492 | 0.011 | 0.000 | 9.91E-01 | 18842 | 0.337 | -0.481 | 1.155 | 0.417 | 0.807 | 0.006 | 4.20E-01 | 18590 |
| 27 | OFCant_L | -0.826 | -1.783 | 0.131 | 0.488 | -1.693 | -0.012 | 9.05E-02 | 18842 | -0.256 | -1.067 | 0.556 | 0.414 | -0.618 | -0.005 | 5.37E-01 | 18590 |
| 28 | OFCant_R | -0.888 | -2.152 | 0.376 | 0.645 | -1.377 | -0.010 | 1.69E-01 | 18842 | 0.764 | -0.308 | 1.836 | 0.547 | 1.397 | 0.010 | 1.62E-01 | 18590 |
| 29 | OFCpost_L | -0.743 | -1.940 | 0.455 | 0.611 | -1.215 | -0.009 | 2.24E-01 | 18842 | 0.094 | -0.922 | 1.110 | 0.518 | 0.181 | 0.001 | 8.57E-01 | 18590 |
| 30 | OFCpost_R | -0.941 | -2.048 | 0.166 | 0.565 | -1.666 | -0.012 | 9.58E-02 | 18842 | 0.416 | -0.522 | 1.354 | 0.479 | 0.869 | 0.006 | 3.85E-01 | 18590 |
| 31 | OFClat_L | -0.100 | -0.504 | 0.304 | 0.206 | -0.484 | -0.004 | 6.28E-01 | 18842 | 0.811 | 0.468 | 1.154 | 0.175 | 4.639 | 0.034 | 3.52E-06 | 18590 |
| 32 | OFClat_R | -0.177 | -0.518 | 0.164 | 0.174 | -1.016 | -0.007 | 3.10E-01 | 18842 | 0.735 | 0.446 | 1.024 | 0.148 | 4.980 | 0.037 | 6.40E-07 | 18590 |
| 33 | Insula_L | -5.642 | -9.346 | -1.938 | 1.890 | -2.986 | -0.022 | 2.83E-03 | 18842 | 3.166 | 0.027 | 6.305 | 1.602 | 1.977 | 0.014 | 4.81E-02 | 18590 |
| 34 | Insula_R | -4.540 | -8.150 | -0.930 | 1.842 | -2.465 | -0.018 | 1.37E-02 | 18842 | 3.709 | 0.648 | 6.769 | 1.561 | 2.375 | 0.017 | 1.76E-02 | 18590 |
| 37 | Cingulate_Mid_L | -2.491 | -5.628 | 0.646 | 1.600 | -1.557 | -0.011 | 1.20E-01 | 18842 | -1.022 | -3.679 | 1.635 | 1.355 | -0.754 | -0.006 | 4.51E-01 | 18590 |
| 38 | Cingulate_Mid_R | -3.145 | -6.488 | 0.198 | 1.706 | -1.844 | -0.013 | 6.52E-02 | 18842 | -1.884 | -4.715 | 0.947 | 1.444 | -1.304 | -0.010 | 1.92E-01 | 18590 |
| 39 | Cingulate_Post_L | -1.079 | -2.036 | -0.122 | 0.488 | -2.211 | -0.016 | 2.71E-02 | 18842 | -0.705 | -1.515 | 0.105 | 0.413 | -1.705 | -0.013 | 8.81E-02 | 18590 |
| 40 | Cingulate_Post_R | -0.606 | -1.137 | -0.075 | 0.271 | -2.238 | -0.016 | 2.52E-02 | 18842 | -0.571 | -1.020 | -0.121 | 0.229 | -2.487 | -0.018 | 1.29E-02 | 18590 |

| | | | | | | | | | | | | | | | | | |
|---|---|---|---|---|---|---|---|---|---|---|---|---|---|---|---|---|---|
| 41 | Hippocampus_L | 0.465 | -1.082 | 2.012 | 0.789 | 0.589 | 0.004 | 5.56E-01 | 18842 | 0.686 | -0.626 | 1.999 | 0.670 | 1.025 | 0.008 | 3.05E-01 | 18590 |
| 42 | Hippocampus_R | 1.501 | -0.104 | 3.106 | 0.819 | 1.833 | 0.013 | 6.68E-02 | 18842 | 0.954 | -0.408 | 2.315 | 0.695 | 1.373 | 0.010 | 1.70E-01 | 18590 |
| 43 | ParaHippocampal_L | -0.063 | -1.438 | 1.311 | 0.701 | -0.090 | -0.001 | 9.28E-01 | 18842 | 0.604 | -0.561 | 1.769 | 0.594 | 1.016 | 0.007 | 3.09E-01 | 18590 |
| 44 | ParaHippocampal_R | 0.254 | -1.482 | 1.990 | 0.886 | 0.287 | 0.002 | 7.74E-01 | 18842 | 0.934 | -0.536 | 2.405 | 0.750 | 1.246 | 0.009 | 2.13E-01 | 18590 |
| 45 | Amygdala_L | -0.698 | -1.232 | -0.163 | 0.272 | -2.560 | -0.019 | 1.05E-02 | 18842 | 0.402 | -0.050 | 0.855 | 0.231 | 1.743 | 0.013 | 8.14E-02 | 18590 |
| 46 | Amygdala_R | -0.727 | -1.274 | -0.181 | 0.279 | -2.608 | -0.019 | 9.12E-03 | 18842 | 0.224 | -0.239 | 0.688 | 0.236 | 0.949 | 0.007 | 3.43E-01 | 18590 |
| 47 | Calcarine_L | -0.507 | -5.603 | 4.590 | 2.600 | -0.195 | -0.001 | 8.45E-01 | 18842 | -12.257 | -16.576 | -7.939 | 2.203 | -5.563 | -0.041 | 2.68E-08 | 18590 |
| 48 | Calcarine_R | -1.104 | -5.363 | 3.154 | 2.173 | -0.508 | -0.004 | 6.11E-01 | 18842 | -9.123 | -12.729 | -5.517 | 1.840 | -4.959 | -0.036 | 7.14E-07 | 18590 |
| 49 | Cuneus_L | 0.283 | -2.918 | 3.483 | 1.633 | 0.173 | 0.001 | 8.63E-01 | 18842 | -6.879 | -9.590 | -4.168 | 1.383 | -4.973 | -0.036 | 6.64E-07 | 18590 |
| 50 | Cuneus_R | -1.570 | -4.018 | 0.877 | 1.249 | -1.257 | -0.009 | 2.09E-01 | 18842 | -4.814 | -6.887 | -2.740 | 1.058 | -4.550 | -0.033 | 5.39E-06 | 18590 |
| 51 | Lingual_L | -1.124 | -4.192 | 1.943 | 1.565 | -0.719 | -0.005 | 4.72E-01 | 18842 | -3.514 | -6.115 | -0.912 | 1.327 | -2.648 | -0.019 | 8.12E-03 | 18590 |
| 52 | Lingual_R | 0.043 | -3.170 | 3.257 | 1.639 | 0.026 | 0.000 | 9.79E-01 | 18842 | -4.295 | -7.017 | -1.572 | 1.389 | -3.092 | -0.023 | 1.99E-03 | 18590 |
| 53 | Occipital_Sup_L | -0.210 | -2.319 | 1.899 | 1.076 | -0.195 | -0.001 | 8.45E-01 | 18842 | -2.185 | -3.972 | -0.398 | 0.912 | -2.397 | -0.018 | 1.65E-02 | 18590 |
| 54 | Occipital_Sup_R | -1.383 | -3.668 | 0.903 | 1.166 | -1.186 | -0.009 | 2.36E-01 | 18842 | -0.745 | -2.681 | 1.192 | 0.988 | -0.754 | -0.006 | 4.51E-01 | 18590 |
| 55 | Occipital_Mid_L | 0.211 | -4.730 | 5.152 | 2.521 | 0.084 | 0.001 | 9.33E-01 | 18842 | -4.166 | -8.348 | 0.016 | 2.134 | -1.952 | -0.014 | 5.09E-02 | 18590 |
| 56 | Occipital_Mid_R | -3.778 | -7.217 | -0.339 | 1.755 | -2.153 | -0.016 | 3.13E-02 | 18842 | -1.819 | -4.733 | 1.095 | 1.487 | -1.224 | -0.009 | 2.21E-01 | 18590 |
| 57 | Occipital_Inf_L | -0.820 | -2.459 | 0.819 | 0.836 | -0.981 | -0.007 | 3.27E-01 | 18842 | -2.768 | -4.156 | -1.380 | 0.708 | -3.909 | -0.029 | 9.29E-05 | 18590 |
| 58 | Occipital_Inf_R | -3.354 | -5.248 | -1.459 | 0.967 | -3.470 | -0.025 | 5.22E-04 | 18842 | -3.354 | -4.960 | -1.749 | 0.819 | -4.095 | -0.030 | 4.24E-05 | 18590 |
| 59 | Fusiform_L | -1.264 | -4.854 | 2.325 | 1.831 | -0.690 | -0.005 | 4.90E-01 | 18842 | -1.021 | -4.057 | 2.016 | 1.549 | -0.659 | -0.005 | 5.10E-01 | 18590 |
| 60 | Fusiform_R | -0.343 | -4.288 | 3.601 | 2.012 | -0.171 | -0.001 | 8.65E-01 | 18842 | 2.133 | -1.210 | 5.476 | 1.706 | 1.251 | 0.009 | 2.11E-01 | 18590 |
| 61 | Postcentral_L | 1.743 | -3.659 | 7.145 | 2.756 | 0.632 | 0.005 | 5.27E-01 | 18842 | 7.468 | 2.901 | 12.036 | 2.330 | 3.205 | 0.024 | 1.35E-03 | 18590 |
| 62 | Postcentral_R | 1.746 | -3.786 | 7.279 | 2.822 | 0.619 | 0.005 | 5.36E-01 | 18842 | 4.301 | -0.379 | 8.982 | 2.388 | 1.801 | 0.013 | 7.17E-02 | 18590 |
| 63 | Parietal_Sup_L | -2.228 | -5.340 | 0.885 | 1.588 | -1.403 | -0.010 | 1.61E-01 | 18842 | -3.039 | -5.675 | -0.402 | 1.345 | -2.259 | -0.017 | 2.39E-02 | 18590 |
| 64 | Parietal_Sup_R | -0.163 | -2.922 | 2.597 | 1.408 | -0.116 | -0.001 | 9.08E-01 | 18842 | -2.632 | -4.968 | -0.295 | 1.192 | -2.208 | -0.016 | 2.73E-02 | 18590 |
| 65 | Parietal_Inf_L | -3.987 | -8.061 | 0.086 | 2.078 | -1.919 | -0.014 | 5.51E-02 | 18842 | 0.224 | -3.225 | 3.674 | 1.760 | 0.127 | 0.001 | 8.99E-01 | 18590 |
| 66 | Parietal_Inf_R | -1.196 | -3.837 | 1.445 | 1.347 | -0.887 | -0.006 | 3.75E-01 | 18842 | -0.646 | -2.880 | 1.588 | 1.140 | -0.567 | -0.004 | 5.71E-01 | 18590 |
| 67 | SupraMarginal_L | -1.544 | -3.931 | 0.843 | 1.218 | -1.268 | -0.009 | 2.05E-01 | 18842 | 2.435 | 0.413 | 4.458 | 1.032 | 2.360 | 0.017 | 1.83E-02 | 18590 |
| 68 | SupraMarginal_R | -1.673 | -5.054 | 1.708 | 1.725 | -0.970 | -0.007 | 3.32E-01 | 18842 | 3.727 | 0.865 | 6.590 | 1.460 | 2.552 | 0.019 | 1.07E-02 | 18590 |
| 69 | Angular_L | -0.285 | -2.631 | 2.062 | 1.197 | -0.238 | -0.002 | 8.12E-01 | 18842 | -0.126 | -2.114 | 1.863 | 1.014 | -0.124 | -0.001 | 9.01E-01 | 18590 |
| 70 | Angular_R | -0.567 | -3.711 | 2.577 | 1.604 | -0.353 | -0.003 | 7.24E-01 | 18842 | -1.011 | -3.676 | 1.655 | 1.360 | -0.743 | -0.005 | 4.57E-01 | 18590 |
| 71 | Precuneus_L | -3.325 | -9.046 | 2.396 | 2.919 | -1.139 | -0.008 | 2.55E-01 | 18842 | -6.195 | -11.039 | -1.351 | 2.471 | -2.507 | -0.018 | 1.22E-02 | 18590 |
| 72 | Precuneus_R | -6.062 | -11.156 | -0.969 | 2.599 | -2.333 | -0.017 | 1.97E-02 | 18842 | -6.633 | -10.951 | -2.315 | 2.203 | -3.011 | -0.022 | 2.61E-03 | 18590 |
| 73 | Paracentral_Lobule_L | 0.154 | -2.464 | 2.772 | 1.336 | 0.115 | 0.001 | 9.08E-01 | 18842 | -3.062 | -5.278 | -0.847 | 1.130 | -2.710 | -0.020 | 6.74E-03 | 18590 |
| 74 | Paracentral_Lobule_R | -0.321 | -2.196 | 1.554 | 0.957 | -0.336 | -0.002 | 7.37E-01 | 18842 | -1.719 | -3.308 | -0.130 | 0.810 | -2.121 | -0.016 | 3.39E-02 | 18590 |
| 75 | Caudate_L | 0.426 | -1.664 | 2.516 | 1.066 | 0.399 | 0.003 | 6.90E-01 | 18842 | 0.190 | -1.577 | 1.957 | 0.902 | 0.211 | 0.002 | 8.33E-01 | 18590 |
| 76 | Caudate_R | 0.237 | -2.130 | 2.605 | 1.208 | 0.197 | 0.001 | 8.44E-01 | 18842 | 0.219 | -1.783 | 2.221 | 1.021 | 0.215 | 0.002 | 8.30E-01 | 18590 |
| 77 | Putamen_L | -4.997 | -8.171 | -1.823 | 1.619 | -3.086 | -0.022 | 2.03E-03 | 18842 | 0.743 | -1.945 | 3.431 | 1.372 | 0.542 | 0.004 | 5.88E-01 | 18590 |
| 78 | Putamen_R | -5.569 | -8.802 | -2.336 | 1.649 | -3.377 | -0.025 | 7.35E-04 | 18842 | 0.939 | -1.799 | 3.678 | 1.397 | 0.672 | 0.005 | 5.01E-01 | 18590 |
| 79 | Pallidum_L | -0.224 | -0.507 | 0.058 | 0.144 | -1.558 | -0.011 | 1.19E-01 | 18842 | 0.081 | -0.158 | 0.320 | 0.122 | 0.664 | 0.005 | 5.07E-01 | 18590 |
| 80 | Pallidum_R | -0.298 | -0.608 | 0.012 | 0.158 | -1.885 | -0.014 | 5.95E-02 | 18842 | 0.041 | -0.221 | 0.303 | 0.134 | 0.308 | 0.002 | 7.58E-01 | 18590 |
| 83 | Heschl_L | -0.437 | -1.010 | 0.135 | 0.292 | -1.498 | -0.011 | 1.34E-01 | 18842 | 0.119 | -0.366 | 0.604 | 0.247 | 0.480 | 0.004 | 6.31E-01 | 18590 |
| 84 | Heschl_R | -0.180 | -0.820 | 0.459 | 0.326 | -0.553 | -0.004 | 5.80E-01 | 18842 | 0.220 | -0.322 | 0.761 | 0.276 | 0.795 | 0.006 | 4.27E-01 | 18590 |
| 85 | Temporal_Sup_L | -4.186 | -8.315 | -0.057 | 2.107 | -1.987 | -0.014 | 4.69E-02 | 18842 | 4.056 | 0.556 | 7.556 | 1.786 | 2.272 | 0.017 | 2.31E-02 | 18590 |
| 86 | Temporal_Sup_R | -1.618 | -6.190 | 2.955 | 2.333 | -0.693 | -0.005 | 4.88E-01 | 18842 | 4.755 | 0.885 | 8.626 | 1.975 | 2.408 | 0.018 | 1.60E-02 | 18590 |
| 87 | Temporal_Pole_Sup_L | -1.812 | -3.768 | 0.145 | 0.998 | -1.815 | -0.013 | 6.95E-02 | 18842 | 0.763 | -0.896 | 2.421 | 0.846 | 0.901 | 0.007 | 3.67E-01 | 18590 |
| 88 | Temporal_Pole_Sup_R | -1.488 | -3.548 | 0.573 | 1.051 | -1.415 | -0.010 | 1.57E-01 | 18842 | 0.979 | -0.767 | 2.725 | 0.891 | 1.099 | 0.008 | 2.72E-01 | 18590 |
| 89 | Temporal_Mid_L | -2.328 | -9.216 | 4.560 | 3.514 | -0.662 | -0.005 | 5.08E-01 | 18842 | 2.642 | -3.188 | 8.473 | 2.975 | 0.888 | 0.007 | 3.74E-01 | 18590 |
| 90 | Temporal_Mid_R | -2.788 | -8.904 | 3.328 | 3.120 | -0.894 | -0.007 | 3.72E-01 | 18842 | 1.221 | -3.962 | 6.403 | 2.644 | 0.462 | 0.003 | 6.44E-01 | 18590 |
| 91 | Temporal_Pole_Mid_L | -0.335 | -1.980 | 1.310 | 0.839 | -0.399 | -0.003 | 6.90E-01 | 18842 | 0.084 | -1.310 | 1.478 | 0.711 | 0.118 | 0.001 | 9.06E-01 | 18590 |

| | | | | | | | | | | | | | | | | | |
|---|---|---|---|---|---|---|---|---|---|---|---|---|---|---|---|---|---|
| 92 | Temporal_Pole_Mid_R | 0.567 | -1.431 | 2.565 | 1.019 | 0.556 | 0.004 | 5.78E-01 | 18842 | 1.986 | 0.295 | 3.678 | 0.863 | 2.301 | 0.017 | 2.14E-02 | 18590 |
| 93 | Temporal_Inf_L | -1.988 | -6.721 | 2.744 | 2.415 | -0.824 | -0.006 | 4.10E-01 | 18842 | -2.776 | -6.786 | 1.234 | 2.046 | -1.357 | -0.010 | 1.75E-01 | 18590 |
| 94 | Temporal_Inf_R | -1.023 | -6.185 | 4.139 | 2.634 | -0.389 | -0.003 | 6.98E-01 | 18842 | -0.252 | -4.620 | 4.115 | 2.228 | -0.113 | -0.001 | 9.10E-01 | 18590 |
| 95 | Cerebellum_Crus1_L | 1.024 | -4.599 | 6.648 | 2.869 | 0.357 | 0.003 | 7.21E-01 | 18842 | -2.249 | -7.011 | 2.512 | 2.429 | -0.926 | -0.007 | 3.55E-01 | 18590 |
| 96 | Cerebellum_Crus1_R | 0.392 | -5.341 | 6.126 | 2.925 | 0.134 | 0.001 | 8.93E-01 | 18842 | 1.588 | -3.269 | 6.446 | 2.478 | 0.641 | 0.005 | 5.22E-01 | 18590 |
| 97 | Cerebellum_Crus2_L | -2.911 | -7.369 | 1.547 | 2.274 | -1.280 | -0.009 | 2.01E-01 | 18842 | 1.930 | -1.850 | 5.710 | 1.928 | 1.001 | 0.007 | 3.17E-01 | 18590 |
| 98 | Cerebellum_Crus2_R | 0.247 | -3.874 | 4.369 | 2.103 | 0.118 | 0.001 | 9.06E-01 | 18842 | 2.570 | -0.923 | 6.064 | 1.782 | 1.442 | 0.011 | 1.49E-01 | 18590 |
| 99 | Cerebellum_3_L | -0.213 | -0.512 | 0.086 | 0.153 | -1.397 | -0.010 | 1.62E-01 | 18842 | -0.414 | -0.667 | -0.161 | 0.129 | -3.203 | -0.023 | 1.36E-03 | 18590 |
| 100 | Cerebellum_3_R | -0.178 | -0.582 | 0.226 | 0.206 | -0.864 | -0.006 | 3.88E-01 | 18842 | -0.505 | -0.846 | -0.163 | 0.174 | -2.896 | -0.021 | 3.78E-03 | 18590 |
| 101 | Cerebellum_4_5_L | -1.036 | -3.456 | 1.384 | 1.235 | -0.839 | -0.006 | 4.01E-01 | 18842 | -1.449 | -3.499 | 0.600 | 1.046 | -1.386 | -0.010 | 1.66E-01 | 18590 |
| 102 | Cerebellum_4_5_R | -0.543 | -2.515 | 1.429 | 1.006 | -0.540 | -0.004 | 5.89E-01 | 18842 | -1.383 | -3.053 | 0.287 | 0.852 | -1.624 | -0.012 | 1.04E-01 | 18590 |
| 103 | Cerebellum_6_L | -0.612 | -4.799 | 3.575 | 2.136 | -0.286 | -0.002 | 7.75E-01 | 18842 | -2.529 | -6.074 | 1.016 | 1.809 | -1.398 | -0.010 | 1.62E-01 | 18590 |
| 104 | Cerebellum_6_R | -0.701 | -5.110 | 3.708 | 2.250 | -0.312 | -0.002 | 7.55E-01 | 18842 | -1.929 | -5.664 | 1.805 | 1.905 | -1.013 | -0.007 | 3.11E-01 | 18590 |
| 105 | Cerebellum_7b_L | -1.021 | -2.384 | 0.342 | 0.696 | -1.468 | -0.011 | 1.42E-01 | 18842 | 1.191 | 0.035 | 2.347 | 0.590 | 2.020 | 0.015 | 4.34E-02 | 18590 |
| 106 | Cerebellum_7b_R | -0.803 | -2.153 | 0.548 | 0.689 | -1.165 | -0.008 | 2.44E-01 | 18842 | 1.946 | 0.802 | 3.091 | 0.584 | 3.334 | 0.024 | 8.58E-04 | 18590 |
| 107 | Cerebellum_8_L | -4.045 | -8.376 | 0.285 | 2.209 | -1.831 | -0.013 | 6.71E-02 | 18842 | 0.361 | -3.310 | 4.033 | 1.873 | 0.193 | 0.001 | 8.47E-01 | 18590 |
| 108 | Cerebellum_8_R | -4.958 | -10.592 | 0.677 | 2.875 | -1.725 | -0.013 | 8.46E-02 | 18842 | 5.932 | 1.157 | 10.706 | 2.436 | 2.435 | 0.018 | 1.49E-02 | 18590 |
| 109 | Cerebellum_9_L | -3.321 | -5.769 | -0.874 | 1.249 | -2.660 | -0.019 | 7.82E-03 | 18842 | -0.695 | -2.769 | 1.379 | 1.058 | -0.657 | -0.005 | 5.11E-01 | 18590 |
| 110 | Cerebellum_9_R | -3.987 | -6.392 | -1.581 | 1.227 | -3.249 | -0.024 | 1.16E-03 | 18842 | -0.529 | -2.567 | 1.509 | 1.040 | -0.509 | -0.004 | 6.11E-01 | 18590 |
| 111 | Cerebellum_10_L | -0.100 | -0.273 | 0.074 | 0.089 | -1.129 | -0.008 | 2.59E-01 | 18842 | -0.160 | -0.307 | -0.013 | 0.075 | -2.129 | -0.016 | 3.33E-02 | 18590 |
| 112 | Cerebellum_10_R | 0.019 | -0.162 | 0.199 | 0.092 | 0.202 | 0.001 | 8.40E-01 | 18842 | 0.009 | -0.144 | 0.162 | 0.078 | 0.116 | 0.001 | 9.08E-01 | 18590 |
| 113 | Vermis_1_2 | -0.009 | -0.122 | 0.104 | 0.058 | -0.154 | -0.001 | 8.77E-01 | 18842 | -0.212 | -0.307 | -0.117 | 0.049 | -4.358 | -0.032 | 1.32E-05 | 18590 |
| 114 | Vermis_3 | -0.146 | -0.595 | 0.302 | 0.229 | -0.639 | -0.005 | 5.23E-01 | 18842 | -0.312 | -0.692 | 0.067 | 0.194 | -1.613 | -0.012 | 1.07E-01 | 18590 |
| 115 | Vermis_4_5 | -0.545 | -1.863 | 0.773 | 0.672 | -0.810 | -0.006 | 4.18E-01 | 18842 | -0.156 | -1.273 | 0.960 | 0.570 | -0.274 | -0.002 | 7.84E-01 | 18590 |
| 116 | Vermis_6 | -0.119 | -0.854 | 0.615 | 0.375 | -0.319 | -0.002 | 7.50E-01 | 18842 | -1.038 | -1.660 | -0.416 | 0.317 | -3.273 | -0.024 | 1.07E-03 | 18590 |
| 117 | Vermis_7 | -0.157 | -0.572 | 0.258 | 0.212 | -0.742 | -0.005 | 4.58E-01 | 18842 | -0.414 | -0.766 | -0.063 | 0.179 | -2.313 | -0.017 | 2.08E-02 | 18590 |
| 118 | Vermis_8 | -0.227 | -0.910 | 0.455 | 0.348 | -0.654 | -0.005 | 5.13E-01 | 18842 | -0.774 | -1.352 | -0.196 | 0.295 | -2.627 | -0.019 | 8.63E-03 | 18590 |
| 119 | Vermis_9 | -0.653 | -1.206 | -0.100 | 0.282 | -2.315 | -0.017 | 2.06E-02 | 18842 | -0.146 | -0.615 | 0.322 | 0.239 | -0.611 | -0.004 | 5.41E-01 | 18590 |
| 120 | Vermis_10 | -0.090 | -0.244 | 0.064 | 0.079 | -1.143 | -0.008 | 2.53E-01 | 18842 | -0.129 | -0.259 | 0.002 | 0.067 | -1.931 | -0.014 | 5.35E-02 | 18590 |
| 121 | Thal_AV_L | 0.046 | -0.016 | 0.107 | 0.031 | 1.451 | 0.011 | 1.47E-01 | 18842 | 0.098 | 0.046 | 0.150 | 0.027 | 3.711 | 0.027 | 2.07E-04 | 18590 |
| 122 | Thal_AV_R | 0.096 | 0.008 | 0.184 | 0.045 | 2.130 | 0.016 | 3.32E-02 | 18842 | 0.115 | 0.041 | 0.189 | 0.038 | 3.040 | 0.022 | 2.37E-03 | 18590 |
| 123 | Thal_LP_L | -0.009 | -0.078 | 0.059 | 0.035 | -0.271 | -0.002 | 7.86E-01 | 18842 | 0.099 | 0.042 | 0.157 | 0.029 | 3.372 | 0.025 | 7.49E-04 | 18590 |
| 124 | Thal_LP_R | 0.013 | -0.077 | 0.102 | 0.045 | 0.276 | 0.002 | 7.82E-01 | 18842 | 0.099 | 0.024 | 0.174 | 0.038 | 2.585 | 0.019 | 9.74E-03 | 18590 |
| 125 | Thal_VA_L | 0.093 | 0.020 | 0.166 | 0.037 | 2.491 | 0.018 | 1.27E-02 | 18842 | 0.060 | -0.001 | 0.122 | 0.031 | 1.913 | 0.014 | 5.58E-02 | 18590 |
| 126 | Thal_VA_R | 0.123 | 0.034 | 0.213 | 0.046 | 2.696 | 0.020 | 7.02E-03 | 18842 | 0.087 | 0.011 | 0.163 | 0.039 | 2.256 | 0.017 | 2.41E-02 | 18590 |
| 127 | Thal_VL_L | 0.014 | -0.193 | 0.222 | 0.106 | 0.135 | 0.001 | 8.93E-01 | 18842 | 0.367 | 0.192 | 0.542 | 0.089 | 4.109 | 0.030 | 3.99E-05 | 18590 |
| 128 | Thal_VL_R | 0.142 | -0.133 | 0.418 | 0.141 | 1.013 | 0.007 | 3.11E-01 | 18842 | 0.538 | 0.305 | 0.771 | 0.119 | 4.530 | 0.033 | 5.93E-06 | 18590 |
| 129 | Thal_VPL_L | -0.275 | -0.522 | -0.027 | 0.126 | -2.176 | -0.016 | 2.95E-02 | 18842 | 0.280 | 0.071 | 0.490 | 0.107 | 2.626 | 0.019 | 8.65E-03 | 18590 |
| 130 | Thal_VPL_R | -0.334 | -0.594 | -0.074 | 0.133 | -2.522 | -0.018 | 1.17E-02 | 18842 | 0.324 | 0.104 | 0.544 | 0.112 | 2.885 | 0.021 | 3.91E-03 | 18590 |
| 131 | Thal_IL_L | -0.107 | -0.214 | 0.000 | 0.055 | -1.968 | -0.014 | 4.90E-02 | 18842 | 0.100 | 0.009 | 0.191 | 0.046 | 2.164 | 0.016 | 3.04E-02 | 18590 |
| 132 | Thal_IL_R | -0.079 | -0.200 | 0.043 | 0.062 | -1.272 | -0.009 | 2.03E-01 | 18842 | 0.107 | 0.004 | 0.210 | 0.052 | 2.041 | 0.015 | 4.12E-02 | 18590 |
| 133 | Thal_Re_L | 0.000 | 0.000 | 0.001 | 0.000 | 1.826 | 0.013 | 6.79E-02 | 18842 | 0.000 | 0.000 | 0.001 | 0.000 | 2.144 | 0.016 | 3.21E-02 | 18590 |
| 134 | Thal_Re_R | 0.002 | -0.002 | 0.006 | 0.002 | 1.045 | 0.008 | 2.96E-01 | 18842 | 0.004 | 0.001 | 0.007 | 0.002 | 2.854 | 0.021 | 4.33E-03 | 18590 |
| 135 | Thal_MDm_L | -0.576 | -1.368 | 0.217 | 0.404 | -1.424 | -0.010 | 1.54E-01 | 18842 | 0.962 | 0.292 | 1.631 | 0.342 | 2.815 | 0.021 | 4.88E-03 | 18590 |
| 136 | Thal_MDm_R | -0.562 | -1.483 | 0.359 | 0.470 | -1.196 | -0.009 | 2.32E-01 | 18842 | 1.122 | 0.345 | 1.899 | 0.397 | 2.830 | 0.021 | 4.67E-03 | 18590 |
| 137 | Thal_MDl_L | -0.113 | -0.250 | 0.024 | 0.070 | -1.612 | -0.012 | 1.07E-01 | 18842 | 0.213 | 0.097 | 0.329 | 0.059 | 3.594 | 0.026 | 3.26E-04 | 18590 |
| 138 | Thal_MDl_R | -0.082 | -0.238 | 0.074 | 0.080 | -1.034 | -0.008 | 3.01E-01 | 18842 | 0.204 | 0.073 | 0.336 | 0.067 | 3.042 | 0.022 | 2.35E-03 | 18590 |
| 139 | Thal_LGN_L | -0.009 | -0.030 | 0.012 | 0.011 | -0.836 | -0.006 | 4.03E-01 | 18842 | 0.010 | -0.007 | 0.028 | 0.009 | 1.163 | 0.009 | 2.45E-01 | 18590 |
| 140 | Thal_LGN_R | -0.046 | -0.090 | -0.001 | 0.023 | -2.025 | -0.015 | 4.29E-02 | 18842 | 0.048 | 0.011 | 0.086 | 0.019 | 2.528 | 0.019 | 1.15E-02 | 18590 |

| | | | | | | | | | | | | | | | | | |
|---|---|---|---|---|---|---|---|---|---|---|---|---|---|---|---|---|---|
| 141 | Thal_MGN_L | -0.071 | -0.108 | -0.033 | 0.019 | -3.724 | -0.027 | 1.96E-04 | 18842 | -0.002 | -0.034 | 0.029 | 0.016 | -0.155 | -0.001 | 8.77E-01 | 18590 |
| 142 | Thal_MGN_R | -0.062 | -0.102 | -0.021 | 0.021 | -2.992 | -0.022 | 2.78E-03 | 18842 | 0.019 | -0.015 | 0.053 | 0.017 | 1.086 | 0.008 | 2.78E-01 | 18590 |
| 143 | Thal_PuA_L | -0.052 | -0.095 | -0.008 | 0.022 | -2.300 | -0.017 | 2.14E-02 | 18842 | 0.000 | -0.037 | 0.038 | 0.019 | 0.025 | 0.000 | 9.80E-01 | 18590 |
| 144 | Thal_PuA_R | -0.122 | -0.213 | -0.031 | 0.047 | -2.623 | -0.019 | 8.71E-03 | 18842 | 0.034 | -0.043 | 0.111 | 0.039 | 0.862 | 0.006 | 3.89E-01 | 18590 |
| 145 | Thal_PuM_L | -0.790 | -1.662 | 0.083 | 0.445 | -1.774 | -0.013 | 7.60E-02 | 18842 | 0.288 | -0.451 | 1.026 | 0.377 | 0.763 | 0.006 | 4.45E-01 | 18590 |
| 146 | Thal_PuM_R | -1.162 | -2.249 | -0.076 | 0.554 | -2.097 | -0.015 | 3.60E-02 | 18842 | 0.479 | -0.441 | 1.399 | 0.469 | 1.020 | 0.007 | 3.08E-01 | 18590 |
| 147 | Thal_PuL_L | -0.161 | -0.320 | -0.001 | 0.081 | -1.977 | -0.014 | 4.81E-02 | 18842 | 0.101 | -0.033 | 0.236 | 0.069 | 1.475 | 0.011 | 1.40E-01 | 18590 |
| 148 | Thal_PuL_R | -0.308 | -0.561 | -0.055 | 0.129 | -2.388 | -0.017 | 1.69E-02 | 18842 | 0.145 | -0.069 | 0.359 | 0.109 | 1.328 | 0.010 | 1.84E-01 | 18590 |
| 149 | Thal_PuI_L | -0.004 | -0.047 | 0.040 | 0.022 | -0.171 | -0.001 | 8.64E-01 | 18842 | 0.037 | 0.000 | 0.074 | 0.019 | 1.981 | 0.015 | 4.76E-02 | 18590 |
| 150 | Thal_PuI_R | -0.005 | -0.062 | 0.052 | 0.029 | -0.171 | -0.001 | 8.65E-01 | 18842 | 0.050 | 0.002 | 0.099 | 0.025 | 2.037 | 0.015 | 4.17E-02 | 18590 |
| 151 | ACC_sub_L | -0.174 | -0.673 | 0.325 | 0.255 | -0.685 | -0.005 | 4.94E-01 | 18842 | -0.090 | -0.514 | 0.333 | 0.216 | -0.418 | -0.003 | 6.76E-01 | 18590 |
| 152 | ACC_sub_R | -0.147 | -0.613 | 0.320 | 0.238 | -0.616 | -0.004 | 5.38E-01 | 18842 | 0.001 | -0.394 | 0.396 | 0.202 | 0.005 | 0.000 | 9.96E-01 | 18590 |
| 153 | ACC_pre_L | -0.878 | -2.158 | 0.402 | 0.653 | -1.344 | -0.010 | 1.79E-01 | 18842 | 0.462 | -0.623 | 1.547 | 0.554 | 0.835 | 0.006 | 4.04E-01 | 18590 |
| 154 | ACC_pre_R | -0.772 | -2.005 | 0.460 | 0.629 | -1.228 | -0.009 | 2.19E-01 | 18842 | 0.537 | -0.507 | 1.582 | 0.533 | 1.009 | 0.007 | 3.13E-01 | 18590 |
| 155 | ACC_sup_L | -1.141 | -2.532 | 0.250 | 0.710 | -1.608 | -0.012 | 1.08E-01 | 18842 | 0.089 | -1.090 | 1.268 | 0.601 | 0.148 | 0.001 | 8.82E-01 | 18590 |
| 156 | ACC_sup_R | -1.154 | -2.595 | 0.286 | 0.735 | -1.570 | -0.011 | 1.16E-01 | 18842 | -0.191 | -1.412 | 1.029 | 0.623 | -0.307 | -0.002 | 7.59E-01 | 18590 |
| 157 | N_Acc_L | 0.227 | -0.192 | 0.645 | 0.214 | 1.061 | 0.008 | 2.89E-01 | 18842 | 0.512 | 0.157 | 0.867 | 0.181 | 2.828 | 0.021 | 4.69E-03 | 18590 |
| 158 | N_Acc_R | -0.052 | -0.414 | 0.310 | 0.185 | -0.283 | -0.002 | 7.77E-01 | 18842 | 0.491 | 0.184 | 0.798 | 0.157 | 3.134 | 0.023 | 1.72E-03 | 18590 |
| 159 | VTA_L | 0.000 | -0.014 | 0.015 | 0.008 | 0.043 | 0.000 | 9.66E-01 | 18842 | 0.005 | -0.007 | 0.017 | 0.006 | 0.785 | 0.006 | 4.32E-01 | 18590 |
| 160 | VTA_R | -0.002 | -0.015 | 0.012 | 0.007 | -0.225 | -0.002 | 8.22E-01 | 18842 | 0.003 | -0.008 | 0.014 | 0.006 | 0.495 | 0.004 | 6.21E-01 | 18590 |
| 161 | SN_pc_L | -0.027 | -0.050 | -0.003 | 0.012 | -2.211 | -0.016 | 2.70E-02 | 18842 | -0.003 | -0.023 | 0.017 | 0.010 | -0.269 | -0.002 | 7.88E-01 | 18590 |
| 162 | SN_pc_R | -0.036 | -0.067 | -0.005 | 0.016 | -2.284 | -0.017 | 2.24E-02 | 18842 | -0.002 | -0.028 | 0.025 | 0.013 | -0.126 | -0.001 | 8.99E-01 | 18590 |
| 163 | SN_pr_L | -0.036 | -0.068 | -0.004 | 0.016 | -2.215 | -0.016 | 2.68E-02 | 18842 | 0.021 | -0.006 | 0.049 | 0.014 | 1.541 | 0.011 | 1.23E-01 | 18590 |
| 164 | SN_pr_R | -0.036 | -0.072 | 0.000 | 0.018 | -1.954 | -0.014 | 5.07E-02 | 18842 | 0.021 | -0.010 | 0.052 | 0.016 | 1.344 | 0.010 | 1.79E-01 | 18590 |
| 165 | Red_N_L | -0.024 | -0.064 | 0.015 | 0.020 | -1.198 | -0.009 | 2.31E-01 | 18842 | 0.012 | -0.022 | 0.045 | 0.017 | 0.699 | 0.005 | 4.85E-01 | 18590 |
| 166 | Red_N_R | -0.025 | -0.068 | 0.018 | 0.022 | -1.128 | -0.008 | 2.59E-01 | 18842 | 0.010 | -0.027 | 0.046 | 0.019 | 0.531 | 0.004 | 5.95E-01 | 18590 |
| 167 | LC_L | 0.011 | -0.004 | 0.026 | 0.008 | 1.425 | 0.010 | 1.54E-01 | 18842 | -0.018 | -0.030 | -0.005 | 0.006 | -2.813 | -0.021 | 4.91E-03 | 18590 |
| 168 | LC_R | 0.010 | -0.006 | 0.026 | 0.008 | 1.229 | 0.009 | 2.19E-01 | 18842 | -0.022 | -0.035 | -0.008 | 0.007 | -3.087 | -0.023 | 2.02E-03 | 18590 |
| 169 | Raphe_D | -0.026 | -0.083 | 0.031 | 0.029 | -0.890 | -0.006 | 3.74E-01 | 18842 | 0.026 | -0.023 | 0.075 | 0.025 | 1.052 | 0.008 | 2.93E-01 | 18590 |
| 170 | Raphe_M | 0.012 | -0.006 | 0.029 | 0.009 | 1.341 | 0.010 | 1.80E-01 | 18842 | -0.019 | -0.034 | -0.004 | 0.008 | -2.511 | -0.018 | 1.20E-02 | 18590 |

| Non-oily fish intake | | | | | | | | | |
|---|---|---|---|---|---|---|---|---|---|
| Number | AAL 3 region | beta | 95%CI_l | 95%CI_h | se | t | r | p | df.e |
| 0 | Total brain | -112.462 | -283.786 | 58.863 | 87.406 | -1.287 | -0.009 | 1.98E-01 | 18837 |
| 1 | Precentral_L | -5.509 | -11.532 | 0.515 | 3.073 | -1.793 | -0.013 | 7.31E-02 | 18837 |
| 2 | Precentral_R | 0.462 | -5.058 | 5.981 | 2.816 | 0.164 | 0.001 | 8.70E-01 | 18837 |
| 3 | Frontal_Sup_2_L | -5.124 | -11.484 | 1.235 | 3.245 | -1.579 | -0.012 | 1.14E-01 | 18837 |
| 4 | Frontal_Sup_2_R | -10.058 | -17.067 | -3.050 | 3.576 | -2.813 | -0.020 | 4.91E-03 | 18837 |
| 5 | Frontal_Mid_2_L | -5.484 | -12.165 | 1.196 | 3.408 | -1.609 | -0.012 | 1.08E-01 | 18837 |
| 6 | Frontal_Mid_2_R | -8.823 | -16.382 | -1.264 | 3.856 | -2.288 | -0.017 | 2.22E-02 | 18837 |
| 7 | Frontal_Inf_Oper_L | -2.278 | -4.251 | -0.305 | 1.006 | -2.263 | -0.016 | 2.36E-02 | 18837 |
| 8 | Frontal_Inf_Oper_R | -2.558 | -5.033 | -0.083 | 1.263 | -2.026 | -0.015 | 4.28E-02 | 18837 |
| 9 | Frontal_Inf_Tri_L | -1.971 | -6.161 | 2.219 | 2.138 | -0.922 | -0.007 | 3.56E-01 | 18837 |
| 10 | Frontal_Inf_Tri_R | -3.562 | -6.853 | -0.271 | 1.679 | -2.122 | -0.015 | 3.39E-02 | 18837 |
| 11 | Frontal_Inf_Orb_2_L | -0.804 | -2.057 | 0.449 | 0.639 | -1.258 | -0.009 | 2.09E-01 | 18837 |
| 12 | Frontal_Inf_Orb_2_R | -1.156 | -2.466 | 0.153 | 0.668 | -1.731 | -0.013 | 8.35E-02 | 18837 |
| 13 | Rolandic_Oper_L | -0.055 | -1.950 | 1.839 | 0.967 | -0.057 | 0.000 | 9.54E-01 | 18837 |
| 14 | Rolandic_Oper_R | -0.200 | -2.912 | 2.511 | 1.383 | -0.145 | -0.001 | 8.85E-01 | 18837 |
| 15 | Supp_Motor_Area_L | -2.597 | -7.104 | 1.910 | 2.299 | -1.129 | -0.008 | 2.59E-01 | 18837 |
| 16 | Supp_Motor_Area_R | -1.335 | -6.315 | 3.646 | 2.541 | -0.525 | -0.004 | 5.99E-01 | 18837 |

| | | | | | | | | | |
|---|---|---|---|---|---|---|---|---|---|
| 17 | Olfactory_L | 0.088 | -0.582 | 0.757 | 0.342 | 0.256 | 0.002 | 7.98E-01 | 18837 |
| 18 | Olfactory_R | 0.132 | -0.540 | 0.803 | 0.343 | 0.385 | 0.003 | 7.01E-01 | 18837 |
| 19 | Frontal_Sup_Medial_L | -4.852 | -9.076 | -0.627 | 2.155 | -2.251 | -0.016 | 2.44E-02 | 18837 |
| 20 | Frontal_Sup_Medial_R | -3.762 | -6.836 | -0.689 | 1.568 | -2.399 | -0.017 | 1.64E-02 | 18837 |
| 21 | Frontal_Med_Orb_L | -0.834 | -2.412 | 0.745 | 0.805 | -1.035 | -0.008 | 3.01E-01 | 18837 |
| 22 | Frontal_Med_Orb_R | -1.331 | -3.271 | 0.608 | 0.989 | -1.346 | -0.010 | 1.78E-01 | 18837 |
| 23 | Rectus_L | -0.873 | -2.507 | 0.761 | 0.833 | -1.048 | -0.008 | 2.95E-01 | 18837 |
| 24 | Rectus_R | -0.562 | -2.074 | 0.949 | 0.771 | -0.729 | -0.005 | 4.66E-01 | 18837 |
| 25 | OFCmed_L | -1.395 | -2.342 | -0.448 | 0.483 | -2.888 | -0.021 | 3.88E-03 | 18837 |
| 26 | OFCmed_R | -0.765 | -1.868 | 0.338 | 0.563 | -1.360 | -0.010 | 1.74E-01 | 18837 |
| 27 | OFCant_L | -0.758 | -1.853 | 0.336 | 0.558 | -1.358 | -0.010 | 1.74E-01 | 18837 |
| 28 | OFCant_R | -1.297 | -2.742 | 0.149 | 0.738 | -1.758 | -0.013 | 7.88E-02 | 18837 |
| 29 | OFCpost_L | -0.542 | -1.912 | 0.827 | 0.699 | -0.776 | -0.006 | 4.38E-01 | 18837 |
| 30 | OFCpost_R | -0.719 | -1.985 | 0.547 | 0.646 | -1.113 | -0.008 | 2.66E-01 | 18837 |
| 31 | OFClat_L | -0.344 | -0.806 | 0.119 | 0.236 | -1.457 | -0.011 | 1.45E-01 | 18837 |
| 32 | OFClat_R | -0.616 | -1.007 | -0.226 | 0.199 | -3.095 | -0.023 | 1.97E-03 | 18837 |
| 33 | Insula_L | -1.948 | -6.186 | 2.290 | 2.162 | -0.901 | -0.007 | 3.68E-01 | 18837 |
| 34 | Insula_R | -2.809 | -6.939 | 1.320 | 2.107 | -1.333 | -0.010 | 1.82E-01 | 18837 |
| 37 | Cingulate_Mid_L | -3.665 | -7.251 | -0.078 | 1.830 | -2.003 | -0.015 | 4.52E-02 | 18837 |
| 38 | Cingulate_Mid_R | -2.341 | -6.165 | 1.482 | 1.951 | -1.200 | -0.009 | 2.30E-01 | 18837 |
| 39 | Cingulate_Post_L | -0.023 | -1.117 | 1.070 | 0.558 | -0.042 | 0.000 | 9.67E-01 | 18837 |
| 40 | Cingulate_Post_R | -0.037 | -0.644 | 0.570 | 0.310 | -0.119 | -0.001 | 9.06E-01 | 18837 |
| 41 | Hippocampus_L | 0.690 | -1.079 | 2.460 | 0.903 | 0.765 | 0.006 | 4.44E-01 | 18837 |
| 42 | Hippocampus_R | 1.257 | -0.580 | 3.093 | 0.937 | 1.341 | 0.010 | 1.80E-01 | 18837 |
| 43 | ParaHippocampal_L | 0.264 | -1.308 | 1.836 | 0.802 | 0.329 | 0.002 | 7.42E-01 | 18837 |
| 44 | ParaHippocampal_R | -0.250 | -2.236 | 1.735 | 1.013 | -0.247 | -0.002 | 8.05E-01 | 18837 |
| 45 | Amygdala_L | -0.147 | -0.758 | 0.464 | 0.312 | -0.471 | -0.003 | 6.37E-01 | 18837 |
| 46 | Amygdala_R | -0.419 | -1.045 | 0.206 | 0.319 | -1.315 | -0.010 | 1.89E-01 | 18837 |
| 47 | Calcarine_L | 4.371 | -1.459 | 10.200 | 2.974 | 1.470 | 0.011 | 1.42E-01 | 18837 |
| 48 | Calcarine_R | 3.389 | -1.482 | 8.259 | 2.485 | 1.364 | 0.010 | 1.73E-01 | 18837 |
| 49 | Cuneus_L | 0.688 | -2.972 | 4.348 | 1.867 | 0.368 | 0.003 | 7.13E-01 | 18837 |
| 50 | Cuneus_R | -0.583 | -3.382 | 2.216 | 1.428 | -0.408 | -0.003 | 6.83E-01 | 18837 |
| 51 | Lingual_L | 1.385 | -2.124 | 4.894 | 1.790 | 0.774 | 0.006 | 4.39E-01 | 18837 |
| 52 | Lingual_R | 0.746 | -2.929 | 4.421 | 1.875 | 0.398 | 0.003 | 6.91E-01 | 18837 |
| 53 | Occipital_Sup_L | -2.255 | -4.668 | 0.157 | 1.231 | -1.833 | -0.013 | 6.69E-02 | 18837 |
| 54 | Occipital_Sup_R | -1.560 | -4.175 | 1.054 | 1.334 | -1.170 | -0.009 | 2.42E-01 | 18837 |
| 55 | Occipital_Mid_L | 1.396 | -4.253 | 7.045 | 2.882 | 0.484 | 0.004 | 6.28E-01 | 18837 |
| 56 | Occipital_Mid_R | -2.791 | -6.725 | 1.143 | 2.007 | -1.390 | -0.010 | 1.64E-01 | 18837 |
| 57 | Occipital_Inf_L | 1.882 | 0.008 | 3.757 | 0.956 | 1.968 | 0.014 | 4.90E-02 | 18837 |
| 58 | Occipital_Inf_R | -1.525 | -3.692 | 0.641 | 1.105 | -1.380 | -0.010 | 1.68E-01 | 18837 |
| 59 | Fusiform_L | 1.774 | -2.332 | 5.880 | 2.095 | 0.847 | 0.006 | 3.97E-01 | 18837 |
| 60 | Fusiform_R | 0.820 | -3.692 | 5.331 | 2.302 | 0.356 | 0.003 | 7.22E-01 | 18837 |
| 61 | Postcentral_L | 0.878 | -5.302 | 7.057 | 3.153 | 0.278 | 0.002 | 7.81E-01 | 18837 |
| 62 | Postcentral_R | 2.792 | -3.533 | 9.118 | 3.227 | 0.865 | 0.006 | 3.87E-01 | 18837 |
| 63 | Parietal_Sup_L | -2.997 | -6.556 | 0.562 | 1.816 | -1.650 | -0.012 | 9.89E-02 | 18837 |
| 64 | Parietal_Sup_R | -2.187 | -5.343 | 0.969 | 1.610 | -1.358 | -0.010 | 1.74E-01 | 18837 |
| 65 | Parietal_Inf_L | -0.151 | -4.811 | 4.508 | 2.377 | -0.064 | 0.000 | 9.49E-01 | 18837 |
| 66 | Parietal_Inf_R | -0.946 | -3.966 | 2.074 | 1.541 | -0.614 | -0.004 | 5.39E-01 | 18837 |
| 67 | SupraMarginal_L | -1.142 | -3.873 | 1.590 | 1.394 | -0.819 | -0.006 | 4.13E-01 | 18837 |

| | | | | | | | | | |
|---|---|---|---|---|---|---|---|---|---|
| 68 | SupraMarginal_R | -3.258 | -7.125 | 0.609 | 1.973 | -1.652 | -0.012 | 9.86E-02 | 18837 |
| 69 | Angular_L | 1.551 | -1.132 | 4.235 | 1.369 | 1.133 | 0.008 | 2.57E-01 | 18837 |
| 70 | Angular_R | 2.325 | -1.270 | 5.921 | 1.834 | 1.268 | 0.009 | 2.05E-01 | 18837 |
| 71 | Precuneus_L | 1.055 | -5.484 | 7.595 | 3.336 | 0.316 | 0.002 | 7.52E-01 | 18837 |
| 72 | Precuneus_R | -1.278 | -7.103 | 4.546 | 2.971 | -0.430 | -0.003 | 6.67E-01 | 18837 |
| 73 | Paracentral_Lobule_L | -1.156 | -4.150 | 1.839 | 1.528 | -0.757 | -0.006 | 4.49E-01 | 18837 |
| 74 | Paracentral_Lobule_R | 0.129 | -2.015 | 2.274 | 1.094 | 0.118 | 0.001 | 9.06E-01 | 18837 |
| 75 | Caudate_L | -0.380 | -2.771 | 2.011 | 1.220 | -0.312 | -0.002 | 7.55E-01 | 18837 |
| 76 | Caudate_R | -1.240 | -3.950 | 1.469 | 1.382 | -0.897 | -0.007 | 3.70E-01 | 18837 |
| 77 | Putamen_L | -2.868 | -6.499 | 0.764 | 1.853 | -1.548 | -0.011 | 1.22E-01 | 18837 |
| 78 | Putamen_R | -3.558 | -7.257 | 0.140 | 1.887 | -1.886 | -0.014 | 5.94E-02 | 18837 |
| 79 | Pallidum_L | -0.117 | -0.440 | 0.206 | 0.165 | -0.712 | -0.005 | 4.76E-01 | 18837 |
| 80 | Pallidum_R | -0.122 | -0.476 | 0.233 | 0.181 | -0.673 | -0.005 | 5.01E-01 | 18837 |
| 83 | Heschl_L | -0.094 | -0.748 | 0.560 | 0.334 | -0.282 | -0.002 | 7.78E-01 | 18837 |
| 84 | Heschl_R | -0.189 | -0.920 | 0.542 | 0.373 | -0.506 | -0.004 | 6.13E-01 | 18837 |
| 85 | Temporal_Sup_L | -2.232 | -6.954 | 2.490 | 2.409 | -0.926 | -0.007 | 3.54E-01 | 18837 |
| 86 | Temporal_Sup_R | -0.753 | -5.982 | 4.475 | 2.668 | -0.282 | -0.002 | 7.78E-01 | 18837 |
| 87 | Temporal_Pole_Sup_L | -0.506 | -2.744 | 1.733 | 1.142 | -0.443 | -0.003 | 6.58E-01 | 18837 |
| 88 | Temporal_Pole_Sup_R | -1.007 | -3.363 | 1.349 | 1.202 | -0.838 | -0.006 | 4.02E-01 | 18837 |
| 89 | Temporal_Mid_L | -1.844 | -9.724 | 6.036 | 4.020 | -0.459 | -0.003 | 6.46E-01 | 18837 |
| 90 | Temporal_Mid_R | 2.158 | -4.835 | 9.150 | 3.567 | 0.605 | 0.004 | 5.45E-01 | 18837 |
| 91 | Temporal_Pole_Mid_L | 0.878 | -1.003 | 2.759 | 0.960 | 0.915 | 0.007 | 3.60E-01 | 18837 |
| 92 | Temporal_Pole_Mid_R | -0.112 | -2.396 | 2.173 | 1.166 | -0.096 | -0.001 | 9.24E-01 | 18837 |
| 93 | Temporal_Inf_L | -0.543 | -5.957 | 4.871 | 2.762 | -0.197 | -0.001 | 8.44E-01 | 18837 |
| 94 | Temporal_Inf_R | -0.407 | -6.311 | 5.496 | 3.012 | -0.135 | -0.001 | 8.92E-01 | 18837 |
| 95 | Cerebellum_Crus1_L | 2.752 | -3.679 | 9.183 | 3.281 | 0.839 | 0.006 | 4.02E-01 | 18837 |
| 96 | Cerebellum_Crus1_R | -0.187 | -6.743 | 6.369 | 3.345 | -0.056 | 0.000 | 9.56E-01 | 18837 |
| 97 | Cerebellum_Crus2_L | -2.459 | -7.559 | 2.640 | 2.602 | -0.945 | -0.007 | 3.45E-01 | 18837 |
| 98 | Cerebellum_Crus2_R | -2.112 | -6.826 | 2.602 | 2.405 | -0.878 | -0.006 | 3.80E-01 | 18837 |
| 99 | Cerebellum_3_L | 0.014 | -0.328 | 0.356 | 0.174 | 0.079 | 0.001 | 9.37E-01 | 18837 |
| 100 | Cerebellum_3_R | -0.097 | -0.558 | 0.365 | 0.235 | -0.411 | -0.003 | 6.81E-01 | 18837 |
| 101 | Cerebellum_4_5_L | -0.753 | -3.521 | 2.015 | 1.412 | -0.533 | -0.004 | 5.94E-01 | 18837 |
| 102 | Cerebellum_4_5_R | -0.908 | -3.164 | 1.347 | 1.151 | -0.789 | -0.006 | 4.30E-01 | 18837 |
| 103 | Cerebellum_6_L | 1.147 | -3.642 | 5.936 | 2.443 | 0.469 | 0.003 | 6.39E-01 | 18837 |
| 104 | Cerebellum_6_R | 0.947 | -4.097 | 5.991 | 2.573 | 0.368 | 0.003 | 7.13E-01 | 18837 |
| 105 | Cerebellum_7b_L | -0.447 | -2.007 | 1.112 | 0.795 | -0.562 | -0.004 | 5.74E-01 | 18837 |
| 106 | Cerebellum_7b_R | -1.125 | -2.670 | 0.420 | 0.788 | -1.427 | -0.010 | 1.53E-01 | 18837 |
| 107 | Cerebellum_8_L | -2.296 | -7.248 | 2.656 | 2.526 | -0.909 | -0.007 | 3.63E-01 | 18837 |
| 108 | Cerebellum_8_R | -4.445 | -10.890 | 2.000 | 3.288 | -1.352 | -0.010 | 1.76E-01 | 18837 |
| 109 | Cerebellum_9_L | -0.105 | -2.903 | 2.694 | 1.428 | -0.074 | -0.001 | 9.41E-01 | 18837 |
| 110 | Cerebellum_9_R | -0.145 | -2.896 | 2.606 | 1.404 | -0.104 | -0.001 | 9.18E-01 | 18837 |
| 111 | Cerebellum_10_L | 0.065 | -0.133 | 0.264 | 0.101 | 0.644 | 0.005 | 5.20E-01 | 18837 |
| 112 | Cerebellum_10_R | 0.008 | -0.198 | 0.215 | 0.105 | 0.079 | 0.001 | 9.37E-01 | 18837 |
| 113 | Vermis_1_2 | 0.040 | -0.089 | 0.169 | 0.066 | 0.609 | 0.004 | 5.42E-01 | 18837 |
| 114 | Vermis_3 | -0.014 | -0.527 | 0.498 | 0.262 | -0.055 | 0.000 | 9.56E-01 | 18837 |
| 115 | Vermis_4_5 | -0.019 | -1.526 | 1.488 | 0.769 | -0.025 | 0.000 | 9.80E-01 | 18837 |
| 116 | Vermis_6 | 0.365 | -0.475 | 1.204 | 0.428 | 0.851 | 0.006 | 3.95E-01 | 18837 |
| 117 | Vermis_7 | 0.160 | -0.314 | 0.635 | 0.242 | 0.662 | 0.005 | 5.08E-01 | 18837 |
| 118 | Vermis_8 | 0.349 | -0.431 | 1.129 | 0.398 | 0.878 | 0.006 | 3.80E-01 | 18837 |

| | | | | | | | | | |
|---|---|---|---|---|---|---|---|---|---|
| 119 | Vermis_9 | -0.128 | -0.761 | 0.504 | 0.323 | -0.398 | -0.003 | 6.91E-01 | 18837 |
| 120 | Vermis_10 | 0.016 | -0.160 | 0.192 | 0.090 | 0.180 | 0.001 | 8.57E-01 | 18837 |
| 121 | Thal_AV_L | 0.006 | -0.065 | 0.076 | 0.036 | 0.160 | 0.001 | 8.73E-01 | 18837 |
| 122 | Thal_AV_R | 0.042 | -0.059 | 0.142 | 0.051 | 0.811 | 0.006 | 4.17E-01 | 18837 |
| 123 | Thal_LP_L | -0.032 | -0.111 | 0.046 | 0.040 | -0.811 | -0.006 | 4.18E-01 | 18837 |
| 124 | Thal_LP_R | -0.037 | -0.139 | 0.065 | 0.052 | -0.706 | -0.005 | 4.80E-01 | 18837 |
| 125 | Thal_VA_L | 0.026 | -0.057 | 0.109 | 0.043 | 0.612 | 0.004 | 5.40E-01 | 18837 |
| 126 | Thal_VA_R | 0.048 | -0.055 | 0.150 | 0.052 | 0.912 | 0.007 | 3.62E-01 | 18837 |
| 127 | Thal_VL_L | -0.127 | -0.364 | 0.111 | 0.121 | -1.044 | -0.008 | 2.96E-01 | 18837 |
| 128 | Thal_VL_R | -0.205 | -0.521 | 0.110 | 0.161 | -1.277 | -0.009 | 2.02E-01 | 18837 |
| 129 | Thal_VPL_L | -0.114 | -0.396 | 0.169 | 0.144 | -0.788 | -0.006 | 4.31E-01 | 18837 |
| 130 | Thal_VPL_R | -0.193 | -0.490 | 0.105 | 0.152 | -1.270 | -0.009 | 2.04E-01 | 18837 |
| 131 | Thal_IL_L | -0.111 | -0.233 | 0.011 | 0.062 | -1.777 | -0.013 | 7.56E-02 | 18837 |
| 132 | Thal_IL_R | -0.082 | -0.221 | 0.057 | 0.071 | -1.162 | -0.008 | 2.45E-01 | 18837 |
| 133 | Thal_Re_L | -0.001 | -0.001 | 0.000 | 0.000 | -2.328 | -0.017 | 1.99E-02 | 18837 |
| 134 | Thal_Re_R | -0.001 | -0.005 | 0.003 | 0.002 | -0.616 | -0.004 | 5.38E-01 | 18837 |
| 135 | Thal_MDm_L | -0.636 | -1.542 | 0.270 | 0.462 | -1.376 | -0.010 | 1.69E-01 | 18837 |
| 136 | Thal_MDm_R | -0.413 | -1.466 | 0.640 | 0.537 | -0.768 | -0.006 | 4.42E-01 | 18837 |
| 137 | Thal_MDl_L | -0.168 | -0.325 | -0.011 | 0.080 | -2.098 | -0.015 | 3.59E-02 | 18837 |
| 138 | Thal_MDl_R | -0.093 | -0.271 | 0.085 | 0.091 | -1.024 | -0.007 | 3.06E-01 | 18837 |
| 139 | Thal_LGN_L | -0.017 | -0.041 | 0.007 | 0.012 | -1.416 | -0.010 | 1.57E-01 | 18837 |
| 140 | Thal_LGN_R | -0.064 | -0.114 | -0.013 | 0.026 | -2.462 | -0.018 | 1.38E-02 | 18837 |
| 141 | Thal_MGN_L | -0.058 | -0.100 | -0.015 | 0.022 | -2.674 | -0.019 | 7.51E-03 | 18837 |
| 142 | Thal_MGN_R | -0.067 | -0.113 | -0.021 | 0.024 | -2.844 | -0.021 | 4.46E-03 | 18837 |
| 143 | Thal_PuA_L | -0.062 | -0.112 | -0.012 | 0.026 | -2.423 | -0.018 | 1.54E-02 | 18837 |
| 144 | Thal_PuA_R | -0.134 | -0.238 | -0.030 | 0.053 | -2.519 | -0.018 | 1.18E-02 | 18837 |
| 145 | Thal_PuM_L | -0.967 | -1.964 | 0.031 | 0.509 | -1.900 | -0.014 | 5.75E-02 | 18837 |
| 146 | Thal_PuM_R | -0.977 | -2.220 | 0.266 | 0.634 | -1.541 | -0.011 | 1.23E-01 | 18837 |
| 147 | Thal_PuL_L | -0.157 | -0.339 | 0.025 | 0.093 | -1.686 | -0.012 | 9.18E-02 | 18837 |
| 148 | Thal_PuL_R | -0.212 | -0.501 | 0.078 | 0.148 | -1.435 | -0.010 | 1.51E-01 | 18837 |
| 149 | Thal_PuI_L | -0.012 | -0.062 | 0.037 | 0.025 | -0.487 | -0.004 | 6.26E-01 | 18837 |
| 150 | Thal_PuI_R | -0.011 | -0.077 | 0.054 | 0.033 | -0.331 | -0.002 | 7.41E-01 | 18837 |
| 151 | ACC_sub_L | -0.539 | -1.110 | 0.032 | 0.291 | -1.850 | -0.013 | 6.43E-02 | 18837 |
| 152 | ACC_sub_R | -0.384 | -0.917 | 0.149 | 0.272 | -1.413 | -0.010 | 1.58E-01 | 18837 |
| 153 | ACC_pre_L | -1.458 | -2.922 | 0.006 | 0.747 | -1.953 | -0.014 | 5.09E-02 | 18837 |
| 154 | ACC_pre_R | -1.297 | -2.707 | 0.113 | 0.719 | -1.803 | -0.013 | 7.14E-02 | 18837 |
| 155 | ACC_sup_L | -3.080 | -4.670 | -1.490 | 0.811 | -3.797 | -0.028 | 1.47E-04 | 18837 |
| 156 | ACC_sup_R | -3.014 | -4.661 | -1.367 | 0.840 | -3.588 | -0.026 | 3.35E-04 | 18837 |
| 157 | N_Acc_L | 0.114 | -0.365 | 0.593 | 0.244 | 0.467 | 0.003 | 6.40E-01 | 18837 |
| 158 | N_Acc_R | 0.067 | -0.347 | 0.482 | 0.211 | 0.319 | 0.002 | 7.50E-01 | 18837 |
| 159 | VTA_L | -0.004 | -0.021 | 0.013 | 0.009 | -0.485 | -0.004 | 6.28E-01 | 18837 |
| 160 | VTA_R | -0.002 | -0.017 | 0.014 | 0.008 | -0.211 | -0.002 | 8.33E-01 | 18837 |
| 161 | SN_pc_L | -0.021 | -0.048 | 0.006 | 0.014 | -1.491 | -0.011 | 1.36E-01 | 18837 |
| 162 | SN_pc_R | -0.021 | -0.056 | 0.015 | 0.018 | -1.151 | -0.008 | 2.50E-01 | 18837 |
| 163 | SN_pr_L | -0.030 | -0.066 | 0.007 | 0.019 | -1.577 | -0.011 | 1.15E-01 | 18837 |
| 164 | SN_pr_R | -0.009 | -0.051 | 0.032 | 0.021 | -0.450 | -0.003 | 6.52E-01 | 18837 |
| 165 | Red_N_L | -0.047 | -0.093 | -0.002 | 0.023 | -2.047 | -0.015 | 4.06E-02 | 18837 |
| 166 | Red_N_R | -0.020 | -0.069 | 0.029 | 0.025 | -0.797 | -0.006 | 4.25E-01 | 18837 |
| 167 | LC_L | 0.006 | -0.011 | 0.023 | 0.009 | 0.663 | 0.005 | 5.07E-01 | 18837 |

| | | | | | | | | | |
|---|---|---|---|---|---|---|---|---|---|
| 168 | LC_R | 0.006 | -0.012 | 0.025 | 0.009 | 0.680 | 0.005 | 4.96E-01 | 18837 |
| 169 | Raphe_D | -0.029 | -0.094 | 0.037 | 0.033 | -0.861 | -0.006 | 3.89E-01 | 18837 |
| 170 | Raphe_M | 0.006 | -0.014 | 0.025 | 0.010 | 0.545 | 0.004 | 5.86E-01 | 18837 |

**Table S4: Associations between grey matter volumes and diet phenotypes (baseline), with follow-up diets as covariates.**

| Number | AAL 3 region | beta | 95%CI_l | 95%CI_h | se | t | r | p | df.e | beta | 95%CI_l | 95%CI_h | se | t | r | p | df.e |
|---|---|---|---|---|---|---|---|---|---|---|---|---|---|---|---|---|---|
| | | Coffee intake | | | | | | | | Cereal intake | | | | | | | |
| 0 | Total brain | -163.196 | -261.077 | -65.316 | 49.937 | -3.268 | -0.024 | 1.08E-03 | 18620 | 152.006 | 92.479 | 211.534 | 30.370 | 5.005 | 0.037 | 5.63E-07 | 18704 |
| 1 | Precentral_L | -3.072 | -6.522 | 0.377 | 1.760 | -1.746 | -0.013 | 8.09E-02 | 18620 | 3.426 | 1.324 | 5.528 | 1.073 | 3.195 | 0.023 | 1.40E-03 | 18704 |
| 2 | Precentral_R | -2.628 | -5.789 | 0.533 | 1.613 | -1.630 | -0.012 | 1.03E-01 | 18620 | 2.830 | 0.905 | 4.755 | 0.982 | 2.882 | 0.021 | 3.96E-03 | 18704 |
| 3 | Frontal_Sup_2_L | -5.921 | -9.560 | -2.281 | 1.857 | -3.189 | -0.023 | 1.43E-03 | 18620 | 4.030 | 1.814 | 6.247 | 1.131 | 3.564 | 0.026 | 3.67E-04 | 18704 |
| 4 | Frontal_Sup_2_R | -5.199 | -9.209 | -1.189 | 2.046 | -2.541 | -0.019 | 1.11E-02 | 18620 | 5.093 | 2.651 | 7.535 | 1.246 | 4.087 | 0.030 | 4.38E-05 | 18704 |
| 5 | Frontal_Mid_2_L | -4.645 | -8.468 | -0.822 | 1.950 | -2.381 | -0.017 | 1.73E-02 | 18620 | 2.109 | -0.221 | 4.438 | 1.188 | 1.774 | 0.013 | 7.60E-02 | 18704 |
| 6 | Frontal_Mid_2_R | -4.718 | -9.043 | -0.394 | 2.206 | -2.139 | -0.016 | 3.25E-02 | 18620 | 3.981 | 1.347 | 6.614 | 1.343 | 2.963 | 0.022 | 3.05E-03 | 18704 |
| 7 | Frontal_Inf_Oper_L | -0.640 | -1.770 | 0.490 | 0.577 | -1.110 | -0.008 | 2.67E-01 | 18620 | 1.015 | 0.327 | 1.704 | 0.351 | 2.889 | 0.021 | 3.86E-03 | 18704 |
| 8 | Frontal_Inf_Oper_R | -1.717 | -3.135 | -0.299 | 0.723 | -2.373 | -0.017 | 1.76E-02 | 18620 | 0.854 | -0.009 | 1.717 | 0.440 | 1.940 | 0.014 | 5.24E-02 | 18704 |
| 9 | Frontal_Inf_Tri_L | -1.113 | -3.513 | 1.287 | 1.224 | -0.909 | -0.007 | 3.63E-01 | 18620 | 1.912 | 0.451 | 3.373 | 0.746 | 2.565 | 0.019 | 1.03E-02 | 18704 |
| 10 | Frontal_Inf_Tri_R | -1.091 | -2.976 | 0.794 | 0.962 | -1.135 | -0.008 | 2.56E-01 | 18620 | 1.614 | 0.467 | 2.762 | 0.586 | 2.757 | 0.020 | 5.84E-03 | 18704 |
| 11 | Frontal_Inf_Orb_2_L | -0.502 | -1.219 | 0.215 | 0.366 | -1.372 | -0.010 | 1.70E-01 | 18620 | 0.387 | -0.050 | 0.824 | 0.223 | 1.737 | 0.013 | 8.24E-02 | 18704 |
| 12 | Frontal_Inf_Orb_2_R | -1.072 | -1.823 | -0.322 | 0.383 | -2.801 | -0.021 | 5.11E-03 | 18620 | 0.618 | 0.161 | 1.075 | 0.233 | 2.652 | 0.019 | 8.01E-03 | 18704 |
| 13 | Rolandic_Oper_L | -0.514 | -1.600 | 0.572 | 0.554 | -0.928 | -0.007 | 3.53E-01 | 18620 | 1.001 | 0.340 | 1.663 | 0.337 | 2.967 | 0.022 | 3.01E-03 | 18704 |
| 14 | Rolandic_Oper_R | -1.834 | -3.387 | -0.280 | 0.793 | -2.314 | -0.017 | 2.07E-02 | 18620 | 1.446 | 0.500 | 2.392 | 0.483 | 2.997 | 0.022 | 2.73E-03 | 18704 |
| 15 | Supp_Motor_Area_L | -2.107 | -4.689 | 0.474 | 1.317 | -1.600 | -0.012 | 1.10E-01 | 18620 | 1.007 | -0.566 | 2.580 | 0.802 | 1.255 | 0.009 | 2.10E-01 | 18704 |
| 16 | Supp_Motor_Area_R | -2.287 | -5.138 | 0.564 | 1.455 | -1.572 | -0.012 | 1.16E-01 | 18620 | 1.702 | -0.036 | 3.439 | 0.886 | 1.920 | 0.014 | 5.49E-02 | 18704 |
| 17 | Olfactory_L | -0.215 | -0.599 | 0.169 | 0.196 | -1.098 | -0.008 | 2.72E-01 | 18620 | 0.298 | 0.065 | 0.532 | 0.119 | 2.502 | 0.018 | 1.24E-02 | 18704 |
| 18 | Olfactory_R | -0.226 | -0.611 | 0.159 | 0.196 | -1.149 | -0.008 | 2.50E-01 | 18620 | 0.246 | 0.012 | 0.480 | 0.120 | 2.058 | 0.015 | 3.96E-02 | 18704 |
| 19 | Frontal_Sup_Medial_L | -3.966 | -6.383 | -1.550 | 1.233 | -3.217 | -0.024 | 1.30E-03 | 18620 | 3.105 | 1.635 | 4.575 | 0.750 | 4.140 | 0.030 | 3.48E-05 | 18704 |
| 20 | Frontal_Sup_Medial_R | -2.677 | -4.436 | -0.918 | 0.898 | -2.983 | -0.022 | 2.86E-03 | 18620 | 1.932 | 0.861 | 3.002 | 0.546 | 3.535 | 0.026 | 4.08E-04 | 18704 |
| 21 | Frontal_Med_Orb_L | -0.879 | -1.784 | 0.025 | 0.461 | -1.907 | -0.014 | 5.66E-02 | 18620 | 1.122 | 0.572 | 1.671 | 0.280 | 4.001 | 0.029 | 6.33E-05 | 18704 |
| 22 | Frontal_Med_Orb_R | -0.789 | -1.900 | 0.322 | 0.567 | -1.391 | -0.010 | 1.64E-01 | 18620 | 1.439 | 0.763 | 2.114 | 0.345 | 4.175 | 0.031 | 2.99E-05 | 18704 |
| 23 | Rectus_L | -0.844 | -1.780 | 0.091 | 0.477 | -1.769 | -0.013 | 7.69E-02 | 18620 | 0.950 | 0.381 | 1.519 | 0.290 | 3.272 | 0.024 | 1.07E-03 | 18704 |
| 24 | Rectus_R | -0.676 | -1.541 | 0.189 | 0.441 | -1.533 | -0.011 | 1.25E-01 | 18620 | 0.993 | 0.467 | 1.519 | 0.269 | 3.698 | 0.027 | 2.18E-04 | 18704 |
| 25 | OFCmed_L | -0.473 | -1.015 | 0.069 | 0.276 | -1.711 | -0.013 | 8.70E-02 | 18620 | 0.426 | 0.096 | 0.756 | 0.168 | 2.529 | 0.018 | 1.15E-02 | 18704 |
| 26 | OFCmed_R | -0.304 | -0.936 | 0.328 | 0.322 | -0.942 | -0.007 | 3.46E-01 | 18620 | 0.528 | 0.143 | 0.913 | 0.196 | 2.691 | 0.020 | 7.13E-03 | 18704 |
| 27 | OFCant_L | -0.320 | -0.947 | 0.308 | 0.320 | -0.998 | -0.007 | 3.18E-01 | 18620 | 0.355 | -0.027 | 0.737 | 0.195 | 1.820 | 0.013 | 6.88E-02 | 18704 |
| 28 | OFCant_R | -0.376 | -1.205 | 0.453 | 0.423 | -0.890 | -0.007 | 3.73E-01 | 18620 | 1.004 | 0.499 | 1.508 | 0.257 | 3.902 | 0.029 | 9.57E-05 | 18704 |
| 29 | OFCpost_L | -0.314 | -1.099 | 0.471 | 0.401 | -0.784 | -0.006 | 4.33E-01 | 18620 | 0.397 | -0.082 | 0.875 | 0.244 | 1.626 | 0.012 | 1.04E-01 | 18704 |
| 30 | OFCpost_R | -0.336 | -1.062 | 0.390 | 0.370 | -0.907 | -0.007 | 3.64E-01 | 18620 | 0.629 | 0.187 | 1.071 | 0.225 | 2.792 | 0.020 | 5.25E-03 | 18704 |
| 31 | OFClat_L | -0.216 | -0.480 | 0.049 | 0.135 | -1.596 | -0.012 | 1.11E-01 | 18620 | 0.174 | 0.013 | 0.335 | 0.082 | 2.115 | 0.015 | 3.45E-02 | 18704 |
| 32 | OFClat_R | -0.281 | -0.505 | -0.057 | 0.114 | -2.464 | -0.018 | 1.37E-02 | 18620 | 0.306 | 0.170 | 0.442 | 0.069 | 4.407 | 0.032 | 1.05E-05 | 18704 |
| 33 | Insula_L | -4.212 | -6.634 | -1.789 | 1.236 | -3.408 | -0.025 | 6.56E-04 | 18620 | 2.523 | 1.046 | 4.001 | 0.754 | 3.347 | 0.024 | 8.18E-04 | 18704 |
| 34 | Insula_R | -4.248 | -6.610 | -1.886 | 1.205 | -3.525 | -0.026 | 4.25E-04 | 18620 | 2.653 | 1.215 | 4.092 | 0.734 | 3.615 | 0.026 | 3.02E-04 | 18704 |
| 37 | Cingulate_Mid_L | -3.984 | -6.033 | -1.935 | 1.045 | -3.811 | -0.028 | 1.39E-04 | 18620 | 3.045 | 1.798 | 4.292 | 0.636 | 4.785 | 0.035 | 1.73E-06 | 18704 |
| 38 | Cingulate_Mid_R | -3.903 | -6.088 | -1.717 | 1.115 | -3.500 | -0.026 | 4.67E-04 | 18620 | 3.003 | 1.673 | 4.333 | 0.679 | 4.425 | 0.032 | 9.69E-06 | 18704 |
| 39 | Cingulate_Post_L | -0.180 | -0.808 | 0.447 | 0.320 | -0.563 | -0.004 | 5.73E-01 | 18620 | 0.488 | 0.106 | 0.870 | 0.195 | 2.504 | 0.018 | 1.23E-02 | 18704 |
| 40 | Cingulate_Post_R | -0.153 | -0.501 | 0.195 | 0.178 | -0.861 | -0.006 | 3.89E-01 | 18620 | 0.156 | -0.056 | 0.368 | 0.108 | 1.442 | 0.011 | 1.49E-01 | 18704 |
| 41 | Hippocampus_L | -0.657 | -1.671 | 0.358 | 0.518 | -1.269 | -0.009 | 2.05E-01 | 18620 | 1.084 | 0.466 | 1.701 | 0.315 | 3.440 | 0.025 | 5.82E-04 | 18704 |
| 42 | Hippocampus_R | -0.302 | -1.355 | 0.750 | 0.537 | -0.563 | -0.004 | 5.73E-01 | 18620 | 0.643 | 0.002 | 1.283 | 0.327 | 1.966 | 0.014 | 4.93E-02 | 18704 |
| 43 | ParaHippocampal_L | -0.943 | -1.845 | -0.042 | 0.460 | -2.052 | -0.015 | 4.02E-02 | 18620 | 0.902 | 0.354 | 1.451 | 0.280 | 3.224 | 0.024 | 1.27E-03 | 18704 |
| 44 | ParaHippocampal_R | -0.861 | -1.998 | 0.277 | 0.580 | -1.483 | -0.011 | 1.38E-01 | 18620 | 0.980 | 0.288 | 1.672 | 0.353 | 2.776 | 0.020 | 5.50E-03 | 18704 |
| 45 | Amygdala_L | -0.478 | -0.828 | -0.128 | 0.179 | -2.675 | -0.020 | 7.48E-03 | 18620 | 0.314 | 0.100 | 0.527 | 0.109 | 2.881 | 0.021 | 3.97E-03 | 18704 |
| 46 | Amygdala_R | -0.475 | -0.833 | -0.117 | 0.183 | -2.603 | -0.019 | 9.25E-03 | 18620 | 0.284 | 0.066 | 0.502 | 0.111 | 2.550 | 0.019 | 1.08E-02 | 18704 |

| | | | | | | | | | | | | | | | | | |
|---|---|---|---|---|---|---|---|---|---|---|---|---|---|---|---|---|---|
| 47 | Calcarine_L | -1.981 | -5.321 | 1.358 | 1.704 | -1.163 | -0.009 | 2.45E-01 | 18620 | 0.318 | -1.718 | 2.355 | 1.039 | 0.306 | 0.002 | 7.59E-01 | 18704 |
| 48 | Calcarine_R | -1.633 | -4.424 | 1.157 | 1.424 | -1.147 | -0.008 | 2.51E-01 | 18620 | 0.657 | -1.045 | 2.359 | 0.868 | 0.756 | 0.006 | 4.49E-01 | 18704 |
| 49 | Cuneus_L | -1.922 | -4.018 | 0.174 | 1.069 | -1.798 | -0.013 | 7.22E-02 | 18620 | 0.303 | -0.974 | 1.580 | 0.652 | 0.465 | 0.003 | 6.42E-01 | 18704 |
| 50 | Cuneus_R | -1.290 | -2.893 | 0.313 | 0.818 | -1.577 | -0.012 | 1.15E-01 | 18620 | 0.594 | -0.383 | 1.571 | 0.499 | 1.192 | 0.009 | 2.33E-01 | 18704 |
| 51 | Lingual_L | -1.137 | -3.146 | 0.872 | 1.025 | -1.110 | -0.008 | 2.67E-01 | 18620 | 0.045 | -1.179 | 1.269 | 0.625 | 0.072 | 0.001 | 9.42E-01 | 18704 |
| 52 | Lingual_R | -0.690 | -2.796 | 1.416 | 1.074 | -0.642 | -0.005 | 5.21E-01 | 18620 | 0.736 | -0.547 | 2.019 | 0.655 | 1.124 | 0.008 | 2.61E-01 | 18704 |
| 53 | Occipital_Sup_L | -1.160 | -2.541 | 0.222 | 0.705 | -1.646 | -0.012 | 9.99E-02 | 18620 | 0.815 | -0.026 | 1.657 | 0.429 | 1.899 | 0.014 | 5.76E-02 | 18704 |
| 54 | Occipital_Sup_R | -1.389 | -2.887 | 0.109 | 0.764 | -1.817 | -0.013 | 6.92E-02 | 18620 | 0.907 | -0.005 | 1.819 | 0.465 | 1.949 | 0.014 | 5.13E-02 | 18704 |
| 55 | Occipital_Mid_L | 0.748 | -2.490 | 3.985 | 1.652 | 0.453 | 0.003 | 6.51E-01 | 18620 | 0.163 | -1.811 | 2.136 | 1.007 | 0.161 | 0.001 | 8.72E-01 | 18704 |
| 56 | Occipital_Mid_R | 0.124 | -2.128 | 2.376 | 1.149 | 0.108 | 0.001 | 9.14E-01 | 18620 | -0.141 | -1.514 | 1.232 | 0.700 | -0.201 | -0.001 | 8.40E-01 | 18704 |
| 57 | Occipital_Inf_L | 0.660 | -0.415 | 1.734 | 0.548 | 1.203 | 0.009 | 2.29E-01 | 18620 | -0.455 | -1.111 | 0.200 | 0.334 | -1.361 | -0.010 | 1.73E-01 | 18704 |
| 58 | Occipital_Inf_R | 0.994 | -0.248 | 2.236 | 0.634 | 1.569 | 0.011 | 1.17E-01 | 18620 | -0.234 | -0.990 | 0.523 | 0.386 | -0.605 | -0.004 | 5.45E-01 | 18704 |
| 59 | Fusiform_L | -0.888 | -3.240 | 1.464 | 1.200 | -0.740 | -0.005 | 4.59E-01 | 18620 | 2.018 | 0.586 | 3.450 | 0.731 | 2.762 | 0.020 | 5.74E-03 | 18704 |
| 60 | Fusiform_R | -1.955 | -4.543 | 0.633 | 1.320 | -1.480 | -0.011 | 1.39E-01 | 18620 | 1.015 | -0.558 | 2.588 | 0.803 | 1.265 | 0.009 | 2.06E-01 | 18704 |
| 61 | Postcentral_L | -5.064 | -8.598 | -1.530 | 1.803 | -2.809 | -0.021 | 4.98E-03 | 18620 | 4.358 | 2.206 | 6.510 | 1.098 | 3.970 | 0.029 | 7.22E-05 | 18704 |
| 62 | Postcentral_R | -3.280 | -6.908 | 0.347 | 1.851 | -1.773 | -0.013 | 7.63E-02 | 18620 | 3.221 | 1.015 | 5.428 | 1.126 | 2.862 | 0.021 | 4.22E-03 | 18704 |
| 63 | Parietal_Sup_L | -2.911 | -4.949 | -0.873 | 1.040 | -2.800 | -0.021 | 5.12E-03 | 18620 | -0.579 | -1.821 | 0.663 | 0.634 | -0.914 | -0.007 | 3.61E-01 | 18704 |
| 64 | Parietal_Sup_R | -0.663 | -2.471 | 1.145 | 0.922 | -0.719 | -0.005 | 4.72E-01 | 18620 | 0.161 | -0.941 | 1.263 | 0.562 | 0.286 | 0.002 | 7.75E-01 | 18704 |
| 65 | Parietal_Inf_L | -2.804 | -5.470 | -0.137 | 1.360 | -2.061 | -0.015 | 3.93E-02 | 18620 | 2.212 | 0.587 | 3.836 | 0.829 | 2.668 | 0.020 | 7.63E-03 | 18704 |
| 66 | Parietal_Inf_R | -1.308 | -3.037 | 0.420 | 0.882 | -1.483 | -0.011 | 1.38E-01 | 18620 | 2.229 | 1.176 | 3.282 | 0.537 | 4.150 | 0.030 | 3.34E-05 | 18704 |
| 67 | SupraMarginal_L | -0.576 | -2.140 | 0.988 | 0.798 | -0.722 | -0.005 | 4.70E-01 | 18620 | 1.372 | 0.421 | 2.324 | 0.486 | 2.826 | 0.021 | 4.72E-03 | 18704 |
| 68 | SupraMarginal_R | -1.975 | -4.189 | 0.238 | 1.129 | -1.749 | -0.013 | 8.03E-02 | 18620 | 3.567 | 2.222 | 4.912 | 0.686 | 5.197 | 0.038 | 2.05E-07 | 18704 |
| 69 | Angular_L | -1.069 | -2.605 | 0.467 | 0.784 | -1.364 | -0.010 | 1.73E-01 | 18620 | 0.244 | -0.692 | 1.181 | 0.478 | 0.511 | 0.004 | 6.09E-01 | 18704 |
| 70 | Angular_R | -0.331 | -2.392 | 1.730 | 1.051 | -0.315 | -0.002 | 7.53E-01 | 18620 | 0.726 | -0.530 | 1.981 | 0.641 | 1.133 | 0.008 | 2.57E-01 | 18704 |
| 71 | Precuneus_L | -3.909 | -7.654 | -0.165 | 1.910 | -2.046 | -0.015 | 4.07E-02 | 18620 | 0.651 | -1.630 | 2.932 | 1.164 | 0.559 | 0.004 | 5.76E-01 | 18704 |
| 72 | Precuneus_R | -3.090 | -6.426 | 0.246 | 1.702 | -1.816 | -0.013 | 6.94E-02 | 18620 | 0.155 | -1.877 | 2.187 | 1.037 | 0.150 | 0.001 | 8.81E-01 | 18704 |
| 73 | Paracentral_Lobule_L | -0.902 | -2.617 | 0.814 | 0.875 | -1.030 | -0.008 | 3.03E-01 | 18620 | -0.173 | -1.219 | 0.872 | 0.533 | -0.324 | -0.002 | 7.46E-01 | 18704 |
| 74 | Paracentral_Lobule_R | -0.767 | -1.996 | 0.462 | 0.627 | -1.224 | -0.009 | 2.21E-01 | 18620 | -0.148 | -0.897 | 0.600 | 0.382 | -0.389 | -0.003 | 6.98E-01 | 18704 |
| 75 | Caudate_L | -1.347 | -2.718 | 0.023 | 0.699 | -1.927 | -0.014 | 5.40E-02 | 18620 | -0.383 | -1.218 | 0.452 | 0.426 | -0.899 | -0.007 | 3.69E-01 | 18704 |
| 76 | Caudate_R | -1.439 | -2.991 | 0.112 | 0.792 | -1.818 | -0.013 | 6.91E-02 | 18620 | -0.586 | -1.531 | 0.360 | 0.482 | -1.214 | -0.009 | 2.25E-01 | 18704 |
| 77 | Putamen_L | -3.512 | -5.590 | -1.435 | 1.060 | -3.313 | -0.024 | 9.23E-04 | 18620 | 2.349 | 1.083 | 3.616 | 0.646 | 3.636 | 0.027 | 2.77E-04 | 18704 |
| 78 | Putamen_R | -3.776 | -5.892 | -1.661 | 1.079 | -3.499 | -0.026 | 4.69E-04 | 18620 | 2.157 | 0.866 | 3.447 | 0.658 | 3.277 | 0.024 | 1.05E-03 | 18704 |
| 79 | Pallidum_L | -0.090 | -0.275 | 0.095 | 0.094 | -0.952 | -0.007 | 3.41E-01 | 18620 | 0.114 | 0.001 | 0.226 | 0.057 | 1.975 | 0.014 | 4.83E-02 | 18704 |
| 80 | Pallidum_R | -0.185 | -0.388 | 0.017 | 0.104 | -1.792 | -0.013 | 7.32E-02 | 18620 | 0.090 | -0.034 | 0.213 | 0.063 | 1.421 | 0.010 | 1.55E-01 | 18704 |
| 83 | Heschl_L | -0.372 | -0.747 | 0.003 | 0.191 | -1.944 | -0.014 | 5.19E-02 | 18620 | 0.324 | 0.095 | 0.552 | 0.117 | 2.779 | 0.020 | 5.46E-03 | 18704 |
| 84 | Heschl_R | -0.357 | -0.776 | 0.062 | 0.214 | -1.671 | -0.012 | 9.46E-02 | 18620 | 0.417 | 0.162 | 0.672 | 0.130 | 3.207 | 0.023 | 1.35E-03 | 18704 |
| 85 | Temporal_Sup_L | -3.083 | -5.788 | -0.378 | 1.380 | -2.234 | -0.016 | 2.55E-02 | 18620 | 1.714 | 0.066 | 3.362 | 0.841 | 2.039 | 0.015 | 4.15E-02 | 18704 |
| 86 | Temporal_Sup_R | -1.834 | -4.827 | 1.159 | 1.527 | -1.201 | -0.009 | 2.30E-01 | 18620 | 4.530 | 2.708 | 6.352 | 0.929 | 4.874 | 0.036 | 1.10E-06 | 18704 |
| 87 | Temporal_Pole_Sup_L | -1.497 | -2.778 | -0.215 | 0.654 | -2.289 | -0.017 | 2.21E-02 | 18620 | 1.150 | 0.370 | 1.931 | 0.398 | 2.888 | 0.021 | 3.88E-03 | 18704 |
| 88 | Temporal_Pole_Sup_R | -1.376 | -2.725 | -0.027 | 0.688 | -1.999 | -0.015 | 4.57E-02 | 18620 | 0.906 | 0.084 | 1.728 | 0.419 | 2.161 | 0.016 | 3.07E-02 | 18704 |
| 89 | Temporal_Mid_L | -2.187 | -6.699 | 2.325 | 2.302 | -0.950 | -0.007 | 3.42E-01 | 18620 | 4.717 | 1.970 | 7.464 | 1.402 | 3.366 | 0.025 | 7.66E-04 | 18704 |
| 90 | Temporal_Mid_R | -1.548 | -5.552 | 2.456 | 2.043 | -0.758 | -0.006 | 4.49E-01 | 18620 | 3.994 | 1.555 | 6.434 | 1.245 | 3.209 | 0.023 | 1.33E-03 | 18704 |
| 91 | Temporal_Pole_Mid_L | -0.304 | -1.382 | 0.774 | 0.550 | -0.553 | -0.004 | 5.80E-01 | 18620 | 0.827 | 0.171 | 1.484 | 0.335 | 2.469 | 0.018 | 1.35E-02 | 18704 |
| 92 | Temporal_Pole_Mid_R | -1.010 | -2.320 | 0.299 | 0.668 | -1.512 | -0.011 | 1.30E-01 | 18620 | 1.342 | 0.545 | 2.139 | 0.407 | 3.300 | 0.024 | 9.67E-04 | 18704 |
| 93 | Temporal_Inf_L | 0.534 | -2.569 | 3.636 | 1.583 | 0.337 | 0.002 | 7.36E-01 | 18620 | 1.162 | -0.728 | 3.052 | 0.964 | 1.205 | 0.009 | 2.28E-01 | 18704 |
| 94 | Temporal_Inf_R | -2.347 | -5.730 | 1.035 | 1.726 | -1.360 | -0.010 | 1.74E-01 | 18620 | 1.526 | -0.534 | 3.587 | 1.051 | 1.452 | 0.011 | 1.47E-01 | 18704 |
| 95 | Cerebellum_Crus1_L | -2.425 | -6.108 | 1.258 | 1.879 | -1.291 | -0.009 | 1.97E-01 | 18620 | 2.629 | 0.385 | 4.873 | 1.145 | 2.296 | 0.017 | 2.17E-02 | 18704 |
| 96 | Cerebellum_Crus1_R | -4.372 | -8.129 | -0.615 | 1.917 | -2.281 | -0.017 | 2.26E-02 | 18620 | 3.106 | 0.818 | 5.394 | 1.167 | 2.661 | 0.019 | 7.80E-03 | 18704 |
| 97 | Cerebellum_Crus2_L | -3.519 | -6.443 | -0.595 | 1.492 | -2.359 | -0.017 | 1.83E-02 | 18620 | 3.041 | 1.263 | 4.820 | 0.907 | 3.352 | 0.025 | 8.04E-04 | 18704 |

| | | | | | | | | | | | | | | | | | |
|---|---|---|---|---|---|---|---|---|---|---|---|---|---|---|---|---|---|
| 98 | Cerebellum_Crus2_R | -2.321 | -5.026 | 0.384 | 1.380 | -1.682 | -0.012 | 9.27E-02 | 18620 | 3.445 | 1.799 | 5.090 | 0.839 | 4.104 | 0.030 | 4.08E-05 | 18704 |
| 99 | Cerebellum_3_L | -0.004 | -0.200 | 0.193 | 0.100 | -0.035 | 0.000 | 9.72E-01 | 18620 | 0.089 | -0.030 | 0.209 | 0.061 | 1.464 | 0.011 | 1.43E-01 | 18704 |
| 100 | Cerebellum_3_R | -0.112 | -0.377 | 0.153 | 0.135 | -0.828 | -0.006 | 4.08E-01 | 18620 | 0.136 | -0.025 | 0.298 | 0.082 | 1.659 | 0.012 | 9.72E-02 | 18704 |
| 101 | Cerebellum_4_5_L | 0.844 | -0.744 | 2.432 | 0.810 | 1.042 | 0.008 | 2.97E-01 | 18620 | 0.395 | -0.571 | 1.362 | 0.493 | 0.802 | 0.006 | 4.23E-01 | 18704 |
| 102 | Cerebellum_4_5_R | -0.026 | -1.320 | 1.267 | 0.660 | -0.040 | 0.000 | 9.68E-01 | 18620 | 0.519 | -0.268 | 1.306 | 0.402 | 1.292 | 0.009 | 1.96E-01 | 18704 |
| 103 | Cerebellum_6_L | -0.725 | -3.467 | 2.017 | 1.399 | -0.518 | -0.004 | 6.04E-01 | 18620 | 1.659 | -0.011 | 3.329 | 0.852 | 1.947 | 0.014 | 5.15E-02 | 18704 |
| 104 | Cerebellum_6_R | -1.153 | -4.043 | 1.737 | 1.474 | -0.782 | -0.006 | 4.34E-01 | 18620 | 1.779 | 0.021 | 3.538 | 0.897 | 1.983 | 0.015 | 4.74E-02 | 18704 |
| 105 | Cerebellum_7b_L | -0.411 | -1.305 | 0.483 | 0.456 | -0.901 | -0.007 | 3.68E-01 | 18620 | 0.917 | 0.373 | 1.461 | 0.277 | 3.306 | 0.024 | 9.47E-04 | 18704 |
| 106 | Cerebellum_7b_R | -0.714 | -1.600 | 0.172 | 0.452 | -1.579 | -0.012 | 1.14E-01 | 18620 | 1.161 | 0.622 | 1.699 | 0.275 | 4.224 | 0.031 | 2.42E-05 | 18704 |
| 107 | Cerebellum_8_L | 0.145 | -2.693 | 2.982 | 1.448 | 0.100 | 0.001 | 9.20E-01 | 18620 | 2.756 | 1.029 | 4.483 | 0.881 | 3.128 | 0.023 | 1.76E-03 | 18704 |
| 108 | Cerebellum_8_R | -2.561 | -6.252 | 1.130 | 1.883 | -1.360 | -0.010 | 1.74E-01 | 18620 | 4.365 | 2.119 | 6.612 | 1.146 | 3.808 | 0.028 | 1.40E-04 | 18704 |
| 109 | Cerebellum_9_L | 0.962 | -0.640 | 2.564 | 0.817 | 1.177 | 0.009 | 2.39E-01 | 18620 | 0.938 | -0.039 | 1.916 | 0.499 | 1.881 | 0.014 | 6.00E-02 | 18704 |
| 110 | Cerebellum_9_R | 0.893 | -0.681 | 2.468 | 0.803 | 1.112 | 0.008 | 2.66E-01 | 18620 | 0.736 | -0.225 | 1.697 | 0.490 | 1.502 | 0.011 | 1.33E-01 | 18704 |
| 111 | Cerebellum_10_L | -0.025 | -0.138 | 0.089 | 0.058 | -0.423 | -0.003 | 6.73E-01 | 18620 | -0.008 | -0.077 | 0.061 | 0.035 | -0.226 | -0.002 | 8.21E-01 | 18704 |
| 112 | Cerebellum_10_R | -0.030 | -0.148 | 0.089 | 0.060 | -0.489 | -0.004 | 6.25E-01 | 18620 | -0.020 | -0.092 | 0.052 | 0.037 | -0.545 | -0.004 | 5.85E-01 | 18704 |
| 113 | Vermis_1_2 | -0.067 | -0.141 | 0.007 | 0.038 | -1.778 | -0.013 | 7.54E-02 | 18620 | 0.036 | -0.009 | 0.081 | 0.023 | 1.583 | 0.012 | 1.13E-01 | 18704 |
| 114 | Vermis_3 | -0.071 | -0.365 | 0.223 | 0.150 | -0.471 | -0.003 | 6.38E-01 | 18620 | 0.185 | 0.006 | 0.364 | 0.091 | 2.021 | 0.015 | 4.33E-02 | 18704 |
| 115 | Vermis_4_5 | 0.649 | -0.216 | 1.514 | 0.441 | 1.471 | 0.011 | 1.41E-01 | 18620 | 0.170 | -0.357 | 0.696 | 0.269 | 0.631 | 0.005 | 5.28E-01 | 18704 |
| 116 | Vermis_6 | 0.383 | -0.098 | 0.864 | 0.245 | 1.562 | 0.011 | 1.18E-01 | 18620 | 0.014 | -0.279 | 0.307 | 0.150 | 0.096 | 0.001 | 9.24E-01 | 18704 |
| 117 | Vermis_7 | 0.090 | -0.182 | 0.362 | 0.139 | 0.650 | 0.005 | 5.16E-01 | 18620 | 0.085 | -0.081 | 0.251 | 0.085 | 1.004 | 0.007 | 3.15E-01 | 18704 |
| 118 | Vermis_8 | 0.265 | -0.182 | 0.712 | 0.228 | 1.162 | 0.009 | 2.45E-01 | 18620 | 0.165 | -0.107 | 0.438 | 0.139 | 1.189 | 0.009 | 2.34E-01 | 18704 |
| 119 | Vermis_9 | 0.196 | -0.166 | 0.558 | 0.185 | 1.061 | 0.008 | 2.89E-01 | 18620 | 0.287 | 0.067 | 0.508 | 0.113 | 2.551 | 0.019 | 1.08E-02 | 18704 |
| 120 | Vermis_10 | 0.027 | -0.074 | 0.128 | 0.052 | 0.520 | 0.004 | 6.03E-01 | 18620 | 0.043 | -0.019 | 0.104 | 0.031 | 1.360 | 0.010 | 1.74E-01 | 18704 |
| 121 | Thal_AV_L | -0.063 | -0.104 | -0.023 | 0.021 | -3.073 | -0.023 | 2.12E-03 | 18620 | 0.013 | -0.012 | 0.037 | 0.013 | 1.001 | 0.007 | 3.17E-01 | 18704 |
| 122 | Thal_AV_R | -0.077 | -0.135 | -0.019 | 0.029 | -2.612 | -0.019 | 9.00E-03 | 18620 | 0.015 | -0.020 | 0.050 | 0.018 | 0.851 | 0.006 | 3.95E-01 | 18704 |
| 123 | Thal_LP_L | -0.077 | -0.122 | -0.033 | 0.023 | -3.387 | -0.025 | 7.09E-04 | 18620 | 0.051 | 0.023 | 0.078 | 0.014 | 3.645 | 0.027 | 2.68E-04 | 18704 |
| 124 | Thal_LP_R | -0.108 | -0.166 | -0.049 | 0.030 | -3.617 | -0.026 | 2.99E-04 | 18620 | 0.074 | 0.038 | 0.109 | 0.018 | 4.053 | 0.030 | 5.09E-05 | 18704 |
| 125 | Thal_VA_L | -0.011 | -0.059 | 0.037 | 0.024 | -0.440 | -0.003 | 6.60E-01 | 18620 | -0.031 | -0.060 | -0.002 | 0.015 | -2.064 | -0.015 | 3.90E-02 | 18704 |
| 126 | Thal_VA_R | -0.006 | -0.065 | 0.053 | 0.030 | -0.205 | -0.002 | 8.37E-01 | 18620 | -0.039 | -0.075 | -0.003 | 0.018 | -2.150 | -0.016 | 3.15E-02 | 18704 |
| 127 | Thal_VL_L | -0.154 | -0.290 | -0.018 | 0.069 | -2.212 | -0.016 | 2.70E-02 | 18620 | 0.082 | -0.001 | 0.164 | 0.042 | 1.927 | 0.014 | 5.40E-02 | 18704 |
| 128 | Thal_VL_R | -0.247 | -0.427 | -0.066 | 0.092 | -2.674 | -0.020 | 7.51E-03 | 18620 | 0.137 | 0.027 | 0.247 | 0.056 | 2.436 | 0.018 | 1.48E-02 | 18704 |
| 129 | Thal_VPL_L | -0.148 | -0.310 | 0.014 | 0.083 | -1.796 | -0.013 | 7.25E-02 | 18620 | 0.238 | 0.139 | 0.337 | 0.050 | 4.726 | 0.035 | 2.31E-06 | 18704 |
| 130 | Thal_VPL_R | -0.258 | -0.428 | -0.088 | 0.087 | -2.970 | -0.022 | 2.98E-03 | 18620 | 0.227 | 0.124 | 0.331 | 0.053 | 4.295 | 0.031 | 1.75E-05 | 18704 |
| 131 | Thal_IL_L | -0.131 | -0.201 | -0.061 | 0.036 | -3.669 | -0.027 | 2.44E-04 | 18620 | 0.100 | 0.057 | 0.142 | 0.022 | 4.577 | 0.033 | 4.76E-06 | 18704 |
| 132 | Thal_IL_R | -0.200 | -0.280 | -0.121 | 0.040 | -4.948 | -0.036 | 7.56E-07 | 18620 | 0.116 | 0.067 | 0.164 | 0.025 | 4.684 | 0.034 | 2.84E-06 | 18704 |
| 133 | Thal_Re_L | 0.000 | 0.000 | 0.000 | 0.000 | -0.513 | -0.004 | 6.08E-01 | 18620 | 0.000 | 0.000 | 0.000 | 0.000 | 0.298 | 0.002 | 7.66E-01 | 18704 |
| 134 | Thal_Re_R | -0.003 | -0.005 | -0.001 | 0.001 | -2.434 | -0.018 | 1.49E-02 | 18620 | 0.001 | 0.000 | 0.003 | 0.001 | 1.878 | 0.014 | 6.04E-02 | 18704 |
| 135 | Thal_MDm_L | -1.291 | -1.810 | -0.773 | 0.264 | -4.884 | -0.036 | 1.05E-06 | 18620 | 0.760 | 0.444 | 1.076 | 0.161 | 4.714 | 0.034 | 2.44E-06 | 18704 |
| 136 | Thal_MDm_R | -1.568 | -2.170 | -0.966 | 0.307 | -5.105 | -0.037 | 3.34E-07 | 18620 | 0.864 | 0.497 | 1.231 | 0.187 | 4.614 | 0.034 | 3.97E-06 | 18704 |
| 137 | Thal_MDl_L | -0.233 | -0.323 | -0.144 | 0.046 | -5.096 | -0.037 | 3.51E-07 | 18620 | 0.151 | 0.096 | 0.205 | 0.028 | 5.395 | 0.039 | 6.92E-08 | 18704 |
| 138 | Thal_MDl_R | -0.263 | -0.365 | -0.161 | 0.052 | -5.052 | -0.037 | 4.41E-07 | 18620 | 0.163 | 0.101 | 0.226 | 0.032 | 5.156 | 0.038 | 2.55E-07 | 18704 |
| 139 | Thal_LGN_L | -0.007 | -0.020 | 0.007 | 0.007 | -0.943 | -0.007 | 3.46E-01 | 18620 | 0.009 | 0.001 | 0.018 | 0.004 | 2.176 | 0.016 | 2.96E-02 | 18704 |
| 140 | Thal_LGN_R | -0.021 | -0.050 | 0.008 | 0.015 | -1.412 | -0.010 | 1.58E-01 | 18620 | 0.034 | 0.016 | 0.052 | 0.009 | 3.785 | 0.028 | 1.54E-04 | 18704 |
| 141 | Thal_MGN_L | -0.024 | -0.049 | 0.000 | 0.012 | -1.962 | -0.014 | 4.98E-02 | 18620 | 0.037 | 0.022 | 0.052 | 0.008 | 4.918 | 0.036 | 8.80E-07 | 18704 |
| 142 | Thal_MGN_R | -0.041 | -0.068 | -0.015 | 0.013 | -3.049 | -0.022 | 2.30E-03 | 18620 | 0.037 | 0.021 | 0.053 | 0.008 | 4.551 | 0.033 | 5.37E-06 | 18704 |
| 143 | Thal_PuA_L | -0.030 | -0.059 | -0.002 | 0.015 | -2.075 | -0.015 | 3.80E-02 | 18620 | 0.042 | 0.025 | 0.060 | 0.009 | 4.736 | 0.035 | 2.20E-06 | 18704 |
| 144 | Thal_PuA_R | -0.101 | -0.161 | -0.041 | 0.030 | -3.315 | -0.024 | 9.18E-04 | 18620 | 0.083 | 0.047 | 0.120 | 0.019 | 4.488 | 0.033 | 7.23E-06 | 18704 |
| 145 | Thal_PuM_L | -0.935 | -1.505 | -0.364 | 0.291 | -3.211 | -0.024 | 1.33E-03 | 18620 | 0.810 | 0.463 | 1.158 | 0.177 | 4.568 | 0.033 | 4.95E-06 | 18704 |
| 146 | Thal_PuM_R | -1.457 | -2.168 | -0.746 | 0.363 | -4.017 | -0.029 | 5.92E-05 | 18620 | 0.934 | 0.501 | 1.368 | 0.221 | 4.226 | 0.031 | 2.39E-05 | 18704 |

| | | | | | | | | | | | | | | | | | |
|---|---|---|---|---|---|---|---|---|---|---|---|---|---|---|---|---|---|
| 147 | Thal_PuL_L | -0.193 | -0.297 | -0.088 | 0.053 | -3.626 | -0.027 | 2.89E-04 | 18620 | 0.163 | 0.100 | 0.227 | 0.032 | 5.039 | 0.037 | 4.72E-07 | 18704 |
| 148 | Thal_PuL_R | -0.370 | -0.536 | -0.205 | 0.084 | -4.382 | -0.032 | 1.18E-05 | 18620 | 0.244 | 0.143 | 0.345 | 0.051 | 4.745 | 0.035 | 2.10E-06 | 18704 |
| 149 | Thal_Pul_L | -0.015 | -0.044 | 0.013 | 0.014 | -1.068 | -0.008 | 2.86E-01 | 18620 | 0.028 | 0.011 | 0.046 | 0.009 | 3.204 | 0.023 | 1.36E-03 | 18704 |
| 150 | Thal_Pul_R | -0.024 | -0.061 | 0.014 | 0.019 | -1.229 | -0.009 | 2.19E-01 | 18620 | 0.030 | 0.007 | 0.053 | 0.012 | 2.549 | 0.019 | 1.08E-02 | 18704 |
| 151 | ACC_sub_L | 0.052 | -0.275 | 0.379 | 0.167 | 0.314 | 0.002 | 7.54E-01 | 18620 | 0.089 | -0.111 | 0.288 | 0.102 | 0.870 | 0.006 | 3.84E-01 | 18704 |
| 152 | ACC_sub_R | -0.050 | -0.355 | 0.256 | 0.156 | -0.319 | -0.002 | 7.50E-01 | 18620 | 0.068 | -0.118 | 0.255 | 0.095 | 0.720 | 0.005 | 4.71E-01 | 18704 |
| 153 | ACC_pre_L | -0.508 | -1.346 | 0.330 | 0.428 | -1.189 | -0.009 | 2.34E-01 | 18620 | 1.053 | 0.542 | 1.563 | 0.260 | 4.043 | 0.030 | 5.30E-05 | 18704 |
| 154 | ACC_pre_R | -0.636 | -1.444 | 0.171 | 0.412 | -1.545 | -0.011 | 1.22E-01 | 18620 | 0.816 | 0.324 | 1.307 | 0.251 | 3.251 | 0.024 | 1.15E-03 | 18704 |
| 155 | ACC_sup_L | -0.798 | -1.708 | 0.112 | 0.464 | -1.719 | -0.013 | 8.57E-02 | 18620 | 0.772 | 0.217 | 1.327 | 0.283 | 2.728 | 0.020 | 6.38E-03 | 18704 |
| 156 | ACC_sup_R | -0.077 | -1.020 | 0.866 | 0.481 | -0.160 | -0.001 | 8.73E-01 | 18620 | 0.426 | -0.149 | 1.002 | 0.294 | 1.453 | 0.011 | 1.46E-01 | 18704 |
| 157 | N_Acc_L | -0.268 | -0.542 | 0.007 | 0.140 | -1.911 | -0.014 | 5.60E-02 | 18620 | 0.266 | 0.099 | 0.433 | 0.085 | 3.120 | 0.023 | 1.81E-03 | 18704 |
| 158 | N_Acc_R | -0.214 | -0.451 | 0.024 | 0.121 | -1.766 | -0.013 | 7.75E-02 | 18620 | 0.261 | 0.117 | 0.406 | 0.074 | 3.545 | 0.026 | 3.94E-04 | 18704 |
| 159 | VTA_L | -0.012 | -0.022 | -0.002 | 0.005 | -2.431 | -0.018 | 1.51E-02 | 18620 | 0.008 | 0.002 | 0.014 | 0.003 | 2.706 | 0.020 | 6.82E-03 | 18704 |
| 160 | VTA_R | -0.014 | -0.022 | -0.005 | 0.004 | -3.070 | -0.022 | 2.14E-03 | 18620 | 0.008 | 0.003 | 0.013 | 0.003 | 2.999 | 0.022 | 2.71E-03 | 18704 |
| 161 | SN_pc_L | -0.025 | -0.041 | -0.010 | 0.008 | -3.190 | -0.023 | 1.42E-03 | 18620 | 0.013 | 0.003 | 0.022 | 0.005 | 2.623 | 0.019 | 8.73E-03 | 18704 |
| 162 | SN_pc_R | -0.033 | -0.054 | -0.013 | 0.010 | -3.204 | -0.023 | 1.36E-03 | 18620 | 0.020 | 0.008 | 0.033 | 0.006 | 3.190 | 0.023 | 1.43E-03 | 18704 |
| 163 | SN_pr_L | -0.021 | -0.042 | 0.000 | 0.011 | -1.970 | -0.014 | 4.88E-02 | 18620 | 0.011 | -0.001 | 0.024 | 0.007 | 1.741 | 0.013 | 8.17E-02 | 18704 |
| 164 | SN_pr_R | -0.026 | -0.050 | -0.003 | 0.012 | -2.179 | -0.016 | 2.93E-02 | 18620 | 0.016 | 0.002 | 0.030 | 0.007 | 2.181 | 0.016 | 2.92E-02 | 18704 |
| 165 | Red_N_L | -0.035 | -0.061 | -0.009 | 0.013 | -2.642 | -0.019 | 8.24E-03 | 18620 | 0.019 | 0.003 | 0.035 | 0.008 | 2.341 | 0.017 | 1.93E-02 | 18704 |
| 166 | Red_N_R | -0.050 | -0.078 | -0.021 | 0.014 | -3.442 | -0.025 | 5.78E-04 | 18620 | 0.023 | 0.005 | 0.040 | 0.009 | 2.565 | 0.019 | 1.03E-02 | 18704 |
| 167 | LC_L | -0.009 | -0.019 | 0.001 | 0.005 | -1.856 | -0.014 | 6.35E-02 | 18620 | -0.001 | -0.007 | 0.005 | 0.003 | -0.379 | -0.003 | 7.05E-01 | 18704 |
| 168 | LC_R | -0.005 | -0.016 | 0.005 | 0.005 | -0.968 | -0.007 | 3.33E-01 | 18620 | -0.003 | -0.009 | 0.004 | 0.003 | -0.866 | -0.006 | 3.87E-01 | 18704 |
| 169 | Raphe_D | -0.070 | -0.107 | -0.032 | 0.019 | -3.627 | -0.027 | 2.88E-04 | 18620 | 0.043 | 0.020 | 0.066 | 0.012 | 3.671 | 0.027 | 2.42E-04 | 18704 |
| 170 | Raphe_M | -0.011 | -0.022 | 0.001 | 0.006 | -1.818 | -0.013 | 6.91E-02 | 18620 | 0.003 | -0.004 | 0.010 | 0.004 | 0.840 | 0.006 | 4.01E-01 | 18704 |

| | | Tea intake | | | | | | | | Water intake | | | | | | | |
|---|---|---|---|---|---|---|---|---|---|---|---|---|---|---|---|---|---|
| **Number** | **AAL 3 region** | **beta** | **95%CI_l** | **95%CI_h** | **se** | **t** | **r** | **p** | **df.e** | **beta** | **95%CI_l** | **95%CI_h** | **se** | **t** | **r** | **p** | **df.e** |
| 0 | Total brain | -56.022 | -131.197 | 19.154 | 38.353 | -1.461 | -0.011 | 1.44E-01 | 18590 | -36.753 | -123.744 | 50.238 | 44.381 | -0.828 | -0.006 | 4.08E-01 | 18531 |
| 1 | Precentral_L | -1.083 | -3.729 | 1.564 | 1.350 | -0.802 | -0.006 | 4.23E-01 | 18590 | -1.344 | -4.409 | 1.722 | 1.564 | -0.859 | -0.006 | 3.90E-01 | 18531 |
| 2 | Precentral_R | -0.206 | -2.632 | 2.220 | 1.238 | -0.166 | -0.001 | 8.68E-01 | 18590 | -1.718 | -4.528 | 1.091 | 1.433 | -1.199 | -0.009 | 2.31E-01 | 18531 |
| 3 | Frontal_Sup_2_L | -0.375 | -3.169 | 2.418 | 1.425 | -0.263 | -0.002 | 7.92E-01 | 18590 | -2.760 | -5.999 | 0.479 | 1.652 | -1.670 | -0.012 | 9.48E-02 | 18531 |
| 4 | Frontal_Sup_2_R | -0.262 | -3.342 | 2.818 | 1.571 | -0.167 | -0.001 | 8.68E-01 | 18590 | -2.328 | -5.898 | 1.243 | 1.822 | -1.278 | -0.009 | 2.01E-01 | 18531 |
| 5 | Frontal_Mid_2_L | -1.750 | -4.684 | 1.184 | 1.497 | -1.169 | -0.009 | 2.42E-01 | 18590 | -0.076 | -3.478 | 3.325 | 1.736 | -0.044 | 0.000 | 9.65E-01 | 18531 |
| 6 | Frontal_Mid_2_R | -0.394 | -3.714 | 2.926 | 1.694 | -0.233 | -0.002 | 8.16E-01 | 18590 | -0.972 | -4.819 | 2.876 | 1.963 | -0.495 | -0.004 | 6.21E-01 | 18531 |
| 7 | Frontal_Inf_Oper_L | -0.297 | -1.164 | 0.570 | 0.442 | -0.671 | -0.005 | 5.02E-01 | 18590 | -0.940 | -1.944 | 0.064 | 0.512 | -1.835 | -0.013 | 6.66E-02 | 18531 |
| 8 | Frontal_Inf_Oper_R | -0.816 | -1.902 | 0.271 | 0.554 | -1.471 | -0.011 | 1.41E-01 | 18590 | -1.013 | -2.273 | 0.247 | 0.643 | -1.576 | -0.012 | 1.15E-01 | 18531 |
| 9 | Frontal_Inf_Tri_L | -1.185 | -3.028 | 0.658 | 0.940 | -1.260 | -0.009 | 2.08E-01 | 18590 | -1.062 | -3.196 | 1.073 | 1.089 | -0.975 | -0.007 | 3.30E-01 | 18531 |
| 10 | Frontal_Inf_Tri_R | -0.989 | -2.434 | 0.457 | 0.738 | -1.340 | -0.010 | 1.80E-01 | 18590 | -1.137 | -2.812 | 0.538 | 0.855 | -1.330 | -0.010 | 1.84E-01 | 18531 |
| 11 | Frontal_Inf_Orb_2_L | -0.209 | -0.761 | 0.342 | 0.281 | -0.745 | -0.005 | 4.56E-01 | 18590 | -0.218 | -0.856 | 0.420 | 0.326 | -0.669 | -0.005 | 5.03E-01 | 18531 |
| 12 | Frontal_Inf_Orb_2_R | -0.188 | -0.764 | 0.388 | 0.294 | -0.640 | -0.005 | 5.22E-01 | 18590 | -0.059 | -0.726 | 0.608 | 0.340 | -0.173 | -0.001 | 8.62E-01 | 18531 |
| 13 | Rolandic_Oper_L | -0.819 | -1.652 | 0.014 | 0.425 | -1.928 | -0.014 | 5.39E-02 | 18590 | 0.205 | -0.760 | 1.169 | 0.492 | 0.416 | 0.003 | 6.78E-01 | 18531 |
| 14 | Rolandic_Oper_R | 0.134 | -1.057 | 1.324 | 0.607 | 0.220 | 0.002 | 8.26E-01 | 18590 | -0.385 | -1.764 | 0.994 | 0.704 | -0.547 | -0.004 | 5.84E-01 | 18531 |
| 15 | Supp_Motor_Area_L | -0.839 | -2.818 | 1.140 | 1.010 | -0.831 | -0.006 | 4.06E-01 | 18590 | -1.603 | -3.897 | 0.692 | 1.170 | -1.369 | -0.010 | 1.71E-01 | 18531 |
| 16 | Supp_Motor_Area_R | -1.301 | -3.489 | 0.886 | 1.116 | -1.166 | -0.009 | 2.44E-01 | 18590 | -1.498 | -4.034 | 1.037 | 1.294 | -1.158 | -0.009 | 2.47E-01 | 18531 |
| 17 | Olfactory_L | -0.108 | -0.402 | 0.187 | 0.150 | -0.717 | -0.005 | 4.73E-01 | 18590 | -0.273 | -0.614 | 0.068 | 0.174 | -1.570 | -0.012 | 1.16E-01 | 18531 |
| 18 | Olfactory_R | -0.165 | -0.460 | 0.130 | 0.151 | -1.095 | -0.008 | 2.74E-01 | 18590 | -0.369 | -0.711 | -0.027 | 0.174 | -2.116 | -0.016 | 3.44E-02 | 18531 |
| 19 | Frontal_Sup_Medial_L | -2.239 | -4.094 | -0.384 | 0.947 | -2.366 | -0.017 | 1.80E-02 | 18590 | -1.787 | -3.937 | 0.363 | 1.097 | -1.629 | -0.012 | 1.03E-01 | 18531 |
| 20 | Frontal_Sup_Medial_R | -1.455 | -2.806 | -0.103 | 0.689 | -2.110 | -0.015 | 3.49E-02 | 18590 | -0.797 | -2.362 | 0.768 | 0.798 | -0.999 | -0.007 | 3.18E-01 | 18531 |
| 21 | Frontal_Med_Orb_L | -0.686 | -1.379 | 0.008 | 0.354 | -1.938 | -0.014 | 5.26E-02 | 18590 | -0.183 | -0.986 | 0.619 | 0.409 | -0.447 | -0.003 | 6.55E-01 | 18531 |
| 22 | Frontal_Med_Orb_R | -0.596 | -1.448 | 0.256 | 0.435 | -1.371 | -0.010 | 1.70E-01 | 18590 | -0.340 | -1.326 | 0.645 | 0.503 | -0.677 | -0.005 | 4.98E-01 | 18531 |

| | | | | | | | | | | | | | | | | | |
|---|---|---|---|---|---|---|---|---|---|---|---|---|---|---|---|---|---|
| 23 | Rectus_L | -0.593 | -1.311 | 0.124 | 0.366 | -1.621 | -0.012 | 1.05E-01 | 18590 | 0.025 | -0.805 | 0.856 | 0.424 | 0.059 | 0.000 | 9.53E-01 | 18531 |
| 24 | Rectus_R | -0.722 | -1.386 | -0.059 | 0.339 | -2.133 | -0.016 | 3.29E-02 | 18590 | -0.238 | -1.006 | 0.531 | 0.392 | -0.606 | -0.004 | 5.44E-01 | 18531 |
| 25 | OFCmed_L | -0.389 | -0.805 | 0.027 | 0.212 | -1.832 | -0.013 | 6.69E-02 | 18590 | -0.154 | -0.635 | 0.328 | 0.246 | -0.625 | -0.005 | 5.32E-01 | 18531 |
| 26 | OFCmed_R | -0.320 | -0.804 | 0.165 | 0.247 | -1.293 | -0.009 | 1.96E-01 | 18590 | -0.536 | -1.097 | 0.026 | 0.286 | -1.870 | -0.014 | 6.15E-02 | 18531 |
| 27 | OFCant_L | -0.212 | -0.692 | 0.269 | 0.245 | -0.863 | -0.006 | 3.88E-01 | 18590 | -0.549 | -1.106 | 0.008 | 0.284 | -1.932 | -0.014 | 5.33E-02 | 18531 |
| 28 | OFCant_R | 0.050 | -0.586 | 0.685 | 0.324 | 0.153 | 0.001 | 8.79E-01 | 18590 | -0.690 | -1.426 | 0.046 | 0.376 | -1.837 | -0.013 | 6.63E-02 | 18531 |
| 29 | OFCpost_L | -0.120 | -0.722 | 0.482 | 0.307 | -0.390 | -0.003 | 6.97E-01 | 18590 | -0.578 | -1.276 | 0.119 | 0.356 | -1.625 | -0.012 | 1.04E-01 | 18531 |
| 30 | OFCpost_R | -0.156 | -0.713 | 0.400 | 0.284 | -0.550 | -0.004 | 5.82E-01 | 18590 | -0.302 | -0.947 | 0.343 | 0.329 | -0.917 | -0.007 | 3.59E-01 | 18531 |
| 31 | OFClat_L | -0.071 | -0.274 | 0.132 | 0.104 | -0.684 | -0.005 | 4.94E-01 | 18590 | -0.235 | -0.471 | 0.000 | 0.120 | -1.959 | -0.014 | 5.01E-02 | 18531 |
| 32 | OFClat_R | -0.115 | -0.286 | 0.057 | 0.088 | -1.312 | -0.010 | 1.90E-01 | 18590 | -0.049 | -0.248 | 0.150 | 0.101 | -0.481 | -0.004 | 6.30E-01 | 18531 |
| 33 | Insula_L | -0.317 | -2.180 | 1.546 | 0.950 | -0.334 | -0.002 | 7.39E-01 | 18590 | -0.965 | -3.121 | 1.191 | 1.100 | -0.878 | -0.006 | 3.80E-01 | 18531 |
| 34 | Insula_R | -0.578 | -2.393 | 1.236 | 0.926 | -0.624 | -0.005 | 5.32E-01 | 18590 | -1.347 | -3.447 | 0.753 | 1.071 | -1.257 | -0.009 | 2.09E-01 | 18531 |
| 37 | Cingulate_Mid_L | -1.268 | -2.841 | 0.305 | 0.803 | -1.580 | -0.012 | 1.14E-01 | 18590 | -1.063 | -2.884 | 0.758 | 0.929 | -1.144 | -0.008 | 2.53E-01 | 18531 |
| 38 | Cingulate_Mid_R | -1.516 | -3.195 | 0.162 | 0.856 | -1.771 | -0.013 | 7.67E-02 | 18590 | -0.583 | -2.526 | 1.359 | 0.991 | -0.589 | -0.004 | 5.56E-01 | 18531 |
| 39 | Cingulate_Post_L | -0.074 | -0.556 | 0.407 | 0.245 | -0.303 | -0.002 | 7.62E-01 | 18590 | -0.314 | -0.871 | 0.244 | 0.284 | -1.104 | -0.008 | 2.70E-01 | 18531 |
| 40 | Cingulate_Post_R | -0.024 | -0.291 | 0.243 | 0.136 | -0.177 | -0.001 | 8.60E-01 | 18590 | -0.230 | -0.539 | 0.080 | 0.158 | -1.456 | -0.011 | 1.45E-01 | 18531 |
| 41 | Hippocampus_L | 0.135 | -0.643 | 0.913 | 0.397 | 0.339 | 0.002 | 7.34E-01 | 18590 | 0.071 | -0.829 | 0.971 | 0.459 | 0.155 | 0.001 | 8.77E-01 | 18531 |
| 42 | Hippocampus_R | 0.171 | -0.636 | 0.978 | 0.412 | 0.416 | 0.003 | 6.78E-01 | 18590 | 0.141 | -0.793 | 1.075 | 0.477 | 0.296 | 0.002 | 7.67E-01 | 18531 |
| 43 | ParaHippocampal_L | 0.413 | -0.278 | 1.104 | 0.353 | 1.172 | 0.009 | 2.41E-01 | 18590 | 0.030 | -0.769 | 0.829 | 0.408 | 0.074 | 0.001 | 9.41E-01 | 18531 |
| 44 | ParaHippocampal_R | 0.092 | -0.780 | 0.963 | 0.445 | 0.206 | 0.002 | 8.37E-01 | 18590 | 0.699 | -0.310 | 1.708 | 0.515 | 1.357 | 0.010 | 1.75E-01 | 18531 |
| 45 | Amygdala_L | 0.217 | -0.052 | 0.485 | 0.137 | 1.581 | 0.012 | 1.14E-01 | 18590 | -0.097 | -0.407 | 0.214 | 0.158 | -0.611 | -0.004 | 5.41E-01 | 18531 |
| 46 | Amygdala_R | 0.198 | -0.077 | 0.473 | 0.140 | 1.413 | 0.010 | 1.58E-01 | 18590 | -0.022 | -0.340 | 0.296 | 0.162 | -0.137 | -0.001 | 8.91E-01 | 18531 |
| 47 | Calcarine_L | -0.175 | -2.739 | 2.389 | 1.308 | -0.134 | -0.001 | 8.93E-01 | 18590 | -0.054 | -3.021 | 2.913 | 1.514 | -0.036 | 0.000 | 9.72E-01 | 18531 |
| 48 | Calcarine_R | -0.316 | -2.458 | 1.826 | 1.093 | -0.289 | -0.002 | 7.73E-01 | 18590 | 0.674 | -1.807 | 3.154 | 1.266 | 0.532 | 0.004 | 5.95E-01 | 18531 |
| 49 | Cuneus_L | -0.544 | -2.151 | 1.063 | 0.820 | -0.663 | -0.005 | 5.07E-01 | 18590 | 1.546 | -0.317 | 3.408 | 0.950 | 1.627 | 0.012 | 1.04E-01 | 18531 |
| 50 | Cuneus_R | -0.177 | -1.406 | 1.053 | 0.627 | -0.281 | -0.002 | 7.78E-01 | 18590 | -0.706 | -2.131 | 0.720 | 0.727 | -0.971 | -0.007 | 3.32E-01 | 18531 |
| 51 | Lingual_L | -0.087 | -1.627 | 1.454 | 0.786 | -0.110 | -0.001 | 9.12E-01 | 18590 | 0.155 | -1.632 | 1.942 | 0.912 | 0.170 | 0.001 | 8.65E-01 | 18531 |
| 52 | Lingual_R | -0.478 | -2.093 | 1.137 | 0.824 | -0.580 | -0.004 | 5.62E-01 | 18590 | 0.300 | -1.569 | 2.169 | 0.954 | 0.315 | 0.002 | 7.53E-01 | 18531 |
| 53 | Occipital_Sup_L | 0.207 | -0.851 | 1.266 | 0.540 | 0.384 | 0.003 | 7.01E-01 | 18590 | 0.351 | -0.876 | 1.579 | 0.626 | 0.561 | 0.004 | 5.75E-01 | 18531 |
| 54 | Occipital_Sup_R | 0.481 | -0.667 | 1.629 | 0.586 | 0.821 | 0.006 | 4.12E-01 | 18590 | -1.330 | -2.660 | 0.001 | 0.679 | -1.959 | -0.014 | 5.01E-02 | 18531 |
| 55 | Occipital_Mid_L | -1.069 | -3.551 | 1.414 | 1.266 | -0.844 | -0.006 | 3.99E-01 | 18590 | -2.021 | -4.893 | 0.852 | 1.465 | -1.379 | -0.010 | 1.68E-01 | 18531 |
| 56 | Occipital_Mid_R | -0.613 | -2.339 | 1.113 | 0.881 | -0.696 | -0.005 | 4.87E-01 | 18590 | -1.701 | -3.702 | 0.300 | 1.021 | -1.666 | -0.012 | 9.57E-02 | 18531 |
| 57 | Occipital_Inf_L | -0.905 | -1.729 | -0.081 | 0.420 | -2.152 | -0.016 | 3.14E-02 | 18590 | -0.878 | -1.832 | 0.076 | 0.487 | -1.803 | -0.013 | 7.13E-02 | 18531 |
| 58 | Occipital_Inf_R | -0.043 | -0.996 | 0.909 | 0.486 | -0.089 | -0.001 | 9.29E-01 | 18590 | -1.392 | -2.495 | -0.289 | 0.563 | -2.474 | -0.018 | 1.34E-02 | 18531 |
| 59 | Fusiform_L | -0.545 | -2.349 | 1.258 | 0.920 | -0.592 | -0.004 | 5.54E-01 | 18590 | -0.289 | -2.377 | 1.799 | 1.065 | -0.271 | -0.002 | 7.86E-01 | 18531 |
| 60 | Fusiform_R | -1.503 | -3.486 | 0.481 | 1.012 | -1.485 | -0.011 | 1.38E-01 | 18590 | 0.666 | -1.630 | 2.962 | 1.171 | 0.568 | 0.004 | 5.70E-01 | 18531 |
| 61 | Postcentral_L | -1.257 | -3.970 | 1.457 | 1.384 | -0.908 | -0.007 | 3.64E-01 | 18590 | 1.608 | -1.534 | 4.750 | 1.603 | 1.003 | 0.007 | 3.16E-01 | 18531 |
| 62 | Postcentral_R | -1.362 | -4.146 | 1.421 | 1.420 | -0.960 | -0.007 | 3.37E-01 | 18590 | 2.074 | -1.148 | 5.296 | 1.644 | 1.262 | 0.009 | 2.07E-01 | 18531 |
| 63 | Parietal_Sup_L | -1.711 | -3.274 | -0.147 | 0.798 | -2.144 | -0.016 | 3.20E-02 | 18590 | -0.075 | -1.886 | 1.736 | 0.924 | -0.081 | -0.001 | 9.35E-01 | 18531 |
| 64 | Parietal_Sup_R | -1.706 | -3.095 | -0.318 | 0.708 | -2.409 | -0.018 | 1.60E-02 | 18590 | 0.025 | -1.582 | 1.633 | 0.820 | 0.031 | 0.000 | 9.75E-01 | 18531 |
| 65 | Parietal_Inf_L | -1.998 | -4.045 | 0.050 | 1.045 | -1.912 | -0.014 | 5.58E-02 | 18590 | 0.245 | -2.126 | 2.616 | 1.210 | 0.203 | 0.001 | 8.39E-01 | 18531 |
| 66 | Parietal_Inf_R | -0.547 | -1.874 | 0.780 | 0.677 | -0.808 | -0.006 | 4.19E-01 | 18590 | -0.148 | -1.684 | 1.388 | 0.784 | -0.188 | -0.001 | 8.51E-01 | 18531 |
| 67 | SupraMarginal_L | -0.736 | -1.936 | 0.463 | 0.612 | -1.203 | -0.009 | 2.29E-01 | 18590 | -0.077 | -1.467 | 1.313 | 0.709 | -0.108 | -0.001 | 9.14E-01 | 18531 |
| 68 | SupraMarginal_R | -0.728 | -2.424 | 0.969 | 0.866 | -0.841 | -0.006 | 4.01E-01 | 18590 | 1.291 | -0.674 | 3.255 | 1.002 | 1.288 | 0.009 | 1.98E-01 | 18531 |
| 69 | Angular_L | -1.504 | -2.683 | -0.325 | 0.601 | -2.501 | -0.018 | 1.24E-02 | 18590 | 0.407 | -0.958 | 1.773 | 0.697 | 0.585 | 0.004 | 5.59E-01 | 18531 |
| 70 | Angular_R | -2.022 | -3.603 | -0.441 | 0.807 | -2.507 | -0.018 | 1.22E-02 | 18590 | -0.573 | -2.403 | 1.258 | 0.934 | -0.613 | -0.005 | 5.40E-01 | 18531 |
| 71 | Precuneus_L | -2.962 | -5.835 | -0.089 | 1.466 | -2.021 | -0.015 | 4.33E-02 | 18590 | -0.435 | -3.760 | 2.891 | 1.696 | -0.256 | -0.002 | 7.98E-01 | 18531 |
| 72 | Precuneus_R | -2.096 | -4.656 | 0.463 | 1.306 | -1.605 | -0.012 | 1.08E-01 | 18590 | -2.011 | -4.972 | 0.950 | 1.511 | -1.331 | -0.010 | 1.83E-01 | 18531 |
| 73 | Paracentral_Lobule_L | -0.922 | -2.237 | 0.393 | 0.671 | -1.374 | -0.010 | 1.69E-01 | 18590 | -0.395 | -1.918 | 1.129 | 0.777 | -0.508 | -0.004 | 6.12E-01 | 18531 |

| | | | | | | | | | | | | | | | | | |
|---|---|---|---|---|---|---|---|---|---|---|---|---|---|---|---|---|---|
| 74 | Paracentral_Lobule_R | -0.803 | -1.745 | 0.139 | 0.481 | -1.670 | -0.012 | 9.49E-02 | 18590 | 0.040 | -1.051 | 1.132 | 0.557 | 0.072 | 0.001 | 9.42E-01 | 18531 |
| 75 | Caudate_L | 1.132 | 0.081 | 2.183 | 0.536 | 2.110 | 0.015 | 3.48E-02 | 18590 | 0.890 | -0.327 | 2.107 | 0.621 | 1.433 | 0.011 | 1.52E-01 | 18531 |
| 76 | Caudate_R | 0.688 | -0.502 | 1.877 | 0.607 | 1.133 | 0.008 | 2.57E-01 | 18590 | 0.785 | -0.593 | 2.162 | 0.703 | 1.116 | 0.008 | 2.64E-01 | 18531 |
| 77 | Putamen_L | 0.343 | -1.252 | 1.938 | 0.814 | 0.421 | 0.003 | 6.73E-01 | 18590 | 0.302 | -1.546 | 2.150 | 0.943 | 0.321 | 0.002 | 7.49E-01 | 18531 |
| 78 | Putamen_R | 0.055 | -1.570 | 1.680 | 0.829 | 0.067 | 0.000 | 9.47E-01 | 18590 | 0.467 | -1.415 | 2.350 | 0.961 | 0.487 | 0.004 | 6.27E-01 | 18531 |
| 79 | Pallidum_L | -0.054 | -0.196 | 0.088 | 0.072 | -0.745 | -0.005 | 4.56E-01 | 18590 | -0.043 | -0.207 | 0.122 | 0.084 | -0.510 | -0.004 | 6.10E-01 | 18531 |
| 80 | Pallidum_R | -0.017 | -0.173 | 0.138 | 0.079 | -0.217 | -0.002 | 8.29E-01 | 18590 | -0.051 | -0.231 | 0.130 | 0.092 | -0.552 | -0.004 | 5.81E-01 | 18531 |
| 83 | Heschl_L | -0.137 | -0.424 | 0.151 | 0.147 | -0.931 | -0.007 | 3.52E-01 | 18590 | -0.018 | -0.351 | 0.315 | 0.170 | -0.105 | -0.001 | 9.16E-01 | 18531 |
| 84 | Heschl_R | 0.171 | -0.150 | 0.492 | 0.164 | 1.041 | 0.008 | 2.98E-01 | 18590 | -0.099 | -0.471 | 0.273 | 0.190 | -0.521 | -0.004 | 6.02E-01 | 18531 |
| 85 | Temporal_Sup_L | -0.036 | -2.113 | 2.040 | 1.059 | -0.034 | 0.000 | 9.73E-01 | 18590 | -0.755 | -3.161 | 1.652 | 1.228 | -0.615 | -0.005 | 5.39E-01 | 18531 |
| 86 | Temporal_Sup_R | 0.454 | -1.844 | 2.752 | 1.172 | 0.387 | 0.003 | 6.99E-01 | 18590 | -2.368 | -5.028 | 0.291 | 1.357 | -1.746 | -0.013 | 8.09E-02 | 18531 |
| 87 | Temporal_Pole_Sup_L | 0.019 | -0.966 | 1.003 | 0.502 | 0.037 | 0.000 | 9.70E-01 | 18590 | -0.744 | -1.881 | 0.394 | 0.580 | -1.281 | -0.009 | 2.00E-01 | 18531 |
| 88 | Temporal_Pole_Sup_R | 0.837 | -0.199 | 1.873 | 0.529 | 1.583 | 0.012 | 1.13E-01 | 18590 | -1.132 | -2.330 | 0.066 | 0.611 | -1.852 | -0.014 | 6.41E-02 | 18531 |
| 89 | Temporal_Mid_L | -1.568 | -5.030 | 1.894 | 1.766 | -0.888 | -0.007 | 3.75E-01 | 18590 | -2.979 | -6.984 | 1.027 | 2.044 | -1.458 | -0.011 | 1.45E-01 | 18531 |
| 90 | Temporal_Mid_R | -2.793 | -5.866 | 0.280 | 1.568 | -1.782 | -0.013 | 7.48E-02 | 18590 | -1.673 | -5.228 | 1.882 | 1.814 | -0.923 | -0.007 | 3.56E-01 | 18531 |
| 91 | Temporal_Pole_Mid_L | 0.180 | -0.646 | 1.006 | 0.422 | 0.427 | 0.003 | 6.69E-01 | 18590 | -0.195 | -1.151 | 0.762 | 0.488 | -0.399 | -0.003 | 6.90E-01 | 18531 |
| 92 | Temporal_Pole_Mid_R | 0.224 | -0.779 | 1.228 | 0.512 | 0.438 | 0.003 | 6.61E-01 | 18590 | -0.695 | -1.858 | 0.467 | 0.593 | -1.172 | -0.009 | 2.41E-01 | 18531 |
| 93 | Temporal_Inf_L | -1.976 | -4.356 | 0.403 | 1.214 | -1.628 | -0.012 | 1.03E-01 | 18590 | -1.093 | -3.847 | 1.661 | 1.405 | -0.778 | -0.006 | 4.37E-01 | 18531 |
| 94 | Temporal_Inf_R | -2.483 | -5.077 | 0.111 | 1.324 | -1.876 | -0.014 | 6.06E-02 | 18590 | 0.276 | -2.727 | 3.280 | 1.532 | 0.180 | 0.001 | 8.57E-01 | 18531 |
| 95 | Cerebellum_Crus1_L | -0.016 | -2.842 | 2.810 | 1.442 | -0.011 | 0.000 | 9.91E-01 | 18590 | 0.865 | -2.408 | 4.138 | 1.670 | 0.518 | 0.004 | 6.04E-01 | 18531 |
| 96 | Cerebellum_Crus1_R | -0.237 | -3.118 | 2.643 | 1.470 | -0.161 | -0.001 | 8.72E-01 | 18590 | -0.168 | -3.507 | 3.172 | 1.704 | -0.099 | -0.001 | 9.21E-01 | 18531 |
| 97 | Cerebellum_Crus2_L | 0.288 | -1.952 | 2.529 | 1.143 | 0.252 | 0.002 | 8.01E-01 | 18590 | 1.087 | -1.506 | 3.681 | 1.323 | 0.822 | 0.006 | 4.11E-01 | 18531 |
| 98 | Cerebellum_Crus2_R | -0.412 | -2.484 | 1.660 | 1.057 | -0.390 | -0.003 | 6.97E-01 | 18590 | 0.248 | -2.152 | 2.648 | 1.224 | 0.202 | 0.001 | 8.40E-01 | 18531 |
| 99 | Cerebellum_3_L | -0.051 | -0.201 | 0.099 | 0.077 | -0.665 | -0.005 | 5.06E-01 | 18590 | -0.009 | -0.183 | 0.165 | 0.089 | -0.101 | -0.001 | 9.20E-01 | 18531 |
| 100 | Cerebellum_3_R | -0.004 | -0.207 | 0.199 | 0.104 | -0.042 | 0.000 | 9.67E-01 | 18590 | -0.016 | -0.251 | 0.220 | 0.120 | -0.130 | -0.001 | 8.97E-01 | 18531 |
| 101 | Cerebellum_4_5_L | 0.092 | -1.124 | 1.309 | 0.621 | 0.149 | 0.001 | 8.82E-01 | 18590 | 0.431 | -0.978 | 1.841 | 0.719 | 0.600 | 0.004 | 5.49E-01 | 18531 |
| 102 | Cerebellum_4_5_R | -0.461 | -1.452 | 0.530 | 0.506 | -0.912 | -0.007 | 3.62E-01 | 18590 | 0.782 | -0.367 | 1.931 | 0.586 | 1.334 | 0.010 | 1.82E-01 | 18531 |
| 103 | Cerebellum_6_L | -0.340 | -2.441 | 1.762 | 1.072 | -0.317 | -0.002 | 7.51E-01 | 18590 | 1.019 | -1.415 | 3.453 | 1.242 | 0.821 | 0.006 | 4.12E-01 | 18531 |
| 104 | Cerebellum_6_R | -0.698 | -2.912 | 1.515 | 1.129 | -0.618 | -0.005 | 5.36E-01 | 18590 | 0.819 | -1.748 | 3.386 | 1.310 | 0.625 | 0.005 | 5.32E-01 | 18531 |
| 105 | Cerebellum_7b_L | -0.247 | -0.932 | 0.438 | 0.350 | -0.706 | -0.005 | 4.80E-01 | 18590 | 0.181 | -0.612 | 0.974 | 0.404 | 0.448 | 0.003 | 6.54E-01 | 18531 |
| 106 | Cerebellum_7b_R | -0.040 | -0.719 | 0.639 | 0.346 | -0.115 | -0.001 | 9.09E-01 | 18590 | 0.191 | -0.595 | 0.977 | 0.401 | 0.476 | 0.003 | 6.34E-01 | 18531 |
| 107 | Cerebellum_8_L | -1.283 | -3.458 | 0.891 | 1.109 | -1.157 | -0.008 | 2.47E-01 | 18590 | 0.676 | -1.841 | 3.194 | 1.284 | 0.527 | 0.004 | 5.98E-01 | 18531 |
| 108 | Cerebellum_8_R | -1.788 | -4.617 | 1.042 | 1.444 | -1.238 | -0.009 | 2.16E-01 | 18590 | 0.045 | -3.233 | 3.323 | 1.673 | 0.027 | 0.000 | 9.79E-01 | 18531 |
| 109 | Cerebellum_9_L | 0.561 | -0.669 | 1.791 | 0.627 | 0.894 | 0.007 | 3.71E-01 | 18590 | -0.165 | -1.589 | 1.259 | 0.727 | -0.227 | -0.002 | 8.21E-01 | 18531 |
| 110 | Cerebellum_9_R | 0.522 | -0.686 | 1.731 | 0.617 | 0.847 | 0.006 | 3.97E-01 | 18590 | -0.741 | -2.142 | 0.660 | 0.715 | -1.037 | -0.008 | 3.00E-01 | 18531 |
| 111 | Cerebellum_10_L | 0.033 | -0.055 | 0.120 | 0.045 | 0.731 | 0.005 | 4.65E-01 | 18590 | 0.077 | -0.024 | 0.178 | 0.052 | 1.497 | 0.011 | 1.35E-01 | 18531 |
| 112 | Cerebellum_10_R | -0.045 | -0.136 | 0.046 | 0.046 | -0.974 | -0.007 | 3.30E-01 | 18590 | 0.062 | -0.043 | 0.167 | 0.054 | 1.150 | 0.008 | 2.50E-01 | 18531 |
| 113 | Vermis_1_2 | -0.029 | -0.085 | 0.028 | 0.029 | -0.992 | -0.007 | 3.21E-01 | 18590 | -0.027 | -0.093 | 0.039 | 0.034 | -0.799 | -0.006 | 4.24E-01 | 18531 |
| 114 | Vermis_3 | -0.150 | -0.375 | 0.075 | 0.115 | -1.306 | -0.010 | 1.92E-01 | 18590 | -0.045 | -0.306 | 0.217 | 0.133 | -0.336 | -0.002 | 7.37E-01 | 18531 |
| 115 | Vermis_4_5 | -0.308 | -0.970 | 0.354 | 0.338 | -0.911 | -0.007 | 3.62E-01 | 18590 | 0.200 | -0.569 | 0.969 | 0.392 | 0.510 | 0.004 | 6.10E-01 | 18531 |
| 116 | Vermis_6 | -0.133 | -0.502 | 0.235 | 0.188 | -0.709 | -0.005 | 4.78E-01 | 18590 | 0.140 | -0.288 | 0.568 | 0.218 | 0.641 | 0.005 | 5.22E-01 | 18531 |
| 117 | Vermis_7 | -0.002 | -0.211 | 0.206 | 0.106 | -0.020 | 0.000 | 9.84E-01 | 18590 | 0.093 | -0.149 | 0.335 | 0.123 | 0.755 | 0.006 | 4.50E-01 | 18531 |
| 118 | Vermis_8 | -0.077 | -0.420 | 0.266 | 0.175 | -0.439 | -0.003 | 6.61E-01 | 18590 | 0.041 | -0.357 | 0.438 | 0.203 | 0.200 | 0.001 | 8.41E-01 | 18531 |
| 119 | Vermis_9 | 0.200 | -0.078 | 0.478 | 0.142 | 1.410 | 0.010 | 1.59E-01 | 18590 | -0.050 | -0.372 | 0.272 | 0.164 | -0.304 | -0.002 | 7.61E-01 | 18531 |
| 120 | Vermis_10 | 0.026 | -0.051 | 0.104 | 0.040 | 0.662 | 0.005 | 5.08E-01 | 18590 | -0.045 | -0.135 | 0.045 | 0.046 | -0.982 | -0.007 | 3.26E-01 | 18531 |
| 121 | Thal_AV_L | 0.010 | -0.021 | 0.041 | 0.016 | 0.623 | 0.005 | 5.33E-01 | 18590 | 0.011 | -0.025 | 0.047 | 0.018 | 0.618 | 0.005 | 5.37E-01 | 18531 |
| 122 | Thal_AV_R | 0.009 | -0.035 | 0.053 | 0.023 | 0.403 | 0.003 | 6.87E-01 | 18590 | 0.021 | -0.031 | 0.072 | 0.026 | 0.795 | 0.006 | 4.27E-01 | 18531 |
| 123 | Thal_LP_L | -0.006 | -0.040 | 0.029 | 0.018 | -0.330 | -0.002 | 7.42E-01 | 18590 | 0.016 | -0.024 | 0.056 | 0.020 | 0.793 | 0.006 | 4.28E-01 | 18531 |
| 124 | Thal_LP_R | 0.029 | -0.016 | 0.074 | 0.023 | 1.262 | 0.009 | 2.07E-01 | 18590 | 0.022 | -0.030 | 0.074 | 0.026 | 0.817 | 0.006 | 4.14E-01 | 18531 |

| | | | | | | | | | | | | | | | | | |
|---|---|---|---|---|---|---|---|---|---|---|---|---|---|---|---|---|---|
| 125 | Thal_VA_L | 0.004 | -0.032 | 0.041 | 0.019 | 0.231 | 0.002 | 8.17E-01 | 18590 | 0.024 | -0.019 | 0.066 | 0.022 | 1.085 | 0.008 | 2.78E-01 | 18531 |
| 126 | Thal_VA_R | -0.014 | -0.059 | 0.031 | 0.023 | -0.600 | -0.004 | 5.48E-01 | 18590 | 0.010 | -0.043 | 0.062 | 0.027 | 0.362 | 0.003 | 7.17E-01 | 18531 |
| 127 | Thal_VL_L | 0.023 | -0.081 | 0.127 | 0.053 | 0.431 | 0.003 | 6.66E-01 | 18590 | -0.049 | -0.170 | 0.072 | 0.062 | -0.798 | -0.006 | 4.25E-01 | 18531 |
| 128 | Thal_VL_R | 0.027 | -0.112 | 0.165 | 0.071 | 0.379 | 0.003 | 7.05E-01 | 18590 | -0.039 | -0.200 | 0.122 | 0.082 | -0.477 | -0.004 | 6.33E-01 | 18531 |
| 129 | Thal_VPL_L | 0.037 | -0.087 | 0.161 | 0.063 | 0.587 | 0.004 | 5.57E-01 | 18590 | -0.062 | -0.206 | 0.081 | 0.073 | -0.851 | -0.006 | 3.95E-01 | 18531 |
| 130 | Thal_VPL_R | 0.076 | -0.055 | 0.207 | 0.067 | 1.139 | 0.008 | 2.55E-01 | 18590 | 0.022 | -0.130 | 0.173 | 0.077 | 0.282 | 0.002 | 7.78E-01 | 18531 |
| 131 | Thal_IL_L | 0.029 | -0.025 | 0.083 | 0.027 | 1.050 | 0.008 | 2.94E-01 | 18590 | -0.008 | -0.070 | 0.055 | 0.032 | -0.244 | -0.002 | 8.07E-01 | 18531 |
| 132 | Thal_IL_R | 0.037 | -0.024 | 0.098 | 0.031 | 1.177 | 0.009 | 2.39E-01 | 18590 | 0.007 | -0.064 | 0.077 | 0.036 | 0.191 | 0.001 | 8.48E-01 | 18531 |
| 133 | Thal_Re_L | 0.000 | 0.000 | 0.000 | 0.000 | -0.801 | -0.006 | 4.23E-01 | 18590 | 0.000 | 0.000 | 0.000 | 0.000 | 1.464 | 0.011 | 1.43E-01 | 18531 |
| 134 | Thal_Re_R | -0.001 | -0.003 | 0.001 | 0.001 | -0.801 | -0.006 | 4.23E-01 | 18590 | 0.000 | -0.002 | 0.003 | 0.001 | 0.384 | 0.003 | 7.01E-01 | 18531 |
| 135 | Thal_MDm_L | 0.200 | -0.198 | 0.598 | 0.203 | 0.985 | 0.007 | 3.25E-01 | 18590 | -0.046 | -0.507 | 0.415 | 0.235 | -0.195 | -0.001 | 8.45E-01 | 18531 |
| 136 | Thal_MDm_R | 0.290 | -0.173 | 0.753 | 0.236 | 1.228 | 0.009 | 2.19E-01 | 18590 | -0.025 | -0.561 | 0.511 | 0.273 | -0.091 | -0.001 | 9.27E-01 | 18531 |
| 137 | Thal_MDl_L | 0.032 | -0.037 | 0.101 | 0.035 | 0.916 | 0.007 | 3.60E-01 | 18590 | -0.005 | -0.085 | 0.075 | 0.041 | -0.120 | -0.001 | 9.05E-01 | 18531 |
| 138 | Thal_MDl_R | 0.041 | -0.038 | 0.119 | 0.040 | 1.019 | 0.007 | 3.08E-01 | 18590 | -0.007 | -0.098 | 0.084 | 0.046 | -0.148 | -0.001 | 8.82E-01 | 18531 |
| 139 | Thal_LGN_L | -0.013 | -0.023 | -0.002 | 0.005 | -2.348 | -0.017 | 1.89E-02 | 18590 | -0.004 | -0.016 | 0.008 | 0.006 | -0.644 | -0.005 | 5.19E-01 | 18531 |
| 140 | Thal_LGN_R | -0.017 | -0.039 | 0.005 | 0.011 | -1.494 | -0.011 | 1.35E-01 | 18590 | -0.015 | -0.041 | 0.011 | 0.013 | -1.131 | -0.008 | 2.58E-01 | 18531 |
| 141 | Thal_MGN_L | -0.007 | -0.025 | 0.012 | 0.010 | -0.683 | -0.005 | 4.95E-01 | 18590 | -0.012 | -0.034 | 0.010 | 0.011 | -1.081 | -0.008 | 2.80E-01 | 18531 |
| 142 | Thal_MGN_R | -0.001 | -0.021 | 0.020 | 0.010 | -0.051 | 0.000 | 9.59E-01 | 18590 | -0.013 | -0.037 | 0.010 | 0.012 | -1.092 | -0.008 | 2.75E-01 | 18531 |
| 143 | Thal_PuA_L | -0.007 | -0.029 | 0.015 | 0.011 | -0.647 | -0.005 | 5.18E-01 | 18590 | -0.013 | -0.039 | 0.012 | 0.013 | -1.014 | -0.007 | 3.11E-01 | 18531 |
| 144 | Thal_PuA_R | 0.011 | -0.035 | 0.057 | 0.023 | 0.456 | 0.003 | 6.48E-01 | 18590 | -0.027 | -0.080 | 0.026 | 0.027 | -0.987 | -0.007 | 3.24E-01 | 18531 |
| 145 | Thal_PuM_L | 0.064 | -0.374 | 0.503 | 0.224 | 0.287 | 0.002 | 7.74E-01 | 18590 | -0.178 | -0.685 | 0.330 | 0.259 | -0.686 | -0.005 | 4.93E-01 | 18531 |
| 146 | Thal_PuM_R | 0.298 | -0.248 | 0.845 | 0.279 | 1.070 | 0.008 | 2.85E-01 | 18590 | -0.071 | -0.704 | 0.561 | 0.323 | -0.221 | -0.002 | 8.25E-01 | 18531 |
| 147 | Thal_PuL_L | 0.024 | -0.056 | 0.104 | 0.041 | 0.582 | 0.004 | 5.61E-01 | 18590 | -0.013 | -0.106 | 0.079 | 0.047 | -0.281 | -0.002 | 7.79E-01 | 18531 |
| 148 | Thal_PuL_R | 0.076 | -0.051 | 0.204 | 0.065 | 1.177 | 0.009 | 2.39E-01 | 18590 | 0.015 | -0.132 | 0.163 | 0.075 | 0.204 | 0.001 | 8.39E-01 | 18531 |
| 149 | Thal_PuI_L | -0.006 | -0.027 | 0.016 | 0.011 | -0.503 | -0.004 | 6.15E-01 | 18590 | -0.001 | -0.026 | 0.024 | 0.013 | -0.082 | -0.001 | 9.35E-01 | 18531 |
| 150 | Thal_PuI_R | 0.002 | -0.026 | 0.031 | 0.015 | 0.165 | 0.001 | 8.69E-01 | 18590 | 0.016 | -0.017 | 0.050 | 0.017 | 0.955 | 0.007 | 3.40E-01 | 18531 |
| 151 | ACC_sub_L | -0.019 | -0.270 | 0.232 | 0.128 | -0.150 | -0.001 | 8.81E-01 | 18590 | -0.227 | -0.517 | 0.064 | 0.148 | -1.529 | -0.011 | 1.26E-01 | 18531 |
| 152 | ACC_sub_R | -0.010 | -0.245 | 0.224 | 0.120 | -0.085 | -0.001 | 9.32E-01 | 18590 | -0.257 | -0.528 | 0.014 | 0.138 | -1.856 | -0.014 | 6.35E-02 | 18531 |
| 153 | ACC_pre_L | -0.401 | -1.044 | 0.242 | 0.328 | -1.223 | -0.009 | 2.21E-01 | 18590 | -0.722 | -1.466 | 0.022 | 0.380 | -1.902 | -0.014 | 5.72E-02 | 18531 |
| 154 | ACC_pre_R | -0.393 | -1.012 | 0.226 | 0.316 | -1.244 | -0.009 | 2.14E-01 | 18590 | -0.418 | -1.135 | 0.299 | 0.366 | -1.144 | -0.008 | 2.53E-01 | 18531 |
| 155 | ACC_sup_L | 0.074 | -0.625 | 0.773 | 0.357 | 0.208 | 0.002 | 8.35E-01 | 18590 | -0.465 | -1.275 | 0.344 | 0.413 | -1.127 | -0.008 | 2.60E-01 | 18531 |
| 156 | ACC_sup_R | 0.183 | -0.541 | 0.907 | 0.369 | 0.496 | 0.004 | 6.20E-01 | 18590 | -0.500 | -1.338 | 0.338 | 0.428 | -1.170 | -0.009 | 2.42E-01 | 18531 |
| 157 | N_Acc_L | 0.021 | -0.189 | 0.232 | 0.107 | 0.199 | 0.001 | 8.42E-01 | 18590 | 0.117 | -0.126 | 0.361 | 0.124 | 0.944 | 0.007 | 3.45E-01 | 18531 |
| 158 | N_Acc_R | -0.054 | -0.236 | 0.128 | 0.093 | -0.581 | -0.004 | 5.62E-01 | 18590 | 0.035 | -0.176 | 0.246 | 0.108 | 0.323 | 0.002 | 7.47E-01 | 18531 |
| 159 | VTA_L | 0.005 | -0.003 | 0.012 | 0.004 | 1.275 | 0.009 | 2.02E-01 | 18590 | -0.004 | -0.013 | 0.004 | 0.004 | -0.971 | -0.007 | 3.31E-01 | 18531 |
| 160 | VTA_R | 0.006 | 0.000 | 0.013 | 0.003 | 1.884 | 0.014 | 5.96E-02 | 18590 | -0.003 | -0.011 | 0.005 | 0.004 | -0.713 | -0.005 | 4.76E-01 | 18531 |
| 161 | SN_pc_L | 0.010 | -0.002 | 0.022 | 0.006 | 1.606 | 0.012 | 1.08E-01 | 18590 | -0.010 | -0.024 | 0.003 | 0.007 | -1.483 | -0.011 | 1.38E-01 | 18531 |
| 162 | SN_pc_R | 0.012 | -0.004 | 0.027 | 0.008 | 1.464 | 0.011 | 1.43E-01 | 18590 | -0.008 | -0.026 | 0.010 | 0.009 | -0.916 | -0.007 | 3.59E-01 | 18531 |
| 163 | SN_pr_L | 0.005 | -0.012 | 0.021 | 0.008 | 0.556 | 0.004 | 5.78E-01 | 18590 | -0.014 | -0.032 | 0.005 | 0.010 | -1.448 | -0.011 | 1.48E-01 | 18531 |
| 164 | SN_pr_R | 0.004 | -0.014 | 0.022 | 0.009 | 0.411 | 0.003 | 6.81E-01 | 18590 | -0.016 | -0.037 | 0.005 | 0.011 | -1.492 | -0.011 | 1.36E-01 | 18531 |
| 165 | Red_N_L | 0.000 | -0.020 | 0.020 | 0.010 | 0.031 | 0.000 | 9.75E-01 | 18590 | -0.018 | -0.041 | 0.005 | 0.012 | -1.558 | -0.011 | 1.19E-01 | 18531 |
| 166 | Red_N_R | -0.003 | -0.024 | 0.019 | 0.011 | -0.241 | -0.002 | 8.09E-01 | 18590 | -0.007 | -0.032 | 0.018 | 0.013 | -0.541 | -0.004 | 5.89E-01 | 18531 |
| 167 | LC_L | 0.001 | -0.007 | 0.008 | 0.004 | 0.209 | 0.002 | 8.34E-01 | 18590 | -0.007 | -0.016 | 0.002 | 0.004 | -1.609 | -0.012 | 1.08E-01 | 18531 |
| 168 | LC_R | 0.002 | -0.007 | 0.010 | 0.004 | 0.368 | 0.003 | 7.13E-01 | 18590 | -0.008 | -0.017 | 0.001 | 0.005 | -1.693 | -0.012 | 9.04E-02 | 18531 |
| 169 | Raphe_D | -0.003 | -0.031 | 0.026 | 0.015 | -0.176 | -0.001 | 8.61E-01 | 18590 | -0.017 | -0.050 | 0.016 | 0.017 | -0.994 | -0.007 | 3.20E-01 | 18531 |
| 170 | Raphe_M | -0.002 | -0.011 | 0.006 | 0.004 | -0.544 | -0.004 | 5.87E-01 | 18590 | -0.006 | -0.016 | 0.005 | 0.005 | -1.086 | -0.008 | 2.77E-01 | 18531 |
| | | **Cooked vegetable intake** | | | | | | | | **Processed meat intake** | | | | | | | |
| **Number** | **AAL 3 region** | **beta** | **95%CI_l** | **95%CI_h** | **se** | **t** | **r** | **p** | **df.e** | **beta** | **95%CI_l** | **95%CI_h** | **se** | **t** | **r** | **p** | **df.e** |
| 0 | Total brain | -122.865 | -235.993 | -9.737 | 57.716 | -2.129 | -0.016 | 3.33E-02 | 18391 | -59.112 | -216.010 | 97.786 | 80.046 | -0.738 | -0.005 | 4.60E-01 | 18724 |

| | | | | | | | | | | | | | | | | | |
|---|---|---|---|---|---|---|---|---|---|---|---|---|---|---|---|---|---|
| 1 | Precentral_L | -2.191 | -6.173 | 1.790 | 2.031 | -1.079 | -0.008 | 2.81E-01 | 18391 | 0.714 | -4.815 | 6.243 | 2.821 | 0.253 | 0.002 | 8.00E-01 | 18724 |
| 2 | Precentral_R | -0.792 | -4.437 | 2.852 | 1.859 | -0.426 | -0.003 | 6.70E-01 | 18391 | -0.571 | -5.638 | 4.497 | 2.585 | -0.221 | -0.002 | 8.25E-01 | 18724 |
| 3 | Frontal_Sup_2_L | -4.682 | -8.887 | -0.478 | 2.145 | -2.183 | -0.016 | 2.91E-02 | 18391 | 2.874 | -2.961 | 8.710 | 2.977 | 0.965 | 0.007 | 3.34E-01 | 18724 |
| 4 | Frontal_Sup_2_R | -3.170 | -7.805 | 1.465 | 2.365 | -1.340 | -0.010 | 1.80E-01 | 18391 | 1.705 | -4.724 | 8.135 | 3.280 | 0.520 | 0.004 | 6.03E-01 | 18724 |
| 5 | Frontal_Mid_2_L | -1.044 | -5.459 | 3.370 | 2.252 | -0.464 | -0.003 | 6.43E-01 | 18391 | 5.394 | -0.737 | 11.524 | 3.128 | 1.724 | 0.013 | 8.46E-02 | 18724 |
| 6 | Frontal_Mid_2_R | -1.555 | -6.545 | 3.435 | 2.546 | -0.611 | -0.005 | 5.41E-01 | 18391 | 2.646 | -4.290 | 9.581 | 3.538 | 0.748 | 0.005 | 4.55E-01 | 18724 |
| 7 | Frontal_Inf_Oper_L | -0.349 | -1.652 | 0.954 | 0.665 | -0.525 | -0.004 | 5.99E-01 | 18391 | 0.754 | -1.057 | 2.565 | 0.924 | 0.816 | 0.006 | 4.14E-01 | 18724 |
| 8 | Frontal_Inf_Oper_R | -0.829 | -2.464 | 0.807 | 0.834 | -0.993 | -0.007 | 3.21E-01 | 18391 | 1.756 | -0.514 | 4.027 | 1.158 | 1.516 | 0.011 | 1.29E-01 | 18724 |
| 9 | Frontal_Inf_Tri_L | -1.795 | -4.562 | 0.972 | 1.412 | -1.272 | -0.009 | 2.03E-01 | 18391 | 2.561 | -1.284 | 6.406 | 1.962 | 1.305 | 0.010 | 1.92E-01 | 18724 |
| 10 | Frontal_Inf_Tri_R | -0.577 | -2.750 | 1.597 | 1.109 | -0.520 | -0.004 | 6.03E-01 | 18391 | 1.145 | -1.875 | 4.165 | 1.541 | 0.743 | 0.005 | 4.57E-01 | 18724 |
| 11 | Frontal_Inf_Orb_2_L | -1.154 | -1.983 | -0.326 | 0.423 | -2.732 | -0.020 | 6.31E-03 | 18391 | 1.287 | 0.137 | 2.437 | 0.587 | 2.194 | 0.016 | 2.82E-02 | 18724 |
| 12 | Frontal_Inf_Orb_2_R | -1.106 | -1.971 | -0.240 | 0.442 | -2.503 | -0.018 | 1.23E-02 | 18391 | 0.559 | -0.644 | 1.761 | 0.614 | 0.910 | 0.007 | 3.63E-01 | 18724 |
| 13 | Rolandic_Oper_L | -1.682 | -2.935 | -0.430 | 0.639 | -2.632 | -0.019 | 8.49E-03 | 18391 | 0.625 | -1.114 | 2.364 | 0.887 | 0.705 | 0.005 | 4.81E-01 | 18724 |
| 14 | Rolandic_Oper_R | -3.284 | -5.076 | -1.492 | 0.914 | -3.593 | -0.026 | 3.28E-04 | 18391 | 1.013 | -1.476 | 3.502 | 1.270 | 0.798 | 0.006 | 4.25E-01 | 18724 |
| 15 | Supp_Motor_Area_L | -1.899 | -4.874 | 1.076 | 1.518 | -1.251 | -0.009 | 2.11E-01 | 18391 | 0.526 | -3.609 | 4.661 | 2.110 | 0.249 | 0.002 | 8.03E-01 | 18724 |
| 16 | Supp_Motor_Area_R | -2.111 | -5.400 | 1.177 | 1.678 | -1.258 | -0.009 | 2.08E-01 | 18391 | -0.050 | -4.619 | 4.519 | 2.331 | -0.021 | 0.000 | 9.83E-01 | 18724 |
| 17 | Olfactory_L | -0.401 | -0.845 | 0.042 | 0.226 | -1.775 | -0.013 | 7.59E-02 | 18391 | 0.438 | -0.176 | 1.052 | 0.313 | 1.399 | 0.010 | 1.62E-01 | 18724 |
| 18 | Olfactory_R | -0.328 | -0.773 | 0.116 | 0.227 | -1.448 | -0.011 | 1.48E-01 | 18391 | 0.442 | -0.174 | 1.058 | 0.314 | 1.405 | 0.010 | 1.60E-01 | 18724 |
| 19 | Frontal_Sup_Medial_L | -2.608 | -5.398 | 0.183 | 1.424 | -1.832 | -0.014 | 6.70E-02 | 18391 | 1.401 | -2.471 | 5.273 | 1.975 | 0.709 | 0.005 | 4.78E-01 | 18724 |
| 20 | Frontal_Sup_Medial_R | -2.111 | -4.144 | -0.078 | 1.037 | -2.035 | -0.015 | 4.18E-02 | 18391 | 1.169 | -1.650 | 3.988 | 1.438 | 0.813 | 0.006 | 4.16E-01 | 18724 |
| 21 | Frontal_Med_Orb_L | -1.074 | -2.117 | -0.031 | 0.532 | -2.019 | -0.015 | 4.35E-02 | 18391 | 1.195 | -0.252 | 2.642 | 0.738 | 1.618 | 0.012 | 1.06E-01 | 18724 |
| 22 | Frontal_Med_Orb_R | -1.027 | -2.309 | 0.255 | 0.654 | -1.570 | -0.012 | 1.16E-01 | 18391 | 1.451 | -0.328 | 3.229 | 0.907 | 1.599 | 0.012 | 1.10E-01 | 18724 |
| 23 | Rectus_L | -1.187 | -2.266 | -0.108 | 0.551 | -2.156 | -0.016 | 3.11E-02 | 18391 | 0.477 | -1.020 | 1.974 | 0.764 | 0.624 | 0.005 | 5.32E-01 | 18724 |
| 24 | Rectus_R | -0.863 | -1.861 | 0.135 | 0.509 | -1.695 | -0.012 | 9.01E-02 | 18391 | 0.717 | -0.668 | 2.103 | 0.707 | 1.015 | 0.007 | 3.10E-01 | 18724 |
| 25 | OFCmed_L | -1.170 | -1.795 | -0.544 | 0.319 | -3.663 | -0.027 | 2.50E-04 | 18391 | 0.096 | -0.772 | 0.964 | 0.443 | 0.217 | 0.002 | 8.28E-01 | 18724 |
| 26 | OFCmed_R | -0.547 | -1.276 | 0.181 | 0.372 | -1.472 | -0.011 | 1.41E-01 | 18391 | 0.758 | -0.253 | 1.770 | 0.516 | 1.469 | 0.011 | 1.42E-01 | 18724 |
| 27 | OFCant_L | -1.011 | -1.734 | -0.287 | 0.369 | -2.736 | -0.020 | 6.22E-03 | 18391 | 0.290 | -0.715 | 1.294 | 0.512 | 0.566 | 0.004 | 5.72E-01 | 18724 |
| 28 | OFCant_R | -0.749 | -1.705 | 0.208 | 0.488 | -1.534 | -0.011 | 1.25E-01 | 18391 | 0.704 | -0.622 | 2.031 | 0.677 | 1.041 | 0.008 | 2.98E-01 | 18724 |
| 29 | OFCpost_L | -1.504 | -2.410 | -0.597 | 0.462 | -3.252 | -0.024 | 1.15E-03 | 18391 | 0.842 | -0.416 | 2.100 | 0.642 | 1.312 | 0.010 | 1.90E-01 | 18724 |
| 30 | OFCpost_R | -1.423 | -2.261 | -0.585 | 0.427 | -3.329 | -0.025 | 8.74E-04 | 18391 | 0.362 | -0.800 | 1.525 | 0.593 | 0.611 | 0.004 | 5.41E-01 | 18724 |
| 31 | OFClat_L | -0.350 | -0.656 | -0.045 | 0.156 | -2.246 | -0.017 | 2.47E-02 | 18391 | 0.106 | -0.318 | 0.531 | 0.216 | 0.492 | 0.004 | 6.23E-01 | 18724 |
| 32 | OFClat_R | -0.469 | -0.728 | -0.211 | 0.132 | -3.562 | -0.026 | 3.69E-04 | 18391 | 0.076 | -0.282 | 0.434 | 0.183 | 0.416 | 0.003 | 6.77E-01 | 18724 |
| 33 | Insula_L | -2.600 | -5.401 | 0.202 | 1.429 | -1.819 | -0.013 | 6.89E-02 | 18391 | 1.430 | -2.458 | 5.318 | 1.984 | 0.721 | 0.005 | 4.71E-01 | 18724 |
| 34 | Insula_R | -3.187 | -5.917 | -0.458 | 1.392 | -2.289 | -0.017 | 2.21E-02 | 18391 | 1.405 | -2.383 | 5.193 | 1.933 | 0.727 | 0.005 | 4.67E-01 | 18724 |
| 37 | Cingulate_Mid_L | -2.248 | -4.615 | 0.118 | 1.207 | -1.862 | -0.014 | 6.26E-02 | 18391 | -2.356 | -5.639 | 0.928 | 1.675 | -1.406 | -0.010 | 1.60E-01 | 18724 |
| 38 | Cingulate_Mid_R | -1.621 | -4.144 | 0.902 | 1.287 | -1.259 | -0.009 | 2.08E-01 | 18391 | -0.979 | -4.480 | 2.521 | 1.786 | -0.548 | -0.004 | 5.84E-01 | 18724 |
| 39 | Cingulate_Post_L | -0.713 | -1.436 | 0.010 | 0.369 | -1.932 | -0.014 | 5.34E-02 | 18391 | -1.403 | -2.407 | -0.398 | 0.512 | -2.737 | -0.020 | 6.20E-03 | 18724 |
| 40 | Cingulate_Post_R | -0.420 | -0.821 | -0.018 | 0.205 | -2.050 | -0.015 | 4.04E-02 | 18391 | -0.557 | -1.115 | 0.000 | 0.284 | -1.961 | -0.014 | 4.99E-02 | 18724 |
| 41 | Hippocampus_L | -0.628 | -1.797 | 0.541 | 0.596 | -1.053 | -0.008 | 2.92E-01 | 18391 | 0.615 | -1.010 | 2.240 | 0.829 | 0.742 | 0.005 | 4.58E-01 | 18724 |
| 42 | Hippocampus_R | -0.568 | -1.781 | 0.645 | 0.619 | -0.918 | -0.007 | 3.59E-01 | 18391 | 1.103 | -0.582 | 2.788 | 0.860 | 1.283 | 0.009 | 1.99E-01 | 18724 |
| 43 | ParaHippocampal_L | -0.840 | -1.879 | 0.199 | 0.530 | -1.585 | -0.012 | 1.13E-01 | 18391 | -0.816 | -2.259 | 0.628 | 0.737 | -1.107 | -0.008 | 2.68E-01 | 18724 |
| 44 | ParaHippocampal_R | -0.228 | -1.539 | 1.084 | 0.669 | -0.340 | -0.003 | 7.34E-01 | 18391 | -1.039 | -2.860 | 0.783 | 0.929 | -1.117 | -0.008 | 2.64E-01 | 18724 |
| 45 | Amygdala_L | -0.373 | -0.777 | 0.031 | 0.206 | -1.811 | -0.013 | 7.02E-02 | 18391 | 0.366 | -0.195 | 0.927 | 0.286 | 1.277 | 0.009 | 2.01E-01 | 18724 |
| 46 | Amygdala_R | -0.261 | -0.674 | 0.152 | 0.211 | -1.240 | -0.009 | 2.15E-01 | 18391 | 0.233 | -0.341 | 0.807 | 0.293 | 0.796 | 0.006 | 4.26E-01 | 18724 |
| 47 | Calcarine_L | -0.486 | -4.341 | 3.369 | 1.967 | -0.247 | -0.002 | 8.05E-01 | 18391 | -3.325 | -8.680 | 2.029 | 2.732 | -1.217 | -0.009 | 2.23E-01 | 18724 |
| 48 | Calcarine_R | -0.357 | -3.580 | 2.865 | 1.644 | -0.217 | -0.002 | 8.28E-01 | 18391 | -1.432 | -5.906 | 3.042 | 2.283 | -0.627 | -0.005 | 5.31E-01 | 18724 |
| 49 | Cuneus_L | -0.817 | -3.235 | 1.601 | 1.233 | -0.662 | -0.005 | 5.08E-01 | 18391 | -3.562 | -6.920 | -0.204 | 1.713 | -2.079 | -0.015 | 3.76E-02 | 18724 |
| 50 | Cuneus_R | -0.284 | -2.135 | 1.566 | 0.944 | -0.301 | -0.002 | 7.63E-01 | 18391 | -1.931 | -4.500 | 0.638 | 1.311 | -1.473 | -0.011 | 1.41E-01 | 18724 |
| 51 | Lingual_L | -1.150 | -3.469 | 1.169 | 1.183 | -0.972 | -0.007 | 3.31E-01 | 18391 | -1.496 | -4.714 | 1.722 | 1.642 | -0.911 | -0.007 | 3.62E-01 | 18724 |

| | | | | | | | | | | | | | | | | | |
|---|---|---|---|---|---|---|---|---|---|---|---|---|---|---|---|---|---|
| 52 | Lingual_R | -1.752 | -4.185 | 0.680 | 1.241 | -1.412 | -0.010 | 1.58E-01 | 18391 | -2.135 | -5.509 | 1.238 | 1.721 | -1.241 | -0.009 | 2.15E-01 | 18724 |
| 53 | Occipital_Sup_L | -0.885 | -2.481 | 0.710 | 0.814 | -1.088 | -0.008 | 2.77E-01 | 18391 | -3.326 | -5.537 | -1.114 | 1.128 | -2.948 | -0.022 | 3.20E-03 | 18724 |
| 54 | Occipital_Sup_R | -1.372 | -3.101 | 0.358 | 0.882 | -1.555 | -0.011 | 1.20E-01 | 18391 | -1.129 | -3.527 | 1.270 | 1.224 | -0.922 | -0.007 | 3.56E-01 | 18724 |
| 55 | Occipital_Mid_L | -1.303 | -5.036 | 2.431 | 1.905 | -0.684 | -0.005 | 4.94E-01 | 18391 | -1.931 | -7.119 | 3.256 | 2.647 | -0.730 | -0.005 | 4.66E-01 | 18724 |
| 56 | Occipital_Mid_R | -0.962 | -3.560 | 1.636 | 1.325 | -0.726 | -0.005 | 4.68E-01 | 18391 | -0.495 | -4.105 | 3.116 | 1.842 | -0.269 | -0.002 | 7.88E-01 | 18724 |
| 57 | Occipital_Inf_L | -1.132 | -2.372 | 0.109 | 0.633 | -1.788 | -0.013 | 7.37E-02 | 18391 | -0.070 | -1.793 | 1.653 | 0.879 | -0.080 | -0.001 | 9.36E-01 | 18724 |
| 58 | Occipital_Inf_R | -1.864 | -3.298 | -0.431 | 0.731 | -2.549 | -0.019 | 1.08E-02 | 18391 | 1.322 | -0.667 | 3.311 | 1.015 | 1.303 | 0.010 | 1.93E-01 | 18724 |
| 59 | Fusiform_L | -0.494 | -3.209 | 2.220 | 1.385 | -0.357 | -0.003 | 7.21E-01 | 18391 | -5.167 | -8.936 | -1.399 | 1.923 | -2.688 | -0.020 | 7.20E-03 | 18724 |
| 60 | Fusiform_R | -2.464 | -5.447 | 0.519 | 1.522 | -1.619 | -0.012 | 1.05E-01 | 18391 | -3.991 | -8.134 | 0.153 | 2.114 | -1.888 | -0.014 | 5.91E-02 | 18724 |
| 61 | Postcentral_L | -4.066 | -8.149 | 0.018 | 2.083 | -1.951 | -0.014 | 5.10E-02 | 18391 | -0.821 | -6.484 | 4.842 | 2.889 | -0.284 | -0.002 | 7.76E-01 | 18724 |
| 62 | Postcentral_R | -3.282 | -7.470 | 0.905 | 2.136 | -1.537 | -0.011 | 1.24E-01 | 18391 | -5.196 | -11.002 | 0.611 | 2.962 | -1.754 | -0.013 | 7.95E-02 | 18724 |
| 63 | Parietal_Sup_L | -0.988 | -3.340 | 1.364 | 1.200 | -0.823 | -0.006 | 4.10E-01 | 18391 | -2.126 | -5.392 | 1.139 | 1.666 | -1.276 | -0.009 | 2.02E-01 | 18724 |
| 64 | Parietal_Sup_R | 0.111 | -1.977 | 2.199 | 1.065 | 0.104 | 0.001 | 9.17E-01 | 18391 | -1.941 | -4.839 | 0.956 | 1.478 | -1.314 | -0.010 | 1.89E-01 | 18724 |
| 65 | Parietal_Inf_L | -1.426 | -4.508 | 1.655 | 1.572 | -0.907 | -0.007 | 3.64E-01 | 18391 | -4.430 | -8.702 | -0.158 | 2.179 | -2.033 | -0.015 | 4.21E-02 | 18724 |
| 66 | Parietal_Inf_R | -0.226 | -2.218 | 1.766 | 1.017 | -0.222 | -0.002 | 8.24E-01 | 18391 | -1.211 | -3.980 | 1.559 | 1.413 | -0.857 | -0.006 | 3.91E-01 | 18724 |
| 67 | SupraMarginal_L | -2.210 | -4.015 | -0.404 | 0.921 | -2.399 | -0.018 | 1.65E-02 | 18391 | -0.495 | -3.000 | 2.009 | 1.278 | -0.388 | -0.003 | 6.98E-01 | 18724 |
| 68 | SupraMarginal_R | -1.809 | -4.364 | 0.745 | 1.303 | -1.388 | -0.010 | 1.65E-01 | 18391 | -0.275 | -3.819 | 3.269 | 1.808 | -0.152 | -0.001 | 8.79E-01 | 18724 |
| 69 | Angular_L | -0.448 | -2.221 | 1.325 | 0.905 | -0.495 | -0.004 | 6.21E-01 | 18391 | -2.340 | -4.802 | 0.121 | 1.256 | -1.863 | -0.014 | 6.24E-02 | 18724 |
| 70 | Angular_R | -0.641 | -3.020 | 1.739 | 1.214 | -0.528 | -0.004 | 5.98E-01 | 18391 | -2.207 | -5.508 | 1.095 | 1.684 | -1.310 | -0.010 | 1.90E-01 | 18724 |
| 71 | Precuneus_L | 0.457 | -3.868 | 4.781 | 2.206 | 0.207 | 0.002 | 8.36E-01 | 18391 | -12.310 | -18.306 | -6.315 | 3.059 | -4.025 | -0.029 | 5.73E-05 | 18724 |
| 72 | Precuneus_R | 0.269 | -3.585 | 4.123 | 1.966 | 0.137 | 0.001 | 8.91E-01 | 18391 | -6.566 | -11.909 | -1.222 | 2.726 | -2.408 | -0.018 | 1.60E-02 | 18724 |
| 73 | Paracentral_Lobule_L | -0.862 | -2.839 | 1.115 | 1.009 | -0.854 | -0.006 | 3.93E-01 | 18391 | -1.035 | -3.783 | 1.713 | 1.402 | -0.738 | -0.005 | 4.60E-01 | 18724 |
| 74 | Paracentral_Lobule_R | -1.191 | -2.606 | 0.225 | 0.722 | -1.649 | -0.012 | 9.92E-02 | 18391 | -1.174 | -3.141 | 0.794 | 1.004 | -1.169 | -0.009 | 2.42E-01 | 18724 |
| 75 | Caudate_L | -0.538 | -2.117 | 1.041 | 0.806 | -0.668 | -0.005 | 5.04E-01 | 18391 | 0.621 | -1.573 | 2.816 | 1.120 | 0.555 | 0.004 | 5.79E-01 | 18724 |
| 76 | Caudate_R | -0.416 | -2.204 | 1.372 | 0.912 | -0.456 | -0.003 | 6.48E-01 | 18391 | 1.137 | -1.348 | 3.621 | 1.268 | 0.897 | 0.007 | 3.70E-01 | 18724 |
| 77 | Putamen_L | 0.377 | -2.021 | 2.776 | 1.224 | 0.308 | 0.002 | 7.58E-01 | 18391 | 0.449 | -2.884 | 3.782 | 1.700 | 0.264 | 0.002 | 7.92E-01 | 18724 |
| 78 | Putamen_R | -0.005 | -2.448 | 2.438 | 1.246 | -0.004 | 0.000 | 9.97E-01 | 18391 | 0.264 | -3.130 | 3.658 | 1.731 | 0.152 | 0.001 | 8.79E-01 | 18724 |
| 79 | Pallidum_L | 0.015 | -0.198 | 0.228 | 0.109 | 0.138 | 0.001 | 8.90E-01 | 18391 | 0.062 | -0.234 | 0.358 | 0.151 | 0.409 | 0.003 | 6.82E-01 | 18724 |
| 80 | Pallidum_R | -0.096 | -0.331 | 0.138 | 0.119 | -0.808 | -0.006 | 4.19E-01 | 18391 | 0.074 | -0.252 | 0.399 | 0.166 | 0.444 | 0.003 | 6.57E-01 | 18724 |
| 83 | Heschl_L | -0.326 | -0.759 | 0.106 | 0.221 | -1.479 | -0.011 | 1.39E-01 | 18391 | -0.033 | -0.634 | 0.567 | 0.306 | -0.109 | -0.001 | 9.13E-01 | 18724 |
| 84 | Heschl_R | -0.636 | -1.120 | -0.153 | 0.247 | -2.579 | -0.019 | 9.92E-03 | 18391 | 0.140 | -0.530 | 0.811 | 0.342 | 0.410 | 0.003 | 6.82E-01 | 18724 |
| 85 | Temporal_Sup_L | -3.943 | -7.068 | -0.817 | 1.595 | -2.472 | -0.018 | 1.34E-02 | 18391 | 1.193 | -3.141 | 5.527 | 2.211 | 0.540 | 0.004 | 5.90E-01 | 18724 |
| 86 | Temporal_Sup_R | -4.955 | -8.412 | -1.498 | 1.764 | -2.809 | -0.021 | 4.97E-03 | 18391 | 4.280 | -0.514 | 9.074 | 2.446 | 1.750 | 0.013 | 8.02E-02 | 18724 |
| 87 | Temporal_Pole_Sup_L | -0.685 | -2.166 | 0.796 | 0.755 | -0.907 | -0.007 | 3.64E-01 | 18391 | 0.358 | -1.698 | 2.413 | 1.049 | 0.341 | 0.002 | 7.33E-01 | 18724 |
| 88 | Temporal_Pole_Sup_R | -0.714 | -2.272 | 0.843 | 0.795 | -0.899 | -0.007 | 3.69E-01 | 18391 | 0.341 | -1.823 | 2.504 | 1.104 | 0.309 | 0.002 | 7.58E-01 | 18724 |
| 89 | Temporal_Mid_L | -5.940 | -11.147 | -0.732 | 2.657 | -2.236 | -0.016 | 2.54E-02 | 18391 | 0.454 | -6.774 | 7.682 | 3.688 | 0.123 | 0.001 | 9.02E-01 | 18724 |
| 90 | Temporal_Mid_R | -2.201 | -6.824 | 2.422 | 2.359 | -0.933 | -0.007 | 3.51E-01 | 18391 | -1.099 | -7.519 | 5.321 | 3.275 | -0.335 | -0.002 | 7.37E-01 | 18724 |
| 91 | Temporal_Pole_Mid_L | 0.130 | -1.114 | 1.374 | 0.635 | 0.205 | 0.002 | 8.38E-01 | 18391 | -1.534 | -3.262 | 0.193 | 0.881 | -1.741 | -0.013 | 8.17E-02 | 18724 |
| 92 | Temporal_Pole_Mid_R | -0.477 | -1.988 | 1.033 | 0.770 | -0.620 | -0.005 | 5.36E-01 | 18391 | 0.044 | -2.056 | 2.143 | 1.071 | 0.041 | 0.000 | 9.68E-01 | 18724 |
| 93 | Temporal_Inf_L | -4.624 | -8.204 | -1.043 | 1.827 | -2.531 | -0.019 | 1.14E-02 | 18391 | -0.692 | -5.664 | 4.280 | 2.537 | -0.273 | -0.002 | 7.85E-01 | 18724 |
| 94 | Temporal_Inf_R | -3.076 | -6.976 | 0.823 | 1.989 | -1.546 | -0.011 | 1.22E-01 | 18391 | -1.492 | -6.914 | 3.931 | 2.766 | -0.539 | -0.004 | 5.90E-01 | 18724 |
| 95 | Cerebellum_Crus1_L | -1.888 | -6.142 | 2.366 | 2.170 | -0.870 | -0.006 | 3.84E-01 | 18391 | -6.037 | -11.936 | -0.138 | 3.009 | -2.006 | -0.015 | 4.49E-02 | 18724 |
| 96 | Cerebellum_Crus1_R | 0.681 | -3.653 | 5.016 | 2.211 | 0.308 | 0.002 | 7.58E-01 | 18391 | -3.996 | -10.014 | 2.022 | 3.070 | -1.301 | -0.010 | 1.93E-01 | 18724 |
| 97 | Cerebellum_Crus2_L | -2.213 | -5.583 | 1.157 | 1.719 | -1.287 | -0.009 | 1.98E-01 | 18391 | -5.704 | -10.384 | -1.024 | 2.388 | -2.389 | -0.017 | 1.69E-02 | 18724 |
| 98 | Cerebellum_Crus2_R | -0.890 | -4.004 | 2.224 | 1.589 | -0.560 | -0.004 | 5.75E-01 | 18391 | -2.702 | -7.032 | 1.628 | 2.209 | -1.223 | -0.009 | 2.21E-01 | 18724 |
| 99 | Cerebellum_3_L | 0.274 | 0.048 | 0.500 | 0.115 | 2.373 | 0.017 | 1.77E-02 | 18391 | -0.131 | -0.445 | 0.183 | 0.160 | -0.818 | -0.006 | 4.14E-01 | 18724 |
| 100 | Cerebellum_3_R | 0.229 | -0.076 | 0.534 | 0.156 | 1.470 | 0.011 | 1.41E-01 | 18391 | -0.138 | -0.562 | 0.286 | 0.216 | -0.639 | -0.005 | 5.23E-01 | 18724 |
| 101 | Cerebellum_4_5_L | 1.987 | 0.158 | 3.815 | 0.933 | 2.129 | 0.016 | 3.33E-02 | 18391 | -0.680 | -3.222 | 1.861 | 1.297 | -0.525 | -0.004 | 6.00E-01 | 18724 |
| 102 | Cerebellum_4_5_R | 0.719 | -0.773 | 2.210 | 0.761 | 0.945 | 0.007 | 3.45E-01 | 18391 | -0.727 | -2.798 | 1.343 | 1.056 | -0.688 | -0.005 | 4.91E-01 | 18724 |

| | | | | | | | | | | | | | | | | | |
|---|---|---|---|---|---|---|---|---|---|---|---|---|---|---|---|---|---|
| 103 | Cerebellum_6_L | 2.434 | -0.728 | 5.597 | 1.613 | 1.509 | 0.011 | 1.31E-01 | 18391 | -3.569 | -7.960 | 0.823 | 2.241 | -1.593 | -0.012 | 1.11E-01 | 18724 |
| 104 | Cerebellum_6_R | 2.211 | -1.119 | 5.541 | 1.699 | 1.301 | 0.010 | 1.93E-01 | 18391 | -2.964 | -7.592 | 1.664 | 2.361 | -1.255 | -0.009 | 2.09E-01 | 18724 |
| 105 | Cerebellum_7b_L | -0.444 | -1.473 | 0.585 | 0.525 | -0.846 | -0.006 | 3.98E-01 | 18391 | -0.711 | -2.142 | 0.720 | 0.730 | -0.974 | -0.007 | 3.30E-01 | 18724 |
| 106 | Cerebellum_7b_R | -0.524 | -1.544 | 0.496 | 0.520 | -1.007 | -0.007 | 3.14E-01 | 18391 | -0.430 | -1.848 | 0.988 | 0.724 | -0.594 | -0.004 | 5.52E-01 | 18724 |
| 107 | Cerebellum_8_L | -1.967 | -5.233 | 1.298 | 1.666 | -1.181 | -0.009 | 2.38E-01 | 18391 | -0.662 | -5.206 | 3.881 | 2.318 | -0.286 | -0.002 | 7.75E-01 | 18724 |
| 108 | Cerebellum_8_R | -2.246 | -6.501 | 2.009 | 2.171 | -1.035 | -0.008 | 3.01E-01 | 18391 | -1.301 | -7.214 | 4.612 | 3.017 | -0.431 | -0.003 | 6.66E-01 | 18724 |
| 109 | Cerebellum_9_L | -0.464 | -2.312 | 1.383 | 0.943 | -0.493 | -0.004 | 6.22E-01 | 18391 | 1.200 | -1.369 | 3.769 | 1.311 | 0.916 | 0.007 | 3.60E-01 | 18724 |
| 110 | Cerebellum_9_R | -0.566 | -2.382 | 1.251 | 0.927 | -0.610 | -0.004 | 5.42E-01 | 18391 | 0.763 | -1.762 | 3.289 | 1.288 | 0.593 | 0.004 | 5.53E-01 | 18724 |
| 111 | Cerebellum_10_L | 0.077 | -0.055 | 0.208 | 0.067 | 1.144 | 0.008 | 2.53E-01 | 18391 | 0.139 | -0.043 | 0.321 | 0.093 | 1.496 | 0.011 | 1.35E-01 | 18724 |
| 112 | Cerebellum_10_R | 0.096 | -0.040 | 0.233 | 0.070 | 1.382 | 0.010 | 1.67E-01 | 18391 | 0.101 | -0.088 | 0.291 | 0.097 | 1.046 | 0.008 | 2.95E-01 | 18724 |
| 113 | Vermis_1_2 | 0.085 | 0.000 | 0.171 | 0.044 | 1.963 | 0.014 | 4.97E-02 | 18391 | -0.026 | -0.144 | 0.093 | 0.060 | -0.429 | -0.003 | 6.68E-01 | 18724 |
| 114 | Vermis_3 | 0.184 | -0.155 | 0.523 | 0.173 | 1.062 | 0.008 | 2.88E-01 | 18391 | -0.201 | -0.672 | 0.270 | 0.240 | -0.836 | -0.006 | 4.03E-01 | 18724 |
| 115 | Vermis_4_5 | -0.421 | -1.417 | 0.575 | 0.508 | -0.828 | -0.006 | 4.08E-01 | 18391 | -0.939 | -2.324 | 0.445 | 0.706 | -1.330 | -0.010 | 1.83E-01 | 18724 |
| 116 | Vermis_6 | 0.298 | -0.256 | 0.852 | 0.283 | 1.054 | 0.008 | 2.92E-01 | 18391 | -0.925 | -1.696 | -0.155 | 0.393 | -2.354 | -0.017 | 1.86E-02 | 18724 |
| 117 | Vermis_7 | 0.285 | -0.029 | 0.599 | 0.160 | 1.781 | 0.013 | 7.49E-02 | 18391 | -0.248 | -0.684 | 0.187 | 0.222 | -1.117 | -0.008 | 2.64E-01 | 18724 |
| 118 | Vermis_8 | 0.517 | 0.002 | 1.032 | 0.263 | 1.966 | 0.014 | 4.93E-02 | 18391 | -0.043 | -0.759 | 0.673 | 0.365 | -0.118 | -0.001 | 9.06E-01 | 18724 |
| 119 | Vermis_9 | 0.195 | -0.222 | 0.612 | 0.213 | 0.916 | 0.007 | 3.60E-01 | 18391 | 0.064 | -0.517 | 0.644 | 0.296 | 0.215 | 0.002 | 8.30E-01 | 18724 |
| 120 | Vermis_10 | 0.014 | -0.102 | 0.130 | 0.059 | 0.235 | 0.002 | 8.14E-01 | 18391 | 0.104 | -0.058 | 0.266 | 0.083 | 1.261 | 0.009 | 2.07E-01 | 18724 |
| 121 | Thal_AV_L | -0.023 | -0.070 | 0.024 | 0.024 | -0.959 | -0.007 | 3.38E-01 | 18391 | -0.039 | -0.104 | 0.026 | 0.033 | -1.175 | -0.009 | 2.40E-01 | 18724 |
| 122 | Thal_AV_R | 0.003 | -0.064 | 0.070 | 0.034 | 0.093 | 0.001 | 9.26E-01 | 18391 | -0.046 | -0.139 | 0.046 | 0.047 | -0.979 | -0.007 | 3.27E-01 | 18724 |
| 123 | Thal_LP_L | -0.013 | -0.065 | 0.039 | 0.026 | -0.482 | -0.004 | 6.30E-01 | 18391 | -0.037 | -0.108 | 0.035 | 0.037 | -0.998 | -0.007 | 3.18E-01 | 18724 |
| 124 | Thal_LP_R | 0.011 | -0.056 | 0.079 | 0.034 | 0.331 | 0.002 | 7.41E-01 | 18391 | -0.024 | -0.118 | 0.070 | 0.048 | -0.502 | -0.004 | 6.16E-01 | 18724 |
| 125 | Thal_VA_L | -0.025 | -0.081 | 0.030 | 0.028 | -0.894 | -0.007 | 3.71E-01 | 18391 | -0.033 | -0.110 | 0.043 | 0.039 | -0.855 | -0.006 | 3.93E-01 | 18724 |
| 126 | Thal_VA_R | -0.007 | -0.075 | 0.061 | 0.035 | -0.190 | -0.001 | 8.49E-01 | 18391 | -0.030 | -0.124 | 0.064 | 0.048 | -0.629 | -0.005 | 5.29E-01 | 18724 |
| 127 | Thal_VL_L | 0.094 | -0.064 | 0.251 | 0.080 | 1.168 | 0.009 | 2.43E-01 | 18391 | -0.118 | -0.336 | 0.100 | 0.111 | -1.063 | -0.008 | 2.88E-01 | 18724 |
| 128 | Thal_VL_R | 0.167 | -0.041 | 0.376 | 0.106 | 1.571 | 0.012 | 1.16E-01 | 18391 | -0.149 | -0.439 | 0.140 | 0.148 | -1.011 | -0.007 | 3.12E-01 | 18724 |
| 129 | Thal_VPL_L | 0.190 | 0.003 | 0.377 | 0.096 | 1.992 | 0.015 | 4.64E-02 | 18391 | 0.026 | -0.234 | 0.286 | 0.132 | 0.197 | 0.001 | 8.44E-01 | 18724 |
| 130 | Thal_VPL_R | 0.149 | -0.048 | 0.346 | 0.100 | 1.485 | 0.011 | 1.38E-01 | 18391 | 0.104 | -0.169 | 0.377 | 0.139 | 0.744 | 0.005 | 4.57E-01 | 18724 |
| 131 | Thal_IL_L | 0.000 | -0.080 | 0.081 | 0.041 | 0.009 | 0.000 | 9.92E-01 | 18391 | 0.044 | -0.068 | 0.156 | 0.057 | 0.767 | 0.006 | 4.43E-01 | 18724 |
| 132 | Thal_IL_R | -0.016 | -0.107 | 0.076 | 0.047 | -0.332 | -0.002 | 7.40E-01 | 18391 | 0.050 | -0.077 | 0.177 | 0.065 | 0.770 | 0.006 | 4.42E-01 | 18724 |
| 133 | Thal_Re_L | 0.000 | -0.001 | 0.000 | 0.000 | -1.057 | -0.008 | 2.90E-01 | 18391 | 0.000 | 0.000 | 0.001 | 0.000 | 1.753 | 0.013 | 7.96E-02 | 18724 |
| 134 | Thal_Re_R | 0.000 | -0.003 | 0.003 | 0.001 | 0.121 | 0.001 | 9.04E-01 | 18391 | 0.001 | -0.003 | 0.005 | 0.002 | 0.626 | 0.005 | 5.31E-01 | 18724 |
| 135 | Thal_MDm_L | -0.186 | -0.785 | 0.414 | 0.306 | -0.607 | -0.004 | 5.44E-01 | 18391 | 0.323 | -0.508 | 1.155 | 0.424 | 0.762 | 0.006 | 4.46E-01 | 18724 |
| 136 | Thal_MDm_R | -0.131 | -0.827 | 0.566 | 0.355 | -0.368 | -0.003 | 7.13E-01 | 18391 | 0.383 | -0.583 | 1.350 | 0.493 | 0.778 | 0.006 | 4.37E-01 | 18724 |
| 137 | Thal_MDl_L | -0.017 | -0.121 | 0.087 | 0.053 | -0.318 | -0.002 | 7.51E-01 | 18391 | 0.023 | -0.121 | 0.167 | 0.073 | 0.313 | 0.002 | 7.55E-01 | 18724 |
| 138 | Thal_MDl_R | 0.004 | -0.114 | 0.122 | 0.060 | 0.066 | 0.000 | 9.47E-01 | 18391 | 0.038 | -0.126 | 0.201 | 0.083 | 0.451 | 0.003 | 6.52E-01 | 18724 |
| 139 | Thal_LGN_L | -0.001 | -0.016 | 0.015 | 0.008 | -0.074 | -0.001 | 9.41E-01 | 18391 | -0.009 | -0.031 | 0.012 | 0.011 | -0.845 | -0.006 | 3.98E-01 | 18724 |
| 140 | Thal_LGN_R | -0.003 | -0.037 | 0.030 | 0.017 | -0.178 | -0.001 | 8.59E-01 | 18391 | -0.013 | -0.060 | 0.033 | 0.024 | -0.557 | -0.004 | 5.77E-01 | 18724 |
| 141 | Thal_MGN_L | -0.013 | -0.041 | 0.015 | 0.014 | -0.897 | -0.007 | 3.69E-01 | 18391 | -0.001 | -0.040 | 0.038 | 0.020 | -0.047 | 0.000 | 9.62E-01 | 18724 |
| 142 | Thal_MGN_R | -0.020 | -0.050 | 0.011 | 0.016 | -1.269 | -0.009 | 2.05E-01 | 18391 | -0.003 | -0.045 | 0.040 | 0.022 | -0.121 | -0.001 | 9.03E-01 | 18724 |
| 143 | Thal_PuA_L | -0.014 | -0.047 | 0.020 | 0.017 | -0.808 | -0.006 | 4.19E-01 | 18391 | -0.009 | -0.055 | 0.037 | 0.023 | -0.393 | -0.003 | 6.94E-01 | 18724 |
| 144 | Thal_PuA_R | -0.034 | -0.103 | 0.035 | 0.035 | -0.974 | -0.007 | 3.30E-01 | 18391 | 0.007 | -0.089 | 0.102 | 0.049 | 0.137 | 0.001 | 8.91E-01 | 18724 |
| 145 | Thal_PuM_L | -0.223 | -0.883 | 0.436 | 0.337 | -0.664 | -0.005 | 5.07E-01 | 18391 | -0.035 | -0.950 | 0.880 | 0.467 | -0.075 | -0.001 | 9.40E-01 | 18724 |
| 146 | Thal_PuM_R | -0.186 | -1.007 | 0.635 | 0.419 | -0.444 | -0.003 | 6.57E-01 | 18391 | 0.200 | -0.940 | 1.341 | 0.582 | 0.345 | 0.003 | 7.30E-01 | 18724 |
| 147 | Thal_PuL_L | -0.015 | -0.135 | 0.106 | 0.061 | -0.239 | -0.002 | 8.11E-01 | 18391 | -0.002 | -0.169 | 0.165 | 0.085 | -0.028 | 0.000 | 9.78E-01 | 18724 |
| 148 | Thal_PuL_R | -0.026 | -0.217 | 0.166 | 0.098 | -0.262 | -0.002 | 7.94E-01 | 18391 | 0.072 | -0.194 | 0.338 | 0.135 | 0.532 | 0.004 | 5.95E-01 | 18724 |
| 149 | Thal_PuI_L | 0.026 | -0.007 | 0.059 | 0.017 | 1.555 | 0.011 | 1.20E-01 | 18391 | 0.000 | -0.046 | 0.045 | 0.023 | -0.011 | 0.000 | 9.91E-01 | 18724 |
| 150 | Thal_PuI_R | 0.023 | -0.020 | 0.066 | 0.022 | 1.042 | 0.008 | 2.97E-01 | 18391 | 0.004 | -0.056 | 0.064 | 0.031 | 0.131 | 0.001 | 8.96E-01 | 18724 |
| 151 | ACC_sub_L | -0.446 | -0.823 | -0.068 | 0.192 | -2.315 | -0.017 | 2.06E-02 | 18391 | 0.404 | -0.120 | 0.928 | 0.267 | 1.512 | 0.011 | 1.30E-01 | 18724 |

| | | | | | | | | | | | | | | | | | |
|---|---|---|---|---|---|---|---|---|---|---|---|---|---|---|---|---|---|
| 152 | ACC_sub_R | -0.463 | -0.815 | -0.110 | 0.180 | -2.573 | -0.019 | 1.01E-02 | 18391 | 0.469 | -0.021 | 0.958 | 0.250 | 1.875 | 0.014 | 6.08E-02 | 18724 |
| 153 | ACC_pre_L | -0.505 | -1.473 | 0.462 | 0.493 | -1.024 | -0.008 | 3.06E-01 | 18391 | 0.817 | -0.527 | 2.160 | 0.685 | 1.192 | 0.009 | 2.33E-01 | 18724 |
| 154 | ACC_pre_R | -0.365 | -1.296 | 0.566 | 0.475 | -0.768 | -0.006 | 4.43E-01 | 18391 | 0.924 | -0.370 | 2.217 | 0.660 | 1.400 | 0.010 | 1.62E-01 | 18724 |
| 155 | ACC_sup_L | -0.695 | -1.746 | 0.356 | 0.536 | -1.296 | -0.010 | 1.95E-01 | 18391 | 0.720 | -0.740 | 2.179 | 0.745 | 0.967 | 0.007 | 3.34E-01 | 18724 |
| 156 | ACC_sup_R | -0.115 | -1.204 | 0.974 | 0.556 | -0.207 | -0.002 | 8.36E-01 | 18391 | 0.861 | -0.651 | 2.373 | 0.771 | 1.116 | 0.008 | 2.64E-01 | 18724 |
| 157 | N_Acc_L | -0.209 | -0.525 | 0.108 | 0.161 | -1.294 | -0.010 | 1.96E-01 | 18391 | 0.210 | -0.229 | 0.649 | 0.224 | 0.937 | 0.007 | 3.49E-01 | 18724 |
| 158 | N_Acc_R | -0.100 | -0.374 | 0.173 | 0.140 | -0.720 | -0.005 | 4.72E-01 | 18391 | 0.185 | -0.195 | 0.565 | 0.194 | 0.956 | 0.007 | 3.39E-01 | 18724 |
| 159 | VTA_L | 0.016 | 0.005 | 0.027 | 0.006 | 2.779 | 0.020 | 5.46E-03 | 18391 | 0.003 | -0.012 | 0.019 | 0.008 | 0.441 | 0.003 | 6.59E-01 | 18724 |
| 160 | VTA_R | 0.013 | 0.003 | 0.024 | 0.005 | 2.633 | 0.019 | 8.48E-03 | 18391 | 0.006 | -0.008 | 0.020 | 0.007 | 0.837 | 0.006 | 4.02E-01 | 18724 |
| 161 | SN_pc_L | 0.001 | -0.017 | 0.019 | 0.009 | 0.152 | 0.001 | 8.80E-01 | 18391 | 0.004 | -0.021 | 0.029 | 0.013 | 0.295 | 0.002 | 7.68E-01 | 18724 |
| 162 | SN_pc_R | 0.001 | -0.023 | 0.024 | 0.012 | 0.047 | 0.000 | 9.62E-01 | 18391 | 0.001 | -0.031 | 0.034 | 0.017 | 0.090 | 0.001 | 9.28E-01 | 18724 |
| 163 | SN_pr_L | 0.001 | -0.024 | 0.025 | 0.012 | 0.049 | 0.000 | 9.61E-01 | 18391 | -0.005 | -0.039 | 0.028 | 0.017 | -0.315 | -0.002 | 7.53E-01 | 18724 |
| 164 | SN_pr_R | 0.000 | -0.028 | 0.027 | 0.014 | -0.029 | 0.000 | 9.77E-01 | 18391 | 0.005 | -0.033 | 0.043 | 0.019 | 0.245 | 0.002 | 8.06E-01 | 18724 |
| 165 | Red_N_L | 0.023 | -0.007 | 0.053 | 0.015 | 1.491 | 0.011 | 1.36E-01 | 18391 | 0.014 | -0.028 | 0.055 | 0.021 | 0.638 | 0.005 | 5.23E-01 | 18724 |
| 166 | Red_N_R | 0.024 | -0.009 | 0.057 | 0.017 | 1.443 | 0.011 | 1.49E-01 | 18391 | 0.021 | -0.024 | 0.066 | 0.023 | 0.907 | 0.007 | 3.64E-01 | 18724 |
| 167 | LC_L | 0.012 | 0.001 | 0.023 | 0.006 | 2.084 | 0.015 | 3.71E-02 | 18391 | 0.005 | -0.010 | 0.021 | 0.008 | 0.656 | 0.005 | 5.12E-01 | 18724 |
| 168 | LC_R | 0.001 | -0.011 | 0.013 | 0.006 | 0.171 | 0.001 | 8.64E-01 | 18391 | 0.006 | -0.011 | 0.023 | 0.009 | 0.680 | 0.005 | 4.97E-01 | 18724 |
| 169 | Raphe_D | 0.019 | -0.024 | 0.062 | 0.022 | 0.858 | 0.006 | 3.91E-01 | 18391 | 0.033 | -0.028 | 0.093 | 0.031 | 1.060 | 0.008 | 2.89E-01 | 18724 |
| 170 | Raphe_M | 0.017 | 0.004 | 0.030 | 0.007 | 2.489 | 0.018 | 1.28E-02 | 18391 | 0.008 | -0.010 | 0.027 | 0.009 | 0.878 | 0.006 | 3.80E-01 | 18724 |

| | | Salad / raw vegetable intake | | | | | | | | Poultry intake | | | | | | | |
|---|---|---|---|---|---|---|---|---|---|---|---|---|---|---|---|---|---|
| Number | AAL 3 region | beta | 95%CI_l | 95%CI_h | se | t | r | p | df.e | beta | 95%CI_l | 95%CI_h | se | t | r | p | df.e |
| 0 | Total brain | -54.236 | -142.943 | 34.471 | 45.257 | -1.198 | -0.009 | 2.31E-01 | 18404 | -39.525 | -235.836 | 156.786 | 100.154 | -0.395 | -0.003 | 6.93E-01 | 18727 |
| 1 | Precentral_L | -1.149 | -4.267 | 1.969 | 1.591 | -0.722 | -0.005 | 4.70E-01 | 18404 | -0.515 | -7.422 | 6.392 | 3.524 | -0.146 | -0.001 | 8.84E-01 | 18727 |
| 2 | Precentral_R | 0.523 | -2.336 | 3.381 | 1.458 | 0.359 | 0.003 | 7.20E-01 | 18404 | -1.155 | -7.485 | 5.174 | 3.229 | -0.358 | -0.003 | 7.21E-01 | 18727 |
| 3 | Frontal_Sup_2_L | -1.776 | -5.071 | 1.519 | 1.681 | -1.057 | -0.008 | 2.91E-01 | 18404 | 0.456 | -6.838 | 7.750 | 3.721 | 0.122 | 0.001 | 9.03E-01 | 18727 |
| 4 | Frontal_Sup_2_R | -0.148 | -3.781 | 3.484 | 1.853 | -0.080 | -0.001 | 9.36E-01 | 18404 | 1.375 | -6.661 | 9.411 | 4.100 | 0.335 | 0.002 | 7.37E-01 | 18727 |
| 5 | Frontal_Mid_2_L | 0.029 | -3.432 | 3.490 | 1.766 | 0.017 | 0.000 | 9.87E-01 | 18404 | 4.960 | -2.697 | 12.617 | 3.907 | 1.270 | 0.009 | 2.04E-01 | 18727 |
| 6 | Frontal_Mid_2_R | -0.107 | -4.020 | 3.805 | 1.996 | -0.054 | 0.000 | 9.57E-01 | 18404 | -2.817 | -11.483 | 5.850 | 4.421 | -0.637 | -0.005 | 5.24E-01 | 18727 |
| 7 | Frontal_Inf_Oper_L | -0.272 | -1.294 | 0.750 | 0.522 | -0.521 | -0.004 | 6.02E-01 | 18404 | 0.122 | -2.141 | 2.385 | 1.155 | 0.106 | 0.001 | 9.16E-01 | 18727 |
| 8 | Frontal_Inf_Oper_R | -0.691 | -1.972 | 0.591 | 0.654 | -1.056 | -0.008 | 2.91E-01 | 18404 | 0.561 | -2.276 | 3.399 | 1.448 | 0.388 | 0.003 | 6.98E-01 | 18727 |
| 9 | Frontal_Inf_Tri_L | -0.605 | -2.777 | 1.567 | 1.108 | -0.546 | -0.004 | 5.85E-01 | 18404 | -0.422 | -5.228 | 4.385 | 2.452 | -0.172 | -0.001 | 8.64E-01 | 18727 |
| 10 | Frontal_Inf_Tri_R | 0.875 | -0.829 | 2.579 | 0.869 | 1.006 | 0.007 | 3.14E-01 | 18404 | 0.968 | -2.805 | 4.741 | 1.925 | 0.503 | 0.004 | 6.15E-01 | 18727 |
| 11 | Frontal_Inf_Orb_2_L | -0.551 | -1.200 | 0.099 | 0.331 | -1.661 | -0.012 | 9.67E-02 | 18404 | 0.132 | -1.305 | 1.569 | 0.733 | 0.180 | 0.001 | 8.57E-01 | 18727 |
| 12 | Frontal_Inf_Orb_2_R | -0.151 | -0.830 | 0.529 | 0.347 | -0.435 | -0.003 | 6.64E-01 | 18404 | 0.525 | -0.978 | 2.028 | 0.767 | 0.685 | 0.005 | 4.94E-01 | 18727 |
| 13 | Rolandic_Oper_L | -0.877 | -1.859 | 0.104 | 0.501 | -1.752 | -0.013 | 7.99E-02 | 18404 | -0.221 | -2.394 | 1.952 | 1.109 | -0.199 | -0.001 | 8.42E-01 | 18727 |
| 14 | Rolandic_Oper_R | -0.378 | -1.785 | 1.028 | 0.718 | -0.527 | -0.004 | 5.98E-01 | 18404 | -0.294 | -3.404 | 2.816 | 1.587 | -0.185 | -0.001 | 8.53E-01 | 18727 |
| 15 | Supp_Motor_Area_L | -0.404 | -2.737 | 1.928 | 1.190 | -0.340 | -0.003 | 7.34E-01 | 18404 | -1.807 | -6.972 | 3.359 | 2.635 | -0.686 | -0.005 | 4.93E-01 | 18727 |
| 16 | Supp_Motor_Area_R | 0.695 | -1.881 | 3.271 | 1.314 | 0.529 | 0.004 | 5.97E-01 | 18404 | -1.474 | -7.181 | 4.234 | 2.912 | -0.506 | -0.004 | 6.13E-01 | 18727 |
| 17 | Olfactory_L | -0.231 | -0.578 | 0.117 | 0.177 | -1.303 | -0.010 | 1.93E-01 | 18404 | 0.484 | -0.284 | 1.252 | 0.392 | 1.235 | 0.009 | 2.17E-01 | 18727 |
| 18 | Olfactory_R | -0.250 | -0.598 | 0.099 | 0.178 | -1.405 | -0.010 | 1.60E-01 | 18404 | 0.339 | -0.431 | 1.109 | 0.393 | 0.862 | 0.006 | 3.88E-01 | 18727 |
| 19 | Frontal_Sup_Medial_L | -1.720 | -3.908 | 0.468 | 1.116 | -1.541 | -0.011 | 1.23E-01 | 18404 | -1.448 | -6.288 | 3.392 | 2.469 | -0.586 | -0.004 | 5.58E-01 | 18727 |
| 20 | Frontal_Sup_Medial_R | -0.747 | -2.340 | 0.847 | 0.813 | -0.918 | -0.007 | 3.58E-01 | 18404 | -1.221 | -4.744 | 2.303 | 1.798 | -0.679 | -0.005 | 4.97E-01 | 18727 |
| 21 | Frontal_Med_Orb_L | -0.968 | -1.786 | -0.151 | 0.417 | -2.321 | -0.017 | 2.03E-02 | 18404 | 0.196 | -1.613 | 2.006 | 0.923 | 0.212 | 0.002 | 8.32E-01 | 18727 |
| 22 | Frontal_Med_Orb_R | -1.154 | -2.159 | -0.150 | 0.513 | -2.252 | -0.017 | 2.43E-02 | 18404 | -0.959 | -3.183 | 1.264 | 1.134 | -0.846 | -0.006 | 3.98E-01 | 18727 |
| 23 | Rectus_L | -0.517 | -1.364 | 0.330 | 0.432 | -1.197 | -0.009 | 2.31E-01 | 18404 | -0.849 | -2.721 | 1.023 | 0.955 | -0.889 | -0.006 | 3.74E-01 | 18727 |
| 24 | Rectus_R | -0.270 | -1.053 | 0.513 | 0.399 | -0.675 | -0.005 | 5.00E-01 | 18404 | -1.144 | -2.876 | 0.587 | 0.884 | -1.295 | -0.009 | 1.95E-01 | 18727 |
| 25 | OFCmed_L | -0.366 | -0.857 | 0.125 | 0.250 | -1.460 | -0.011 | 1.44E-01 | 18404 | -0.521 | -1.606 | 0.564 | 0.554 | -0.942 | -0.007 | 3.46E-01 | 18727 |
| 26 | OFCmed_R | -0.593 | -1.164 | -0.021 | 0.292 | -2.032 | -0.015 | 4.22E-02 | 18404 | -0.286 | -1.550 | 0.979 | 0.645 | -0.443 | -0.003 | 6.58E-01 | 18727 |
| 27 | OFCant_L | -0.533 | -1.101 | 0.034 | 0.289 | -1.842 | -0.014 | 6.55E-02 | 18404 | 0.013 | -1.242 | 1.268 | 0.640 | 0.020 | 0.000 | 9.84E-01 | 18727 |

| | | | | | | | | | | | | | | | | | |
|---|---|---|---|---|---|---|---|---|---|---|---|---|---|---|---|---|---|
| 28 | OFCant_R | -0.674 | -1.424 | 0.076 | 0.383 | -1.762 | -0.013 | 7.82E-02 | 18404 | -0.122 | -1.779 | 1.536 | 0.846 | -0.144 | -0.001 | 8.85E-01 | 18727 |
| 29 | OFCpost_L | -0.709 | -1.419 | 0.001 | 0.362 | -1.958 | -0.014 | 5.02E-02 | 18404 | 0.295 | -1.277 | 1.867 | 0.802 | 0.368 | 0.003 | 7.13E-01 | 18727 |
| 30 | OFCpost_R | -0.757 | -1.413 | -0.100 | 0.335 | -2.259 | -0.017 | 2.39E-02 | 18404 | 0.189 | -1.263 | 1.642 | 0.741 | 0.255 | 0.002 | 7.98E-01 | 18727 |
| 31 | OFClat_L | -0.182 | -0.422 | 0.058 | 0.122 | -1.489 | -0.011 | 1.37E-01 | 18404 | -0.133 | -0.663 | 0.397 | 0.270 | -0.493 | -0.004 | 6.22E-01 | 18727 |
| 32 | OFClat_R | -0.093 | -0.295 | 0.110 | 0.103 | -0.897 | -0.007 | 3.70E-01 | 18404 | -0.043 | -0.491 | 0.405 | 0.228 | -0.189 | -0.001 | 8.50E-01 | 18727 |
| 33 | Insula_L | -1.182 | -3.379 | 1.015 | 1.121 | -1.055 | -0.008 | 2.92E-01 | 18404 | -1.567 | -6.426 | 3.292 | 2.479 | -0.632 | -0.005 | 5.27E-01 | 18727 |
| 34 | Insula_R | -0.720 | -2.860 | 1.421 | 1.092 | -0.659 | -0.005 | 5.10E-01 | 18404 | -2.220 | -6.954 | 2.515 | 2.416 | -0.919 | -0.007 | 3.58E-01 | 18727 |
| 37 | Cingulate_Mid_L | 1.467 | -0.390 | 3.324 | 0.947 | 1.549 | 0.011 | 1.21E-01 | 18404 | -1.145 | -5.250 | 2.960 | 2.094 | -0.547 | -0.004 | 5.85E-01 | 18727 |
| 38 | Cingulate_Mid_R | 0.912 | -1.067 | 2.892 | 1.010 | 0.903 | 0.007 | 3.66E-01 | 18404 | -1.943 | -6.322 | 2.435 | 2.234 | -0.870 | -0.006 | 3.84E-01 | 18727 |
| 39 | Cingulate_Post_L | -0.433 | -1.000 | 0.134 | 0.289 | -1.496 | -0.011 | 1.35E-01 | 18404 | -0.580 | -1.835 | 0.674 | 0.640 | -0.907 | -0.007 | 3.65E-01 | 18727 |
| 40 | Cingulate_Post_R | -0.252 | -0.566 | 0.063 | 0.161 | -1.567 | -0.012 | 1.17E-01 | 18404 | -0.214 | -0.910 | 0.482 | 0.355 | -0.602 | -0.004 | 5.47E-01 | 18727 |
| 41 | Hippocampus_L | 0.573 | -0.344 | 1.490 | 0.468 | 1.226 | 0.009 | 2.20E-01 | 18404 | -1.267 | -3.296 | 0.763 | 1.035 | -1.223 | -0.009 | 2.21E-01 | 18727 |
| 42 | Hippocampus_R | 0.692 | -0.259 | 1.643 | 0.485 | 1.426 | 0.011 | 1.54E-01 | 18404 | -0.288 | -2.392 | 1.816 | 1.073 | -0.268 | -0.002 | 7.88E-01 | 18727 |
| 43 | ParaHippocampal_L | 0.432 | -0.382 | 1.246 | 0.415 | 1.041 | 0.008 | 2.98E-01 | 18404 | -1.301 | -3.104 | 0.503 | 0.920 | -1.414 | -0.010 | 1.57E-01 | 18727 |
| 44 | ParaHippocampal_R | 0.901 | -0.128 | 1.930 | 0.525 | 1.717 | 0.013 | 8.60E-02 | 18404 | -1.122 | -3.397 | 1.154 | 1.161 | -0.966 | -0.007 | 3.34E-01 | 18727 |
| 45 | Amygdala_L | -0.031 | -0.348 | 0.286 | 0.162 | -0.193 | -0.001 | 8.47E-01 | 18404 | -0.365 | -1.066 | 0.336 | 0.358 | -1.021 | -0.007 | 3.07E-01 | 18727 |
| 46 | Amygdala_R | -0.089 | -0.413 | 0.235 | 0.165 | -0.537 | -0.004 | 5.91E-01 | 18404 | -0.254 | -0.971 | 0.463 | 0.366 | -0.695 | -0.005 | 4.87E-01 | 18727 |
| 47 | Calcarine_L | -0.965 | -3.986 | 2.056 | 1.541 | -0.626 | -0.005 | 5.31E-01 | 18404 | 6.349 | -0.337 | 13.035 | 3.411 | 1.861 | 0.014 | 6.27E-02 | 18727 |
| 48 | Calcarine_R | -1.052 | -3.576 | 1.473 | 1.288 | -0.817 | -0.006 | 4.14E-01 | 18404 | 3.568 | -2.020 | 9.156 | 2.851 | 1.252 | 0.009 | 2.11E-01 | 18727 |
| 49 | Cuneus_L | 0.128 | -1.767 | 2.023 | 0.967 | 0.132 | 0.001 | 8.95E-01 | 18404 | 2.921 | -1.274 | 7.115 | 2.140 | 1.365 | 0.010 | 1.72E-01 | 18727 |
| 50 | Cuneus_R | -0.729 | -2.180 | 0.723 | 0.740 | -0.984 | -0.007 | 3.25E-01 | 18404 | 0.601 | -2.609 | 3.811 | 1.638 | 0.367 | 0.003 | 7.14E-01 | 18727 |
| 51 | Lingual_L | 0.500 | -1.319 | 2.318 | 0.928 | 0.539 | 0.004 | 5.90E-01 | 18404 | 3.331 | -0.689 | 7.351 | 2.051 | 1.624 | 0.012 | 1.04E-01 | 18727 |
| 52 | Lingual_R | -0.328 | -2.233 | 1.576 | 0.971 | -0.338 | -0.002 | 7.35E-01 | 18404 | 2.079 | -2.134 | 6.293 | 2.150 | 0.967 | 0.007 | 3.33E-01 | 18727 |
| 53 | Occipital_Sup_L | 0.344 | -0.906 | 1.594 | 0.638 | 0.540 | 0.004 | 5.89E-01 | 18404 | 1.488 | -1.276 | 4.252 | 1.410 | 1.055 | 0.008 | 2.91E-01 | 18727 |
| 54 | Occipital_Sup_R | -0.392 | -1.747 | 0.963 | 0.691 | -0.567 | -0.004 | 5.71E-01 | 18404 | 1.514 | -1.482 | 4.511 | 1.529 | 0.991 | 0.007 | 3.22E-01 | 18727 |
| 55 | Occipital_Mid_L | -0.940 | -3.868 | 1.988 | 1.494 | -0.629 | -0.005 | 5.29E-01 | 18404 | 2.639 | -3.841 | 9.120 | 3.306 | 0.798 | 0.006 | 4.25E-01 | 18727 |
| 56 | Occipital_Mid_R | -1.773 | -3.810 | 0.263 | 1.039 | -1.707 | -0.013 | 8.79E-02 | 18404 | 2.027 | -2.482 | 6.536 | 2.300 | 0.881 | 0.006 | 3.78E-01 | 18727 |
| 57 | Occipital_Inf_L | -1.438 | -2.411 | -0.466 | 0.496 | -2.900 | -0.021 | 3.74E-03 | 18404 | 1.124 | -1.027 | 3.276 | 1.098 | 1.024 | 0.007 | 3.06E-01 | 18727 |
| 58 | Occipital_Inf_R | -1.268 | -2.391 | -0.145 | 0.573 | -2.213 | -0.016 | 2.69E-02 | 18404 | -0.114 | -2.599 | 2.370 | 1.268 | -0.090 | -0.001 | 9.28E-01 | 18727 |
| 59 | Fusiform_L | -0.043 | -2.172 | 2.086 | 1.086 | -0.039 | 0.000 | 9.69E-01 | 18404 | -1.225 | -5.934 | 3.485 | 2.403 | -0.510 | -0.004 | 6.10E-01 | 18727 |
| 60 | Fusiform_R | -0.272 | -2.611 | 2.067 | 1.193 | -0.228 | -0.002 | 8.20E-01 | 18404 | -1.375 | -6.553 | 3.804 | 2.642 | -0.520 | -0.004 | 6.03E-01 | 18727 |
| 61 | Postcentral_L | 1.419 | -1.783 | 4.620 | 1.633 | 0.869 | 0.006 | 3.85E-01 | 18404 | -4.022 | -11.100 | 3.056 | 3.611 | -1.114 | -0.008 | 2.65E-01 | 18727 |
| 62 | Postcentral_R | 2.770 | -0.514 | 6.053 | 1.675 | 1.653 | 0.012 | 9.83E-02 | 18404 | -3.563 | -10.820 | 3.694 | 3.702 | -0.962 | -0.007 | 3.36E-01 | 18727 |
| 63 | Parietal_Sup_L | -0.291 | -2.136 | 1.553 | 0.941 | -0.309 | -0.002 | 7.57E-01 | 18404 | -1.194 | -5.274 | 2.886 | 2.081 | -0.574 | -0.004 | 5.66E-01 | 18727 |
| 64 | Parietal_Sup_R | 0.312 | -1.324 | 1.949 | 0.835 | 0.374 | 0.003 | 7.08E-01 | 18404 | 1.494 | -2.124 | 5.113 | 1.846 | 0.809 | 0.006 | 4.18E-01 | 18727 |
| 65 | Parietal_Inf_L | -1.729 | -4.145 | 0.687 | 1.233 | -1.403 | -0.010 | 1.61E-01 | 18404 | -1.453 | -6.795 | 3.890 | 2.726 | -0.533 | -0.004 | 5.94E-01 | 18727 |
| 66 | Parietal_Inf_R | 0.080 | -1.485 | 1.644 | 0.798 | 0.100 | 0.001 | 9.21E-01 | 18404 | -2.064 | -5.525 | 1.397 | 1.766 | -1.169 | -0.009 | 2.42E-01 | 18727 |
| 67 | SupraMarginal_L | -0.979 | -2.394 | 0.436 | 0.722 | -1.356 | -0.010 | 1.75E-01 | 18404 | -2.793 | -5.923 | 0.337 | 1.597 | -1.749 | -0.013 | 8.03E-02 | 18727 |
| 68 | SupraMarginal_R | 0.509 | -1.493 | 2.511 | 1.021 | 0.498 | 0.004 | 6.18E-01 | 18404 | -4.157 | -8.585 | 0.272 | 2.259 | -1.840 | -0.013 | 6.58E-02 | 18727 |
| 69 | Angular_L | -0.465 | -1.856 | 0.925 | 0.710 | -0.656 | -0.005 | 5.12E-01 | 18404 | -1.707 | -4.783 | 1.368 | 1.569 | -1.088 | -0.008 | 2.77E-01 | 18727 |
| 70 | Angular_R | 0.013 | -1.850 | 1.877 | 0.951 | 0.014 | 0.000 | 9.89E-01 | 18404 | 0.510 | -3.616 | 4.635 | 2.105 | 0.242 | 0.002 | 8.09E-01 | 18727 |
| 71 | Precuneus_L | 0.290 | -3.099 | 3.679 | 1.729 | 0.168 | 0.001 | 8.67E-01 | 18404 | -1.969 | -9.461 | 5.523 | 3.822 | -0.515 | -0.004 | 6.06E-01 | 18727 |
| 72 | Precuneus_R | -0.929 | -3.951 | 2.092 | 1.541 | -0.603 | -0.004 | 5.47E-01 | 18404 | -0.468 | -7.144 | 6.208 | 3.406 | -0.137 | -0.001 | 8.91E-01 | 18727 |
| 73 | Paracentral_Lobule_L | 0.929 | -0.621 | 2.478 | 0.791 | 1.174 | 0.009 | 2.40E-01 | 18404 | -2.894 | -6.326 | 0.538 | 1.751 | -1.653 | -0.012 | 9.84E-02 | 18727 |
| 74 | Paracentral_Lobule_R | 0.256 | -0.856 | 1.367 | 0.567 | 0.451 | 0.003 | 6.52E-01 | 18404 | -0.631 | -3.089 | 1.826 | 1.254 | -0.503 | -0.004 | 6.15E-01 | 18727 |
| 75 | Caudate_L | 0.294 | -0.947 | 1.535 | 0.633 | 0.464 | 0.003 | 6.42E-01 | 18404 | 0.092 | -2.649 | 2.834 | 1.399 | 0.066 | 0.000 | 9.47E-01 | 18727 |
| 76 | Caudate_R | 0.085 | -1.318 | 1.488 | 0.716 | 0.119 | 0.001 | 9.06E-01 | 18404 | 0.561 | -2.543 | 3.665 | 1.583 | 0.354 | 0.003 | 7.23E-01 | 18727 |
| 77 | Putamen_L | -1.065 | -2.948 | 0.817 | 0.960 | -1.109 | -0.008 | 2.67E-01 | 18404 | 1.825 | -2.340 | 5.990 | 2.125 | 0.859 | 0.006 | 3.90E-01 | 18727 |
| 78 | Putamen_R | -1.129 | -3.046 | 0.787 | 0.978 | -1.155 | -0.009 | 2.48E-01 | 18404 | -0.037 | -4.278 | 4.204 | 2.164 | -0.017 | 0.000 | 9.86E-01 | 18727 |

| | | | | | | | | | | | | | | | | | |
|---|---|---|---|---|---|---|---|---|---|---|---|---|---|---|---|---|---|
| 79 | Pallidum_L | 0.059 | -0.108 | 0.227 | 0.085 | 0.695 | 0.005 | 4.87E-01 | 18404 | 0.091 | -0.279 | 0.461 | 0.189 | 0.483 | 0.004 | 6.29E-01 | 18727 |
| 80 | Pallidum_R | -0.017 | -0.200 | 0.167 | 0.094 | -0.177 | -0.001 | 8.59E-01 | 18404 | 0.038 | -0.369 | 0.444 | 0.207 | 0.181 | 0.001 | 8.56E-01 | 18727 |
| 83 | Heschl_L | -0.341 | -0.680 | -0.002 | 0.173 | -1.970 | -0.015 | 4.88E-02 | 18404 | -0.053 | -0.804 | 0.697 | 0.383 | -0.139 | -0.001 | 8.89E-01 | 18727 |
| 84 | Heschl_R | -0.021 | -0.400 | 0.358 | 0.193 | -0.110 | -0.001 | 9.12E-01 | 18404 | 0.154 | -0.685 | 0.992 | 0.428 | 0.360 | 0.003 | 7.19E-01 | 18727 |
| 85 | Temporal_Sup_L | -3.054 | -5.504 | -0.604 | 1.250 | -2.444 | -0.018 | 1.46E-02 | 18404 | -3.242 | -8.660 | 2.175 | 2.764 | -1.173 | -0.009 | 2.41E-01 | 18727 |
| 86 | Temporal_Sup_R | -2.036 | -4.745 | 0.674 | 1.382 | -1.472 | -0.011 | 1.41E-01 | 18404 | -2.307 | -8.300 | 3.686 | 3.057 | -0.755 | -0.006 | 4.51E-01 | 18727 |
| 87 | Temporal_Pole_Sup_L | -0.789 | -1.948 | 0.370 | 0.591 | -1.335 | -0.010 | 1.82E-01 | 18404 | -1.454 | -4.022 | 1.114 | 1.310 | -1.110 | -0.008 | 2.67E-01 | 18727 |
| 88 | Temporal_Pole_Sup_R | -1.157 | -2.378 | 0.064 | 0.623 | -1.857 | -0.014 | 6.33E-02 | 18404 | -0.739 | -3.443 | 1.965 | 1.380 | -0.536 | -0.004 | 5.92E-01 | 18727 |
| 89 | Temporal_Mid_L | -1.600 | -5.682 | 2.483 | 2.083 | -0.768 | -0.006 | 4.42E-01 | 18404 | -6.205 | -15.237 | 2.826 | 4.608 | -1.347 | -0.010 | 1.78E-01 | 18727 |
| 90 | Temporal_Mid_R | -1.400 | -5.025 | 2.226 | 1.850 | -0.757 | -0.006 | 4.49E-01 | 18404 | -3.066 | -11.088 | 4.957 | 4.093 | -0.749 | -0.005 | 4.54E-01 | 18727 |
| 91 | Temporal_Pole_Mid_L | -0.047 | -1.022 | 0.927 | 0.497 | -0.095 | -0.001 | 9.24E-01 | 18404 | -2.051 | -4.209 | 0.107 | 1.101 | -1.863 | -0.014 | 6.25E-02 | 18727 |
| 92 | Temporal_Pole_Mid_R | -0.401 | -1.586 | 0.783 | 0.604 | -0.664 | -0.005 | 5.07E-01 | 18404 | -0.749 | -3.371 | 1.874 | 1.338 | -0.559 | -0.004 | 5.76E-01 | 18727 |
| 93 | Temporal_Inf_L | -0.697 | -3.504 | 2.111 | 1.432 | -0.486 | -0.004 | 6.27E-01 | 18404 | -3.162 | -9.371 | 3.048 | 3.168 | -0.998 | -0.007 | 3.18E-01 | 18727 |
| 94 | Temporal_Inf_R | -0.984 | -4.047 | 2.078 | 1.563 | -0.630 | -0.005 | 5.29E-01 | 18404 | -2.232 | -9.005 | 4.541 | 3.455 | -0.646 | -0.005 | 5.18E-01 | 18727 |
| 95 | Cerebellum_Crus1_L | -3.535 | -6.866 | -0.203 | 1.700 | -2.080 | -0.015 | 3.76E-02 | 18404 | 3.433 | -3.938 | 10.804 | 3.761 | 0.913 | 0.007 | 3.61E-01 | 18727 |
| 96 | Cerebellum_Crus1_R | -2.262 | -5.658 | 1.135 | 1.733 | -1.305 | -0.010 | 1.92E-01 | 18404 | 4.269 | -3.248 | 11.786 | 3.835 | 1.113 | 0.008 | 2.66E-01 | 18727 |
| 97 | Cerebellum_Crus2_L | -3.216 | -5.859 | -0.574 | 1.348 | -2.386 | -0.018 | 1.71E-02 | 18404 | -2.330 | -8.180 | 3.520 | 2.985 | -0.781 | -0.006 | 4.35E-01 | 18727 |
| 98 | Cerebellum_Crus2_R | -1.172 | -3.616 | 1.272 | 1.247 | -0.940 | -0.007 | 3.47E-01 | 18404 | -2.530 | -7.940 | 2.880 | 2.760 | -0.917 | -0.007 | 3.59E-01 | 18727 |
| 99 | Cerebellum_3_L | 0.065 | -0.113 | 0.242 | 0.091 | 0.717 | 0.005 | 4.73E-01 | 18404 | 0.408 | 0.015 | 0.800 | 0.200 | 2.035 | 0.015 | 4.18E-02 | 18727 |
| 100 | Cerebellum_3_R | 0.001 | -0.238 | 0.241 | 0.122 | 0.011 | 0.000 | 9.91E-01 | 18404 | 0.342 | -0.187 | 0.872 | 0.270 | 1.267 | 0.009 | 2.05E-01 | 18727 |
| 101 | Cerebellum_4_5_L | 0.167 | -1.266 | 1.601 | 0.731 | 0.229 | 0.002 | 8.19E-01 | 18404 | 2.202 | -0.972 | 5.377 | 1.619 | 1.360 | 0.010 | 1.74E-01 | 18727 |
| 102 | Cerebellum_4_5_R | -0.037 | -1.206 | 1.132 | 0.597 | -0.062 | 0.000 | 9.51E-01 | 18404 | 0.824 | -1.762 | 3.411 | 1.320 | 0.625 | 0.005 | 5.32E-01 | 18727 |
| 103 | Cerebellum_6_L | -0.395 | -2.874 | 2.084 | 1.265 | -0.312 | -0.002 | 7.55E-01 | 18404 | 0.559 | -4.926 | 6.045 | 2.799 | 0.200 | 0.001 | 8.42E-01 | 18727 |
| 104 | Cerebellum_6_R | -0.194 | -2.806 | 2.418 | 1.332 | -0.146 | -0.001 | 8.84E-01 | 18404 | 2.191 | -3.589 | 7.971 | 2.949 | 0.743 | 0.005 | 4.58E-01 | 18727 |
| 105 | Cerebellum_7b_L | -0.796 | -1.604 | 0.011 | 0.412 | -1.932 | -0.014 | 5.33E-02 | 18404 | -0.556 | -2.345 | 1.232 | 0.912 | -0.610 | -0.004 | 5.42E-01 | 18727 |
| 106 | Cerebellum_7b_R | -0.494 | -1.295 | 0.307 | 0.409 | -1.209 | -0.009 | 2.27E-01 | 18404 | -1.407 | -3.180 | 0.365 | 0.904 | -1.557 | -0.011 | 1.20E-01 | 18727 |
| 107 | Cerebellum_8_L | -2.100 | -4.663 | 0.462 | 1.307 | -1.607 | -0.012 | 1.08E-01 | 18404 | -1.851 | -7.525 | 3.823 | 2.895 | -0.640 | -0.005 | 5.22E-01 | 18727 |
| 108 | Cerebellum_8_R | -3.914 | -7.251 | -0.576 | 1.703 | -2.298 | -0.017 | 2.16E-02 | 18404 | -5.455 | -12.843 | 1.934 | 3.769 | -1.447 | -0.011 | 1.48E-01 | 18727 |
| 109 | Cerebellum_9_L | -1.276 | -2.725 | 0.173 | 0.739 | -1.726 | -0.013 | 8.44E-02 | 18404 | -0.012 | -3.219 | 3.195 | 1.636 | -0.008 | 0.000 | 9.94E-01 | 18727 |
| 110 | Cerebellum_9_R | -1.244 | -2.669 | 0.182 | 0.727 | -1.710 | -0.013 | 8.74E-02 | 18404 | 0.401 | -2.752 | 3.554 | 1.609 | 0.249 | 0.002 | 8.03E-01 | 18727 |
| 111 | Cerebellum_10_L | -0.155 | -0.257 | -0.052 | 0.052 | -2.951 | -0.022 | 3.17E-03 | 18404 | -0.013 | -0.241 | 0.214 | 0.116 | -0.114 | -0.001 | 9.09E-01 | 18727 |
| 112 | Cerebellum_10_R | -0.083 | -0.190 | 0.024 | 0.055 | -1.514 | -0.011 | 1.30E-01 | 18404 | 0.101 | -0.136 | 0.337 | 0.121 | 0.832 | 0.006 | 4.05E-01 | 18727 |
| 113 | Vermis_1_2 | 0.047 | -0.020 | 0.114 | 0.034 | 1.379 | 0.010 | 1.68E-01 | 18404 | 0.132 | -0.016 | 0.280 | 0.075 | 1.743 | 0.013 | 8.13E-02 | 18727 |
| 114 | Vermis_3 | 0.103 | -0.163 | 0.369 | 0.136 | 0.762 | 0.006 | 4.46E-01 | 18404 | 0.408 | -0.180 | 0.996 | 0.300 | 1.359 | 0.010 | 1.74E-01 | 18727 |
| 115 | Vermis_4_5 | 0.230 | -0.551 | 1.012 | 0.399 | 0.578 | 0.004 | 5.64E-01 | 18404 | 0.766 | -0.963 | 2.495 | 0.882 | 0.868 | 0.006 | 3.85E-01 | 18727 |
| 116 | Vermis_6 | 0.054 | -0.381 | 0.489 | 0.222 | 0.244 | 0.002 | 8.07E-01 | 18404 | 0.724 | -0.238 | 1.687 | 0.491 | 1.475 | 0.011 | 1.40E-01 | 18727 |
| 117 | Vermis_7 | -0.057 | -0.303 | 0.189 | 0.125 | -0.454 | -0.003 | 6.50E-01 | 18404 | 0.430 | -0.114 | 0.974 | 0.277 | 1.551 | 0.011 | 1.21E-01 | 18727 |
| 118 | Vermis_8 | 0.049 | -0.355 | 0.453 | 0.206 | 0.238 | 0.002 | 8.12E-01 | 18404 | 1.074 | 0.179 | 1.968 | 0.456 | 2.353 | 0.017 | 1.86E-02 | 18727 |
| 119 | Vermis_9 | -0.006 | -0.333 | 0.322 | 0.167 | -0.034 | 0.000 | 9.73E-01 | 18404 | 0.369 | -0.356 | 1.094 | 0.370 | 0.997 | 0.007 | 3.19E-01 | 18727 |
| 120 | Vermis_10 | 0.007 | -0.084 | 0.098 | 0.047 | 0.149 | 0.001 | 8.81E-01 | 18404 | 0.191 | -0.011 | 0.393 | 0.103 | 1.853 | 0.014 | 6.39E-02 | 18727 |
| 121 | Thal_AV_L | 0.015 | -0.022 | 0.052 | 0.019 | 0.809 | 0.006 | 4.18E-01 | 18404 | -0.029 | -0.110 | 0.052 | 0.041 | -0.698 | -0.005 | 4.85E-01 | 18727 |
| 122 | Thal_AV_R | 0.056 | 0.004 | 0.108 | 0.027 | 2.107 | 0.016 | 3.51E-02 | 18404 | -0.063 | -0.179 | 0.052 | 0.059 | -1.073 | -0.008 | 2.83E-01 | 18727 |
| 123 | Thal_LP_L | -0.007 | -0.048 | 0.033 | 0.021 | -0.345 | -0.003 | 7.30E-01 | 18404 | -0.111 | -0.201 | -0.021 | 0.046 | -2.420 | -0.018 | 1.55E-02 | 18727 |
| 124 | Thal_LP_R | 0.017 | -0.035 | 0.070 | 0.027 | 0.648 | 0.005 | 5.17E-01 | 18404 | -0.109 | -0.226 | 0.008 | 0.060 | -1.824 | -0.013 | 6.82E-02 | 18727 |
| 125 | Thal_VA_L | 0.015 | -0.028 | 0.059 | 0.022 | 0.694 | 0.005 | 4.88E-01 | 18404 | -0.020 | -0.116 | 0.075 | 0.049 | -0.419 | -0.003 | 6.75E-01 | 18727 |
| 126 | Thal_VA_R | 0.028 | -0.025 | 0.082 | 0.027 | 1.045 | 0.008 | 2.96E-01 | 18404 | -0.023 | -0.140 | 0.095 | 0.060 | -0.378 | -0.003 | 7.05E-01 | 18727 |
| 127 | Thal_VL_L | -0.018 | -0.141 | 0.106 | 0.063 | -0.280 | -0.002 | 7.80E-01 | 18404 | -0.317 | -0.589 | -0.045 | 0.139 | -2.281 | -0.017 | 2.25E-02 | 18727 |
| 128 | Thal_VL_R | 0.058 | -0.106 | 0.221 | 0.083 | 0.692 | 0.005 | 4.89E-01 | 18404 | -0.469 | -0.830 | -0.107 | 0.185 | -2.539 | -0.019 | 1.11E-02 | 18727 |
| 129 | Thal_VPL_L | -0.084 | -0.231 | 0.062 | 0.075 | -1.125 | -0.008 | 2.61E-01 | 18404 | -0.252 | -0.577 | 0.072 | 0.165 | -1.524 | -0.011 | 1.27E-01 | 18727 |

| | | | | | | | | | | | | | | | | | |
|---|---|---|---|---|---|---|---|---|---|---|---|---|---|---|---|---|---|
| 130 | Thal_VPL_R | -0.061 | -0.215 | 0.093 | 0.079 | -0.775 | -0.006 | 4.39E-01 | 18404 | -0.357 | -0.698 | -0.016 | 0.174 | -2.053 | -0.015 | 4.01E-02 | 18727 |
| 131 | Thal_IL_L | -0.001 | -0.064 | 0.063 | 0.032 | -0.019 | 0.000 | 9.84E-01 | 18404 | -0.094 | -0.235 | 0.046 | 0.072 | -1.318 | -0.010 | 1.87E-01 | 18727 |
| 132 | Thal_IL_R | 0.027 | -0.045 | 0.099 | 0.037 | 0.734 | 0.005 | 4.63E-01 | 18404 | -0.131 | -0.291 | 0.028 | 0.081 | -1.619 | -0.012 | 1.06E-01 | 18727 |
| 133 | Thal_Re_L | 0.000 | 0.000 | 0.000 | 0.000 | 1.330 | 0.010 | 1.83E-01 | 18404 | 0.000 | -0.001 | 0.000 | 0.000 | -0.980 | -0.007 | 3.27E-01 | 18727 |
| 134 | Thal_Re_R | 0.002 | 0.000 | 0.004 | 0.001 | 1.826 | 0.013 | 6.79E-02 | 18404 | -0.004 | -0.009 | 0.001 | 0.002 | -1.584 | -0.012 | 1.13E-01 | 18727 |
| 135 | Thal_MDm_L | 0.087 | -0.383 | 0.556 | 0.240 | 0.362 | 0.003 | 7.18E-01 | 18404 | -0.933 | -1.972 | 0.107 | 0.530 | -1.759 | -0.013 | 7.86E-02 | 18727 |
| 136 | Thal_MDm_R | 0.251 | -0.295 | 0.796 | 0.278 | 0.901 | 0.007 | 3.67E-01 | 18404 | -0.910 | -2.118 | 0.298 | 0.616 | -1.476 | -0.011 | 1.40E-01 | 18727 |
| 137 | Thal_MDl_L | 0.003 | -0.078 | 0.084 | 0.041 | 0.067 | 0.000 | 9.46E-01 | 18404 | -0.218 | -0.398 | -0.038 | 0.092 | -2.370 | -0.017 | 1.78E-02 | 18727 |
| 138 | Thal_MDl_R | 0.048 | -0.044 | 0.140 | 0.047 | 1.017 | 0.008 | 3.09E-01 | 18404 | -0.191 | -0.395 | 0.014 | 0.104 | -1.828 | -0.013 | 6.76E-02 | 18727 |
| 139 | Thal_LGN_L | 0.001 | -0.012 | 0.013 | 0.006 | 0.113 | 0.001 | 9.10E-01 | 18404 | -0.019 | -0.047 | 0.008 | 0.014 | -1.394 | -0.010 | 1.63E-01 | 18727 |
| 140 | Thal_LGN_R | 0.000 | -0.026 | 0.026 | 0.013 | -0.003 | 0.000 | 9.98E-01 | 18404 | -0.045 | -0.103 | 0.014 | 0.030 | -1.503 | -0.011 | 1.33E-01 | 18727 |
| 141 | Thal_MGN_L | -0.020 | -0.043 | 0.002 | 0.011 | -1.823 | -0.013 | 6.83E-02 | 18404 | -0.037 | -0.086 | 0.011 | 0.025 | -1.500 | -0.011 | 1.34E-01 | 18727 |
| 142 | Thal_MGN_R | -0.011 | -0.035 | 0.013 | 0.012 | -0.916 | -0.007 | 3.60E-01 | 18404 | -0.045 | -0.098 | 0.008 | 0.027 | -1.667 | -0.012 | 9.55E-02 | 18727 |
| 143 | Thal_PuA_L | -0.015 | -0.041 | 0.011 | 0.013 | -1.137 | -0.008 | 2.55E-01 | 18404 | -0.043 | -0.101 | 0.015 | 0.029 | -1.465 | -0.011 | 1.43E-01 | 18727 |
| 144 | Thal_PuA_R | -0.021 | -0.075 | 0.033 | 0.028 | -0.752 | -0.006 | 4.52E-01 | 18404 | -0.099 | -0.219 | 0.020 | 0.061 | -1.625 | -0.012 | 1.04E-01 | 18727 |
| 145 | Thal_PuM_L | -0.103 | -0.620 | 0.413 | 0.264 | -0.392 | -0.003 | 6.95E-01 | 18404 | -0.862 | -2.006 | 0.282 | 0.583 | -1.477 | -0.011 | 1.40E-01 | 18727 |
| 146 | Thal_PuM_R | -0.107 | -0.751 | 0.537 | 0.328 | -0.326 | -0.002 | 7.44E-01 | 18404 | -1.210 | -2.635 | 0.215 | 0.727 | -1.664 | -0.012 | 9.61E-02 | 18727 |
| 147 | Thal_PuL_L | -0.019 | -0.113 | 0.075 | 0.048 | -0.399 | -0.003 | 6.90E-01 | 18404 | -0.162 | -0.371 | 0.046 | 0.106 | -1.526 | -0.011 | 1.27E-01 | 18727 |
| 148 | Thal_PuL_R | -0.021 | -0.170 | 0.129 | 0.076 | -0.269 | -0.002 | 7.88E-01 | 18404 | -0.276 | -0.608 | 0.056 | 0.169 | -1.628 | -0.012 | 1.03E-01 | 18727 |
| 149 | Thal_Pul_L | -0.008 | -0.033 | 0.018 | 0.013 | -0.603 | -0.004 | 5.47E-01 | 18404 | -0.069 | -0.125 | -0.012 | 0.029 | -2.366 | -0.017 | 1.80E-02 | 18727 |
| 150 | Thal_Pul_R | -0.006 | -0.040 | 0.028 | 0.017 | -0.345 | -0.003 | 7.30E-01 | 18404 | -0.073 | -0.148 | 0.002 | 0.038 | -1.908 | -0.014 | 5.64E-02 | 18727 |
| 151 | ACC_sub_L | -0.313 | -0.609 | -0.017 | 0.151 | -2.072 | -0.015 | 3.82E-02 | 18404 | 0.091 | -0.564 | 0.745 | 0.334 | 0.272 | 0.002 | 7.86E-01 | 18727 |
| 152 | ACC_sub_R | -0.248 | -0.524 | 0.029 | 0.141 | -1.755 | -0.013 | 7.92E-02 | 18404 | 0.157 | -0.454 | 0.769 | 0.312 | 0.504 | 0.004 | 6.14E-01 | 18727 |
| 153 | ACC_pre_L | -0.495 | -1.253 | 0.264 | 0.387 | -1.278 | -0.009 | 2.01E-01 | 18404 | 0.742 | -0.936 | 2.421 | 0.856 | 0.867 | 0.006 | 3.86E-01 | 18727 |
| 154 | ACC_pre_R | -0.412 | -1.142 | 0.319 | 0.373 | -1.104 | -0.008 | 2.70E-01 | 18404 | 0.870 | -0.747 | 2.486 | 0.825 | 1.055 | 0.008 | 2.92E-01 | 18727 |
| 155 | ACC_sup_L | 0.378 | -0.447 | 1.202 | 0.421 | 0.897 | 0.007 | 3.70E-01 | 18404 | -0.034 | -1.858 | 1.790 | 0.931 | -0.036 | 0.000 | 9.71E-01 | 18727 |
| 156 | ACC_sup_R | -0.077 | -0.931 | 0.776 | 0.436 | -0.178 | -0.001 | 8.59E-01 | 18404 | 0.741 | -1.147 | 2.630 | 0.964 | 0.769 | 0.006 | 4.42E-01 | 18727 |
| 157 | N_Acc_L | -0.147 | -0.395 | 0.102 | 0.127 | -1.158 | -0.009 | 2.47E-01 | 18404 | 0.175 | -0.374 | 0.724 | 0.280 | 0.625 | 0.005 | 5.32E-01 | 18727 |
| 158 | N_Acc_R | -0.048 | -0.262 | 0.167 | 0.110 | -0.435 | -0.003 | 6.64E-01 | 18404 | 0.026 | -0.449 | 0.501 | 0.242 | 0.109 | 0.001 | 9.14E-01 | 18727 |
| 159 | VTA_L | 0.004 | -0.004 | 0.013 | 0.004 | 1.006 | 0.007 | 3.14E-01 | 18404 | -0.008 | -0.027 | 0.011 | 0.010 | -0.813 | -0.006 | 4.16E-01 | 18727 |
| 160 | VTA_R | 0.002 | -0.006 | 0.010 | 0.004 | 0.456 | 0.003 | 6.48E-01 | 18404 | -0.007 | -0.024 | 0.010 | 0.009 | -0.798 | -0.006 | 4.25E-01 | 18727 |
| 161 | SN_pc_L | -0.003 | -0.017 | 0.011 | 0.007 | -0.478 | -0.004 | 6.33E-01 | 18404 | -0.030 | -0.061 | 0.001 | 0.016 | -1.897 | -0.014 | 5.78E-02 | 18727 |
| 162 | SN_pc_R | -0.002 | -0.020 | 0.016 | 0.009 | -0.218 | -0.002 | 8.28E-01 | 18404 | -0.033 | -0.074 | 0.007 | 0.021 | -1.606 | -0.012 | 1.08E-01 | 18727 |
| 163 | SN_pr_L | -0.006 | -0.025 | 0.014 | 0.010 | -0.570 | -0.004 | 5.69E-01 | 18404 | -0.066 | -0.108 | -0.024 | 0.021 | -3.090 | -0.023 | 2.00E-03 | 18727 |
| 164 | SN_pr_R | -0.008 | -0.030 | 0.013 | 0.011 | -0.760 | -0.006 | 4.47E-01 | 18404 | -0.045 | -0.093 | 0.002 | 0.024 | -1.878 | -0.014 | 6.04E-02 | 18727 |
| 165 | Red_N_L | 0.017 | -0.006 | 0.041 | 0.012 | 1.443 | 0.011 | 1.49E-01 | 18404 | -0.035 | -0.087 | 0.017 | 0.027 | -1.333 | -0.010 | 1.83E-01 | 18727 |
| 166 | Red_N_R | 0.015 | -0.011 | 0.041 | 0.013 | 1.155 | 0.009 | 2.48E-01 | 18404 | -0.012 | -0.069 | 0.044 | 0.029 | -0.424 | -0.003 | 6.71E-01 | 18727 |
| 167 | LC_L | 0.007 | -0.002 | 0.016 | 0.004 | 1.522 | 0.011 | 1.28E-01 | 18404 | 0.021 | 0.002 | 0.040 | 0.010 | 2.112 | 0.015 | 3.47E-02 | 18727 |
| 168 | LC_R | 0.005 | -0.005 | 0.014 | 0.005 | 0.970 | 0.007 | 3.32E-01 | 18404 | 0.021 | 0.000 | 0.042 | 0.011 | 1.939 | 0.014 | 5.25E-02 | 18727 |
| 169 | Raphe_D | 0.017 | -0.017 | 0.051 | 0.017 | 0.960 | 0.007 | 3.37E-01 | 18404 | -0.011 | -0.086 | 0.065 | 0.038 | -0.275 | -0.002 | 7.84E-01 | 18727 |
| 170 | Raphe_M | 0.006 | -0.004 | 0.017 | 0.005 | 1.215 | 0.009 | 2.25E-01 | 18404 | 0.015 | -0.008 | 0.038 | 0.012 | 1.280 | 0.009 | 2.01E-01 | 18727 |

| | | Fresh fruit intake | | | | | | | | Beef intake | | | | | | | |
|---|---|---|---|---|---|---|---|---|---|---|---|---|---|---|---|---|---|
| **Number** | **AAL 3 region** | **beta** | **95%CI_l** | **95%CI_h** | **se** | **t** | **r** | **p** | **df.e** | **beta** | **95%CI_l** | **95%CI_h** | **se** | **t** | **r** | **p** | **df.e** |
| 0 | Total brain | -27.850 | -146.381 | 90.681 | 60.472 | -0.461 | -0.003 | 6.45E-01 | 18540 | -280.805 | -478.983 | -82.627 | 101.107 | -2.777 | -0.020 | 5.49E-03 | 18702 |
| 1 | Precentral_L | -0.341 | -4.516 | 3.834 | 2.130 | -0.160 | -0.001 | 8.73E-01 | 18540 | -2.654 | -9.636 | 4.328 | 3.562 | -0.745 | -0.005 | 4.56E-01 | 18702 |
| 2 | Precentral_R | 1.090 | -2.737 | 4.917 | 1.952 | 0.558 | 0.004 | 5.77E-01 | 18540 | -4.637 | -11.033 | 1.759 | 3.263 | -1.421 | -0.010 | 1.55E-01 | 18702 |
| 3 | Frontal_Sup_2_L | -0.380 | -4.788 | 4.028 | 2.249 | -0.169 | -0.001 | 8.66E-01 | 18540 | -10.652 | -18.025 | -3.279 | 3.762 | -2.832 | -0.021 | 4.63E-03 | 18702 |
| 4 | Frontal_Sup_2_R | -1.798 | -6.658 | 3.062 | 2.479 | -0.725 | -0.005 | 4.68E-01 | 18540 | -8.667 | -16.793 | -0.542 | 4.145 | -2.091 | -0.015 | 3.66E-02 | 18702 |
| 5 | Frontal_Mid_2_L | -2.141 | -6.771 | 2.489 | 2.362 | -0.906 | -0.007 | 3.65E-01 | 18540 | -6.598 | -14.338 | 1.142 | 3.949 | -1.671 | -0.012 | 9.48E-02 | 18702 |

| | | | | | | | | | | | | | | | | | |
|---|---|---|---|---|---|---|---|---|---|---|---|---|---|---|---|---|---|
| 6 | Frontal_Mid_2_R | -2.790 | -8.024 | 2.444 | 2.670 | -1.045 | -0.008 | 2.96E-01 | 18540 | -8.915 | -17.672 | -0.158 | 4.468 | -1.996 | -0.015 | 4.60E-02 | 18702 |
| 7 | Frontal_Inf_Oper_L | 0.002 | -1.365 | 1.369 | 0.698 | 0.003 | 0.000 | 9.97E-01 | 18540 | -3.395 | -5.682 | -1.109 | 1.167 | -2.911 | -0.021 | 3.61E-03 | 18702 |
| 8 | Frontal_Inf_Oper_R | -0.079 | -1.792 | 1.635 | 0.874 | -0.090 | -0.001 | 9.28E-01 | 18540 | -4.401 | -7.268 | -1.533 | 1.463 | -3.008 | -0.022 | 2.63E-03 | 18702 |
| 9 | Frontal_Inf_Tri_L | -0.762 | -3.665 | 2.141 | 1.481 | -0.514 | -0.004 | 6.07E-01 | 18540 | -5.461 | -10.318 | -0.603 | 2.478 | -2.204 | -0.016 | 2.76E-02 | 18702 |
| 10 | Frontal_Inf_Tri_R | -0.545 | -2.825 | 1.736 | 1.164 | -0.468 | -0.003 | 6.40E-01 | 18540 | -5.326 | -9.140 | -1.512 | 1.946 | -2.737 | -0.020 | 6.20E-03 | 18702 |
| 11 | Frontal_Inf_Orb_2_L | -0.542 | -1.409 | 0.326 | 0.443 | -1.224 | -0.009 | 2.21E-01 | 18540 | -1.705 | -3.157 | -0.252 | 0.741 | -2.300 | -0.017 | 2.14E-02 | 18702 |
| 12 | Frontal_Inf_Orb_2_R | 0.124 | -0.783 | 1.032 | 0.463 | 0.269 | 0.002 | 7.88E-01 | 18540 | -3.002 | -4.521 | -1.484 | 0.775 | -3.875 | -0.028 | 1.07E-04 | 18702 |
| 13 | Rolandic_Oper_L | -1.521 | -2.835 | -0.208 | 0.670 | -2.271 | -0.017 | 2.32E-02 | 18540 | -0.828 | -3.024 | 1.369 | 1.121 | -0.739 | -0.005 | 4.60E-01 | 18702 |
| 14 | Rolandic_Oper_R | -0.018 | -1.896 | 1.861 | 0.958 | -0.018 | 0.000 | 9.85E-01 | 18540 | -4.920 | -8.063 | -1.777 | 1.603 | -3.068 | -0.022 | 2.16E-03 | 18702 |
| 15 | Supp_Motor_Area_L | 1.348 | -1.777 | 4.472 | 1.594 | 0.845 | 0.006 | 3.98E-01 | 18540 | -1.583 | -6.806 | 3.640 | 2.665 | -0.594 | -0.004 | 5.53E-01 | 18702 |
| 16 | Supp_Motor_Area_R | 1.575 | -1.876 | 5.027 | 1.761 | 0.895 | 0.007 | 3.71E-01 | 18540 | -1.443 | -7.214 | 4.327 | 2.944 | -0.490 | -0.004 | 6.24E-01 | 18702 |
| 17 | Olfactory_L | -0.342 | -0.805 | 0.122 | 0.237 | -1.445 | -0.011 | 1.48E-01 | 18540 | -0.395 | -1.171 | 0.381 | 0.396 | -0.997 | -0.007 | 3.19E-01 | 18702 |
| 18 | Olfactory_R | -0.370 | -0.835 | 0.095 | 0.237 | -1.561 | -0.011 | 1.19E-01 | 18540 | -0.436 | -1.214 | 0.342 | 0.397 | -1.098 | -0.008 | 2.72E-01 | 18702 |
| 19 | Frontal_Sup_Medial_L | 0.355 | -2.572 | 3.282 | 1.493 | 0.238 | 0.002 | 8.12E-01 | 18540 | -6.769 | -11.660 | -1.877 | 2.496 | -2.712 | -0.020 | 6.69E-03 | 18702 |
| 20 | Frontal_Sup_Medial_R | 0.194 | -1.937 | 2.324 | 1.087 | 0.178 | 0.001 | 8.59E-01 | 18540 | -6.187 | -9.747 | -2.627 | 1.816 | -3.407 | -0.025 | 6.59E-04 | 18702 |
| 21 | Frontal_Med_Orb_L | -0.081 | -1.174 | 1.012 | 0.558 | -0.145 | -0.001 | 8.84E-01 | 18540 | -2.294 | -4.123 | -0.466 | 0.933 | -2.460 | -0.018 | 1.39E-02 | 18702 |
| 22 | Frontal_Med_Orb_R | 0.267 | -1.076 | 1.609 | 0.685 | 0.389 | 0.003 | 6.97E-01 | 18540 | -2.319 | -4.566 | -0.072 | 1.146 | -2.023 | -0.015 | 4.31E-02 | 18702 |
| 23 | Rectus_L | -0.099 | -1.230 | 1.031 | 0.577 | -0.172 | -0.001 | 8.63E-01 | 18540 | -1.790 | -3.682 | 0.102 | 0.965 | -1.855 | -0.014 | 6.37E-02 | 18702 |
| 24 | Rectus_R | -0.140 | -1.186 | 0.906 | 0.534 | -0.262 | -0.002 | 7.93E-01 | 18540 | -2.066 | -3.816 | -0.316 | 0.893 | -2.314 | -0.017 | 2.07E-02 | 18702 |
| 25 | OFCmed_L | -0.212 | -0.867 | 0.444 | 0.334 | -0.633 | -0.005 | 5.27E-01 | 18540 | -1.229 | -2.326 | -0.133 | 0.560 | -2.197 | -0.016 | 2.80E-02 | 18702 |
| 26 | OFCmed_R | -0.383 | -1.147 | 0.380 | 0.390 | -0.984 | -0.007 | 3.25E-01 | 18540 | -1.545 | -2.823 | -0.267 | 0.652 | -2.370 | -0.017 | 1.78E-02 | 18702 |
| 27 | OFCant_L | -0.525 | -1.283 | 0.233 | 0.387 | -1.358 | -0.010 | 1.74E-01 | 18540 | -1.241 | -2.509 | 0.028 | 0.647 | -1.917 | -0.014 | 5.52E-02 | 18702 |
| 28 | OFCant_R | -1.061 | -2.062 | -0.060 | 0.511 | -2.077 | -0.015 | 3.78E-02 | 18540 | -1.859 | -3.534 | -0.184 | 0.855 | -2.175 | -0.016 | 2.96E-02 | 18702 |
| 29 | OFCpost_L | -0.548 | -1.496 | 0.401 | 0.484 | -1.132 | -0.008 | 2.58E-01 | 18540 | -1.680 | -3.269 | -0.092 | 0.810 | -2.074 | -0.015 | 3.81E-02 | 18702 |
| 30 | OFCpost_R | -0.499 | -1.376 | 0.378 | 0.447 | -1.116 | -0.008 | 2.65E-01 | 18540 | -2.236 | -3.704 | -0.769 | 0.749 | -2.987 | -0.022 | 2.82E-03 | 18702 |
| 31 | OFClat_L | -0.325 | -0.645 | -0.005 | 0.163 | -1.994 | -0.015 | 4.62E-02 | 18540 | -0.865 | -1.400 | -0.329 | 0.273 | -3.163 | -0.023 | 1.56E-03 | 18702 |
| 32 | OFClat_R | -0.100 | -0.371 | 0.170 | 0.138 | -0.725 | -0.005 | 4.68E-01 | 18540 | -0.989 | -1.441 | -0.537 | 0.231 | -4.285 | -0.031 | 1.83E-05 | 18702 |
| 33 | Insula_L | -2.680 | -5.611 | 0.252 | 1.496 | -1.792 | -0.013 | 7.32E-02 | 18540 | -5.023 | -9.933 | -0.114 | 2.505 | -2.006 | -0.015 | 4.49E-02 | 18702 |
| 34 | Insula_R | -2.306 | -5.163 | 0.551 | 1.458 | -1.582 | -0.012 | 1.14E-01 | 18540 | -8.027 | -12.810 | -3.244 | 2.440 | -3.290 | -0.024 | 1.00E-03 | 18702 |
| 37 | Cingulate_Mid_L | 1.603 | -0.879 | 4.084 | 1.266 | 1.266 | 0.009 | 2.06E-01 | 18540 | -5.319 | -9.466 | -1.172 | 2.116 | -2.514 | -0.018 | 1.20E-02 | 18702 |
| 38 | Cingulate_Mid_R | 0.766 | -1.879 | 3.411 | 1.350 | 0.568 | 0.004 | 5.70E-01 | 18540 | -5.196 | -9.617 | -0.774 | 2.256 | -2.303 | -0.017 | 2.13E-02 | 18702 |
| 39 | Cingulate_Post_L | 0.084 | -0.675 | 0.843 | 0.387 | 0.217 | 0.002 | 8.28E-01 | 18540 | -1.530 | -2.798 | -0.262 | 0.647 | -2.366 | -0.017 | 1.80E-02 | 18702 |
| 40 | Cingulate_Post_R | -0.138 | -0.559 | 0.283 | 0.215 | -0.642 | -0.005 | 5.21E-01 | 18540 | -0.613 | -1.316 | 0.090 | 0.359 | -1.708 | -0.012 | 8.76E-02 | 18702 |
| 41 | Hippocampus_L | -0.028 | -1.255 | 1.199 | 0.626 | -0.045 | 0.000 | 9.64E-01 | 18540 | -1.978 | -4.030 | 0.073 | 1.047 | -1.890 | -0.014 | 5.88E-02 | 18702 |
| 42 | Hippocampus_R | 0.107 | -1.164 | 1.379 | 0.649 | 0.166 | 0.001 | 8.68E-01 | 18540 | -2.339 | -4.466 | -0.212 | 1.085 | -2.155 | -0.016 | 3.11E-02 | 18702 |
| 43 | ParaHippocampal_L | 0.272 | -0.818 | 1.362 | 0.556 | 0.489 | 0.004 | 6.25E-01 | 18540 | -1.304 | -3.126 | 0.519 | 0.930 | -1.402 | -0.010 | 1.61E-01 | 18702 |
| 44 | ParaHippocampal_R | -0.410 | -1.784 | 0.965 | 0.701 | -0.584 | -0.004 | 5.59E-01 | 18540 | -2.234 | -4.533 | 0.065 | 1.173 | -1.905 | -0.014 | 5.68E-02 | 18702 |
| 45 | Amygdala_L | 0.017 | -0.406 | 0.441 | 0.216 | 0.080 | 0.001 | 9.36E-01 | 18540 | -0.633 | -1.342 | 0.076 | 0.362 | -1.751 | -0.013 | 8.00E-02 | 18702 |
| 46 | Amygdala_R | -0.022 | -0.455 | 0.411 | 0.221 | -0.098 | -0.001 | 9.22E-01 | 18540 | -1.127 | -1.852 | -0.403 | 0.370 | -3.051 | -0.022 | 2.28E-03 | 18702 |
| 47 | Calcarine_L | -0.265 | -4.311 | 3.782 | 2.064 | -0.128 | -0.001 | 8.98E-01 | 18540 | 4.125 | -2.637 | 10.888 | 3.450 | 1.196 | 0.009 | 2.32E-01 | 18702 |
| 48 | Calcarine_R | -0.471 | -3.850 | 2.908 | 1.724 | -0.273 | -0.002 | 7.85E-01 | 18540 | 3.888 | -1.761 | 9.538 | 2.882 | 1.349 | 0.010 | 1.77E-01 | 18702 |
| 49 | Cuneus_L | 2.271 | -0.266 | 4.808 | 1.294 | 1.755 | 0.013 | 7.93E-02 | 18540 | -0.178 | -4.418 | 4.062 | 2.163 | -0.082 | -0.001 | 9.34E-01 | 18702 |
| 50 | Cuneus_R | -0.648 | -2.587 | 1.291 | 0.989 | -0.655 | -0.005 | 5.12E-01 | 18540 | 0.434 | -2.810 | 3.677 | 1.655 | 0.262 | 0.002 | 7.93E-01 | 18702 |
| 51 | Lingual_L | -0.741 | -3.174 | 1.691 | 1.241 | -0.597 | -0.004 | 5.50E-01 | 18540 | 0.381 | -3.685 | 4.446 | 2.074 | 0.183 | 0.001 | 8.54E-01 | 18702 |
| 52 | Lingual_R | -0.992 | -3.540 | 1.556 | 1.300 | -0.763 | -0.006 | 4.45E-01 | 18540 | 0.756 | -3.502 | 5.014 | 2.172 | 0.348 | 0.003 | 7.28E-01 | 18702 |
| 53 | Occipital_Sup_L | 0.591 | -1.080 | 2.262 | 0.853 | 0.693 | 0.005 | 4.88E-01 | 18540 | -3.812 | -6.606 | -1.018 | 1.425 | -2.674 | -0.020 | 7.50E-03 | 18702 |
| 54 | Occipital_Sup_R | -0.187 | -1.999 | 1.625 | 0.924 | -0.202 | -0.001 | 8.40E-01 | 18540 | -3.070 | -6.097 | -0.043 | 1.544 | -1.988 | -0.015 | 4.68E-02 | 18702 |
| 55 | Occipital_Mid_L | -1.141 | -5.056 | 2.773 | 1.997 | -0.571 | -0.004 | 5.68E-01 | 18540 | -4.205 | -10.754 | 2.345 | 3.341 | -1.258 | -0.009 | 2.08E-01 | 18702 |
| 56 | Occipital_Mid_R | 0.382 | -2.344 | 3.108 | 1.391 | 0.275 | 0.002 | 7.84E-01 | 18540 | -3.744 | -8.301 | 0.812 | 2.325 | -1.611 | -0.012 | 1.07E-01 | 18702 |

| | | | | | | | | | | | | | | | | | |
|---|---|---|---|---|---|---|---|---|---|---|---|---|---|---|---|---|---|
| 57 | Occipital_Inf_L | -1.529 | -2.829 | -0.228 | 0.664 | -2.304 | -0.017 | 2.13E-02 | 18540 | 0.800 | -1.376 | 2.975 | 1.110 | 0.720 | 0.005 | 4.71E-01 | 18702 |
| 58 | Occipital_Inf_R | -1.540 | -3.043 | -0.036 | 0.767 | -2.008 | -0.015 | 4.47E-02 | 18540 | 0.992 | -1.519 | 3.504 | 1.281 | 0.775 | 0.006 | 4.39E-01 | 18702 |
| 59 | Fusiform_L | 0.632 | -2.212 | 3.476 | 1.451 | 0.436 | 0.003 | 6.63E-01 | 18540 | 2.343 | -2.416 | 7.102 | 2.428 | 0.965 | 0.007 | 3.35E-01 | 18702 |
| 60 | Fusiform_R | 0.883 | -2.242 | 4.009 | 1.594 | 0.554 | 0.004 | 5.80E-01 | 18540 | -1.406 | -6.635 | 3.822 | 2.667 | -0.527 | -0.004 | 5.98E-01 | 18702 |
| 61 | Postcentral_L | 7.158 | 2.878 | 11.438 | 2.184 | 3.278 | 0.024 | 1.05E-03 | 18540 | -2.173 | -9.329 | 4.983 | 3.651 | -0.595 | -0.004 | 5.52E-01 | 18702 |
| 62 | Postcentral_R | 7.793 | 3.406 | 12.179 | 2.238 | 3.482 | 0.026 | 4.98E-04 | 18540 | -0.334 | -7.671 | 7.003 | 3.743 | -0.089 | -0.001 | 9.29E-01 | 18702 |
| 63 | Parietal_Sup_L | -0.650 | -3.117 | 1.817 | 1.259 | -0.516 | -0.004 | 6.06E-01 | 18540 | -4.734 | -8.859 | -0.610 | 2.104 | -2.250 | -0.016 | 2.45E-02 | 18702 |
| 64 | Parietal_Sup_R | 0.157 | -2.033 | 2.347 | 1.117 | 0.141 | 0.001 | 8.88E-01 | 18540 | -2.269 | -5.928 | 1.390 | 1.867 | -1.216 | -0.009 | 2.24E-01 | 18702 |
| 65 | Parietal_Inf_L | 2.066 | -1.166 | 5.298 | 1.649 | 1.253 | 0.009 | 2.10E-01 | 18540 | -2.812 | -8.210 | 2.587 | 2.754 | -1.021 | -0.007 | 3.07E-01 | 18702 |
| 66 | Parietal_Inf_R | 2.456 | 0.363 | 4.548 | 1.068 | 2.300 | 0.017 | 2.14E-02 | 18540 | -2.300 | -5.798 | 1.198 | 1.784 | -1.289 | -0.009 | 1.97E-01 | 18702 |
| 67 | SupraMarginal_L | 1.201 | -0.693 | 3.094 | 0.966 | 1.243 | 0.009 | 2.14E-01 | 18540 | -3.785 | -6.949 | -0.622 | 1.614 | -2.345 | -0.017 | 1.90E-02 | 18702 |
| 68 | SupraMarginal_R | 4.123 | 1.445 | 6.801 | 1.366 | 3.018 | 0.022 | 2.55E-03 | 18540 | -7.421 | -11.897 | -2.945 | 2.284 | -3.250 | -0.024 | 1.16E-03 | 18702 |
| 69 | Angular_L | 0.291 | -1.570 | 2.152 | 0.949 | 0.306 | 0.002 | 7.59E-01 | 18540 | -0.298 | -3.407 | 2.811 | 1.586 | -0.188 | -0.001 | 8.51E-01 | 18702 |
| 70 | Angular_R | 0.878 | -1.617 | 3.372 | 1.273 | 0.690 | 0.005 | 4.90E-01 | 18540 | -1.244 | -5.412 | 2.924 | 2.127 | -0.585 | -0.004 | 5.59E-01 | 18702 |
| 71 | Precuneus_L | 3.558 | -0.971 | 8.087 | 2.311 | 1.540 | 0.011 | 1.24E-01 | 18540 | -3.911 | -11.485 | 3.664 | 3.864 | -1.012 | -0.007 | 3.12E-01 | 18702 |
| 72 | Precuneus_R | -0.044 | -4.081 | 3.993 | 2.059 | -0.021 | 0.000 | 9.83E-01 | 18540 | -0.838 | -7.585 | 5.908 | 3.442 | -0.244 | -0.002 | 8.08E-01 | 18702 |
| 73 | Paracentral_Lobule_L | 2.093 | 0.018 | 4.169 | 1.059 | 1.977 | 0.015 | 4.80E-02 | 18540 | -1.015 | -4.485 | 2.456 | 1.771 | -0.573 | -0.004 | 5.67E-01 | 18702 |
| 74 | Paracentral_Lobule_R | 0.136 | -1.350 | 1.622 | 0.758 | 0.180 | 0.001 | 8.57E-01 | 18540 | -0.566 | -3.051 | 1.919 | 1.268 | -0.446 | -0.003 | 6.55E-01 | 18702 |
| 75 | Caudate_L | -0.239 | -1.895 | 1.417 | 0.845 | -0.283 | -0.002 | 7.77E-01 | 18540 | -3.534 | -6.307 | -0.761 | 1.415 | -2.498 | -0.018 | 1.25E-02 | 18702 |
| 76 | Caudate_R | -1.051 | -2.925 | 0.823 | 0.956 | -1.099 | -0.008 | 2.72E-01 | 18540 | -3.312 | -6.450 | -0.173 | 1.601 | -2.068 | -0.015 | 3.86E-02 | 18702 |
| 77 | Putamen_L | -2.249 | -4.762 | 0.264 | 1.282 | -1.754 | -0.013 | 7.94E-02 | 18540 | -6.989 | -11.197 | -2.781 | 2.147 | -3.255 | -0.024 | 1.13E-03 | 18702 |
| 78 | Putamen_R | -2.244 | -4.803 | 0.316 | 1.306 | -1.718 | -0.013 | 8.58E-02 | 18540 | -6.746 | -11.032 | -2.461 | 2.186 | -3.086 | -0.023 | 2.03E-03 | 18702 |
| 79 | Pallidum_L | -0.128 | -0.352 | 0.096 | 0.114 | -1.122 | -0.008 | 2.62E-01 | 18540 | -0.647 | -1.021 | -0.273 | 0.191 | -3.389 | -0.025 | 7.02E-04 | 18702 |
| 80 | Pallidum_R | -0.170 | -0.415 | 0.076 | 0.125 | -1.356 | -0.010 | 1.75E-01 | 18540 | -0.731 | -1.142 | -0.321 | 0.209 | -3.490 | -0.026 | 4.84E-04 | 18702 |
| 83 | Heschl_L | -0.439 | -0.893 | 0.014 | 0.231 | -1.898 | -0.014 | 5.78E-02 | 18540 | -0.676 | -1.434 | 0.083 | 0.387 | -1.746 | -0.013 | 8.09E-02 | 18702 |
| 84 | Heschl_R | -0.036 | -0.542 | 0.471 | 0.259 | -0.138 | -0.001 | 8.91E-01 | 18540 | -1.617 | -2.464 | -0.770 | 0.432 | -3.740 | -0.027 | 1.84E-04 | 18702 |
| 85 | Temporal_Sup_L | -1.746 | -5.019 | 1.527 | 1.670 | -1.046 | -0.008 | 2.96E-01 | 18540 | -6.283 | -11.759 | -0.807 | 2.794 | -2.249 | -0.016 | 2.45E-02 | 18702 |
| 86 | Temporal_Sup_R | 0.903 | -2.719 | 4.524 | 1.848 | 0.488 | 0.004 | 6.25E-01 | 18540 | -11.303 | -17.359 | -5.247 | 3.090 | -3.658 | -0.027 | 2.54E-04 | 18702 |
| 87 | Temporal_Pole_Sup_L | -0.878 | -2.428 | 0.673 | 0.791 | -1.110 | -0.008 | 2.67E-01 | 18540 | -2.637 | -5.231 | -0.042 | 1.324 | -1.992 | -0.015 | 4.64E-02 | 18702 |
| 88 | Temporal_Pole_Sup_R | -1.062 | -2.694 | 0.570 | 0.833 | -1.275 | -0.009 | 2.02E-01 | 18540 | -3.390 | -6.120 | -0.659 | 1.393 | -2.433 | -0.018 | 1.50E-02 | 18702 |
| 89 | Temporal_Mid_L | -0.509 | -5.968 | 4.950 | 2.785 | -0.183 | -0.001 | 8.55E-01 | 18540 | -11.673 | -20.797 | -2.549 | 4.655 | -2.508 | -0.018 | 1.22E-02 | 18702 |
| 90 | Temporal_Mid_R | -0.953 | -5.802 | 3.895 | 2.474 | -0.385 | -0.003 | 7.00E-01 | 18540 | -3.769 | -11.871 | 4.334 | 4.134 | -0.912 | -0.007 | 3.62E-01 | 18702 |
| 91 | Temporal_Pole_Mid_L | -0.028 | -1.332 | 1.276 | 0.665 | -0.042 | 0.000 | 9.66E-01 | 18540 | 0.139 | -2.042 | 2.320 | 1.113 | 0.125 | 0.001 | 9.01E-01 | 18702 |
| 92 | Temporal_Pole_Mid_R | -0.923 | -2.506 | 0.660 | 0.807 | -1.143 | -0.008 | 2.53E-01 | 18540 | -1.418 | -4.068 | 1.232 | 1.352 | -1.049 | -0.008 | 2.94E-01 | 18702 |
| 93 | Temporal_Inf_L | -1.773 | -5.522 | 1.976 | 1.913 | -0.927 | -0.007 | 3.54E-01 | 18540 | -4.110 | -10.381 | 2.162 | 3.200 | -1.284 | -0.009 | 1.99E-01 | 18702 |
| 94 | Temporal_Inf_R | -1.252 | -5.341 | 2.838 | 2.086 | -0.600 | -0.004 | 5.49E-01 | 18540 | -1.658 | -8.496 | 5.179 | 3.488 | -0.475 | -0.003 | 6.34E-01 | 18702 |
| 95 | Cerebellum_Crus1_L | -0.431 | -4.887 | 4.025 | 2.273 | -0.190 | -0.001 | 8.49E-01 | 18540 | 0.983 | -6.468 | 8.435 | 3.802 | 0.259 | 0.002 | 7.96E-01 | 18702 |
| 96 | Cerebellum_Crus1_R | -2.409 | -6.955 | 2.137 | 2.319 | -1.039 | -0.008 | 2.99E-01 | 18540 | -1.761 | -9.361 | 5.839 | 3.877 | -0.454 | -0.003 | 6.50E-01 | 18702 |
| 97 | Cerebellum_Crus2_L | -2.610 | -6.144 | 0.924 | 1.803 | -1.447 | -0.011 | 1.48E-01 | 18540 | -3.869 | -9.780 | 2.043 | 3.016 | -1.283 | -0.009 | 2.00E-01 | 18702 |
| 98 | Cerebellum_Crus2_R | -2.248 | -5.515 | 1.019 | 1.667 | -1.349 | -0.010 | 1.77E-01 | 18540 | -3.750 | -9.218 | 1.717 | 2.789 | -1.344 | -0.010 | 1.79E-01 | 18702 |
| 99 | Cerebellum_3_L | -0.147 | -0.384 | 0.090 | 0.121 | -1.215 | -0.009 | 2.24E-01 | 18540 | 0.756 | 0.359 | 1.153 | 0.202 | 3.735 | 0.027 | 1.88E-04 | 18702 |
| 100 | Cerebellum_3_R | -0.082 | -0.402 | 0.239 | 0.163 | -0.500 | -0.004 | 6.17E-01 | 18540 | 0.695 | 0.159 | 1.230 | 0.273 | 2.542 | 0.019 | 1.10E-02 | 18702 |
| 101 | Cerebellum_4_5_L | -0.455 | -2.374 | 1.465 | 0.979 | -0.464 | -0.003 | 6.42E-01 | 18540 | 2.846 | -0.364 | 6.056 | 1.638 | 1.738 | 0.013 | 8.22E-02 | 18702 |
| 102 | Cerebellum_4_5_R | 0.540 | -1.024 | 2.104 | 0.798 | 0.677 | 0.005 | 4.99E-01 | 18540 | 1.963 | -0.653 | 4.579 | 1.335 | 1.471 | 0.011 | 1.41E-01 | 18702 |
| 103 | Cerebellum_6_L | 1.022 | -2.294 | 4.337 | 1.692 | 0.604 | 0.004 | 5.46E-01 | 18540 | 0.582 | -4.965 | 6.130 | 2.830 | 0.206 | 0.002 | 8.37E-01 | 18702 |
| 104 | Cerebellum_6_R | 0.854 | -2.641 | 4.348 | 1.783 | 0.479 | 0.004 | 6.32E-01 | 18540 | -0.046 | -5.890 | 5.799 | 2.982 | -0.015 | 0.000 | 9.88E-01 | 18702 |
| 105 | Cerebellum_7b_L | -0.956 | -2.036 | 0.124 | 0.551 | -1.735 | -0.013 | 8.27E-02 | 18540 | -1.338 | -3.144 | 0.468 | 0.921 | -1.453 | -0.011 | 1.46E-01 | 18702 |
| 106 | Cerebellum_7b_R | -0.929 | -2.000 | 0.142 | 0.546 | -1.701 | -0.012 | 8.90E-02 | 18540 | -1.995 | -3.785 | -0.204 | 0.913 | -2.184 | -0.016 | 2.90E-02 | 18702 |
| 107 | Cerebellum_8_L | -2.983 | -6.415 | 0.448 | 1.751 | -1.704 | -0.013 | 8.84E-02 | 18540 | -2.486 | -8.222 | 3.250 | 2.926 | -0.849 | -0.006 | 3.96E-01 | 18702 |

| | | | | | | | | | | | | | | | | | |
|---|---|---|---|---|---|---|---|---|---|---|---|---|---|---|---|---|---|
| 108 | Cerebellum_8_R | -5.079 | -9.544 | -0.614 | 2.278 | -2.230 | -0.016 | 2.58E-02 | 18540 | -7.227 | -14.693 | 0.239 | 3.809 | -1.897 | -0.014 | 5.78E-02 | 18702 |
| 109 | Cerebellum_9_L | -2.067 | -4.007 | -0.127 | 0.990 | -2.088 | -0.015 | 3.68E-02 | 18540 | -0.035 | -3.280 | 3.209 | 1.655 | -0.021 | 0.000 | 9.83E-01 | 18702 |
| 110 | Cerebellum_9_R | -2.316 | -4.223 | -0.408 | 0.973 | -2.379 | -0.017 | 1.74E-02 | 18540 | 0.576 | -2.613 | 3.765 | 1.627 | 0.354 | 0.003 | 7.23E-01 | 18702 |
| 111 | Cerebellum_10_L | 0.001 | -0.137 | 0.139 | 0.070 | 0.012 | 0.000 | 9.90E-01 | 18540 | 0.024 | -0.207 | 0.254 | 0.117 | 0.201 | 0.001 | 8.41E-01 | 18702 |
| 112 | Cerebellum_10_R | 0.061 | -0.082 | 0.204 | 0.073 | 0.833 | 0.006 | 4.05E-01 | 18540 | 0.156 | -0.083 | 0.395 | 0.122 | 1.277 | 0.009 | 2.02E-01 | 18702 |
| 113 | Vermis_1_2 | -0.098 | -0.188 | -0.009 | 0.046 | -2.158 | -0.016 | 3.09E-02 | 18540 | 0.202 | 0.052 | 0.352 | 0.076 | 2.647 | 0.019 | 8.12E-03 | 18702 |
| 114 | Vermis_3 | -0.415 | -0.770 | -0.060 | 0.181 | -2.290 | -0.017 | 2.20E-02 | 18540 | 0.622 | 0.028 | 1.217 | 0.303 | 2.051 | 0.015 | 4.02E-02 | 18702 |
| 115 | Vermis_4_5 | -0.753 | -1.798 | 0.292 | 0.533 | -1.412 | -0.010 | 1.58E-01 | 18540 | 0.976 | -0.773 | 2.725 | 0.892 | 1.094 | 0.008 | 2.74E-01 | 18702 |
| 116 | Vermis_6 | -0.280 | -0.862 | 0.302 | 0.297 | -0.944 | -0.007 | 3.45E-01 | 18540 | 0.319 | -0.654 | 1.293 | 0.497 | 0.643 | 0.005 | 5.20E-01 | 18702 |
| 117 | Vermis_7 | -0.239 | -0.568 | 0.090 | 0.168 | -1.425 | -0.010 | 1.54E-01 | 18540 | -0.191 | -0.742 | 0.359 | 0.281 | -0.682 | -0.005 | 4.95E-01 | 18702 |
| 118 | Vermis_8 | -0.174 | -0.715 | 0.367 | 0.276 | -0.631 | -0.005 | 5.28E-01 | 18540 | 0.001 | -0.903 | 0.905 | 0.461 | 0.002 | 0.000 | 9.98E-01 | 18702 |
| 119 | Vermis_9 | -0.288 | -0.727 | 0.150 | 0.224 | -1.290 | -0.009 | 1.97E-01 | 18540 | 0.178 | -0.555 | 0.911 | 0.374 | 0.476 | 0.003 | 6.34E-01 | 18702 |
| 120 | Vermis_10 | -0.143 | -0.266 | -0.021 | 0.062 | -2.301 | -0.017 | 2.14E-02 | 18540 | 0.244 | 0.040 | 0.448 | 0.104 | 2.343 | 0.017 | 1.92E-02 | 18702 |
| 121 | Thal_AV_L | 0.019 | -0.030 | 0.068 | 0.025 | 0.747 | 0.005 | 4.55E-01 | 18540 | -0.065 | -0.147 | 0.017 | 0.042 | -1.557 | -0.011 | 1.20E-01 | 18702 |
| 122 | Thal_AV_R | -0.032 | -0.102 | 0.038 | 0.036 | -0.890 | -0.007 | 3.74E-01 | 18540 | -0.049 | -0.166 | 0.068 | 0.060 | -0.819 | -0.006 | 4.13E-01 | 18702 |
| 123 | Thal_LP_L | 0.015 | -0.040 | 0.069 | 0.028 | 0.527 | 0.004 | 5.99E-01 | 18540 | -0.085 | -0.176 | 0.006 | 0.046 | -1.841 | -0.013 | 6.56E-02 | 18702 |
| 124 | Thal_LP_R | -0.025 | -0.096 | 0.045 | 0.036 | -0.707 | -0.005 | 4.79E-01 | 18540 | -0.078 | -0.196 | 0.040 | 0.060 | -1.296 | -0.009 | 1.95E-01 | 18702 |
| 125 | Thal_VA_L | 0.018 | -0.040 | 0.076 | 0.030 | 0.613 | 0.004 | 5.40E-01 | 18540 | -0.050 | -0.147 | 0.047 | 0.049 | -1.012 | -0.007 | 3.12E-01 | 18702 |
| 126 | Thal_VA_R | -0.007 | -0.078 | 0.064 | 0.036 | -0.195 | -0.001 | 8.46E-01 | 18540 | -0.015 | -0.134 | 0.104 | 0.061 | -0.245 | -0.002 | 8.07E-01 | 18702 |
| 127 | Thal_VL_L | 0.056 | -0.108 | 0.221 | 0.084 | 0.673 | 0.005 | 5.01E-01 | 18540 | -0.257 | -0.532 | 0.018 | 0.140 | -1.830 | -0.013 | 6.72E-02 | 18702 |
| 128 | Thal_VL_R | -0.050 | -0.269 | 0.169 | 0.112 | -0.448 | -0.003 | 6.54E-01 | 18540 | -0.270 | -0.636 | 0.096 | 0.187 | -1.448 | -0.011 | 1.48E-01 | 18702 |
| 129 | Thal_VPL_L | -0.075 | -0.271 | 0.121 | 0.100 | -0.750 | -0.006 | 4.53E-01 | 18540 | -0.218 | -0.546 | 0.110 | 0.167 | -1.303 | -0.010 | 1.93E-01 | 18702 |
| 130 | Thal_VPL_R | -0.050 | -0.256 | 0.156 | 0.105 | -0.474 | -0.003 | 6.36E-01 | 18540 | -0.397 | -0.741 | -0.052 | 0.176 | -2.257 | -0.016 | 2.40E-02 | 18702 |
| 131 | Thal_IL_L | -0.019 | -0.104 | 0.066 | 0.043 | -0.436 | -0.003 | 6.63E-01 | 18540 | -0.224 | -0.365 | -0.082 | 0.072 | -3.095 | -0.023 | 1.97E-03 | 18702 |
| 132 | Thal_IL_R | -0.034 | -0.130 | 0.062 | 0.049 | -0.691 | -0.005 | 4.89E-01 | 18540 | -0.254 | -0.415 | -0.093 | 0.082 | -3.097 | -0.023 | 1.96E-03 | 18702 |
| 133 | Thal_Re_L | 0.000 | 0.000 | 0.000 | 0.000 | -0.253 | -0.002 | 8.00E-01 | 18540 | 0.000 | -0.001 | 0.001 | 0.000 | 0.337 | 0.002 | 7.36E-01 | 18702 |
| 134 | Thal_Re_R | -0.002 | -0.004 | 0.001 | 0.001 | -1.102 | -0.008 | 2.71E-01 | 18540 | -0.001 | -0.006 | 0.004 | 0.002 | -0.519 | -0.004 | 6.03E-01 | 18702 |
| 135 | Thal_MDm_L | -0.350 | -0.978 | 0.278 | 0.320 | -1.094 | -0.008 | 2.74E-01 | 18540 | -1.795 | -2.844 | -0.745 | 0.536 | -3.351 | -0.024 | 8.08E-04 | 18702 |
| 136 | Thal_MDm_R | -0.510 | -1.240 | 0.220 | 0.372 | -1.370 | -0.010 | 1.71E-01 | 18540 | -1.871 | -3.091 | -0.650 | 0.623 | -3.004 | -0.022 | 2.66E-03 | 18702 |
| 137 | Thal_MDl_L | -0.012 | -0.120 | 0.097 | 0.055 | -0.209 | -0.002 | 8.35E-01 | 18540 | -0.305 | -0.486 | -0.123 | 0.093 | -3.287 | -0.024 | 1.02E-03 | 18702 |
| 138 | Thal_MDl_R | -0.053 | -0.177 | 0.070 | 0.063 | -0.847 | -0.006 | 3.97E-01 | 18540 | -0.310 | -0.517 | -0.103 | 0.105 | -2.942 | -0.022 | 3.27E-03 | 18702 |
| 139 | Thal_LGN_L | -0.004 | -0.020 | 0.013 | 0.008 | -0.447 | -0.003 | 6.55E-01 | 18540 | -0.025 | -0.053 | 0.002 | 0.014 | -1.791 | -0.013 | 7.33E-02 | 18702 |
| 140 | Thal_LGN_R | -0.019 | -0.054 | 0.016 | 0.018 | -1.049 | -0.008 | 2.94E-01 | 18540 | -0.070 | -0.128 | -0.011 | 0.030 | -2.328 | -0.017 | 1.99E-02 | 18702 |
| 141 | Thal_MGN_L | -0.025 | -0.055 | 0.004 | 0.015 | -1.688 | -0.012 | 9.15E-02 | 18540 | -0.072 | -0.121 | -0.023 | 0.025 | -2.860 | -0.021 | 4.24E-03 | 18702 |
| 142 | Thal_MGN_R | -0.024 | -0.056 | 0.008 | 0.016 | -1.465 | -0.011 | 1.43E-01 | 18540 | -0.100 | -0.154 | -0.047 | 0.027 | -3.664 | -0.027 | 2.49E-04 | 18702 |
| 143 | Thal_PuA_L | -0.028 | -0.063 | 0.007 | 0.018 | -1.567 | -0.012 | 1.17E-01 | 18540 | -0.085 | -0.144 | -0.027 | 0.030 | -2.880 | -0.021 | 3.98E-03 | 18702 |
| 144 | Thal_PuA_R | -0.043 | -0.115 | 0.029 | 0.037 | -1.163 | -0.009 | 2.45E-01 | 18540 | -0.225 | -0.346 | -0.105 | 0.062 | -3.654 | -0.027 | 2.59E-04 | 18702 |
| 145 | Thal_PuM_L | -0.514 | -1.205 | 0.177 | 0.353 | -1.457 | -0.011 | 1.45E-01 | 18540 | -1.667 | -2.823 | -0.511 | 0.590 | -2.827 | -0.021 | 4.70E-03 | 18702 |
| 146 | Thal_PuM_R | -0.377 | -1.238 | 0.485 | 0.440 | -0.857 | -0.006 | 3.92E-01 | 18540 | -2.305 | -3.746 | -0.864 | 0.735 | -3.136 | -0.023 | 1.71E-03 | 18702 |
| 147 | Thal_PuL_L | -0.053 | -0.179 | 0.073 | 0.064 | -0.828 | -0.006 | 4.07E-01 | 18540 | -0.293 | -0.503 | -0.082 | 0.108 | -2.721 | -0.020 | 6.51E-03 | 18702 |
| 148 | Thal_PuL_R | -0.079 | -0.279 | 0.122 | 0.102 | -0.768 | -0.006 | 4.43E-01 | 18540 | -0.504 | -0.840 | -0.169 | 0.171 | -2.947 | -0.022 | 3.22E-03 | 18702 |
| 149 | Thal_PuI_L | -0.010 | -0.045 | 0.024 | 0.018 | -0.599 | -0.004 | 5.49E-01 | 18540 | -0.030 | -0.088 | 0.027 | 0.029 | -1.035 | -0.008 | 3.01E-01 | 18702 |
| 150 | Thal_PuI_R | -0.003 | -0.048 | 0.043 | 0.023 | -0.126 | -0.001 | 9.00E-01 | 18540 | -0.048 | -0.124 | 0.028 | 0.039 | -1.236 | -0.009 | 2.16E-01 | 18702 |
| 151 | ACC_sub_L | -0.190 | -0.585 | 0.206 | 0.202 | -0.940 | -0.007 | 3.47E-01 | 18540 | -0.265 | -0.927 | 0.396 | 0.338 | -0.786 | -0.006 | 4.32E-01 | 18702 |
| 152 | ACC_sub_R | -0.273 | -0.643 | 0.096 | 0.188 | -1.451 | -0.011 | 1.47E-01 | 18540 | -0.220 | -0.838 | 0.398 | 0.315 | -0.697 | -0.005 | 4.86E-01 | 18702 |
| 153 | ACC_pre_L | -0.703 | -1.716 | 0.311 | 0.517 | -1.358 | -0.010 | 1.74E-01 | 18540 | -1.972 | -3.667 | -0.276 | 0.865 | -2.279 | -0.017 | 2.27E-02 | 18702 |
| 154 | ACC_pre_R | -0.586 | -1.562 | 0.390 | 0.498 | -1.177 | -0.009 | 2.39E-01 | 18540 | -1.644 | -3.278 | -0.010 | 0.834 | -1.972 | -0.014 | 4.86E-02 | 18702 |
| 155 | ACC_sup_L | -0.207 | -1.309 | 0.895 | 0.562 | -0.368 | -0.003 | 7.13E-01 | 18540 | -1.166 | -3.010 | 0.678 | 0.941 | -1.240 | -0.009 | 2.15E-01 | 18702 |
| 156 | ACC_sup_R | -0.418 | -1.559 | 0.723 | 0.582 | -0.718 | -0.005 | 4.73E-01 | 18540 | -0.872 | -2.782 | 1.038 | 0.974 | -0.895 | -0.007 | 3.71E-01 | 18702 |

| | | | | | | | | | | | | | | | | | |
|---|---|---|---|---|---|---|---|---|---|---|---|---|---|---|---|---|---|
| 157 | N_Acc_L | -0.313 | -0.644 | 0.019 | 0.169 | -1.848 | -0.014 | 6.46E-02 | 18540 | -0.784 | -1.339 | -0.229 | 0.283 | -2.768 | -0.020 | 5.64E-03 | 18702 |
| 158 | N_Acc_R | -0.328 | -0.614 | -0.042 | 0.146 | -2.245 | -0.016 | 2.48E-02 | 18540 | -0.573 | -1.053 | -0.092 | 0.245 | -2.338 | -0.017 | 1.94E-02 | 18702 |
| 159 | VTA_L | -0.002 | -0.013 | 0.010 | 0.006 | -0.264 | -0.002 | 7.92E-01 | 18540 | 0.013 | -0.007 | 0.032 | 0.010 | 1.295 | 0.009 | 1.95E-01 | 18702 |
| 160 | VTA_R | -0.003 | -0.014 | 0.007 | 0.005 | -0.595 | -0.004 | 5.52E-01 | 18540 | 0.020 | 0.002 | 0.037 | 0.009 | 2.208 | 0.016 | 2.72E-02 | 18702 |
| 161 | SN_pc_L | -0.004 | -0.023 | 0.015 | 0.010 | -0.445 | -0.003 | 6.57E-01 | 18540 | -0.011 | -0.042 | 0.021 | 0.016 | -0.664 | -0.005 | 5.07E-01 | 18702 |
| 162 | SN_pc_R | -0.020 | -0.045 | 0.005 | 0.013 | -1.601 | -0.012 | 1.09E-01 | 18540 | 0.016 | -0.025 | 0.057 | 0.021 | 0.757 | 0.006 | 4.49E-01 | 18702 |
| 163 | SN_pr_L | -0.018 | -0.044 | 0.007 | 0.013 | -1.391 | -0.010 | 1.64E-01 | 18540 | -0.019 | -0.061 | 0.024 | 0.022 | -0.860 | -0.006 | 3.90E-01 | 18702 |
| 164 | SN_pr_R | -0.035 | -0.064 | -0.006 | 0.015 | -2.391 | -0.018 | 1.68E-02 | 18540 | 0.015 | -0.032 | 0.063 | 0.024 | 0.629 | 0.005 | 5.29E-01 | 18702 |
| 165 | Red_N_L | -0.029 | -0.060 | 0.003 | 0.016 | -1.784 | -0.013 | 7.44E-02 | 18540 | 0.003 | -0.050 | 0.056 | 0.027 | 0.114 | 0.001 | 9.09E-01 | 18702 |
| 166 | Red_N_R | -0.038 | -0.072 | -0.004 | 0.017 | -2.181 | -0.016 | 2.92E-02 | 18540 | 0.021 | -0.036 | 0.078 | 0.029 | 0.714 | 0.005 | 4.75E-01 | 18702 |
| 167 | LC_L | -0.008 | -0.020 | 0.004 | 0.006 | -1.353 | -0.010 | 1.76E-01 | 18540 | 0.016 | -0.004 | 0.035 | 0.010 | 1.566 | 0.011 | 1.17E-01 | 18702 |
| 168 | LC_R | -0.015 | -0.028 | -0.002 | 0.007 | -2.317 | -0.017 | 2.05E-02 | 18540 | 0.014 | -0.007 | 0.036 | 0.011 | 1.325 | 0.010 | 1.85E-01 | 18702 |
| 169 | Raphe_D | -0.042 | -0.088 | 0.003 | 0.023 | -1.823 | -0.013 | 6.84E-02 | 18540 | -0.010 | -0.086 | 0.066 | 0.039 | -0.254 | -0.002 | 8.00E-01 | 18702 |
| 170 | Raphe_M | -0.008 | -0.022 | 0.006 | 0.007 | -1.145 | -0.008 | 2.52E-01 | 18540 | 0.012 | -0.011 | 0.035 | 0.012 | 1.033 | 0.008 | 3.02E-01 | 18702 |

| | | Dried fruit intake | | | | | | | | Lamb/mutton intake | | | | | | | |
|---|---|---|---|---|---|---|---|---|---|---|---|---|---|---|---|---|---|
| Number | AAL 3 region | beta | 95%CI_l | 95%CI_h | se | t | r | p | df.e | beta | 95%CI_l | 95%CI_h | se | t | r | p | df.e |
| 0 | Total brain | 163.394 | 40.297 | 286.492 | 62.802 | 2.602 | 0.019 | 9.28E-03 | 18314 | -341.713 | -586.037 | -97.388 | 124.650 | -2.741 | -0.020 | 6.12E-03 | 18664 |
| 1 | Precentral_L | -0.382 | -4.723 | 3.960 | 2.215 | -0.172 | -0.001 | 8.63E-01 | 18314 | -3.116 | -11.725 | 5.493 | 4.392 | -0.709 | -0.005 | 4.78E-01 | 18664 |
| 2 | Precentral_R | 2.677 | -1.291 | 6.645 | 2.024 | 1.322 | 0.010 | 1.86E-01 | 18314 | -2.946 | -10.828 | 4.935 | 4.021 | -0.733 | -0.005 | 4.64E-01 | 18664 |
| 3 | Frontal_Sup_2_L | 3.981 | -0.590 | 8.552 | 2.332 | 1.707 | 0.013 | 8.79E-02 | 18314 | -9.679 | -18.767 | -0.591 | 4.637 | -2.088 | -0.015 | 3.68E-02 | 18664 |
| 4 | Frontal_Sup_2_R | 5.028 | -0.015 | 10.072 | 2.573 | 1.954 | 0.014 | 5.07E-02 | 18314 | -5.127 | -15.143 | 4.888 | 5.110 | -1.003 | -0.007 | 3.16E-01 | 18664 |
| 5 | Frontal_Mid_2_L | 4.752 | -0.051 | 9.556 | 2.451 | 1.939 | 0.014 | 5.25E-02 | 18314 | -6.516 | -16.059 | 3.027 | 4.869 | -1.338 | -0.010 | 1.81E-01 | 18664 |
| 6 | Frontal_Mid_2_R | 6.563 | 1.129 | 11.998 | 2.773 | 2.367 | 0.017 | 1.79E-02 | 18314 | -11.810 | -22.607 | -1.012 | 5.509 | -2.144 | -0.016 | 3.21E-02 | 18664 |
| 7 | Frontal_Inf_Oper_L | 1.984 | 0.564 | 3.403 | 0.724 | 2.739 | 0.020 | 6.16E-03 | 18314 | -2.771 | -5.590 | 0.048 | 1.438 | -1.927 | -0.014 | 5.40E-02 | 18664 |
| 8 | Frontal_Inf_Oper_R | 1.572 | -0.208 | 3.352 | 0.908 | 1.731 | 0.013 | 8.35E-02 | 18314 | -4.805 | -8.340 | -1.269 | 1.804 | -2.664 | -0.019 | 7.73E-03 | 18664 |
| 9 | Frontal_Inf_Tri_L | 4.153 | 1.139 | 7.168 | 1.538 | 2.700 | 0.020 | 6.93E-03 | 18314 | -0.813 | -6.803 | 5.177 | 3.056 | -0.266 | -0.002 | 7.90E-01 | 18664 |
| 10 | Frontal_Inf_Tri_R | 2.552 | 0.185 | 4.920 | 1.208 | 2.113 | 0.016 | 3.46E-02 | 18314 | -3.678 | -8.383 | 1.026 | 2.400 | -1.532 | -0.011 | 1.25E-01 | 18664 |
| 11 | Frontal_Inf_Orb_2_L | 0.567 | -0.335 | 1.468 | 0.460 | 1.232 | 0.009 | 2.18E-01 | 18314 | -0.335 | -2.127 | 1.456 | 0.914 | -0.367 | -0.003 | 7.14E-01 | 18664 |
| 12 | Frontal_Inf_Orb_2_R | 0.842 | -0.100 | 1.785 | 0.481 | 1.752 | 0.013 | 7.98E-02 | 18314 | -0.409 | -2.281 | 1.463 | 0.955 | -0.428 | -0.003 | 6.69E-01 | 18664 |
| 13 | Rolandic_Oper_L | 0.053 | -1.311 | 1.417 | 0.696 | 0.077 | 0.001 | 9.39E-01 | 18314 | -1.638 | -4.346 | 1.070 | 1.382 | -1.186 | -0.009 | 2.36E-01 | 18664 |
| 14 | Rolandic_Oper_R | 0.205 | -1.746 | 2.157 | 0.996 | 0.206 | 0.002 | 8.37E-01 | 18314 | -2.810 | -6.685 | 1.065 | 1.977 | -1.421 | -0.010 | 1.55E-01 | 18664 |
| 15 | Supp_Motor_Area_L | -0.400 | -3.638 | 2.839 | 1.652 | -0.242 | -0.002 | 8.09E-01 | 18314 | -2.156 | -8.591 | 4.278 | 3.283 | -0.657 | -0.005 | 5.11E-01 | 18664 |
| 16 | Supp_Motor_Area_R | -0.087 | -3.669 | 3.494 | 1.827 | -0.048 | 0.000 | 9.62E-01 | 18314 | -4.246 | -11.360 | 2.868 | 3.629 | -1.170 | -0.009 | 2.42E-01 | 18664 |
| 17 | Olfactory_L | 0.361 | -0.121 | 0.844 | 0.246 | 1.469 | 0.011 | 1.42E-01 | 18314 | 0.280 | -0.677 | 1.237 | 0.488 | 0.573 | 0.004 | 5.67E-01 | 18664 |
| 18 | Olfactory_R | 0.315 | -0.169 | 0.799 | 0.247 | 1.276 | 0.009 | 2.02E-01 | 18314 | -0.251 | -1.211 | 0.708 | 0.489 | -0.514 | -0.004 | 6.08E-01 | 18664 |
| 19 | Frontal_Sup_Medial_L | 5.266 | 2.230 | 8.303 | 1.549 | 3.399 | 0.025 | 6.77E-04 | 18314 | -6.896 | -12.926 | -0.866 | 3.076 | -2.242 | -0.016 | 2.50E-02 | 18664 |
| 20 | Frontal_Sup_Medial_R | 3.155 | 0.945 | 5.364 | 1.127 | 2.799 | 0.021 | 5.14E-03 | 18314 | -3.159 | -7.548 | 1.230 | 2.239 | -1.411 | -0.010 | 1.58E-01 | 18664 |
| 21 | Frontal_Med_Orb_L | 1.651 | 0.515 | 2.788 | 0.580 | 2.848 | 0.021 | 4.40E-03 | 18314 | -2.925 | -5.180 | -0.670 | 1.150 | -2.542 | -0.019 | 1.10E-02 | 18664 |
| 22 | Frontal_Med_Orb_R | 1.874 | 0.477 | 3.271 | 0.713 | 2.630 | 0.019 | 8.55E-03 | 18314 | -2.440 | -5.210 | 0.330 | 1.413 | -1.726 | -0.013 | 8.43E-02 | 18664 |
| 23 | Rectus_L | 1.292 | 0.117 | 2.468 | 0.600 | 2.155 | 0.016 | 3.12E-02 | 18314 | -1.345 | -3.679 | 0.988 | 1.190 | -1.130 | -0.008 | 2.59E-01 | 18664 |
| 24 | Rectus_R | 1.618 | 0.531 | 2.706 | 0.555 | 2.917 | 0.022 | 3.54E-03 | 18314 | -1.063 | -3.221 | 1.095 | 1.101 | -0.966 | -0.007 | 3.34E-01 | 18664 |
| 25 | OFCmed_L | 0.467 | -0.213 | 1.148 | 0.347 | 1.347 | 0.010 | 1.78E-01 | 18314 | -0.219 | -1.571 | 1.133 | 0.690 | -0.318 | -0.002 | 7.50E-01 | 18664 |
| 26 | OFCmed_R | 0.846 | 0.053 | 1.639 | 0.405 | 2.090 | 0.015 | 3.66E-02 | 18314 | -0.392 | -1.967 | 1.184 | 0.804 | -0.487 | -0.004 | 6.26E-01 | 18664 |
| 27 | OFCant_L | 0.145 | -0.643 | 0.933 | 0.402 | 0.361 | 0.003 | 7.18E-01 | 18314 | -1.075 | -2.640 | 0.489 | 0.798 | -1.347 | -0.010 | 1.78E-01 | 18664 |
| 28 | OFCant_R | 0.794 | -0.245 | 1.833 | 0.530 | 1.497 | 0.011 | 1.34E-01 | 18314 | -1.634 | -3.699 | 0.432 | 1.054 | -1.550 | -0.011 | 1.21E-01 | 18664 |
| 29 | OFCpost_L | 0.339 | -0.646 | 1.324 | 0.503 | 0.674 | 0.005 | 5.00E-01 | 18314 | -0.341 | -2.299 | 1.618 | 0.999 | -0.341 | -0.002 | 7.33E-01 | 18664 |
| 30 | OFCpost_R | 0.398 | -0.514 | 1.309 | 0.465 | 0.855 | 0.006 | 3.93E-01 | 18314 | -0.367 | -2.176 | 1.441 | 0.923 | -0.398 | -0.003 | 6.90E-01 | 18664 |
| 31 | OFClat_L | 0.459 | 0.127 | 0.792 | 0.170 | 2.709 | 0.020 | 6.76E-03 | 18314 | -0.090 | -0.751 | 0.571 | 0.337 | -0.266 | -0.002 | 7.90E-01 | 18664 |
| 32 | OFClat_R | 0.494 | 0.213 | 0.774 | 0.143 | 3.449 | 0.025 | 5.64E-04 | 18314 | -0.303 | -0.860 | 0.255 | 0.285 | -1.064 | -0.008 | 2.88E-01 | 18664 |

| | | | | | | | | | | | | | | | | | |
|---|---|---|---|---|---|---|---|---|---|---|---|---|---|---|---|---|---|
| 33 | Insula_L | 0.547 | -2.505 | 3.600 | 1.557 | 0.351 | 0.003 | 7.25E-01 | 18314 | -3.229 | -9.282 | 2.825 | 3.088 | -1.045 | -0.008 | 2.96E-01 | 18664 |
| 34 | Insula_R | -0.089 | -3.063 | 2.886 | 1.517 | -0.058 | 0.000 | 9.53E-01 | 18314 | -3.991 | -9.887 | 1.905 | 3.008 | -1.327 | -0.010 | 1.85E-01 | 18664 |
| 37 | Cingulate_Mid_L | 3.746 | 1.167 | 6.326 | 1.316 | 2.847 | 0.021 | 4.42E-03 | 18314 | -8.235 | -13.346 | -3.124 | 2.608 | -3.158 | -0.023 | 1.59E-03 | 18664 |
| 38 | Cingulate_Mid_R | 1.703 | -1.049 | 4.455 | 1.404 | 1.213 | 0.009 | 2.25E-01 | 18314 | -6.565 | -12.016 | -1.114 | 2.781 | -2.360 | -0.017 | 1.83E-02 | 18664 |
| 39 | Cingulate_Post_L | -0.306 | -1.094 | 0.481 | 0.402 | -0.762 | -0.006 | 4.46E-01 | 18314 | -1.081 | -2.643 | 0.481 | 0.797 | -1.356 | -0.010 | 1.75E-01 | 18664 |
| 40 | Cingulate_Post_R | -0.367 | -0.804 | 0.070 | 0.223 | -1.646 | -0.012 | 9.98E-02 | 18314 | -0.025 | -0.892 | 0.841 | 0.442 | -0.057 | 0.000 | 9.54E-01 | 18664 |
| 41 | Hippocampus_L | 1.077 | -0.195 | 2.349 | 0.649 | 1.659 | 0.012 | 9.71E-02 | 18314 | -0.921 | -3.450 | 1.608 | 1.290 | -0.714 | -0.005 | 4.76E-01 | 18664 |
| 42 | Hippocampus_R | 1.496 | 0.175 | 2.816 | 0.674 | 2.221 | 0.016 | 2.64E-02 | 18314 | -1.329 | -3.950 | 1.293 | 1.337 | -0.993 | -0.007 | 3.21E-01 | 18664 |
| 43 | ParaHippocampal_L | 1.458 | 0.327 | 2.589 | 0.577 | 2.528 | 0.019 | 1.15E-02 | 18314 | -1.591 | -3.837 | 0.656 | 1.146 | -1.388 | -0.010 | 1.65E-01 | 18664 |
| 44 | ParaHippocampal_R | 1.281 | -0.148 | 2.709 | 0.729 | 1.757 | 0.013 | 7.90E-02 | 18314 | -1.545 | -4.379 | 1.289 | 1.446 | -1.069 | -0.008 | 2.85E-01 | 18664 |
| 45 | Amygdala_L | -0.022 | -0.462 | 0.418 | 0.224 | -0.097 | -0.001 | 9.23E-01 | 18314 | -0.236 | -1.110 | 0.637 | 0.446 | -0.530 | -0.004 | 5.96E-01 | 18664 |
| 46 | Amygdala_R | -0.008 | -0.458 | 0.442 | 0.230 | -0.035 | 0.000 | 9.72E-01 | 18314 | -0.586 | -1.478 | 0.307 | 0.455 | -1.287 | -0.009 | 1.98E-01 | 18664 |
| 47 | Calcarine_L | 1.594 | -2.599 | 5.787 | 2.139 | 0.745 | 0.006 | 4.56E-01 | 18314 | -0.396 | -8.732 | 7.939 | 4.253 | -0.093 | -0.001 | 9.26E-01 | 18664 |
| 48 | Calcarine_R | 0.362 | -3.142 | 3.867 | 1.788 | 0.203 | 0.001 | 8.39E-01 | 18314 | -0.049 | -7.016 | 6.917 | 3.554 | -0.014 | 0.000 | 9.89E-01 | 18664 |
| 49 | Cuneus_L | 1.238 | -1.395 | 3.872 | 1.343 | 0.922 | 0.007 | 3.57E-01 | 18314 | -1.208 | -6.433 | 4.017 | 2.666 | -0.453 | -0.003 | 6.50E-01 | 18664 |
| 50 | Cuneus_R | -0.605 | -2.620 | 1.410 | 1.028 | -0.588 | -0.004 | 5.56E-01 | 18314 | -1.798 | -5.795 | 2.198 | 2.039 | -0.882 | -0.006 | 3.78E-01 | 18664 |
| 51 | Lingual_L | 2.541 | 0.020 | 5.063 | 1.287 | 1.975 | 0.015 | 4.82E-02 | 18314 | -1.736 | -6.748 | 3.275 | 2.557 | -0.679 | -0.005 | 4.97E-01 | 18664 |
| 52 | Lingual_R | 1.046 | -1.597 | 3.688 | 1.348 | 0.776 | 0.006 | 4.38E-01 | 18314 | -0.199 | -5.450 | 5.053 | 2.679 | -0.074 | -0.001 | 9.41E-01 | 18664 |
| 53 | Occipital_Sup_L | 1.701 | -0.034 | 3.435 | 0.885 | 1.922 | 0.014 | 5.46E-02 | 18314 | -3.699 | -7.145 | -0.254 | 1.758 | -2.105 | -0.015 | 3.53E-02 | 18664 |
| 54 | Occipital_Sup_R | -0.075 | -1.956 | 1.806 | 0.960 | -0.078 | -0.001 | 9.38E-01 | 18314 | -2.155 | -5.887 | 1.577 | 1.904 | -1.132 | -0.008 | 2.58E-01 | 18664 |
| 55 | Occipital_Mid_L | -3.786 | -7.855 | 0.283 | 2.076 | -1.824 | -0.013 | 6.82E-02 | 18314 | -5.070 | -13.145 | 3.005 | 4.120 | -1.231 | -0.009 | 2.18E-01 | 18664 |
| 56 | Occipital_Mid_R | -0.108 | -2.940 | 2.723 | 1.445 | -0.075 | -0.001 | 9.40E-01 | 18314 | -0.613 | -6.232 | 5.006 | 2.867 | -0.214 | -0.002 | 8.31E-01 | 18664 |
| 57 | Occipital_Inf_L | -1.152 | -2.503 | 0.199 | 0.689 | -1.671 | -0.012 | 9.47E-02 | 18314 | -2.588 | -5.269 | 0.094 | 1.368 | -1.892 | -0.014 | 5.85E-02 | 18664 |
| 58 | Occipital_Inf_R | -1.999 | -3.558 | -0.439 | 0.796 | -2.512 | -0.019 | 1.20E-02 | 18314 | 0.606 | -2.489 | 3.700 | 1.579 | 0.384 | 0.003 | 7.01E-01 | 18664 |
| 59 | Fusiform_L | 2.513 | -0.440 | 5.465 | 1.506 | 1.668 | 0.012 | 9.53E-02 | 18314 | -7.308 | -13.176 | -1.441 | 2.993 | -2.442 | -0.018 | 1.46E-02 | 18664 |
| 60 | Fusiform_R | 2.792 | -0.455 | 6.039 | 1.657 | 1.686 | 0.012 | 9.19E-02 | 18314 | -4.606 | -11.051 | 1.839 | 3.288 | -1.401 | -0.010 | 1.61E-01 | 18664 |
| 61 | Postcentral_L | 5.700 | 1.260 | 10.140 | 2.265 | 2.516 | 0.019 | 1.19E-02 | 18314 | -7.322 | -16.143 | 1.499 | 4.500 | -1.627 | -0.012 | 1.04E-01 | 18664 |
| 62 | Postcentral_R | 6.082 | 1.528 | 10.636 | 2.323 | 2.618 | 0.019 | 8.86E-03 | 18314 | -5.895 | -14.940 | 3.150 | 4.615 | -1.277 | -0.009 | 2.01E-01 | 18664 |
| 63 | Parietal_Sup_L | 0.065 | -2.494 | 2.624 | 1.306 | 0.050 | 0.000 | 9.60E-01 | 18314 | -0.511 | -5.595 | 4.573 | 2.594 | -0.197 | -0.001 | 8.44E-01 | 18664 |
| 64 | Parietal_Sup_R | 0.699 | -1.574 | 2.972 | 1.160 | 0.603 | 0.004 | 5.47E-01 | 18314 | -0.647 | -5.154 | 3.861 | 2.300 | -0.281 | -0.002 | 7.79E-01 | 18664 |
| 65 | Parietal_Inf_L | 2.158 | -1.193 | 5.510 | 1.710 | 1.262 | 0.009 | 2.07E-01 | 18314 | -6.360 | -13.014 | 0.294 | 3.395 | -1.873 | -0.014 | 6.10E-02 | 18664 |
| 66 | Parietal_Inf_R | 1.011 | -1.161 | 3.183 | 1.108 | 0.912 | 0.007 | 3.62E-01 | 18314 | -2.856 | -7.167 | 1.455 | 2.199 | -1.299 | -0.010 | 1.94E-01 | 18664 |
| 67 | SupraMarginal_L | 1.185 | -0.779 | 3.149 | 1.002 | 1.183 | 0.009 | 2.37E-01 | 18314 | -5.101 | -9.003 | -1.199 | 1.991 | -2.562 | -0.019 | 1.04E-02 | 18664 |
| 68 | SupraMarginal_R | 1.964 | -0.812 | 4.739 | 1.416 | 1.387 | 0.010 | 1.65E-01 | 18314 | -9.279 | -14.795 | -3.762 | 2.814 | -3.297 | -0.024 | 9.79E-04 | 18664 |
| 69 | Angular_L | 1.490 | -0.440 | 3.420 | 0.985 | 1.513 | 0.011 | 1.30E-01 | 18314 | -4.570 | -8.401 | -0.739 | 1.954 | -2.338 | -0.017 | 1.94E-02 | 18664 |
| 70 | Angular_R | 1.951 | -0.640 | 4.542 | 1.322 | 1.476 | 0.011 | 1.40E-01 | 18314 | -2.327 | -7.465 | 2.811 | 2.621 | -0.888 | -0.006 | 3.75E-01 | 18664 |
| 71 | Precuneus_L | 2.070 | -2.638 | 6.779 | 2.402 | 0.862 | 0.006 | 3.89E-01 | 18314 | -1.834 | -11.168 | 7.500 | 4.762 | -0.385 | -0.003 | 7.00E-01 | 18664 |
| 72 | Precuneus_R | -0.037 | -4.232 | 4.158 | 2.140 | -0.017 | 0.000 | 9.86E-01 | 18314 | -2.575 | -10.887 | 5.738 | 4.241 | -0.607 | -0.004 | 5.44E-01 | 18664 |
| 73 | Paracentral_Lobule_L | -1.060 | -3.213 | 1.092 | 1.098 | -0.965 | -0.007 | 3.34E-01 | 18314 | 0.285 | -3.993 | 4.563 | 2.183 | 0.131 | 0.001 | 8.96E-01 | 18664 |
| 74 | Paracentral_Lobule_R | -0.550 | -2.093 | 0.992 | 0.787 | -0.699 | -0.005 | 4.84E-01 | 18314 | -0.349 | -3.413 | 2.716 | 1.563 | -0.223 | -0.002 | 8.24E-01 | 18664 |
| 75 | Caudate_L | 0.752 | -0.964 | 2.468 | 0.875 | 0.859 | 0.006 | 3.90E-01 | 18314 | -3.253 | -6.670 | 0.165 | 1.744 | -1.865 | -0.014 | 6.21E-02 | 18664 |
| 76 | Caudate_R | 1.167 | -0.777 | 3.110 | 0.992 | 1.177 | 0.009 | 2.39E-01 | 18314 | -5.146 | -9.013 | -1.278 | 1.973 | -2.608 | -0.019 | 9.11E-03 | 18664 |
| 77 | Putamen_L | 2.133 | -0.479 | 4.744 | 1.332 | 1.601 | 0.012 | 1.09E-01 | 18314 | -5.996 | -11.184 | -0.809 | 2.646 | -2.266 | -0.017 | 2.35E-02 | 18664 |
| 78 | Putamen_R | 1.749 | -0.910 | 4.408 | 1.356 | 1.290 | 0.010 | 1.97E-01 | 18314 | -6.072 | -11.355 | -0.789 | 2.695 | -2.253 | -0.016 | 2.43E-02 | 18664 |
| 79 | Pallidum_L | 0.049 | -0.183 | 0.282 | 0.119 | 0.417 | 0.003 | 6.76E-01 | 18314 | -0.216 | -0.677 | 0.245 | 0.235 | -0.918 | -0.007 | 3.59E-01 | 18664 |
| 80 | Pallidum_R | 0.011 | -0.244 | 0.265 | 0.130 | 0.082 | 0.001 | 9.35E-01 | 18314 | -0.434 | -0.941 | 0.072 | 0.258 | -1.682 | -0.012 | 9.27E-02 | 18664 |
| 83 | Heschl_L | -0.154 | -0.625 | 0.317 | 0.240 | -0.640 | -0.005 | 5.22E-01 | 18314 | -0.314 | -1.249 | 0.621 | 0.477 | -0.657 | -0.005 | 5.11E-01 | 18664 |
| 84 | Heschl_R | -0.061 | -0.587 | 0.466 | 0.269 | -0.226 | -0.002 | 8.21E-01 | 18314 | -0.756 | -1.801 | 0.289 | 0.533 | -1.419 | -0.010 | 1.56E-01 | 18664 |
| 85 | Temporal_Sup_L | -0.312 | -3.710 | 3.087 | 1.734 | -0.180 | -0.001 | 8.57E-01 | 18314 | -5.050 | -11.801 | 1.701 | 3.444 | -1.466 | -0.011 | 1.43E-01 | 18664 |

| | | | | | | | | | | | | | | | | | |
|---|---|---|---|---|---|---|---|---|---|---|---|---|---|---|---|---|---|
| 86 | Temporal_Sup_R | 1.106 | -2.655 | 4.867 | 1.919 | 0.576 | 0.004 | 5.64E-01 | 18314 | -8.642 | -16.107 | -1.177 | 3.809 | -2.269 | -0.017 | 2.33E-02 | 18664 |
| 87 | Temporal_Pole_Sup_L | 0.665 | -0.948 | 2.278 | 0.823 | 0.808 | 0.006 | 4.19E-01 | 18314 | -0.924 | -4.121 | 2.272 | 1.631 | -0.567 | -0.004 | 5.71E-01 | 18664 |
| 88 | Temporal_Pole_Sup_R | 1.060 | -0.639 | 2.758 | 0.867 | 1.223 | 0.009 | 2.21E-01 | 18314 | -2.556 | -5.920 | 0.809 | 1.716 | -1.489 | -0.011 | 1.37E-01 | 18664 |
| 89 | Temporal_Mid_L | 3.722 | -1.942 | 9.386 | 2.890 | 1.288 | 0.010 | 1.98E-01 | 18314 | -7.990 | -19.240 | 3.260 | 5.739 | -1.392 | -0.010 | 1.64E-01 | 18664 |
| 90 | Temporal_Mid_R | 1.196 | -3.838 | 6.231 | 2.568 | 0.466 | 0.003 | 6.41E-01 | 18314 | -8.937 | -18.916 | 1.041 | 5.091 | -1.756 | -0.013 | 7.92E-02 | 18664 |
| 91 | Temporal_Pole_Mid_L | 0.907 | -0.448 | 2.261 | 0.691 | 1.312 | 0.010 | 1.90E-01 | 18314 | -1.090 | -3.778 | 1.599 | 1.372 | -0.795 | -0.006 | 4.27E-01 | 18664 |
| 92 | Temporal_Pole_Mid_R | 1.496 | -0.150 | 3.142 | 0.840 | 1.782 | 0.013 | 7.48E-02 | 18314 | -2.328 | -5.592 | 0.936 | 1.665 | -1.398 | -0.010 | 1.62E-01 | 18664 |
| 93 | Temporal_Inf_L | 0.217 | -3.677 | 4.111 | 1.986 | 0.109 | 0.001 | 9.13E-01 | 18314 | -5.242 | -12.975 | 2.491 | 3.945 | -1.329 | -0.010 | 1.84E-01 | 18664 |
| 94 | Temporal_Inf_R | -0.763 | -5.016 | 3.489 | 2.169 | -0.352 | -0.003 | 7.25E-01 | 18314 | -5.526 | -13.948 | 2.896 | 4.297 | -1.286 | -0.009 | 1.98E-01 | 18664 |
| 95 | Cerebellum_Crus1_L | 4.483 | -0.145 | 9.111 | 2.361 | 1.899 | 0.014 | 5.76E-02 | 18314 | -6.450 | -15.634 | 2.734 | 4.685 | -1.377 | -0.010 | 1.69E-01 | 18664 |
| 96 | Cerebellum_Crus1_R | 5.323 | 0.605 | 10.040 | 2.407 | 2.212 | 0.016 | 2.70E-02 | 18314 | -5.279 | -14.649 | 4.092 | 4.780 | -1.104 | -0.008 | 2.70E-01 | 18664 |
| 97 | Cerebellum_Crus2_L | 4.306 | 0.642 | 7.971 | 1.870 | 2.303 | 0.017 | 2.13E-02 | 18314 | -6.333 | -13.625 | 0.959 | 3.720 | -1.702 | -0.012 | 8.87E-02 | 18664 |
| 98 | Cerebellum_Crus2_R | 4.195 | 0.805 | 7.585 | 1.730 | 2.425 | 0.018 | 1.53E-02 | 18314 | -4.644 | -11.389 | 2.102 | 3.442 | -1.349 | -0.010 | 1.77E-01 | 18664 |
| 99 | Cerebellum_3_L | 0.113 | -0.133 | 0.360 | 0.126 | 0.899 | 0.007 | 3.69E-01 | 18314 | 0.234 | -0.255 | 0.724 | 0.250 | 0.938 | 0.007 | 3.48E-01 | 18664 |
| 100 | Cerebellum_3_R | 0.188 | -0.145 | 0.520 | 0.170 | 1.105 | 0.008 | 2.69E-01 | 18314 | 0.330 | -0.330 | 0.991 | 0.337 | 0.981 | 0.007 | 3.27E-01 | 18664 |
| 101 | Cerebellum_4_5_L | 2.236 | 0.242 | 4.230 | 1.017 | 2.198 | 0.016 | 2.80E-02 | 18314 | 0.038 | -3.919 | 3.995 | 2.019 | 0.019 | 0.000 | 9.85E-01 | 18664 |
| 102 | Cerebellum_4_5_R | 1.698 | 0.073 | 3.322 | 0.829 | 2.048 | 0.015 | 4.05E-02 | 18314 | 0.385 | -2.840 | 3.610 | 1.645 | 0.234 | 0.002 | 8.15E-01 | 18664 |
| 103 | Cerebellum_6_L | 3.956 | 0.507 | 7.404 | 1.759 | 2.248 | 0.017 | 2.46E-02 | 18314 | -8.186 | -15.024 | -1.349 | 3.488 | -2.347 | -0.017 | 1.89E-02 | 18664 |
| 104 | Cerebellum_6_R | 4.539 | 0.909 | 8.169 | 1.852 | 2.451 | 0.018 | 1.43E-02 | 18314 | -4.678 | -11.884 | 2.528 | 3.676 | -1.272 | -0.009 | 2.03E-01 | 18664 |
| 105 | Cerebellum_7b_L | 1.703 | 0.582 | 2.823 | 0.572 | 2.978 | 0.022 | 2.90E-03 | 18314 | -1.399 | -3.628 | 0.829 | 1.137 | -1.231 | -0.009 | 2.18E-01 | 18664 |
| 106 | Cerebellum_7b_R | 2.087 | 0.977 | 3.198 | 0.566 | 3.685 | 0.027 | 2.29E-04 | 18314 | -2.015 | -4.224 | 0.195 | 1.127 | -1.788 | -0.013 | 7.39E-02 | 18664 |
| 107 | Cerebellum_8_L | 3.463 | -0.095 | 7.020 | 1.815 | 1.908 | 0.014 | 5.64E-02 | 18314 | -3.543 | -10.616 | 3.530 | 3.609 | -0.982 | -0.007 | 3.26E-01 | 18664 |
| 108 | Cerebellum_8_R | 7.546 | 2.912 | 12.180 | 2.364 | 3.192 | 0.024 | 1.42E-03 | 18314 | -7.960 | -17.167 | 1.247 | 4.697 | -1.695 | -0.012 | 9.02E-02 | 18664 |
| 109 | Cerebellum_9_L | -1.412 | -3.425 | 0.602 | 1.027 | -1.374 | -0.010 | 1.69E-01 | 18314 | -2.618 | -6.617 | 1.380 | 2.040 | -1.283 | -0.009 | 1.99E-01 | 18664 |
| 110 | Cerebellum_9_R | -1.261 | -3.242 | 0.721 | 1.011 | -1.247 | -0.009 | 2.12E-01 | 18314 | -1.890 | -5.821 | 2.041 | 2.005 | -0.943 | -0.007 | 3.46E-01 | 18664 |
| 111 | Cerebellum_10_L | -0.139 | -0.282 | 0.004 | 0.073 | -1.902 | -0.014 | 5.72E-02 | 18314 | -0.086 | -0.370 | 0.198 | 0.145 | -0.593 | -0.004 | 5.53E-01 | 18664 |
| 112 | Cerebellum_10_R | -0.019 | -0.168 | 0.129 | 0.076 | -0.254 | -0.002 | 7.99E-01 | 18314 | -0.072 | -0.368 | 0.223 | 0.151 | -0.481 | -0.004 | 6.31E-01 | 18664 |
| 113 | Vermis_1_2 | 0.003 | -0.090 | 0.096 | 0.047 | 0.058 | 0.000 | 9.54E-01 | 18314 | 0.083 | -0.102 | 0.267 | 0.094 | 0.877 | 0.006 | 3.80E-01 | 18664 |
| 114 | Vermis_3 | 0.245 | -0.125 | 0.614 | 0.188 | 1.298 | 0.010 | 1.94E-01 | 18314 | 0.014 | -0.720 | 0.747 | 0.374 | 0.037 | 0.000 | 9.70E-01 | 18664 |
| 115 | Vermis_4_5 | 0.925 | -0.160 | 2.010 | 0.553 | 1.671 | 0.012 | 9.48E-02 | 18314 | 0.113 | -2.043 | 2.269 | 1.100 | 0.103 | 0.001 | 9.18E-01 | 18664 |
| 116 | Vermis_6 | 0.030 | -0.574 | 0.634 | 0.308 | 0.097 | 0.001 | 9.22E-01 | 18314 | -0.596 | -1.796 | 0.604 | 0.612 | -0.973 | -0.007 | 3.30E-01 | 18664 |
| 117 | Vermis_7 | 0.097 | -0.245 | 0.439 | 0.174 | 0.554 | 0.004 | 5.80E-01 | 18314 | -0.362 | -1.040 | 0.317 | 0.346 | -1.045 | -0.008 | 2.96E-01 | 18664 |
| 118 | Vermis_8 | -0.047 | -0.608 | 0.515 | 0.286 | -0.163 | -0.001 | 8.71E-01 | 18314 | -1.228 | -2.343 | -0.113 | 0.569 | -2.159 | -0.016 | 3.08E-02 | 18664 |
| 119 | Vermis_9 | -0.263 | -0.718 | 0.192 | 0.232 | -1.132 | -0.008 | 2.58E-01 | 18314 | -0.419 | -1.323 | 0.484 | 0.461 | -0.910 | -0.007 | 3.63E-01 | 18664 |
| 120 | Vermis_10 | -0.111 | -0.238 | 0.016 | 0.065 | -1.709 | -0.013 | 8.74E-02 | 18314 | 0.083 | -0.169 | 0.335 | 0.129 | 0.645 | 0.005 | 5.19E-01 | 18664 |
| 121 | Thal_AV_L | 0.062 | 0.011 | 0.112 | 0.026 | 2.385 | 0.018 | 1.71E-02 | 18314 | -0.072 | -0.173 | 0.029 | 0.052 | -1.391 | -0.010 | 1.64E-01 | 18664 |
| 122 | Thal_AV_R | 0.104 | 0.031 | 0.176 | 0.037 | 2.807 | 0.021 | 5.01E-03 | 18314 | -0.054 | -0.199 | 0.090 | 0.074 | -0.738 | -0.005 | 4.60E-01 | 18664 |
| 123 | Thal_LP_L | 0.089 | 0.032 | 0.145 | 0.029 | 3.087 | 0.023 | 2.03E-03 | 18314 | -0.087 | -0.199 | 0.024 | 0.057 | -1.531 | -0.011 | 1.26E-01 | 18664 |
| 124 | Thal_LP_R | 0.119 | 0.046 | 0.192 | 0.037 | 3.191 | 0.024 | 1.42E-03 | 18314 | -0.113 | -0.259 | 0.033 | 0.074 | -1.521 | -0.011 | 1.28E-01 | 18664 |
| 125 | Thal_VA_L | 0.039 | -0.021 | 0.099 | 0.031 | 1.275 | 0.009 | 2.02E-01 | 18314 | -0.029 | -0.149 | 0.090 | 0.061 | -0.480 | -0.004 | 6.31E-01 | 18664 |
| 126 | Thal_VA_R | 0.063 | -0.010 | 0.137 | 0.038 | 1.683 | 0.012 | 9.23E-02 | 18314 | -0.040 | -0.186 | 0.107 | 0.075 | -0.529 | -0.004 | 5.97E-01 | 18664 |
| 127 | Thal_VL_L | 0.179 | 0.009 | 0.350 | 0.087 | 2.060 | 0.015 | 3.94E-02 | 18314 | -0.321 | -0.660 | 0.018 | 0.173 | -1.855 | -0.014 | 6.36E-02 | 18664 |
| 128 | Thal_VL_R | 0.358 | 0.131 | 0.584 | 0.116 | 3.097 | 0.023 | 1.95E-03 | 18314 | -0.386 | -0.837 | 0.065 | 0.230 | -1.680 | -0.012 | 9.31E-02 | 18664 |
| 129 | Thal_VPL_L | 0.135 | -0.069 | 0.338 | 0.104 | 1.295 | 0.010 | 1.95E-01 | 18314 | -0.166 | -0.570 | 0.238 | 0.206 | -0.807 | -0.006 | 4.19E-01 | 18664 |
| 130 | Thal_VPL_R | 0.299 | 0.085 | 0.513 | 0.109 | 2.740 | 0.020 | 6.14E-03 | 18314 | -0.269 | -0.693 | 0.156 | 0.217 | -1.239 | -0.009 | 2.15E-01 | 18664 |
| 131 | Thal_IL_L | 0.153 | 0.065 | 0.241 | 0.045 | 3.420 | 0.025 | 6.29E-04 | 18314 | -0.187 | -0.361 | -0.012 | 0.089 | -2.095 | -0.015 | 3.62E-02 | 18664 |
| 132 | Thal_IL_R | 0.206 | 0.107 | 0.306 | 0.051 | 4.062 | 0.030 | 4.88E-05 | 18314 | -0.201 | -0.399 | -0.002 | 0.101 | -1.985 | -0.015 | 4.72E-02 | 18664 |
| 133 | Thal_Re_L | 0.000 | 0.000 | 0.001 | 0.000 | 1.961 | 0.014 | 4.98E-02 | 18314 | 0.000 | -0.001 | 0.001 | 0.000 | 0.288 | 0.002 | 7.73E-01 | 18664 |
| 134 | Thal_Re_R | 0.004 | 0.001 | 0.007 | 0.002 | 2.672 | 0.020 | 7.54E-03 | 18314 | -0.003 | -0.009 | 0.003 | 0.003 | -0.933 | -0.007 | 3.51E-01 | 18664 |

| | | | | | | | | | | | | | | | | | |
|---|---|---|---|---|---|---|---|---|---|---|---|---|---|---|---|---|---|
| 135 | Thal_MDm_L | 1.256 | 0.606 | 1.907 | 0.332 | 3.785 | 0.028 | 1.54E-04 | 18314 | -1.523 | -2.817 | -0.228 | 0.660 | -2.306 | -0.017 | 2.11E-02 | 18664 |
| 136 | Thal_MDm_R | 1.518 | 0.763 | 2.273 | 0.385 | 3.940 | 0.029 | 8.16E-05 | 18314 | -1.691 | -3.195 | -0.186 | 0.768 | -2.202 | -0.016 | 2.77E-02 | 18664 |
| 137 | Thal_MDl_L | 0.229 | 0.116 | 0.342 | 0.057 | 3.986 | 0.029 | 6.74E-05 | 18314 | -0.280 | -0.504 | -0.056 | 0.114 | -2.448 | -0.018 | 1.44E-02 | 18664 |
| 138 | Thal_MDl_R | 0.264 | 0.136 | 0.391 | 0.065 | 4.041 | 0.030 | 5.35E-05 | 18314 | -0.292 | -0.547 | -0.037 | 0.130 | -2.247 | -0.016 | 2.47E-02 | 18664 |
| 139 | Thal_LGN_L | 0.024 | 0.007 | 0.041 | 0.009 | 2.780 | 0.021 | 5.44E-03 | 18314 | -0.016 | -0.050 | 0.018 | 0.017 | -0.923 | -0.007 | 3.56E-01 | 18664 |
| 140 | Thal_LGN_R | 0.073 | 0.037 | 0.110 | 0.019 | 3.952 | 0.029 | 7.80E-05 | 18314 | -0.083 | -0.156 | -0.011 | 0.037 | -2.253 | -0.016 | 2.42E-02 | 18664 |
| 141 | Thal_MGN_L | 0.042 | 0.011 | 0.072 | 0.016 | 2.682 | 0.020 | 7.32E-03 | 18314 | -0.061 | -0.122 | 0.000 | 0.031 | -1.970 | -0.014 | 4.88E-02 | 18664 |
| 142 | Thal_MGN_R | 0.059 | 0.026 | 0.092 | 0.017 | 3.477 | 0.026 | 5.08E-04 | 18314 | -0.072 | -0.138 | -0.006 | 0.034 | -2.130 | -0.016 | 3.32E-02 | 18664 |
| 143 | Thal_PuA_L | 0.057 | 0.021 | 0.093 | 0.018 | 3.074 | 0.023 | 2.11E-03 | 18314 | -0.070 | -0.141 | 0.002 | 0.037 | -1.905 | -0.014 | 5.68E-02 | 18664 |
| 144 | Thal_PuA_R | 0.144 | 0.069 | 0.219 | 0.038 | 3.762 | 0.028 | 1.69E-04 | 18314 | -0.151 | -0.300 | -0.002 | 0.076 | -1.984 | -0.015 | 4.73E-02 | 18664 |
| 145 | Thal_PuM_L | 1.279 | 0.561 | 1.997 | 0.366 | 3.494 | 0.026 | 4.77E-04 | 18314 | -1.127 | -2.550 | 0.297 | 0.726 | -1.551 | -0.011 | 1.21E-01 | 18664 |
| 146 | Thal_PuM_R | 1.656 | 0.763 | 2.550 | 0.456 | 3.634 | 0.027 | 2.80E-04 | 18314 | -1.368 | -3.143 | 0.406 | 0.905 | -1.511 | -0.011 | 1.31E-01 | 18664 |
| 147 | Thal_PuL_L | 0.224 | 0.093 | 0.355 | 0.067 | 3.348 | 0.025 | 8.15E-04 | 18314 | -0.208 | -0.467 | 0.052 | 0.133 | -1.566 | -0.011 | 1.17E-01 | 18664 |
| 148 | Thal_PuL_R | 0.377 | 0.169 | 0.585 | 0.106 | 3.550 | 0.026 | 3.86E-04 | 18314 | -0.366 | -0.779 | 0.048 | 0.211 | -1.734 | -0.013 | 8.29E-02 | 18664 |
| 149 | Thal_PuI_L | 0.018 | -0.018 | 0.054 | 0.018 | 0.996 | 0.007 | 3.19E-01 | 18314 | -0.009 | -0.080 | 0.062 | 0.036 | -0.247 | -0.002 | 8.05E-01 | 18664 |
| 150 | Thal_PuI_R | 0.062 | 0.015 | 0.110 | 0.024 | 2.604 | 0.019 | 9.23E-03 | 18314 | -0.036 | -0.130 | 0.057 | 0.048 | -0.761 | -0.006 | 4.47E-01 | 18664 |
| 151 | ACC_sub_L | -0.010 | -0.422 | 0.401 | 0.210 | -0.049 | 0.000 | 9.61E-01 | 18314 | -0.719 | -1.535 | 0.097 | 0.416 | -1.727 | -0.013 | 8.41E-02 | 18664 |
| 152 | ACC_sub_R | 0.059 | -0.325 | 0.443 | 0.196 | 0.301 | 0.002 | 7.64E-01 | 18314 | -0.602 | -1.364 | 0.161 | 0.389 | -1.546 | -0.011 | 1.22E-01 | 18664 |
| 153 | ACC_pre_L | 0.621 | -0.434 | 1.675 | 0.538 | 1.154 | 0.009 | 2.49E-01 | 18314 | -2.927 | -5.017 | -0.837 | 1.066 | -2.745 | -0.020 | 6.05E-03 | 18664 |
| 154 | ACC_pre_R | 0.521 | -0.494 | 1.537 | 0.518 | 1.006 | 0.007 | 3.14E-01 | 18314 | -2.418 | -4.431 | -0.405 | 1.027 | -2.354 | -0.017 | 1.86E-02 | 18664 |
| 155 | ACC_sup_L | 0.503 | -0.644 | 1.650 | 0.585 | 0.859 | 0.006 | 3.90E-01 | 18314 | -1.904 | -4.177 | 0.369 | 1.160 | -1.642 | -0.012 | 1.01E-01 | 18664 |
| 156 | ACC_sup_R | 0.207 | -0.980 | 1.393 | 0.605 | 0.342 | 0.003 | 7.32E-01 | 18314 | -1.615 | -3.970 | 0.739 | 1.201 | -1.345 | -0.010 | 1.79E-01 | 18664 |
| 157 | N_Acc_L | 0.404 | 0.060 | 0.748 | 0.176 | 2.304 | 0.017 | 2.13E-02 | 18314 | -0.279 | -0.963 | 0.405 | 0.349 | -0.799 | -0.006 | 4.24E-01 | 18664 |
| 158 | N_Acc_R | 0.160 | -0.138 | 0.458 | 0.152 | 1.055 | 0.008 | 2.92E-01 | 18314 | -0.330 | -0.922 | 0.261 | 0.302 | -1.095 | -0.008 | 2.74E-01 | 18664 |
| 159 | VTA_L | 0.008 | -0.004 | 0.020 | 0.006 | 1.256 | 0.009 | 2.09E-01 | 18314 | 0.003 | -0.021 | 0.027 | 0.012 | 0.276 | 0.002 | 7.82E-01 | 18664 |
| 160 | VTA_R | 0.007 | -0.004 | 0.018 | 0.006 | 1.272 | 0.009 | 2.03E-01 | 18314 | -0.001 | -0.023 | 0.021 | 0.011 | -0.087 | -0.001 | 9.31E-01 | 18664 |
| 161 | SN_pc_L | 0.006 | -0.014 | 0.026 | 0.010 | 0.603 | 0.004 | 5.47E-01 | 18314 | -0.032 | -0.071 | 0.006 | 0.020 | -1.648 | -0.012 | 9.94E-02 | 18664 |
| 162 | SN_pc_R | 0.009 | -0.016 | 0.035 | 0.013 | 0.708 | 0.005 | 4.79E-01 | 18314 | -0.035 | -0.086 | 0.015 | 0.026 | -1.368 | -0.010 | 1.71E-01 | 18664 |
| 163 | SN_pr_L | 0.003 | -0.024 | 0.029 | 0.014 | 0.209 | 0.002 | 8.34E-01 | 18314 | -0.019 | -0.071 | 0.034 | 0.027 | -0.696 | -0.005 | 4.86E-01 | 18664 |
| 164 | SN_pr_R | 0.003 | -0.026 | 0.033 | 0.015 | 0.216 | 0.002 | 8.29E-01 | 18314 | -0.017 | -0.076 | 0.042 | 0.030 | -0.556 | -0.004 | 5.78E-01 | 18664 |
| 165 | Red_N_L | 0.033 | 0.001 | 0.066 | 0.017 | 2.003 | 0.015 | 4.52E-02 | 18314 | -0.036 | -0.101 | 0.029 | 0.033 | -1.081 | -0.008 | 2.80E-01 | 18664 |
| 166 | Red_N_R | 0.027 | -0.008 | 0.063 | 0.018 | 1.508 | 0.011 | 1.31E-01 | 18314 | -0.015 | -0.086 | 0.055 | 0.036 | -0.428 | -0.003 | 6.68E-01 | 18664 |
| 167 | LC_L | 0.001 | -0.011 | 0.013 | 0.006 | 0.123 | 0.001 | 9.02E-01 | 18314 | 0.015 | -0.009 | 0.039 | 0.012 | 1.207 | 0.009 | 2.28E-01 | 18664 |
| 168 | LC_R | -0.004 | -0.017 | 0.009 | 0.007 | -0.615 | -0.005 | 5.39E-01 | 18314 | 0.011 | -0.015 | 0.038 | 0.013 | 0.827 | 0.006 | 4.08E-01 | 18664 |
| 169 | Raphe_D | 0.066 | 0.019 | 0.113 | 0.024 | 2.746 | 0.020 | 6.04E-03 | 18314 | -0.007 | -0.101 | 0.087 | 0.048 | -0.151 | -0.001 | 8.80E-01 | 18664 |
| 170 | Raphe_M | 0.008 | -0.006 | 0.023 | 0.007 | 1.141 | 0.008 | 2.54E-01 | 18314 | 0.013 | -0.016 | 0.041 | 0.015 | 0.889 | 0.007 | 3.74E-01 | 18664 |

| | | Bread intake | | | | | | | | Pork intake | | | | | | | |
|---|---|---|---|---|---|---|---|---|---|---|---|---|---|---|---|---|---|
| **Number** | **AAL 3 region** | **beta** | **95%CI_l** | **95%CI_h** | **se** | **t** | **r** | **p** | **df.e** | **beta** | **95%CI_l** | **95%CI_h** | **se** | **t** | **r** | **p** | **df.e** |
| 0 | Total brain | 16.782 | -3.635 | 37.198 | 10.416 | 1.611 | 0.012 | 1.07E-01 | 18371 | -234.354 | -465.599 | -3.108 | 117.977 | -1.986 | -0.015 | 4.70E-02 | 18656 |
| 1 | Precentral_L | 0.105 | -0.615 | 0.824 | 0.367 | 0.285 | 0.002 | 7.76E-01 | 18371 | -2.089 | -10.235 | 6.058 | 4.156 | -0.502 | -0.004 | 6.15E-01 | 18656 |
| 2 | Precentral_R | 0.162 | -0.497 | 0.821 | 0.336 | 0.482 | 0.004 | 6.30E-01 | 18371 | -3.443 | -10.904 | 4.018 | 3.806 | -0.904 | -0.007 | 3.66E-01 | 18656 |
| 3 | Frontal_Sup_2_L | 0.886 | 0.127 | 1.646 | 0.388 | 2.286 | 0.017 | 2.22E-02 | 18371 | -2.165 | -10.768 | 6.438 | 4.389 | -0.493 | -0.004 | 6.22E-01 | 18656 |
| 4 | Frontal_Sup_2_R | 0.632 | -0.206 | 1.469 | 0.427 | 1.479 | 0.011 | 1.39E-01 | 18371 | -6.947 | -16.430 | 2.536 | 4.838 | -1.436 | -0.011 | 1.51E-01 | 18656 |
| 5 | Frontal_Mid_2_L | 0.098 | -0.699 | 0.895 | 0.407 | 0.241 | 0.002 | 8.09E-01 | 18371 | -7.211 | -16.243 | 1.820 | 4.607 | -1.565 | -0.011 | 1.18E-01 | 18656 |
| 6 | Frontal_Mid_2_R | 0.810 | -0.092 | 1.712 | 0.460 | 1.760 | 0.013 | 7.84E-02 | 18371 | -6.625 | -16.845 | 3.596 | 5.214 | -1.270 | -0.009 | 2.04E-01 | 18656 |
| 7 | Frontal_Inf_Oper_L | 0.156 | -0.079 | 0.392 | 0.120 | 1.303 | 0.010 | 1.93E-01 | 18371 | -0.819 | -3.489 | 1.850 | 1.362 | -0.602 | -0.004 | 5.47E-01 | 18656 |
| 8 | Frontal_Inf_Oper_R | 0.251 | -0.044 | 0.547 | 0.151 | 1.666 | 0.012 | 9.57E-02 | 18371 | -2.285 | -5.633 | 1.062 | 1.708 | -1.338 | -0.010 | 1.81E-01 | 18656 |
| 9 | Frontal_Inf_Tri_L | 0.302 | -0.199 | 0.802 | 0.255 | 1.181 | 0.009 | 2.38E-01 | 18371 | -3.927 | -9.594 | 1.740 | 2.891 | -1.358 | -0.010 | 1.74E-01 | 18656 |
| 10 | Frontal_Inf_Tri_R | 0.225 | -0.168 | 0.618 | 0.200 | 1.121 | 0.008 | 2.62E-01 | 18371 | -1.025 | -5.477 | 3.426 | 2.271 | -0.452 | -0.003 | 6.52E-01 | 18656 |

| | | | | | | | | | | | | | | | | | |
|---|---|---|---|---|---|---|---|---|---|---|---|---|---|---|---|---|---|
| 11 | Frontal_Inf_Orb_2_L | 0.132 | -0.018 | 0.281 | 0.076 | 1.726 | 0.013 | 8.44E-02 | 18371 | -1.241 | -2.936 | 0.454 | 0.865 | -1.436 | -0.011 | 1.51E-01 | 18656 |
| 12 | Frontal_Inf_Orb_2_R | 0.092 | -0.065 | 0.248 | 0.080 | 1.148 | 0.008 | 2.51E-01 | 18371 | -1.153 | -2.925 | 0.620 | 0.904 | -1.275 | -0.009 | 2.02E-01 | 18656 |
| 13 | Rolandic_Oper_L | 0.094 | -0.132 | 0.321 | 0.116 | 0.817 | 0.006 | 4.14E-01 | 18371 | -0.783 | -3.347 | 1.781 | 1.308 | -0.599 | -0.004 | 5.49E-01 | 18656 |
| 14 | Rolandic_Oper_R | 0.189 | -0.135 | 0.514 | 0.165 | 1.146 | 0.008 | 2.52E-01 | 18371 | -2.643 | -6.312 | 1.026 | 1.872 | -1.412 | -0.010 | 1.58E-01 | 18656 |
| 15 | Supp_Motor_Area_L | 0.293 | -0.245 | 0.831 | 0.275 | 1.066 | 0.008 | 2.86E-01 | 18371 | 1.029 | -5.064 | 7.122 | 3.109 | 0.331 | 0.002 | 7.41E-01 | 18656 |
| 16 | Supp_Motor_Area_R | 0.289 | -0.305 | 0.884 | 0.303 | 0.955 | 0.007 | 3.40E-01 | 18371 | -2.636 | -9.370 | 4.097 | 3.435 | -0.768 | -0.006 | 4.43E-01 | 18656 |
| 17 | Olfactory_L | 0.072 | -0.008 | 0.152 | 0.041 | 1.767 | 0.013 | 7.73E-02 | 18371 | -0.401 | -1.307 | 0.505 | 0.462 | -0.868 | -0.006 | 3.85E-01 | 18656 |
| 18 | Olfactory_R | 0.075 | -0.005 | 0.155 | 0.041 | 1.839 | 0.014 | 6.59E-02 | 18371 | -0.856 | -1.764 | 0.053 | 0.463 | -1.847 | -0.014 | 6.48E-02 | 18656 |
| 19 | Frontal_Sup_Medial_L | 0.581 | 0.077 | 1.085 | 0.257 | 2.259 | 0.017 | 2.39E-02 | 18371 | -1.693 | -7.405 | 4.020 | 2.914 | -0.581 | -0.004 | 5.61E-01 | 18656 |
| 20 | Frontal_Sup_Medial_R | 0.097 | -0.270 | 0.464 | 0.187 | 0.519 | 0.004 | 6.04E-01 | 18371 | -3.969 | -8.126 | 0.187 | 2.121 | -1.872 | -0.014 | 6.12E-02 | 18656 |
| 21 | Frontal_Med_Orb_L | 0.140 | -0.049 | 0.328 | 0.096 | 1.454 | 0.011 | 1.46E-01 | 18371 | -1.449 | -3.581 | 0.684 | 1.088 | -1.332 | -0.010 | 1.83E-01 | 18656 |
| 22 | Frontal_Med_Orb_R | -0.004 | -0.235 | 0.227 | 0.118 | -0.033 | 0.000 | 9.74E-01 | 18371 | -1.237 | -3.859 | 1.384 | 1.337 | -0.925 | -0.007 | 3.55E-01 | 18656 |
| 23 | Rectus_L | 0.153 | -0.042 | 0.348 | 0.099 | 1.541 | 0.011 | 1.23E-01 | 18371 | -1.628 | -3.836 | 0.580 | 1.126 | -1.445 | -0.011 | 1.48E-01 | 18656 |
| 24 | Rectus_R | 0.145 | -0.036 | 0.325 | 0.092 | 1.573 | 0.012 | 1.16E-01 | 18371 | -0.768 | -2.811 | 1.274 | 1.042 | -0.737 | -0.005 | 4.61E-01 | 18656 |
| 25 | OFCmed_L | -0.001 | -0.114 | 0.112 | 0.058 | -0.014 | 0.000 | 9.89E-01 | 18371 | -0.694 | -1.973 | 0.586 | 0.653 | -1.062 | -0.008 | 2.88E-01 | 18656 |
| 26 | OFCmed_R | 0.054 | -0.078 | 0.185 | 0.067 | 0.799 | 0.006 | 4.24E-01 | 18371 | 0.258 | -1.234 | 1.750 | 0.761 | 0.339 | 0.002 | 7.34E-01 | 18656 |
| 27 | OFCant_L | -0.045 | -0.176 | 0.085 | 0.067 | -0.679 | -0.005 | 4.97E-01 | 18371 | -0.827 | -2.307 | 0.653 | 0.755 | -1.096 | -0.008 | 2.73E-01 | 18656 |
| 28 | OFCant_R | 0.094 | -0.078 | 0.267 | 0.088 | 1.071 | 0.008 | 2.84E-01 | 18371 | -0.626 | -2.582 | 1.329 | 0.998 | -0.628 | -0.005 | 5.30E-01 | 18656 |
| 29 | OFCpost_L | 0.217 | 0.053 | 0.380 | 0.083 | 2.597 | 0.019 | 9.41E-03 | 18371 | -1.148 | -3.000 | 0.705 | 0.945 | -1.214 | -0.009 | 2.25E-01 | 18656 |
| 30 | OFCpost_R | 0.176 | 0.025 | 0.327 | 0.077 | 2.283 | 0.017 | 2.24E-02 | 18371 | -2.005 | -3.717 | -0.293 | 0.874 | -2.295 | -0.017 | 2.17E-02 | 18656 |
| 31 | OFClat_L | 0.022 | -0.033 | 0.077 | 0.028 | 0.784 | 0.006 | 4.33E-01 | 18371 | -0.265 | -0.890 | 0.360 | 0.319 | -0.831 | -0.006 | 4.06E-01 | 18656 |
| 32 | OFClat_R | 0.021 | -0.026 | 0.067 | 0.024 | 0.868 | 0.006 | 3.85E-01 | 18371 | -0.334 | -0.862 | 0.194 | 0.269 | -1.239 | -0.009 | 2.15E-01 | 18656 |
| 33 | Insula_L | 0.671 | 0.165 | 1.177 | 0.258 | 2.599 | 0.019 | 9.37E-03 | 18371 | -7.378 | -13.105 | -1.651 | 2.922 | -2.525 | -0.018 | 1.16E-02 | 18656 |
| 34 | Insula_R | 0.699 | 0.206 | 1.192 | 0.251 | 2.781 | 0.021 | 5.42E-03 | 18371 | -6.198 | -11.780 | -0.616 | 2.848 | -2.176 | -0.016 | 2.95E-02 | 18656 |
| 37 | Cingulate_Mid_L | 0.351 | -0.076 | 0.779 | 0.218 | 1.612 | 0.012 | 1.07E-01 | 18371 | -4.242 | -9.080 | 0.596 | 2.468 | -1.719 | -0.013 | 8.57E-02 | 18656 |
| 38 | Cingulate_Mid_R | 0.499 | 0.044 | 0.954 | 0.232 | 2.149 | 0.016 | 3.17E-02 | 18371 | -3.892 | -9.051 | 1.266 | 2.632 | -1.479 | -0.011 | 1.39E-01 | 18656 |
| 39 | Cingulate_Post_L | 0.102 | -0.029 | 0.233 | 0.067 | 1.531 | 0.011 | 1.26E-01 | 18371 | -1.489 | -2.969 | -0.010 | 0.755 | -1.973 | -0.014 | 4.85E-02 | 18656 |
| 40 | Cingulate_Post_R | 0.031 | -0.042 | 0.103 | 0.037 | 0.836 | 0.006 | 4.03E-01 | 18371 | -0.240 | -1.060 | 0.581 | 0.419 | -0.572 | -0.004 | 5.67E-01 | 18656 |
| 41 | Hippocampus_L | 0.268 | 0.056 | 0.479 | 0.108 | 2.483 | 0.018 | 1.30E-02 | 18371 | -2.585 | -4.977 | -0.193 | 1.221 | -2.118 | -0.016 | 3.42E-02 | 18656 |
| 42 | Hippocampus_R | 0.178 | -0.041 | 0.397 | 0.112 | 1.591 | 0.012 | 1.12E-01 | 18371 | -3.828 | -6.307 | -1.349 | 1.265 | -3.027 | -0.022 | 2.48E-03 | 18656 |
| 43 | ParaHippocampal_L | 0.208 | 0.020 | 0.396 | 0.096 | 2.173 | 0.016 | 2.98E-02 | 18371 | -2.299 | -4.424 | -0.173 | 1.084 | -2.120 | -0.016 | 3.40E-02 | 18656 |
| 44 | ParaHippocampal_R | 0.284 | 0.047 | 0.520 | 0.121 | 2.348 | 0.017 | 1.89E-02 | 18371 | -4.637 | -7.318 | -1.956 | 1.368 | -3.390 | -0.025 | 7.00E-04 | 18656 |
| 45 | Amygdala_L | 0.126 | 0.053 | 0.199 | 0.037 | 3.393 | 0.025 | 6.92E-04 | 18371 | -0.492 | -1.318 | 0.334 | 0.421 | -1.168 | -0.009 | 2.43E-01 | 18656 |
| 46 | Amygdala_R | 0.116 | 0.042 | 0.191 | 0.038 | 3.057 | 0.023 | 2.24E-03 | 18371 | -0.891 | -1.736 | -0.046 | 0.431 | -2.067 | -0.015 | 3.87E-02 | 18656 |
| 47 | Calcarine_L | -0.159 | -0.855 | 0.538 | 0.355 | -0.446 | -0.003 | 6.55E-01 | 18371 | -0.852 | -8.743 | 7.038 | 4.026 | -0.212 | -0.002 | 8.32E-01 | 18656 |
| 48 | Calcarine_R | -0.230 | -0.812 | 0.351 | 0.297 | -0.776 | -0.006 | 4.38E-01 | 18371 | -0.888 | -7.484 | 5.708 | 3.365 | -0.264 | -0.002 | 7.92E-01 | 18656 |
| 49 | Cuneus_L | -0.337 | -0.774 | 0.100 | 0.223 | -1.511 | -0.011 | 1.31E-01 | 18371 | 1.259 | -3.686 | 6.203 | 2.523 | 0.499 | 0.004 | 6.18E-01 | 18656 |
| 50 | Cuneus_R | -0.131 | -0.465 | 0.203 | 0.170 | -0.771 | -0.006 | 4.41E-01 | 18371 | 0.261 | -3.523 | 4.045 | 1.930 | 0.135 | 0.001 | 8.92E-01 | 18656 |
| 51 | Lingual_L | -0.046 | -0.465 | 0.373 | 0.214 | -0.216 | -0.002 | 8.29E-01 | 18371 | -0.582 | -5.327 | 4.164 | 2.421 | -0.240 | -0.002 | 8.10E-01 | 18656 |
| 52 | Lingual_R | 0.109 | -0.329 | 0.548 | 0.224 | 0.489 | 0.004 | 6.25E-01 | 18371 | -1.838 | -6.811 | 3.134 | 2.537 | -0.725 | -0.005 | 4.69E-01 | 18656 |
| 53 | Occipital_Sup_L | -0.040 | -0.328 | 0.248 | 0.147 | -0.274 | -0.002 | 7.84E-01 | 18371 | -0.870 | -4.131 | 2.392 | 1.664 | -0.523 | -0.004 | 6.01E-01 | 18656 |
| 54 | Occipital_Sup_R | 0.214 | -0.099 | 0.526 | 0.159 | 1.342 | 0.010 | 1.80E-01 | 18371 | -0.783 | -4.318 | 2.751 | 1.803 | -0.434 | -0.003 | 6.64E-01 | 18656 |
| 55 | Occipital_Mid_L | 0.279 | -0.396 | 0.954 | 0.344 | 0.811 | 0.006 | 4.18E-01 | 18371 | -1.344 | -8.986 | 6.299 | 3.899 | -0.345 | -0.003 | 7.30E-01 | 18656 |
| 56 | Occipital_Mid_R | 0.398 | -0.071 | 0.867 | 0.239 | 1.662 | 0.012 | 9.65E-02 | 18371 | -1.061 | -6.381 | 4.259 | 2.714 | -0.391 | -0.003 | 6.96E-01 | 18656 |
| 57 | Occipital_Inf_L | 0.135 | -0.090 | 0.359 | 0.114 | 1.176 | 0.009 | 2.40E-01 | 18371 | -2.079 | -4.618 | 0.459 | 1.295 | -1.605 | -0.012 | 1.08E-01 | 18656 |
| 58 | Occipital_Inf_R | 0.362 | 0.103 | 0.621 | 0.132 | 2.738 | 0.020 | 6.19E-03 | 18371 | 0.719 | -2.212 | 3.649 | 1.495 | 0.481 | 0.004 | 6.31E-01 | 18656 |
| 59 | Fusiform_L | -0.196 | -0.687 | 0.294 | 0.250 | -0.785 | -0.006 | 4.33E-01 | 18371 | -3.653 | -9.205 | 1.900 | 2.833 | -1.289 | -0.009 | 1.97E-01 | 18656 |
| 60 | Fusiform_R | 0.139 | -0.400 | 0.678 | 0.275 | 0.504 | 0.004 | 6.14E-01 | 18371 | -4.605 | -10.702 | 1.493 | 3.111 | -1.480 | -0.011 | 1.39E-01 | 18656 |
| 61 | Postcentral_L | 0.213 | -0.525 | 0.952 | 0.377 | 0.567 | 0.004 | 5.71E-01 | 18371 | 0.709 | -7.635 | 9.054 | 4.257 | 0.167 | 0.001 | 8.68E-01 | 18656 |

| | | | | | | | | | | | | | | | | | |
|---|---|---|---|---|---|---|---|---|---|---|---|---|---|---|---|---|---|
| 62 | Postcentral_R | -0.072 | -0.829 | 0.685 | 0.386 | -0.185 | -0.001 | 8.53E-01 | 18371 | -1.083 | -9.647 | 7.481 | 4.369 | -0.248 | -0.002 | 8.04E-01 | 18656 |
| 63 | Parietal_Sup_L | -0.004 | -0.430 | 0.421 | 0.217 | -0.021 | 0.000 | 9.83E-01 | 18371 | -0.901 | -5.713 | 3.911 | 2.455 | -0.367 | -0.003 | 7.14E-01 | 18656 |
| 64 | Parietal_Sup_R | 0.352 | -0.026 | 0.729 | 0.193 | 1.827 | 0.013 | 6.77E-02 | 18371 | -3.874 | -8.141 | 0.393 | 2.177 | -1.780 | -0.013 | 7.52E-02 | 18656 |
| 65 | Parietal_Inf_L | -0.025 | -0.582 | 0.532 | 0.284 | -0.088 | -0.001 | 9.30E-01 | 18371 | -2.086 | -8.384 | 4.212 | 3.213 | -0.649 | -0.005 | 5.16E-01 | 18656 |
| 66 | Parietal_Inf_R | -0.002 | -0.362 | 0.359 | 0.184 | -0.009 | 0.000 | 9.93E-01 | 18371 | -0.859 | -4.941 | 3.223 | 2.083 | -0.412 | -0.003 | 6.80E-01 | 18656 |
| 67 | SupraMarginal_L | 0.189 | -0.137 | 0.515 | 0.166 | 1.134 | 0.008 | 2.57E-01 | 18371 | -2.054 | -5.745 | 1.636 | 1.883 | -1.091 | -0.008 | 2.75E-01 | 18656 |
| 68 | SupraMarginal_R | 0.206 | -0.255 | 0.667 | 0.235 | 0.876 | 0.006 | 3.81E-01 | 18371 | -3.088 | -8.314 | 2.139 | 2.667 | -1.158 | -0.008 | 2.47E-01 | 18656 |
| 69 | Angular_L | 0.053 | -0.267 | 0.374 | 0.164 | 0.326 | 0.002 | 7.44E-01 | 18371 | -2.322 | -5.946 | 1.302 | 1.849 | -1.256 | -0.009 | 2.09E-01 | 18656 |
| 70 | Angular_R | -0.261 | -0.690 | 0.169 | 0.219 | -1.189 | -0.009 | 2.34E-01 | 18371 | -1.299 | -6.163 | 3.564 | 2.481 | -0.524 | -0.004 | 6.01E-01 | 18656 |
| 71 | Precuneus_L | 0.051 | -0.730 | 0.832 | 0.399 | 0.127 | 0.001 | 8.99E-01 | 18371 | -3.705 | -12.545 | 5.135 | 4.510 | -0.822 | -0.006 | 4.11E-01 | 18656 |
| 72 | Precuneus_R | 0.061 | -0.634 | 0.757 | 0.355 | 0.173 | 0.001 | 8.63E-01 | 18371 | -3.664 | -11.540 | 4.212 | 4.018 | -0.912 | -0.007 | 3.62E-01 | 18656 |
| 73 | Paracentral_Lobule_L | -0.075 | -0.432 | 0.283 | 0.182 | -0.409 | -0.003 | 6.82E-01 | 18371 | -0.190 | -4.240 | 3.860 | 2.066 | -0.092 | -0.001 | 9.27E-01 | 18656 |
| 74 | Paracentral_Lobule_R | 0.046 | -0.210 | 0.302 | 0.130 | 0.354 | 0.003 | 7.23E-01 | 18371 | 0.941 | -1.961 | 3.842 | 1.480 | 0.635 | 0.005 | 5.25E-01 | 18656 |
| 75 | Caudate_L | 0.419 | 0.134 | 0.704 | 0.145 | 2.880 | 0.021 | 3.98E-03 | 18371 | -2.544 | -5.778 | 0.691 | 1.650 | -1.542 | -0.011 | 1.23E-01 | 18656 |
| 76 | Caudate_R | 0.484 | 0.161 | 0.807 | 0.165 | 2.938 | 0.022 | 3.31E-03 | 18371 | -3.450 | -7.112 | 0.211 | 1.868 | -1.847 | -0.014 | 6.47E-02 | 18656 |
| 77 | Putamen_L | 0.550 | 0.117 | 0.983 | 0.221 | 2.490 | 0.018 | 1.28E-02 | 18371 | -2.174 | -7.084 | 2.736 | 2.505 | -0.868 | -0.006 | 3.85E-01 | 18656 |
| 78 | Putamen_R | 0.434 | -0.008 | 0.875 | 0.225 | 1.927 | 0.014 | 5.40E-02 | 18371 | -2.791 | -7.791 | 2.208 | 2.551 | -1.094 | -0.008 | 2.74E-01 | 18656 |
| 79 | Pallidum_L | 0.025 | -0.014 | 0.064 | 0.020 | 1.273 | 0.009 | 2.03E-01 | 18371 | -0.204 | -0.641 | 0.233 | 0.223 | -0.915 | -0.007 | 3.60E-01 | 18656 |
| 80 | Pallidum_R | 0.034 | -0.009 | 0.076 | 0.022 | 1.555 | 0.011 | 1.20E-01 | 18371 | -0.364 | -0.844 | 0.115 | 0.245 | -1.490 | -0.011 | 1.36E-01 | 18656 |
| 83 | Heschl_L | 0.046 | -0.033 | 0.124 | 0.040 | 1.142 | 0.008 | 2.53E-01 | 18371 | -0.381 | -1.266 | 0.504 | 0.451 | -0.844 | -0.006 | 3.99E-01 | 18656 |
| 84 | Heschl_R | 0.049 | -0.039 | 0.136 | 0.045 | 1.089 | 0.008 | 2.76E-01 | 18371 | -1.030 | -2.019 | -0.041 | 0.505 | -2.041 | -0.015 | 4.12E-02 | 18656 |
| 85 | Temporal_Sup_L | 0.847 | 0.282 | 1.411 | 0.288 | 2.940 | 0.022 | 3.29E-03 | 18371 | -5.581 | -11.967 | 0.806 | 3.258 | -1.713 | -0.013 | 8.68E-02 | 18656 |
| 86 | Temporal_Sup_R | 0.407 | -0.217 | 1.031 | 0.318 | 1.278 | 0.009 | 2.01E-01 | 18371 | -7.445 | -14.510 | -0.380 | 3.605 | -2.065 | -0.015 | 3.89E-02 | 18656 |
| 87 | Temporal_Pole_Sup_L | 0.371 | 0.105 | 0.638 | 0.136 | 2.729 | 0.020 | 6.36E-03 | 18371 | -3.372 | -6.397 | -0.347 | 1.543 | -2.185 | -0.016 | 2.89E-02 | 18656 |
| 88 | Temporal_Pole_Sup_R | 0.483 | 0.202 | 0.764 | 0.143 | 3.370 | 0.025 | 7.53E-04 | 18371 | -3.867 | -7.051 | -0.683 | 1.624 | -2.381 | -0.017 | 1.73E-02 | 18656 |
| 89 | Temporal_Mid_L | 0.240 | -0.700 | 1.180 | 0.480 | 0.501 | 0.004 | 6.17E-01 | 18371 | -13.626 | -24.269 | -2.984 | 5.430 | -2.510 | -0.018 | 1.21E-02 | 18656 |
| 90 | Temporal_Mid_R | 0.406 | -0.430 | 1.242 | 0.427 | 0.952 | 0.007 | 3.41E-01 | 18371 | -5.832 | -15.278 | 3.614 | 4.819 | -1.210 | -0.009 | 2.26E-01 | 18656 |
| 91 | Temporal_Pole_Mid_L | 0.097 | -0.128 | 0.321 | 0.114 | 0.845 | 0.006 | 3.98E-01 | 18371 | -2.832 | -5.376 | -0.288 | 1.298 | -2.182 | -0.016 | 2.91E-02 | 18656 |
| 92 | Temporal_Pole_Mid_R | 0.360 | 0.088 | 0.633 | 0.139 | 2.590 | 0.019 | 9.60E-03 | 18371 | -3.182 | -6.272 | -0.091 | 1.577 | -2.018 | -0.015 | 4.36E-02 | 18656 |
| 93 | Temporal_Inf_L | -0.265 | -0.912 | 0.382 | 0.330 | -0.802 | -0.006 | 4.23E-01 | 18371 | -6.230 | -13.544 | 1.084 | 3.731 | -1.670 | -0.012 | 9.50E-02 | 18656 |
| 94 | Temporal_Inf_R | 0.307 | -0.399 | 1.012 | 0.360 | 0.852 | 0.006 | 3.94E-01 | 18371 | -2.722 | -10.706 | 5.262 | 4.073 | -0.668 | -0.005 | 5.04E-01 | 18656 |
| 95 | Cerebellum_Crus1_L | -0.415 | -1.183 | 0.352 | 0.392 | -1.061 | -0.008 | 2.89E-01 | 18371 | -1.093 | -9.793 | 7.606 | 4.438 | -0.246 | -0.002 | 8.05E-01 | 18656 |
| 96 | Cerebellum_Crus1_R | -0.301 | -1.084 | 0.482 | 0.399 | -0.753 | -0.006 | 4.51E-01 | 18371 | 2.093 | -6.779 | 10.966 | 4.527 | 0.462 | 0.003 | 6.44E-01 | 18656 |
| 97 | Cerebellum_Crus2_L | -0.102 | -0.711 | 0.507 | 0.311 | -0.329 | -0.002 | 7.42E-01 | 18371 | -5.931 | -12.830 | 0.967 | 3.519 | -1.685 | -0.012 | 9.19E-02 | 18656 |
| 98 | Cerebellum_Crus2_R | 0.001 | -0.561 | 0.564 | 0.287 | 0.005 | 0.000 | 9.96E-01 | 18371 | -2.641 | -9.023 | 3.741 | 3.256 | -0.811 | -0.006 | 4.17E-01 | 18656 |
| 99 | Cerebellum_3_L | 0.030 | -0.011 | 0.070 | 0.021 | 1.419 | 0.010 | 1.56E-01 | 18371 | 0.359 | -0.105 | 0.822 | 0.236 | 1.517 | 0.011 | 1.29E-01 | 18656 |
| 100 | Cerebellum_3_R | 0.029 | -0.027 | 0.084 | 0.028 | 1.015 | 0.007 | 3.10E-01 | 18371 | 0.295 | -0.330 | 0.920 | 0.319 | 0.925 | 0.007 | 3.55E-01 | 18656 |
| 101 | Cerebellum_4_5_L | 0.088 | -0.242 | 0.419 | 0.169 | 0.524 | 0.004 | 6.00E-01 | 18371 | 3.527 | -0.221 | 7.274 | 1.912 | 1.845 | 0.014 | 6.51E-02 | 18656 |
| 102 | Cerebellum_4_5_R | 0.169 | -0.100 | 0.438 | 0.137 | 1.232 | 0.009 | 2.18E-01 | 18371 | 2.005 | -1.048 | 5.057 | 1.557 | 1.287 | 0.009 | 1.98E-01 | 18656 |
| 103 | Cerebellum_6_L | -0.390 | -0.962 | 0.181 | 0.291 | -1.340 | -0.010 | 1.80E-01 | 18371 | 0.075 | -6.401 | 6.551 | 3.304 | 0.023 | 0.000 | 9.82E-01 | 18656 |
| 104 | Cerebellum_6_R | -0.195 | -0.797 | 0.407 | 0.307 | -0.636 | -0.005 | 5.24E-01 | 18371 | 2.503 | -4.319 | 9.325 | 3.481 | 0.719 | 0.005 | 4.72E-01 | 18656 |
| 105 | Cerebellum_7b_L | -0.061 | -0.247 | 0.125 | 0.095 | -0.642 | -0.005 | 5.21E-01 | 18371 | -1.233 | -3.341 | 0.876 | 1.076 | -1.146 | -0.008 | 2.52E-01 | 18656 |
| 106 | Cerebellum_7b_R | -0.012 | -0.196 | 0.172 | 0.094 | -0.127 | -0.001 | 8.99E-01 | 18371 | -1.668 | -3.758 | 0.422 | 1.066 | -1.565 | -0.011 | 1.18E-01 | 18656 |
| 107 | Cerebellum_8_L | -0.143 | -0.733 | 0.447 | 0.301 | -0.475 | -0.004 | 6.35E-01 | 18371 | -2.409 | -9.104 | 4.286 | 3.415 | -0.705 | -0.005 | 4.81E-01 | 18656 |
| 108 | Cerebellum_8_R | 0.113 | -0.655 | 0.882 | 0.392 | 0.288 | 0.002 | 7.73E-01 | 18371 | -4.257 | -12.971 | 4.458 | 4.446 | -0.957 | -0.007 | 3.38E-01 | 18656 |
| 109 | Cerebellum_9_L | -0.025 | -0.359 | 0.308 | 0.170 | -0.148 | -0.001 | 8.83E-01 | 18371 | -2.030 | -5.817 | 1.756 | 1.932 | -1.051 | -0.008 | 2.93E-01 | 18656 |
| 110 | Cerebellum_9_R | 0.081 | -0.247 | 0.409 | 0.167 | 0.484 | 0.004 | 6.29E-01 | 18371 | -1.150 | -4.872 | 2.572 | 1.899 | -0.606 | -0.004 | 5.45E-01 | 18656 |
| 111 | Cerebellum_10_L | 0.015 | -0.009 | 0.039 | 0.012 | 1.233 | 0.009 | 2.18E-01 | 18371 | 0.085 | -0.184 | 0.354 | 0.137 | 0.622 | 0.005 | 5.34E-01 | 18656 |
| 112 | Cerebellum_10_R | 0.023 | -0.002 | 0.048 | 0.013 | 1.831 | 0.014 | 6.71E-02 | 18371 | 0.199 | -0.080 | 0.479 | 0.143 | 1.399 | 0.010 | 1.62E-01 | 18656 |

| | | | | | | | | | | | | | | | | | |
|---|---|---|---|---|---|---|---|---|---|---|---|---|---|---|---|---|---|
| 113 | Vermis_1_2 | 0.014 | -0.001 | 0.030 | 0.008 | 1.840 | 0.014 | 6.58E-02 | 18371 | 0.042 | -0.133 | 0.217 | 0.089 | 0.469 | 0.003 | 6.39E-01 | 18656 |
| 114 | Vermis_3 | 0.064 | 0.002 | 0.125 | 0.031 | 2.033 | 0.015 | 4.21E-02 | 18371 | 0.213 | -0.482 | 0.907 | 0.354 | 0.600 | 0.004 | 5.48E-01 | 18656 |
| 115 | Vermis_4_5 | 0.149 | -0.031 | 0.329 | 0.092 | 1.618 | 0.012 | 1.06E-01 | 18371 | 2.281 | 0.241 | 4.322 | 1.041 | 2.192 | 0.016 | 2.84E-02 | 18656 |
| 116 | Vermis_6 | -0.011 | -0.111 | 0.090 | 0.051 | -0.210 | -0.002 | 8.34E-01 | 18371 | 0.430 | -0.706 | 1.567 | 0.580 | 0.742 | 0.005 | 4.58E-01 | 18656 |
| 117 | Vermis_7 | -0.017 | -0.074 | 0.040 | 0.029 | -0.587 | -0.004 | 5.57E-01 | 18371 | 0.102 | -0.540 | 0.744 | 0.328 | 0.310 | 0.002 | 7.56E-01 | 18656 |
| 118 | Vermis_8 | -0.044 | -0.137 | 0.049 | 0.047 | -0.922 | -0.007 | 3.57E-01 | 18371 | -0.493 | -1.548 | 0.563 | 0.538 | -0.915 | -0.007 | 3.60E-01 | 18656 |
| 119 | Vermis_9 | 0.000 | -0.075 | 0.076 | 0.038 | 0.006 | 0.000 | 9.96E-01 | 18371 | -0.207 | -1.062 | 0.648 | 0.436 | -0.474 | -0.003 | 6.35E-01 | 18656 |
| 120 | Vermis_10 | 0.012 | -0.009 | 0.033 | 0.011 | 1.122 | 0.008 | 2.62E-01 | 18371 | -0.050 | -0.288 | 0.189 | 0.122 | -0.409 | -0.003 | 6.83E-01 | 18656 |
| 121 | Thal_AV_L | 0.008 | 0.000 | 0.017 | 0.004 | 1.968 | 0.015 | 4.91E-02 | 18371 | -0.065 | -0.161 | 0.030 | 0.049 | -1.342 | -0.010 | 1.80E-01 | 18656 |
| 122 | Thal_AV_R | 0.007 | -0.005 | 0.019 | 0.006 | 1.109 | 0.008 | 2.67E-01 | 18371 | -0.120 | -0.256 | 0.017 | 0.070 | -1.718 | -0.013 | 8.59E-02 | 18656 |
| 123 | Thal_LP_L | 0.012 | 0.003 | 0.021 | 0.005 | 2.520 | 0.019 | 1.17E-02 | 18371 | -0.085 | -0.191 | 0.020 | 0.054 | -1.582 | -0.012 | 1.14E-01 | 18656 |
| 124 | Thal_LP_R | 0.006 | -0.006 | 0.018 | 0.006 | 0.979 | 0.007 | 3.27E-01 | 18371 | -0.172 | -0.310 | -0.034 | 0.070 | -2.451 | -0.018 | 1.43E-02 | 18656 |
| 125 | Thal_VA_L | 0.007 | -0.003 | 0.017 | 0.005 | 1.336 | 0.010 | 1.82E-01 | 18371 | -0.060 | -0.172 | 0.053 | 0.058 | -1.035 | -0.008 | 3.01E-01 | 18656 |
| 126 | Thal_VA_R | 0.005 | -0.007 | 0.017 | 0.006 | 0.821 | 0.006 | 4.12E-01 | 18371 | -0.099 | -0.237 | 0.040 | 0.071 | -1.397 | -0.010 | 1.63E-01 | 18656 |
| 127 | Thal_VL_L | 0.051 | 0.023 | 0.079 | 0.014 | 3.523 | 0.026 | 4.28E-04 | 18371 | -0.243 | -0.564 | 0.078 | 0.164 | -1.484 | -0.011 | 1.38E-01 | 18656 |
| 128 | Thal_VL_R | 0.038 | 0.000 | 0.075 | 0.019 | 1.962 | 0.014 | 4.98E-02 | 18371 | -0.483 | -0.909 | -0.056 | 0.218 | -2.216 | -0.016 | 2.67E-02 | 18656 |
| 129 | Thal_VPL_L | 0.037 | 0.003 | 0.071 | 0.017 | 2.136 | 0.016 | 3.27E-02 | 18371 | -0.283 | -0.665 | 0.100 | 0.195 | -1.449 | -0.011 | 1.47E-01 | 18656 |
| 130 | Thal_VPL_R | 0.019 | -0.017 | 0.054 | 0.018 | 1.039 | 0.008 | 2.99E-01 | 18371 | -0.359 | -0.761 | 0.044 | 0.205 | -1.748 | -0.013 | 8.05E-02 | 18656 |
| 131 | Thal_IL_L | 0.021 | 0.006 | 0.036 | 0.007 | 2.818 | 0.021 | 4.83E-03 | 18371 | -0.277 | -0.443 | -0.112 | 0.084 | -3.283 | -0.024 | 1.03E-03 | 18656 |
| 132 | Thal_IL_R | 0.015 | -0.002 | 0.031 | 0.008 | 1.742 | 0.013 | 8.15E-02 | 18371 | -0.260 | -0.448 | -0.073 | 0.096 | -2.720 | -0.020 | 6.54E-03 | 18656 |
| 133 | Thal_Re_L | 0.000 | 0.000 | 0.000 | 0.000 | 0.526 | 0.004 | 5.99E-01 | 18371 | 0.000 | -0.001 | 0.001 | 0.000 | -0.349 | -0.003 | 7.27E-01 | 18656 |
| 134 | Thal_Re_R | 0.000 | 0.000 | 0.001 | 0.000 | 0.984 | 0.007 | 3.25E-01 | 18371 | -0.004 | -0.010 | 0.001 | 0.003 | -1.463 | -0.011 | 1.44E-01 | 18656 |
| 135 | Thal_MDm_L | 0.121 | 0.013 | 0.229 | 0.055 | 2.200 | 0.016 | 2.78E-02 | 18371 | -1.650 | -2.875 | -0.424 | 0.625 | -2.638 | -0.019 | 8.34E-03 | 18656 |
| 136 | Thal_MDm_R | 0.097 | -0.029 | 0.222 | 0.064 | 1.510 | 0.011 | 1.31E-01 | 18371 | -1.740 | -3.164 | -0.316 | 0.726 | -2.395 | -0.018 | 1.66E-02 | 18656 |
| 137 | Thal_MDl_L | 0.027 | 0.009 | 0.046 | 0.010 | 2.864 | 0.021 | 4.18E-03 | 18371 | -0.288 | -0.500 | -0.076 | 0.108 | -2.663 | -0.019 | 7.76E-03 | 18656 |
| 138 | Thal_MDl_R | 0.018 | -0.003 | 0.040 | 0.011 | 1.686 | 0.012 | 9.18E-02 | 18371 | -0.302 | -0.543 | -0.061 | 0.123 | -2.454 | -0.018 | 1.41E-02 | 18656 |
| 139 | Thal_LGN_L | 0.002 | -0.001 | 0.005 | 0.001 | 1.371 | 0.010 | 1.70E-01 | 18371 | -0.042 | -0.074 | -0.010 | 0.016 | -2.565 | -0.019 | 1.03E-02 | 18656 |
| 140 | Thal_LGN_R | 0.006 | 0.000 | 0.012 | 0.003 | 1.943 | 0.014 | 5.20E-02 | 18371 | -0.109 | -0.177 | -0.040 | 0.035 | -3.102 | -0.023 | 1.92E-03 | 18656 |
| 141 | Thal_MGN_L | 0.003 | -0.002 | 0.008 | 0.003 | 1.288 | 0.010 | 1.98E-01 | 18371 | -0.078 | -0.135 | -0.020 | 0.029 | -2.658 | -0.019 | 7.86E-03 | 18656 |
| 142 | Thal_MGN_R | 0.004 | -0.002 | 0.009 | 0.003 | 1.332 | 0.010 | 1.83E-01 | 18371 | -0.098 | -0.161 | -0.036 | 0.032 | -3.085 | -0.023 | 2.04E-03 | 18656 |
| 143 | Thal_PuA_L | 0.005 | -0.001 | 0.011 | 0.003 | 1.591 | 0.012 | 1.12E-01 | 18371 | -0.109 | -0.177 | -0.041 | 0.035 | -3.148 | -0.023 | 1.65E-03 | 18656 |
| 144 | Thal_PuA_R | 0.009 | -0.003 | 0.021 | 0.006 | 1.411 | 0.010 | 1.58E-01 | 18371 | -0.225 | -0.366 | -0.084 | 0.072 | -3.122 | -0.023 | 1.80E-03 | 18656 |
| 145 | Thal_PuM_L | 0.092 | -0.027 | 0.211 | 0.061 | 1.519 | 0.011 | 1.29E-01 | 18371 | -1.590 | -2.939 | -0.241 | 0.688 | -2.311 | -0.017 | 2.09E-02 | 18656 |
| 146 | Thal_PuM_R | 0.095 | -0.053 | 0.244 | 0.076 | 1.257 | 0.009 | 2.09E-01 | 18371 | -2.223 | -3.904 | -0.543 | 0.857 | -2.593 | -0.019 | 9.51E-03 | 18656 |
| 147 | Thal_PuL_L | 0.019 | -0.002 | 0.041 | 0.011 | 1.757 | 0.013 | 7.89E-02 | 18371 | -0.286 | -0.532 | -0.040 | 0.126 | -2.279 | -0.017 | 2.27E-02 | 18656 |
| 148 | Thal_PuL_R | 0.020 | -0.015 | 0.054 | 0.018 | 1.117 | 0.008 | 2.64E-01 | 18371 | -0.473 | -0.864 | -0.082 | 0.200 | -2.369 | -0.017 | 1.79E-02 | 18656 |
| 149 | Thal_PuI_L | 0.003 | -0.003 | 0.009 | 0.003 | 1.012 | 0.007 | 3.12E-01 | 18371 | -0.016 | -0.083 | 0.051 | 0.034 | -0.460 | -0.003 | 6.46E-01 | 18656 |
| 150 | Thal_PuI_R | 0.002 | -0.006 | 0.009 | 0.004 | 0.379 | 0.003 | 7.05E-01 | 18371 | -0.067 | -0.156 | 0.022 | 0.045 | -1.483 | -0.011 | 1.38E-01 | 18656 |
| 151 | ACC_sub_L | -0.023 | -0.092 | 0.045 | 0.035 | -0.668 | -0.005 | 5.04E-01 | 18371 | -0.404 | -1.176 | 0.368 | 0.394 | -1.026 | -0.008 | 3.05E-01 | 18656 |
| 152 | ACC_sub_R | -0.008 | -0.072 | 0.056 | 0.033 | -0.253 | -0.002 | 8.00E-01 | 18371 | -0.427 | -1.149 | 0.295 | 0.368 | -1.160 | -0.008 | 2.46E-01 | 18656 |
| 153 | ACC_pre_L | 0.004 | -0.170 | 0.179 | 0.089 | 0.047 | 0.000 | 9.63E-01 | 18371 | -1.327 | -3.305 | 0.651 | 1.009 | -1.315 | -0.010 | 1.88E-01 | 18656 |
| 154 | ACC_pre_R | 0.050 | -0.118 | 0.218 | 0.086 | 0.581 | 0.004 | 5.61E-01 | 18371 | -1.367 | -3.273 | 0.539 | 0.972 | -1.406 | -0.010 | 1.60E-01 | 18656 |
| 155 | ACC_sup_L | 0.133 | -0.057 | 0.323 | 0.097 | 1.372 | 0.010 | 1.70E-01 | 18371 | -0.628 | -2.777 | 1.522 | 1.097 | -0.572 | -0.004 | 5.67E-01 | 18656 |
| 156 | ACC_sup_R | 0.058 | -0.138 | 0.255 | 0.100 | 0.581 | 0.004 | 5.61E-01 | 18371 | -1.084 | -3.313 | 1.145 | 1.137 | -0.953 | -0.007 | 3.40E-01 | 18656 |
| 157 | N_Acc_L | 0.080 | 0.023 | 0.137 | 0.029 | 2.753 | 0.020 | 5.91E-03 | 18371 | -0.422 | -1.069 | 0.225 | 0.330 | -1.278 | -0.009 | 2.01E-01 | 18656 |
| 158 | N_Acc_R | 0.058 | 0.009 | 0.107 | 0.025 | 2.308 | 0.017 | 2.10E-02 | 18371 | -0.561 | -1.121 | -0.001 | 0.286 | -1.965 | -0.014 | 4.95E-02 | 18656 |
| 159 | VTA_L | 0.001 | -0.001 | 0.003 | 0.001 | 0.977 | 0.007 | 3.29E-01 | 18371 | 0.006 | -0.017 | 0.029 | 0.012 | 0.498 | 0.004 | 6.18E-01 | 18656 |
| 160 | VTA_R | 0.001 | -0.001 | 0.003 | 0.001 | 1.021 | 0.008 | 3.07E-01 | 18371 | 0.002 | -0.018 | 0.023 | 0.010 | 0.202 | 0.001 | 8.40E-01 | 18656 |
| 161 | SN_pc_L | 0.002 | -0.001 | 0.006 | 0.002 | 1.381 | 0.010 | 1.67E-01 | 18371 | -0.012 | -0.049 | 0.024 | 0.019 | -0.665 | -0.005 | 5.06E-01 | 18656 |

| | | | | | | | | | | | | | | | | | |
|---|---|---|---|---|---|---|---|---|---|---|---|---|---|---|---|---|---|
| 162 | SN_pc_R | 0.005 | 0.001 | 0.009 | 0.002 | 2.379 | 0.018 | 1.73E-02 | 18371 | -0.035 | -0.083 | 0.013 | 0.024 | -1.432 | -0.010 | 1.52E-01 | 18656 |
| 163 | SN_pr_L | 0.005 | 0.000 | 0.009 | 0.002 | 2.039 | 0.015 | 4.15E-02 | 18371 | -0.022 | -0.071 | 0.028 | 0.025 | -0.861 | -0.006 | 3.90E-01 | 18656 |
| 164 | SN_pr_R | 0.008 | 0.003 | 0.013 | 0.003 | 3.049 | 0.022 | 2.30E-03 | 18371 | -0.049 | -0.104 | 0.007 | 0.028 | -1.708 | -0.013 | 8.76E-02 | 18656 |
| 165 | Red_N_L | 0.004 | -0.001 | 0.009 | 0.003 | 1.469 | 0.011 | 1.42E-01 | 18371 | -0.039 | -0.100 | 0.023 | 0.031 | -1.238 | -0.009 | 2.16E-01 | 18656 |
| 166 | Red_N_R | 0.005 | -0.001 | 0.011 | 0.003 | 1.705 | 0.013 | 8.83E-02 | 18371 | -0.016 | -0.083 | 0.051 | 0.034 | -0.476 | -0.003 | 6.34E-01 | 18656 |
| 167 | LC_L | 0.002 | 0.000 | 0.004 | 0.001 | 1.780 | 0.013 | 7.50E-02 | 18371 | 0.007 | -0.016 | 0.030 | 0.012 | 0.607 | 0.004 | 5.44E-01 | 18656 |
| 168 | LC_R | 0.002 | 0.000 | 0.004 | 0.001 | 1.647 | 0.012 | 9.96E-02 | 18371 | 0.008 | -0.017 | 0.033 | 0.013 | 0.618 | 0.005 | 5.36E-01 | 18656 |
| 169 | Raphe_D | 0.005 | -0.003 | 0.012 | 0.004 | 1.131 | 0.008 | 2.58E-01 | 18371 | -0.039 | -0.128 | 0.050 | 0.045 | -0.855 | -0.006 | 3.93E-01 | 18656 |
| 170 | Raphe_M | 0.002 | 0.000 | 0.004 | 0.001 | 1.726 | 0.013 | 8.44E-02 | 18371 | 0.009 | -0.018 | 0.036 | 0.014 | 0.688 | 0.005 | 4.92E-01 | 18656 |

| | | Oily fish intake | | | | | | | | Cheese intake | | | | | | | |
|---|---|---|---|---|---|---|---|---|---|---|---|---|---|---|---|---|---|
| **Number** | **AAL 3 region** | **beta** | **95%CI_l** | **95%CI_h** | **se** | **t** | **r** | **p** | **df.e** | **beta** | **95%CI_l** | **95%CI_h** | **se** | **t** | **r** | **p** | **df.e** |
| 0 | Total brain | -237.794 | -423.876 | -51.711 | 94.936 | -2.505 | -0.018 | 1.23E-02 | 18696 | 63.960 | -91.971 | 219.890 | 79.553 | 0.804 | 0.006 | 4.21E-01 | 18245 |
| 1 | Precentral_L | -3.397 | -9.944 | 3.150 | 3.340 | -1.017 | -0.007 | 3.09E-01 | 18696 | 0.909 | -4.570 | 6.388 | 2.795 | 0.325 | 0.002 | 7.45E-01 | 18245 |
| 2 | Precentral_R | -3.944 | -9.945 | 2.057 | 3.062 | -1.288 | -0.009 | 1.98E-01 | 18696 | 1.176 | -3.841 | 6.194 | 2.560 | 0.459 | 0.003 | 6.46E-01 | 18245 |
| 3 | Frontal_Sup_2_L | -6.332 | -13.247 | 0.582 | 3.528 | -1.795 | -0.013 | 7.27E-02 | 18696 | 3.333 | -2.454 | 9.120 | 2.952 | 1.129 | 0.008 | 2.59E-01 | 18245 |
| 4 | Frontal_Sup_2_R | -6.144 | -13.765 | 1.476 | 3.888 | -1.581 | -0.012 | 1.14E-01 | 18696 | 2.023 | -4.363 | 8.410 | 3.258 | 0.621 | 0.005 | 5.35E-01 | 18245 |
| 5 | Frontal_Mid_2_L | -4.111 | -11.372 | 3.149 | 3.704 | -1.110 | -0.008 | 2.67E-01 | 18696 | 3.539 | -2.548 | 9.627 | 3.106 | 1.140 | 0.008 | 2.54E-01 | 18245 |
| 6 | Frontal_Mid_2_R | -6.958 | -15.177 | 1.261 | 4.193 | -1.659 | -0.012 | 9.70E-02 | 18696 | -0.205 | -7.084 | 6.674 | 3.509 | -0.058 | 0.000 | 9.53E-01 | 18245 |
| 7 | Frontal_Inf_Oper_L | -2.156 | -4.300 | -0.011 | 1.094 | -1.970 | -0.014 | 4.88E-02 | 18696 | 1.590 | -0.207 | 3.386 | 0.917 | 1.734 | 0.013 | 8.29E-02 | 18245 |
| 8 | Frontal_Inf_Oper_R | -1.688 | -4.378 | 1.002 | 1.372 | -1.230 | -0.009 | 2.19E-01 | 18696 | 2.529 | 0.273 | 4.784 | 1.151 | 2.197 | 0.016 | 2.80E-02 | 18245 |
| 9 | Frontal_Inf_Tri_L | -3.606 | -8.161 | 0.950 | 2.324 | -1.551 | -0.011 | 1.21E-01 | 18696 | 5.234 | 1.417 | 9.052 | 1.948 | 2.687 | 0.020 | 7.21E-03 | 18245 |
| 10 | Frontal_Inf_Tri_R | -0.187 | -3.766 | 3.391 | 1.826 | -0.103 | -0.001 | 9.18E-01 | 18696 | 4.945 | 1.944 | 7.947 | 1.531 | 3.229 | 0.024 | 1.24E-03 | 18245 |
| 11 | Frontal_Inf_Orb_2_L | -1.284 | -2.646 | 0.078 | 0.695 | -1.848 | -0.014 | 6.46E-02 | 18696 | 1.872 | 0.729 | 3.015 | 0.583 | 3.211 | 0.024 | 1.33E-03 | 18245 |
| 12 | Frontal_Inf_Orb_2_R | -0.641 | -2.066 | 0.783 | 0.727 | -0.882 | -0.006 | 3.78E-01 | 18696 | 2.247 | 1.053 | 3.441 | 0.609 | 3.688 | 0.027 | 2.26E-04 | 18245 |
| 13 | Rolandic_Oper_L | -3.027 | -5.087 | -0.968 | 1.051 | -2.882 | -0.021 | 3.96E-03 | 18696 | 1.692 | -0.035 | 3.420 | 0.882 | 1.920 | 0.014 | 5.49E-02 | 18245 |
| 14 | Rolandic_Oper_R | -1.698 | -4.646 | 1.249 | 1.504 | -1.129 | -0.008 | 2.59E-01 | 18696 | 1.402 | -1.071 | 3.876 | 1.262 | 1.111 | 0.008 | 2.67E-01 | 18245 |
| 15 | Supp_Motor_Area_L | -0.938 | -5.835 | 3.959 | 2.499 | -0.375 | -0.003 | 7.07E-01 | 18696 | -1.282 | -5.381 | 2.818 | 2.091 | -0.613 | -0.005 | 5.40E-01 | 18245 |
| 16 | Supp_Motor_Area_R | 1.556 | -3.857 | 6.969 | 2.762 | 0.563 | 0.004 | 5.73E-01 | 18696 | -0.604 | -5.135 | 3.927 | 2.312 | -0.261 | -0.002 | 7.94E-01 | 18245 |
| 17 | Olfactory_L | -0.395 | -1.123 | 0.333 | 0.371 | -1.063 | -0.008 | 2.88E-01 | 18696 | 0.574 | -0.037 | 1.185 | 0.312 | 1.841 | 0.014 | 6.56E-02 | 18245 |
| 18 | Olfactory_R | -0.229 | -0.959 | 0.501 | 0.372 | -0.616 | -0.005 | 5.38E-01 | 18696 | 0.818 | 0.206 | 1.430 | 0.312 | 2.619 | 0.019 | 8.82E-03 | 18245 |
| 19 | Frontal_Sup_Medial_L | -1.417 | -6.009 | 3.174 | 2.342 | -0.605 | -0.004 | 5.45E-01 | 18696 | 3.438 | -0.409 | 7.286 | 1.963 | 1.752 | 0.013 | 7.99E-02 | 18245 |
| 20 | Frontal_Sup_Medial_R | -1.306 | -4.648 | 2.036 | 1.705 | -0.766 | -0.006 | 4.44E-01 | 18696 | 0.804 | -1.997 | 3.604 | 1.429 | 0.562 | 0.004 | 5.74E-01 | 18245 |
| 21 | Frontal_Med_Orb_L | -1.228 | -2.943 | 0.488 | 0.875 | -1.403 | -0.010 | 1.61E-01 | 18696 | 0.864 | -0.574 | 2.302 | 0.734 | 1.177 | 0.009 | 2.39E-01 | 18245 |
| 22 | Frontal_Med_Orb_R | -1.476 | -3.584 | 0.632 | 1.076 | -1.372 | -0.010 | 1.70E-01 | 18696 | 1.317 | -0.449 | 3.084 | 0.901 | 1.462 | 0.011 | 1.44E-01 | 18245 |
| 23 | Rectus_L | -2.165 | -3.939 | -0.390 | 0.906 | -2.390 | -0.017 | 1.68E-02 | 18696 | 1.469 | -0.020 | 2.957 | 0.759 | 1.934 | 0.014 | 5.31E-02 | 18245 |
| 24 | Rectus_R | -1.447 | -3.088 | 0.195 | 0.838 | -1.727 | -0.013 | 8.41E-02 | 18696 | 1.678 | 0.302 | 3.055 | 0.702 | 2.389 | 0.018 | 1.69E-02 | 18245 |
| 25 | OFCmed_L | -1.115 | -2.144 | -0.087 | 0.525 | -2.125 | -0.016 | 3.36E-02 | 18696 | 0.171 | -0.692 | 1.034 | 0.441 | 0.388 | 0.003 | 6.98E-01 | 18245 |
| 26 | OFCmed_R | -0.422 | -1.620 | 0.777 | 0.611 | -0.690 | -0.005 | 4.90E-01 | 18696 | 0.137 | -0.868 | 1.143 | 0.513 | 0.268 | 0.002 | 7.89E-01 | 18245 |
| 27 | OFCant_L | -0.992 | -2.182 | 0.197 | 0.607 | -1.635 | -0.012 | 1.02E-01 | 18696 | 0.164 | -0.834 | 1.162 | 0.509 | 0.322 | 0.002 | 7.47E-01 | 18245 |
| 28 | OFCant_R | -0.638 | -2.210 | 0.934 | 0.802 | -0.795 | -0.006 | 4.26E-01 | 18696 | 0.488 | -0.831 | 1.807 | 0.673 | 0.726 | 0.005 | 4.68E-01 | 18245 |
| 29 | OFCpost_L | -1.538 | -3.027 | -0.050 | 0.760 | -2.025 | -0.015 | 4.28E-02 | 18696 | 0.063 | -1.188 | 1.313 | 0.638 | 0.098 | 0.001 | 9.22E-01 | 18245 |
| 30 | OFCpost_R | -0.990 | -2.367 | 0.386 | 0.702 | -1.410 | -0.010 | 1.58E-01 | 18696 | 0.184 | -0.969 | 1.338 | 0.589 | 0.313 | 0.002 | 7.54E-01 | 18245 |
| 31 | OFClat_L | -0.153 | -0.656 | 0.349 | 0.256 | -0.599 | -0.004 | 5.49E-01 | 18696 | 0.817 | 0.395 | 1.238 | 0.215 | 3.798 | 0.028 | 1.46E-04 | 18245 |
| 32 | OFClat_R | -0.038 | -0.462 | 0.386 | 0.217 | -0.175 | -0.001 | 8.61E-01 | 18696 | 0.428 | 0.072 | 0.784 | 0.181 | 2.358 | 0.017 | 1.84E-02 | 18245 |
| 33 | Insula_L | -9.303 | -13.906 | -4.700 | 2.348 | -3.961 | -0.029 | 7.48E-05 | 18696 | 3.061 | -0.799 | 6.921 | 1.969 | 1.554 | 0.012 | 1.20E-01 | 18245 |
| 34 | Insula_R | -7.436 | -11.922 | -2.950 | 2.289 | -3.249 | -0.024 | 1.16E-03 | 18696 | 5.222 | 1.460 | 8.984 | 1.919 | 2.721 | 0.020 | 6.52E-03 | 18245 |
| 37 | Cingulate_Mid_L | -1.905 | -5.800 | 1.989 | 1.987 | -0.959 | -0.007 | 3.38E-01 | 18696 | -0.652 | -3.916 | 2.612 | 1.665 | -0.391 | -0.003 | 6.95E-01 | 18245 |
| 38 | Cingulate_Mid_R | -3.302 | -7.455 | 0.851 | 2.119 | -1.559 | -0.011 | 1.19E-01 | 18696 | -1.042 | -4.523 | 2.439 | 1.776 | -0.587 | -0.004 | 5.57E-01 | 18245 |
| 39 | Cingulate_Post_L | -0.815 | -2.005 | 0.374 | 0.607 | -1.343 | -0.010 | 1.79E-01 | 18696 | -0.194 | -1.189 | 0.802 | 0.508 | -0.381 | -0.003 | 7.03E-01 | 18245 |

| | | | | | | | | | | | | | | | | | |
|---|---|---|---|---|---|---|---|---|---|---|---|---|---|---|---|---|---|
| 40 | Cingulate_Post_R | -0.540 | -1.200 | 0.120 | 0.337 | -1.604 | -0.012 | 1.09E-01 | 18696 | -0.162 | -0.715 | 0.391 | 0.282 | -0.574 | -0.004 | 5.66E-01 | 18245 |
| 41 | Hippocampus_L | -0.754 | -2.678 | 1.169 | 0.981 | -0.769 | -0.006 | 4.42E-01 | 18696 | 1.196 | -0.419 | 2.811 | 0.824 | 1.452 | 0.011 | 1.47E-01 | 18245 |
| 42 | Hippocampus_R | 0.590 | -1.404 | 2.584 | 1.017 | 0.580 | 0.004 | 5.62E-01 | 18696 | 1.400 | -0.275 | 3.075 | 0.855 | 1.638 | 0.012 | 1.01E-01 | 18245 |
| 43 | ParaHippocampal_L | -0.588 | -2.297 | 1.121 | 0.872 | -0.674 | -0.005 | 5.00E-01 | 18696 | 0.857 | -0.576 | 2.291 | 0.732 | 1.172 | 0.009 | 2.41E-01 | 18245 |
| 44 | ParaHippocampal_R | -0.475 | -2.631 | 1.682 | 1.100 | -0.431 | -0.003 | 6.66E-01 | 18696 | 0.833 | -0.977 | 2.642 | 0.923 | 0.902 | 0.007 | 3.67E-01 | 18245 |
| 45 | Amygdala_L | -0.700 | -1.365 | -0.036 | 0.339 | -2.066 | -0.015 | 3.88E-02 | 18696 | 0.445 | -0.112 | 1.001 | 0.284 | 1.566 | 0.012 | 1.17E-01 | 18245 |
| 46 | Amygdala_R | -0.914 | -1.593 | -0.234 | 0.347 | -2.636 | -0.019 | 8.40E-03 | 18696 | 0.170 | -0.400 | 0.740 | 0.291 | 0.584 | 0.004 | 5.59E-01 | 18245 |
| 47 | Calcarine_L | -3.105 | -9.444 | 3.234 | 3.234 | -0.960 | -0.007 | 3.37E-01 | 18696 | -7.137 | -12.446 | -1.827 | 2.709 | -2.635 | -0.020 | 8.43E-03 | 18245 |
| 48 | Calcarine_R | -3.554 | -8.850 | 1.742 | 2.702 | -1.315 | -0.010 | 1.88E-01 | 18696 | -5.104 | -9.539 | -0.669 | 2.263 | -2.256 | -0.017 | 2.41E-02 | 18245 |
| 49 | Cuneus_L | -1.166 | -5.143 | 2.812 | 2.029 | -0.574 | -0.004 | 5.66E-01 | 18696 | -4.551 | -7.883 | -1.218 | 1.700 | -2.677 | -0.020 | 7.44E-03 | 18245 |
| 50 | Cuneus_R | -2.605 | -5.648 | 0.438 | 1.552 | -1.678 | -0.012 | 9.33E-02 | 18696 | -2.498 | -5.046 | 0.051 | 1.300 | -1.921 | -0.014 | 5.48E-02 | 18245 |
| 51 | Lingual_L | -2.372 | -6.182 | 1.439 | 1.944 | -1.220 | -0.009 | 2.23E-01 | 18696 | -0.641 | -3.839 | 2.557 | 1.632 | -0.393 | -0.003 | 6.94E-01 | 18245 |
| 52 | Lingual_R | -0.693 | -4.688 | 3.302 | 2.038 | -0.340 | -0.002 | 7.34E-01 | 18696 | -2.617 | -5.966 | 0.733 | 1.709 | -1.531 | -0.011 | 1.26E-01 | 18245 |
| 53 | Occipital_Sup_L | -1.244 | -3.866 | 1.377 | 1.337 | -0.931 | -0.007 | 3.52E-01 | 18696 | -1.150 | -3.348 | 1.048 | 1.121 | -1.026 | -0.008 | 3.05E-01 | 18245 |
| 54 | Occipital_Sup_R | -1.895 | -4.737 | 0.947 | 1.450 | -1.307 | -0.010 | 1.91E-01 | 18696 | 0.624 | -1.759 | 3.007 | 1.216 | 0.513 | 0.004 | 6.08E-01 | 18245 |
| 55 | Occipital_Mid_L | -0.327 | -6.472 | 5.818 | 3.135 | -0.104 | -0.001 | 9.17E-01 | 18696 | -0.401 | -5.545 | 4.742 | 2.624 | -0.153 | -0.001 | 8.78E-01 | 18245 |
| 56 | Occipital_Mid_R | -3.588 | -7.862 | 0.686 | 2.181 | -1.646 | -0.012 | 9.99E-02 | 18696 | 0.986 | -2.595 | 4.568 | 1.827 | 0.540 | 0.004 | 5.89E-01 | 18245 |
| 57 | Occipital_Inf_L | -1.909 | -3.948 | 0.130 | 1.040 | -1.835 | -0.013 | 6.65E-02 | 18696 | -1.298 | -3.008 | 0.411 | 0.872 | -1.489 | -0.011 | 1.37E-01 | 18245 |
| 58 | Occipital_Inf_R | -2.054 | -4.409 | 0.301 | 1.201 | -1.710 | -0.013 | 8.74E-02 | 18696 | -0.379 | -2.352 | 1.594 | 1.007 | -0.377 | -0.003 | 7.06E-01 | 18245 |
| 59 | Fusiform_L | -2.679 | -7.142 | 1.784 | 2.277 | -1.177 | -0.009 | 2.39E-01 | 18696 | -0.041 | -3.776 | 3.695 | 1.906 | -0.021 | 0.000 | 9.83E-01 | 18245 |
| 60 | Fusiform_R | -2.530 | -7.437 | 2.378 | 2.504 | -1.010 | -0.007 | 3.12E-01 | 18696 | 2.913 | -1.203 | 7.028 | 2.099 | 1.387 | 0.010 | 1.65E-01 | 18245 |
| 61 | Postcentral_L | 0.578 | -6.137 | 7.293 | 3.426 | 0.169 | 0.001 | 8.66E-01 | 18696 | 8.290 | 2.676 | 13.903 | 2.864 | 2.894 | 0.021 | 3.80E-03 | 18245 |
| 62 | Postcentral_R | -0.822 | -7.702 | 6.058 | 3.510 | -0.234 | -0.002 | 8.15E-01 | 18696 | 5.121 | -0.639 | 10.881 | 2.939 | 1.743 | 0.013 | 8.14E-02 | 18245 |
| 63 | Parietal_Sup_L | -0.406 | -4.275 | 3.463 | 1.974 | -0.205 | -0.002 | 8.37E-01 | 18696 | -1.196 | -4.439 | 2.048 | 1.655 | -0.723 | -0.005 | 4.70E-01 | 18245 |
| 64 | Parietal_Sup_R | 1.311 | -2.120 | 4.743 | 1.751 | 0.749 | 0.005 | 4.54E-01 | 18696 | 0.321 | -2.552 | 3.195 | 1.466 | 0.219 | 0.002 | 8.26E-01 | 18245 |
| 65 | Parietal_Inf_L | -4.278 | -9.342 | 0.787 | 2.584 | -1.656 | -0.012 | 9.78E-02 | 18696 | 0.656 | -3.584 | 4.896 | 2.163 | 0.303 | 0.002 | 7.62E-01 | 18245 |
| 66 | Parietal_Inf_R | -0.556 | -3.838 | 2.725 | 1.674 | -0.332 | -0.002 | 7.40E-01 | 18696 | 0.695 | -2.050 | 3.440 | 1.400 | 0.496 | 0.004 | 6.20E-01 | 18245 |
| 67 | SupraMarginal_L | -2.743 | -5.711 | 0.224 | 1.514 | -1.812 | -0.013 | 7.00E-02 | 18696 | 2.551 | 0.063 | 5.039 | 1.269 | 2.010 | 0.015 | 4.45E-02 | 18245 |
| 68 | SupraMarginal_R | -1.924 | -6.124 | 2.276 | 2.143 | -0.898 | -0.007 | 3.69E-01 | 18696 | 3.905 | 0.386 | 7.424 | 1.795 | 2.175 | 0.016 | 2.96E-02 | 18245 |
| 69 | Angular_L | -1.422 | -4.336 | 1.492 | 1.487 | -0.956 | -0.007 | 3.39E-01 | 18696 | 0.908 | -1.534 | 3.350 | 1.246 | 0.729 | 0.005 | 4.66E-01 | 18245 |
| 70 | Angular_R | -1.118 | -5.028 | 2.792 | 1.995 | -0.560 | -0.004 | 5.75E-01 | 18696 | 0.619 | -2.662 | 3.901 | 1.674 | 0.370 | 0.003 | 7.11E-01 | 18245 |
| 71 | Precuneus_L | -3.261 | -10.366 | 3.845 | 3.625 | -0.899 | -0.007 | 3.68E-01 | 18696 | -3.208 | -9.159 | 2.743 | 3.036 | -1.057 | -0.008 | 2.91E-01 | 18245 |
| 72 | Precuneus_R | -5.649 | -11.979 | 0.681 | 3.229 | -1.749 | -0.013 | 8.03E-02 | 18696 | -2.730 | -8.037 | 2.576 | 2.707 | -1.008 | -0.007 | 3.13E-01 | 18245 |
| 73 | Paracentral_Lobule_L | 0.284 | -2.970 | 3.537 | 1.660 | 0.171 | 0.001 | 8.64E-01 | 18696 | -0.974 | -3.697 | 1.748 | 1.389 | -0.701 | -0.005 | 4.83E-01 | 18245 |
| 74 | Paracentral_Lobule_R | 0.248 | -2.081 | 2.577 | 1.188 | 0.209 | 0.002 | 8.35E-01 | 18696 | -0.879 | -2.831 | 1.074 | 0.996 | -0.882 | -0.007 | 3.78E-01 | 18245 |
| 75 | Caudate_L | 0.867 | -1.729 | 3.463 | 1.324 | 0.654 | 0.005 | 5.13E-01 | 18696 | -0.091 | -2.264 | 2.082 | 1.109 | -0.082 | -0.001 | 9.35E-01 | 18245 |
| 76 | Caudate_R | 0.814 | -2.125 | 3.752 | 1.499 | 0.543 | 0.004 | 5.87E-01 | 18696 | 0.255 | -2.203 | 2.714 | 1.254 | 0.203 | 0.002 | 8.39E-01 | 18245 |
| 77 | Putamen_L | -4.906 | -8.851 | -0.960 | 2.013 | -2.437 | -0.018 | 1.48E-02 | 18696 | 0.119 | -3.185 | 3.424 | 1.686 | 0.071 | 0.001 | 9.44E-01 | 18245 |
| 78 | Putamen_R | -5.469 | -9.487 | -1.451 | 2.050 | -2.668 | -0.020 | 7.64E-03 | 18696 | 0.892 | -2.474 | 4.258 | 1.717 | 0.519 | 0.004 | 6.04E-01 | 18245 |
| 79 | Pallidum_L | -0.106 | -0.457 | 0.244 | 0.179 | -0.595 | -0.004 | 5.52E-01 | 18696 | 0.028 | -0.265 | 0.322 | 0.150 | 0.189 | 0.001 | 8.50E-01 | 18245 |
| 80 | Pallidum_R | -0.222 | -0.607 | 0.163 | 0.196 | -1.132 | -0.008 | 2.58E-01 | 18696 | 0.021 | -0.301 | 0.344 | 0.165 | 0.129 | 0.001 | 8.97E-01 | 18245 |
| 83 | Heschl_L | -0.913 | -1.624 | -0.201 | 0.363 | -2.515 | -0.018 | 1.19E-02 | 18696 | 0.258 | -0.338 | 0.854 | 0.304 | 0.847 | 0.006 | 3.97E-01 | 18245 |
| 84 | Heschl_R | -0.591 | -1.386 | 0.203 | 0.405 | -1.459 | -0.011 | 1.45E-01 | 18696 | 0.455 | -0.212 | 1.121 | 0.340 | 1.337 | 0.010 | 1.81E-01 | 18245 |
| 85 | Temporal_Sup_L | -8.637 | -13.772 | -3.502 | 2.620 | -3.297 | -0.024 | 9.80E-04 | 18696 | 4.188 | -0.118 | 8.495 | 2.197 | 1.906 | 0.014 | 5.66E-02 | 18245 |
| 86 | Temporal_Sup_R | -5.567 | -11.250 | 0.116 | 2.899 | -1.920 | -0.014 | 5.49E-02 | 18696 | 5.859 | 1.097 | 10.621 | 2.429 | 2.412 | 0.018 | 1.59E-02 | 18245 |
| 87 | Temporal_Pole_Sup_L | -2.372 | -4.805 | 0.061 | 1.241 | -1.911 | -0.014 | 5.60E-02 | 18696 | 1.108 | -0.931 | 3.147 | 1.040 | 1.065 | 0.008 | 2.87E-01 | 18245 |
| 88 | Temporal_Pole_Sup_R | -3.004 | -5.566 | -0.441 | 1.307 | -2.297 | -0.017 | 2.16E-02 | 18696 | 0.768 | -1.381 | 2.916 | 1.096 | 0.700 | 0.005 | 4.84E-01 | 18245 |
| 89 | Temporal_Mid_L | -7.999 | -16.561 | 0.563 | 4.368 | -1.831 | -0.013 | 6.71E-02 | 18696 | 2.490 | -4.681 | 9.660 | 3.658 | 0.681 | 0.005 | 4.96E-01 | 18245 |
| 90 | Temporal_Mid_R | -3.378 | -10.982 | 4.226 | 3.880 | -0.871 | -0.006 | 3.84E-01 | 18696 | 3.131 | -3.245 | 9.507 | 3.253 | 0.963 | 0.007 | 3.36E-01 | 18245 |

| | | | | | | | | | | | | | | | | | |
|---|---|---|---|---|---|---|---|---|---|---|---|---|---|---|---|---|---|
| 91 | Temporal_Pole_Mid_L | -1.090 | -3.135 | 0.954 | 1.043 | -1.045 | -0.008 | 2.96E-01 | 18696 | 0.134 | -1.581 | 1.848 | 0.875 | 0.153 | 0.001 | 8.78E-01 | 18245 |
| 92 | Temporal_Pole_Mid_R | -1.210 | -3.696 | 1.276 | 1.268 | -0.954 | -0.007 | 3.40E-01 | 18696 | 1.881 | -0.201 | 3.963 | 1.062 | 1.771 | 0.013 | 7.65E-02 | 18245 |
| 93 | Temporal_Inf_L | -3.366 | -9.251 | 2.518 | 3.002 | -1.121 | -0.008 | 2.62E-01 | 18696 | -1.244 | -6.175 | 3.687 | 2.516 | -0.494 | -0.004 | 6.21E-01 | 18245 |
| 94 | Temporal_Inf_R | -3.482 | -9.902 | 2.938 | 3.275 | -1.063 | -0.008 | 2.88E-01 | 18696 | 1.457 | -3.912 | 6.826 | 2.739 | 0.532 | 0.004 | 5.95E-01 | 18245 |
| 95 | Cerebellum_Crus1_L | -3.112 | -10.099 | 3.874 | 3.564 | -0.873 | -0.006 | 3.83E-01 | 18696 | -3.975 | -9.820 | 1.870 | 2.982 | -1.333 | -0.010 | 1.83E-01 | 18245 |
| 96 | Cerebellum_Crus1_R | 0.011 | -7.114 | 7.136 | 3.635 | 0.003 | 0.000 | 9.98E-01 | 18696 | 2.292 | -3.672 | 8.256 | 3.042 | 0.753 | 0.006 | 4.51E-01 | 18245 |
| 97 | Cerebellum_Crus2_L | -4.368 | -9.910 | 1.174 | 2.827 | -1.545 | -0.011 | 1.22E-01 | 18696 | -1.662 | -6.306 | 2.982 | 2.369 | -0.701 | -0.005 | 4.83E-01 | 18245 |
| 98 | Cerebellum_Crus2_R | -1.526 | -6.652 | 3.600 | 2.615 | -0.583 | -0.004 | 5.60E-01 | 18696 | 0.604 | -3.689 | 4.897 | 2.190 | 0.276 | 0.002 | 7.83E-01 | 18245 |
| 99 | Cerebellum_3_L | -0.450 | -0.822 | -0.078 | 0.190 | -2.371 | -0.017 | 1.78E-02 | 18696 | -0.336 | -0.647 | -0.024 | 0.159 | -2.114 | -0.016 | 3.45E-02 | 18245 |
| 100 | Cerebellum_3_R | -0.281 | -0.783 | 0.221 | 0.256 | -1.097 | -0.008 | 2.73E-01 | 18696 | -0.265 | -0.684 | 0.155 | 0.214 | -1.235 | -0.009 | 2.17E-01 | 18245 |
| 101 | Cerebellum_4_5_L | -1.860 | -4.868 | 1.149 | 1.535 | -1.212 | -0.009 | 2.26E-01 | 18696 | -0.434 | -2.952 | 2.085 | 1.285 | -0.338 | -0.002 | 7.36E-01 | 18245 |
| 102 | Cerebellum_4_5_R | -0.875 | -3.327 | 1.577 | 1.251 | -0.699 | -0.005 | 4.84E-01 | 18696 | -0.510 | -2.563 | 1.543 | 1.047 | -0.487 | -0.004 | 6.26E-01 | 18245 |
| 103 | Cerebellum_6_L | -2.795 | -7.995 | 2.406 | 2.653 | -1.053 | -0.008 | 2.92E-01 | 18696 | -1.937 | -6.290 | 2.416 | 2.221 | -0.872 | -0.006 | 3.83E-01 | 18245 |
| 104 | Cerebellum_6_R | -1.610 | -7.090 | 3.869 | 2.796 | -0.576 | -0.004 | 5.65E-01 | 18696 | -1.553 | -6.143 | 3.036 | 2.342 | -0.663 | -0.005 | 5.07E-01 | 18245 |
| 105 | Cerebellum_7b_L | -1.650 | -3.344 | 0.045 | 0.864 | -1.908 | -0.014 | 5.63E-02 | 18696 | -0.066 | -1.486 | 1.353 | 0.724 | -0.092 | -0.001 | 9.27E-01 | 18245 |
| 106 | Cerebellum_7b_R | -1.904 | -3.583 | -0.225 | 0.857 | -2.223 | -0.016 | 2.62E-02 | 18696 | 0.843 | -0.564 | 2.250 | 0.718 | 1.174 | 0.009 | 2.40E-01 | 18245 |
| 107 | Cerebellum_8_L | -4.540 | -9.918 | 0.839 | 2.744 | -1.654 | -0.012 | 9.81E-02 | 18696 | -1.960 | -6.465 | 2.545 | 2.298 | -0.853 | -0.006 | 3.94E-01 | 18245 |
| 108 | Cerebellum_8_R | -6.791 | -13.790 | 0.208 | 3.571 | -1.902 | -0.014 | 5.72E-02 | 18696 | 2.543 | -3.320 | 8.407 | 2.992 | 0.850 | 0.006 | 3.95E-01 | 18245 |
| 109 | Cerebellum_9_L | -3.331 | -6.372 | -0.289 | 1.552 | -2.146 | -0.016 | 3.19E-02 | 18696 | -1.163 | -3.712 | 1.385 | 1.300 | -0.895 | -0.007 | 3.71E-01 | 18245 |
| 110 | Cerebellum_9_R | -3.434 | -6.423 | -0.445 | 1.525 | -2.252 | -0.016 | 2.43E-02 | 18696 | -1.215 | -3.719 | 1.289 | 1.278 | -0.951 | -0.007 | 3.41E-01 | 18245 |
| 111 | Cerebellum_10_L | -0.074 | -0.290 | 0.142 | 0.110 | -0.671 | -0.005 | 5.02E-01 | 18696 | -0.115 | -0.296 | 0.066 | 0.092 | -1.242 | -0.009 | 2.14E-01 | 18245 |
| 112 | Cerebellum_10_R | -0.017 | -0.242 | 0.207 | 0.115 | -0.150 | -0.001 | 8.81E-01 | 18696 | 0.014 | -0.174 | 0.202 | 0.096 | 0.149 | 0.001 | 8.82E-01 | 18245 |
| 113 | Vermis_1_2 | -0.064 | -0.205 | 0.076 | 0.072 | -0.901 | -0.007 | 3.68E-01 | 18696 | -0.192 | -0.309 | -0.074 | 0.060 | -3.201 | -0.024 | 1.37E-03 | 18245 |
| 114 | Vermis_3 | -0.462 | -1.019 | 0.096 | 0.284 | -1.624 | -0.012 | 1.04E-01 | 18696 | -0.349 | -0.816 | 0.117 | 0.238 | -1.467 | -0.011 | 1.42E-01 | 18245 |
| 115 | Vermis_4_5 | -0.633 | -2.272 | 1.006 | 0.836 | -0.757 | -0.006 | 4.49E-01 | 18696 | -0.216 | -1.589 | 1.157 | 0.700 | -0.308 | -0.002 | 7.58E-01 | 18245 |
| 116 | Vermis_6 | 0.064 | -0.849 | 0.976 | 0.466 | 0.137 | 0.001 | 8.91E-01 | 18696 | -1.138 | -1.902 | -0.374 | 0.390 | -2.919 | -0.022 | 3.52E-03 | 18245 |
| 117 | Vermis_7 | -0.169 | -0.685 | 0.347 | 0.263 | -0.641 | -0.005 | 5.21E-01 | 18696 | -0.387 | -0.818 | 0.045 | 0.220 | -1.756 | -0.013 | 7.91E-02 | 18245 |
| 118 | Vermis_8 | -0.315 | -1.163 | 0.532 | 0.433 | -0.729 | -0.005 | 4.66E-01 | 18696 | -1.044 | -1.754 | -0.334 | 0.362 | -2.882 | -0.021 | 3.95E-03 | 18245 |
| 119 | Vermis_9 | -0.542 | -1.229 | 0.145 | 0.351 | -1.547 | -0.011 | 1.22E-01 | 18696 | -0.391 | -0.967 | 0.184 | 0.293 | -1.334 | -0.010 | 1.82E-01 | 18245 |
| 120 | Vermis_10 | -0.034 | -0.226 | 0.158 | 0.098 | -0.347 | -0.003 | 7.28E-01 | 18696 | -0.113 | -0.274 | 0.047 | 0.082 | -1.384 | -0.010 | 1.66E-01 | 18245 |
| 121 | Thal_AV_L | 0.038 | -0.039 | 0.114 | 0.039 | 0.960 | 0.007 | 3.37E-01 | 18696 | 0.082 | 0.018 | 0.146 | 0.033 | 2.498 | 0.018 | 1.25E-02 | 18245 |
| 122 | Thal_AV_R | 0.045 | -0.065 | 0.155 | 0.056 | 0.806 | 0.006 | 4.20E-01 | 18696 | 0.084 | -0.008 | 0.175 | 0.047 | 1.799 | 0.013 | 7.21E-02 | 18245 |
| 123 | Thal_LP_L | -0.009 | -0.094 | 0.076 | 0.043 | -0.201 | -0.001 | 8.41E-01 | 18696 | 0.078 | 0.007 | 0.149 | 0.036 | 2.142 | 0.016 | 3.22E-02 | 18245 |
| 124 | Thal_LP_R | -0.018 | -0.129 | 0.093 | 0.057 | -0.315 | -0.002 | 7.53E-01 | 18696 | 0.065 | -0.027 | 0.158 | 0.047 | 1.386 | 0.010 | 1.66E-01 | 18245 |
| 125 | Thal_VA_L | 0.106 | 0.015 | 0.197 | 0.046 | 2.290 | 0.017 | 2.21E-02 | 18696 | 0.070 | -0.006 | 0.146 | 0.039 | 1.808 | 0.013 | 7.06E-02 | 18245 |
| 126 | Thal_VA_R | 0.096 | -0.015 | 0.208 | 0.057 | 1.694 | 0.012 | 9.03E-02 | 18696 | 0.093 | 0.000 | 0.186 | 0.048 | 1.958 | 0.014 | 5.03E-02 | 18245 |
| 127 | Thal_VL_L | 0.086 | -0.172 | 0.345 | 0.132 | 0.656 | 0.005 | 5.12E-01 | 18696 | 0.285 | 0.070 | 0.501 | 0.110 | 2.594 | 0.019 | 9.50E-03 | 18245 |
| 128 | Thal_VL_R | 0.095 | -0.248 | 0.438 | 0.175 | 0.543 | 0.004 | 5.87E-01 | 18696 | 0.387 | 0.101 | 0.674 | 0.146 | 2.647 | 0.020 | 8.12E-03 | 18245 |
| 129 | Thal_VPL_L | -0.351 | -0.659 | -0.044 | 0.157 | -2.240 | -0.016 | 2.51E-02 | 18696 | 0.175 | -0.083 | 0.433 | 0.131 | 1.329 | 0.010 | 1.84E-01 | 18245 |
| 130 | Thal_VPL_R | -0.351 | -0.674 | -0.027 | 0.165 | -2.127 | -0.016 | 3.35E-02 | 18696 | 0.276 | 0.005 | 0.547 | 0.138 | 1.996 | 0.015 | 4.59E-02 | 18245 |
| 131 | Thal_IL_L | -0.149 | -0.282 | -0.016 | 0.068 | -2.194 | -0.016 | 2.82E-02 | 18696 | 0.060 | -0.051 | 0.172 | 0.057 | 1.060 | 0.008 | 2.89E-01 | 18245 |
| 132 | Thal_IL_R | -0.116 | -0.266 | 0.035 | 0.077 | -1.501 | -0.011 | 1.33E-01 | 18696 | 0.062 | -0.064 | 0.189 | 0.064 | 0.970 | 0.007 | 3.32E-01 | 18245 |
| 133 | Thal_Re_L | 0.000 | 0.000 | 0.001 | 0.000 | 1.603 | 0.012 | 1.09E-01 | 18696 | 0.001 | 0.000 | 0.001 | 0.000 | 2.133 | 0.016 | 3.29E-02 | 18245 |
| 134 | Thal_Re_R | 0.001 | -0.004 | 0.005 | 0.002 | 0.422 | 0.003 | 6.73E-01 | 18696 | 0.004 | 0.000 | 0.008 | 0.002 | 2.004 | 0.015 | 4.51E-02 | 18245 |
| 135 | Thal_MDm_L | -1.059 | -2.044 | -0.073 | 0.503 | -2.106 | -0.015 | 3.52E-02 | 18696 | 0.497 | -0.327 | 1.321 | 0.421 | 1.182 | 0.009 | 2.37E-01 | 18245 |
| 136 | Thal_MDm_R | -0.971 | -2.116 | 0.175 | 0.584 | -1.661 | -0.012 | 9.67E-02 | 18696 | 0.512 | -0.445 | 1.469 | 0.488 | 1.048 | 0.008 | 2.95E-01 | 18245 |
| 137 | Thal_MDl_L | -0.153 | -0.323 | 0.018 | 0.087 | -1.753 | -0.013 | 7.96E-02 | 18696 | 0.131 | -0.012 | 0.273 | 0.073 | 1.793 | 0.013 | 7.30E-02 | 18245 |
| 138 | Thal_MDl_R | -0.140 | -0.334 | 0.054 | 0.099 | -1.416 | -0.010 | 1.57E-01 | 18696 | 0.097 | -0.066 | 0.259 | 0.083 | 1.168 | 0.009 | 2.43E-01 | 18245 |
| 139 | Thal_LGN_L | -0.012 | -0.038 | 0.013 | 0.013 | -0.946 | -0.007 | 3.44E-01 | 18696 | 0.016 | -0.005 | 0.038 | 0.011 | 1.490 | 0.011 | 1.36E-01 | 18245 |

| | | | | | | | | | | | | | | | | | |
|---|---|---|---|---|---|---|---|---|---|---|---|---|---|---|---|---|---|
| 140 | Thal_LGN_R | -0.046 | -0.101 | 0.009 | 0.028 | -1.631 | -0.012 | 1.03E-01 | 18696 | 0.058 | 0.012 | 0.104 | 0.024 | 2.450 | 0.018 | 1.43E-02 | 18245 |
| 141 | Thal_MGN_L | -0.090 | -0.137 | -0.044 | 0.024 | -3.830 | -0.028 | 1.28E-04 | 18696 | 0.008 | -0.031 | 0.047 | 0.020 | 0.403 | 0.003 | 6.87E-01 | 18245 |
| 142 | Thal_MGN_R | -0.069 | -0.119 | -0.019 | 0.026 | -2.700 | -0.020 | 6.95E-03 | 18696 | 0.029 | -0.013 | 0.071 | 0.021 | 1.343 | 0.010 | 1.79E-01 | 18245 |
| 143 | Thal_PuA_L | -0.080 | -0.134 | -0.025 | 0.028 | -2.860 | -0.021 | 4.25E-03 | 18696 | 0.010 | -0.036 | 0.055 | 0.023 | 0.408 | 0.003 | 6.83E-01 | 18245 |
| 144 | Thal_PuA_R | -0.140 | -0.254 | -0.027 | 0.058 | -2.427 | -0.018 | 1.52E-02 | 18696 | 0.053 | -0.042 | 0.149 | 0.049 | 1.102 | 0.008 | 2.70E-01 | 18245 |
| 145 | Thal_PuM_L | -1.456 | -2.540 | -0.372 | 0.553 | -2.632 | -0.019 | 8.48E-03 | 18696 | 0.090 | -0.820 | 0.999 | 0.464 | 0.194 | 0.001 | 8.46E-01 | 18245 |
| 146 | Thal_PuM_R | -1.589 | -2.940 | -0.238 | 0.689 | -2.306 | -0.017 | 2.11E-02 | 18696 | 0.411 | -0.722 | 1.544 | 0.578 | 0.712 | 0.005 | 4.77E-01 | 18245 |
| 147 | Thal_PuL_L | -0.249 | -0.447 | -0.051 | 0.101 | -2.467 | -0.018 | 1.36E-02 | 18696 | 0.042 | -0.124 | 0.208 | 0.085 | 0.498 | 0.004 | 6.19E-01 | 18245 |
| 148 | Thal_PuL_R | -0.388 | -0.703 | -0.074 | 0.160 | -2.420 | -0.018 | 1.55E-02 | 18696 | 0.091 | -0.173 | 0.355 | 0.135 | 0.677 | 0.005 | 4.98E-01 | 18245 |
| 149 | Thal_PuI_L | -0.014 | -0.068 | 0.039 | 0.027 | -0.526 | -0.004 | 5.99E-01 | 18696 | 0.022 | -0.023 | 0.067 | 0.023 | 0.942 | 0.007 | 3.46E-01 | 18245 |
| 150 | Thal_PuI_R | -0.011 | -0.082 | 0.060 | 0.036 | -0.304 | -0.002 | 7.61E-01 | 18696 | 0.046 | -0.013 | 0.106 | 0.030 | 1.528 | 0.011 | 1.26E-01 | 18245 |
| 151 | ACC_sub_L | -0.105 | -0.725 | 0.516 | 0.317 | -0.331 | -0.002 | 7.41E-01 | 18696 | 0.195 | -0.326 | 0.715 | 0.266 | 0.733 | 0.005 | 4.64E-01 | 18245 |
| 152 | ACC_sub_R | 0.011 | -0.569 | 0.591 | 0.296 | 0.037 | 0.000 | 9.71E-01 | 18696 | 0.201 | -0.285 | 0.687 | 0.248 | 0.810 | 0.006 | 4.18E-01 | 18245 |
| 153 | ACC_pre_L | -0.578 | -2.170 | 1.014 | 0.812 | -0.712 | -0.005 | 4.77E-01 | 18696 | 0.876 | -0.459 | 2.211 | 0.681 | 1.286 | 0.010 | 1.98E-01 | 18245 |
| 154 | ACC_pre_R | -0.768 | -2.301 | 0.765 | 0.782 | -0.982 | -0.007 | 3.26E-01 | 18696 | 0.881 | -0.404 | 2.166 | 0.656 | 1.343 | 0.010 | 1.79E-01 | 18245 |
| 155 | ACC_sup_L | -0.613 | -2.342 | 1.116 | 0.882 | -0.695 | -0.005 | 4.87E-01 | 18696 | 0.072 | -1.380 | 1.523 | 0.740 | 0.097 | 0.001 | 9.23E-01 | 18245 |
| 156 | ACC_sup_R | -1.348 | -3.140 | 0.443 | 0.914 | -1.475 | -0.011 | 1.40E-01 | 18696 | 0.080 | -1.421 | 1.582 | 0.766 | 0.105 | 0.001 | 9.17E-01 | 18245 |
| 157 | N_Acc_L | 0.077 | -0.443 | 0.598 | 0.266 | 0.290 | 0.002 | 7.72E-01 | 18696 | 0.499 | 0.063 | 0.936 | 0.223 | 2.241 | 0.017 | 2.50E-02 | 18245 |
| 158 | N_Acc_R | -0.098 | -0.548 | 0.352 | 0.230 | -0.429 | -0.003 | 6.68E-01 | 18696 | 0.572 | 0.195 | 0.950 | 0.193 | 2.970 | 0.022 | 2.98E-03 | 18245 |
| 159 | VTA_L | -0.005 | -0.023 | 0.014 | 0.009 | -0.503 | -0.004 | 6.15E-01 | 18696 | 0.006 | -0.009 | 0.021 | 0.008 | 0.788 | 0.006 | 4.31E-01 | 18245 |
| 160 | VTA_R | -0.002 | -0.019 | 0.014 | 0.008 | -0.254 | -0.002 | 8.00E-01 | 18696 | 0.000 | -0.014 | 0.014 | 0.007 | 0.018 | 0.000 | 9.86E-01 | 18245 |
| 161 | SN_pc_L | -0.036 | -0.066 | -0.007 | 0.015 | -2.424 | -0.018 | 1.54E-02 | 18696 | -0.006 | -0.031 | 0.019 | 0.013 | -0.482 | -0.004 | 6.30E-01 | 18245 |
| 162 | SN_pc_R | -0.032 | -0.071 | 0.006 | 0.020 | -1.649 | -0.012 | 9.92E-02 | 18696 | -0.007 | -0.039 | 0.025 | 0.017 | -0.425 | -0.003 | 6.71E-01 | 18245 |
| 163 | SN_pr_L | -0.040 | -0.080 | 0.000 | 0.020 | -1.984 | -0.015 | 4.73E-02 | 18696 | 0.011 | -0.023 | 0.044 | 0.017 | 0.623 | 0.005 | 5.33E-01 | 18245 |
| 164 | SN_pr_R | -0.034 | -0.079 | 0.011 | 0.023 | -1.497 | -0.011 | 1.34E-01 | 18696 | 0.024 | -0.014 | 0.062 | 0.019 | 1.248 | 0.009 | 2.12E-01 | 18245 |
| 165 | Red_N_L | -0.042 | -0.091 | 0.008 | 0.025 | -1.659 | -0.012 | 9.70E-02 | 18696 | 0.021 | -0.021 | 0.062 | 0.021 | 0.986 | 0.007 | 3.24E-01 | 18245 |
| 166 | Red_N_R | -0.024 | -0.078 | 0.029 | 0.027 | -0.891 | -0.007 | 3.73E-01 | 18696 | 0.005 | -0.040 | 0.050 | 0.023 | 0.225 | 0.002 | 8.22E-01 | 18245 |
| 167 | LC_L | 0.003 | -0.015 | 0.021 | 0.009 | 0.321 | 0.002 | 7.49E-01 | 18696 | -0.015 | -0.030 | 0.001 | 0.008 | -1.860 | -0.014 | 6.29E-02 | 18245 |
| 168 | LC_R | 0.011 | -0.009 | 0.032 | 0.010 | 1.120 | 0.008 | 2.63E-01 | 18696 | -0.014 | -0.031 | 0.003 | 0.009 | -1.643 | -0.012 | 1.00E-01 | 18245 |
| 169 | Raphe_D | -0.056 | -0.127 | 0.015 | 0.036 | -1.535 | -0.011 | 1.25E-01 | 18696 | 0.021 | -0.039 | 0.080 | 0.031 | 0.678 | 0.005 | 4.98E-01 | 18245 |
| 170 | Raphe_M | 0.000 | -0.022 | 0.022 | 0.011 | -0.002 | 0.000 | 9.98E-01 | 18696 | -0.016 | -0.034 | 0.002 | 0.009 | -1.710 | -0.013 | 8.73E-02 | 18245 |

Non-oily fish intake

| Number | AAL 3 region | beta | 95%CI_l | 95%CI_h | se | t | r | p | df.e |
|---|---|---|---|---|---|---|---|---|---|
| 0 | Total brain | -159.545 | -356.492 | 37.402 | 100.478 | -1.588 | -0.012 | 1.12E-01 | 18667 |
| 1 | Precentral_L | -5.435 | -12.369 | 1.499 | 3.538 | -1.536 | -0.011 | 1.24E-01 | 18667 |
| 2 | Precentral_R | 1.551 | -4.803 | 7.906 | 3.242 | 0.478 | 0.004 | 6.32E-01 | 18667 |
| 3 | Frontal_Sup_2_L | -3.548 | -10.869 | 3.773 | 3.735 | -0.950 | -0.007 | 3.42E-01 | 18667 |
| 4 | Frontal_Sup_2_R | -9.608 | -17.675 | -1.540 | 4.116 | -2.334 | -0.017 | 1.96E-02 | 18667 |
| 5 | Frontal_Mid_2_L | -6.405 | -14.097 | 1.287 | 3.924 | -1.632 | -0.012 | 1.03E-01 | 18667 |
| 6 | Frontal_Mid_2_R | -10.148 | -18.849 | -1.447 | 4.439 | -2.286 | -0.017 | 2.23E-02 | 18667 |
| 7 | Frontal_Inf_Oper_L | -1.219 | -3.489 | 1.052 | 1.158 | -1.052 | -0.008 | 2.93E-01 | 18667 |
| 8 | Frontal_Inf_Oper_R | -1.622 | -4.469 | 1.226 | 1.453 | -1.116 | -0.008 | 2.64E-01 | 18667 |
| 9 | Frontal_Inf_Tri_L | -2.550 | -7.375 | 2.276 | 2.462 | -1.036 | -0.008 | 3.00E-01 | 18667 |
| 10 | Frontal_Inf_Tri_R | -3.531 | -7.320 | 0.257 | 1.933 | -1.827 | -0.013 | 6.77E-02 | 18667 |
| 11 | Frontal_Inf_Orb_2_L | -1.121 | -2.563 | 0.321 | 0.736 | -1.523 | -0.011 | 1.28E-01 | 18667 |
| 12 | Frontal_Inf_Orb_2_R | -1.208 | -2.716 | 0.301 | 0.770 | -1.569 | -0.011 | 1.17E-01 | 18667 |
| 13 | Rolandic_Oper_L | -0.218 | -2.400 | 1.963 | 1.113 | -0.196 | -0.001 | 8.44E-01 | 18667 |
| 14 | Rolandic_Oper_R | -0.897 | -4.019 | 2.224 | 1.592 | -0.564 | -0.004 | 5.73E-01 | 18667 |
| 15 | Supp_Motor_Area_L | -1.761 | -6.945 | 3.423 | 2.645 | -0.666 | -0.005 | 5.06E-01 | 18667 |

| | | | | | | | | | |
|---|---|---|---|---|---|---|---|---|---|
| 16 | Supp_Motor_Area_R | -1.986 | -7.717 | 3.745 | 2.924 | -0.679 | -0.005 | 4.97E-01 | 18667 |
| 17 | Olfactory_L | -0.289 | -1.060 | 0.482 | 0.393 | -0.734 | -0.005 | 4.63E-01 | 18667 |
| 18 | Olfactory_R | -0.009 | -0.782 | 0.764 | 0.394 | -0.023 | 0.000 | 9.82E-01 | 18667 |
| 19 | Frontal_Sup_Medial_L | -3.503 | -8.364 | 1.358 | 2.480 | -1.412 | -0.010 | 1.58E-01 | 18667 |
| 20 | Frontal_Sup_Medial_R | -3.454 | -6.992 | 0.084 | 1.805 | -1.913 | -0.014 | 5.57E-02 | 18667 |
| 21 | Frontal_Med_Orb_L | -0.530 | -2.346 | 1.285 | 0.926 | -0.572 | -0.004 | 5.67E-01 | 18667 |
| 22 | Frontal_Med_Orb_R | -0.832 | -3.063 | 1.399 | 1.138 | -0.731 | -0.005 | 4.65E-01 | 18667 |
| 23 | Rectus_L | -1.311 | -3.190 | 0.568 | 0.959 | -1.368 | -0.010 | 1.71E-01 | 18667 |
| 24 | Rectus_R | -0.802 | -2.540 | 0.936 | 0.887 | -0.904 | -0.007 | 3.66E-01 | 18667 |
| 25 | OFCmed_L | -1.438 | -2.527 | -0.348 | 0.556 | -2.587 | -0.019 | 9.69E-03 | 18667 |
| 26 | OFCmed_R | -0.562 | -1.831 | 0.708 | 0.648 | -0.867 | -0.006 | 3.86E-01 | 18667 |
| 27 | OFCant_L | -0.493 | -1.753 | 0.766 | 0.643 | -0.768 | -0.006 | 4.43E-01 | 18667 |
| 28 | OFCant_R | -0.678 | -2.342 | 0.986 | 0.849 | -0.799 | -0.006 | 4.25E-01 | 18667 |
| 29 | OFCpost_L | -0.587 | -2.165 | 0.990 | 0.805 | -0.730 | -0.005 | 4.66E-01 | 18667 |
| 30 | OFCpost_R | -0.264 | -1.721 | 1.194 | 0.744 | -0.354 | -0.003 | 7.23E-01 | 18667 |
| 31 | OFClat_L | -0.354 | -0.886 | 0.178 | 0.271 | -1.303 | -0.010 | 1.93E-01 | 18667 |
| 32 | OFClat_R | -0.536 | -0.985 | -0.086 | 0.229 | -2.336 | -0.017 | 1.95E-02 | 18667 |
| 33 | Insula_L | -2.595 | -7.473 | 2.284 | 2.489 | -1.042 | -0.008 | 2.97E-01 | 18667 |
| 34 | Insula_R | -3.375 | -8.127 | 1.377 | 2.424 | -1.392 | -0.010 | 1.64E-01 | 18667 |
| 37 | Cingulate_Mid_L | -3.748 | -7.870 | 0.375 | 2.103 | -1.782 | -0.013 | 7.48E-02 | 18667 |
| 38 | Cingulate_Mid_R | -1.774 | -6.170 | 2.623 | 2.243 | -0.791 | -0.006 | 4.29E-01 | 18667 |
| 39 | Cingulate_Post_L | 0.194 | -1.065 | 1.453 | 0.642 | 0.302 | 0.002 | 7.63E-01 | 18667 |
| 40 | Cingulate_Post_R | -0.007 | -0.705 | 0.692 | 0.357 | -0.019 | 0.000 | 9.85E-01 | 18667 |
| 41 | Hippocampus_L | 0.609 | -1.428 | 2.646 | 1.039 | 0.586 | 0.004 | 5.58E-01 | 18667 |
| 42 | Hippocampus_R | 1.316 | -0.796 | 3.428 | 1.078 | 1.222 | 0.009 | 2.22E-01 | 18667 |
| 43 | ParaHippocampal_L | 0.161 | -1.648 | 1.971 | 0.923 | 0.175 | 0.001 | 8.61E-01 | 18667 |
| 44 | ParaHippocampal_R | 0.013 | -2.269 | 2.295 | 1.164 | 0.011 | 0.000 | 9.91E-01 | 18667 |
| 45 | Amygdala_L | -0.096 | -0.800 | 0.608 | 0.359 | -0.267 | -0.002 | 7.90E-01 | 18667 |
| 46 | Amygdala_R | -0.233 | -0.952 | 0.487 | 0.367 | -0.634 | -0.005 | 5.26E-01 | 18667 |
| 47 | Calcarine_L | 0.554 | -6.160 | 7.268 | 3.425 | 0.162 | 0.001 | 8.71E-01 | 18667 |
| 48 | Calcarine_R | 0.792 | -4.817 | 6.402 | 2.862 | 0.277 | 0.002 | 7.82E-01 | 18667 |
| 49 | Cuneus_L | -1.820 | -6.030 | 2.389 | 2.148 | -0.848 | -0.006 | 3.97E-01 | 18667 |
| 50 | Cuneus_R | -1.319 | -4.540 | 1.901 | 1.643 | -0.803 | -0.006 | 4.22E-01 | 18667 |
| 51 | Lingual_L | -1.710 | -5.744 | 2.325 | 2.058 | -0.831 | -0.006 | 4.06E-01 | 18667 |
| 52 | Lingual_R | -1.288 | -5.516 | 2.941 | 2.157 | -0.597 | -0.004 | 5.51E-01 | 18667 |
| 53 | Occipital_Sup_L | -3.139 | -5.914 | -0.364 | 1.416 | -2.217 | -0.016 | 2.66E-02 | 18667 |
| 54 | Occipital_Sup_R | -0.195 | -3.204 | 2.814 | 1.535 | -0.127 | -0.001 | 8.99E-01 | 18667 |
| 55 | Occipital_Mid_L | 0.092 | -6.408 | 6.591 | 3.316 | 0.028 | 0.000 | 9.78E-01 | 18667 |
| 56 | Occipital_Mid_R | -2.396 | -6.923 | 2.132 | 2.310 | -1.037 | -0.008 | 3.00E-01 | 18667 |
| 57 | Occipital_Inf_L | 0.771 | -1.387 | 2.930 | 1.101 | 0.700 | 0.005 | 4.84E-01 | 18667 |
| 58 | Occipital_Inf_R | -1.983 | -4.476 | 0.510 | 1.272 | -1.559 | -0.011 | 1.19E-01 | 18667 |
| 59 | Fusiform_L | 0.572 | -4.154 | 5.297 | 2.411 | 0.237 | 0.002 | 8.13E-01 | 18667 |
| 60 | Fusiform_R | -1.912 | -7.103 | 3.279 | 2.648 | -0.722 | -0.005 | 4.70E-01 | 18667 |
| 61 | Postcentral_L | 3.686 | -3.421 | 10.794 | 3.626 | 1.017 | 0.007 | 3.09E-01 | 18667 |
| 62 | Postcentral_R | 5.411 | -1.873 | 12.695 | 3.716 | 1.456 | 0.011 | 1.45E-01 | 18667 |
| 63 | Parietal_Sup_L | -1.381 | -5.477 | 2.715 | 2.090 | -0.661 | -0.005 | 5.09E-01 | 18667 |
| 64 | Parietal_Sup_R | -1.626 | -5.261 | 2.008 | 1.854 | -0.877 | -0.006 | 3.80E-01 | 18667 |
| 65 | Parietal_Inf_L | 1.257 | -4.105 | 6.620 | 2.736 | 0.460 | 0.003 | 6.46E-01 | 18667 |
| 66 | Parietal_Inf_R | -0.384 | -3.856 | 3.088 | 1.771 | -0.217 | -0.002 | 8.28E-01 | 18667 |

| | | | | | | | | | |
|---|---|---|---|---|---|---|---|---|---|
| 67 | SupraMarginal_L | -0.474 | -3.617 | 2.670 | 1.604 | -0.295 | -0.002 | 7.68E-01 | 18667 |
| 68 | SupraMarginal_R | -2.144 | -6.590 | 2.302 | 2.268 | -0.945 | -0.007 | 3.45E-01 | 18667 |
| 69 | Angular_L | 0.990 | -2.096 | 4.076 | 1.575 | 0.629 | 0.005 | 5.30E-01 | 18667 |
| 70 | Angular_R | 0.838 | -3.299 | 4.976 | 2.111 | 0.397 | 0.003 | 6.91E-01 | 18667 |
| 71 | Precuneus_L | 2.319 | -5.201 | 9.839 | 3.837 | 0.604 | 0.004 | 5.46E-01 | 18667 |
| 72 | Precuneus_R | -0.450 | -7.149 | 6.249 | 3.418 | -0.132 | -0.001 | 8.95E-01 | 18667 |
| 73 | Paracentral_Lobule_L | -0.849 | -4.292 | 2.594 | 1.757 | -0.483 | -0.004 | 6.29E-01 | 18667 |
| 74 | Paracentral_Lobule_R | 1.678 | -0.789 | 4.144 | 1.258 | 1.333 | 0.010 | 1.83E-01 | 18667 |
| 75 | Caudate_L | -1.090 | -3.842 | 1.662 | 1.404 | -0.776 | -0.006 | 4.38E-01 | 18667 |
| 76 | Caudate_R | -2.415 | -5.531 | 0.701 | 1.590 | -1.519 | -0.011 | 1.29E-01 | 18667 |
| 77 | Putamen_L | -2.824 | -7.003 | 1.355 | 2.132 | -1.324 | -0.010 | 1.85E-01 | 18667 |
| 78 | Putamen_R | -3.469 | -7.725 | 0.787 | 2.171 | -1.598 | -0.012 | 1.10E-01 | 18667 |
| 79 | Pallidum_L | -0.041 | -0.412 | 0.331 | 0.189 | -0.214 | -0.002 | 8.30E-01 | 18667 |
| 80 | Pallidum_R | -0.045 | -0.453 | 0.363 | 0.208 | -0.216 | -0.002 | 8.29E-01 | 18667 |
| 83 | Heschl_L | -0.138 | -0.891 | 0.615 | 0.384 | -0.359 | -0.003 | 7.19E-01 | 18667 |
| 84 | Heschl_R | -0.486 | -1.327 | 0.355 | 0.429 | -1.132 | -0.008 | 2.58E-01 | 18667 |
| 85 | Temporal_Sup_L | -2.691 | -8.130 | 2.748 | 2.775 | -0.970 | -0.007 | 3.32E-01 | 18667 |
| 86 | Temporal_Sup_R | -2.232 | -8.249 | 3.785 | 3.070 | -0.727 | -0.005 | 4.67E-01 | 18667 |
| 87 | Temporal_Pole_Sup_L | -0.208 | -2.784 | 2.369 | 1.314 | -0.158 | -0.001 | 8.74E-01 | 18667 |
| 88 | Temporal_Pole_Sup_R | -0.974 | -3.686 | 1.738 | 1.384 | -0.704 | -0.005 | 4.81E-01 | 18667 |
| 89 | Temporal_Mid_L | -1.480 | -10.543 | 7.583 | 4.624 | -0.320 | -0.002 | 7.49E-01 | 18667 |
| 90 | Temporal_Mid_R | 5.093 | -2.948 | 13.135 | 4.103 | 1.241 | 0.009 | 2.14E-01 | 18667 |
| 91 | Temporal_Pole_Mid_L | 1.159 | -1.005 | 3.323 | 1.104 | 1.050 | 0.008 | 2.94E-01 | 18667 |
| 92 | Temporal_Pole_Mid_R | 0.730 | -1.900 | 3.361 | 1.342 | 0.544 | 0.004 | 5.86E-01 | 18667 |
| 93 | Temporal_Inf_L | -1.126 | -7.354 | 5.101 | 3.177 | -0.355 | -0.003 | 7.23E-01 | 18667 |
| 94 | Temporal_Inf_R | -0.672 | -7.461 | 6.117 | 3.463 | -0.194 | -0.001 | 8.46E-01 | 18667 |
| 95 | Cerebellum_Crus1_L | -4.147 | -11.538 | 3.243 | 3.771 | -1.100 | -0.008 | 2.71E-01 | 18667 |
| 96 | Cerebellum_Crus1_R | -5.836 | -13.373 | 1.701 | 3.845 | -1.518 | -0.011 | 1.29E-01 | 18667 |
| 97 | Cerebellum_Crus2_L | -5.062 | -10.927 | 0.802 | 2.992 | -1.692 | -0.012 | 9.07E-02 | 18667 |
| 98 | Cerebellum_Crus2_R | -4.654 | -10.078 | 0.770 | 2.767 | -1.682 | -0.012 | 9.26E-02 | 18667 |
| 99 | Cerebellum_3_L | -0.202 | -0.595 | 0.192 | 0.201 | -1.003 | -0.007 | 3.16E-01 | 18667 |
| 100 | Cerebellum_3_R | -0.341 | -0.872 | 0.190 | 0.271 | -1.258 | -0.009 | 2.08E-01 | 18667 |
| 101 | Cerebellum_4_5_L | -3.050 | -6.235 | 0.134 | 1.625 | -1.878 | -0.014 | 6.05E-02 | 18667 |
| 102 | Cerebellum_4_5_R | -2.441 | -5.036 | 0.154 | 1.324 | -1.844 | -0.013 | 6.52E-02 | 18667 |
| 103 | Cerebellum_6_L | -2.938 | -8.441 | 2.565 | 2.807 | -1.047 | -0.008 | 2.95E-01 | 18667 |
| 104 | Cerebellum_6_R | -3.061 | -8.858 | 2.737 | 2.958 | -1.035 | -0.008 | 3.01E-01 | 18667 |
| 105 | Cerebellum_7b_L | -1.197 | -2.990 | 0.595 | 0.915 | -1.309 | -0.010 | 1.91E-01 | 18667 |
| 106 | Cerebellum_7b_R | -2.150 | -3.927 | -0.373 | 0.907 | -2.372 | -0.017 | 1.77E-02 | 18667 |
| 107 | Cerebellum_8_L | -5.488 | -11.177 | 0.201 | 2.903 | -1.891 | -0.014 | 5.87E-02 | 18667 |
| 108 | Cerebellum_8_R | -9.115 | -16.519 | -1.712 | 3.777 | -2.413 | -0.018 | 1.58E-02 | 18667 |
| 109 | Cerebellum_9_L | -1.242 | -4.461 | 1.977 | 1.642 | -0.756 | -0.006 | 4.50E-01 | 18667 |
| 110 | Cerebellum_9_R | -0.898 | -4.063 | 2.266 | 1.614 | -0.556 | -0.004 | 5.78E-01 | 18667 |
| 111 | Cerebellum_10_L | -0.029 | -0.257 | 0.200 | 0.117 | -0.247 | -0.002 | 8.05E-01 | 18667 |
| 112 | Cerebellum_10_R | -0.133 | -0.371 | 0.104 | 0.121 | -1.099 | -0.008 | 2.72E-01 | 18667 |
| 113 | Vermis_1_2 | 0.016 | -0.133 | 0.164 | 0.076 | 0.205 | 0.002 | 8.37E-01 | 18667 |
| 114 | Vermis_3 | -0.155 | -0.746 | 0.435 | 0.301 | -0.516 | -0.004 | 6.06E-01 | 18667 |
| 115 | Vermis_4_5 | -0.543 | -2.278 | 1.192 | 0.885 | -0.613 | -0.004 | 5.40E-01 | 18667 |
| 116 | Vermis_6 | -0.021 | -0.988 | 0.945 | 0.493 | -0.043 | 0.000 | 9.65E-01 | 18667 |
| 117 | Vermis_7 | -0.115 | -0.661 | 0.431 | 0.279 | -0.414 | -0.003 | 6.79E-01 | 18667 |

| | | | | | | | | | |
|---|---|---|---|---|---|---|---|---|---|
| 118 | Vermis_8 | -0.149 | -1.045 | 0.748 | 0.457 | -0.325 | -0.002 | 7.45E-01 | 18667 |
| 119 | Vermis_9 | -0.320 | -1.047 | 0.407 | 0.371 | -0.863 | -0.006 | 3.88E-01 | 18667 |
| 120 | Vermis_10 | -0.017 | -0.220 | 0.186 | 0.103 | -0.166 | -0.001 | 8.68E-01 | 18667 |
| 121 | Thal_AV_L | 0.018 | -0.063 | 0.100 | 0.042 | 0.438 | 0.003 | 6.61E-01 | 18667 |
| 122 | Thal_AV_R | 0.072 | -0.044 | 0.188 | 0.059 | 1.217 | 0.009 | 2.24E-01 | 18667 |
| 123 | Thal_LP_L | -0.007 | -0.098 | 0.083 | 0.046 | -0.161 | -0.001 | 8.72E-01 | 18667 |
| 124 | Thal_LP_R | -0.016 | -0.133 | 0.101 | 0.060 | -0.267 | -0.002 | 7.90E-01 | 18667 |
| 125 | Thal_VA_L | 0.018 | -0.078 | 0.114 | 0.049 | 0.365 | 0.003 | 7.15E-01 | 18667 |
| 126 | Thal_VA_R | 0.057 | -0.061 | 0.176 | 0.060 | 0.953 | 0.007 | 3.40E-01 | 18667 |
| 127 | Thal_VL_L | -0.062 | -0.336 | 0.211 | 0.140 | -0.445 | -0.003 | 6.56E-01 | 18667 |
| 128 | Thal_VL_R | -0.204 | -0.567 | 0.160 | 0.185 | -1.099 | -0.008 | 2.72E-01 | 18667 |
| 129 | Thal_VPL_L | -0.051 | -0.376 | 0.275 | 0.166 | -0.306 | -0.002 | 7.60E-01 | 18667 |
| 130 | Thal_VPL_R | -0.174 | -0.516 | 0.169 | 0.175 | -0.994 | -0.007 | 3.20E-01 | 18667 |
| 131 | Thal_IL_L | -0.114 | -0.255 | 0.027 | 0.072 | -1.589 | -0.012 | 1.12E-01 | 18667 |
| 132 | Thal_IL_R | -0.056 | -0.216 | 0.104 | 0.082 | -0.688 | -0.005 | 4.91E-01 | 18667 |
| 133 | Thal_Re_L | -0.001 | -0.002 | 0.000 | 0.000 | -2.853 | -0.021 | 4.33E-03 | 18667 |
| 134 | Thal_Re_R | 0.000 | -0.004 | 0.005 | 0.002 | 0.140 | 0.001 | 8.89E-01 | 18667 |
| 135 | Thal_MDm_L | -0.453 | -1.497 | 0.591 | 0.533 | -0.851 | -0.006 | 3.95E-01 | 18667 |
| 136 | Thal_MDm_R | -0.014 | -1.228 | 1.199 | 0.619 | -0.023 | 0.000 | 9.81E-01 | 18667 |
| 137 | Thal_MDl_L | -0.157 | -0.338 | 0.024 | 0.092 | -1.702 | -0.012 | 8.88E-02 | 18667 |
| 138 | Thal_MDl_R | -0.039 | -0.245 | 0.166 | 0.105 | -0.377 | -0.003 | 7.06E-01 | 18667 |
| 139 | Thal_LGN_L | -0.001 | -0.028 | 0.027 | 0.014 | -0.046 | 0.000 | 9.63E-01 | 18667 |
| 140 | Thal_LGN_R | -0.036 | -0.095 | 0.022 | 0.030 | -1.217 | -0.009 | 2.24E-01 | 18667 |
| 141 | Thal_MGN_L | -0.054 | -0.102 | -0.005 | 0.025 | -2.146 | -0.016 | 3.19E-02 | 18667 |
| 142 | Thal_MGN_R | -0.064 | -0.118 | -0.011 | 0.027 | -2.376 | -0.017 | 1.75E-02 | 18667 |
| 143 | Thal_PuA_L | -0.067 | -0.125 | -0.009 | 0.029 | -2.268 | -0.017 | 2.34E-02 | 18667 |
| 144 | Thal_PuA_R | -0.127 | -0.247 | -0.006 | 0.061 | -2.065 | -0.015 | 3.89E-02 | 18667 |
| 145 | Thal_PuM_L | -1.033 | -2.181 | 0.115 | 0.586 | -1.763 | -0.013 | 7.79E-02 | 18667 |
| 146 | Thal_PuM_R | -0.757 | -2.188 | 0.674 | 0.730 | -1.037 | -0.008 | 3.00E-01 | 18667 |
| 147 | Thal_PuL_L | -0.161 | -0.370 | 0.049 | 0.107 | -1.502 | -0.011 | 1.33E-01 | 18667 |
| 148 | Thal_PuL_R | -0.168 | -0.502 | 0.165 | 0.170 | -0.990 | -0.007 | 3.22E-01 | 18667 |
| 149 | Thal_PuI_L | -0.010 | -0.067 | 0.047 | 0.029 | -0.359 | -0.003 | 7.20E-01 | 18667 |
| 150 | Thal_PuI_R | 0.000 | -0.076 | 0.075 | 0.038 | -0.008 | 0.000 | 9.94E-01 | 18667 |
| 151 | ACC_sub_L | -0.660 | -1.318 | -0.003 | 0.335 | -1.969 | -0.014 | 4.90E-02 | 18667 |
| 152 | ACC_sub_R | -0.474 | -1.088 | 0.141 | 0.313 | -1.511 | -0.011 | 1.31E-01 | 18667 |
| 153 | ACC_pre_L | -1.766 | -3.450 | -0.081 | 0.860 | -2.054 | -0.015 | 4.00E-02 | 18667 |
| 154 | ACC_pre_R | -1.559 | -3.182 | 0.064 | 0.828 | -1.882 | -0.014 | 5.98E-02 | 18667 |
| 155 | ACC_sup_L | -2.970 | -4.800 | -1.139 | 0.934 | -3.180 | -0.023 | 1.48E-03 | 18667 |
| 156 | ACC_sup_R | -3.108 | -5.004 | -1.211 | 0.967 | -3.212 | -0.024 | 1.32E-03 | 18667 |
| 157 | N_Acc_L | 0.006 | -0.545 | 0.557 | 0.281 | 0.021 | 0.000 | 9.83E-01 | 18667 |
| 158 | N_Acc_R | 0.017 | -0.460 | 0.493 | 0.243 | 0.069 | 0.001 | 9.45E-01 | 18667 |
| 159 | VTA_L | -0.006 | -0.025 | 0.013 | 0.010 | -0.604 | -0.004 | 5.46E-01 | 18667 |
| 160 | VTA_R | 0.000 | -0.017 | 0.018 | 0.009 | 0.048 | 0.000 | 9.62E-01 | 18667 |
| 161 | SN_pc_L | -0.029 | -0.060 | 0.002 | 0.016 | -1.856 | -0.014 | 6.35E-02 | 18667 |
| 162 | SN_pc_R | -0.022 | -0.063 | 0.019 | 0.021 | -1.060 | -0.008 | 2.89E-01 | 18667 |
| 163 | SN_pr_L | -0.024 | -0.066 | 0.018 | 0.022 | -1.104 | -0.008 | 2.69E-01 | 18667 |
| 164 | SN_pr_R | 0.001 | -0.046 | 0.049 | 0.024 | 0.054 | 0.000 | 9.57E-01 | 18667 |
| 165 | Red_N_L | -0.056 | -0.108 | -0.004 | 0.027 | -2.099 | -0.015 | 3.58E-02 | 18667 |
| 166 | Red_N_R | -0.006 | -0.062 | 0.051 | 0.029 | -0.191 | -0.001 | 8.48E-01 | 18667 |

| | | | | | | | | | |
|---|---|---|---|---|---|---|---|---|---|
| 167 | LC_L | -0.001 | -0.020 | 0.019 | 0.010 | -0.054 | 0.000 | 9.57E-01 | 18667 |
| 168 | LC_R | 0.001 | -0.020 | 0.022 | 0.011 | 0.101 | 0.001 | 9.19E-01 | 18667 |
| 169 | Raphe_D | -0.040 | -0.115 | 0.036 | 0.039 | -1.033 | -0.008 | 3.02E-01 | 18667 |
| 170 | Raphe_M | -0.003 | -0.026 | 0.020 | 0.012 | -0.259 | -0.002 | 7.96E-01 | 18667 |

Table S5: Associations between diet phenotypes (follow-up) and alternative brain structure measures.

| Field ID | Variable | beta | 95%CI_l | 95%CI_h | se | t | r | p | df.e | beta | 95%CI_l | 95%CI_h | se | t | r | p | df.e |
|---|---|---|---|---|---|---|---|---|---|---|---|---|---|---|---|---|---|
| | | Coffee intake | | | | | | | | Cereal intake | | | | | | | |
| 25005 | Volume of grey matter (normalised for head size) | -1424.739 | -1721.605 | -1127.872 | 151.455 | -9.407 | -0.069 | 5.68E-21 | 18652 | 760.627 | 572.932 | 948.321 | 95.758 | 7.943 | 0.058 | 2.08E-15 | 18722 |
| 25006 | Volume of grey matter | -771.503 | -1142.405 | -400.601 | 189.227 | -4.077 | -0.030 | 4.58E-05 | 18652 | 1143.483 | 909.710 | 1377.257 | 119.267 | 9.588 | 0.070 | 1.01E-21 | 18722 |
| 26518 | Volume of TotalGray (whole brain) | -371.902 | -611.399 | -132.404 | 122.187 | -3.044 | -0.022 | 2.34E-03 | 18411 | 641.751 | 490.591 | 792.912 | 77.119 | 8.322 | 0.061 | 9.28E-17 | 18481 |
| | | Tea intake | | | | | | | | Water intake | | | | | | | |
| 25005 | Volume of grey matter (normalised for head size) | 332.224 | 121.948 | 542.500 | 107.279 | 3.097 | 0.023 | 1.96E-03 | 18673 | -706.304 | -984.868 | -427.740 | 142.118 | -4.970 | -0.036 | 6.76E-07 | 18638 |
| 25006 | Volume of grey matter | 331.933 | 69.959 | 593.907 | 133.654 | 2.484 | 0.018 | 1.30E-02 | 18673 | -43.933 | -391.337 | 303.470 | 177.238 | -0.248 | -0.002 | 8.04E-01 | 18638 |
| 26518 | Volume of TotalGray (whole brain) | -163.522 | -332.764 | 5.721 | 86.344 | -1.894 | -0.014 | 5.83E-02 | 18432 | -101.702 | -326.176 | 122.773 | 114.522 | -0.888 | -0.007 | 3.75E-01 | 18398 |
| | | Cooked vegetable intake | | | | | | | | Processed meat intake | | | | | | | |
| 25005 | Volume of grey matter (normalised for head size) | -372.602 | -722.403 | -22.802 | 178.461 | -2.088 | -0.015 | 3.68E-02 | 18582 | -687.292 | -1203.363 | -171.221 | 263.289 | -2.610 | -0.019 | 9.05E-03 | 18734 |
| 25006 | Volume of grey matter | 232.576 | -203.868 | 669.020 | 222.665 | 1.045 | 0.008 | 2.96E-01 | 18582 | -1356.663 | -1999.748 | -713.578 | 328.089 | -4.135 | -0.030 | 3.56E-05 | 18734 |
| 26518 | Volume of TotalGray (whole brain) | -36.527 | -318.325 | 245.272 | 143.768 | -0.254 | -0.002 | 7.99E-01 | 18344 | -314.469 | -730.199 | 101.261 | 212.097 | -1.483 | -0.011 | 1.38E-01 | 18493 |
| | | Salad / raw vegetable intake | | | | | | | | Poultry intake | | | | | | | |
| 25005 | Volume of grey matter (normalised for head size) | -256.220 | -561.706 | 49.266 | 155.853 | -1.644 | -0.012 | 1.00E-01 | 18550 | 629.399 | 66.272 | 1192.526 | 287.296 | 2.191 | 0.016 | 2.85E-02 | 18739 |
| 25006 | Volume of grey matter | -20.031 | -400.838 | 360.776 | 194.280 | -0.103 | -0.001 | 9.18E-01 | 18550 | -895.312 | -1597.243 | -193.380 | 358.112 | -2.500 | -0.018 | 1.24E-02 | 18739 |
| 26518 | Volume of TotalGray (whole brain) | -76.955 | -323.503 | 169.594 | 125.784 | -0.612 | -0.005 | 5.41E-01 | 18312 | -236.534 | -690.103 | 217.035 | 231.402 | -1.022 | -0.008 | 3.07E-01 | 18498 |
| | | Fresh fruit intake | | | | | | | | Beef intake | | | | | | | |
| 25005 | Volume of grey matter (normalised for head size) | -103.837 | -479.250 | 271.575 | 191.528 | -0.542 | -0.004 | 5.88E-01 | 18642 | -388.421 | -1039.824 | 262.982 | 332.333 | -1.169 | -0.009 | 2.43E-01 | 18720 |
| 25006 | Volume of grey matter | 1642.923 | 1175.225 | 2110.622 | 238.610 | 6.885 | 0.050 | 5.95E-12 | 18642 | -1987.895 | -2798.565 | -1177.226 | 413.588 | -4.806 | -0.035 | 1.55E-06 | 18720 |
| 26518 | Volume of TotalGray (whole brain) | 499.420 | 197.309 | 801.531 | 154.131 | 3.240 | 0.024 | 1.20E-03 | 18402 | -25.217 | -550.301 | 499.867 | 267.887 | -0.094 | -0.001 | 9.25E-01 | 18480 |
| | | Dried fruit intake | | | | | | | | Lamb/mutton intake | | | | | | | |
| 25005 | Volume of grey matter (normalised for head size) | 470.420 | 31.853 | 908.987 | 223.748 | 2.102 | 0.015 | 3.55E-02 | 18519 | -1658.252 | -2486.879 | -829.626 | 422.749 | -3.923 | -0.029 | 8.79E-05 | 18710 |
| 25006 | Volume of grey matter | 1557.090 | 1010.311 | 2103.869 | 278.956 | 5.582 | 0.041 | 2.41E-08 | 18519 | -1028.257 | -2059.694 | 3.181 | 526.219 | -1.954 | -0.014 | 5.07E-02 | 18710 |
| 26518 | Volume of TotalGray (whole brain) | 694.765 | 341.810 | 1047.719 | 180.070 | 3.858 | 0.029 | 1.15E-04 | 18279 | -599.485 | -1266.132 | 67.161 | 340.110 | -1.763 | -0.013 | 7.80E-02 | 18472 |
| | | Bread intake | | | | | | | | Pork intake | | | | | | | |
| 25005 | Volume of grey matter (normalised for head size) | 41.042 | -35.596 | 117.680 | 39.099 | 1.050 | 0.008 | 2.94E-01 | 18608 | -846.020 | -1612.953 | -79.087 | 391.274 | -2.162 | -0.016 | 3.06E-02 | 18698 |
| 25006 | Volume of grey matter | 67.872 | -27.637 | 163.381 | 48.727 | 1.393 | 0.010 | 1.64E-01 | 18608 | -2315.315 | -3269.217 | -1361.413 | 486.662 | -4.758 | -0.035 | 1.97E-06 | 18698 |
| 26518 | Volume of TotalGray (whole brain) | 40.993 | -20.737 | 102.723 | 31.493 | 1.302 | 0.010 | 1.93E-01 | 18368 | -554.095 | -1171.163 | 62.974 | 314.816 | -1.760 | -0.013 | 7.84E-02 | 18459 |
| | | Oily fish intake | | | | | | | | Cheese intake | | | | | | | |
| 25005 | Volume of grey matter (normalised for head size) | -805.187 | -1383.092 | -227.282 | 294.836 | -2.731 | -0.020 | 6.32E-03 | 18726 | -1156.874 | -1646.154 | -667.594 | 249.621 | -4.635 | -0.034 | 3.60E-06 | 18411 |
| 25006 | Volume of grey matter | 1258.133 | 538.303 | 1977.964 | 367.244 | 3.426 | 0.025 | 6.14E-04 | 18726 | 1774.561 | 1164.066 | 2385.056 | 311.462 | 5.698 | 0.042 | 1.23E-08 | 18411 |
| 26518 | Volume of TotalGray (whole brain) | -481.721 | -947.312 | -16.130 | 237.535 | -2.028 | -0.015 | 4.26E-02 | 18485 | 316.898 | -78.120 | 711.915 | 201.530 | 1.572 | 0.012 | 1.16E-01 | 18174 |
| | | Non-oily fish intake | | | | | | | | | | | | | | | |
| 25005 | Volume of grey matter (normalised for head size) | -60.147 | -739.056 | 618.762 | 346.366 | -0.174 | -0.001 | 8.62E-01 | 18701 | | | | | | | | |
| 25006 | Volume of grey matter | 408.881 | -436.977 | 1254.738 | 431.540 | 0.947 | 0.007 | 3.43E-01 | 18701 | | | | | | | | |
| 26518 | Volume of TotalGray (whole brain) | -127.781 | -674.740 | 419.179 | 279.048 | -0.458 | -0.003 | 6.47E-01 | 18460 | | | | | | | | |

**Table S6: Associations between diet phenotypes (baseline) and alternative brain structure measures.**

| Field ID | Variable | beta | 95%CI_l | 95%CI_h | se | t | r | p | df.e |
|---|---|---|---|---|---|---|---|---|---|
| | | **Coffee intake** | | | | | | | |
| 25005 | Volume of grey matter (normalised for head size) | -1244.918 | -1514.548 | -975.288 | 137.560 | -9.050 | -0.066 | 1.57E-19 | 18835 |
| 25006 | Volume of grey matter | -787.633 | -1124.012 | -451.254 | 171.614 | -4.590 | -0.033 | 4.47E-06 | 18835 |
| 26518 | Volume of TotalGray (whole brain) | -326.831 | -544.356 | -109.306 | 110.977 | -2.945 | -0.022 | 3.23E-03 | 18593 |
| | | **Tea intake** | | | | | | | |
| 25005 | Volume of grey matter (normalised for head size) | 193.235 | -10.976 | 397.445 | 104.184 | 1.855 | 0.014 | 6.36E-02 | 18776 |
| 25006 | Volume of grey matter | 206.552 | -47.688 | 460.792 | 129.708 | 1.592 | 0.012 | 1.11E-01 | 18776 |
| 26518 | Volume of TotalGray (whole brain) | -255.293 | -419.458 | -91.127 | 83.754 | -3.048 | -0.022 | 2.31E-03 | 18536 |
| | | **Cooked vegetable intake** | | | | | | | |
| 25005 | Volume of grey matter (normalised for head size) | -608.938 | -1000.666 | -217.210 | 199.852 | -3.047 | -0.022 | 2.32E-03 | 18662 |
| 25006 | Volume of grey matter | 183.021 | -304.667 | 670.709 | 248.809 | 0.736 | 0.005 | 4.62E-01 | 18662 |
| 26518 | Volume of TotalGray (whole brain) | -298.414 | -613.893 | 17.065 | 160.951 | -1.854 | -0.014 | 6.37E-02 | 18424 |
| | | **Salad / raw vegetable intake** | | | | | | | |
| 25005 | Volume of grey matter (normalised for head size) | -294.460 | -604.037 | 15.117 | 157.940 | -1.864 | -0.014 | 6.23E-02 | 18702 |
| 25006 | Volume of grey matter | 45.336 | -339.850 | 430.521 | 196.514 | 0.231 | 0.002 | 8.18E-01 | 18702 |
| 26518 | Volume of TotalGray (whole brain) | -43.430 | -292.669 | 205.810 | 127.157 | -0.342 | -0.003 | 7.33E-01 | 18461 |
| | | **Fresh fruit intake** | | | | | | | |
| 25005 | Volume of grey matter (normalised for head size) | -445.746 | -835.692 | -55.800 | 198.943 | -2.241 | -0.016 | 2.51E-02 | 18762 |
| 25006 | Volume of grey matter | 1475.899 | 990.716 | 1961.082 | 247.531 | 5.962 | 0.043 | 2.53E-09 | 18762 |
| 26518 | Volume of TotalGray (whole brain) | 292.492 | -21.378 | 606.363 | 160.130 | 1.827 | 0.013 | 6.78E-02 | 18524 |
| | | **Dried fruit intake** | | | | | | | |
| 25005 | Volume of grey matter (normalised for head size) | 167.201 | -271.553 | 605.955 | 223.843 | 0.747 | 0.005 | 4.55E-01 | 18646 |
| 25006 | Volume of grey matter | 1724.994 | 1179.381 | 2270.607 | 278.361 | 6.197 | 0.045 | 5.88E-10 | 18646 |
| 26518 | Volume of TotalGray (whole brain) | 732.298 | 379.025 | 1085.572 | 180.233 | 4.063 | 0.030 | 4.86E-05 | 18407 |
| | | **Bread intake** | | | | | | | |
| 25005 | Volume of grey matter (normalised for head size) | 95.144 | 27.103 | 163.184 | 34.713 | 2.741 | 0.020 | 6.13E-03 | 18600 |
| 25006 | Volume of grey matter | 151.751 | 67.042 | 236.460 | 43.217 | 3.511 | 0.026 | 4.47E-04 | 18600 |
| 26518 | Volume of TotalGray (whole brain) | 56.657 | 1.953 | 111.361 | 27.909 | 2.030 | 0.015 | 4.24E-02 | 18363 |
| | | **Oily fish intake** | | | | | | | |
| 25005 | Volume of grey matter (normalised for head size) | -1229.155 | -1818.533 | -639.777 | 300.689 | -4.088 | -0.030 | 4.37E-05 | 18843 |
| 25006 | Volume of grey matter | 795.099 | 60.778 | 1529.419 | 374.636 | 2.122 | 0.015 | 3.38E-02 | 18843 |
| 26518 | Volume of TotalGray (whole brain) | -620.405 | -1094.933 | -145.876 | 242.095 | -2.563 | -0.019 | 1.04E-02 | 18601 |
| | | **Non-oily fish intake** | | | | | | | |
| 25005 | Volume of grey matter (normalised for head size) | -400.031 | -1074.846 | 274.785 | 344.278 | -1.162 | -0.008 | 2.45E-01 | 18838 |
| 25006 | Volume of grey matter | 671.114 | -168.946 | 1511.175 | 428.583 | 1.566 | 0.011 | 1.17E-01 | 18838 |
| 26518 | Volume of TotalGray (whole brain) | -539.435 | -1082.023 | 3.154 | 276.818 | -1.949 | -0.014 | 5.13E-02 | 18596 |

| Field ID | Variable | beta | 95%CI_l | 95%CI_h | se | t | r | p | df.e |
|---|---|---|---|---|---|---|---|---|---|
| | | **Cereal intake** | | | | | | | |
| 25005 | Volume of grey matter (normalised for head size) | 717.845 | 522.853 | 912.837 | 99.481 | 7.216 | 0.052 | 5.56E-13 | 18853 |
| 25006 | Volume of grey matter | 1230.264 | 987.825 | 1472.703 | 123.688 | 9.947 | 0.072 | 2.98E-23 | 18853 |
| 26518 | Volume of TotalGray (whole brain) | 492.145 | 335.206 | 649.083 | 80.067 | 6.147 | 0.045 | 8.07E-10 | 18611 |
| | | **Water intake** | | | | | | | |
| 25005 | Volume of grey matter (normalised for head size) | -664.794 | -930.073 | -399.515 | 135.340 | -4.912 | -0.036 | 9.09E-07 | 18751 |
| 25006 | Volume of grey matter | 299.377 | -30.967 | 629.721 | 168.535 | 1.776 | 0.013 | 7.57E-02 | 18751 |
| 26518 | Volume of TotalGray (whole brain) | 87.225 | -126.676 | 301.125 | 109.128 | 0.799 | 0.006 | 4.24E-01 | 18510 |
| | | **Processed meat intake** | | | | | | | |
| 25005 | Volume of grey matter (normalised for head size) | -91.974 | -607.675 | 423.727 | 263.101 | -0.350 | -0.003 | 7.27E-01 | 18866 |
| 25006 | Volume of grey matter | -977.930 | -1620.167 | -335.694 | 327.657 | -2.985 | -0.022 | 2.84E-03 | 18866 |
| 26518 | Volume of TotalGray (whole brain) | -284.719 | -699.352 | 129.914 | 211.538 | -1.346 | -0.010 | 1.78E-01 | 18624 |
| | | **Poultry intake** | | | | | | | |
| 25005 | Volume of grey matter (normalised for head size) | 424.216 | -154.184 | 1002.617 | 295.089 | 1.438 | 0.010 | 1.51E-01 | 18864 |
| 25006 | Volume of grey matter | -558.128 | -1278.317 | 162.062 | 367.427 | -1.519 | -0.011 | 1.29E-01 | 18864 |
| 26518 | Volume of TotalGray (whole brain) | -207.959 | -672.779 | 256.862 | 237.142 | -0.877 | -0.006 | 3.81E-01 | 18622 |
| | | **Beef intake** | | | | | | | |
| 25005 | Volume of grey matter (normalised for head size) | -431.630 | -1065.664 | 202.404 | 323.472 | -1.334 | -0.010 | 1.82E-01 | 18857 |
| 25006 | Volume of grey matter | -2020.125 | -2809.062 | -1231.189 | 402.500 | -5.019 | -0.037 | 5.24E-07 | 18857 |
| 26518 | Volume of TotalGray (whole brain) | -203.376 | -713.750 | 306.998 | 260.383 | -0.781 | -0.006 | 4.35E-01 | 18615 |
| | | **Lamb/mutton intake** | | | | | | | |
| 25005 | Volume of grey matter (normalised for head size) | -1239.419 | -2022.040 | -456.798 | 399.278 | -3.104 | -0.023 | 1.91E-03 | 18829 |
| 25006 | Volume of grey matter | -1028.445 | -2003.602 | -53.288 | 497.506 | -2.067 | -0.015 | 3.87E-02 | 18829 |
| 26518 | Volume of TotalGray (whole brain) | -462.097 | -1091.833 | 167.639 | 321.279 | -1.438 | -0.011 | 1.50E-01 | 18587 |
| | | **Pork intake** | | | | | | | |
| 25005 | Volume of grey matter (normalised for head size) | -592.477 | -1359.919 | 174.964 | 391.534 | -1.513 | -0.011 | 1.30E-01 | 18828 |
| 25006 | Volume of grey matter | -1924.296 | -2879.792 | -968.800 | 487.476 | -3.947 | -0.029 | 7.93E-05 | 18828 |
| 26518 | Volume of TotalGray (whole brain) | -674.555 | -1291.708 | -57.403 | 314.859 | -2.142 | -0.016 | 3.22E-02 | 18586 |
| | | **Cheese intake** | | | | | | | |
| 25005 | Volume of grey matter (normalised for head size) | -951.753 | -1450.426 | -453.080 | 254.413 | -3.741 | -0.027 | 1.84E-04 | 18591 |
| 25006 | Volume of grey matter | 1962.234 | 1341.028 | 2583.439 | 316.927 | 6.191 | 0.045 | 6.09E-10 | 18591 |
| 26518 | Volume of TotalGray (whole brain) | 283.424 | -119.300 | 686.147 | 205.461 | 1.379 | 0.010 | 1.68E-01 | 18351 |

**Table S7: Genome-wide independent significant variants of cereal intake.**

| CHR | SNP | BP | A1 | A2 | N | BETA | STAT | P | Is lead SNP |
|---|---|---|---|---|---|---|---|---|---|
| 1 | rs17030613 | 113190807 | C | A | 335090 | 0.04525 | 5.465 | 4.63E-08 | yes |
| 2 | rs10190898 | 60171757 | G | A | 334076 | 0.04518 | 5.758 | 8.54E-09 | yes |
| 3 | rs7618975 | 25086328 | C | A | 334112 | 0.03832 | 5.593 | 2.24E-08 | no |
| 3 | rs4858694 | 25097736 | A | G | 334974 | 0.04652 | 6.351 | 2.14E-10 | yes |
| 3 | rs7643390 | 25174059 | T | C | 334844 | 0.03853 | 5.757 | 8.56E-09 | no |
| 3 | rs720822 | 25232155 | A | G | 334631 | 0.03745 | 5.602 | 2.12E-08 | no |
| 3 | rs1483848 | 25316283 | A | G | 334342 | -0.03749 | -5.59 | 2.28E-08 | no |
| 3 | rs1562502 | 35735596 | T | C | 334961 | 0.0374 | 5.483 | 4.20E-08 | yes |
| 3 | rs2236950 | 50420554 | A | C | 335339 | 0.04909 | 5.607 | 2.06E-08 | yes |
| 5 | rs358288 | 87495504 | G | A | 335175 | -0.06738 | -6.725 | 1.76E-11 | yes |
| 5 | rs11748798 | 87650742 | T | G | 334620 | -0.04261 | -5.523 | 3.34E-08 | no |
| 6 | rs9381903 | 50773228 | A | G | 334734 | -0.03654 | -5.46 | 4.76E-08 | yes |
| 6 | rs13202780 | 51180392 | C | T | 334584 | -0.04685 | -6.363 | 1.99E-10 | yes |
| 6 | rs2504706 | 51258579 | C | T | 334485 | 0.05759 | 7.298 | 2.94E-13 | yes |
| 6 | rs1342613 | 51302471 | T | G | 334625 | -0.04519 | -5.622 | 1.89E-08 | no |
| 6 | rs9401196 | 98274551 | G | A | 333167 | -0.04266 | -6.312 | 2.76E-10 | no |
| 6 | rs7770226 | 98320402 | T | C | 334723 | -0.04334 | -6.362 | 1.99E-10 | no |
| 6 | rs9401305 | 98331937 | C | A | 334610 | 0.04835 | 7.204 | 5.87E-13 | yes |
| 6 | rs9482094 | 98364895 | A | G | 334516 | 0.03891 | 5.612 | 2.00E-08 | no |
| 6 | rs4524616 | 98445008 | C | A | 334599 | 0.04724 | 7 | 2.57E-12 | no |
| 6 | rs4557524 | 98466195 | T | G | 334289 | -0.04112 | -6.128 | 8.93E-10 | no |
| 6 | rs4601136 | 98488771 | G | A | 334855 | -0.04232 | -6.27 | 3.61E-10 | no |
| 6 | rs62422661 | 98545611 | T | G | 334865 | 0.04294 | 6.272 | 3.58E-10 | no |
| 7 | rs10278591 | 1921362 | T | C | 334728 | 0.05088 | 5.84 | 5.23E-09 | no |
| 7 | rs7791299 | 1935598 | C | T | 330905 | 0.04897 | 5.458 | 4.83E-08 | no |
| 7 | rs56259105 | 1978002 | C | T | 334205 | 0.0514 | 5.966 | 2.43E-09 | yes |
| 7 | rs34809719 | 2028968 | T | G | 334937 | 0.05096 | 5.896 | 3.73E-09 | no |
| 7 | rs56226325 | 2078981 | T | C | 335026 | 0.0526 | 5.668 | 1.44E-08 | no |
| 7 | rs4410790 | 17284577 | C | T | 334375 | -0.03806 | -5.474 | 4.40E-08 | yes |
| 8 | rs2670028 | 57423673 | G | A | 335051 | -0.04246 | -5.474 | 4.42E-08 | yes |
| 9 | rs7038814 | 86733355 | G | A | 335050 | 0.03847 | 5.567 | 2.60E-08 | no |
| 9 | rs2799849 | 86752641 | C | T | 334839 | 0.04467 | 6.245 | 4.24E-10 | yes |
| 9 | rs1549314 | 127910307 | G | T | 335235 | 0.03781 | 5.644 | 1.66E-08 | yes |

| | | | | | | | | | |
|---|---|---|---|---|---|---|---|---|---|
| 9 | rs501963 | 127962916 | A | G | 335170 | 0.03682 | 5.495 | 3.90E-08 | no |
| 11 | rs566534 | 28730053 | C | T | 334372 | 0.04044 | 5.601 | 2.14E-08 | yes |
| 15 | rs12440952 | 74615292 | G | A | 334998 | 0.04206 | 5.991 | 2.09E-09 | no |
| 15 | rs2959005 | 74618128 | C | T | 335121 | 0.04335 | 6.098 | 1.08E-09 | no |
| 15 | rs1564782 | 74622678 | G | A | 335313 | 0.04226 | 5.791 | 6.99E-09 | no |
| 15 | rs4077582 | 74665622 | C | T | 334477 | 0.04236 | 5.785 | 7.28E-09 | no |
| 15 | rs11856835 | 74716174 | G | A | 333176 | 0.04702 | 7.004 | 2.49E-12 | no |
| 15 | rs750607 | 74734819 | C | T | 331243 | 0.04175 | 6.141 | 8.19E-10 | no |
| 15 | rs2470893 | 75019449 | T | C | 333685 | -0.05338 | -7.474 | 7.78E-14 | no |
| 15 | rs2472297 | 75027880 | T | C | 334995 | -0.05918 | -7.844 | 4.38E-15 | yes |
| 18 | rs12709578 | 1834649 | T | G | 334811 | -0.04464 | -6.543 | 6.05E-11 | no |
| 18 | rs72867256 | 1849194 | A | G | 335530 | -0.08093 | -5.64 | 1.71E-08 | no |
| 18 | rs555692 | 1883692 | T | C | 334338 | -0.04216 | -6.274 | 3.53E-10 | no |
| 18 | rs2289322 | 1884329 | T | G | 334933 | -0.07967 | -8.439 | 3.20E-17 | yes |
| 18 | rs9951259 | 1898350 | A | G | 334936 | -0.04384 | -6.31 | 2.79E-10 | no |
| 19 | rs78816499 | 42596056 | C | T | 335429 | 0.07298 | 5.548 | 2.90E-08 | yes |
| 19 | rs769449 | 45410002 | A | G | 334498 | 0.05718 | 5.685 | 1.31E-08 | yes |
| 19 | rs2287921 | 49228272 | T | C | 335253 | -0.03858 | -5.751 | 8.89E-09 | no |
| 19 | rs2287922 | 49232226 | G | A | 334277 | -0.03893 | -5.791 | 7.00E-09 | yes |

**Table S8: Genome-wide independent significant variants of coffee intake.**

| CHR | SNP | BP | A1 | A2 | N | BETA | STAT | P | Is lead SNP |
|---|---|---|---|---|---|---|---|---|---|
| 2 | rs62106258 | 417167 | C | T | 334871 | -0.06039 | -5.48 | 4.25E-08 | yes |
| 2 | rs2947411 | 614168 | A | G | 334825 | -0.04882 | -7.831 | 4.86E-15 | no |
| 2 | rs2867125 | 622827 | T | C | 334788 | -0.04838 | -7.729 | 1.09E-14 | no |
| 2 | rs11127485 | 632028 | C | T | 333775 | -0.04937 | -7.87 | 3.57E-15 | no |
| 2 | rs6548238 | 634905 | T | C | 334844 | -0.04995 | -7.961 | 1.72E-15 | yes |
| 2 | rs13393304 | 637830 | A | G | 334396 | -0.0495 | -7.862 | 3.81E-15 | no |
| 2 | rs7561317 | 644953 | A | G | 334808 | -0.04936 | -7.884 | 3.17E-15 | no |
| 2 | rs1260326 | 27730940 | T | C | 333668 | -0.03923 | -8.083 | 6.34E-16 | yes |
| 2 | rs780094 | 27741237 | T | C | 334770 | -0.03783 | -7.765 | 8.18E-15 | no |
| 2 | rs780093 | 27742603 | T | C | 334480 | -0.03773 | -7.734 | 1.04E-14 | no |
| 2 | rs780092 | 27743154 | G | A | 334828 | 0.03828 | 5.935 | 2.94E-09 | no |
| 2 | rs1260333 | 27748624 | A | G | 334537 | -0.03502 | -7.337 | 2.19E-13 | no |
| 2 | rs1919127 | 27801493 | C | T | 334293 | -0.03374 | -6.195 | 5.82E-10 | no |
| 2 | rs1919128 | 27801759 | G | A | 334714 | -0.0336 | -6.178 | 6.49E-10 | no |
| 2 | rs13022873 | 27815510 | C | A | 333508 | -0.03247 | -5.933 | 2.97E-09 | no |
| 2 | rs12987055 | 27834152 | C | T | 334340 | -0.03425 | -6.279 | 3.40E-10 | no |
| 2 | rs3749147 | 27851918 | A | G | 334033 | -0.03355 | -6.116 | 9.60E-10 | no |
| 4 | rs58115360 | 2865107 | T | C | 334607 | -0.03048 | -5.832 | 5.47E-09 | yes |
| 4 | rs6824567 | 2883172 | T | C | 334500 | -0.02993 | -5.687 | 1.30E-08 | no |
| 4 | rs2231164 | 89015857 | C | T | 334759 | -0.03846 | -5.472 | 4.46E-08 | no |
| 4 | rs2231142 | 89052323 | T | G | 334794 | -0.05224 | -7.004 | 2.49E-12 | yes |
| 5 | rs12519880 | 7391434 | A | C | 334047 | -0.03241 | -6.158 | 7.39E-10 | yes |
| 5 | rs1422192 | 87959023 | A | G | 333513 | 0.03959 | 6.059 | 1.38E-09 | yes |
| 5 | rs618741 | 88064012 | C | T | 334555 | 0.02698 | 5.627 | 1.84E-08 | no |
| 5 | rs304132 | 88215594 | A | G | 334297 | 0.02629 | 5.487 | 4.09E-08 | yes |
| 6 | rs2517618 | 30158127 | A | G | 334574 | 0.03493 | 5.479 | 4.28E-08 | no |
| 6 | rs3132649 | 30321057 | A | G | 334850 | 0.03683 | 5.517 | 3.44E-08 | no |
| 6 | rs3094061 | 30321189 | C | A | 334728 | 0.03725 | 5.58 | 2.41E-08 | no |
| 6 | rs3130374 | 30321336 | T | C | 334492 | 0.03688 | 5.526 | 3.28E-08 | no |
| 6 | rs3130375 | 30321732 | A | C | 334557 | 0.03704 | 5.542 | 3.00E-08 | no |
| 6 | rs3131038 | 30734080 | A | G | 334432 | 0.03041 | 5.459 | 4.81E-08 | no |
| 6 | rs1264350 | 30796545 | C | T | 334508 | 0.03526 | 5.528 | 3.24E-08 | no |
| 6 | rs3131787 | 30899524 | C | T | 334619 | 0.03198 | 5.558 | 2.73E-08 | no |

| | | | | | | | | | |
|---|---|---|---|---|---|---|---|---|---|
| 6 | rs3095151 | 30900150 | T | G | 334436 | 0.03188 | 5.537 | 3.08E-08 | no |
| 6 | rs3130781 | 30914552 | C | T | 334723 | 0.03179 | 5.534 | 3.13E-08 | no |
| 6 | rs2247056 | 31265490 | T | C | 329376 | 0.02861 | 5.456 | 4.87E-08 | no |
| 6 | rs3094005 | 31465047 | T | G | 334717 | 0.03767 | 5.546 | 2.92E-08 | no |
| 6 | rs3132450 | 31596138 | G | A | 334394 | 0.03919 | 5.647 | 1.63E-08 | no |
| 6 | rs3117582 | 31620520 | G | T | 334928 | 0.03948 | 5.688 | 1.29E-08 | no |
| 6 | rs3132449 | 31626013 | T | C | 334913 | 0.03935 | 5.673 | 1.41E-08 | no |
| 6 | rs805293 | 31688518 | A | T | 334725 | -0.03005 | -6.317 | 2.66E-10 | no |
| 6 | rs3131383 | 31704294 | T | G | 334781 | 0.03933 | 5.666 | 1.46E-08 | no |
| 6 | rs3101018 | 31705864 | T | C | 334725 | 0.03863 | 5.56 | 2.71E-08 | no |
| 6 | rs3130484 | 31715882 | C | T | 334531 | 0.03932 | 5.659 | 1.52E-08 | no |
| 6 | rs3131379 | 31721033 | A | G | 334825 | 0.03925 | 5.654 | 1.57E-08 | no |
| 6 | rs3117574 | 31725230 | A | G | 334841 | 0.04029 | 5.801 | 6.59E-09 | no |
| 6 | rs3131378 | 31725285 | G | A | 334993 | 0.03927 | 5.659 | 1.52E-08 | no |
| 6 | rs3117575 | 31726253 | C | T | 334785 | 0.03926 | 5.659 | 1.52E-08 | no |
| 6 | rs3117577 | 31727474 | G | A | 334706 | 0.0389 | 5.604 | 2.09E-08 | no |
| 6 | rs3115671 | 31734345 | T | G | 334835 | 0.03918 | 5.641 | 1.70E-08 | no |
| 6 | rs3130490 | 31739120 | T | G | 334428 | 0.03985 | 5.714 | 1.10E-08 | no |
| 6 | rs3130679 | 31807540 | G | A | 334074 | 0.04115 | 5.919 | 3.25E-09 | no |
| 6 | rs521977 | 31836827 | T | G | 334634 | 0.03529 | 6.978 | 2.99E-12 | no |
| 6 | rs660594 | 31837250 | G | A | 334649 | 0.03064 | 6.462 | 1.03E-10 | no |
| 6 | rs660550 | 31837277 | C | A | 334773 | 0.03109 | 6.561 | 5.35E-11 | no |
| 6 | rs577272 | 31837963 | G | A | 334268 | 0.03092 | 6.524 | 6.86E-11 | no |
| 6 | rs644827 | 31838441 | T | C | 334745 | 0.03087 | 6.515 | 7.29E-11 | no |
| 6 | rs644774 | 31838490 | C | T | 334692 | 0.03087 | 6.515 | 7.29E-11 | no |
| 6 | rs2242665 | 31839309 | C | T | 334641 | 0.03036 | 6.404 | 1.51E-10 | no |
| 6 | rs3132442 | 31839494 | C | T | 334559 | 0.03035 | 6.409 | 1.47E-10 | no |
| 6 | rs3130482 | 31839782 | C | A | 334577 | 0.0301 | 6.358 | 2.05E-10 | no |
| 6 | rs605203 | 31847012 | C | A | 333769 | 0.03414 | 6.919 | 4.56E-12 | no |
| 6 | rs652888 | 31851234 | G | A | 334334 | 0.03107 | 5.556 | 2.76E-08 | no |
| 6 | rs486416 | 31856070 | G | A | 333954 | 0.03315 | 6.74 | 1.59E-11 | no |
| 6 | rs535586 | 31860337 | T | C | 334532 | 0.03377 | 6.872 | 6.34E-12 | no |
| 6 | rs659445 | 31864304 | G | A | 334444 | 0.03408 | 6.937 | 4.01E-12 | no |
| 6 | rs558702 | 31870326 | A | G | 334803 | 0.03924 | 5.633 | 1.78E-08 | no |
| 6 | rs497309 | 31892484 | C | A | 334859 | 0.03941 | 5.655 | 1.56E-08 | no |

| | | | | | | | | | |
|---|---|---|---|---|---|---|---|---|---|
| 6 | rs537160 | 31916400 | A | G | 333501 | 0.0305 | 6.165 | 7.04E-10 | no |
| 6 | rs1270942 | 31918860 | G | A | 334507 | 0.03839 | 5.512 | 3.55E-08 | no |
| 6 | rs630379 | 31922254 | A | C | 334128 | 0.03109 | 6.127 | 8.96E-10 | no |
| 6 | rs440454 | 31927342 | A | G | 333113 | 0.03034 | 5.991 | 2.09E-09 | no |
| 6 | rs419788 | 31928799 | T | C | 334363 | 0.03071 | 6.078 | 1.22E-09 | no |
| 6 | rs437179 | 31929014 | A | C | 333753 | 0.02981 | 5.892 | 3.81E-09 | no |
| 6 | rs410851 | 31936668 | T | C | 334732 | 0.03175 | 6.279 | 3.41E-10 | no |
| 6 | rs389884 | 31940897 | G | A | 333863 | 0.03956 | 5.665 | 1.48E-08 | no |
| 6 | rs389883 | 31947460 | G | T | 334545 | 0.03094 | 6.123 | 9.21E-10 | no |
| 6 | rs1150753 | 32059867 | G | A | 334784 | 0.0398 | 5.702 | 1.18E-08 | no |
| 6 | rs1150752 | 32064726 | C | T | 334792 | 0.03917 | 5.609 | 2.04E-08 | no |
| 6 | rs1269852 | 32080191 | C | G | 334624 | 0.04111 | 5.87 | 4.36E-09 | no |
| 6 | rs3130288 | 32096001 | A | C | 334496 | 0.04071 | 5.808 | 6.32E-09 | no |
| 6 | rs3130349 | 32147696 | A | G | 334807 | 0.03251 | 5.495 | 3.90E-08 | no |
| 6 | rs1062070 | 32148031 | G | A | 334144 | 0.03274 | 5.501 | 3.77E-08 | no |
| 6 | rs3134605 | 32159956 | C | T | 334787 | 0.03295 | 5.759 | 8.45E-09 | no |
| 6 | Affx-28468489 | 32165444 | G | A | 332971 | 0.03604 | 5.642 | 1.68E-08 | no |
| 6 | rs3132947 | 32176782 | T | G | 334211 | 0.03253 | 5.66 | 1.51E-08 | no |
| 6 | rs3096690 | 32194630 | C | G | 334354 | 0.03875 | 5.579 | 2.42E-08 | no |
| 6 | rs6936204 | 32217092 | T | C | 334472 | 0.02707 | 5.527 | 3.27E-08 | no |
| 6 | rs3130320 | 32223258 | T | C | 334395 | 0.0271 | 5.556 | 2.77E-08 | no |
| 6 | rs7756177 | 32289358 | A | G | 334437 | 0.03392 | 5.774 | 7.77E-09 | no |
| 6 | rs3129949 | 32298814 | A | C | 334232 | 0.03078 | 5.55 | 2.86E-08 | no |
| 6 | rs1003878 | 32299822 | A | G | 334584 | 0.03105 | 5.605 | 2.08E-08 | no |
| 6 | rs1265757 | 32302382 | T | C | 334838 | 0.03853 | 5.497 | 3.87E-08 | no |
| 6 | rs2395149 | 32325562 | A | G | 334798 | 0.03821 | 5.456 | 4.87E-08 | no |
| 6 | rs3129927 | 32333827 | C | A | 334725 | 0.03825 | 5.464 | 4.66E-08 | no |
| 6 | rs2858324 | 32660375 | A | G | 332865 | 0.02839 | 5.859 | 4.66E-09 | no |
| 6 | rs2647012 | 32664458 | T | C | 334466 | 0.02876 | 5.963 | 2.48E-09 | no |
| 6 | rs2647003 | 32664880 | T | G | 332652 | 0.02934 | 6.07 | 1.28E-09 | no |
| 6 | rs2647046 | 32668336 | A | C | 334679 | 0.02812 | 5.828 | 5.60E-09 | no |
| 6 | rs2856717 | 32670308 | A | G | 334223 | 0.02792 | 5.787 | 7.15E-09 | no |
| 6 | rs2858305 | 32670464 | G | T | 332198 | 0.02661 | 5.473 | 4.42E-08 | no |
| 6 | rs9275572 | 32678999 | A | G | 334631 | 0.02617 | 5.466 | 4.61E-08 | no |
| 6 | rs9275596 | 32681631 | C | T | 329449 | 0.02714 | 5.535 | 3.11E-08 | no |

| | | | | | | | | | |
|---|---|---|---|---|---|---|---|---|---|
| 6 | rs2504706 | 51258579 | C | T | 333860 | -0.03371 | -6.027 | 1.67E-09 | yes |
| 7 | rs7805128 | 17106017 | G | T | 333705 | -0.02893 | -6.062 | 1.35E-09 | yes |
| 7 | rs6973364 | 17160612 | T | C | 333541 | 0.02894 | 5.902 | 3.59E-09 | no |
| 7 | rs10950647 | 17196529 | G | A | 334015 | -0.04465 | -9.378 | 6.79E-21 | yes |
| 7 | rs112476491 | 17204040 | A | G | 334464 | -0.07753 | -5.812 | 6.17E-09 | no |
| 7 | rs4721583 | 17215812 | G | A | 334196 | 0.0335 | 5.934 | 2.95E-09 | no |
| 7 | rs13328338 | 17221091 | G | A | 333272 | 0.02655 | 5.455 | 4.90E-08 | no |
| 7 | rs4476901 | 17243954 | G | A | 334362 | 0.05193 | 7.54 | 4.70E-14 | yes |
| 7 | rs74919529 | 17269707 | T | C | 327881 | -0.07873 | -6.436 | 1.23E-10 | yes |
| 7 | Affx-30120430 | 17273374 | CT | C | 334700 | -0.05107 | -9.319 | 1.18E-20 | no |
| 7 | rs12539940 | 17275597 | C | T | 334196 | 0.0532 | 5.903 | 3.56E-09 | no |
| 7 | rs78707595 | 17279393 | T | C | 334876 | 0.111 | 7.263 | 3.80E-13 | no |
| 7 | rs4410790 | 17284577 | C | T | 333749 | 0.12 | 24.37 | 4.71E-131 | yes |
| 7 | rs1117203 | 17303549 | A | G | 334210 | -0.05868 | -11.03 | 2.74E-28 | no |
| 7 | rs12670403 | 17309279 | C | A | 334606 | -0.09552 | -20.18 | 1.72E-90 | no |
| 7 | rs111577430 | 17319257 | G | A | 332704 | -0.1144 | -6.143 | 8.12E-10 | no |
| 7 | rs10272066 | 17374902 | C | T | 333280 | -0.07016 | -12.48 | 1.01E-35 | yes |
| 7 | rs2066853 | 17379110 | A | G | 334344 | -0.08551 | -11.1 | 1.23E-28 | no |
| 7 | rs113070557 | 17382217 | G | T | 334679 | -0.1239 | -7.813 | 5.60E-15 | yes |
| 7 | rs62446370 | 17404160 | A | G | 334181 | 0.06413 | 5.972 | 2.35E-09 | yes |
| 7 | rs2198497 | 17414133 | G | A | 325214 | -0.05438 | -8.801 | 1.36E-18 | no |
| 7 | rs11764350 | 17422113 | T | A | 331955 | -0.03137 | -5.977 | 2.28E-09 | no |
| 7 | rs80265229 | 17465828 | T | C | 334506 | 0.05936 | 7.855 | 4.02E-15 | yes |
| 7 | rs2034684 | 17475607 | C | T | 333131 | 0.04152 | 7.626 | 2.44E-14 | no |
| 7 | rs74789644 | 17488791 | G | C | 333474 | -0.07512 | -7.709 | 1.27E-14 | yes |
| 7 | rs606056 | 17501082 | C | A | 333974 | -0.03453 | -7.242 | 4.44E-13 | no |
| 7 | rs4721617 | 17507948 | G | A | 334420 | 0.04506 | 8.25 | 1.58E-16 | no |
| 7 | rs6963451 | 17517644 | G | T | 334350 | -0.06862 | -9.524 | 1.68E-21 | no |
| 7 | rs11761325 | 17552884 | A | G | 332012 | -0.03797 | -6.615 | 3.71E-11 | yes |
| 7 | rs4717763 | 72825125 | A | G | 334128 | 0.02666 | 5.535 | 3.11E-08 | no |
| 7 | rs71556711 | 72854549 | T | C | 334600 | 0.04771 | 5.788 | 7.13E-09 | no |
| 7 | rs2240466 | 72856269 | A | G | 334858 | 0.05365 | 7.475 | 7.75E-14 | no |
| 7 | rs1178979 | 72856430 | C | T | 334695 | 0.04142 | 6.97 | 3.18E-12 | no |
| 7 | rs714052 | 72864869 | G | A | 333967 | 0.05312 | 7.411 | 1.26E-13 | no |
| 7 | rs12056034 | 72878645 | G | A | 332370 | 0.05353 | 7.467 | 8.25E-14 | no |

| | | | | | | | | | |
|---|---|---|---|---|---|---|---|---|---|
| 7 | rs17145713 | 72904810 | T | C | 334453 | 0.04116 | 6.922 | 4.47E-12 | no |
| 7 | rs13233571 | 72971231 | T | C | 334401 | 0.05561 | 7.744 | 9.63E-15 | no |
| 7 | rs17145738 | 72982874 | T | C | 334610 | 0.05516 | 7.665 | 1.79E-14 | no |
| 7 | rs2286276 | 72987354 | T | C | 334711 | 0.03813 | 7.281 | 3.32E-13 | no |
| 7 | rs1051921 | 73007943 | A | G | 334795 | 0.04321 | 7.29 | 3.12E-13 | no |
| 7 | rs35332062 | 73012042 | A | G | 334765 | 0.05382 | 7.609 | 2.77E-14 | no |
| 7 | rs35368205 | 73017657 | T | C | 334026 | 0.04288 | 7.25 | 4.17E-13 | no |
| 7 | rs3812316 | 73020337 | G | C | 334937 | 0.05593 | 7.924 | 2.31E-15 | yes |
| 7 | rs17145750 | 73026378 | T | C | 334165 | 0.04697 | 7.305 | 2.78E-13 | no |
| 7 | rs34958196 | 73043101 | A | G | 334255 | 0.05462 | 7.751 | 9.13E-15 | no |
| 7 | rs41299460 | 75600951 | A | G | 333784 | 0.03619 | 7.414 | 1.23E-13 | no |
| 7 | rs1057868 | 75615006 | T | C | 334836 | 0.06098 | 11.65 | 2.36E-31 | yes |
| 7 | rs17685 | 75616105 | A | G | 334742 | 0.0589 | 11.16 | 6.37E-29 | no |
| 7 | rs8565 | 75630274 | T | C | 334610 | 0.05392 | 10.08 | 7.05E-24 | no |
| 7 | Affx-30886472 | 75659815 | T | C | 334235 | 0.04213 | 8.163 | 3.29E-16 | no |
| 7 | rs61303167 | 75671175 | T | G | 334555 | 0.04507 | 7.09 | 1.35E-12 | no |
| 7 | rs79859948 | 75679099 | G | A | 334402 | 0.04716 | 5.991 | 2.08E-09 | no |
| 7 | rs74999892 | 75756464 | C | T | 333917 | 0.04668 | 5.929 | 3.06E-09 | no |
| 7 | rs56343450 | 75796235 | T | C | 334424 | 0.04232 | 8.209 | 2.24E-16 | no |
| 7 | rs78663390 | 75845837 | A | C | 333340 | 0.04387 | 6.671 | 2.55E-11 | no |
| 12 | rs2599415 | 11217237 | G | A | 331414 | 0.03613 | 5.635 | 1.75E-08 | no |
| 12 | rs61928609 | 11316437 | A | C | 334259 | 0.03771 | 5.893 | 3.81E-09 | yes |
| 12 | rs1669413 | 11338781 | C | A | 334631 | 0.03721 | 5.832 | 5.48E-09 | no |
| 12 | rs1669424 | 11346562 | T | C | 334480 | 0.03636 | 5.696 | 1.23E-08 | no |
| 15 | rs12914489 | 74187937 | A | G | 334708 | 0.06171 | 8.106 | 5.25E-16 | yes |
| 15 | rs5742914 | 74286929 | T | C | 334713 | 0.06184 | 8.463 | 2.61E-17 | no |
| 15 | Affx-11962945 | 74467796 | T | C | 334262 | 0.05894 | 10.3 | 6.99E-25 | no |
| 15 | rs1052622 | 74467856 | G | A | 334191 | 0.04713 | 9.292 | 1.52E-20 | yes |
| 15 | rs34868798 | 74469716 | C | T | 334744 | 0.05902 | 10.34 | 4.93E-25 | no |
| 15 | rs351224 | 74487036 | A | T | 333823 | -0.03089 | -6.476 | 9.42E-11 | no |
| 15 | rs11857410 | 74488424 | A | G | 334671 | 0.06163 | 10.76 | 5.37E-27 | no |
| 15 | rs74922218 | 74516615 | A | C | 331201 | 0.09201 | 5.543 | 2.98E-08 | no |
| 15 | rs11635191 | 74550050 | A | C | 334055 | -0.03848 | -6.468 | 9.95E-11 | no |
| 15 | rs12916871 | 74552742 | C | A | 334352 | 0.04682 | 9.796 | 1.18E-22 | no |
| 15 | rs28395230 | 74557835 | C | T | 327973 | -0.04784 | -8.068 | 7.17E-16 | yes |

| | | | | | | | | | |
|---|---|---|---|---|---|---|---|---|---|
| 15 | rs1038436 | 74560566 | C | T | 333734 | -0.03851 | -7.445 | 9.69E-14 | no |
| 15 | rs16967313 | 74561301 | A | G | 333238 | -0.04426 | -6.706 | 2.00E-11 | no |
| 15 | rs901130 | 74573907 | G | C | 334384 | -0.0339 | -6.429 | 1.28E-10 | no |
| 15 | rs4887131 | 74598757 | A | C | 334065 | -0.04562 | -9.224 | 2.88E-20 | no |
| 15 | rs1009176 | 74603484 | T | C | 334865 | -0.05065 | -7.546 | 4.50E-14 | no |
| 15 | rs12440952 | 74615292 | G | A | 334370 | -0.05075 | -10.2 | 1.99E-24 | no |
| 15 | rs2959005 | 74618128 | C | T | 334493 | -0.05076 | -10.07 | 7.26E-24 | no |
| 15 | rs1564782 | 74622678 | G | A | 334687 | -0.04932 | -9.534 | 1.52E-21 | no |
| 15 | rs4077582 | 74665622 | C | T | 333853 | -0.04874 | -9.388 | 6.12E-21 | no |
| 15 | rs28362906 | 74711790 | A | G | 334407 | 0.1034 | 10.49 | 1.02E-25 | no |
| 15 | rs11856835 | 74716174 | G | A | 332558 | -0.07035 | -14.79 | 1.85E-49 | no |
| 15 | rs8036030 | 74716609 | A | G | 334554 | -0.06242 | -12.78 | 2.11E-37 | no |
| 15 | rs750607 | 74734819 | C | T | 330624 | -0.05822 | -12.08 | 1.32E-33 | no |
| 15 | rs35032603 | 74753425 | C | T | 334391 | 0.1195 | 9.549 | 1.32E-21 | yes |
| 15 | rs80025017 | 74845566 | A | C | 333788 | -0.05471 | -6.15 | 7.77E-10 | no |
| 15 | rs62005806 | 74883323 | A | G | 334239 | 0.1111 | 12.28 | 1.25E-34 | no |
| 15 | rs4128436 | 74935894 | T | C | 334374 | -0.05794 | -6.667 | 2.62E-11 | yes |
| 15 | rs145923939 | 74961238 | C | T | 334541 | 0.1073 | 5.874 | 4.27E-09 | yes |
| 15 | rs2606345 | 75017176 | C | A | 333843 | -0.05203 | -10.28 | 8.67E-25 | no |
| 15 | rs2470893 | 75019449 | T | C | 333057 | 0.1174 | 23.19 | 7.37E-119 | no |
| 15 | rs2472297 | 75027880 | T | C | 334365 | 0.1419 | 26.55 | 4.20E-155 | yes |
| 15 | rs2472299 | 75033400 | A | G | 334699 | -0.06438 | -12.1 | 1.13E-33 | no |
| 15 | rs762551 | 75041917 | C | A | 334695 | -0.06331 | -11.89 | 1.33E-32 | no |
| 15 | rs45468096 | 75043539 | T | C | 334638 | -0.4812 | -9.012 | 2.04E-19 | yes |
| 15 | rs28399419 | 75043669 | T | C | 334414 | -0.157 | -6.45 | 1.12E-10 | yes |
| 15 | rs2472304 | 75044238 | G | A | 334693 | -0.06658 | -13.22 | 6.90E-40 | no |
| 15 | rs2470890 | 75047426 | C | T | 334575 | -0.06623 | -13.15 | 1.78E-39 | no |
| 15 | rs936226 | 75069282 | C | T | 333752 | -0.06121 | -11.31 | 1.15E-29 | no |
| 15 | rs1378942 | 75077367 | C | A | 334640 | -0.06221 | -12.26 | 1.56E-34 | no |
| 15 | rs34862454 | 75101530 | C | T | 333910 | -0.06001 | -11.87 | 1.79E-32 | no |
| 15 | rs6495122 | 75125645 | A | C | 334609 | -0.0615 | -12.74 | 3.73E-37 | no |
| 15 | rs55973697 | 75129584 | C | A | 334682 | 0.1299 | 7.295 | 3.00E-13 | yes |
| 15 | rs12898397 | 75130093 | T | C | 334075 | -0.05828 | -11.85 | 2.26E-32 | no |
| 15 | rs4886615 | 75131661 | A | G | 334646 | -0.05062 | -9.538 | 1.47E-21 | no |
| 15 | rs6938 | 75136261 | C | G | 332531 | -0.05281 | -10.16 | 2.91E-24 | yes |

| | | | | | | | | | |
|---|---|---|---|---|---|---|---|---|---|
| 15 | rs2305668 | 75142761 | G | T | 334547 | -0.04528 | -6.004 | 1.93E-09 | no |
| 15 | rs11630918 | 75155896 | C | T | 334268 | -0.05996 | -12.53 | 5.51E-36 | no |
| 15 | rs12911254 | 75166335 | G | A | 334247 | -0.05757 | -12.04 | 2.11E-33 | no |
| 15 | rs1130741 | 75189930 | A | G | 334515 | -0.05355 | -11.17 | 5.71E-29 | no |
| 15 | rs6495127 | 75194490 | C | T | 334241 | -0.04989 | -9.522 | 1.71E-21 | no |
| 15 | rs12907898 | 75207872 | C | T | 332523 | -0.05264 | -10.94 | 7.82E-28 | no |
| 15 | rs12912839 | 75221245 | C | T | 331594 | -0.05774 | -12.04 | 2.17E-33 | no |
| 15 | rs12912343 | 75251041 | C | G | 334644 | -0.0632 | -13.32 | 1.76E-40 | no |
| 15 | rs6495133 | 75255458 | G | C | 334278 | -0.04834 | -7.635 | 2.26E-14 | yes |
| 15 | rs62029217 | 75261899 | C | T | 334406 | -0.06298 | -13.27 | 3.56E-40 | no |
| 15 | rs12050759 | 75265191 | A | G | 334337 | -0.0411 | -8.011 | 1.14E-15 | no |
| 15 | rs11636892 | 75266049 | G | C | 333799 | -0.06189 | -13.04 | 7.30E-39 | no |
| 15 | rs34359839 | 75274669 | G | A | 334369 | -0.049 | -7.005 | 2.48E-12 | no |
| 15 | rs3812945 | 75289722 | C | T | 334093 | 0.06489 | 13.69 | 1.26E-42 | no |
| 15 | rs8030089 | 75290377 | T | C | 334530 | -0.03527 | -5.815 | 6.07E-09 | no |
| 15 | rs2289582 | 75310394 | C | T | 334353 | -0.03371 | -5.543 | 2.98E-08 | no |
| 15 | rs8042558 | 75320433 | T | G | 333921 | -0.04071 | -7.276 | 3.44E-13 | no |
| 15 | rs12916473 | 75321999 | A | G | 334474 | 0.1124 | 8.899 | 5.67E-19 | no |
| 15 | rs34835684 | 75331805 | G | T | 334240 | -0.03366 | -5.564 | 2.64E-08 | no |
| 15 | rs8043181 | 75380424 | C | T | 334740 | -0.03479 | -5.946 | 2.75E-09 | no |
| 15 | rs12148488 | 75382542 | T | G | 330604 | -0.0604 | -12.62 | 1.77E-36 | no |
| 15 | rs8034281 | 75384227 | T | C | 334596 | -0.04834 | -9.869 | 5.73E-23 | no |
| 15 | rs11638746 | 75403108 | C | A | 334380 | -0.04516 | -8.635 | 5.92E-18 | no |
| 15 | rs1602360 | 75420761 | G | A | 320676 | -0.04432 | -8.21 | 2.22E-16 | no |
| 15 | rs34511319 | 75429717 | A | C | 334246 | -0.04708 | -8.739 | 2.36E-18 | no |
| 15 | rs8025412 | 75444033 | C | T | 334634 | -0.04602 | -9.127 | 7.08E-20 | no |
| 15 | rs58201389 | 75446835 | G | A | 334383 | -0.04643 | -9.201 | 3.56E-20 | no |
| 15 | rs8040372 | 75450557 | A | G | 334113 | -0.04843 | -9.847 | 7.11E-23 | no |
| 15 | rs7170378 | 75457490 | T | C | 333733 | -0.04431 | -9.065 | 1.25E-19 | no |
| 15 | rs11072530 | 75458850 | G | A | 334606 | -0.04472 | -9.091 | 9.85E-20 | no |
| 15 | rs2201002 | 75463150 | G | A | 334109 | -0.04453 | -9.127 | 7.05E-20 | yes |
| 15 | rs1873379 | 75498744 | C | G | 330117 | 0.05366 | 10.2 | 1.99E-24 | no |
| 15 | rs1984586 | 75607479 | A | G | 333858 | 0.04335 | 8.16 | 3.37E-16 | no |
| 15 | rs55653892 | 75610684 | C | G | 334603 | 0.0418 | 7.816 | 5.45E-15 | no |
| 15 | rs111858375 | 75614821 | A | C | 334649 | 0.04288 | 8.089 | 6.02E-16 | no |

| | | | | | | | | | |
|---|---|---|---|---|---|---|---|---|---|
| 15 | rs115991344 | 75614973 | T | C | 334625 | 0.04218 | 7.96 | 1.73E-15 | no |
| 15 | rs77532275 | 75690292 | C | A | 334767 | 0.04236 | 7.932 | 2.16E-15 | no |
| 15 | rs75782768 | 75692186 | T | C | 334498 | 0.04237 | 7.923 | 2.33E-15 | no |
| 15 | rs112994348 | 75698201 | T | C | 332588 | 0.06587 | 6.421 | 1.36E-10 | no |
| 15 | rs34608804 | 75718459 | C | T | 333648 | 0.06845 | 5.749 | 8.99E-09 | no |
| 15 | rs78428611 | 75738292 | A | C | 334565 | 0.04254 | 7.956 | 1.79E-15 | no |
| 15 | rs117174465 | 75753810 | G | A | 334669 | 0.1026 | 5.941 | 2.83E-09 | no |
| 15 | rs76550488 | 75757050 | C | G | 334164 | 0.04422 | 8.27 | 1.35E-16 | no |
| 15 | rs80163632 | 75763377 | G | A | 334120 | 0.04364 | 8.164 | 3.26E-16 | no |
| 15 | rs78521534 | 75769347 | T | C | 322729 | 0.05104 | 7.273 | 3.51E-13 | yes |
| 15 | rs79471369 | 75771103 | C | T | 334515 | 0.04391 | 8.22 | 2.04E-16 | no |
| 15 | rs76063558 | 75775476 | A | G | 334488 | 0.04374 | 8.181 | 2.82E-16 | no |
| 15 | rs77716675 | 75781554 | G | A | 334320 | 0.04352 | 8.135 | 4.14E-16 | no |
| 15 | rs76621678 | 75844953 | T | C | 334424 | 0.03877 | 6.164 | 7.09E-10 | no |
| 15 | rs118028692 | 75847217 | A | C | 334272 | 0.08481 | 6.081 | 1.20E-09 | no |
| 15 | rs74845979 | 75912699 | T | C | 334730 | 0.04398 | 8.098 | 5.59E-16 | no |
| 15 | rs78911138 | 76275641 | T | G | 333859 | 0.06926 | 6.872 | 6.35E-12 | yes |
| 15 | rs868299 | 77873549 | A | G | 333986 | -0.02731 | -5.534 | 3.14E-08 | yes |
| 15 | rs11330240 | 91428521 | C | CT | 334276 | -0.02757 | -5.459 | 4.80E-08 | no |
| 15 | rs7497304 | 91429176 | T | G | 334505 | -0.02761 | -5.467 | 4.57E-08 | yes |
| 16 | rs9940128 | 53800754 | A | G | 334223 | 0.03791 | 7.899 | 2.83E-15 | no |
| 16 | rs1421085 | 53800954 | C | T | 334254 | 0.03859 | 7.987 | 1.39E-15 | no |
| 16 | rs1121980 | 53809247 | A | G | 334456 | 0.03797 | 7.916 | 2.45E-15 | no |
| 16 | rs17817449 | 53813367 | G | T | 334387 | 0.03819 | 7.876 | 3.39E-15 | no |
| 16 | rs8050136 | 53816275 | A | C | 334609 | 0.03826 | 7.887 | 3.10E-15 | no |
| 16 | rs3751812 | 53818460 | T | G | 334637 | 0.03817 | 7.873 | 3.49E-15 | no |
| 16 | rs9939609 | 53820527 | A | T | 334878 | 0.03806 | 7.856 | 3.99E-15 | no |
| 16 | rs7202116 | 53821615 | G | A | 334500 | 0.03805 | 7.847 | 4.28E-15 | no |
| 16 | rs9941349 | 53825488 | T | C | 333785 | 0.03866 | 8.004 | 1.21E-15 | yes |
| 16 | rs9930506 | 53830465 | G | A | 331183 | 0.03627 | 7.558 | 4.12E-14 | no |
| 16 | rs9922619 | 53831771 | T | G | 333782 | 0.03659 | 7.64 | 2.18E-14 | no |
| 16 | rs12149832 | 53842908 | A | G | 334593 | 0.03723 | 7.739 | 1.01E-14 | no |
| 16 | rs11642841 | 53845487 | A | C | 334752 | 0.03354 | 6.938 | 3.97E-12 | no |
| 17 | Affx-13612776 | 17579861 | G | C | 334321 | -0.02699 | -5.701 | 1.20E-08 | no |
| 17 | rs11649804 | 17696755 | A | C | 334655 | -0.03184 | -6.105 | 1.03E-09 | yes |

| | | | | | | | | | |
|---|---|---|---|---|---|---|---|---|---|
| 17 | rs12936927 | 17726965 | C | T | 334125 | -0.02897 | -5.802 | 6.56E-09 | no |
| 17 | rs3744115 | 17749822 | G | A | 334674 | -0.02754 | -5.534 | 3.13E-08 | no |
| 17 | rs11078405 | 17824978 | T | G | 334446 | -0.02901 | -5.914 | 3.35E-09 | no |
| 18 | rs4121765 | 57730096 | G | A | 334400 | 0.02963 | 5.747 | 9.07E-09 | no |
| 18 | rs36030660 | 57735945 | C | T | 334167 | 0.03778 | 7.066 | 1.60E-12 | no |
| 18 | rs7240566 | 57744189 | G | A | 334589 | 0.04109 | 7.498 | 6.49E-14 | no |
| 18 | rs35981845 | 57758860 | G | A | 334533 | 0.02991 | 5.961 | 2.50E-09 | no |
| 18 | rs633265 | 57831468 | T | G | 334409 | 0.02662 | 5.579 | 2.42E-08 | no |
| 18 | rs571312 | 57839769 | A | C | 334744 | 0.04742 | 8.489 | 2.09E-17 | no |
| 18 | rs589850 | 57841891 | A | G | 334385 | 0.02726 | 5.718 | 1.08E-08 | no |
| 18 | rs1350341 | 57842533 | A | G | 334624 | 0.02708 | 5.683 | 1.33E-08 | no |
| 18 | rs17782313 | 57851097 | C | T | 334156 | 0.04803 | 8.576 | 9.86E-18 | no |
| 18 | rs10871777 | 57851763 | G | A | 334610 | 0.04878 | 8.774 | 1.74E-18 | yes |
| 18 | rs489693 | 57882787 | A | C | 334587 | 0.03563 | 7.052 | 1.77E-12 | no |
| 18 | rs12970134 | 57884750 | A | G | 334334 | 0.0418 | 7.815 | 5.50E-15 | no |
| 19 | rs3810291 | 47569003 | G | A | 334674 | -0.0284 | -5.611 | 2.01E-08 | yes |
| 20 | rs6062359 | 62900380 | G | T | 333934 | 0.03034 | 6.344 | 2.24E-10 | yes |

**Table S9: Shared lead SNPs of the cereal and coffee intake.**

| SNP information | | | | | Cereal intake | | | | | | | | Coffee intake | | | | | | | |
|---|---|---|---|---|---|---|---|---|---|---|---|---|---|---|---|---|---|---|---|---|
| CHR | SNP | BP | A1 | A2 | beta | 95%CI_l | 95%CI_h | se | t | r | p | df.e | beta | 95%CI_l | 95%CI_h | se | t | r | p | df.e |
| 6 | rs2504706 | 51258579 | C | T | 0.058 | 0.042 | 0.073 | 0.008 | 7.30 | 0.013 | 2.94E-13 | 334441 | -0.034 | -0.045 | -0.023 | 0.006 | -6.03 | -0.010 | 1.67E-09 | 333816 |
| 7 | rs4410790 | 17284577 | C | T | -0.038 | -0.052 | -0.024 | 0.007 | -5.47 | -0.010 | 4.40E-08 | 334331 | 0.120 | 0.110 | 0.130 | 0.005 | 24.37 | 0.042 | 4.71E-131 | 333705 |
| 15 | rs2472297 | 75027880 | T | C | -0.059 | -0.074 | -0.044 | 0.008 | -7.84 | -0.014 | 4.38E-15 | 334951 | 0.142 | 0.131 | 0.152 | 0.005 | 26.55 | 0.046 | 4.20E-155 | 334321 |

**Table S10: Associations between SNPs and diets.**

| | rs2504706_C | | | | | | | | rs4410790_C | | | | | | | | rs2472297_T | | | | | | | |
|---|---|---|---|---|---|---|---|---|---|---|---|---|---|---|---|---|---|---|---|---|---|---|---|---|
| **Variable** | **beta** | **95%CI_l** | **95%CI_h** | **se** | **t** | **r** | **p** | **df.e** | **beta** | **95%CI_l** | **95%CI_h** | **se** | **t** | **r** | **p** | **df.e** | **beta** | **95%CI_l** | **95%CI_h** | **se** | **t** | **r** | **p** | **df.e** |
| Water intake | 0.012 | 0.001 | 0.024 | 0.006 | 2.13 | 0.004 | 3.28E-02 | 331990 | -0.075 | -0.085 | -0.065 | 0.005 | -14.64 | -0.025 | 1.65E-48 | 331879 | -0.086 | -0.097 | -0.076 | 0.006 | -15.62 | -0.027 | 5.53E-55 | 332497 |
| Tea intake | 0.057 | 0.043 | 0.071 | 0.007 | 7.73 | 0.013 | 1.05E-14 | 332621 | 0.111 | 0.098 | 0.123 | 0.006 | 17.04 | 0.030 | 4.58E-65 | 332509 | 0.148 | 0.134 | 0.162 | 0.007 | 21.03 | 0.036 | 3.82E-98 | 333124 |
| Cooked vegetable intake | -0.001 | -0.008 | 0.007 | 0.004 | -0.16 | 0.000 | 8.70E-01 | 329048 | 0.003 | -0.004 | 0.010 | 0.003 | 0.85 | 0.001 | 3.94E-01 | 328938 | -0.005 | -0.012 | 0.003 | 0.004 | -1.27 | -0.002 | 2.06E-01 | 329549 |
| Salad / raw vegetable intake | 0.011 | 0.001 | 0.020 | 0.005 | 2.25 | 0.004 | 2.42E-02 | 330099 | -0.009 | -0.017 | 0.000 | 0.004 | -2.01 | -0.004 | 4.42E-02 | 329995 | -0.005 | -0.014 | 0.004 | 0.005 | -1.12 | -0.002 | 2.62E-01 | 330603 |
| Fresh fruit intake | 0.034 | 0.026 | 0.042 | 0.004 | 8.53 | 0.015 | 1.53E-17 | 333101 | -0.012 | -0.019 | -0.005 | 0.004 | -3.45 | -0.006 | 5.59E-04 | 332986 | -0.020 | -0.027 | -0.013 | 0.004 | -5.24 | -0.009 | 1.56E-07 | 333615 |
| Dried fruit intake | 0.012 | 0.005 | 0.018 | 0.003 | 3.44 | 0.006 | 5.86E-04 | 329367 | -0.003 | -0.009 | 0.003 | 0.003 | -0.94 | -0.002 | 3.45E-01 | 329257 | -0.003 | -0.009 | 0.003 | 0.003 | -0.91 | -0.002 | 3.63E-01 | 329872 |
| Bread intake | -0.042 | -0.085 | 0.002 | 0.022 | -1.89 | -0.003 | 5.92E-02 | 329979 | 0.041 | 0.003 | 0.080 | 0.020 | 2.12 | 0.004 | 3.43E-02 | 329867 | 0.051 | 0.009 | 0.092 | 0.021 | 2.39 | 0.004 | 1.70E-02 | 330482 |
| Oily fish intake | 0.002 | -0.003 | 0.007 | 0.003 | 0.82 | 0.001 | 4.10E-01 | 333791 | -0.001 | -0.005 | 0.004 | 0.002 | -0.25 | 0.000 | 8.04E-01 | 333675 | 0.002 | -0.003 | 0.007 | 0.002 | 0.85 | 0.001 | 3.95E-01 | 334293 |
| Non-oily fish intake | -0.008 | -0.012 | -0.003 | 0.002 | -3.49 | -0.006 | 4.82E-04 | 333746 | -0.002 | -0.006 | 0.001 | 0.002 | -1.22 | -0.002 | 2.22E-01 | 333631 | -0.003 | -0.007 | 0.001 | 0.002 | -1.56 | -0.003 | 1.19E-01 | 334248 |
| Processed meat intake | 0.004 | -0.002 | 0.010 | 0.003 | 1.41 | 0.002 | 1.60E-01 | 334794 | 0.007 | 0.002 | 0.012 | 0.003 | 2.73 | 0.005 | 6.36E-03 | 334680 | 0.006 | 0.001 | 0.011 | 0.003 | 2.19 | 0.004 | 2.89E-02 | 335299 |
| Poultry intake | 0.001 | -0.004 | 0.006 | 0.002 | 0.54 | 0.001 | 5.89E-01 | 334720 | 0.003 | -0.002 | 0.007 | 0.002 | 1.15 | 0.002 | 2.51E-01 | 334607 | -0.001 | -0.006 | 0.003 | 0.002 | -0.54 | -0.001 | 5.87E-01 | 335227 |
| Beef intake | 0.005 | 0.001 | 0.010 | 0.002 | 2.25 | 0.004 | 2.41E-02 | 334023 | 0.002 | -0.002 | 0.007 | 0.002 | 1.19 | 0.002 | 2.35E-01 | 333914 | 0.003 | -0.002 | 0.007 | 0.002 | 1.19 | 0.002 | 2.33E-01 | 334529 |
| Lamb/mutton intake | 0.011 | 0.007 | 0.015 | 0.002 | 5.64 | 0.010 | 1.67E-08 | 333335 | 0.001 | -0.003 | 0.004 | 0.002 | 0.36 | 0.001 | 7.16E-01 | 333226 | 0.001 | -0.002 | 0.005 | 0.002 | 0.77 | 0.001 | 4.40E-01 | 333842 |
| Pork intake | 0.005 | 0.001 | 0.009 | 0.002 | 2.51 | 0.004 | 1.21E-02 | 333277 | 0.002 | -0.002 | 0.005 | 0.002 | 1.02 | 0.002 | 3.08E-01 | 333170 | 0.000 | -0.004 | 0.004 | 0.002 | 0.04 | 0.000 | 9.69E-01 | 333787 |
| Cheese intake | -0.011 | -0.017 | -0.005 | 0.003 | -3.47 | -0.006 | 5.20E-04 | 327491 | -0.003 | -0.008 | 0.002 | 0.003 | -1.11 | -0.002 | 2.67E-01 | 327380 | 0.000 | -0.006 | 0.005 | 0.003 | -0.16 | 0.000 | 8.69E-01 | 327987 |

**Table S11: Associations between SNP and lifestyle phenotypes.**

| | rs2504706_C | | | | | | | | rs4410790_C | | | | | | | | rs2472297_T | | | | | | | |
|---|---|---|---|---|---|---|---|---|---|---|---|---|---|---|---|---|---|---|---|---|---|---|---|---|
| **Variable** | **beta** | **95%CI_l** | **95%CI_h** | **se** | **t** | **r** | **p** | **df.e** | **beta** | **95%CI_l** | **95%CI_h** | **se** | **t** | **r** | **p** | **df.e** | **beta** | **95%CI_l** | **95%CI_h** | **se** | **t** | **r** | **p** | **df.e** |
| Time spent watching television (TV) | -0.012 | -0.021 | -0.003 | 0.005 | -2.65 | -0.005 | 8.03E-03 | 332902 | 0.012 | 0.004 | 0.020 | 0.004 | 3.02 | 0.005 | 2.54E-03 | 332793 | 0.003 | -0.005 | 0.012 | 0.004 | 0.78 | 0.001 | 4.32E-01 | 333404 |
| Usual walking pace | 0.004 | 0.001 | 0.008 | 0.002 | 2.43 | 0.004 | 1.51E-02 | 333424 | -0.001 | -0.003 | 0.002 | 0.002 | -0.33 | -0.001 | 7.41E-01 | 333312 | -0.002 | -0.005 | 0.001 | 0.002 | -1.17 | -0.002 | 2.43E-01 | 333933 |
| Summed MET minutes per week for all activity | -12.803 | -29.777 | 4.171 | 8.660 | -1.48 | -0.003 | 1.39E-01 | 271649 | 0.548 | -14.417 | 15.514 | 7.635 | 0.07 | 0.000 | 9.43E-01 | 271577 | -9.714 | ##### | 6.517 | 8.281 | -1.17 | -0.002 | 2.41E-01 | 272093 |
| MET minutes per week for moderate activity | -5.049 | -12.741 | 2.644 | 3.925 | -1.29 | -0.002 | 1.98E-01 | 271649 | -0.914 | -7.695 | 5.866 | 3.460 | -0.26 | -0.001 | 7.92E-01 | 271577 | -3.288 | ##### | 4.067 | 3.753 | -0.88 | -0.002 | 3.81E-01 | 272093 |
| MET minutes per week for vigorous activity | -0.026 | -7.492 | 7.440 | 3.809 | -0.01 | 0.000 | 9.95E-01 | 271649 | -1.891 | -8.475 | 4.694 | 3.360 | -0.56 | -0.001 | 5.74E-01 | 271577 | -4.525 | ##### | 2.616 | 3.643 | -1.24 | -0.002 | 2.14E-01 | 272093 |
| MET minutes per week for walking | -7.728 | -14.566 | -0.890 | 3.489 | -2.22 | -0.004 | 2.68E-02 | 271649 | 3.353 | -2.675 | 9.381 | 3.076 | 1.09 | 0.002 | 2.76E-01 | 271577 | -1.901 | -8.438 | 4.636 | 3.335 | -0.57 | -0.001 | 5.69E-01 | 272093 |
| Sleep duration | 0.007 | 0.000 | 0.013 | 0.003 | 2.09 | 0.004 | 3.62E-02 | 333487 | -0.001 | -0.007 | 0.004 | 0.003 | -0.51 | -0.001 | 6.11E-01 | 333372 | 0.000 | -0.005 | 0.006 | 0.003 | 0.14 | 0.000 | 8.86E-01 | 333993 |
| Getting up in morning | -0.010 | -0.014 | -0.006 | 0.002 | -4.64 | -0.008 | 3.56E-06 | 334568 | -0.008 | -0.012 | -0.004 | 0.002 | -4.11 | -0.007 | 3.89E-05 | 334451 | -0.003 | -0.007 | 0.001 | 0.002 | -1.24 | -0.002 | 2.15E-01 | 335072 |
| Morning/evening person (chronotype) | 0.012 | 0.007 | 0.018 | 0.003 | 4.39 | 0.008 | 1.12E-05 | 299433 | 0.001 | -0.003 | 0.006 | 0.002 | 0.59 | 0.001 | 5.55E-01 | 299344 | 0.002 | -0.004 | 0.007 | 0.003 | 0.56 | 0.001 | 5.76E-01 | 299893 |
| Nap during day | 0.003 | 0.000 | 0.006 | 0.002 | 1.73 | 0.003 | 8.33E-02 | 335137 | -0.003 | -0.006 | -0.001 | 0.001 | -2.31 | -0.004 | 2.10E-02 | 335022 | -0.005 | -0.008 | -0.002 | 0.002 | -3.23 | -0.006 | 1.22E-03 | 335644 |
| Sleeplessness / insomnia | -0.001 | -0.005 | 0.003 | 0.002 | -0.64 | -0.001 | 5.19E-01 | 335030 | 0.001 | -0.003 | 0.004 | 0.002 | 0.43 | 0.001 | 6.65E-01 | 334913 | 0.001 | -0.003 | 0.005 | 0.002 | 0.42 | 0.001 | 6.72E-01 | 335536 |
| Daytime dozing / sleeping (narcolepsy) | 0.001 | -0.002 | 0.003 | 0.001 | 0.36 | 0.001 | 7.22E-01 | 334152 | -0.004 | -0.007 | -0.002 | 0.001 | -3.60 | -0.006 | 3.17E-04 | 334034 | -0.006 | -0.008 | -0.003 | 0.001 | -4.28 | -0.007 | 1.91E-05 | 334656 |
| Ever smoked | 0.001 | -0.002 | 0.003 | 0.001 | 0.44 | 0.001 | 6.59E-01 | 334140 | -0.005 | -0.007 | -0.002 | 0.001 | -3.87 | -0.007 | 1.11E-04 | 334030 | -0.002 | -0.005 | 0.001 | 0.001 | -1.56 | -0.003 | 1.18E-01 | 334645 |
| Number of cigarettes currently smoked daily (current cigarette smokers) | -0.041 | -0.219 | 0.138 | 0.091 | -0.44 | -0.003 | 6.57E-01 | 23024 | 0.008 | -0.148 | 0.163 | 0.079 | 0.09 | 0.001 | 9.25E-01 | 23040 | 0.036 | -0.133 | 0.205 | 0.086 | 0.42 | 0.003 | 6.77E-01 | 23068 |
| Alcohol intake frequency | -0.007 | -0.015 | 0.002 | 0.004 | -1.56 | -0.003 | 1.19E-01 | 335029 | 0.001 | -0.007 | 0.008 | 0.004 | 0.19 | 0.000 | 8.50E-01 | 334915 | 0.013 | 0.005 | 0.021 | 0.004 | 3.28 | 0.006 | 1.02E-03 | 335536 |
| Average intake of alcoholic units per week | -0.057 | -0.163 | 0.049 | 0.054 | -1.06 | -0.002 | 2.90E-01 | 331227 | 0.085 | -0.008 | 0.179 | 0.048 | 1.79 | 0.003 | 7.27E-02 | 331114 | 0.161 | 0.060 | 0.262 | 0.052 | 3.12 | 0.005 | 1.82E-03 | 331734 |

**Table S12: Associations between the cereal/coffee intake and lifestyle phenotypes.**

| | Cereal intake | | | | | | | | Coffee intake | | | | | | | |
|---|---|---|---|---|---|---|---|---|---|---|---|---|---|---|---|---|
| Variable | beta | 95%CI_l | 95%CI_h | se | t | r | p | df.e | beta | 95%CI_l | 95%CI_h | se | t | r | p | df.e |
| Time spent watching television (TV) | -0.034 | -0.036 | -0.032 | 0.001 | -33.21 | -0.057 | 2.11E-241 | 333481 | 0.003 | 0.001 | 0.006 | 0.001 | 2.35 | 0.004 | 1.89E-02 | 332848 |
| Usual walking pace | 0.015 | 0.014 | 0.015 | 0.000 | 38.53 | 0.067 | 0.00E+00 | 333879 | -0.003 | -0.004 | -0.002 | 0.001 | -6.24 | -0.011 | 4.30E-10 | 333261 |
| Summed MET minutes per week for all activity | 16.055 | 12.339 | 19.772 | 1.896 | 8.47 | 0.016 | 2.52E-17 | 272389 | -15.916 | -21.186 | -10.647 | 2.689 | -5.92 | -0.011 | 3.23E-09 | 271875 |
| MET minutes per week for moderate activity | 3.943 | 2.259 | 5.628 | 0.859 | 4.59 | 0.009 | 4.46E-06 | 272389 | -8.399 | -10.789 | -6.010 | 1.219 | -6.89 | -0.013 | 5.63E-12 | 271875 |
| MET minutes per week for vigorous activity | 13.333 | 11.700 | 14.966 | 0.833 | 16.00 | 0.031 | 1.36E-57 | 272389 | -5.675 | -7.991 | -3.359 | 1.182 | -4.80 | -0.009 | 1.56E-06 | 271875 |
| MET minutes per week for walking | -1.221 | -2.718 | 0.277 | 0.764 | -1.60 | -0.003 | 1.10E-01 | 272389 | -1.842 | -3.967 | 0.282 | 1.084 | -1.70 | -0.003 | 8.92E-02 | 271875 |
| Sleep duration | 0.012 | 0.011 | 0.013 | 0.001 | 17.54 | 0.030 | 7.62E-69 | 334015 | -0.016 | -0.018 | -0.014 | 0.001 | -16.93 | -0.029 | 2.67E-64 | 333400 |
| Getting up in morning | 0.004 | 0.003 | 0.005 | 0.000 | 8.22 | 0.014 | 2.10E-16 | 335031 | 0.000 | -0.002 | 0.001 | 0.001 | -0.48 | -0.001 | 6.30E-01 | 334401 |
| Morning/evening person (chronotype) | -0.013 | -0.015 | -0.012 | 0.001 | -21.70 | -0.040 | 2.57E-104 | 299974 | 0.015 | 0.014 | 0.017 | 0.001 | 17.59 | 0.032 | 3.05E-69 | 299387 |
| Nap during day | -0.001 | -0.002 | 0.000 | 0.000 | -3.30 | -0.006 | 9.75E-04 | 335581 | -0.001 | -0.002 | 0.000 | 0.001 | -1.66 | -0.003 | 9.62E-02 | 334951 |
| Sleeplessness / insomnia | -0.007 | -0.008 | -0.006 | 0.000 | -15.23 | -0.026 | 2.24E-52 | 335475 | -0.003 | -0.004 | -0.001 | 0.001 | -3.96 | -0.007 | 7.47E-05 | 334845 |
| Daytime dozing / sleeping (narcolepsy) | 0.002 | 0.001 | 0.002 | 0.000 | 5.93 | 0.010 | 3.05E-09 | 334626 | 0.000 | -0.001 | 0.001 | 0.000 | -0.70 | -0.001 | 4.86E-01 | 333999 |
| Ever smoked | -0.016 | -0.016 | -0.015 | 0.000 | -50.78 | -0.087 | 0.00E+00 | 334593 | 0.018 | 0.017 | 0.019 | 0.000 | 40.80 | 0.070 | 0.00E+00 | 333973 |
| Number of cigarettes currently smoked daily (current cigarette smokers) | -0.380 | -0.418 | -0.342 | 0.019 | -19.77 | -0.129 | 2.65E-86 | 23037 | 0.282 | 0.241 | 0.322 | 0.021 | 13.59 | 0.090 | 6.69E-42 | 22727 |
| Alcohol intake frequency | -0.054 | -0.056 | -0.052 | 0.001 | -58.81 | -0.101 | 0.00E+00 | 335482 | 0.038 | 0.035 | 0.040 | 0.001 | 28.94 | 0.050 | 5.77E-184 | 334856 |
| Average intake of alcoholic units per week | -1.245 | -1.268 | -1.222 | 0.012 | -106.90 | -0.182 | 0.00E+00 | 331798 | 0.326 | 0.293 | 0.359 | 0.017 | 19.48 | 0.034 | 1.68E-84 | 331168 |

**Table S13: Significant mediation models of SNPs, lifestyle phenotypes, and the intake of cereal and coffee.**

| | | | Mediation model: X~Y~Z | | | | | | | Mediation model: X~Z~Y | | | | | | |
|---|---|---|---|---|---|---|---|---|---|---|---|---|---|---|---|---|
| | | | X~Z | | | | | | | X~Y | | | | | | |
| X | Y | Z | Beta | 95%CI_l | 95%CI_h | se | T | P | R | Beta | 95%CI_l | 95%CI_h | se | T | P | R |
| rs4410790_C | Cereal intake | Getting up in morning | -0.008 | -0.012 | -0.004 | 0.002 | -4.188 | 2.82E-05 | -0.007 | -0.038 | -0.052 | -0.024 | 0.007 | -5.447 | 5.13E-08 | -0.009 |
| rs4410790_C | Cereal intake | Daytime dozing / sleeping | -0.004 | -0.007 | -0.002 | 0.001 | -3.615 | 3.00E-04 | -0.006 | -0.038 | -0.051 | -0.024 | 0.007 | -5.395 | 6.87E-08 | -0.009 |
| rs2472297_T | Cereal intake | Daytime dozing / sleeping | -0.006 | -0.008 | -0.003 | 0.001 | -4.240 | 2.23E-05 | -0.007 | -0.059 | -0.073 | -0.044 | 0.008 | -7.761 | 8.48E-15 | -0.013 |
| rs2472297_T | Cereal intake | Alcohol intake frequency | 0.013 | 0.005 | 0.021 | 0.004 | 3.243 | 1.18E-03 | 0.006 | -0.060 | -0.074 | -0.045 | 0.008 | -7.891 | 3.01E-15 | -0.014 |
| rs2472297_T | Coffee intake | Alcohol intake frequency | 0.014 | 0.006 | 0.022 | 0.004 | 3.418 | 6.32E-04 | 0.006 | 0.142 | 0.131 | 0.152 | 0.005 | 26.544 | 4.36E-155 | 0.046 |
| | | | X~Y | | | | | | | X~Z | | | | | | |
| X | Y | Z | Beta | 95%CI_l | 95%CI_h | se | T | P | R | Beta | 95%CI_l | 95%CI_h | se | T | P | R |
| rs4410790_C | Cereal intake | Getting up in morning | -0.038 | -0.052 | -0.024 | 0.007 | -5.447 | 5.13E-08 | -0.009 | -0.008 | -0.012 | -0.004 | 0.002 | -4.188 | 2.82E-05 | -0.007 |
| rs4410790_C | Cereal intake | Daytime dozing / sleeping | -0.038 | -0.051 | -0.024 | 0.007 | -5.395 | 6.87E-08 | -0.009 | -0.004 | -0.007 | -0.002 | 0.001 | -3.615 | 3.00E-04 | -0.006 |
| rs2472297_T | Cereal intake | Daytime dozing / sleeping | -0.059 | -0.073 | -0.044 | 0.008 | -7.761 | 8.48E-15 | -0.013 | -0.006 | -0.008 | -0.003 | 0.001 | -4.240 | 2.23E-05 | -0.007 |
| rs2472297_T | Cereal intake | Alcohol intake frequency | -0.060 | -0.074 | -0.045 | 0.008 | -7.891 | 3.01E-15 | -0.014 | 0.013 | 0.005 | 0.021 | 0.004 | 3.243 | 1.18E-03 | 0.006 |
| rs2472297_T | Coffee intake | Alcohol intake frequency | 0.142 | 0.131 | 0.152 | 0.005 | 26.544 | 4.36E-155 | 0.046 | 0.014 | 0.006 | 0.022 | 0.004 | 3.418 | 6.32E-04 | 0.006 |
| | | | Y~Z/X | | | | | | | Z~Y/X | | | | | | |
| X | Y | Z | Beta | 95%CI_l | 95%CI_h | se | T | P | R | Beta | 95%CI_l | 95%CI_h | se | T | P | R |
| rs4410790_C | Cereal intake | Getting up in morning | 0.004 | 0.003 | 0.005 | 0.000 | 7.588 | 3.25E-14 | 0.013 | 0.048 | 0.036 | 0.060 | 0.006 | 7.588 | 3.25E-14 | 0.013 |
| rs4410790_C | Cereal intake | Daytime dozing / sleeping | 0.002 | 0.001 | 0.003 | 0.000 | 6.288 | 3.23E-10 | 0.011 | 0.061 | 0.042 | 0.080 | 0.010 | 6.288 | 3.23E-10 | 0.011 |
| rs2472297_T | Cereal intake | Daytime dozing / sleeping | 0.002 | 0.001 | 0.003 | 0.000 | 6.241 | 4.36E-10 | 0.011 | 0.060 | 0.041 | 0.079 | 0.010 | 6.241 | 4.36E-10 | 0.011 |
| rs2472297_T | Cereal intake | Alcohol intake frequency | -0.054 | -0.056 | -0.052 | 0.001 | -59.152 | 0.00E+00 | -0.102 | -0.191 | -0.197 | -0.185 | 0.003 | -59.152 | 0.00E+00 | -0.102 |
| rs2472297_T | Coffee intake | Alcohol intake frequency | 0.037 | 0.034 | 0.040 | 0.001 | 28.421 | 1.79E-177 | 0.049 | 0.065 | 0.061 | 0.070 | 0.002 | 28.421 | 1.79E-177 | 0.049 |
| | | | X~Z/Y | | | | | | | X~Y/Z | | | | | | |
| X | Y | Z | Beta | 95%CI_l | 95%CI_h | se | T | P | R | Beta | 95%CI_l | 95%CI_h | se | T | P | R |
| rs4410790_C | Cereal intake | Getting up in morning | -0.008 | -0.012 | -0.004 | 0.002 | -4.116 | 3.85E-05 | -0.007 | -0.038 | -0.051 | -0.024 | 0.007 | -5.392 | 6.96E-08 | -0.009 |
| rs4410790_C | Cereal intake | Daytime dozing / sleeping | -0.004 | -0.007 | -0.002 | 0.001 | -3.556 | 3.76E-04 | -0.006 | -0.037 | -0.051 | -0.024 | 0.007 | -5.355 | 8.54E-08 | -0.009 |
| rs2472297_T | Cereal intake | Daytime dozing / sleeping | -0.006 | -0.008 | -0.003 | 0.001 | -4.156 | 3.24E-05 | -0.007 | -0.058 | -0.073 | -0.043 | 0.008 | -7.715 | 1.21E-14 | -0.013 |
| rs2472297_T | Cereal intake | Alcohol intake frequency | 0.010 | 0.002 | 0.018 | 0.004 | 2.453 | 1.42E-02 | 0.004 | -0.057 | -0.072 | -0.042 | 0.008 | -7.600 | 2.97E-14 | -0.013 |
| rs2472297_T | Coffee intake | Alcohol intake frequency | 0.009 | 0.001 | 0.016 | 0.004 | 2.114 | 3.45E-02 | 0.004 | 0.141 | 0.131 | 0.151 | 0.005 | 26.408 | 1.61E-153 | 0.046 |

Model selection

| | | | X~Y~Z | | | X~Z~Y | | | | |
|---|---|---|---|---|---|---|---|---|---|---|
| X | Y | Z | Bootsrap P value | Theoretic P value | Proportion of Mediation (PM) | Bootsrap P value | Theoretic P value | Proportion of Mediation (PM) | Excess PM of X~Y~Z than X~Z~Y | P value of Excess PM |
| rs4410790_C | Cereal intake | Getting up in morning | 0 | 1.55E-05 | 1.71% | 0 | 2.28E-04 | 1.01% | 0.70% | 0.38 |
| rs4410790_C | Cereal intake | Daytime dozing / sleeping | 0 | 1.07E-04 | 1.63% | 0 | 1.89E-03 | 0.73% | 0.90% | 0.214 |
| rs2472297_T | Cereal intake | Daytime dozing / sleeping | 0 | 2.82E-06 | 1.98% | 0 | 4.55E-04 | 0.59% | 1.39% | 0.018 |
| rs2472297_T | Cereal intake | Alcohol intake frequency | 0 | 5.47E-15 | 24.75% | 0 | 1.54E-03 | 4.18% | 20.57% | 0.002 |
| rs2472297_T | Coffee intake | Alcohol intake frequency | 0 | 1.37E-82 | 38.14% | 0 | 8.85E-04 | 0.63% | 37.51% | 0 |

**Table S14: Associations between SNPs and body/blood fat levels.**

| | rs2504706_C | | | | | | | | rs4410790_C | | | | | | | | rs2472297_T | | | | | | | |
|---|---|---|---|---|---|---|---|---|---|---|---|---|---|---|---|---|---|---|---|---|---|---|---|---|
| Variable | beta | 95%CI_l | 95%CI_h | se | t | r | p | df.e | beta | 95%CI_l | 95%CI_h | se | t | r | p | df.e | beta | 95%CI_l | 95%CI_h | se | t | r | p | df.e |
| Body mass index (BMI) | -0.080 | -0.107 | -0.053 | 0.014 | -5.88 | -0.010 | 4.21E-09 | 334176 | 0.046 | 0.022 | 0.070 | 0.012 | 3.83 | 0.007 | 1.30E-04 | 334063 | 0.070 | 0.044 | 0.095 | 0.013 | 5.34 | 0.009 | 9.43E-08 | 334680 |
| Cholesterol | -0.004 | -0.011 | 0.002 | 0.003 | -1.22 | -0.002 | 2.21E-01 | 319620 | 0.005 | -0.001 | 0.011 | 0.003 | 1.64 | 0.003 | 1.02E-01 | 319519 | 0.014 | 0.008 | 0.020 | 0.003 | 4.38 | 0.008 | 1.19E-05 | 320107 |
| HDL cholesterol | -0.001 | -0.003 | 0.001 | 0.001 | -1.08 | -0.002 | 2.81E-01 | 292598 | -0.005 | -0.007 | -0.003 | 0.001 | -4.93 | -0.009 | 8.07E-07 | 292504 | 0.003 | 0.001 | 0.005 | 0.001 | 2.64 | 0.005 | 8.32E-03 | 293041 |
| LDL direct | -0.001 | -0.006 | 0.004 | 0.003 | -0.47 | -0.001 | 6.38E-01 | 319014 | 0.005 | 0.000 | 0.009 | 0.002 | 2.15 | 0.004 | 3.17E-02 | 318915 | 0.010 | 0.006 | 0.015 | 0.002 | 4.23 | 0.007 | 2.39E-05 | 319502 |
| Triglycerides | -0.002 | -0.008 | 0.003 | 0.003 | -0.78 | -0.001 | 4.36E-01 | 319357 | 0.015 | 0.010 | 0.021 | 0.003 | 5.94 | 0.011 | 2.84E-09 | 319256 | -0.005 | -0.011 | 0.000 | 0.003 | -1.88 | -0.003 | 5.98E-02 | 319843 |

Table S15: Associations between the cereal/coffee intake and body/blood fat levels.

| | Cereal intake | | | | | | | | Coffee intake | | | | | | | |
|---|---|---|---|---|---|---|---|---|---|---|---|---|---|---|---|---|
| Variable | beta | 95%CI_l | 95%CI_h | se | t | r | p | df.e | beta | 95%CI_l | 95%CI_h | se | t | r | p | df.e |
| Body mass index (BMI) | -0.131 | -0.137 | -0.125 | 0.003 | -43.92 | -0.076 | 0.00E+00 | 334613 | 0.129 | 0.121 | 0.138 | 0.004 | 30.66 | 0.053 | 3.84E-206 | 333987 |
| Cholesterol | -0.027 | -0.029 | -0.026 | 0.001 | -37.66 | -0.066 | 9.99E-310 | 320045 | 0.026 | 0.024 | 0.028 | 0.001 | 25.15 | 0.044 | 1.89E-139 | 319454 |
| HDL cholesterol | -0.007 | -0.007 | -0.006 | 0.000 | -29.61 | -0.055 | 2.15E-192 | 292978 | -0.001 | -0.002 | -0.001 | 0.000 | -3.49 | -0.006 | 4.81E-04 | 292451 |
| LDL direct | -0.015 | -0.016 | -0.014 | 0.001 | -27.10 | -0.048 | 1.57E-161 | 319441 | 0.022 | 0.020 | 0.023 | 0.001 | 27.37 | 0.048 | 9.02E-165 | 318853 |
| Triglycerides | -0.010 | -0.011 | -0.009 | 0.001 | -15.67 | -0.028 | 2.60E-55 | 319782 | 0.004 | 0.003 | 0.006 | 0.001 | 4.83 | 0.009 | 1.39E-06 | 319191 |

**Table S16: Significant mediation models of SNPs, body/blood fat levels and the intake of cereal and coffee.**

| | | | Mediation model: X~Y~Z | | | | | | | Mediation model: X~Z~Y | | | | | | |
|---|---|---|---|---|---|---|---|---|---|---|---|---|---|---|---|---|
| | | | X~Z | | | | | | | X~Y | | | | | | |
| X | Y | Z | Beta | 95%CI_l | 95%CI_h | se | T | P | R | Beta | 95%CI_l | 95%CI_h | se | T | P | R |
| rs2504706_C | Cereal intake | Body mass index (BMI) | -0.080 | -0.107 | -0.054 | 0.014 | -5.890 | 3.86E-09 | -0.010 | 0.058 | 0.042 | 0.073 | 0.008 | 7.304 | 2.81E-13 | 0.013 |
| rs4410790_C | Cereal intake | Body mass index (BMI) | 0.047 | 0.023 | 0.070 | 0.012 | 3.896 | 9.77E-05 | 0.007 | -0.038 | -0.052 | -0.025 | 0.007 | -5.529 | 3.23E-08 | -0.010 |
| rs2472297_T | Cereal intake | Body mass index (BMI) | 0.069 | 0.044 | 0.095 | 0.013 | 5.317 | 1.06E-07 | 0.009 | -0.059 | -0.074 | -0.045 | 0.008 | -7.871 | 3.52E-15 | -0.014 |
| rs2504706_C | Coffee intake | Body mass index (BMI) | -0.081 | -0.108 | -0.054 | 0.014 | -5.930 | 3.03E-09 | -0.010 | -0.033 | -0.044 | -0.022 | 0.006 | -5.885 | 3.99E-09 | -0.010 |
| rs4410790_C | Coffee intake | Body mass index (BMI) | 0.045 | 0.021 | 0.068 | 0.012 | 3.721 | 1.98E-04 | 0.006 | 0.120 | 0.110 | 0.130 | 0.005 | 24.338 | 1.00E-130 | 0.042 |
| rs2472297_T | Coffee intake | Body mass index (BMI) | 0.067 | 0.042 | 0.093 | 0.013 | 5.161 | 2.46E-07 | 0.009 | 0.142 | 0.132 | 0.153 | 0.005 | 26.577 | 1.83E-155 | 0.046 |
| rs2472297_T | Cereal intake | Cholesterol | 0.014 | 0.008 | 0.020 | 0.003 | 4.444 | 8.85E-06 | 0.008 | -0.059 | -0.074 | -0.044 | 0.008 | -7.602 | 2.93E-14 | -0.013 |
| rs2472297_T | Coffee intake | Cholesterol | 0.014 | 0.008 | 0.020 | 0.003 | 4.481 | 7.44E-06 | 0.008 | 0.142 | 0.131 | 0.152 | 0.005 | 25.863 | 2.50E-147 | 0.046 |
| rs4410790_C | Coffee intake | HDL cholesterol | -0.005 | -0.006 | -0.003 | 0.001 | -4.851 | 1.23E-06 | -0.009 | 0.120 | 0.110 | 0.130 | 0.005 | 22.754 | 1.67E-114 | 0.042 |
| rs2472297_T | Cereal intake | LDL direct | 0.011 | 0.006 | 0.015 | 0.002 | 4.293 | 1.77E-05 | 0.008 | -0.059 | -0.074 | -0.044 | 0.008 | -7.610 | 2.75E-14 | -0.013 |
| rs2472297_T | Coffee intake | LDL direct | 0.010 | 0.006 | 0.015 | 0.002 | 4.262 | 2.02E-05 | 0.008 | 0.142 | 0.131 | 0.153 | 0.005 | 25.864 | 2.44E-147 | 0.046 |
| rs4410790_C | Cereal intake | Triglycerides | 0.015 | 0.010 | 0.021 | 0.003 | 5.929 | 3.05E-09 | 0.011 | -0.037 | -0.051 | -0.023 | 0.007 | -5.193 | 2.07E-07 | -0.009 |
| rs4410790_C | Coffee intake | Triglycerides | 0.015 | 0.010 | 0.020 | 0.003 | 5.807 | 6.36E-09 | 0.010 | 0.119 | 0.109 | 0.129 | 0.005 | 23.596 | 5.39E-123 | 0.042 |
| | | | X~Y | | | | | | | X~Z | | | | | | |
| X | Y | Z | Beta | 95%CI_l | 95%CI_h | se | T | P | R | Beta | 95%CI_l | 95%CI_h | se | T | P | R |
| rs2504706_C | Cereal intake | Body mass index (BMI) | 0.058 | 0.042 | 0.073 | 0.008 | 7.304 | 2.81E-13 | 0.013 | -0.080 | -0.107 | -0.054 | 0.014 | -5.890 | 3.86E-09 | -0.010 |
| rs4410790_C | Cereal intake | Body mass index (BMI) | -0.038 | -0.052 | -0.025 | 0.007 | -5.529 | 3.23E-08 | -0.010 | 0.047 | 0.023 | 0.070 | 0.012 | 3.896 | 9.77E-05 | 0.007 |
| rs2472297_T | Cereal intake | Body mass index (BMI) | -0.059 | -0.074 | -0.045 | 0.008 | -7.871 | 3.52E-15 | -0.014 | 0.069 | 0.044 | 0.095 | 0.013 | 5.317 | 1.06E-07 | 0.009 |
| rs2504706_C | Coffee intake | Body mass index (BMI) | -0.033 | -0.044 | -0.022 | 0.006 | -5.885 | 3.99E-09 | -0.010 | -0.081 | -0.108 | -0.054 | 0.014 | -5.930 | 3.03E-09 | -0.010 |
| rs4410790_C | Coffee intake | Body mass index (BMI) | 0.120 | 0.110 | 0.130 | 0.005 | 24.338 | 1.00E-130 | 0.042 | 0.045 | 0.021 | 0.068 | 0.012 | 3.721 | 1.98E-04 | 0.006 |
| rs2472297_T | Coffee intake | Body mass index (BMI) | 0.142 | 0.132 | 0.153 | 0.005 | 26.577 | 1.83E-155 | 0.046 | 0.067 | 0.042 | 0.093 | 0.013 | 5.161 | 2.46E-07 | 0.009 |
| rs2472297_T | Cereal intake | Cholesterol | -0.059 | -0.074 | -0.044 | 0.008 | -7.602 | 2.93E-14 | -0.013 | 0.014 | 0.008 | 0.020 | 0.003 | 4.444 | 8.85E-06 | 0.008 |
| rs2472297_T | Coffee intake | Cholesterol | 0.142 | 0.131 | 0.152 | 0.005 | 25.863 | 2.50E-147 | 0.046 | 0.014 | 0.008 | 0.020 | 0.003 | 4.481 | 7.44E-06 | 0.008 |
| rs4410790_C | Coffee intake | HDL cholesterol | 0.120 | 0.110 | 0.130 | 0.005 | 22.754 | 1.67E-114 | 0.042 | -0.005 | -0.006 | -0.003 | 0.001 | -4.851 | 1.23E-06 | -0.009 |
| rs2472297_T | Cereal intake | LDL direct | -0.059 | -0.074 | -0.044 | 0.008 | -7.610 | 2.75E-14 | -0.013 | 0.011 | 0.006 | 0.015 | 0.002 | 4.293 | 1.77E-05 | 0.008 |
| rs2472297_T | Coffee intake | LDL direct | 0.142 | 0.131 | 0.153 | 0.005 | 25.864 | 2.44E-147 | 0.046 | 0.010 | 0.006 | 0.015 | 0.002 | 4.262 | 2.02E-05 | 0.008 |
| rs4410790_C | Cereal intake | Triglycerides | -0.037 | -0.051 | -0.023 | 0.007 | -5.193 | 2.07E-07 | -0.009 | 0.015 | 0.010 | 0.021 | 0.003 | 5.929 | 3.05E-09 | 0.011 |
| rs4410790_C | Coffee intake | Triglycerides | 0.119 | 0.109 | 0.129 | 0.005 | 23.596 | 5.39E-123 | 0.042 | 0.015 | 0.010 | 0.020 | 0.003 | 5.807 | 6.36E-09 | 0.010 |
| | | | Y~Z/X | | | | | | | Z~Y/X | | | | | | |
| X | Y | Z | Beta | 95%CI_l | 95%CI_h | se | T | P | R | Beta | 95%CI_l | 95%CI_h | se | T | P | R |
| rs2504706_C | Cereal intake | Body mass index (BMI) | -0.128 | -0.134 | -0.122 | 0.003 | -42.887 | 0.00E+00 | -0.074 | -0.043 | -0.045 | -0.041 | 0.001 | -42.887 | 0.00E+00 | -0.074 |
| rs4410790_C | Cereal intake | Body mass index (BMI) | -0.128 | -0.134 | -0.122 | 0.003 | -42.964 | 0.00E+00 | -0.074 | -0.043 | -0.045 | -0.041 | 0.001 | -42.964 | 0.00E+00 | -0.074 |
| rs2472297_T | Cereal intake | Body mass index (BMI) | -0.128 | -0.134 | -0.122 | 0.003 | -42.934 | 0.00E+00 | -0.074 | -0.043 | -0.045 | -0.041 | 0.001 | -42.934 | 0.00E+00 | -0.074 |
| rs2504706_C | Coffee intake | Body mass index (BMI) | 0.132 | 0.124 | 0.140 | 0.004 | 31.268 | 2.64E-214 | 0.054 | 0.022 | 0.021 | 0.024 | 0.001 | 31.268 | 2.64E-214 | 0.054 |
| rs4410790_C | Coffee intake | Body mass index (BMI) | 0.132 | 0.123 | 0.140 | 0.004 | 31.181 | 3.96E-213 | 0.054 | 0.022 | 0.021 | 0.024 | 0.001 | 31.181 | 3.96E-213 | 0.054 |
| rs2472297_T | Coffee intake | Body mass index (BMI) | 0.131 | 0.123 | 0.140 | 0.004 | 31.105 | 4.15E-212 | 0.054 | 0.022 | 0.021 | 0.023 | 0.001 | 31.105 | 4.15E-212 | 0.054 |
| rs2472297_T | Cereal intake | Cholesterol | -0.028 | -0.029 | -0.026 | 0.001 | -37.942 | 2.63E-314 | -0.067 | -0.163 | -0.171 | -0.155 | 0.004 | -37.942 | 2.63E-314 | -0.067 |
| rs2472297_T | Coffee intake | Cholesterol | 0.025 | 0.023 | 0.027 | 0.001 | 24.610 | 1.32E-133 | 0.044 | 0.075 | 0.069 | 0.081 | 0.003 | 24.610 | 1.32E-133 | 0.044 |
| rs4410790_C | Coffee intake | HDL cholesterol | -0.001 | -0.002 | -0.001 | 0.000 | -3.550 | 3.85E-04 | -0.007 | -0.037 | -0.057 | -0.016 | 0.010 | -3.550 | 3.85E-04 | -0.007 |

| | | | | | | | | | | | | | | | | |
|---|---|---|---|---|---|---|---|---|---|---|---|---|---|---|---|---|
| rs2472297_T | Cereal intake | LDL direct | -0.015 | -0.016 | -0.014 | 0.001 | -27.281 | 1.10E-163 | -0.048 | -0.152 | -0.163 | -0.141 | 0.006 | -27.281 | 1.10E-163 | -0.048 |
| rs2472297_T | Coffee intake | LDL direct | 0.021 | 0.020 | 0.023 | 0.001 | 26.912 | 2.43E-159 | 0.048 | 0.107 | 0.099 | 0.114 | 0.004 | 26.912 | 2.43E-159 | 0.048 |
| rs4410790_C | Cereal intake | Triglycerides | -0.010 | -0.011 | -0.009 | 0.001 | -15.700 | 1.60E-55 | -0.028 | -0.076 | -0.086 | -0.067 | 0.005 | -15.700 | 1.60E-55 | -0.028 |
| rs4410790_C | Coffee intake | Triglycerides | 0.004 | 0.002 | 0.006 | 0.001 | 4.583 | 4.58E-06 | 0.008 | 0.016 | 0.009 | 0.023 | 0.003 | 4.583 | 4.58E-06 | 0.008 |
| | | | X~Z/Y | | | | | | | X~Y/Z | | | | | | |
| **X** | **Y** | **Z** | **Beta** | **95%CI_l** | **95%CI_h** | **se** | **T** | **P** | **R** | **Beta** | **95%CI_l** | **95%CI_h** | **se** | **T** | **P** | **R** |
| rs2504706_C | Cereal intake | Body mass index (BMI) | -0.073 | -0.100 | -0.046 | 0.014 | -5.363 | 8.18E-08 | -0.009 | 0.054 | 0.039 | 0.070 | 0.008 | 6.886 | 5.76E-12 | 0.012 |
| rs4410790_C | Cereal intake | Body mass index (BMI) | 0.042 | 0.018 | 0.065 | 0.012 | 3.496 | 4.73E-04 | 0.006 | -0.036 | -0.050 | -0.023 | 0.007 | -5.254 | 1.49E-07 | -0.009 |
| rs2472297_T | Cereal intake | Body mass index (BMI) | 0.062 | 0.036 | 0.087 | 0.013 | 4.746 | 2.08E-06 | 0.008 | -0.056 | -0.071 | -0.042 | 0.008 | -7.498 | 6.51E-14 | -0.013 |
| rs2504706_C | Coffee intake | Body mass index (BMI) | -0.077 | -0.103 | -0.050 | 0.014 | -5.619 | 1.92E-08 | -0.010 | -0.031 | -0.042 | -0.020 | 0.006 | -5.572 | 2.52E-08 | -0.010 |
| rs4410790_C | Coffee intake | Body mass index (BMI) | 0.029 | 0.005 | 0.053 | 0.012 | 2.409 | 1.60E-02 | 0.004 | 0.119 | 0.109 | 0.129 | 0.005 | 24.172 | 5.64E-129 | 0.042 |
| rs2472297_T | Coffee intake | Body mass index (BMI) | 0.049 | 0.023 | 0.074 | 0.013 | 3.732 | 1.90E-04 | 0.006 | 0.141 | 0.130 | 0.151 | 0.005 | 26.336 | 1.06E-152 | 0.046 |
| rs2472297_T | Cereal intake | Cholesterol | 0.013 | 0.006 | 0.019 | 0.003 | 3.943 | 8.05E-05 | 0.007 | -0.056 | -0.072 | -0.041 | 0.008 | -7.320 | 2.48E-13 | -0.013 |
| rs2472297_T | Coffee intake | Cholesterol | 0.011 | 0.004 | 0.017 | 0.003 | 3.354 | 7.97E-04 | 0.006 | 0.141 | 0.130 | 0.151 | 0.005 | 25.691 | 2.08E-145 | 0.045 |
| rs4410790_C | Coffee intake | HDL cholesterol | -0.004 | -0.006 | -0.003 | 0.001 | -4.697 | 2.64E-06 | -0.009 | 0.120 | 0.110 | 0.130 | 0.005 | 22.721 | 3.48E-114 | 0.042 |
| rs2472297_T | Cereal intake | LDL direct | 0.010 | 0.005 | 0.014 | 0.002 | 3.930 | 8.51E-05 | 0.007 | -0.057 | -0.072 | -0.042 | 0.008 | -7.411 | 1.26E-13 | -0.013 |
| rs2472297_T | Coffee intake | LDL direct | 0.007 | 0.003 | 0.012 | 0.002 | 3.030 | 2.45E-03 | 0.005 | 0.141 | 0.130 | 0.151 | 0.005 | 25.689 | 2.19E-145 | 0.045 |
| rs4410790_C | Cereal intake | Triglycerides | 0.015 | 0.010 | 0.020 | 0.003 | 5.787 | 7.18E-09 | 0.010 | -0.036 | -0.050 | -0.022 | 0.007 | -5.029 | 4.92E-07 | -0.009 |
| rs4410790_C | Coffee intake | Triglycerides | 0.015 | 0.010 | 0.020 | 0.003 | 5.611 | 2.02E-08 | 0.010 | 0.119 | 0.109 | 0.129 | 0.005 | 23.548 | 1.66E-122 | 0.042 |

**Model selection**

| | | | X~Y~Z | | | X~Z~Y | | | | |
|---|---|---|---|---|---|---|---|---|---|---|
| **X** | **Y** | **Z** | **Bootsrap P value** | **Theoretic P value** | **Proportion of Mediation (PM)** | **Bootsrap P value** | **Theoretic P value** | **Proportion of Mediation (PM)** | **Excess PM of X~Y~Z than X~Z~Y** | **P value of Excess PM** |
| rs2504706_C | Cereal intake | Body mass index (BMI) | 0 | 1.68E-12 | 9.19% | 0 | 4.64E-09 | 5.97% | 3.21% | 0.3 |
| rs4410790_C | Cereal intake | Body mass index (BMI) | 0 | 8.95E-08 | 10.53% | 0 | 8.61E-05 | 5.23% | 5.30% | 0.234 |
| rs2472297_T | Cereal intake | Body mass index (BMI) | 0 | 1.60E-14 | 10.97% | 0 | 2.39E-07 | 5.00% | 5.97% | 0.07 |
| rs2504706_C | Coffee intake | Body mass index (BMI) | 0 | 2.66E-09 | 5.37% | 0 | 6.45E-09 | 5.45% | -0.08% | 0.996 |
| rs4410790_C | Coffee intake | Body mass index (BMI) | 0 | 1.41E-81 | 35.31% | 0 | 1.74E-04 | 0.83% | 34.48% | 0 |
| rs2472297_T | Coffee intake | Body mass index (BMI) | 0 | 6.72E-83 | 27.71% | 0 | 6.47E-07 | 1.04% | 26.66% | 0 |
| rs2472297_T | Cereal intake | Cholesterol | 0 | 6.51E-14 | 11.46% | 0 | 1.17E-05 | 3.92% | 7.54% | 0.024 |
| rs2472297_T | Coffee intake | Cholesterol | 0 | 6.82E-68 | 25.14% | 0 | 1.43E-05 | 0.75% | 24.38% | 0 |
| rs4410790_C | Coffee intake | HDL cholesterol | 0 | 2.28E-04 | 3.09% | 0 | 4.61E-03 | 0.14% | 2.95% | 0 |
| rs2472297_T | Cereal intake | LDL direct | 0 | 1.80E-13 | 8.56% | 0 | 3.04E-05 | 2.72% | 5.83% | 0.012 |
| rs2472297_T | Coffee intake | LDL direct | 0 | 4.65E-75 | 28.92% | 0 | 4.11E-05 | 0.79% | 28.13% | 0 |
| rs4410790_C | Cereal intake | Triglycerides | 0 | 1.33E-06 | 2.44% | 0 | 3.79E-08 | 3.18% | -0.74% | 0.588 |
| rs4410790_C | Coffee intake | Triglycerides | 0 | 1.04E-05 | 3.30% | 0 | 4.52E-04 | 0.20% | 3.10% | 0 |

**Table S17: Associations between the GMV-association patterns of cognitive functions and the GMV-association patterns of the intake of cereal and coffee (follow-up).**

| | Cereal intake | | Coffee intake | |
|---|---|---|---|---|
| Variable | R | P_perm | R | P_perm |
| Fluid intelligence | 0.6292 | 2.00E-04 | -0.4230 | 1.52E-02 |
| Numeric memory | 0.6566 | 0.00E+00 | -0.4553 | 8.20E-03 |
| Prospective memory | 0.4812 | 4.80E-03 | -0.4259 | 1.36E-02 |
| Reaction time | -0.4430 | 9.80E-03 | 0.3069 | 8.47E-02 |
| Pairs matching | -0.4396 | 8.40E-03 | 0.5763 | 2.00E-04 |
| Matrix pattern completion | 0.7104 | 0.00E+00 | -0.6213 | 1.00E-04 |
| Symbol digit substitution | 0.6875 | 0.00E+00 | -0.6187 | 0.00E+00 |
| Tower rearranging | 0.4178 | 1.66E-02 | -0.3914 | 2.71E-02 |
| Numeric trail making | 0.6251 | 0.00E+00 | -0.6043 | 0.00E+00 |
| Alphanumeric trail making | 0.6873 | 0.00E+00 | -0.7018 | 0.00E+00 |

**Table S18: Associations between the GMV-association patterns of cognitive functions and the GMV-association patterns of the intake of cereal and coffee (baseline).**

| | Cereal intake | | Coffee intake | |
|---|---|---|---|---|
| **Variable** | **R** | **P_perm** | **R** | **P_perm** |
| Fluid intelligence | 0.6177 | 0.00E+00 | -0.4353 | 1.24E-02 |
| Numeric memory | 0.0717 | 6.91E-01 | 0.1005 | 5.79E-01 |
| Prospective memory | 0.4122 | 2.00E-02 | -0.3661 | 3.50E-02 |
| Reaction time | -0.3931 | 2.06E-02 | 0.2239 | 2.14E-01 |
| Pairs matching | -0.4448 | 9.30E-03 | 0.3957 | 2.13E-02 |
| Matrix pattern completion | 0.7693 | 0.00E+00 | -0.6601 | 0.00E+00 |
| Symbol digit substitution | 0.7832 | 0.00E+00 | -0.6864 | 0.00E+00 |
| Tower rearranging | 0.4482 | 8.10E-03 | -0.4022 | 2.21E-02 |
| Numeric trail making | 0.6506 | 0.00E+00 | -0.5945 | 2.00E-04 |
| Alphanumeric trail making | 0.7320 | 0.00E+00 | -0.6902 | 1.00E-04 |

**Table S19: Associations between the GMV-association patterns of the cereal/coffee intake and the gene-expression patterns of eQTL genes.**

| Gene information | | | | | | Cereal intake | | | Coffee intake | | |
|---|---|---|---|---|---|---|---|---|---|---|---|
| ensg | symbol | chr | start | end | IndSigSNPs | R | P_perm | FDR | R | P_perm | FDR |
| ENSG00000170915 | PAQR8 | 6 | 52226219 | 52272575 | rs2504706 | -0.1236 | 0.5728 | 0.7731 | 0.0121 | 0.9553 | 0.9818 |
| ENSG00000106546 | AHR | 7 | 17338246 | 17385776 | rs4410790 | 0.2110 | 0.4152 | 0.7107 | -0.5764 | 0.0030 | 0.0333 |
| ENSG00000048052 | HDAC9 | 7 | 18126572 | 19042039 | rs4410790 | 0.3952 | 0.0703 | 0.2724 | -0.5828 | 0.0016 | 0.0333 |
| ENSG00000071189 | SNX13 | 7 | 17830385 | 17980124 | rs4410790 | -0.0009 | 0.9953 | 0.9953 | -0.1543 | 0.2854 | 0.6452 |
| ENSG00000171243 | SOSTDC1 | 7 | 16501106 | 16570205 | rs4410790 | 0.1868 | 0.4992 | 0.7255 | -0.4163 | 0.0946 | 0.3087 |
| ENSG00000106537 | TSPAN13 | 7 | 16793160 | 16824161 | rs4410790 | 0.2506 | 0.3784 | 0.7107 | -0.6340 | 0.0043 | 0.0333 |
| ENSG00000103653 | CSK | 15 | 75074398 | 75095539 | rs2472297 | -0.1701 | 0.4239 | 0.7107 | 0.5100 | 0.0020 | 0.0333 |
| ENSG00000129009 | ISLR | 15 | 74466012 | 74469213 | rs2472297 | 0.1493 | 0.5736 | 0.7731 | -0.4290 | 0.0655 | 0.2707 |
| ENSG00000129038 | LOXL1 | 15 | 74218330 | 74244478 | rs2472297 | 0.1000 | 0.6380 | 0.8070 | -0.2116 | 0.2914 | 0.6452 |
| ENSG00000140400 | MAN2C1 | 15 | 75648133 | 75660971 | rs2472297 | -0.0210 | 0.9234 | 0.9818 | 0.1608 | 0.4241 | 0.7107 |
| ENSG00000178802 | MPI | 15 | 75182346 | 75191798 | rs2472297 | 0.2229 | 0.2847 | 0.6452 | -0.4723 | 0.0035 | 0.0333 |
| ENSG00000140464 | PML | 15 | 74287014 | 74340153 | rs2472297 | 0.2057 | 0.2529 | 0.6272 | -0.4198 | 0.0043 | 0.0333 |
| ENSG00000169410 | PTPN9 | 15 | 75759462 | 75871630 | rs2472297 | 0.1916 | 0.4476 | 0.7116 | -0.3645 | 0.1209 | 0.3569 |
| ENSG00000138623 | SEMA7A | 15 | 74701630 | 74726808 | rs2472297 | -0.1685 | 0.3427 | 0.7107 | -0.0681 | 0.7029 | 0.8070 |
| ENSG00000067221 | STOML1 | 15 | 74275547 | 74286963 | rs2472297 | 0.3875 | 0.1639 | 0.4619 | -0.6437 | 0.0033 | 0.0333 |
| ENSG00000140497 | SCAMP2 | 15 | 75136071 | 75165706 | rs2472297 | -0.0420 | 0.7987 | 0.8843 | -0.1404 | 0.3915 | 0.7107 |
| ENSG00000169371 | SNUPN | 15 | 75890424 | 75918810 | rs2472297 | 0.3137 | 0.2141 | 0.5531 | -0.4634 | 0.0420 | 0.1864 |
| ENSG00000169375 | SIN3A | 15 | 75661720 | 75748183 | rs2472297 | -0.1270 | 0.5032 | 0.7255 | 0.4013 | 0.0164 | 0.0924 |
| ENSG00000140365 | COMMD4 | 15 | 75628232 | 75634268 | rs2472297 | 0.1329 | 0.3881 | 0.7107 | -0.3447 | 0.0074 | 0.0459 |
| ENSG00000177971 | IMP3 | 15 | 75931426 | 75941047 | rs2472297 | 0.1356 | 0.5161 | 0.7272 | -0.2717 | 0.1733 | 0.4672 |
| ENSG00000167173 | C15orf39 | 15 | 75487984 | 75504510 | rs2472297 | 0.0865 | 0.6984 | 0.8070 | -0.1532 | 0.4910 | 0.7255 |
| ENSG00000178761 | FAM219B | 15 | 75192328 | 75199462 | rs2472297 | -0.0090 | 0.9660 | 0.9818 | -0.0842 | 0.6825 | 0.8070 |
| ENSG00000167178 | ISLR2 | 15 | 74392652 | 74430881 | rs2472297 | 0.2473 | 0.3861 | 0.7107 | -0.4618 | 0.0782 | 0.2852 |
| ENSG00000138621 | PPCDC | 15 | 75315896 | 75409803 | rs2472297 | -0.1075 | 0.5919 | 0.7808 | 0.0802 | 0.6957 | 0.8070 |
| ENSG00000140398 | NEIL1 | 15 | 75639296 | 75647592 | rs2472297 | -0.1162 | 0.6314 | 0.8070 | 0.1878 | 0.4364 | 0.7116 |
| ENSG00000140506 | LMAN1L | 15 | 75105057 | 75118099 | rs2472297 | -0.0849 | 0.6943 | 0.8070 | 0.3244 | 0.0937 | 0.3087 |
| ENSG00000179151 | EDC3 | 15 | 74922899 | 74988633 | rs2472297 | 0.0631 | 0.7842 | 0.8840 | 0.0137 | 0.9533 | 0.9818 |
| ENSG00000138629 | UBL7 | 15 | 74738318 | 74753523 | rs2472297 | 0.1920 | 0.4862 | 0.7255 | -0.5072 | 0.0294 | 0.1519 |
| ENSG00000182950 | ODF3L1 | 15 | 76016318 | 76020029 | rs2472297 | -0.0162 | 0.9413 | 0.9818 | 0.3239 | 0.1104 | 0.3422 |
| ENSG00000198794 | SCAMP5 | 15 | 75249560 | 75313837 | rs2472297 | 0.2328 | 0.3772 | 0.7107 | -0.4699 | 0.0421 | 0.1864 |
| ENSG00000213578 | CPLX3 | 15 | 75118888 | 75124141 | rs2472297 | 0.4721 | 0.0029 | 0.0333 | -0.4429 | 0.0072 | 0.0459 |

**Table S20: Association between the GMV-association patterns of the cognitive functions and the gene-expression patterns of eQTL genes.**

| Gene symbol | CPLX3 | | |
|---|---|---|---|
| | R | P_perm | FDR |
| Fluid intelligence | 0.42267375 | 0.0098 | 0.016333333 |
| Numeric memory | 0.490723015 | 0.0013 | 0.0065 |
| Prospective memory | 0.442872557 | 0.0074 | 0.0148 |
| Reaction time | -0.228002002 | 0.2351 | 0.2351 |
| Pairs matching | -0.327496513 | 0.0704 | 0.078222222 |
| Matrix pattern completion | 0.456387352 | 0.0042 | 0.014 |
| Symbol digit substitution | 0.394847484 | 0.0192 | 0.027428571 |
| Tower rearranging | 0.338529308 | 0.0613 | 0.076625 |
| Numeric trail making | 0.441621796 | 0.0064 | 0.0148 |
| Alphanumeric trail making | 0.545641373 | 0 | 0 |

#### Table S21: Main results of the replication sample.

##### A. Associations between total grey matter volume of the brain and the intake of cereal and coffee.

| | Associations between TGMV and the intake of cereal and coffee (baseline). | | | | | | | |
|---|---|---|---|---|---|---|---|---|
| | beta | 95%CI_l | 95%CI_h | se | t | r | $p_{one-tailed}$ | df.e |
| Cereal intake | 209.57 | 157.19 | 261.94 | 26.72 | 7.843 | 0.061 | 2.34E-15 | 16388 |
| Coffee intake | -296.37 | -375.04 | -217.71 | 40.13 | -7.385 | -0.058 | 7.99E-14 | 16307 |
| | Associations between TGMV and the intake of cereal and coffee (Follow-up). | | | | | | | |
| | beta | 95%CI_l | 95%CI_h | se | t | r | $p_{one-tailed}$ | df.e |
| Cereal intake | 272.05 | 222.05 | 322.05 | 25.51 | 10.666 | 0.083 | 9.01E-27 | 16304 |
| Coffee intake | -277.84 | -362.70 | -192.99 | 43.29 | -6.418 | -0.050 | 7.09E-11 | 16244 |
| | Associations between TGMV and the intake of cereal and coffee (Follow-up). | | | | | | | |
| | beta | 95%CI_l | 95%CI_h | se | t | r | $p_{one-tailed}$ | df.e |
| Cereal intake | 76.63 | 15.02 | 138.24 | 31.43 | 2.438 | 0.019 | 7.39E-03 | 16287 |
| Coffee intake | -229.29 | -335.37 | -123.21 | 54.12 | -4.237 | -0.033 | 1.14E-05 | 16159 |

##### B: Associations between the GMV-association patterns of cognitive functions and the GMV-association patterns of the intake of cereal and coffee.

| | Baseline | | | | Follow-up | | | |
|---|---|---|---|---|---|---|---|---|
| | Cereal intake | | Coffee intake | | Cereal intake | | Coffee intake | |
| | R | P_one-tailed | R | P_one-tailed | R | P_one-tailed | R | P_one-tailed |
| Fluid intelligence | 0.538 | 6.0E-04 | NA | | 0.724 | 0.0E+00 | NA | |
| Matrix pattern completion | 0.776 | 0.0E+00 | -0.417 | 7.7E-03 | 0.782 | 0.0E+00 | -0.403 | 1.0E-02 |
| Symbol digit substitution | 0.853 | 0.0E+00 | -0.554 | 7.5E-04 | 0.769 | 0.0E+00 | -0.561 | 3.0E-04 |
| Numeric trail making | 0.687 | 0.0E+00 | -0.547 | 4.5E-04 | 0.578 | 4.5E-04 | -0.536 | 1.2E-03 |
| Alphanumeric trail making | 0.741 | 0.0E+00 | -0.576 | 2.0E-04 | 0.770 | 0.0E+00 | -0.579 | 2.5E-04 |

**C. Associations between the GMV-association patterns of the cereal/coffee intake and the gene-expression patterns of CPLX gene.**

| | Cereal intake | | Coffee intake | |
|---|---|---|---|---|
| | R | P_one-tailed | R | P_one-tailed |
| CPLX gene | 0.396 | 1.1E-02 | -0.360 | 2.0E-02 |

**D. Association between the GMV-association patterns of the cognitive functions and the gene-expression patterns of CPLX gene.**

| | R | P_one-tailed |
|---|---|---|
| Fluid intelligence | 0.476 | 1.4E-03 |
| Numeric memory | 0.466 | 1.8E-03 |
| Prospective memory | 0.388 | 1.1E-02 |
| Matrix pattern completion | 0.474 | 1.1E-03 |
| Symbol digit substitution | 0.468 | 1.8E-03 |
| Numeric trail making | 0.463 | 2.1E-03 |
| Alphanumeric trail making | 0.500 | 7.0E-04 |

Table S22: Results of modified mendelian randomisation analysis.

A. Association between PRS for TGV and TGV.

| PRS | beta | 95%CI_l | 95%CI_h | se | t | r | p_one-tailed | df.e |
|---|---|---|---|---|---|---|---|---|
| TGV PRS | 1.90E-04 | 1.56E-04 | 2.24E-04 | 1.74E-05 | 10.951 | 0.097 | 4.39E-28 | 12736 |
| TGV/Cereal0.05 PRS | 1.80E-04 | 1.45E-04 | 2.14E-04 | 1.77E-05 | 10.117 | 0.089 | 2.87E-24 | 12736 |
| TGV/Cereal0.10 PRS | 1.73E-04 | 1.37E-04 | 2.08E-04 | 1.82E-05 | 9.488 | 0.084 | 1.39E-21 | 12736 |
| TGV/Cereal0.15 PRS | 1.68E-04 | 1.31E-04 | 2.04E-04 | 1.86E-05 | 9.003 | 0.080 | 1.25E-19 | 12736 |
| TGV/Cereal0.20 PRS | 1.67E-04 | 1.30E-04 | 2.05E-04 | 1.91E-05 | 8.744 | 0.077 | 1.27E-18 | 12736 |
| TGV/Cereal0.25 PRS | 1.66E-04 | 1.28E-04 | 2.05E-04 | 1.98E-05 | 8.424 | 0.074 | 2.01E-17 | 12736 |
| TGV/Cereal0.30 PRS | 1.69E-04 | 1.29E-04 | 2.09E-04 | 2.04E-05 | 8.260 | 0.073 | 8.01E-17 | 12736 |
| TGV/Cereal0.35 PRS | 1.63E-04 | 1.21E-04 | 2.04E-04 | 2.11E-05 | 7.725 | 0.068 | 6.00E-15 | 12736 |
| TGV/Cereal0.40 PRS | 1.74E-04 | 1.32E-04 | 2.17E-04 | 2.18E-05 | 7.984 | 0.071 | 7.66E-16 | 12736 |
| TGV/Cereal0.45 PRS | 1.74E-04 | 1.30E-04 | 2.19E-04 | 2.26E-05 | 7.714 | 0.068 | 6.53E-15 | 12736 |
| TGV/Cereal0.50 PRS | 1.83E-04 | 1.37E-04 | 2.29E-04 | 2.36E-05 | 7.763 | 0.069 | 4.46E-15 | 12736 |
| TGV/Coffee0.05 PRS | 1.78E-04 | 1.43E-04 | 2.13E-04 | 1.78E-05 | 10.036 | 0.089 | 6.46E-24 | 12736 |
| TGV/Coffee0.10 PRS | 1.78E-04 | 1.42E-04 | 2.14E-04 | 1.81E-05 | 9.820 | 0.087 | 5.58E-23 | 12736 |
| TGV/Coffee0.15 PRS | 1.70E-04 | 1.34E-04 | 2.07E-04 | 1.86E-05 | 9.155 | 0.081 | 3.11E-20 | 12736 |
| TGV/Coffee0.20 PRS | 1.78E-04 | 1.40E-04 | 2.15E-04 | 1.90E-05 | 9.332 | 0.082 | 6.06E-21 | 12736 |
| TGV/Coffee0.25 PRS | 1.78E-04 | 1.40E-04 | 2.17E-04 | 1.96E-05 | 9.085 | 0.080 | 5.96E-20 | 12736 |
| TGV/Coffee0.30 PRS | 1.76E-04 | 1.36E-04 | 2.16E-04 | 2.02E-05 | 8.708 | 0.077 | 1.74E-18 | 12736 |
| TGV/Coffee0.35 PRS | 1.70E-04 | 1.29E-04 | 2.11E-04 | 2.09E-05 | 8.139 | 0.072 | 2.18E-16 | 12736 |
| TGV/Coffee0.40 PRS | 1.67E-04 | 1.25E-04 | 2.10E-04 | 2.17E-05 | 7.721 | 0.068 | 6.21E-15 | 12736 |
| TGV/Coffee0.45 PRS | 1.57E-04 | 1.13E-04 | 2.01E-04 | 2.26E-05 | 6.950 | 0.061 | 1.92E-12 | 12736 |
| TGV/Coffee0.50 PRS | 1.57E-04 | 1.11E-04 | 2.03E-04 | 2.35E-05 | 6.678 | 0.059 | 1.26E-11 | 12736 |

B. Association between PRS for cereal intake and cereal intake.

| PRS | beta | 95%CI_l | 95%CI_h | se | t | r | p_one-tailed | df.e |
|---|---|---|---|---|---|---|---|---|
| Cereal PRS | 3.37E-05 | 2.65E-05 | 4.09E-05 | 3.67E-06 | 9.197 | 0.081 | 2.13E-20 | 12739 |
| Cereal/TGV0.05 PRS | 3.29E-05 | 2.56E-05 | 4.01E-05 | 3.72E-06 | 8.832 | 0.078 | 5.80E-19 | 12739 |
| Cereal/TGV0.10 PRS | 3.33E-05 | 2.59E-05 | 4.08E-05 | 3.80E-06 | 8.773 | 0.077 | 9.77E-19 | 12739 |

| | | | | | | | | |
|---|---|---|---|---|---|---|---|---|
| Cereal/TGV0.15 PRS | 3.44E-05 | 2.68E-05 | 4.20E-05 | 3.88E-06 | 8.884 | 0.078 | 3.66E-19 | 12739 |
| Cereal/TGV0.20 PRS | 3.44E-05 | 2.67E-05 | 4.22E-05 | 3.96E-06 | 8.688 | 0.077 | 2.08E-18 | 12739 |
| Cereal/TGV0.25 PRS | 3.42E-05 | 2.62E-05 | 4.21E-05 | 4.06E-06 | 8.414 | 0.074 | 2.20E-17 | 12739 |
| Cereal/TGV0.30 PRS | 3.48E-05 | 2.66E-05 | 4.30E-05 | 4.18E-06 | 8.322 | 0.074 | 4.75E-17 | 12739 |
| Cereal/TGV0.35 PRS | 3.34E-05 | 2.50E-05 | 4.19E-05 | 4.31E-06 | 7.755 | 0.069 | 4.75E-15 | 12739 |
| Cereal/TGV0.40 PRS | 3.15E-05 | 2.28E-05 | 4.02E-05 | 4.45E-06 | 7.082 | 0.063 | 7.45E-13 | 12739 |
| Cereal/TGV0.45 PRS | 3.04E-05 | 2.13E-05 | 3.95E-05 | 4.64E-06 | 6.541 | 0.058 | 3.17E-11 | 12739 |
| Cereal/TGV0.50 PRS | 3.14E-05 | 2.18E-05 | 4.10E-05 | 4.88E-06 | 6.441 | 0.057 | 6.16E-11 | 12739 |

**C. Association between PRS for coffee intake and coffee intake.**

| PRS | beta | 95%CI_l | 95%CI_h | se | t | r | p_one-tailed | df.e |
|---|---|---|---|---|---|---|---|---|
| Coffee PRS | 3.52E-05 | 2.70E-05 | 4.33E-05 | 4.15E-06 | 8.475 | 0.075 | 1.31E-17 | 12739 |
| Coffee/TGV0.05 PRS | 3.21E-05 | 2.40E-05 | 4.01E-05 | 4.11E-06 | 7.801 | 0.069 | 3.30E-15 | 12739 |
| Coffee/TGV0.10 PRS | 3.21E-05 | 2.39E-05 | 4.02E-05 | 4.16E-06 | 7.713 | 0.068 | 6.59E-15 | 12739 |
| Coffee/TGV0.15 PRS | 3.04E-05 | 2.20E-05 | 3.87E-05 | 4.25E-06 | 7.142 | 0.063 | 4.85E-13 | 12739 |
| Coffee/TGV0.20 PRS | 3.26E-05 | 2.41E-05 | 4.11E-05 | 4.35E-06 | 7.488 | 0.066 | 3.72E-14 | 12739 |
| Coffee/TGV0.25 PRS | 3.44E-05 | 2.57E-05 | 4.32E-05 | 4.46E-06 | 7.723 | 0.068 | 6.12E-15 | 12739 |
| Coffee/TGV0.30 PRS | 3.44E-05 | 2.54E-05 | 4.33E-05 | 4.58E-06 | 7.508 | 0.066 | 3.20E-14 | 12739 |
| Coffee/TGV0.35 PRS | 3.43E-05 | 2.51E-05 | 4.35E-05 | 4.70E-06 | 7.296 | 0.065 | 1.57E-13 | 12739 |
| Coffee/TGV0.40 PRS | 3.38E-05 | 2.42E-05 | 4.33E-05 | 4.87E-06 | 6.938 | 0.061 | 2.08E-12 | 12739 |
| Coffee/TGV0.45 PRS | 3.39E-05 | 2.40E-05 | 4.38E-05 | 5.06E-06 | 6.702 | 0.059 | 1.07E-11 | 12739 |
| Coffee/TGV0.50 PRS | 3.62E-05 | 2.59E-05 | 4.65E-05 | 5.23E-06 | 6.916 | 0.061 | 2.44E-12 | 12739 |

**D. Association between PRS for the intake of cereal/coffee and TGV.**

| PRS | beta | 95%CI_l | 95%CI_h | se | t | r | p_one-tailed | df.e |
|---|---|---|---|---|---|---|---|---|
| Cereal PRS | 2.67E-09 | 4.90E-10 | 4.85E-09 | 1.11E-09 | 2.400 | 0.021 | 8.20E-03 | 12736 |
| Cereal/TGV0.05 PRS | 2.03E-09 | -1.88E-10 | 4.24E-09 | 1.13E-09 | 1.794 | 0.016 | 3.64E-02 | 12736 |
| Cereal/TGV0.10 PRS | 1.67E-09 | -5.92E-10 | 3.93E-09 | 1.15E-09 | 1.446 | 0.013 | 7.40E-02 | 12736 |
| Cereal/TGV0.15 PRS | 1.82E-09 | -4.82E-10 | 4.13E-09 | 1.18E-09 | 1.551 | 0.014 | 6.05E-02 | 12736 |
| Cereal/TGV0.20 PRS | 1.99E-09 | -3.63E-10 | 4.35E-09 | 1.20E-09 | 1.658 | 0.015 | 4.86E-02 | 12736 |
| Cereal/TGV0.25 PRS | 2.02E-09 | -3.96E-10 | 4.44E-09 | 1.23E-09 | 1.639 | 0.015 | 5.06E-02 | 12736 |

| | | | | | | | | |
|---|---|---|---|---|---|---|---|---|
| Cereal/TGV0.30 PRS | 2.03E-09 | -4.55E-10 | 4.52E-09 | 1.27E-09 | 1.602 | 0.014 | 5.46E-02 | 12736 |
| Cereal/TGV0.35 PRS | 2.29E-09 | -2.72E-10 | 4.85E-09 | 1.31E-09 | 1.752 | 0.016 | 3.99E-02 | 12736 |
| Cereal/TGV0.40 PRS | 2.12E-09 | -5.23E-10 | 4.77E-09 | 1.35E-09 | 1.573 | 0.014 | 5.79E-02 | 12736 |
| Cereal/TGV0.45 PRS | 1.85E-09 | -9.12E-10 | 4.60E-09 | 1.41E-09 | 1.312 | 0.012 | 9.47E-02 | 12736 |
| Cereal/TGV0.50 PRS | 1.94E-09 | -9.54E-10 | 4.84E-09 | 1.48E-09 | 1.315 | 0.012 | 9.43E-02 | 12736 |
| Coffee PRS | -2.50E-09 | -4.15E-09 | -8.52E-10 | 8.42E-10 | -2.973 | -0.026 | 1.48E-03 | 12736 |
| Coffee/TGV0.05 PRS | -2.27E-09 | -3.91E-09 | -6.41E-10 | 8.33E-10 | -2.730 | -0.024 | 3.17E-03 | 12736 |
| Coffee/TGV0.10 PRS | -2.13E-09 | -3.78E-09 | -4.80E-10 | 8.43E-10 | -2.529 | -0.022 | 5.72E-03 | 12736 |
| Coffee/TGV0.15 PRS | -1.97E-09 | -3.66E-09 | -2.77E-10 | 8.62E-10 | -2.281 | -0.020 | 1.13E-02 | 12736 |
| Coffee/TGV0.20 PRS | -1.92E-09 | -3.65E-09 | -1.85E-10 | 8.83E-10 | -2.170 | -0.019 | 1.50E-02 | 12736 |
| Coffee/TGV0.25 PRS | -2.25E-09 | -4.02E-09 | -4.74E-10 | 9.04E-10 | -2.485 | -0.022 | 6.49E-03 | 12736 |
| Coffee/TGV0.30 PRS | -1.94E-09 | -3.76E-09 | -1.21E-10 | 9.28E-10 | -2.091 | -0.019 | 1.83E-02 | 12736 |
| Coffee/TGV0.35 PRS | -2.09E-09 | -3.96E-09 | -2.21E-10 | 9.54E-10 | -2.192 | -0.019 | 1.42E-02 | 12736 |
| Coffee/TGV0.40 PRS | -1.66E-09 | -3.60E-09 | 2.71E-10 | 9.86E-10 | -1.686 | -0.015 | 4.59E-02 | 12736 |
| Coffee/TGV0.45 PRS | -1.87E-09 | -3.88E-09 | 1.37E-10 | 1.03E-09 | -1.827 | -0.016 | 3.39E-02 | 12736 |
| Coffee/TGV0.50 PRS | -1.85E-09 | -3.93E-09 | 2.32E-10 | 1.06E-09 | -1.742 | -0.015 | 4.08E-02 | 12736 |

**E. Association between PRS for TGV and cereal intake.**

| PRS | beta | 95%CI_l | 95%CI_h | se | t | r | p_one-tailed | df.e |
|---|---|---|---|---|---|---|---|---|
| TGV PRS | 4.56E-02 | -6.75E-02 | 1.59E-01 | 5.77E-02 | 0.790 | 0.007 | 2.15E-01 | 12739 |
| TGV/Cereal0.05 PRS | 1.95E-02 | -9.59E-02 | 1.35E-01 | 5.89E-02 | 0.331 | 0.003 | 3.70E-01 | 12739 |
| TGV/Cereal0.10 PRS | 3.24E-04 | -1.18E-01 | 1.19E-01 | 6.03E-02 | 0.005 | 0.000 | 4.98E-01 | 12739 |
| TGV/Cereal0.15 PRS | 1.88E-02 | -1.02E-01 | 1.40E-01 | 6.18E-02 | 0.304 | 0.003 | 3.81E-01 | 12739 |
| TGV/Cereal0.20 PRS | 4.10E-03 | -1.20E-01 | 1.28E-01 | 6.34E-02 | 0.065 | 0.001 | 4.74E-01 | 12739 |
| TGV/Cereal0.25 PRS | 2.32E-04 | -1.28E-01 | 1.29E-01 | 6.55E-02 | 0.004 | 0.000 | 4.99E-01 | 12739 |
| TGV/Cereal0.30 PRS | 1.24E-02 | -1.20E-01 | 1.45E-01 | 6.77E-02 | 0.183 | 0.002 | 4.27E-01 | 12739 |
| TGV/Cereal0.35 PRS | 9.69E-04 | -1.36E-01 | 1.38E-01 | 6.97E-02 | 0.014 | 0.000 | 4.94E-01 | 12739 |
| TGV/Cereal0.40 PRS | -7.60E-03 | -1.49E-01 | 1.34E-01 | 7.24E-02 | -0.105 | -0.001 | 4.58E-01 | 12739 |
| TGV/Cereal0.45 PRS | -6.75E-03 | -1.54E-01 | 1.40E-01 | 7.49E-02 | -0.090 | -0.001 | 4.64E-01 | 12739 |
| TGV/Cereal0.50 PRS | -1.12E-02 | -1.64E-01 | 1.42E-01 | 7.80E-02 | -0.143 | -0.001 | 4.43E-01 | 12739 |

**F. Association between PRS for TGV and coffee intake.**

| PRS | beta | 95%CI_l | 95%CI_h | se | t | r | p_one-tailed | df.e |
|---|---|---|---|---|---|---|---|---|
| TGV PRS | -5.98E-02 | -2.29E-01 | 1.09E-01 | 8.63E-02 | -0.693 | -0.006 | 2.44E-01 | 12739 |
| TGV/Coffee0.05 PRS | -3.35E-02 | -2.06E-01 | 1.39E-01 | 8.82E-02 | -0.380 | -0.003 | 3.52E-01 | 12739 |
| TGV/Coffee0.10 PRS | -5.62E-02 | -2.33E-01 | 1.20E-01 | 9.00E-02 | -0.625 | -0.006 | 2.66E-01 | 12739 |
| TGV/Coffee0.15 PRS | -5.01E-02 | -2.31E-01 | 1.31E-01 | 9.22E-02 | -0.544 | -0.005 | 2.93E-01 | 12739 |
| TGV/Coffee0.20 PRS | -5.19E-02 | -2.37E-01 | 1.33E-01 | 9.45E-02 | -0.549 | -0.005 | 2.92E-01 | 12739 |
| TGV/Coffee0.25 PRS | -4.28E-02 | -2.33E-01 | 1.48E-01 | 9.72E-02 | -0.440 | -0.004 | 3.30E-01 | 12739 |
| TGV/Coffee0.30 PRS | -3.75E-02 | -2.34E-01 | 1.59E-01 | 1.00E-01 | -0.374 | -0.003 | 3.54E-01 | 12739 |
| TGV/Coffee0.35 PRS | -6.02E-02 | -2.63E-01 | 1.43E-01 | 1.03E-01 | -0.582 | -0.005 | 2.80E-01 | 12739 |
| TGV/Coffee0.40 PRS | -5.62E-02 | -2.67E-01 | 1.54E-01 | 1.07E-01 | -0.524 | -0.005 | 3.00E-01 | 12739 |
| TGV/Coffee0.45 PRS | -6.96E-02 | -2.89E-01 | 1.50E-01 | 1.12E-01 | -0.623 | -0.006 | 2.67E-01 | 12739 |
| TGV/Coffee0.50 PRS | -3.06E-02 | -2.59E-01 | 1.98E-01 | 1.16E-01 | -0.263 | -0.002 | 3.96E-01 | 12739 |

**Table S23: GO terms for  genes via the clusterProfiler package.**

**A. GO terms for the overlap between the transcription-related genes and the coffee-intake-related genes.**

| ONTOLOGY | ID | Description | GeneRatio | BgRatio | pvalue | p.adjust | qvalue |
|---|---|---|---|---|---|---|---|
| BP | GO:0001580 | detection of chemical stimulus involved in sensory perception of bitter taste | 3/13 | 29/17913 | 1.08E-06 | 1.22E-04 | 8.73E-05 |
| BP | GO:0050913 | sensory perception of bitter taste | 3/13 | 34/17913 | 1.76E-06 | 1.22E-04 | 8.73E-05 |
| BP | GO:0050912 | detection of chemical stimulus involved in sensory perception of taste | 3/13 | 35/17913 | 1.93E-06 | 1.22E-04 | 8.73E-05 |
| BP | GO:0050909 | sensory perception of taste | 3/13 | 57/17913 | 8.54E-06 | 4.06E-04 | 2.90E-04 |
| MF | GO:0033038 | bitter taste receptor activity | 3/12 | 18/16969 | 2.19E-07 | 5.70E-06 | 3.46E-06 |
| MF | GO:0008527 | taste receptor activity | 3/12 | 24/16969 | 5.42E-07 | 7.05E-06 | 4.28E-06 |

**B. GO terms for the transcription-related specifc genes.**

| ONTOLOGY | ID | Description | GeneRatio | BgRatio | pvalue | p.adjust | qvalue |
|---|---|---|---|---|---|---|---|
| BP | GO:0009150 | purine ribonucleotide metabolic process | 77/1423 | 500/17913 | 1.27E-08 | 6.93E-05 | 6.49E-05 |
| BP | GO:0050808 | synapse organization | 62/1423 | 394/17913 | 1.54E-07 | 4.18E-04 | 3.92E-04 |
| BP | GO:0009199 | ribonucleoside triphosphate metabolic process | 52/1423 | 319/17913 | 5.08E-07 | 6.63E-04 | 6.20E-04 |
| BP | GO:0009141 | nucleoside triphosphate metabolic process | 54/1423 | 337/17913 | 5.39E-07 | 6.63E-04 | 6.20E-04 |
| BP | GO:0009144 | purine nucleoside triphosphate metabolic process | 52/1423 | 321/17913 | 6.19E-07 | 6.63E-04 | 6.20E-04 |
| BP | GO:0009205 | purine ribonucleoside triphosphate metabolic process | 51/1423 | 314/17913 | 7.30E-07 | 6.63E-04 | 6.20E-04 |
| BP | GO:0099132 | ATP hydrolysis coupled cation transmembrane transport | 16/1423 | 54/17913 | 2.68E-06 | 2.09E-03 | 1.95E-03 |
| BP | GO:0046034 | ATP metabolic process | 46/1423 | 287/17913 | 3.64E-06 | 2.48E-03 | 2.32E-03 |
| BP | GO:0009167 | purine ribonucleoside monophosphate metabolic process | 49/1423 | 317/17913 | 5.04E-06 | 3.00E-03 | 2.81E-03 |
| BP | GO:0009126 | purine nucleoside monophosphate metabolic process | 49/1423 | 318/17913 | 5.51E-06 | 3.00E-03 | 2.81E-03 |
| BP | GO:0009161 | ribonucleoside monophosphate metabolic process | 50/1423 | 331/17913 | 7.81E-06 | 3.87E-03 | 3.62E-03 |
| BP | GO:0050807 | regulation of synapse organization | 36/1423 | 218/17913 | 2.08E-05 | 9.45E-03 | 8.85E-03 |
| BP | GO:0099175 | regulation of postsynapse organization | 21/1423 | 98/17913 | 2.30E-05 | 9.62E-03 | 9.00E-03 |
| BP | GO:0099177 | regulation of trans-synaptic signaling | 55/1423 | 394/17913 | 2.90E-05 | 9.99E-03 | 9.35E-03 |

| | | | | | | | |
|---|---|---|---|---|---|---|---|
| BP | GO:0009152 | purine ribonucleotide biosynthetic process | 41/1423 | 266/17913 | 3.10E-05 | 9.99E-03 | 9.35E-03 |
| BP | GO:0050803 | regulation of synapse structure or activity | 36/1423 | 222/17913 | 3.11E-05 | 9.99E-03 | 9.35E-03 |
| BP | GO:0046390 | ribose phosphate biosynthetic process | 43/1423 | 284/17913 | 3.12E-05 | 9.99E-03 | 9.35E-03 |
| BP | GO:0009123 | nucleoside monophosphate metabolic process | 50/1423 | 349/17913 | 3.33E-05 | 1.01E-02 | 9.43E-03 |
| BP | GO:0009260 | ribonucleotide biosynthetic process | 42/1423 | 279/17913 | 4.40E-05 | 1.26E-02 | 1.18E-02 |
| BP | GO:0099173 | postsynapse organization | 28/1423 | 158/17913 | 4.66E-05 | 1.27E-02 | 1.19E-02 |
| BP | GO:0050804 | modulation of chemical synaptic transmission | 54/1423 | 393/17913 | 5.27E-05 | 1.34E-02 | 1.25E-02 |
| BP | GO:0045333 | cellular respiration | 30/1423 | 176/17913 | 5.41E-05 | 1.34E-02 | 1.25E-02 |
| BP | GO:0051963 | regulation of synapse assembly | 21/1423 | 105/17913 | 6.75E-05 | 1.60E-02 | 1.49E-02 |
| BP | GO:0010506 | regulation of autophagy | 45/1423 | 314/17913 | 8.02E-05 | 1.82E-02 | 1.70E-02 |
| BP | GO:0007416 | synapse assembly | 29/1423 | 172/17913 | 8.74E-05 | 1.85E-02 | 1.73E-02 |
| BP | GO:0006090 | pyruvate metabolic process | 26/1423 | 147/17913 | 8.85E-05 | 1.85E-02 | 1.73E-02 |
| BP | GO:0060271 | cilium assembly | 47/1423 | 335/17913 | 9.50E-05 | 1.92E-02 | 1.79E-02 |
| BP | GO:0006164 | purine nucleotide biosynthetic process | 41/1423 | 280/17913 | 1.01E-04 | 1.97E-02 | 1.84E-02 |
| BP | GO:0072522 | purine-containing compound biosynthetic process | 42/1423 | 291/17913 | 1.16E-04 | 2.19E-02 | 2.05E-02 |
| BP | GO:0070646 | protein modification by small protein removal | 40/1423 | 273/17913 | 1.21E-04 | 2.19E-02 | 2.05E-02 |
| BP | GO:0048489 | synaptic vesicle transport | 23/1423 | 126/17913 | 1.36E-04 | 2.24E-02 | 2.10E-02 |
| BP | GO:0097480 | establishment of synaptic vesicle localization | 23/1423 | 126/17913 | 1.36E-04 | 2.24E-02 | 2.10E-02 |
| BP | GO:0007005 | mitochondrion organization | 60/1423 | 466/17913 | 1.36E-04 | 2.24E-02 | 2.10E-02 |
| BP | GO:0099150 | regulation of postsynaptic specialization assembly | 6/1423 | 12/17913 | 1.51E-04 | 2.30E-02 | 2.15E-02 |
| BP | GO:0150052 | regulation of postsynapse assembly | 6/1423 | 12/17913 | 1.51E-04 | 2.30E-02 | 2.15E-02 |
| BP | GO:0010975 | regulation of neuron projection development | 59/1423 | 458/17913 | 1.52E-04 | 2.30E-02 | 2.15E-02 |
| BP | GO:0044782 | cilium organization | 47/1423 | 346/17913 | 2.06E-04 | 3.03E-02 | 2.83E-02 |
| BP | GO:0099003 | vesicle-mediated transport in synapse | 28/1423 | 174/17913 | 2.58E-04 | 3.46E-02 | 3.24E-02 |
| BP | GO:0070296 | sarcoplasmic reticulum calcium ion transport | 11/1423 | 41/17913 | 2.60E-04 | 3.46E-02 | 3.24E-02 |
| BP | GO:0031123 | RNA 3'-end processing | 20/1423 | 107/17913 | 2.61E-04 | 3.46E-02 | 3.24E-02 |
| BP | GO:0051656 | establishment of organelle localization | 57/1423 | 448/17913 | 2.69E-04 | 3.46E-02 | 3.24E-02 |
| BP | GO:0016241 | regulation of macroautophagy | 27/1423 | 166/17913 | 2.76E-04 | 3.46E-02 | 3.24E-02 |
| BP | GO:0014808 | release of sequestered calcium ion into cytosol by sarcoplasmic reticulum | 10/1423 | 35/17913 | 2.82E-04 | 3.46E-02 | 3.24E-02 |

| | | | | | | | |
|---|---|---|---|---|---|---|---|
| BP | GO:0070932 | histone H3 deacetylation | 7/1423 | 18/17913 | 2.86E-04 | 3.46E-02 | 3.24E-02 |
| BP | GO:0098698 | postsynaptic specialization assembly | 7/1423 | 18/17913 | 2.86E-04 | 3.46E-02 | 3.24E-02 |
| BP | GO:0071426 | ribonucleoprotein complex export from nucleus | 21/1423 | 116/17913 | 2.93E-04 | 3.46E-02 | 3.24E-02 |
| BP | GO:0022406 | membrane docking | 25/1423 | 150/17913 | 3.13E-04 | 3.46E-02 | 3.24E-02 |
| BP | GO:0051168 | nuclear export | 28/1423 | 176/17913 | 3.13E-04 | 3.46E-02 | 3.24E-02 |
| BP | GO:0051169 | nuclear transport | 42/1423 | 305/17913 | 3.27E-04 | 3.46E-02 | 3.24E-02 |
| BP | GO:0051208 | sequestering of calcium ion | 21/1423 | 117/17913 | 3.31E-04 | 3.46E-02 | 3.24E-02 |
| BP | GO:0071166 | ribonucleoprotein complex localization | 21/1423 | 117/17913 | 3.31E-04 | 3.46E-02 | 3.24E-02 |
| BP | GO:0034248 | regulation of cellular amide metabolic process | 57/1423 | 452/17913 | 3.38E-04 | 3.46E-02 | 3.24E-02 |
| BP | GO:0097553 | calcium ion transmembrane import into cytosol | 22/1423 | 126/17913 | 3.61E-04 | 3.46E-02 | 3.24E-02 |
| BP | GO:0015682 | ferric iron transport | 10/1423 | 36/17913 | 3.62E-04 | 3.46E-02 | 3.24E-02 |
| BP | GO:0033572 | transferrin transport | 10/1423 | 36/17913 | 3.62E-04 | 3.46E-02 | 3.24E-02 |
| BP | GO:0072512 | trivalent inorganic cation transport | 10/1423 | 36/17913 | 3.62E-04 | 3.46E-02 | 3.24E-02 |
| BP | GO:1903514 | release of sequestered calcium ion into cytosol by endoplasmic reticulum | 10/1423 | 36/17913 | 3.62E-04 | 3.46E-02 | 3.24E-02 |
| BP | GO:1901293 | nucleoside phosphate biosynthetic process | 48/1423 | 365/17913 | 3.78E-04 | 3.52E-02 | 3.30E-02 |
| BP | GO:0016579 | protein deubiquitination | 37/1423 | 260/17913 | 3.82E-04 | 3.52E-02 | 3.30E-02 |
| BP | GO:0097479 | synaptic vesicle localization | 23/1423 | 137/17913 | 4.80E-04 | 4.15E-02 | 3.88E-02 |
| BP | GO:0006611 | protein export from nucleus | 26/1423 | 163/17913 | 4.84E-04 | 4.15E-02 | 3.88E-02 |
| BP | GO:0051209 | release of sequestered calcium ion into cytosol | 20/1423 | 112/17913 | 4.86E-04 | 4.15E-02 | 3.88E-02 |
| BP | GO:0051283 | negative regulation of sequestering of calcium ion | 20/1423 | 112/17913 | 4.86E-04 | 4.15E-02 | 3.88E-02 |
| BP | GO:0015991 | ATP hydrolysis coupled proton transport | 8/1423 | 25/17913 | 4.93E-04 | 4.15E-02 | 3.88E-02 |
| BP | GO:0010880 | regulation of release of sequestered calcium ion into cytosol by sarcoplasmic reticulum | 9/1423 | 31/17913 | 4.97E-04 | 4.15E-02 | 3.88E-02 |
| BP | GO:0051028 | mRNA transport | 24/1423 | 146/17913 | 5.03E-04 | 4.15E-02 | 3.88E-02 |
| BP | GO:0006913 | nucleocytoplasmic transport | 41/1423 | 302/17913 | 5.14E-04 | 4.17E-02 | 3.91E-02 |
| BP | GO:0009201 | ribonucleoside triphosphate biosynthetic process | 26/1423 | 164/17913 | 5.33E-04 | 4.27E-02 | 3.99E-02 |
| BP | GO:0055117 | regulation of cardiac muscle contraction | 16/1423 | 81/17913 | 5.54E-04 | 4.29E-02 | 4.01E-02 |
| BP | GO:0021548 | pons development | 5/1423 | 10/17913 | 5.64E-04 | 4.29E-02 | 4.01E-02 |
| BP | GO:0060456 | positive regulation of digestive system process | 5/1423 | 10/17913 | 5.64E-04 | 4.29E-02 | 4.01E-02 |

| | | | | | | | |
|---|---|---|---|---|---|---|---|
| BP | GO:0031124 | mRNA 3'-end processing | 17/1423 | 89/17913 | 5.67E-04 | 4.29E-02 | 4.01E-02 |
| BP | GO:0006405 | RNA export from nucleus | 21/1423 | 122/17913 | 5.91E-04 | 4.41E-02 | 4.12E-02 |
| BP | GO:0031503 | protein-containing complex localization | 34/1423 | 238/17913 | 6.01E-04 | 4.41E-02 | 4.12E-02 |
| BP | GO:1903902 | positive regulation of viral life cycle | 13/1423 | 59/17913 | 6.07E-04 | 4.41E-02 | 4.12E-02 |
| BP | GO:0051282 | regulation of sequestering of calcium ion | 20/1423 | 114/17913 | 6.15E-04 | 4.41E-02 | 4.13E-02 |
| BP | GO:0023061 | signal release | 54/1423 | 433/17913 | 6.25E-04 | 4.42E-02 | 4.14E-02 |
| BP | GO:0009142 | nucleoside triphosphate biosynthetic process | 27/1423 | 175/17913 | 6.50E-04 | 4.48E-02 | 4.19E-02 |
| BP | GO:1901292 | nucleoside phosphate catabolic process | 27/1423 | 175/17913 | 6.50E-04 | 4.48E-02 | 4.19E-02 |
| BP | GO:0015988 | energy coupled proton transmembrane transport, against electrochemical gradient | 8/1423 | 26/17913 | 6.63E-04 | 4.51E-02 | 4.22E-02 |
| BP | GO:0050657 | nucleic acid transport | 27/1423 | 176/17913 | 7.10E-04 | 4.63E-02 | 4.34E-02 |
| BP | GO:0050658 | RNA transport | 27/1423 | 176/17913 | 7.10E-04 | 4.63E-02 | 4.34E-02 |
| BP | GO:0006406 | mRNA export from nucleus | 19/1423 | 107/17913 | 7.15E-04 | 4.63E-02 | 4.34E-02 |
| BP | GO:0071427 | mRNA-containing ribonucleoprotein complex export from nucleus | 19/1423 | 107/17913 | 7.15E-04 | 4.63E-02 | 4.34E-02 |
| BP | GO:0009206 | purine ribonucleoside triphosphate biosynthetic process | 25/1423 | 159/17913 | 7.64E-04 | 4.85E-02 | 4.54E-02 |
| BP | GO:0009135 | purine nucleoside diphosphate metabolic process | 22/1423 | 133/17913 | 7.75E-04 | 4.85E-02 | 4.54E-02 |
| BP | GO:0009179 | purine ribonucleoside diphosphate metabolic process | 22/1423 | 133/17913 | 7.75E-04 | 4.85E-02 | 4.54E-02 |
| CC | GO:0033267 | axon part | 54/1472 | 354/18678 | 2.03E-06 | 1.31E-03 | 1.08E-03 |
| CC | GO:0098978 | glutamatergic synapse | 51/1472 | 352/18678 | 1.67E-05 | 4.36E-03 | 3.59E-03 |
| CC | GO:0097060 | synaptic membrane | 55/1472 | 395/18678 | 2.52E-05 | 4.36E-03 | 3.59E-03 |
| CC | GO:0043025 | neuronal cell body | 61/1472 | 454/18678 | 2.79E-05 | 4.36E-03 | 3.59E-03 |
| CC | GO:0098793 | presynapse | 59/1472 | 438/18678 | 3.49E-05 | 4.36E-03 | 3.59E-03 |
| CC | GO:0150034 | distal axon | 40/1472 | 262/18678 | 4.05E-05 | 4.36E-03 | 3.59E-03 |
| CC | GO:0045211 | postsynaptic membrane | 43/1472 | 299/18678 | 8.88E-05 | 8.19E-03 | 6.75E-03 |
| CC | GO:0016469 | proton-transporting two-sector ATPase complex | 12/1472 | 44/18678 | 1.07E-04 | 8.68E-03 | 7.16E-03 |
| CC | GO:0043204 | perikaryon | 23/1472 | 127/18678 | 1.37E-04 | 9.71E-03 | 8.01E-03 |
| CC | GO:0044306 | neuron projection terminus | 22/1472 | 120/18678 | 1.57E-04 | 9.71E-03 | 8.01E-03 |
| CC | GO:0098798 | mitochondrial protein complex | 35/1472 | 233/18678 | 1.65E-04 | 9.71E-03 | 8.01E-03 |
| CC | GO:0048786 | presynaptic active zone | 14/1472 | 60/18678 | 1.85E-04 | 9.94E-03 | 8.20E-03 |

| | | | | | | | |
|---|---|---|---|---|---|---|---|
| CC | GO:0036464 | cytoplasmic ribonucleoprotein granule | 28/1472 | 176/18678 | 2.76E-04 | 1.37E-02 | 1.13E-02 |
| CC | GO:0043679 | axon terminus | 19/1472 | 101/18678 | 3.06E-04 | 1.38E-02 | 1.14E-02 |
| CC | GO:0034399 | nuclear periphery | 22/1472 | 126/18678 | 3.24E-04 | 1.38E-02 | 1.14E-02 |
| CC | GO:0000922 | spindle pole | 25/1472 | 152/18678 | 3.42E-04 | 1.38E-02 | 1.14E-02 |
| CC | GO:0035770 | ribonucleoprotein granule | 29/1472 | 188/18678 | 3.69E-04 | 1.40E-02 | 1.16E-02 |
| CC | GO:0016363 | nuclear matrix | 19/1472 | 106/18678 | 5.76E-04 | 2.07E-02 | 1.71E-02 |
| CC | GO:0098800 | inner mitochondrial membrane protein complex | 20/1472 | 115/18678 | 6.26E-04 | 2.13E-02 | 1.75E-02 |
| CC | GO:0005681 | spliceosomal complex | 25/1472 | 160/18678 | 7.50E-04 | 2.42E-02 | 2.00E-02 |
| CC | GO:0010494 | cytoplasmic stress granule | 12/1472 | 55/18678 | 9.96E-04 | 3.02E-02 | 2.49E-02 |
| CC | GO:0098984 | neuron to neuron synapse | 41/1472 | 315/18678 | 1.03E-03 | 3.02E-02 | 2.49E-02 |
| CC | GO:0099240 | intrinsic component of synaptic membrane | 25/1472 | 164/18678 | 1.08E-03 | 3.03E-02 | 2.50E-02 |
| CC | GO:0001725 | stress fiber | 12/1472 | 56/18678 | 1.18E-03 | 3.05E-02 | 2.51E-02 |
| CC | GO:0097517 | contractile actin filament bundle | 12/1472 | 56/18678 | 1.18E-03 | 3.05E-02 | 2.51E-02 |
| CC | GO:0070469 | respiratory chain | 16/1472 | 88/18678 | 1.30E-03 | 3.21E-02 | 2.65E-02 |
| CC | GO:0005747 | mitochondrial respiratory chain complex I | 11/1472 | 50/18678 | 1.49E-03 | 3.21E-02 | 2.65E-02 |
| CC | GO:0030964 | NADH dehydrogenase complex | 11/1472 | 50/18678 | 1.49E-03 | 3.21E-02 | 2.65E-02 |
| CC | GO:0045271 | respiratory chain complex I | 11/1472 | 50/18678 | 1.49E-03 | 3.21E-02 | 2.65E-02 |
| CC | GO:0060198 | clathrin-sculpted vesicle | 5/1472 | 12/18678 | 1.49E-03 | 3.21E-02 | 2.65E-02 |
| CC | GO:0042734 | presynaptic membrane | 22/1472 | 141/18678 | 1.54E-03 | 3.21E-02 | 2.65E-02 |
| CC | GO:0043197 | dendritic spine | 22/1472 | 142/18678 | 1.69E-03 | 3.41E-02 | 2.81E-02 |
| CC | GO:0032432 | actin filament bundle | 12/1472 | 59/18678 | 1.90E-03 | 3.62E-02 | 2.98E-02 |
| CC | GO:0005759 | mitochondrial matrix | 53/1472 | 448/18678 | 1.93E-03 | 3.62E-02 | 2.98E-02 |
| CC | GO:0042641 | actomyosin | 13/1472 | 67/18678 | 1.96E-03 | 3.62E-02 | 2.98E-02 |
| CC | GO:0044309 | neuron spine | 22/1472 | 145/18678 | 2.22E-03 | 3.91E-02 | 3.23E-02 |
| CC | GO:0005635 | nuclear envelope | 50/1472 | 420/18678 | 2.24E-03 | 3.91E-02 | 3.23E-02 |
| CC | GO:0098936 | intrinsic component of postsynaptic membrane | 19/1472 | 120/18678 | 2.63E-03 | 4.37E-02 | 3.60E-02 |
| CC | GO:0031965 | nuclear membrane | 36/1472 | 281/18678 | 2.64E-03 | 4.37E-02 | 3.60E-02 |
| CC | GO:0005743 | mitochondrial inner membrane | 52/1472 | 445/18678 | 2.75E-03 | 4.37E-02 | 3.60E-02 |
| CC | GO:0099634 | postsynaptic specialization membrane | 15/1472 | 86/18678 | 2.77E-03 | 4.37E-02 | 3.60E-02 |
| CC | GO:0032279 | asymmetric synapse | 37/1472 | 294/18678 | 3.15E-03 | 4.64E-02 | 3.83E-02 |

| | | | | | | | |
|---|---|---|---|---|---|---|---|
| CC | GO:0016607 | nuclear speck | 46/1472 | 386/18678 | 3.21E-03 | 4.64E-02 | 3.83E-02 |
| CC | GO:0005746 | mitochondrial respiratory chain | 14/1472 | 79/18678 | 3.25E-03 | 4.64E-02 | 3.83E-02 |
| CC | GO:0000159 | protein phosphatase type 2A complex | 5/1472 | 14/18678 | 3.31E-03 | 4.64E-02 | 3.83E-02 |
| CC | GO:0033178 | proton-transporting two-sector ATPase complex, catalytic domain | 5/1472 | 14/18678 | 3.31E-03 | 4.64E-02 | 3.83E-02 |
| CC | GO:0099572 | postsynaptic specialization | 39/1472 | 316/18678 | 3.49E-03 | 4.76E-02 | 3.92E-02 |
| CC | GO:0005874 | microtubule | 43/1472 | 357/18678 | 3.53E-03 | 4.76E-02 | 3.92E-02 |

**C. GO terms for the coffee-intake-related specifc genes.**

| ONTOLOGY | ID | Description | GeneRatio | BgRatio | pvalue | p.adjust | qvalue |
|---|---|---|---|---|---|---|---|
| BP | GO:0071248 | cellular response to metal ion | 9/161 | 167/17913 | 2.04E-05 | 2.70E-02 | 2.61E-02 |
| BP | GO:0071294 | cellular response to zinc ion | 4/161 | 19/17913 | 2.19E-05 | 2.70E-02 | 2.61E-02 |
| BP | GO:0071241 | cellular response to inorganic substance | 9/161 | 194/17913 | 6.60E-05 | 4.23E-02 | 4.08E-02 |
| BP | GO:0006706 | steroid catabolic process | 4/161 | 25/17913 | 6.86E-05 | 4.23E-02 | 4.08E-02 |
| MF | GO:0033038 | bitter taste receptor activity | 4/161 | 18/16969 | 2.15E-05 | 7.41E-03 | 7.28E-03 |
| MF | GO:0008527 | taste receptor activity | 4/161 | 24/16969 | 7.15E-05 | 1.23E-02 | 1.21E-02 |
